# Effects of ketamine and esketamine on death, suicidal behaviour, and suicidal ideation in psychiatric disorders: A systematic review and meta-analysis

**DOI:** 10.1101/2025.08.19.25333796

**Authors:** Martin Plöderl, Ruth Cooper, Tom Walker, Vibha Shah, Mark Horowitz, Constantin Volkmann, Joanna Moncrieff

**Affiliations:** Center for Inpatient Psychotherapy and Crisis Intervention, University Clinic for Psychiatry/Psychotherapy/Psychosomatics, Paracelsus Medical University, 5020 Salzburg, Austria.; Department of Health Service & Population Research, David Goldberg Centre, 18 De Crespigny Park, Institute of Psychiatry, Psychology and Neuroscience, King’s College London, London, UK, SE5 8AF.; West London NHS Foundation Trust, UB2 4SA London, UK.; North East London Foundation Trust, Goodmayes Hospital, 157 Barley Lane, Goodmayes, Ilford IG3 8XJ, UK.; Research and Development Department, North East London Foundation Trust, Goodmayes Hospital, Ilford IG3 8XJ, UK.; Department of Psychiatry, Charité-Universitätsmedizin Berlin, corporate member of Freie Universität Berlin and Humboldt-Universität zu Berlin, 10117 Berlin, Germany. 8.; Division of Psychiatry, University College London, London WC1E 6BT, and North East London Foundation Trust, Goodmayes Hospital, Essex, IG3 8XJ.

## Abstract

**Obective:** Ketamine and esketamine have been claimed to possess anti-suicidal effects and potentially to transform suicide prevention. This study provides an updated overview of evidence from clinical trials to establish whether ketamine or esketamine reduce death, suicides, suicide attempts or suicidal ideation, compared to active or inert placebo among people with psychiatric disorders.

**Design:** Systematic review and meta-analysis.

**Data sources:** We searched EMBASE, PubMed and PsycINFO from inception until 02.12.2025.

**Eligibility criteria for selecting studies:** Eligible for inclusion were randomised controlled trials which compared the effect of ketamine or esketamine with active or inert placebo for the treatment of people with psychiatric disorders. We included trials with concomitant treatments and excluded those where ketamine/esketamine were used as anaesthics.

**Data synthesis and study quality:** Data were synthesised with meta-analysis, including methods for double-zero events. Risk of bias was assessed using the Cochrane Risk of Bias Tool and quality of the evidence was evaluated using GRADE guidelines.

**Results:** We included 73 trials with a total of 5671 participants. The majority of trials (56) examined ketamine, 16 esketamine, and 1 arketamine. Twelve of the ketamine trials were cross-over trials and the rest were parallel group trials. A single dose was used in 35 trials. Median length of treatment and follow-up was 45 days. Rates of suicidal behaviour were 1.63% for ketamine/esketamine and 1.72% for placebo, with the 95% credible interval including the null-effect, OR = 0.98 [0.58 to 1.60] (Bayesian Analysis with weak priors). There were 42 (1.43%) suicide attempts, 6 (0.20%) suicides and 9 (0.31%) deaths with ketamine/esketamine compared to 41 (1.64%), 2 (0.08%) and 5 (0.20%) on placebo. Ketamine/esketamine significantly reduced suicide ideation up to 4 weeks (standardized mean differences [SMD] at 12h to 24h of -0.31 [-0.45 to -0.18], I^2^ = 26%), but effects were small after 24 hours and dropped to near zero after 4 weeks. Subgroup analysis for suicide ideation revealed that effects of esketamine were close to zero after 24 hours whereas effects for ketamine were small to medium for the first 4 weeks. Effects tended to be larger in trials involving suicidal patients. Repeated dosings were not superior to single doses. Quality assessment revealed unreliable blinding, selective reporting and - especially for suicidal behaviour - imprecision, leading to low or very low certainty ratings for suicidal behaviour and low or moderate certainty ratings for suicide ideation.

**Conclusions:** There is insufficient evidence for a preventive effect of ketamine/esketamine for suicidal behaviour. The observed immediate but short-term effect on suicide ideation may be overestimated due to unblinding bias. Our review is the most comprehensive on suicidality to date, however, more evidence is needed to draw conclusions on suicidal behaviour.

**Other:** No specific funding was involved. The protocol was registered with PROSPERO (<u>CRD42023364156</u>).

**KEY MESSAGES:** *What is already known on this topic:*

⍰ Ketamine and esketamine have been considered to transform suicide prevention with evidence from multiple studies suggesting they may rapidly alleviate depression and suicide ideation.
⍰ There have been numerous calls for studies to investigate if these findings translate to suicidal behaviour. With the many new studies that have emerged only recently, such a review is overdue, together with an update on suicide ideation.

*What this study adds:*

⍰ Despite including studies up to December 2025, evidence on whether ketamine/esketamine can effectively reduce suicidal behaviour remains inconclusive.
⍰ Small to medium effect on suicide ideation within the first 4 weeks (with near zero effects thereafter) were observed for ketamine but for esketamine effects were close to zero already after 24 hours, and unblinding bias is likely involved.
⍰ Repeated doses did not show superiority over single administrations in reducing suicide ideation.

*How this study might affect research, practice, or policy:*

⍰ More studies are needed to judge if ketamine/esketamine can reduce suicidal behaviour, and how unblinding effects might explain the short-term effect on suicide ideation.
⍰ The practice of repeated dosings should be questioned and needs further investigation.

## INTRODUCTION

Suicide is a major public health concern across the world. Overall, 1 in 100 people die by suicide and among people aged 15-29 it is the fourth leading cause of death worldwide [1]. In the UK suicide is the most common cause of death among both men and women aged 20 to 34 [2]. Despite a worldwide decline in the rate of suicide since 2000, rates are increasing in the US and some European countries, including the UK [2,3]. A broad range of psychiatric disorders is associated with an increase of suicides and suicide attempts [4].

Ketamine and its S-enantiomer, esketamine, are increasingly used for the treatment of treatment-resistant depression and also investigated for a broader range of mental health problems such as bipolar depression, substance use disorders and anxiety disorders. Furthermore, they have been suggested to have the potential to offer rapid relief from suicidal thoughts and to help prevent suicide [5,6] and Esketamine was licensed in the US for people with treatment resistant depression and major depressive disorder with suicidal thoughts or actions [7]. Previous summaries of the literature have claimed promising results for both agents [8–10], suggesting that esketamine is “the only FDA-approved antidepressant that works in hours not weeks – thus potentially transforming treatment of suicidal patients” [8]. There is also some evidence that ketamine reduces suicide ideation independent of depression symptoms [11]. On the other hand, post marketing data suggests that esketamine may be associated with an increase in suicidal ideation and suicides [12] and chronic, recreational ketamine misuse is associated with depression [13–15]. While previous systematic and non-systematic reviews have generally suggested that ketamine and esketamine reduce suicidal ideation and behaviour, at least short-term [16–19], they also highlight methodological limitations of the research and a lack of data on medium or long-term effects [17]. However, these limitations have not been explored in detail, and no existing reviews have considered the outcome of deaths by suicide or suicide attempts, even though reducing suicide is the ultimate aim of this intervention. Moreover, there is the repeated request for using harder outcomes in psychiatric research, such as suicide attempts or suicide, because softer outcomes (reduction in symptom scores) are more prone to bias [20]. Reviews on suicidal behaviour are lacking partly due to the difficulty of conducting meta-analysis of rare events, but statistical methods to manage this situation have been developed recently [21,22]. Furthermore, existing reviews, even those of otherwise good quality, had significant limitations, for example, mixing outcomes at different time-points, or errors in data extraction [18,23,24]. Finally, many studies appeared recently and an update is required.

In the current systematic review and meta-analysis we aimed to answer the following question: does ketamine or esketamine reduce suicidal behaviour (suicides, suicide attempts), deaths of any cause (as additional outcome because suicidal intent is sometimes unknown), and suicidal ideation for people with primary psychiatric disorders? We examined short-term and longer-term outcomes. We also explored methodological limitations of the existing studies.

## METHODS

This systematic review and meta-analysis was conducted following guidance from the Preferred Reporting Items for Systematic Reviews and Meta-Analyses (PRISMA) guidelines [25]; see Supplement for the PRISMA Checklist.

### Eligibility criteria

All randomised controlled trials (RCTs), comparing ketamine or esketamine with inert placebo or active placebo in individuals with a primary diagnosis of any major psychiatric disorder, were eligible for inclusion in this review. Data from the first period of cross-over trials were also included (to protect against the carry-over effect). Trials comparing ketamine or esketamine in combination with an antidepressant, psychotherapy or other intervention were included as long as other interventions were equally available to those in the control group. Studies were excluded if the drugs under investigation were intended as an analgesic and/or anaesthetic agent. In addition, although not specified in the protocol due to being unforeseen at the time, studies were excluded if data on suicidality were not available despite contacting the authors.

### Information sources and search strategy

We searched EMBASE, PubMed and PsycINFO from inception until 01.10.2022 using the strategy outlined in supplementary Table S1. The search was updated on 09.01.2025 and again on 02.12.2025. We had no language restrictions. We also searched citations of included studies and previous reviews.

### Selection process

Two reviewers (initial search and first update TW, MP; second update BAM, MP) independently screened titles and abstracts and subsequently full texts. Discrepancies were resolved by discussion (RC, JM, and MP).

### Data collection process

For the initial search and the first search update, authors extracted summary statistics from an assigned portion of selected studies using a standardised form and another author (MP) did an independent extraction. Data from the final search update was extracted by a single author (MP). Uncertainties and discrepancies were resolved by discussion. In cases of missing data or areas of uncertainty, the corresponding author of the paper was contacted with at least one reminder, and we looked up the data on trials registers using the study registration number. Summary statistics from Janssen’s esketamine trials were calculated by one author (MP) based on individual patient data accessed via the Yale Open University Data Access (YODA) project. This was validated by comparison with the available summary statistics in the trial publications.

### Data items and outcome measures

We extracted the following data: trial design, length of follow-up, participant number and characteristics, intervention, control, and Conflicts of Interest (COIs).

Outcomes consisted of death by any cause, suicides, suicide attempts and suicidal ideation. We extracted outcome data at seven time-points, similar to a previous review [18]: within 4 hours, >4 to ⍰12 hours, >12 to ⍰24 hours, >24 to ⍰72 hours, >72 hours to ⍰2 weeks, >2 to ⍰4 weeks, and >4 weeks. If there were more than one measure within a specific time frame, we used the one closest to the end of the time frame.

Suicidal behaviour was made up of a composite of suicides and suicide attempts. We checked the trial registers for suicidal behaviour and mortality, and we assumed that no such events occurred when the paper reported that “no serious adverse events occurred” or when tabulated adverse events did not include suicidal behaviour/mortality.

For suicidal ideation, we used results based on assessments with suicide-specific scales such as the Beck Scale for Suicidal Ideation (BSI) or the Columbia Suicide Severity Rating Scale (C-SSRS). If this was not available, we used the suicidal ideation item of a depression rating scale in this hierarchy: the authors designated primary outcome and if this was not specified the Montgomery-Åsberg Depression Rating Scale (MADRS), followed by the Beck Depression Inventory (BDI), Hamilton Depression Rating Scale (HDRS), and other scales such as the Quick Inventory of Depressive Symptomatology (QIDS).

When examining the trials, it became clear that there were distinct “treatment phases” and “follow-up” phases, although these were not defined consistently in the original studies. Results from follow-up periods were included as long as randomization was maintained, in order to include data from the same time point in studies with different treatment durations. Data from non-randomized open-label treatment phases were excluded.

### Quality assessment

The risk of bias in the included studies was assessed using the Cochrane Risk of Bias tool [26]. This measure rates studies as being at high, low or unclear risk of bias according to various domains. Criteria for the rating of each domain were established by consensus within the research team in accordance with the Cochrane guidelines. Quality assessments for the initial search were carried out by MP, MH, JM, SA, RC, TW, and VS and independently rated by JM or MP. For the two search updates, ratings were done by JM and MP. Discrepancies were resolved by discussion. Quality of the evidence to judge if the confidence that the estimation of the effect is correct was evaluated (JM, MP, RC) using GRADE (Grading of Recommendations, Assessment Development and Evaluation) [27,28] and GRADEPro software.

### Synthesis methods

We reported results for suicide attempts, suicides, and mortality according to whether they occurred in the treatment phase, the follow-up phase, and both phases combined, since further information about timing was generally not available. We included studies without any events to avoid biased results, following recent recommendations and different random effects meta-analytic approaches suggested by Xu et al. (2021). These authors also recommended using more than one method to confirm that results are not dependent on a specific method. Therefore, as outlined in our protocol, we used a random effects two-stage exact method [22] and a one-stage Bayesian method as in a previous publication [30] which was based on a binomial-normal hierarchical model. We used weak priors based on empirically derived suggestions, that is, a normal distribution with mean = 0 and variance = 2.82, with a limitation to delta=250, meaning that odds ratios larger than 250 or smaller than 1/250 are considered unlikely, and a half-normal prior for the heterogeneity tau=0.5 [31]. We also used a Bayesian model with a more informative prior (delta = 15, meaning that the odds ratios are in the range of 1/15 to 15). We also ran sensitivity analysis with very weak/informative priors for the treatment effect and for heterogeneity. In addition, we used one-stage frequentist beta-binomial and GLMM methods [32] and the following two stage models: Peto’s method, Mantel-Haenszel methods with continuity corrections, and two methods based on risk differences (Mantel-Haenszel and arcsine transformation methods). All other methods were based on odds-ratios as measures of effect-size. Not specified in the protocol but for ease of interpretation we prioritize the binomial-normal hierarchical Bayesian model with weak priors because this model has been found suitable [31] and considered other models as sensitivity analysis. We considered the results of different models to be consistent if they lead to the same GRADE ratings for imprecision and heterogeneity.

Heterogeneity was assessed with I^2^ and τ^2^ if the meta-analytic model allowed for this (with R’s default parameters). We combined drug-arms for studies with different doses as recommended [33].

For suicide ideation as outcome, we ran random effects meta-analysis using R’s “meta” package, with Hedges *g* as measure of effect size, I^2^ and τ^2^ as measures of heterogeneity (using Sidik & Jonkman’s method to estimate heterogeneity). Meta-analyses were based on drug-placebo differences in suicide ideation at specific time-points and not on differences in change from baseline, except for one study where only change-scores were available. Publication bias was explored with Egger’s regression test. For the few studies which only reported medians we used the procedure suggested by Wan et al. to calculate means and standard deviations [34]. For one study [35] no information about the standard deviations were available and we imputed these with those from other studies with the same assessment instruments, as recommended [33]. We combined results from multiple arms with different doses following the recommendation by Cochrane [33] or, if possible, by using individual patient data. We planned to conduct various pre-specified subgroup or subset analyses, where sufficient data permitted, namely: inclusion or exclusion of people at significant risk of suicide, different primary psychiatric disorders (including substance misuse), esketamine or ketamine, method of administration (e.g., intravenous, oral), sample size, inert or active placebo, different trial designs (RCT or single blinded trial or open label study) and clinician or self-report measures of suicidal ideation. We also conducted a subgroup analysis of single versus repeated doses which was not included in the protocol but was considered important given the increasing practice of repeated, ongoing dosing. We did not distinguish between the treatment-phase and follow-up phase in the analysis for suicide ideation because this was the only way that longer-term effects from single-administrations could be compared to those of repeated administrations.

We used R 4.3.0 for all analysis [36]. The protocol was developed and registered prior to data extraction (PROSPERO <u>CRD42023364156</u>).

### Patient and public involvement

No patient and public involvement happened at the stage of planning the analysis. However, one member of the review-team has experiences of mental health difficulties and treatments. During the data-analysis, a representative of Austria’s patient groups who is also a member of the expert panel in Austria’s national suicide prevention program SUPRA (Suicide Prevention Austria) was informed about the aims and methods of the review and asked to provide feedback after discussing the study with other representatives of patient groups. This did not result in practical suggestions about the study aims or methodology. Patient representatives felt it was important that the review fulfils quality criteria and will be publicly available. They expressed concerns about the validity of the assessment of suicidality and about potential addiction or dependence problems with longer-term use of ketamine or esketamine. We include these points in the discussion section and intend to publish this review open access.

## RESULTS

### Study selection and characteristics

The search identified 3118 titles via databases and additional 15 titles via reference checking, of which 264 studies were subjected to full-text screening (Figure 1). A total of 191 studies were excluded following full-text screening (Table S2). A total of 73 studies were identified that met eligibility criteria, including 56 involving ketamine, 16 esketamine, and 1 arketamine. Twelve of the ketamine trials were cross-over trials and the rest were parallel group trials. A single dose was used in 35 trials (33 ketamine trials). In repeated dose trials, doses were administered between weekly and two times daily and treatment lasted between three days and ten weeks. Follow-up lasted between 24 hours to 12 months (median 27 days, interquartile range 7 to 31 days). The trials involved 5671 participants in total. Further characteristics are detailed in Table S3.

**Figure 1.**
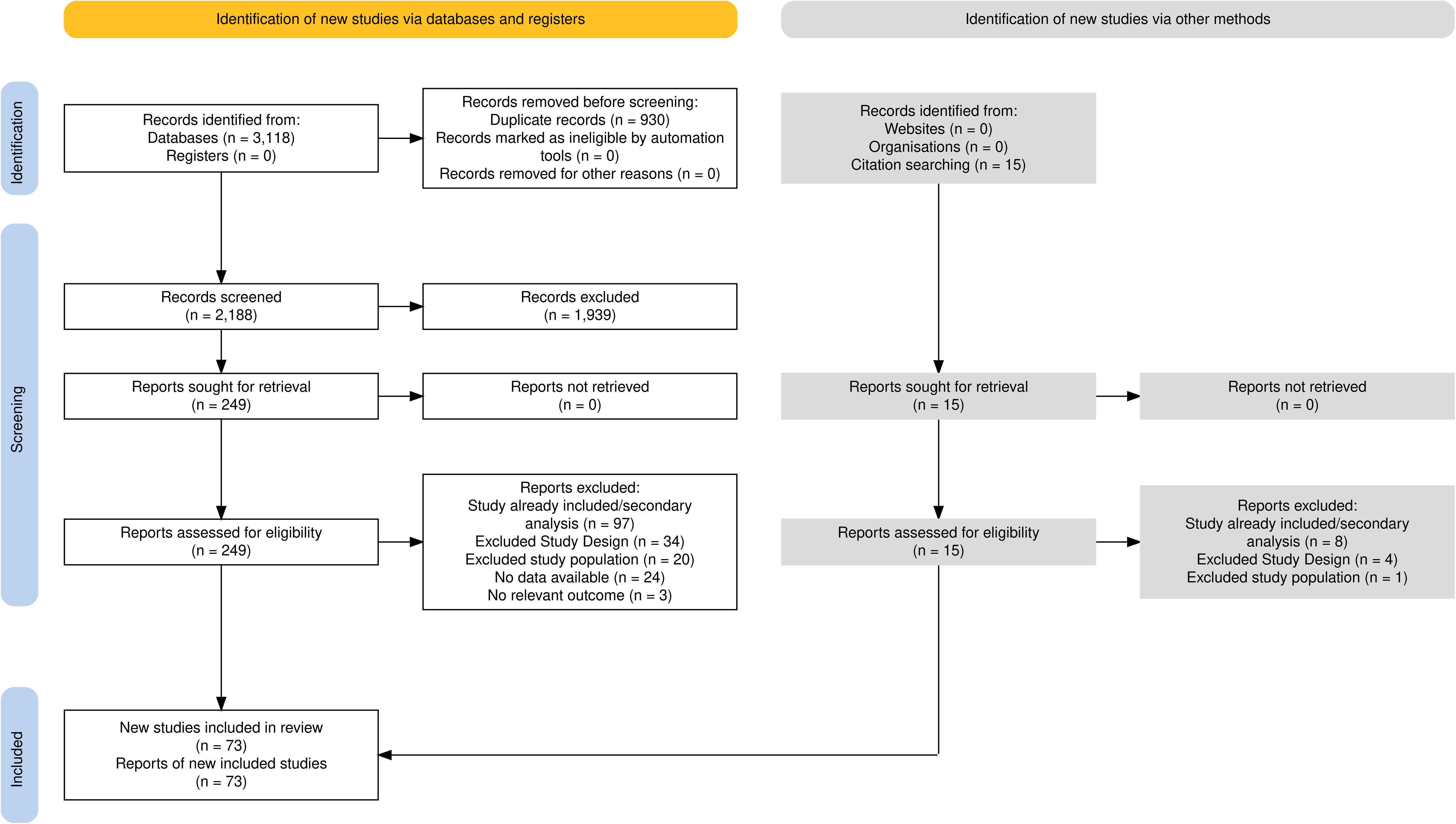
PRISMA flowchart (Created with PRISMA 2020 [62]).

### Risk of bias and GRADE ratings

Full risk of bias ratings are shown in supplementary Figure S2. These revealed that patients and study personnel were rarely likely to be adequately blinded. Only 18 studies tested the integrity of the double blind, of which 13 showed evidence of unblinding among participants and or assessors (Table S4). These included 9 of the 11 studies that employed an active placebo agent such as midazolam or remifentanil, with some of these finding rates of correct guesses by participants and assessors between 80 and 100% [37,38]. Only three out of these 18 studies were considered adequately blinded, two of which involved administering ketamine along with an anaesthetic [39,40]. Furthermore, we rated 21 (29%) of studies as high risk of bias due to selective reporting of outcomes, and one trial publication did not report suicidality as an outcome despite this being planned (Dwyer. et al. 2021). Overall 61 (84%) trials were rated as being at high risk of bias in some domain. Ratings of the certainty of evidence for estimations of effects for the most important outcomes are summarised in Table 1. For suicide ideation, certainty of the evidence ranged from low to moderate, mainly due to risk of bias due to functional unblinding, and due to imprecision and heterogeneity which varied for different time-points. For suicide attempts and suicides combined, certainty of the evidence was rated as very low due to risk of bias and imprecision, and low for suicide because of imprecision.

**Table 1:**
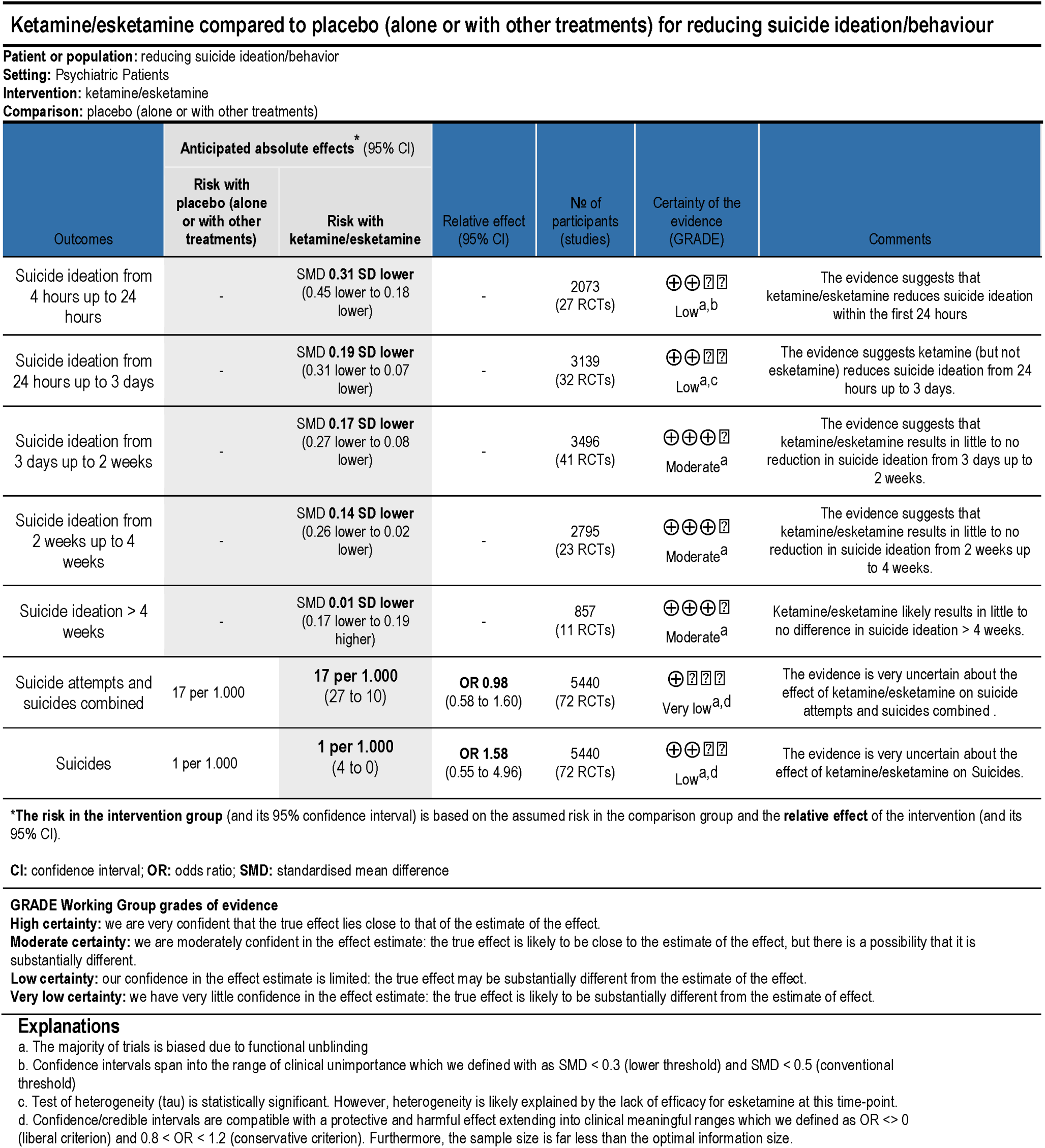
Summary of findings table with GRADE ratings.

### Suicidal Behaviour and Deaths

For suicides and suicide attempt combined, and for both the treatment and follow-up phase combined, there were 48 (1.63%) incidents in 2936 patients randomized to ketamine or esketamine compared to 43 (1.72%) incidents in 2504 patients randomized to placebo (Table S5). The drug-placebo difference included the null effect (OR = 1) and the credible interval was wide, OR = 0.98 [0.58 to 1.60] (Table 2, S6). Results for different priors and other meta-analytic methods lead to comparable findings (Tables 2, S9). In the treatment-period, there were 20 (0.68%) incidents with ketamine/esketamine and 15 (0.60%) with placebo, and for the follow-up period there were 28 (1.96%) and 28 (2.21%) incidents, respectively (Table S5). None of the meta-analytic results were statistically significant (Table 2, forest-plots with results for individual studies Figures S2-S95). For suicide attempts, there were 42 (1.43%) incidences with ketamine or esketamine, compared to 41 (1.64%) with placebo and again the difference overlapped with the null-effect, OR = 0.87 [0.50 to 1.45] (Table S10). For suicides, there were 6 (0.20%) incidences with ketamine or esketamine compared to 2 with placebo (0.08%), OR = 1.58 [0.55 to 4.96] (Table S13). The number of deaths of any cause in combined treatment and follow-up phases was 9 (0.31%) in ketamine or esketamine and 5 (0.20%) in the placebo groups and the credible interval in the meta-analysis overlapped with the null-effect, OR = 1.41 [0.57 to 3.65] (Table S16). Results were comparable with other meta-analytic models and for different treatment phases (Tables S10-S18). None of the drug-placebo differences for deaths, suicides, and suicide attempts were statistically significant. There was no indication of reporting bias in any of the analysis for suicidal behaviour and deaths.

**Table 2:** Meta-analysis of suicides and suicide attempts (Treatment + Follow-Up Phases combined)

| Analysis | OR [95% CI] | p | I<sup>2</sup> [95% CI] | τ<sup>2</sup> [95%CI] | number of studies |
| --- | --- | --- | --- | --- | --- |
| Bayesian - Weak prior (1/250 < OR < 250)* | 0.98 [0.58 - 1.60] |  |  | 1.31 [1.01 - 2.49] | 72 |
| Bayesian - Informative prior (1/15 < OR < 15) | 0.97 [0.58 - 1.58] |  |  | 1.31 [1.01 - 2.50] | 72 |
| Exact (Liu et al. 2012) | 1.06 [0.63 - 1.78] |  |  |  | 72 |
| Binomial-normal GLMM, fixed intercept | 1.01 [0.65, 1.57] |  |  | 0.00 |  |
| Binomial-normal GLMM, fixed intercept, alternative parameterization | 1.01 [0.64, 1.57] |  |  | 0.00 |  |
| Binomial-normal GLMM, random intercept | 1.00 [0.64, 1.55] |  |  | 0.00 |  |
| Binomial-normal GLMM, random intercept, alternative parameterization | 1.00 [0.67, 1.55] |  |  | 0.00 |  |
| Hypergeometric-normal GLMM (Nchg) | 1.01 [0.67, 1.53] |  |  | 0.00 |  |
| Conditional binomial GLMM (Condbin) | 1.01 [0.32, 1.53] |  |  | 0.00 |  |
| Beta-binomial model | 0.75 [0.65, 1.73] |  |  |  |  |
| Peto | 1.01 [0.65 - 1.57] | 0.96 | 25 [0 - 59] | 0.00 [0.00 - 1.95] | 16 |
| Mantel-Haenszel - with continuity correction | 0.92 [0.65 - 1.30] | 0.64 | 0 [0 - 28] | 0.00 [0.00 - 0.00] | 72 |
| Mantel-Haenszel - with continuity/treatment arm correction | 1.00 [0.71 - 1.42] | 0.98 | 0 [0 - 28] | 0.00 [0.00 - 0.00] | 72 |
| Mantel-Haenszel - risk difference | 0.00 [0.00 - 0.01] | 0.59 | 0 [0 - 28] | 0.00 [0.00 - 0.00] | 72 |
| Arcsine square root transformed risk difference | 0.01 [-0.02 - 0.03] | 0.63 | 0 [-] | 0.00 [-] | 72 |

Note: OR: odds-ratio, CI: confidence interval or credible interval for Bayesian analysis, p: p-value of odds-ratio, I<sup>2</sup> and τ<sup>2</sup>: measures of heterogeneity, GLMM: generalized linear mixed models.
All methods are appropriate except Peto's method, because there are both single-arm-zero-events studies and double-arm-zero-events studies and the total event count in each arm is non-zero [29].
\*Prioritized analysis, other methods should be considered as sensitivity analysis (see methods section).

### Suicide Ideation

Suicidal ideation was statistically significantly reduced among people allocated to ketamine/esketamine compared to placebo, with a small to medium effect size up to 24 hours after randomisation (SMD -0.31 [-0.45 to -0.18], p < 0.001 for > 12h up to 24h). At subsequent follow-ups differences were small (point sizes about SMD 0.20) and after 4 weeks follow-up they were close to zero and not statistically significant (Table 3, Figure 2). An end of treatment phase analysis (combining results from studies of all durations) showed a significant effect, SMD = -0.25 [-0.35 to - 0.15]. Heterogeneity was only statistically significant for results in the first 4 hours and for >24h up to 72h. There was no indication of publication bias in any of the analyses for suicidal ideation (Table 3, Funnel-plots Figures S103-109).

**Figure 2.**
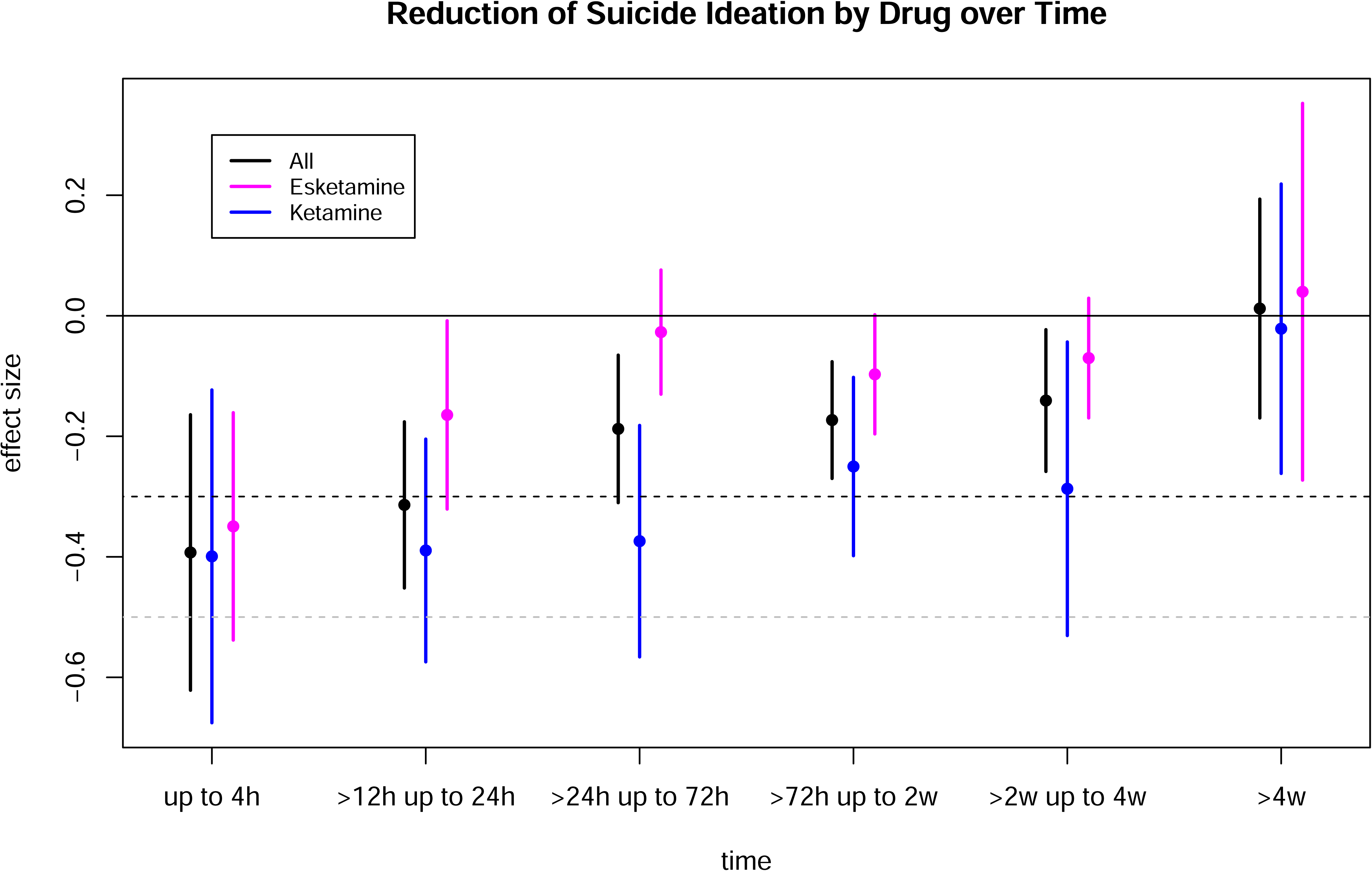
Meta-analytic results for effects of ketamine and esketamine on suicide ideation at different time-points. The y-axis refers to the standardized mean-difference. Negative values correspond to increased reduction in suicide ideation in the drug arms compared to the placebo arms. The horizontal dashed lines are two thresholds for clinical significance at SMD = -0.3 and SMD = -0.5.

**Table 3:** Meta-analysis of suicidal ideation.

| Result | up to 4h | >12h up to 24h | >24h up to 72h | >72h up to 2w | >2w up to 4w | >4w | End of Treatment |
| --- | --- | --- | --- | --- | --- | --- | --- |
| SMD [95%-CI] | -0.39 [-0.62 to -0.16] | -0.31 [-0.45 to -0.18] | -0.19 [-0.31 to -0.07] | -0.17 [-0.27 to -0.08] | -0.14 [-0.26 to -0.02] | 0.01 [-0.17 to 0.19] | -0.25 [-0.35 to -0.15] |
| p | < 0.001 | < 0.001 | < 0.01 | < 0.001 | 0.02 | 0.90 | < 0.001 |
| Heterogeneity | I<sup>2</sup>=58%, T<sup>2</sup>=0.23,<br>p<0.001 | I<sup>2</sup>=26%, T<sup>2</sup>=0.05,<br>p=0.10 | I<sup>2</sup>=41%, T<sup>2</sup>=0.06,<br>p<0.01 | I<sup>2</sup>=0%, T<sup>2</sup>=0.03,<br>p=0.74 | I<sup>2</sup>=12%, T<sup>2</sup>=0.03,<br>p=0.29 | I<sup>2</sup>=13%, T<sup>2</sup>=0.03,<br>p=0.32 | I<sup>2</sup>=25%, T<sup>2</sup>=0.05,<br>p=0.07 |
| Number of trials | 26 | 27 | 32 | 41 | 23 | 11 | 44 |
| Number of observations | 1509 | 2073 | 3139 | 3496 | 2795 | 857 | 3582 |
| Small study effect<br>p-Value of Egger's<br>regression test | 0.43 | 0.06 | 0.04 | 0.30 | 0.51 | 0.91 | < 0.01 |

Note: SMD: standardized mean difference, CI: confidence interval, I<sup>2</sup> and $\tau^2$ : measures of heterogeneity, p: p-value

### Subgroup analysis

Deaths, suicides and suicide attempts were too rare to do subgroup analyses. For suicidal ideation, subgroup analysis for different types of drugs showed that the effects of ketamine were consistently larger than esketamine up to 4 weeks, with some differences being statistically significant, too (Table 4). Esketamine’s efficacy was close to zero after 24 hours, whereas the efficacy of ketamine remained significant up to 4 weeks and end of treatment. The significant heterogeneity at 24h up to 72h for drugs combined disappeared within individual drugs (Ketamine, SMD -0.37 [-0.57 to -0.18], Esketamine SMD -0.03 [-0.13 - 0.08]). Efficacy was numerically larger in trials where suicidality was an inclusion criterion (vs. suicidality as exclusion criteria or unspecified) in the first two weeks and end of treatment with some differences also being statistically significant. Trials with active placebos (defined as an active drug) had larger effects than trials with inert placebos (including those with a bittering agent) with significant differences as several time-points. There were no significant differences between trials where suicide ideation was assessed by a clinician compared with by patients. There was insufficient data for comparing different diagnoses (most studies involved MDD), trial design, or low risk of bias (or post-hoc, exploratory subgroup analysis for age-groups, doses, etc.). In the explorative subgroup analysis, repeated administrations were not statistically significantly superior to a single dose at any time point (we ran analysis after 72h since repeated dosing usually starts after this time point). Forest-plots for subgroup analyses are in the online supplement (Figures S110-S149).

**Table 4:** Meta Analytic Results, Suicide Ideation - Subgroup Analyses.

| Subgroup analysis | up to 4h | >12h up to 24h | >24h up to 72h | >72h up to 2w | >2w up to 4w | >4w | End of Treatment |
| --- | --- | --- | --- | --- | --- | --- | --- |
| PRE-PLANNED |  |  |  |  |  |  |  |
| Drug |  |  |  |  |  |  |  |
| --- Esketamine | -0.35 [-0.54 to -0.16]* | -0.16 [-0.32 to -0.01]* | -0.03 [-0.13 to 0.08] | -0.10 [-0.20 to 0.00] | -0.07 [-0.17 to 0.03] | 0.04 [-0.27 to 0.35] | -0.07 [-0.17 to 0.03] |
| --- Ketamine | -0.40 [-0.68 to -0.12]* | -0.39 [-0.57 to -0.20]* | -0.37 [-0.57 to -0.18]* | -0.25 [-0.40 to -0.10]* | -0.29 [-0.53 to -0.04]* | -0.02 [-0.26 to 0.22] | -0.43 [-0.57 to -0.29]* |
| --- test of difference (p-value) | 0.10 | < 0.01 | < 0.001 | 0.02 | < 0.01 | 0.55 | < 0.001 |
| Suicidality excl/incl criterion |  |  |  |  |  |  |  |
| --- Exclusion criterion or unknown | -0.20 [-0.59 to 0.20] | -0.20 [-0.40 to 0.00] | -0.06 [-0.18 to 0.06] | -0.14 [-0.25 to -0.02]* | -0.12 [-0.28 to 0.03] | 0.13 [-0.13 to 0.38] | -0.16 [-0.28 to -0.04]* |
| --- Inclusion criterion | -0.48 [-0.76 to -0.21]* | -0.39 [-0.58 to -0.21]* | -0.38 [-0.60 to -0.15]* | -0.25 [-0.42 to -0.07]* | -0.17 [-0.36 to 0.01] | -0.09 [-0.32 to 0.13] | -0.38 [-0.56 to -0.21]* |
| --- test of difference (p-value) | 0.11 | 0.02 | < 0.001 | 0.08 | 0.24 | 0.12 | < 0.001 |
| Rating |  |  |  |  |  |  |  |
| --- Clinician | no study | -0.30 [-0.44 to -0.15]* | no study | no study | -0.15 [-0.28 to -0.03]* | -0.05 [-0.25 to 0.16] | no study |
| --- Patient | in one subgroup | -0.47 [-0.94 to -0.01]* | in one subgroup | in one subgroup | 0.05 [-0.45 to 0.56] | -0.01 [-0.41 to 0.40] | in one subgroup |
| --- test of difference (p-value) |  | 0.26 |  |  | 0.43 | 0.86 |  |
| Placebo |  |  |  |  |  |  |  |
| --- Active | -0.27 [-0.62 to 0.09] | -0.40 [-0.63 to -0.18]* | -0.36 [-0.61 to -0.10]* | -0.26 [-0.50 to -0.03]* | -0.16 [-0.47 to 0.16] | 0.02 [-0.36 to 0.40] | -0.46 [-0.65 to -0.27]* |
| --- Non-active | -0.48 [-0.77 to -0.19]* | -0.25 [-0.43 to -0.08]* | -0.13 [-0.27 to 0.00] | -0.15 [-0.24 to -0.05]* | -0.14 [-0.26 to -0.01]* | 0.01 [-0.19 to 0.21] | -0.16 [-0.27 to -0.05]* |
| --- test of difference (p-value) | 0.89 | 0.04 | < 0.01 | 0.14 | 0.67 | 0.93 | < 0.001 |
| EXPLORATIVE |  |  |  |  |  |  |  |
| Doses |  |  |  |  |  |  |  |
| ---Repeated |  |  |  | -0.14 [-0.25 to -0.04]* | -0.12 [-0.22 to -0.01]* | 0.01 [-0.20 to 0.22] |  |
| ---Single |  |  |  | -0.24 [-0.44 to -0.04]* | -0.17 [-0.57 to 0.22] | 0.02 [-0.40 to 0.44] |  |
| --- test of difference (p-value) |  |  |  | 0.12 | 0.16 | 0.96 |  |

Note: Results are standardized mean differences with their 95%-confidence intervals in brackets
\* $p < 0.05$

## DISCUSSION

The evidence that ketamine or esketamine prevent suicidal behaviour or mortality is inconclusive. Numerically, there were fewer suicide attempts and more suicides and deaths in the drug arms than in the placebo arms, but there is much uncertainty in the estimations of differences. Medium-sized protective effects on suicide ideation were only observed up to three days, and these were generally smaller for esketamine than ketamine. Repeated dosings were not superior to single doses.

In general, drawing conclusions is hampered by the lack of long-term data, insufficient statistical power, and selective reporting. Another problem is the acute psychoactive effects of ketamine/esketamine. These may influence suicidal ideation and mood in their own right, and may also undermine the double blind design. This was confirmed by the few trials which tested blinding and found that it was mostly not successful, even for active placebos [42,43]. In two trials in which ketamine was given during an anaesthetic procedure and blinding was successful, the drug-placebo differences were close to zero (see subgroup analysis in Supplement). In one of these, people who guessed they had had ketamine showed substantially lower symptoms at follow-up than people who guessed they had had placebo (p=0.001), despite there being no effect of the actual substance [40]. The assumption that functional unblinding leads to an overestimation of efficacy is also supported by a lack of drug-placebo differences for depressive symptoms in trials where functional unblinding is likely reduced because ketamine/esketamine was given in addition to anaesthics in electroconvulsive therapy [39,44–47], or as an anaesthic agent compared to another anaesthic [48,49]. In addition, selection biases and expectancy effects may interact with functional unblinding. In a recent trial, for example, no effect on depression was found for a course of up to 8 ketamine infusions and the authors argued that this may be explained by the enrollment of patients from an academic center instead of self-selected patients [50].

Furthermore, it remains unknown if the evidence base for ketamine/esketamine is affected by problems seen in antidepressant research where suicidal behaviour was sometimes reclassified as emotional worsening or “drop-out” [51,52]. In addition, it is often not clear if suicidal behaviour was assessed by blinded raters, which is known to result in lower efficacy compared to non-blinded raters [53]. Whereas assessment of suicide ideation was often part of a separate and systematic assessment process, suicidal behaviour was often only identified during the assessment of spontaneously reported adverse events. Consequently, our risk of bias assessment provides a conservative estimation of the likely biases due to rater blinding.

Esketamine is approved by the FDA and increasingly used but appears less effective than ketamine. This was also reported in other reviews with depression symptoms as an outcome [17,23,54,55] and may reflect greater amplified placebo effects of ketamine, which is usually administered intravenously. More head-to-head comparisons are needed for clarification.

A challenge to the clinical practice of repeated dosings for maintaining efficacy [23] is our explorative finding that this is not superior to single doses. This has not been systematically explored in placebo controlled trials, to our knowledge, but is in line with a recent finding of controlled and uncontrolled trials based on pre-post differences [56]. The diminishing effect could be explained by decreasing expectancy effects over time or tolerance effects. This result challenges claims that ketamine or esketamine work by stimulating neurogeneration or other processes that evolve over time.

Although numbers of suicides and deaths of any cause were small, the fact that they were higher in the ketamine/esketamine arms than in the placebo arms may represent a signal of concern, given that post-marketing surveillance data on esketamine suggests it may increase the incidence of suicidal ideation and behaviour [12]. Of note, two additional suicides and one other death occurred in drug-arms of enrichment or open label phases of studies that were excluded from our analysis [57,58]. However, suicide attempts were numerically lower in the drug arms. Concerns about potential addiction problems from longer-term use were also pointed out by our patient representatives and are well known [59,60].

Strengths of this review include the inclusion of suicidal behaviour and mortality and statistical methods to handle these rare events, thus avoiding biases resulting from excluding trials with zero events. Previous reviews were either outdated or did not report on suicidal behaviour as outcome, especially not on suicides and deaths of any cause [16–18]. Furthermore, in contrast to some previous reviews, results for suicide ideation were aggregated for different time points and the review is up to date by including studies up to December 2025. The larger number of included studies replicated the findings of a short-term effect on suicide ideation and the weaker effect for esketamine compared to ketamines [17,19,23,55], but is more precise. Limitations include potential biases resulting from our assumption that no suicidal behaviour occurred when none were mentioned in the reported adverse events. Some authors did not respond to data requests and this may introduce bias if non-response is systematically related to published outcomes. Additional limitations also affecting other reviews include the use of means and standard deviations when suicide ideation has a skewed distribution or when suicide ideation was measured on a Likert-scale with few response options, and that studies did not report results separately by sex/gender, and that follow-up time-frames substantially varied (and that we had no further information about the exact timing of events to be able to restrict time-frames).

In conclusion, there is insufficient data to ascertain whether ketamine or esketamine reduce suicide and suicide attempts. Short-term effects for ketamine on suicide ideation are detected, but these are not sustained and may be the product of acute psychoactive effects or unblinding, contributing to the low quality of efficacy estimations.

## Data Availability

All data used in this study will be made publicly available via the OSF upon publication in a journal.

https://osf.io/xzj7k/

## ACKNOWLEDGEMENTS

We thank the authors who responded to our requests for data. We also thank Janssen and Yale for making patient-level data available via the YODA platform (YODA statement: “This study, carried out under YODA Project [2024–0612], used data obtained from the Yale University Open Data Access Project, which has an agreement with JANSSEN RESEARCH & DEVELOPMENT, L.L.C.. The interpretation and reporting of research using this data are solely the responsibility of the authors and does not necessarily represent the official views of the Yale University Open Data Access Project or JANSSEN RESEARCH & DEVELOPMENT, L.L.C..”). We thank Silvi Mühringer, patient representative of SUPRA Austria for discussions and suggestions for our study. We thank Simone Amendola for assistance in designing the study and data-extraction and Blanca Acimas Müller for independent screening of the search update.

## Research ethics and consent

This is a systematic review of already published studies.

## Contributor and guarantor information

Protocol development: MP, RC, TW, MH, CV, JM; literature search: TC, VS, MP, JM; data extraction, data management, data-cleaning: MP, RC, TW, VS, MH, CV, JM, statistical analysis: MP, drafting: MP, RC, JM; reviewing and revising: MP, RC, TW, VS, MH, CV, JM. Responsible for overall content and guarantor: MP and JM. The corresponding author attests that all listed authors meet authorship criteria and that no others meeting the criteria have been omitted.

## Copyright/licence for publication

The Corresponding Author has the right to grant on behalf of all authors and does grant on behalf of all authors, a worldwide licence to the Publishers and its licensees in perpetuity, in all forms, formats and media (whether known now or created in the future), to i) publish, reproduce, distribute, display and store the Contribution, ii) translate the Contribution into other languages, create adaptations, reprints, include within collections and create summaries, extracts and/or, abstracts of the Contribution, iii) create any other derivative work(s) based on the Contribution, iv) to exploit all subsidiary rights in the Contribution, v) the inclusion of electronic links from the Contribution to third party material where-ever it may be located; and, vi) licence any third party to do any or all of the above.

## Competing interests

All authors have completed the ICMJE uniform disclosure form at http://www.icmje.org/disclosure-of-interest/ and declare: CV, MP, RC, TW, VS no support from any commercial organisation for the submitted work and have no other conflicts of interest to declare; JM has received royalties for books about psychiatric drugs; MH received support from a clinical research fellowship at NELFT (NHS), royalties from the Maudsley Deprescribing Guidelines, and is a co-founder of Outro Health, a digital clinic in the US where he receives consulting fees.

## Data sharing

The data and statistical code are available on the OSF [61]

## Notes

### Clinical Protocols

https://www.crd.york.ac.uk/prospero/display_record.php?ID=CRD42023364156

### Funding Statement

No funding was involved

### Summary of Updates

Table with description of studies now in supplement and supplement is restructured.

