## Supplement for "Effects of ketamine and esketamine on death, suicidal behaviour, and suicidal ideation in psychiatric disorders: A systematic review and meta-analysis" (from another version of this preprint: version 1, 2025-08-21)

#### Ketamine and Esketamine in Suicide Prevention

Martin Plöderl

2025-08-07

### Contents

**Search Strategy** 3

**PRISMA Checklist** 4

**PRISMA Flow Chart** 9

**Excluded Studies** 10

**Risk of Bias Analysis** 13

**Blinding Success** 14

**Meta-Analytic Results - Summary Tables** 15

**Funnel Plots, Suicide Ideation Meta-Analyses** **167**

#### Search Strategy

Table S1: Search Strategy

| Steps | Terms |
|---|---|
| 1 | ketamin* or ketalar OR ketaject OR ketanest OR esketamin* OR spravato |
| 2 | (depression or depressive or mood disorder* or affective disorder* or bipolar or psychosis or psychotic disorder* or schizophren* or schizoaffect* or eating disorder* or anorexi* or bulimi* or EDNOS or anxi* or anxiety disorder* or general* anxiety or GAD or obsess* or OCD or trauma* or PTSD or personality disorder* or paranoi* or schizoid or schizotyp* or antisocial or borderline or histroni* or narcissisti* or avoidant or dependent or substance misuse or heroin or alcohol or alcoholism or cocaine or cannabis or amphetamine or methamphetamine or tobacco or nicotine or opi*).mp. [mp=title, abstract, original title, name of substance word, subject heading word, floating sub-heading word, keyword heading word, organism supplementary concept word, protocol supplementary concept word, rare disease supplementary concept word, unique identifier, synonyms] |
| 3 | 1 and 2 |
| 4 | Limit to randomised controlled trial |
| 5 | Limit to humans |

The strategy in Table S1 was used to search PsychINFO, Embase, MEDLINE

No language restrictions were applied.

#### PRISMA Checklist

| Section and Topic | Item # | Checklist item | Location where item is reported |
|---|---|---|---|
| **TITLE** | | | |
| Title | 1 | Identify the report as a systematic review. | Title |
| **ABSTRACT** | | | |
| Abstract | 2 | See the PRISMA 2020 for Abstracts checklist. | Abstract |
| **INTRODUCTION** | | | |
| Rationale | 3 | Describe the rationale for the review in the context of existing knowledge. | Introduction |
| Objectives | 4 | Provide an explicit statement of the objective(s) or question(s) the review addresses. | Introduction |
| **METHODS** | | | |
| Eligibility criteria | 5 | Specify the inclusion and exclusion criteria for the review and how studies were grouped for the syntheses. | Eligibility criteria |
| Information sources | 6 | Specify all databases, registers, websites, organisations, reference lists and other sources searched or consulted to identify studies. Specify the date when each source was last searched or consulted. | Information sources and search strategy |
| Search strategy | 7 | Present the full search strategies for all databases, registers and websites, including any filters and limits used. | Information sources and search strategy, Table S1 |
| Selection process | 8 | Specify the methods used to decide whether a study met the inclusion criteria of the review, including how many reviewers screened each record and each report retrieved, whether they worked independently, and if applicable, details of automation tools used in the process. | Selection process |
| Data collection process | 9 | Specify the methods used to collect data from reports, including how many reviewers collected data from each report, whether they worked independently, any processes for obtaining or confirming data from study investigators, and if applicable, details of automation tools used in the process. | Data collection process |
| Data items | 10a | List and define all outcomes for which data were sought. Specify whether all results that were compatible with each outcome domain in each study were sought (e.g. for all measures, time points, analyses), and if not, the methods used to decide which results to collect. | Data items and outcome measures |
| | 10b | List and define all other variables for which data were sought (e.g. participant and intervention characteristics, funding sources). Describe any assumptions made about any missing or unclear information. | Data items and outcome measures |

| Section and Topic | Item # | Checklist item | Location where item is reported |
|---|---|---|---|
| Study risk of bias assessment | 11 | Specify the methods used to assess risk of bias in the included studies, including details of the tool(s) used, how many reviewers assessed each study and whether they worked independently, and if applicable, details of automation tools used in the process. | Quality assessment |
| Effect measures | 12 | Specify for each outcome the effect measure(s) (e.g. risk ratio, mean difference) used in the synthesis or presentation of results. | Synthesis methods |
| Synthesis methods | 13a | Describe the processes used to decide which studies were eligible for each synthesis (e.g. tabulating the study intervention characteristics and comparing against the planned groups for each synthesis (item #5)). | Synthesis methods |
| | 13b | Describe any methods required to prepare the data for presentation or synthesis, such as handling of missing summary statistics, or data conversions. | Synthesis methods |
| | 13c | Describe any methods used to tabulate or visually display results of individual studies and syntheses. | Synthesis methods |
| | 13d | Describe any methods used to synthesize results and provide a rationale for the choice(s). If meta-analysis was performed, describe the model(s), method(s) to identify the presence and extent of statistical heterogeneity, and software package(s) used. | Synthesis methods |
| | 13e | Describe any methods used to explore possible causes of heterogeneity among study results (e.g. subgroup analysis, meta-regression). | Synthesis methods |
| | 13f | Describe any sensitivity analyses conducted to assess robustness of the synthesized results. | Synthesis methods |
| Reporting bias assessment | 14 | Describe any methods used to assess risk of bias due to missing results in a synthesis (arising from reporting biases). | Quality assessment |
| Certainty assessment | 15 | Describe any methods used to assess certainty (or confidence) in the body of evidence for an outcome. | Quality assessment |
| **RESULTS** | | | |
| Study selection | 16a | Describe the results of the search and selection process, from the number of records identified in the search to the number of studies included in the review, ideally using a flow diagram. | Results, Figure S1 |
| | 16b | Cite studies that might appear to meet the inclusion criteria, but which were excluded, and explain why they were excluded. | Supplement Table S2 |
| Study characteristics | 17 | Cite each included study and present its characteristics. | Table 1 |
| Risk of bias in studies | 18 | Present assessments of risk of bias for each included study. | Supplement Figure S2 |

| Section and Topic | Item # | Checklist item | Location where item is reported |
|---|---|---|---|
| Results of individual studies | 19 | For all outcomes, present, for each study: (a) summary statistics for each group (where appropriate) and (b) an effect estimate and its precision (e.g. confidence/credible interval), ideally using structured tables or plots. | Forest plots in supplement, also data- |
| Results of syntheses | 20a | For each synthesis, briefly summarise the characteristics and risk of bias among contributing studies. | Results |
| | 20b | Present results of all statistical syntheses conducted. If meta-analysis was done, present for each the summary estimate and its precision (e.g. confidence/credible interval) and measures of statistical heterogeneity. If comparing groups, describe the direction of the effect. | Tables 2-4 in pape and S3-S16 in supplement |
| | 20c | Present results of all investigations of possible causes of heterogeneity among study results. | Results |
| | 20d | Present results of all sensitivity analyses conducted to assess the robustness of the synthesized results. | Results |
| Reporting biases | 21 | Present assessments of risk of bias due to missing results (arising from reporting biases) for each synthesis assessed. | Table 3, Funnel plots in suppl. |
| Certainty of evidence | 22 | Present assessments of certainty (or confidence) in the body of evidence for each outcome assessed. | Results |
| **DISCUSSION** | | | |
| Discussion | 23a | Provide a general interpretation of the results in the context of other evidence. | Discussion |
| | 23b | Discuss any limitations of the evidence included in the review. | Discussion |
| | 23c | Discuss any limitations of the review processes used. | Discussion |
| | 23d | Discuss implications of the results for practice, policy, and future research. | Discussion |
| **OTHER INFORMATION** | | | |
| Registration and protocol | 24a | Provide registration information for the review, including register name and registration number, or state that the review was not registered. | Abstract, Methods |
| | 24b | Indicate where the review protocol can be accessed, or state that a protocol was not prepared. | Abstract, Methods |
| | 24c | Describe and explain any amendments to information provided at registration or in the protocol. | Methods |

| Section and Topic | Item # | Checklist item | Location where item is reported |
|---|---|---|---|
| Support | 25 | Describe sources of financial or non-financial support for the review, and the role of the funders or sponsors in the review. | Abstract, Methods |
| Competing interests | 26 | Declare any competing interests of review authors. | Competing interest statement |
| Availability of data, code and other materials | 27 | Report which of the following are publicly available and where they can be found: template data collection forms; data extracted from included studies; data used for all analyses; analytic code; any other materials used in the review. | Methods, Data-sharing statement |

### PRISMA Flow Chart

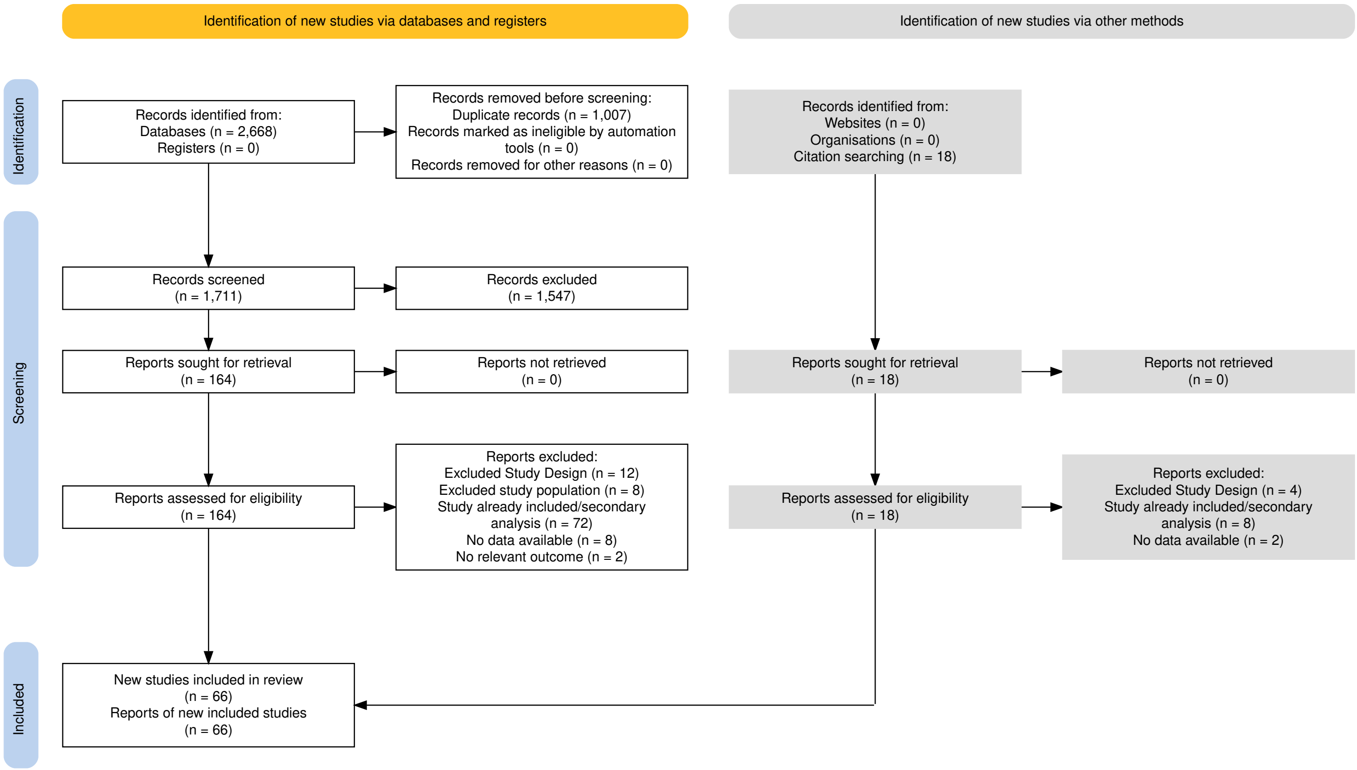

Figure S1: PRISMA flow chart

Created with PRISMA2020

Haddaway, N. R., Page, M. J., Pritchard, C. C., & McGuinness, L. A. (2022). PRISMA2020: An R package and Shiny app for producing PRISMA 2020-compliant flow diagrams, with interactivity for optimised digital transparency and Open Synthesis Campbell Systematic Reviews, 18, e1230. https://doi.org/10.1002/cl2.1230

#### Excluded Studies

Table S2: Excluded studies at the full-text screening stage

Table starts next page

| Author | Reason for exclusion | PRISMA Category |
|---|---|---|
| Abdallah 2022 | overlap with older search / study already included | Trial already included |
| Abdullah 2019 | no placebo group | Excluded trial design |
| Acevedo Diaz 2020 | Study already included/secondary analysis (Zarate) | Trial already included |
| Alexander 2023 | Study already included/secondary analysis (Zarate) | Trial already included |
| Arabzadeh 2018 | Queries for data could not be clarified | No data available |
| Araujo 2021 | No placebo group | Excluded trial design |
| Ballard 2017 | Study already included/secondary analysis (Zarate) | Trial already included |
| Ballard 2018 | Study already included/secondary analysis (Zarate) | Trial already included |
| Ballard 2024 | Study already included/secondary analysis (Zarate) | Trial already included |
| Beanes 2022 | active comparator/no placebo | Excluded trial design |
| Bermann 2000 | Unable to get data | No data available |
| Canuso 2019 | Studies aleardy included (Esketamine ASPIRE) | Trial already included |
| Canuso 2021 | Studies aleardy included (Esketamine ASPIRE) | Trial already included |
| Cao 2018 | Study already included/secondary analysis (Su 2017) | Trial already included |
| Chen 2018 a Cognitive function of patients with TR | Study already included/secondary analysis (Su 2017) | Trial already included |
| Chen 2018 b Rapid inflammation | Study already included/secondary analysis (Su 2017) | Trial already included |
| Chen 2018 c Persistent antidepressant effect | Study already included/secondary analysis (Su 2017) | Trial already included |
| Chen 2019 a Maintenance of | Study already included/secondary analysis (Su 2017) | Trial already included |
| Chen 2019 b Antidepressant and antisuicidal effect | Study already included/secondary analysis (Su 2017) | Trial already included |
| Chen 2020 Functional Dysconnectivity | Study already included/secondary analysis (Su 2017) | Trial already included |
| Chen 2021 a Effects of treatment refractoriness | Study already included/secondary analysis (Su 2017) | Trial already included |
| Chen 2021 b Low-dose ketamine infusion for treati | Study already included/secondary analysis (Su 2017) | Trial already included |
| Chen 2021 c Efficacy oflow-dose ketamine in anxio | Study already included/secondary analysis (Su 2017) | Trial already included |
| Chen 2021 d Is one or two infusions | Study already included/secondary analysis (Su 2017) | Trial already included |
| Chen 2021 e Interest-activity symptom severity | Study already included/secondary analysis (Su 2017) | Trial already included |
| Chen 2022 a Low-dose ketamine infusion in | Study already included/secondary analysis (Su 2017) | Trial already included |
| Chen 2022 b Relationship Between Dissociation | Study already included/secondary analysis (TRANSFORM) | Trial already included |
| Chen 2022 c Baseline Working Memory | Secondary Analysis (Su 2017) | Trial already included |
| Chen 2023 a Cortical inhibition function | Study already included/secondary analysis (Su 2023) | Trial already included |
| Chen 2023 b Effects of low-dose ketamine | Secondary Analysis (Su et al. 2023) | Trial already included |
| Chen 2023 c Effects of melancholic features | Secondary Analysis (Su et al. 2023) | Trial already included |
| Chen 2023 d - Klotho | Study already included/secondary analysis (Su 2023) | Trial already included |
| Citrome 2020 | Study already included/secondary analysis (TRANSFORM) | Trial already included |
| Costi 2019 | Excluded trial design | Excluded trial design |
| Dai 2025 | non-psychiatric sample (prevention of depression) | Excluded study population |
| Daly 2019 | Excluded - enrichment design | Excluded trial design |
| Danböck 2024 | No relevant outcome assessed | No relevant outcome |
| De la Salle 2022 | Study already included/secondary analysis (Phillips 2019) | Trial already included |
| De Salle 2022 | overlap with older search / study already included | Trial already included |
| Diazgrandos 2010 | Study already included/secondary analysis (Zarate) | Trial already included |
| Diekamp | Studies already included (Esketamine ASPIRE) | Trial already included |
| Domany 2022 | Study already included in older search | Trial already included |
| Donel 2024 | Study already included/secondary analysis (Loo 2023) | Trial already included |
| Dong 2023 | Intervention and control group differ in treatments | Excluded trial design |
| Doty 2021 | Studies already included (Esketamine TRANSFORM/SUSTAIN) | Trial already included |
| Downey 2016 | Unable to get data, and nothing available via clinicaltrials.gov | No data available |
| Duncan 2017 | Study already included/secondary analysis (Zarate) | Trial already included |
| Dwyer 2019 | Study already included (Dwyer 2021) | Trial already included |
| Evans 2018 | Study already included/secondary analysis (Zarate) | Trial already included |
| Evans 2024 | Study already included/secondary analysis (Zarate) | Trial already included |
| Fan et al. 2017 | Participants inclusion criteria not met) | Excluded study population |
| Feeney 2021 | Study already included/secondary analysis (Fava 2020) | Trial already included |
| Finnegan 2016 | Protocol | Excluded trial design |
| Finnegan et al. 2019 | Participants inclusion criteria not met) | Excluded study population |
| Fu 2023 | Study already included/secondary analysis (Fu 2020) | Trial already included |
| Galvez 2018 | Unable to get data | No data available |
| Gent 2024 | No outcome of interest, no adverse events mentioned at all | No relevant outcome |
| Gilbert 2020 | Study already included/secondary analysis (Zarate) | Trial already included |
| Gilbert 2022 | Study already included/secondary analysis (Zarate) | Trial already included |
| Glue 2024a | Enrichment design | Excluded trial design |
| Grabski 2022 | overlap with older search / study already included | Trial already included |
| Greenway 2024 | No placebo control group | Excluded trial design |
| Ha 2023 | no control group | Excluded trial design |
| Hack 2023 | Nonclinical sample | Excluded study population |
| Herzog 2024 | Healthy participants | Excluded study population |
| Hess 2024 | Study already included/secondary analysis (Zarate) | Trial already included |
| Hochschild 2022 | Study already included/secondary analysis (Grunebaum 2018) | Trial already included |
| Hochschild 2022 | Already in older search (and secondary, Grunebaum 2018) | Trial already included |
| Jesus-Nunes 2022 | active control, no placebo | Excluded trial design |
| Jiang2021 | Study already included/secondary analysis (Tiger 2020) | Trial already included |

| | | |
|---|---|---|
| Jollant 2024 | Study already included/secondary analysis (Abbar 2023) | Trial already included |
| Keilp2021 | Study already included/secondary analysis (Grunebaum 2018) | Trial already included |
| Kheirabadi2020 | Comparison with ECT | Excluded trial design |
| Kopelman 2023 | Study already included/secondary analysis (Price 2022) | Trial already included |
| Lally 2014 | Study already included/secondary analysis (Zarate) | Trial already included |
| Li 2023 a Right dorsolateral prefrontal | Study already included/secondary analysis (Su 2023) | Trial already included |
| Li 2024 | Not clinical population | Excluded study population |
| Li2016 | Unable to get data | No data available |
| Lijfijt 2022 | overlap with older search / study already included | Trial already included |
| Lin 2023 | secondary analysis on Lin 2016 and Su 2017 | Trial already included |
| Lin 2024 a | Study already included/secondary analysis (Su 2023) | Trial already included |
| Lin 2024 b | No control group | Excluded trial design |
| Lundin 2021 | Study already included/secondary analysis (Zarate) | Trial already included |
| Machado-Vieira 2024 | Study already included/secondary analysis (Jones 2024) | Trial already included |
| McMillan 2020 | Study already included/secondary analysis (Sumner 2019) | Trial already included |
| Moaddel 2018 | Study already included/secondary analysis (Zarate) | Trial already included |
| Morris 2020 | Excluded trial design (is Costi 2019) | Trial already included |
| Murphy 2023 | Study already included/secondary analysis (Lijfijt) | Trial already included |
| Murrough 2015 I | Study already included/secondary analysis (Murrough 2013) | Trial already included |
| Nugent 2018 | Study already included/secondary analysis (Zarate) | Trial already included |
| Nugent 2019 | Study already included/secondary analysis (Zarate) | Trial already included |
| O'Brien 2019 | this is a study protocol | Excluded trial design |
| Ohnishi 2022 | Study already included/secondary analysis (Takahashi 2021) | Trial already included |
| Pathak 2021 | Queries for data could not be clarified | No data available |
| Phillips 2020 | Study already included/secondary analysis (Phillips 2019) | Trial already included |
| Price 2013 | Study already included/secondary analysis (Murrough 2013) | Trial already included |
| Price 2023 | includes FU phase where we get the data anyway | Trial already included |
| Reed 2018 | Study already included/secondary analysis (Zarate) | Trial already included |
| Reed 2019 | Study already included/secondary analysis (Zarate) | Trial already included |
| Ren 2024 | No data on suicide ideation, not certin if suicidal behavior occurred or not | No data available |
| Sajid 2024 | Study already included/secondary analysis (Grunebaum 2018) | Trial already included |
| Salloum 2019 | Study already included/secondary analysis (Fava 2020) | Trial already included |
| Salloum 2020 | Study already included/secondary analysis (Fava 2020) | Trial already included |
| Shen 2023 | non-psychiatric sample (prevention of depression) | Excluded study population |
| Shiroma 2020 | Study already included/secondary analysis (Shiroma 2020) | Trial already included |
| Singh 2022 | Open label trial, no placebo group | Excluded trial design |
| Sumner 2021 | Study already included/secondary analysis (Sumner 2019) | Trial already included |
| Tu 2025 | Study already included/secondary analysis (Su 2017) | Trial already included |
| Veldman 2021 | Study already included/secondary analysis (Tiger 2020) | Trial already included |
| Wasserthal 2023 | Not clinical population | Excluded study population |
| Wilkowska 2021 | No control group | Excluded trial design |
| Wink2021 | Unable to get data | No data available |
| Woolsey 2022 | No data on suicide ideation, not certin if suicidal behavior occurred or not | No data available |
| Xu2015 | Study already included/secondary analysis (Zarate) | Trial already included |
| Yonezawa 2024 | Study already included/secondary analysis (Fava 2020) | Trial already included |
| Zolghadriha 2024 | Queries for data could not be clarified | No data available |

### Risk of Bias Analysis

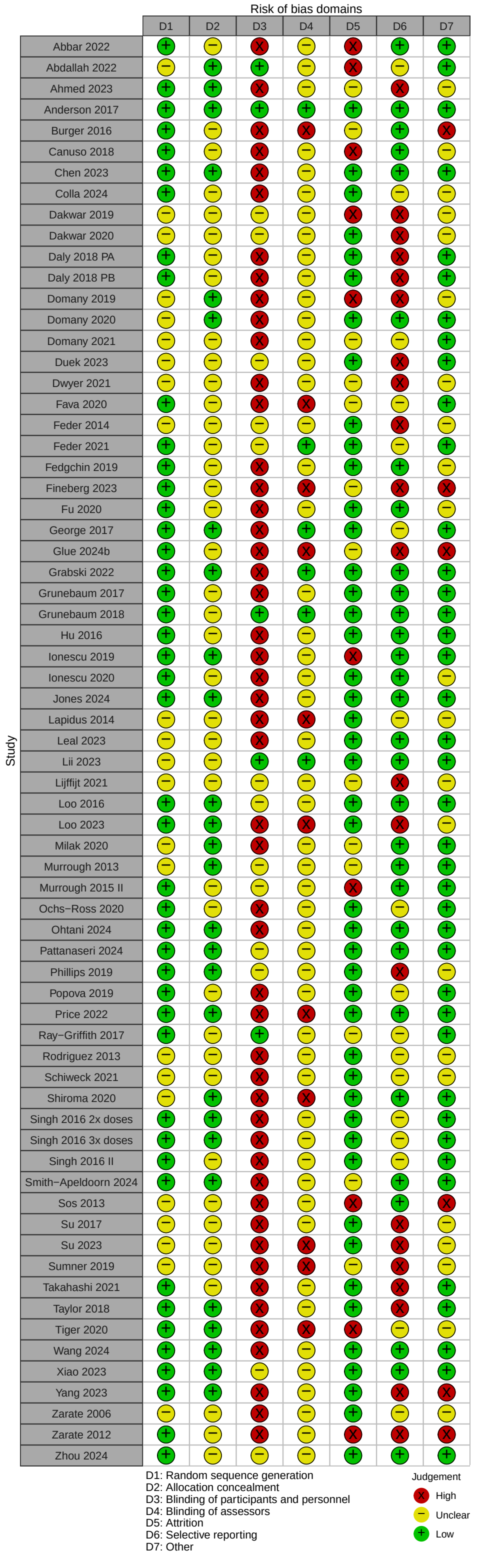

Figure S2: Risk of Bias

### Blinding Success

Table S3: Blinding success

| Study | Interventions.used | Participants | Raters |
|---|---|---|---|
| Abdullah 2022 | Ketamine vs saline placebo | Evidence of unblinding | Not tested |
| Anderson 2017 | Ketamine vs saline placebo both administered along with an anaesthetic for ECT | No evidence of unblinding | No evidence of unblinding |
| Dwyer 2021 | Ketamine vs midazolam | Evidence of unblinding | Not tested |
| Feder 2021 | Ketamine vs midazolam | Unclear** | Not tested |
| Fava 2020 | Ketamine vs midazolam | Evidence of unblinding | Evidence of unblinding |
| Grabski 2022 | Ketamine vs saline placebo | Evidence of unblinding | Not reported |
| Grunbaum 2017 | Ketamine vs midazolam | Evidence of unblinding | No evidence of unblinding |
| Grunbaum 2018 | Ketamine vs midazolam | No evidence of unblinding | No evidence of unblinding |
| Lii 2024 | Ketamine vs. Saline under surgery | No evidence of unblinding | Not tested |
| Lijfijt 2021 | Ketamine vs midazolam | Evidence of unblinding | Evidence of unblinding |
| Loo 2023 | Ketamine vs midazolam | Evidence of unblinding | Evidence of unblinding |
| Price 2022 | Ketamine vs saline placebo | Evidence of unblinding | Evidence of unblinding |
| Shiroma 2020 | Ketamine vs midazolam | Evidence of unblinding | Evidence of unblinding |
| Smith-Appeldorn 2024 | Ketamine vs saline placebo | Evidence of unblinding | Not tested |
| Sumner 2019 | Ketamine vs remifentanil | Evidence of unblinding | Evidence of unblinding |

One study was considered as adequately blinded which did not test the blind. This was the study by Ray-Griffith et al 2017 in which ketamine and an active comparator were administered as part of an anaesthetic procedure for ECT

**correct guesses are less than 50%, but a large number of guesses are not counted because of people having ‘changed their minds during the course of the study’

#### Meta-Analytic Results - Summary Tables

#### Suicidal Behavior and Mortality

##### Suicides and Suicide Attempts Combined

Table S4: Suicidal behavior: Meta analytic results, treatment- and follow-up phases combined

| Analysis | OR [95% CI] | p | I2 [95% CI] | T2 [95%CI] | number of studies |
|---|---|---|---|---|---|
| Exact (Liu et al. 2012) | 0.97 [0.48 - 1.97] | | | | 65 |
| Bayesian - Uninformative prior (delta = 250) | 0.86 [0.45 - 1.56] | | | 1.34 [1.01 - 2.59] | 65 |
| Bayesian - Informative prior (delta = 15) | 0.86 [0.46 - 1.54] | | | 1.33 [1.01 - 2.55] | 65 |
| Binomial-normal GLMM, fixed intercept | 0.91 [0.52, 1.6] | | | 0.00 | |
| Binomial-normal GLMM, fixed intercept, alternative parameterization | 0.91 [0.51, 1.6] | | | 0.00 | |
| Binomial-normal GLMM, random intercept | 0.89 [0.51, 1.56] | | | 0.00 | |
| Binomial-normal GLMM, random intercept, alternative parameterization | 0.89 [0.53, 1.57] | | | 0.00 | |
| Hypergeometric-normal GLMM (Nchg) | 0.92 [0.53, 1.58] | | | 0.00 | |
| Conditional binomial GLMM (Condbin) | 0.92 [0.26, 1.58] | | | 0.00 | |
| Beta-binomial model | 0.66 [0.52, 1.66] | | | | |
| Peto | 0.88 [0.49 - 1.61] | 0.69 | 19 [0 - 57] | 0.27 [0.00 - 2.10] | 13 |
| Mantel-Haenszel - with continuity correction | 0.88 [0.59 - 1.30] | 0.52 | 0 [0 - 29] | 0.00 [0.00 - 0.00] | 65 |
| Mantel-Haenszel - with continuity/treatment arm correction | 0.99 [0.66 - 1.48] | 0.97 | 0 [0 - 29] | 0.00 [0.00 - 0.00] | 65 |
| Mantel-Haenszel - risk difference | 0.00 [-0.01 - 0.01] | 0.99 | 0 [0 - 29] | 0.00 [0.00 - 0.00] | 65 |
| Arcsine square root transformed risk difference | 0.00 [-0.03 - 0.02] | 0.78 | 0 [ - ] | 0.00 [ - ] | 65 |

Table S5: Suicidal behavior: Meta analytic results, treatment phase

| Analysis | OR [95% CI] | p | I2 [95% CI] | T2 [95%CI] | number of studies |
|---|---|---|---|---|---|
| Exact (Liu et al. 2012) | 1.03 [0.21 - 5.19] | | | | 65 |
| Bayesian - Uninformative prior (delta = 250) | 0.94 [0.39 - 2.19] | | | 1.31 [1.01 - 2.54] | 65 |
| Bayesian - Informative prior (delta = 15) | 0.94 [0.41 - 2.15] | | | 1.32 [1.01 - 2.53] | 65 |
| Binomial-normal GLMM, fixed intercept | 0.99 [0.39, 2.52] | | | 0.00 | |
| Binomial-normal GLMM, fixed intercept, alternative parameterization | 0.99 [0.36, 2.52] | | | 0.00 | |
| Binomial-normal GLMM, random intercept | 0.91 [0.36, 2.31] | | | 0.00 | |
| Binomial-normal GLMM, random intercept, alternative parameterization | 0.91 [0.39, 2.31] | | | 0.00 | |
| Hypergeometric-normal GLMM (Nchg) | 0.99 [0.39, 2.5] | | | 0.00 | |
| Conditional binomial GLMM (Condbin) | 0.99 [0.27, 2.5] | | | 0.00 | |
| Beta-binomial model | 0.82 [0.39, 2.49] | | | | |
| Peto | 1.00 [0.40 - 2.53] | 1.00 | 0 [0 - 65] | 0.00 [0.00 - 2.79] | 9 |
| Mantel-Haenszel - with continuity correction | 1.00 [0.63 - 1.58] | 1.00 | 0 [0 - 29] | 0.00 [0.00 - 0.00] | 65 |
| Mantel-Haenszel - with continuity/treatment arm correction | 1.00 [0.63 - 1.58] | 1.00 | 0 [0 - 29] | 0.00 [0.00 - 0.00] | 65 |
| Mantel-Haenszel - risk difference | 0.00 [0.00 - 0.00] | 0.94 | 0 [0 - 29] | 0.00 [0.00 - 0.00] | 65 |
| Arcsine square root transformed risk difference | 0.00 [-0.03 - 0.03] | 0.91 | 0 [ - ] | 0.00 [ - ] | 65 |

Table S6: Suicidal behavior: Meta analytic results, follow-up phase

| Analysis | OR [95% CI] | p | I2 [95% CI] | T2 [95%CI] | number of studies |
|---|---|---|---|---|---|
| Exact (Liu et al. 2012) | 0.94 [0.39 - 2.28] | | | | 44 |
| Bayesian - Uninformative prior (delta = 250) | 0.83 [0.38 - 1.75] | | | 1.38 [1.02 - 2.78] | 44 |
| Bayesian - Informative prior (delta = 15) | 0.84 [0.39 - 1.74] | | | 1.38 [1.02 - 2.76] | 44 |
| Binomial-normal GLMM, fixed intercept | 0.86 [0.43, 1.72] | | | 0.00 | |
| Binomial-normal GLMM, fixed intercept, alternative parameterization | 0.86 [0.43, 1.72] | | | 0.00 | |
| Binomial-normal GLMM, random intercept | 0.86 [0.43, 1.72] | | | 0.00 | |
| Binomial-normal GLMM, random intercept, alternative parameterization | 0.86 [0.44, 1.72] | | | 0.00 | |
| Hypergeometric-normal GLMM (Nchg) | 0.86 [0.44, 1.7] | | | 0.00 | |
| Conditional binomial GLMM (Condbin) | 0.86 [0.2, 1.7] | | | 0.00 | |
| Beta-binomial model | 0.64 [0.43, 1.99] | | | | |
| Peto | 0.78 [0.32 - 1.92] | 0.59 | 26 [0 - 66] | 0.65 [0.00 - 2.76] | 8 |
| Mantel-Haenszel - with continuity correction | 0.88 [0.54 - 1.43] | 0.60 | 0 [0 - 35] | 0.00 [0.00 - 0.00] | 44 |
| Mantel-Haenszel - with continuity/treatment arm correction | 0.94 [0.58 - 1.55] | 0.82 | 0 [0 - 35] | 0.00 [0.00 - 0.00] | 44 |
| Mantel-Haenszel - risk difference | 0.00 [-0.01 - 0.01] | 0.92 | 0 [0 - 35] | 0.00 [0.00 - 0.00] | 44 |
| Arcsine square root transformed risk difference | 0.00 [-0.04 - 0.03] | 0.85 | 0 [ - ] | 0.00 [ - ] | 44 |

Note: All meta-analytic methods are appropriate except Peto's method, because there are both single-arm-zero-events studies and double-arm-zero-events studies and the total event count in each arm is non-zero (Xu et al., 2021)

#### Suicide Attempts

Table S7: Suicide attempts: Meta analytic results, treatment- and follow-up phases combined

| Analysis | OR [95% CI] | p | I2 [95% CI] | T2 [95%CI] | number of studies |
|---|---|---|---|---|---|
| Exact (Liu et al. 2012) | 0.84 [0.39 - 1.79] | | | | 65 |
| Bayesian - Uninformative prior (delta = 250) | 0.75 [0.39 - 1.38] | | | 1.34 [1.01 - 2.55] | 65 |
| Bayesian - Informative prior (delta = 15) | 0.76 [0.39 - 1.39] | | | 1.34 [1.01 - 2.55] | 65 |
| Binomial-normal GLMM, fixed intercept | 0.79 [0.44, 1.43] | | | 0.00 | |
| Binomial-normal GLMM, fixed intercept, alternative parameterization | 0.79 [0.2, 1.43] | | | 0.00 | |
| Binomial-normal GLMM, random intercept | 0.72 [0.3, 2.55] | | | 0.01 | |
| Binomial-normal GLMM, random intercept, alternative parameterization | 0.76 [0.45, 1.91] | | | 0.00 | |
| Hypergeometric-normal GLMM (Nchg) | 0.80 [0.45, 1.42] | | | 0.00 | |
| Conditional binomial GLMM (Condbin) | 0.80 [0.23, 1.42] | | | 0.00 | |
| Beta-binomial model | 0.63 [0.44, 1.68] | | | | |
| Peto | 0.75 [0.39 - 1.43] | 0.38 | 15 [0 - 55] | 0.35 [0.00 - 2.20] | 12 |
| Mantel-Haenszel - with continuity correction | 0.83 [0.55 - 1.24] | 0.36 | 0 [0 - 29] | 0.00 [0.00 - 0.00] | 65 |
| Mantel-Haenszel - with continuity/treatment arm correction | 0.94 [0.62 - 1.41] | 0.77 | 0 [0 - 29] | 0.00 [0.00 - 0.00] | 65 |
| Mantel-Haenszel - risk difference | 0.00 [-0.01 - 0.00] | 0.93 | 0 [0 - 29] | 0.00 [0.00 - 0.00] | 65 |
| Arcsine square root transformed risk difference | -0.01 [-0.04 - 0.02] | 0.59 | 0 [ - ] | 0.00 [ - ] | 65 |

Table S8: Suicide attempts: Meta analytic results, treatment phase

| Analysis | OR [95% CI] | p | I2 [95% CI] | T2 [95%CI] | number of studies |
|---|---|---|---|---|---|
| Exact (Liu et al. 2012) | 0.9 [0.13 - 5.83] | | | | 65 |
| Bayesian - Uninformative prior (delta = 250) | 0.85 [0.34 - 2.12] | | | 1.31 [1.01 - 2.53] | 65 |
| Bayesian - Informative prior (delta = 15) | 0.86 [0.36 - 2.05] | | | 1.32 [1.01 - 2.54] | 65 |
| Binomial-normal GLMM, fixed intercept | 0.87 [0.31, 2.43] | | | 0.00 | |
| Binomial-normal GLMM, fixed intercept, alternative parameterization | 0.87 [0.29, 2.43] | | | 0.00 | |
| Binomial-normal GLMM, random intercept | 0.80 [0.29, 2.22] | | | 0.00 | |
| Binomial-normal GLMM, random intercept, alternative parameterization | 0.80 [0.32, 2.22] | | | 0.00 | |
| Hypergeometric-normal GLMM (Nchg) | 0.87 [0.32, 2.41] | | | 0.00 | |
| Conditional binomial GLMM (Condbin) | 0.87 [0.23, 2.41] | | | 0.00 | |
| Beta-binomial model | 0.78 [0.31, 2.67] | | | | |
| Peto | 0.87 [0.32 - 2.42] | 0.80 | 0 [0 - 68] | 0.00 [0.00 - 3.26] | 8 |
| Mantel-Haenszel - with continuity correction | 0.99 [0.62 - 1.56] | 0.95 | 0 [0 - 29] | 0.00 [0.00 - 0.00] | 65 |
| Mantel-Haenszel - with continuity/treatment arm correction | 0.99 [0.62 - 1.56] | 0.95 | 0 [0 - 29] | 0.00 [0.00 - 0.00] | 65 |
| Mantel-Haenszel - risk difference | 0.00 [0.00 - 0.00] | 0.98 | 0 [0 - 29] | 0.00 [0.00 - 0.00] | 65 |
| Arcsine square root transformed risk difference | 0.00 [-0.03 - 0.03] | 0.98 | 0 [ - ] | 0.00 [ - ] | 65 |

Table S9: Suicide attempts: Meta analytic results, follow-up phase

| Analysis | OR [95% CI] | p | I2 [95% CI] | T2 [95%CI] | number of studies |
|---|---|---|---|---|---|
| Exact (Liu et al. 2012) | 0.82 [0.32 - 2.04] | | | | 44 |
| Bayesian - Uninformative prior (delta = 250) | 0.74 [0.33 - 1.57] | | | 1.38 [1.02 - 2.76] | 44 |
| Bayesian - Informative prior (delta = 15) | 0.75 [0.35 - 1.58] | | | 1.37 [1.02 - 2.74] | 44 |
| Binomial-normal GLMM, fixed intercept | 0.74 [0.36, 1.53] | | | 0.00 | |
| Binomial-normal GLMM, fixed intercept, alternative parameterization | 0.74 [0.36, 1.53] | | | 0.00 | |
| Binomial-normal GLMM, random intercept | 0.74 [0.3, 1.53] | | | 0.00 | |
| Binomial-normal GLMM, random intercept, alternative parameterization | 0.73 [0.37, 1.77] | | | 0.02 | |
| Hypergeometric-normal GLMM (Nchg) | 0.75 [0.37, 1.52] | | | 0.00 | |
| Conditional binomial GLMM (Condbin) | 0.75 [0.2, 1.52] | | | 0.00 | |
| Beta-binomial model | 0.62 [0.36, 1.94] | | | | |
| Peto | 0.70 [0.29 - 1.69] | 0.43 | 21 [0 - 63] | 0.59 [0.00 - 2.66] | 8 |
| Mantel-Haenszel - with continuity correction | 0.83 [0.50 - 1.35] | 0.45 | 0 [0 - 35] | 0.00 [0.00 - 0.00] | 44 |
| Mantel-Haenszel - with continuity/treatment arm correction | 0.89 [0.54 - 1.47] | 0.64 | 0 [0 - 35] | 0.00 [0.00 - 0.00] | 44 |
| Mantel-Haenszel - risk difference | 0.00 [-0.01 - 0.01] | 0.98 | 0 [0 - 35] | 0.00 [0.00 - 0.00] | 44 |
| Arcsine square root transformed risk difference | -0.01 [-0.04 - 0.03] | 0.71 | 0 [ - ] | 0.00 [ - ] | 44 |

Note: All meta-analytic methods are appropriate except Peto's method, because there are both single-arm-zero-events studies and double-arm-zero-events studies and the total event count in each arm is non-zero (Xu et al., 2021)

#### Suicides

Table S10: Suicides: Meta analytic results, treatment- and follow-up phases combined

| Analysis | OR [95% CI] | p | I2 [95% CI] | T2 [95%CI] | number of studies |
|---|---|---|---|---|---|
| Exact (Liu et al. 2012) | 4.57 [0.01 - Inf] | | | | 65 |
| Bayesian - Uninformative prior (delta = 250) | 1.53 [0.47 - 5.55] | | | 1.33 [1.01 - 2.59] | 65 |
| Bayesian - Informative prior (delta = 15) | 1.44 [0.47 - 4.69] | | | 1.33 [1.01 - 2.59] | 65 |
| Binomial-normal GLMM, fixed intercept | 4.06 [0.45, 36.64] | | | 0.00 | |
| Binomial-normal GLMM, fixed intercept, alternative parameterization | 4.06 [0.4, 36.64] | | | 0.00 | |
| Binomial-normal GLMM, random intercept | 3.60 [0.4, 32.66] | | | 0.00 | |
| Binomial-normal GLMM, random intercept, alternative parameterization | 3.60 [0.45, 32.68] | | | 0.00 | |
| Hypergeometric-normal GLMM (Nchg) | 4.01 [0.45, 35.95] | | | 0.00 | |
| Conditional binomial GLMM (Condbin) | 4.01 [0.35, 35.95] | | | 0.00 | |
| Beta-binomial model | 3.16 [0.45, 28.23] | | | | |
| Peto | 3.35 [0.58 - 19.40] | 0.18 | 0 [0 - 85] | 0.00 [0.00 - 3.58] | 4 |
| Mantel-Haenszel - with continuity correction | 0.88 [0.54 - 1.42] | 0.60 | 0 [0 - 29] | 0.00 [0.00 - 0.00] | 65 |
| Mantel-Haenszel - with continuity/treatment arm correction | 1.08 [0.66 - 1.78] | 0.75 | 0 [0 - 29] | 0.00 [0.00 - 0.00] | 65 |
| Mantel-Haenszel - risk difference | 0.00 [0.00 - 0.01] | 0.70 | 0 [0 - 29] | 0.00 [0.00 - 0.00] | 65 |
| Arcsine square root transformed risk difference | 0.01 [-0.02 - 0.04] | 0.37 | 0 [ - ] | 0.00 [ - ] | 65 |

Table S11: Suicides: Meta analytic results, treatment phase

| Analysis | OR [95% CI] | p | I2 [95% CI] | T2 [95%CI] | number of studies |
|---|---|---|---|---|---|
| Exact (Liu et al. 2012) | 2.38 [0 - Inf] | | | | 65 |
| Bayesian - Uninformative prior (delta = 250) | 1.05 [0.28 - 4.13] | | | 1.33 [1.01 - 2.61] | 65 |
| Bayesian - Informative prior (delta = 15) | 1.04 [0.31 - 3.57] | | | 1.32 [1.01 - 2.57] | 65 |
| Binomial-normal GLMM, fixed intercept | 1.95 [0.17, 21.77] | | | 0.00 | |
| Binomial-normal GLMM, fixed intercept, alternative parameterization | 1.95 [0.17, 21.77] | | | 0.00 | |
| Binomial-normal GLMM, random intercept | 1.90 [0.17, 21.38] | | | 0.00 | |
| Binomial-normal GLMM, random intercept, alternative parameterization | 1.90 [0.17, 21.38] | | | 0.00 | |
| Hypergeometric-normal GLMM (Nchg) | 1.93 [0.17, 21.28] | | | 0.00 | |
| Conditional binomial GLMM (Condbin) | 1.93 [0.14, 21.28] | | | 0.00 | |
| Beta-binomial model | 1.58 [0.17, 17.43] | | | | |
| Peto | 1.96 [0.20 - 18.93] | 0.56 | 0 [NA - NA] | 0.00 [NA - NA] | 2 |
| Mantel-Haenszel - with continuity correction | 1.03 [0.63 - 1.67] | 0.92 | 0 [0 - 29] | 0.00 [0.00 - 0.00] | 65 |
| Mantel-Haenszel - with continuity/treatment arm correction | 1.03 [0.63 - 1.67] | 0.92 | 0 [0 - 29] | 0.00 [0.00 - 0.00] | 65 |
| Mantel-Haenszel - risk difference | 0.00 [0.00 - 0.00] | 0.90 | 0 [0 - 29] | 0.00 [0.00 - 0.00] | 65 |
| Arcsine square root transformed risk difference | 0.00 [-0.02 - 0.03] | 0.75 | 0 [ - ] | 0.00 [ - ] | 65 |

Table S12: Suicides: Meta analytic results, follow-up phase

| Analysis | OR [95% CI] | p | I2 [95% CI] | T2 [95%CI] | number of studies |
|---|---|---|---|---|---|
| Exact (Liu et al. 2012) | NA [NA - NA] | | | | |
| Bayesian - Uninformative prior (delta = 250) | 1.73 [0.35 - 10.33] | | | 1.35 [1.01 - 2.74] | 44 |
| Bayesian - Informative prior (delta = 15) | 1.52 [0.37 - 6.81] | | | 1.36 [1.01 - 2.76] | 44 |
| Binomial-normal GLMM, fixed intercept | | | | | |
| Binomial-normal GLMM, fixed intercept, alternative parameterization | | | | | |
| Binomial-normal GLMM, random intercept | | | | | |
| Binomial-normal GLMM, random intercept, alternative parameterization | | | | | |
| Hypergeometric-normal GLMM (Nchg) | | | | | |
| Conditional binomial GLMM (Condbin) | | | | | |
| Beta-binomial model | | | | | |
| Peto | 7.53 [0.47 - 120.96] | 0.15 | 0 [NA - NA] | 0.00 [NA - NA] | 2 |
| Mantel-Haenszel - with continuity correction | 0.93 [0.51 - 1.68] | 0.81 | 0 [0 - 35] | 0.00 [0.00 - 0.00] | 44 |
| Mantel-Haenszel - with continuity/treatment arm correction | 1.08 [0.59 - 1.98] | 0.80 | 0 [0 - 35] | 0.00 [0.00 - 0.00] | 44 |
| Mantel-Haenszel - risk difference | 0.00 [-0.01 - 0.01] | 0.76 | 0 [0 - 35] | 0.00 [0.00 - 0.00] | 44 |
| Arcsine square root transformed risk difference | 0.01 [-0.02 - 0.05] | 0.47 | 0 [ - ] | 0.00 [ - ] | 44 |

Note: For the treatment-phase and both phases combined, all meta-analytic methods are appropriate except Peto's method, because there are both single-arm-zero-events studies and double-arm-zero-events studies and the total event count in each arm is non-zero. For the follow-up phase, Peto's, GLMM, and beta-binomial methods are not recommended and/or not possible to conduct because there were no events in one arm (Xu et al., 2021)

#### Deaths

Table S13: Deaths: Meta analytic results, treatment- and follow-up phases combined

| Analysis | OR [95% CI] | p | I2 [95% CI] | T2 [95%CI] | number of studies |
|---|---|---|---|---|---|
| Exact (Liu et al. 2012) | 3.1 [0.07 - 1688450927.09] | | | | 65 |
| Bayesian - Uninformative prior (delta = 250) | 1.64 [0.56 - 5.27] | | | 1.33 [1.01 - 2.58] | 65 |
| Bayesian - Informative prior (delta = 15) | 1.55 [0.55 - 4.51] | | | 1.33 [1.01 - 2.59] | 65 |
| Binomial-normal GLMM, fixed intercept | 3.06 [0.61, 15.32] | | | 0.00 | |
| Binomial-normal GLMM, fixed intercept, alternative parameterization | 3.06 [0.53, 15.32] | | | 0.00 | |
| Binomial-normal GLMM, random intercept | 2.63 [0.53, 13.21] | | | 0.00 | |
| Binomial-normal GLMM, random intercept, alternative parameterization | 2.63 [0.61, 13.21] | | | 0.00 | |
| Hypergeometric-normal GLMM (Nchg) | 3.01 [0.61, 14.9] | | | 0.00 | |
| Conditional binomial GLMM (Condbin) | 3.01 [0.48, 14.9] | | | 0.00 | |
| Beta-binomial model | 2.37 [0.61, 11.76] | | | | |
| Peto | 2.73 [0.68 - 10.94] | 0.16 | 0 [0 - 71] | 0.00 [0.00 - 2.92] | 7 |
| Mantel-Haenszel - with continuity correction | 0.90 [0.56 - 1.46] | 0.67 | 0 [0 - 29] | 0.00 [0.00 - 0.00] | 65 |
| Mantel-Haenszel - with continuity/treatment arm correction | 1.11 [0.68 - 1.81] | 0.68 | 0 [0 - 29] | 0.00 [0.00 - 0.00] | 65 |
| Mantel-Haenszel - risk difference | 0.00 [0.00 - 0.01] | 0.69 | 0 [0 - 29] | 0.00 [0.00 - 0.00] | 65 |
| Arcsine square root transformed risk difference | 0.01 [-0.01 - 0.04] | 0.33 | 0 [ - ] | 0.00 [ - ] | 65 |

Table S14: Deaths: Meta analytic results, treatment phase

| Analysis | OR [95% CI] | p | I2 [95% CI] | T2 [95%CI] | number of studies |
|---|---|---|---|---|---|
| Exact (Liu et al. 2012) | 1.37 [0 - 1888341390.27] | | | | 65 |
| Bayesian - Uninformative prior (delta = 250) | 0.87 [0.24 - 3.08] | | | 1.33 [1.01 - 2.59] | 65 |
| Bayesian - Informative prior (delta = 15) | 0.88 [0.27 - 2.87] | | | 1.33 [1.01 - 2.59] | 65 |
| Binomial-normal GLMM, fixed intercept | 0.97 [0.14, 7.01] | | | 0.00 | |
| Binomial-normal GLMM, fixed intercept, alternative parameterization | 0.97 [0.13, 7.01] | | | 0.00 | |
| Binomial-normal GLMM, random intercept | 0.94 [0.13, 6.79] | | | 0.00 | |
| Binomial-normal GLMM, random intercept, alternative parameterization | 0.94 [0.14, 6.79] | | | 0.00 | |
| Hypergeometric-normal GLMM (Nchg) | 0.97 [0.14, 6.91] | | | 0.00 | |
| Conditional binomial GLMM (Condbin) | 0.97 [0.11, 6.91] | | | 0.00 | |
| Beta-binomial model | 0.79 [0.14, 5.59] | | | | |
| Peto | 1.00 [0.14 - 7.13] | 1.00 | 0 [0 - 90] | 0.00 [0.00 - 10.00] | 3 |
| Mantel-Haenszel - with continuity correction | 1.00 [0.62 - 1.62] | 1.00 | 0 [0 - 29] | 0.00 [0.00 - 0.00] | 65 |
| Mantel-Haenszel - with continuity/treatment arm correction | 1.00 [0.62 - 1.62] | 1.00 | 0 [0 - 29] | 0.00 [0.00 - 0.00] | 65 |
| Mantel-Haenszel - risk difference | 0.00 [0.00 - 0.00] | 0.92 | 0 [0 - 29] | 0.00 [0.00 - 0.00] | 65 |
| Arcsine square root transformed risk difference | 0.00 [-0.03 - 0.03] | 0.87 | 0 [ - ] | 0.00 [ - ] | 65 |

Table S15: Deaths: Meta analytic results, follow-up phase

| Analysis | OR [95% CI] | p | I2 [95% CI] | T2 [95%CI] | number of studies |
|---|---|---|---|---|---|
| Exact (Liu et al. 2012) | NA [NA - NA] | | | | |
| Bayesian - Uninformative prior (delta = 250) | 1.73 [0.35 - 10.33] | | | 1.35 [1.01 - 2.74] | 44 |
| Bayesian - Informative prior (delta = 15) | 1.52 [0.37 - 6.81] | | | 1.36 [1.01 - 2.76] | 44 |
| Binomial-normal GLMM, fixed intercept | | | | | |
| Binomial-normal GLMM, fixed intercept, alternative parameterization | | | | | |
| Binomial-normal GLMM, random intercept | | | | | |
| Binomial-normal GLMM, random intercept, alternative parameterization | | | | | |
| Hypergeometric-normal GLMM (Nchg) | | | | | |
| Conditional binomial GLMM (Condbin) | | | | | |
| Beta-binomial model | | | | | |
| Peto | 7.53 [0.47 - 120.96] | 0.15 | 0 [NA - NA] | 0.00 [NA - NA] | 2 |
| Mantel-Haenszel - with continuity correction | 0.93 [0.51 - 1.68] | 0.81 | 0 [0 - 35] | 0.00 [0.00 - 0.00] | 44 |
| Mantel-Haenszel - with continuity/treatment arm correction | 1.08 [0.59 - 1.98] | 0.80 | 0 [0 - 35] | 0.00 [0.00 - 0.00] | 44 |
| Mantel-Haenszel - risk difference | 0.00 [-0.01 - 0.01] | 0.76 | 0 [0 - 35] | 0.00 [0.00 - 0.00] | 44 |
| Arcsine square root transformed risk difference | 0.01 [-0.02 - 0.05] | 0.47 | 0 [ - ] | 0.00 [ - ] | 44 |

Note: For the treatment-phase and both phases combined, all meta-analytic methods are appropriate except Peto's method, because there are both single-arm-zero-events studies and double-arm-zero-events studies and the total event count in each arm is non-zero. For the follow-up phase, Peto's, GLMM, and beta-binomial methods are not recommended and/or not possible to conduct because there were no events in one arm (Xu et al., 2021)

##### Frequencies of suicidal behavior incidences

Table S16: Frequencies of Suicidal Behavior

| Outcome | Phase | Arm | N.. | N |
|---|---|---|---|---|
| Suicides and suicide attempts | Treatment | Drug | 9 (0.33) | 2742 |
| | | Placebo | 9 (0.41) | 2201 |
| | FU | Drug | 16 (1.06) | 1513 |
| | | Placebo | 18 (1.42) | 1270 |
| | Treatment + FU | Drug | 25 (0.91) | 2742 |
| | | Placebo | 27 (1.23) | 2201 |
| Suicide attempts | Treatment | Drug | 7 (0.26) | 2742 |
| | | Placebo | 8 (0.36) | 2201 |
| | FU | Drug | 14 (0.93) | 1513 |
| | | Placebo | 18 (1.42) | 1270 |
| | Treatment + FU | Drug | 21 (0.77) | 2742 |
| | | Placebo | 26 (1.18) | 2201 |
| Suicides | Treatment | Drug | 2 (0.07) | 2792 |
| | | Placebo | 1 (0.05) | 2201 |
| | FU | Drug | 2 (0.13) | 1563 |
| | | Placebo | 0 (0.00) | 1269 |
| | Treatment + FU | Drug | 4 (0.14) | 2792 |
| | | Placebo | 1 (0.05) | 2201 |
| Deaths | Treatment | Drug | 2 (0.07) | 2792 |
| | | Placebo | 2 (0.09) | 2201 |
| | FU | Drug | 4 (0.26) | 1563 |
| | | Placebo | 0 (0.00) | 1269 |
| | Treatment + FU | Drug | 6 (0.21) | 2792 |
| | | Placebo | 2 (0.09) | 2201 |

##### Suicide Ideation

#### Table S17: Suicide Ideation: Meta Analytic Results, Main Results

| Result | up to 4h | >12h up to 24h | >24h up to 72h | >72h up to 2w | >2w up to 4w | >4w | End of Treatment |
|---|---|---|---|---|---|---|---|
| SMD [95%-CI] | -0.40 [-0.63 - -0.16] | -0.35 [-0.51 - -0.19] | -0.16 [-0.28 - -0.04] | -0.16 [-0.26 - -0.06] | -0.14 [-0.27 - 0.00] | -0.01 [-0.20 - 0.17] | -0.24 [-0.35 - -0.13] |
| p | < 0.01 | < 0.001 | < 0.01 | < 0.01 | 0.05 | 0.87 | < 0.001 |
| Heterogeneity | I2=52%, T2=0.2, p<0.01 | I2=12%, T2=0.06, p=0.29 | I2=17%, T2=0.04, p=0.21 | I2=0%, T2=0.03, p=0.90 | I2=25%, T2=0.04, p=0.16 | I2=8%, T2=0.02, p=0.37 | I2=24%, T2=0.05, p=0.10 |
| Number of trials | 23 | 25 | 29 | 36 | 19 | 9 | 38 |
| Number of observations | 1261 | 1435 | 2529 | 2858 | 2159 | 792 | 2827 |
| Small study effect p-Value of Egger's regresson test | 0.63 | 0.75 | 0.17 | 0.50 | 0.88 | too few studies | 0.02 |

#### Table S18: Suicide Ideation: Meta Analytic Results, Subgroup-Analyses

| Subgroup analysis | up to 4h | >12h up to 24h | >24h up to 72h | >72h up to 2w | >2w up to 4w | >4w | End of Treatment |
|---|---|---|---|---|---|---|---|
| Drug | | | | | | | |
| —Esketamine | -0.31 [-0.53 - -0.09]* | -0.23 [-0.43 - -0.02]* | -0.05 [-0.16 - 0.07] | -0.10 [-0.21 - 0.01] | -0.06 [-0.18 - 0.05] | 0.04 [-0.27 - 0.35] | -0.06 [-0.18 - 0.05] |
| —Ketamine | -0.41 [-0.69 - -0.13]* | -0.38 [-0.58 - -0.19]* | -0.32 [-0.52 - -0.12]* | -0.22 [-0.38 - -0.07]* | -0.26 [-0.54 - 0.02] | -0.10 [-0.32 - 0.13] | -0.41 [-0.56 - -0.25]* |
| — test of difference (p-value) | 0.09 | 0.05 | < 0.001 | 0.08 | < 0.01 | 0.33 | < 0.001 |
| Suicidality excl/incl criterion | | | | | | | |
| —Exclusion criterion or unknown | -0.32 [-0.69 - 0.06] | -0.24 [-0.48 - 0.00] | -0.06 [-0.19 - 0.07] | -0.14 [-0.27 - -0.01]* | -0.12 [-0.30 - 0.07] | 0.10 [-0.18 - 0.37] | -0.15 [-0.29 - -0.02]* |
| —Inclusion criterion | -0.44 [-0.74 - -0.13]* | -0.42 [-0.61 - -0.23]* | -0.31 [-0.52 - -0.10]* | -0.20 [-0.37 - -0.03]* | -0.17 [-0.38 - 0.04] | -0.09 [-0.32 - 0.13] | -0.37 [-0.56 - -0.18]* |
| — test of difference (p-value) | 0.58 | 0.32 | < 0.01 | 0.31 | 0.25 | 0.21 | < 0.01 |
| Doses | | | | | | | |
| —Repeated | | | | -0.15 [-0.26 - -0.03]* | -0.11 [-0.23 - 0.01] | -0.02 [-0.23 - 0.18] | |
| —Single | | | | -0.19 [-0.39 - 0.01] | -0.17 [-0.57 - 0.22] | 0.02 [-0.40 - 0.44] | |
| — test of difference (p-value) | | | | 0.53 | 0.16 | 0.88 | |
| Rating | | | | | | | |
| —Clinician | -0.38 [-0.59 - -0.17]* | -0.35 [-0.51 - -0.19]* | -0.14 [-0.25 - -0.03]* | -0.16 [-0.27 - -0.05]* | -0.15 [-0.30 - -0.01]* | -0.09 [-0.27 - 0.10] | -0.24 [-0.37 - -0.11]* |
| —Patient | -0.64 [-1.48 - 0.20] | -0.47 [-0.94 - -0.01]* | -0.44 [-1.00 - 0.11] | -0.18 [-0.50 - 0.13] | 0.05 [-0.45 - 0.56] | -0.01 [-0.41 - 0.40] | -0.33 [-0.63 - -0.03]* |
| — test of difference (p-value) | 0.82 | 0.49 | 0.15 | 0.79 | 0.42 | 0.74 | 0.43 |
| Placebo | | | | | | | |
| —Active | -0.24 [-0.62 - 0.14] | -0.43 [-0.70 - -0.16]* | -0.24 [-0.50 - 0.03] | -0.19 [-0.48 - 0.10] | 0.00 [-0.56 - 0.56] | -0.15 [-0.59 - 0.30] | -0.48 [-0.74 - -0.22]* |
| —Non-active | -0.48 [-0.77 - -0.19]* | -0.30 [-0.48 - -0.11]* | -0.15 [-0.29 - -0.01]* | -0.15 [-0.26 - -0.05]* | -0.14 [-0.28 - -0.01]* | 0.01 [-0.19 - 0.21] | -0.17 [-0.28 - -0.05]* |
| — test of difference (p-value) | 0.74 | 0.09 | 0.30 | 0.73 | 0.78 | 0.48 | < 0.001 |

#### Meta-Analytic Results - Forest-Plots

### Suicides and Suicide Attempts Combined

#### Treatment Phase

#### Peto's Method

| Study | Experimental Events | Experimental Total | Control Events | Control Total | OR | 95%-CI | Weight (common) | Weight (random) |
|---|---|---|---|---|---|---|---|---|
| Abbar 2022 | 1 | 73 | 0 | 73 | 7.39 | [0.15; 372.38] | 5.6% | 5.6% |
| Abdallah 2022 | 0 | 108 | 0 | 108 | | | 0.0% | 0.0% |
| Ahmed 2023 | 0 | 18 | 0 | 18 | | | 0.0% | 0.0% |
| Anderson 2017 | 0 | 40 | 1 | 40 | 0.14 | [0.00; 6.82] | 5.6% | 5.6% |
| Burger 2016 | 0 | 3 | 0 | 3 | | | 0.0% | 0.0% |
| Canuso 2018 | 0 | 35 | 0 | 35 | | | 0.0% | 0.0% |
| Chen 2023 | 2 | 126 | 0 | 126 | 7.45 | [0.46; 119.74] | 11.2% | 11.2% |
| Colla 2024 | 0 | 17 | 0 | 17 | | | 0.0% | 0.0% |
| Dakwar 2019 | 0 | 27 | 0 | 27 | | | 0.0% | 0.0% |
| Dakwar 2020 | 0 | 17 | 0 | 17 | | | 0.0% | 0.0% |
| Daly 2018 PA | 0 | 34 | 0 | 34 | | | 0.0% | 0.0% |
| Daly 2018 PB | 0 | 20 | 0 | 20 | | | 0.0% | 0.0% |
| Domany 2019 | 0 | 22 | 0 | 22 | | | 0.0% | 0.0% |
| Domany 2020 | 0 | 9 | 0 | 9 | | | 0.0% | 0.0% |
| Duek 2023 | 0 | 14 | 0 | 14 | | | 0.0% | 0.0% |
| Dwyer 2021 | 0 | 11 | 0 | 11 | | | 0.0% | 0.0% |
| Fava 2020 | 0 | 80 | 0 | 80 | | | 0.0% | 0.0% |
| Feder 2014 | 0 | 22 | 0 | 22 | | | 0.0% | 0.0% |
| Feder 2021 | 0 | 15 | 0 | 15 | | | 0.0% | 0.0% |
| Fedgchin 2019 | 0 | 231 | 0 | 231 | | | 0.0% | 0.0% |
| Fineberg 2023 | 0 | 10 | 0 | 10 | | | 0.0% | 0.0% |
| Fu 2020 | 1 | 113 | 1 | 113 | 1.00 | [0.06; 16.09] | 11.2% | 11.2% |
| George 2017 | 0 | 12 | 0 | 12 | | | 0.0% | 0.0% |
| Glue 2024b | 0 | 17 | 0 | 17 | | | 0.0% | 0.0% |
| Grabski 2022 | 0 | 48 | 0 | 48 | | | 0.0% | 0.0% |
| Grunebaum 2017 | 0 | 9 | 0 | 9 | | | 0.0% | 0.0% |
| Grunebaum 2018 | 0 | 40 | 0 | 40 | | | 0.0% | 0.0% |
| Ionescu 2020 | 3 | 114 | 3 | 114 | 1.00 | [0.20; 5.04] | 32.9% | 32.9% |
| Jones 2024 | 0 | 17 | 0 | 17 | | | 0.0% | 0.0% |
| Lapidus 2014 | 0 | 10 | 0 | 10 | | | 0.0% | 0.0% |
| Leal 2023 | 0 | 5 | 0 | 5 | | | 0.0% | 0.0% |
| Lii 2023 | 0 | 20 | 0 | 20 | | | 0.0% | 0.0% |
| Lijffijt 2021 | 0 | 20 | 0 | 20 | | | 0.0% | 0.0% |
| Loo 2016 | 0 | 9 | 0 | 9 | | | 0.0% | 0.0% |
| Loo 2023 | 0 | 89 | 2 | 89 | 0.13 | [0.01; 2.16] | 11.2% | 11.2% |
| Milak 2020 | 0 | 38 | 0 | 38 | | | 0.0% | 0.0% |
| Murrough 2013 | 0 | 47 | 0 | 47 | | | 0.0% | 0.0% |
| Murrough 2015 II | 0 | 12 | 0 | 12 | | | 0.0% | 0.0% |
| Ochs-Ross 2020 | 0 | 72 | 0 | 72 | | | 0.0% | 0.0% |
| Ohtani 2024 | 0 | 17 | 0 | 17 | | | 0.0% | 0.0% |
| Pattanaseri 2024 | 0 | 11 | 0 | 11 | | | 0.0% | 0.0% |
| Phillips 2019 | 0 | 22 | 0 | 22 | | | 0.0% | 0.0% |
| Popova 2019 | 0 | 115 | 0 | 115 | | | 0.0% | 0.0% |
| Price 2022 | 0 | 53 | 0 | 53 | | | 0.0% | 0.0% |
| Ray-Griffith 2017 | 0 | 8 | 0 | 8 | | | 0.0% | 0.0% |
| Rodriguez 2013 | 0 | 8 | 0 | 8 | | | 0.0% | 0.0% |
| Schiweck 2021 | 0 | 21 | 0 | 21 | | | 0.0% | 0.0% |
| Shiroma 2020 | 0 | 29 | 0 | 29 | | | 0.0% | 0.0% |
| Singh 2016 2x doses | 1 | 18 | 0 | 18 | 7.39 | [0.15; 372.38] | 5.6% | 5.6% |
| Singh 2016 3x doses | 0 | 17 | 0 | 17 | | | 0.0% | 0.0% |
| Singh 2016 II | 0 | 20 | 0 | 20 | | | 0.0% | 0.0% |
| Smith-Apeldoorn 2024 | 1 | 57 | 1 | 57 | 1.00 | [0.06; 16.19] | 11.1% | 11.1% |
| Sos 2013 | 0 | 11 | 0 | 11 | | | 0.0% | 0.0% |
| Su 2017 | 0 | 47 | 0 | 47 | | | 0.0% | 0.0% |
| Su 2023 | 0 | 42 | 0 | 42 | | | 0.0% | 0.0% |
| Sumner 2019 | 0 | 17 | 0 | 17 | | | 0.0% | 0.0% |
| Takahashi 2021 | 0 | 122 | 0 | 122 | | | 0.0% | 0.0% |
| Taylor 2018 | 0 | 9 | 0 | 9 | | | 0.0% | 0.0% |
| Tiger 2020 | 0 | 20 | 0 | 20 | | | 0.0% | 0.0% |
| Wang 2024 | 0 | 182 | 0 | 182 | | | 0.0% | 0.0% |
| Xiao 2023 | 0 | 15 | 0 | 15 | | | 0.0% | 0.0% |
| Yang 2023 | 0 | 200 | 0 | 200 | | | 0.0% | 0.0% |
| Zarate 2006 | 0 | 9 | 0 | 9 | | | 0.0% | 0.0% |
| Zarate 2012 | 0 | 21 | 0 | 21 | | | 0.0% | 0.0% |
| Zhou 2024 | 0 | 27 | 1 | 27 | 0.14 | [0.00; 6.82] | 5.6% | 5.6% |
| **Common effect model** | | **2742** | | **2742** | **1.00** | **[0.40; 2.53]** | **100.0%** | **.** |
| **Random effects model** | | | | | **1.00** | **[0.40; 2.53]** | **.** | **100.0%** |

Heterogeneity: $I^2 = 0.2\%$, $\tau^2 < 0.0001$, $p = 0.4316$

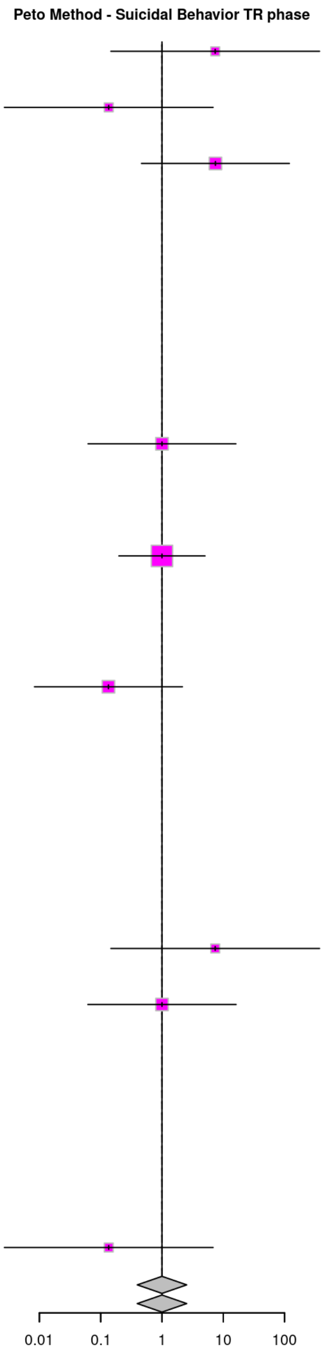

#### Maentel Mantel-Haenszel with CC

| Study | Experimental Events | Experimental Total | Control Events | Control Total | OR | 95%-CI | Weight (common) | Weight (random) |
|---|---|---|---|---|---|---|---|---|
| Abbar 2022 | 1 | 73 | 0 | 73 | 3.04 | [0.12; 75.89] | 1.3% | 2.0% |
| Abdallah 2022 | 0 | 108 | 0 | 108 | 1.00 | [0.02; 50.85] | 1.3% | 1.4% |
| Ahmed 2023 | 0 | 18 | 0 | 18 | 1.00 | [0.02; 53.12] | 1.2% | 1.3% |
| Anderson 2017 | 0 | 40 | 1 | 40 | 0.33 | [0.01; 8.22] | 3.8% | 2.0% |
| Burger 2016 | 0 | 3 | 0 | 3 | 1.00 | [0.02; 66.06] | 1.1% | 1.2% |
| Canuso 2018 | 0 | 35 | 0 | 35 | 1.00 | [0.02; 51.80] | 1.3% | 1.3% |
| Chen 2023 | 2 | 126 | 0 | 126 | 5.08 | [0.24; 106.89] | 1.3% | 2.3% |
| Colla 2024 | 0 | 17 | 0 | 17 | 1.00 | [0.02; 53.28] | 1.2% | 1.3% |
| Dakwar 2019 | 0 | 27 | 0 | 27 | 1.00 | [0.02; 52.22] | 1.3% | 1.3% |
| Dakwar 2020 | 0 | 17 | 0 | 17 | 1.00 | [0.02; 53.28] | 1.2% | 1.3% |
| Daly 2018 PA | 0 | 34 | 0 | 34 | 1.00 | [0.02; 51.84] | 1.3% | 1.3% |
| Daly 2018 PB | 0 | 20 | 0 | 20 | 1.00 | [0.02; 52.85] | 1.3% | 1.3% |
| Domany 2019 | 0 | 22 | 0 | 22 | 1.00 | [0.02; 52.63] | 1.3% | 1.3% |
| Domany 2020 | 0 | 9 | 0 | 9 | 1.00 | [0.02; 55.80] | 1.2% | 1.3% |
| Duek 2023 | 0 | 14 | 0 | 14 | 1.00 | [0.02; 53.89] | 1.2% | 1.3% |
| Dwyer 2021 | 0 | 11 | 0 | 11 | 1.00 | [0.02; 54.83] | 1.2% | 1.3% |
| Fava 2020 | 0 | 80 | 0 | 80 | 1.00 | [0.02; 51.01] | 1.3% | 1.4% |
| Feder 2014 | 0 | 22 | 0 | 22 | 1.00 | [0.02; 52.63] | 1.3% | 1.3% |
| Feder 2021 | 0 | 15 | 0 | 15 | 1.00 | [0.02; 53.66] | 1.2% | 1.3% |
| Fedgchin 2019 | 0 | 231 | 0 | 231 | 1.00 | [0.02; 50.61] | 1.3% | 1.4% |
| Fineberg 2023 | 0 | 10 | 0 | 10 | 1.00 | [0.02; 55.27] | 1.2% | 1.3% |
| Fu 2020 | 1 | 113 | 1 | 113 | 1.00 | [0.06; 16.19] | 2.5% | 2.7% |
| George 2017 | 0 | 12 | 0 | 12 | 1.00 | [0.02; 54.46] | 1.2% | 1.3% |
| Glue 2024b | 0 | 17 | 0 | 17 | 1.00 | [0.02; 53.28] | 1.2% | 1.3% |
| Grabski 2022 | 0 | 48 | 0 | 48 | 1.00 | [0.02; 51.42] | 1.3% | 1.3% |
| Grunebaum 2017 | 0 | 9 | 0 | 9 | 1.00 | [0.02; 55.80] | 1.2% | 1.3% |
| Grunebaum 2018 | 0 | 40 | 0 | 40 | 1.00 | [0.02; 51.63] | 1.3% | 1.3% |
| Ionescu 2020 | 3 | 114 | 3 | 114 | 1.00 | [0.20; 5.06] | 7.5% | 8.0% |
| Jones 2024 | 0 | 17 | 0 | 17 | 1.00 | [0.02; 53.28] | 1.2% | 1.3% |
| Lapidus 2014 | 0 | 10 | 0 | 10 | 1.00 | [0.02; 55.27] | 1.2% | 1.3% |
| Leal 2023 | 0 | 5 | 0 | 5 | 1.00 | [0.02; 59.99] | 1.2% | 1.2% |
| Lii 2023 | 0 | 20 | 0 | 20 | 1.00 | [0.02; 52.85] | 1.3% | 1.3% |
| Lijffijt 2021 | 0 | 20 | 0 | 20 | 1.00 | [0.02; 52.85] | 1.3% | 1.3% |
| Loo 2016 | 0 | 9 | 0 | 9 | 1.00 | [0.02; 55.80] | 1.2% | 1.3% |
| Loo 2023 | 0 | 89 | 2 | 89 | 0.20 | [0.01; 4.13] | 6.4% | 2.3% |
| Milak 2020 | 0 | 38 | 0 | 38 | 1.00 | [0.02; 51.69] | 1.3% | 1.3% |
| Murrough 2013 | 0 | 47 | 0 | 47 | 1.00 | [0.02; 51.44] | 1.3% | 1.3% |
| Murrough 2015 II | 0 | 12 | 0 | 12 | 1.00 | [0.02; 54.46] | 1.2% | 1.3% |
| Ochs-Ross 2020 | 0 | 72 | 0 | 72 | 1.00 | [0.02; 51.08] | 1.3% | 1.4% |
| Ohtani 2024 | 0 | 17 | 0 | 17 | 1.00 | [0.02; 53.28] | 1.2% | 1.3% |
| Pattanaseri 2024 | 0 | 11 | 0 | 11 | 1.00 | [0.02; 54.83] | 1.2% | 1.3% |
| Phillips 2019 | 0 | 22 | 0 | 22 | 1.00 | [0.02; 52.63] | 1.3% | 1.3% |
| Popova 2019 | 0 | 115 | 0 | 115 | 1.00 | [0.02; 50.83] | 1.3% | 1.4% |
| Price 2022 | 0 | 53 | 0 | 53 | 1.00 | [0.02; 51.33] | 1.3% | 1.4% |
| Ray-Griffith 2017 | 0 | 8 | 0 | 8 | 1.00 | [0.02; 56.46] | 1.2% | 1.3% |
| Rodriguez 2013 | 0 | 8 | 0 | 8 | 1.00 | [0.02; 56.46] | 1.2% | 1.3% |
| Schiweck 2021 | 0 | 21 | 0 | 21 | 1.00 | [0.02; 52.73] | 1.3% | 1.3% |
| Shiroma 2020 | 0 | 29 | 0 | 29 | 1.00 | [0.02; 52.09] | 1.3% | 1.3% |
| Singh 2016 2x doses | 1 | 18 | 0 | 18 | 3.17 | [0.12; 83.17] | 1.2% | 2.0% |
| Singh 2016 3x doses | 0 | 17 | 0 | 17 | 1.00 | [0.02; 53.28] | 1.2% | 1.3% |
| Singh 2016 II | 0 | 20 | 0 | 20 | 1.00 | [0.02; 52.85] | 1.3% | 1.3% |
| Smith-Apeldoorn 2024 | 1 | 57 | 1 | 57 | 1.00 | [0.06; 16.39] | 2.5% | 2.7% |
| Sos 2013 | 0 | 11 | 0 | 11 | 1.00 | [0.02; 54.83] | 1.2% | 1.3% |
| Su 2017 | 0 | 47 | 0 | 47 | 1.00 | [0.02; 51.44] | 1.3% | 1.3% |
| Su 2023 | 0 | 42 | 0 | 42 | 1.00 | [0.02; 51.57] | 1.3% | 1.3% |
| Sumner 2019 | 0 | 17 | 0 | 17 | 1.00 | [0.02; 53.28] | 1.2% | 1.3% |
| Takahashi 2021 | 0 | 122 | 0 | 122 | 1.00 | [0.02; 50.80] | 1.3% | 1.4% |
| Taylor 2018 | 0 | 9 | 0 | 9 | 1.00 | [0.02; 55.80] | 1.2% | 1.3% |
| Tiger 2020 | 0 | 20 | 0 | 20 | 1.00 | [0.02; 52.85] | 1.3% | 1.3% |
| Wang 2024 | 0 | 182 | 0 | 182 | 1.00 | [0.02; 50.67] | 1.3% | 1.4% |
| Xiao 2023 | 0 | 15 | 0 | 15 | 1.00 | [0.02; 53.66] | 1.2% | 1.3% |
| Yang 2023 | 0 | 200 | 0 | 200 | 1.00 | [0.02; 50.64] | 1.3% | 1.4% |
| Zarate 2006 | 0 | 9 | 0 | 9 | 1.00 | [0.02; 55.80] | 1.2% | 1.3% |
| Zarate 2012 | 0 | 21 | 0 | 21 | 1.00 | [0.02; 52.73] | 1.3% | 1.3% |
| Zhou 2024 | 0 | 27 | 1 | 27 | 0.32 | [0.01; 8.24] | 3.8% | 2.0% |
| **Common effect model** | | **2742** | | **2742** | **1.00** | **[0.64; 1.56]** | **100.0%** | **.** |
| **Random effects model** | | | | | **1.00** | **[0.63; 1.58]** | **.** | **100.0%** |

Heterogeneity: $I^2 = 0.0\%$, $\tau^2 = 0$, $p = 1.0000$

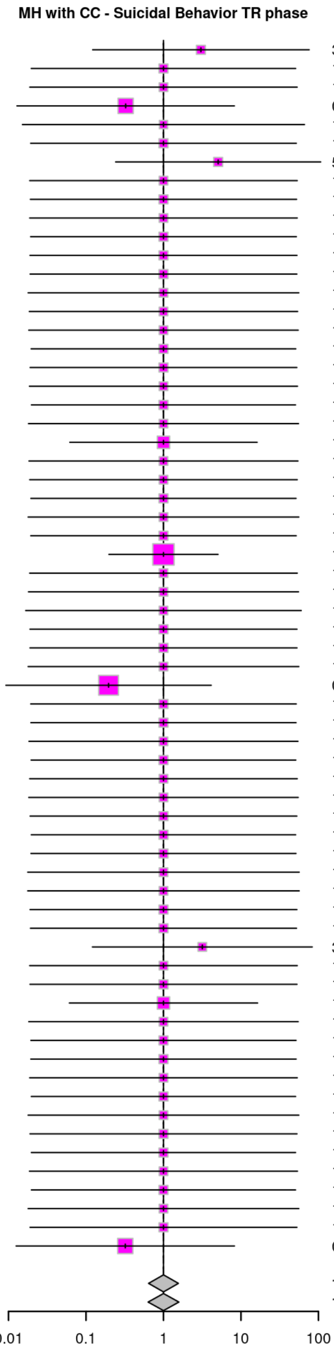

#### Maentel Mantel-Haenszel with Treatment-Arm CC

| Study | Experimental Events | Experimental Total | Control Events | Control Total | OR | 95%-CI | Weight (common) | Weight (random) |
|---|---|---|---|---|---|---|---|---|
| Abbar 2022 | 1 | 73 | 0 | 73 | 3.04 | [0.12; 75.89] | 1.3% | 2.0% |
| Abdallah 2022 | 0 | 108 | 0 | 108 | 1.00 | [0.02; 50.85] | 1.3% | 1.4% |
| Ahmed 2023 | 0 | 18 | 0 | 18 | 1.00 | [0.02; 53.12] | 1.2% | 1.3% |
| Anderson 2017 | 0 | 40 | 1 | 40 | 0.33 | [0.01; 8.22] | 3.8% | 2.0% |
| Burger 2016 | 0 | 3 | 0 | 3 | 1.00 | [0.02; 66.06] | 1.1% | 1.2% |
| Canuso 2018 | 0 | 35 | 0 | 35 | 1.00 | [0.02; 51.80] | 1.3% | 1.3% |
| Chen 2023 | 2 | 126 | 0 | 126 | 5.08 | [0.24; 106.89] | 1.3% | 2.3% |
| Colla 2024 | 0 | 17 | 0 | 17 | 1.00 | [0.02; 53.28] | 1.2% | 1.3% |
| Dakwar 2019 | 0 | 27 | 0 | 27 | 1.00 | [0.02; 52.22] | 1.3% | 1.3% |
| Dakwar 2020 | 0 | 17 | 0 | 17 | 1.00 | [0.02; 53.28] | 1.2% | 1.3% |
| Daly 2018 PA | 0 | 34 | 0 | 34 | 1.00 | [0.02; 51.84] | 1.3% | 1.3% |
| Daly 2018 PB | 0 | 20 | 0 | 20 | 1.00 | [0.02; 52.85] | 1.3% | 1.3% |
| Domany 2019 | 0 | 22 | 0 | 22 | 1.00 | [0.02; 52.63] | 1.3% | 1.3% |
| Domany 2020 | 0 | 9 | 0 | 9 | 1.00 | [0.02; 55.80] | 1.2% | 1.3% |
| Duek 2023 | 0 | 14 | 0 | 14 | 1.00 | [0.02; 53.89] | 1.2% | 1.3% |
| Dwyer 2021 | 0 | 11 | 0 | 11 | 1.00 | [0.02; 54.83] | 1.2% | 1.3% |
| Fava 2020 | 0 | 80 | 0 | 80 | 1.00 | [0.02; 51.01] | 1.3% | 1.4% |
| Feder 2014 | 0 | 22 | 0 | 22 | 1.00 | [0.02; 52.63] | 1.3% | 1.3% |
| Feder 2021 | 0 | 15 | 0 | 15 | 1.00 | [0.02; 53.66] | 1.2% | 1.3% |
| Fedgchin 2019 | 0 | 231 | 0 | 231 | 1.00 | [0.02; 50.61] | 1.3% | 1.4% |
| Fineberg 2023 | 0 | 10 | 0 | 10 | 1.00 | [0.02; 55.27] | 1.2% | 1.3% |
| Fu 2020 | 1 | 113 | 1 | 113 | 1.00 | [0.06; 16.19] | 2.5% | 2.7% |
| George 2017 | 0 | 12 | 0 | 12 | 1.00 | [0.02; 54.46] | 1.2% | 1.3% |
| Glue 2024b | 0 | 17 | 0 | 17 | 1.00 | [0.02; 53.28] | 1.2% | 1.3% |
| Grabski 2022 | 0 | 48 | 0 | 48 | 1.00 | [0.02; 51.42] | 1.3% | 1.3% |
| Grunebaum 2017 | 0 | 9 | 0 | 9 | 1.00 | [0.02; 55.80] | 1.2% | 1.3% |
| Grunebaum 2018 | 0 | 40 | 0 | 40 | 1.00 | [0.02; 51.63] | 1.3% | 1.3% |
| Ionescu 2020 | 3 | 114 | 3 | 114 | 1.00 | [0.20; 5.06] | 7.5% | 8.0% |
| Jones 2024 | 0 | 17 | 0 | 17 | 1.00 | [0.02; 53.28] | 1.2% | 1.3% |
| Lapidus 2014 | 0 | 10 | 0 | 10 | 1.00 | [0.02; 55.27] | 1.2% | 1.3% |
| Leal 2023 | 0 | 5 | 0 | 5 | 1.00 | [0.02; 59.99] | 1.2% | 1.2% |
| Lii 2023 | 0 | 20 | 0 | 20 | 1.00 | [0.02; 52.85] | 1.3% | 1.3% |
| Lijffijt 2021 | 0 | 20 | 0 | 20 | 1.00 | [0.02; 52.85] | 1.3% | 1.3% |
| Loo 2016 | 0 | 9 | 0 | 9 | 1.00 | [0.02; 55.80] | 1.2% | 1.3% |
| Loo 2023 | 0 | 89 | 2 | 89 | 0.20 | [0.01; 4.13] | 6.4% | 2.3% |
| Milak 2020 | 0 | 38 | 0 | 38 | 1.00 | [0.02; 51.69] | 1.3% | 1.3% |
| Murrough 2013 | 0 | 47 | 0 | 47 | 1.00 | [0.02; 51.44] | 1.3% | 1.3% |
| Murrough 2015 II | 0 | 12 | 0 | 12 | 1.00 | [0.02; 54.46] | 1.2% | 1.3% |
| Ochs-Ross 2020 | 0 | 72 | 0 | 72 | 1.00 | [0.02; 51.08] | 1.3% | 1.4% |
| Ohtani 2024 | 0 | 17 | 0 | 17 | 1.00 | [0.02; 53.28] | 1.2% | 1.3% |
| Pattanaseri 2024 | 0 | 11 | 0 | 11 | 1.00 | [0.02; 54.83] | 1.2% | 1.3% |
| Phillips 2019 | 0 | 22 | 0 | 22 | 1.00 | [0.02; 52.63] | 1.3% | 1.3% |
| Popova 2019 | 0 | 115 | 0 | 115 | 1.00 | [0.02; 50.83] | 1.3% | 1.4% |
| Price 2022 | 0 | 53 | 0 | 53 | 1.00 | [0.02; 51.33] | 1.3% | 1.4% |
| Ray-Griffith 2017 | 0 | 8 | 0 | 8 | 1.00 | [0.02; 56.46] | 1.2% | 1.3% |
| Rodriguez 2013 | 0 | 8 | 0 | 8 | 1.00 | [0.02; 56.46] | 1.2% | 1.3% |
| Schiweck 2021 | 0 | 21 | 0 | 21 | 1.00 | [0.02; 52.73] | 1.3% | 1.3% |
| Shiroma 2020 | 0 | 29 | 0 | 29 | 1.00 | [0.02; 52.09] | 1.3% | 1.3% |
| Singh 2016 2x doses | 1 | 18 | 0 | 18 | 3.17 | [0.12; 83.17] | 1.2% | 2.0% |
| Singh 2016 3x doses | 0 | 17 | 0 | 17 | 1.00 | [0.02; 53.28] | 1.2% | 1.3% |
| Singh 2016 II | 0 | 20 | 0 | 20 | 1.00 | [0.02; 52.85] | 1.3% | 1.3% |
| Smith-Apeldoorn 2024 | 1 | 57 | 1 | 57 | 1.00 | [0.06; 16.39] | 2.5% | 2.7% |
| Sos 2013 | 0 | 11 | 0 | 11 | 1.00 | [0.02; 54.83] | 1.2% | 1.3% |
| Su 2017 | 0 | 47 | 0 | 47 | 1.00 | [0.02; 51.44] | 1.3% | 1.3% |
| Su 2023 | 0 | 42 | 0 | 42 | 1.00 | [0.02; 51.57] | 1.3% | 1.3% |
| Sumner 2019 | 0 | 17 | 0 | 17 | 1.00 | [0.02; 53.28] | 1.2% | 1.3% |
| Takahashi 2021 | 0 | 122 | 0 | 122 | 1.00 | [0.02; 50.80] | 1.3% | 1.4% |
| Taylor 2018 | 0 | 9 | 0 | 9 | 1.00 | [0.02; 55.80] | 1.2% | 1.3% |
| Tiger 2020 | 0 | 20 | 0 | 20 | 1.00 | [0.02; 52.85] | 1.3% | 1.3% |
| Wang 2024 | 0 | 182 | 0 | 182 | 1.00 | [0.02; 50.67] | 1.3% | 1.4% |
| Xiao 2023 | 0 | 15 | 0 | 15 | 1.00 | [0.02; 53.66] | 1.2% | 1.3% |
| Yang 2023 | 0 | 200 | 0 | 200 | 1.00 | [0.02; 50.64] | 1.3% | 1.4% |
| Zarate 2006 | 0 | 9 | 0 | 9 | 1.00 | [0.02; 55.80] | 1.2% | 1.3% |
| Zarate 2012 | 0 | 21 | 0 | 21 | 1.00 | [0.02; 52.73] | 1.3% | 1.3% |
| Zhou 2024 | 0 | 27 | 1 | 27 | 0.32 | [0.01; 8.24] | 3.8% | 2.0% |
| **Common effect model** | | **2742** | | **2742** | **1.00** | **[0.64; 1.56]** | **100.0%** | **.** |
| **Random effects model** | | | | | **1.00** | **[0.63; 1.58]** | **.** | **100.0%** |

Heterogeneity: $I^2 = 0.0\%$, $\tau^2 = 0$, $p = 1.0000$

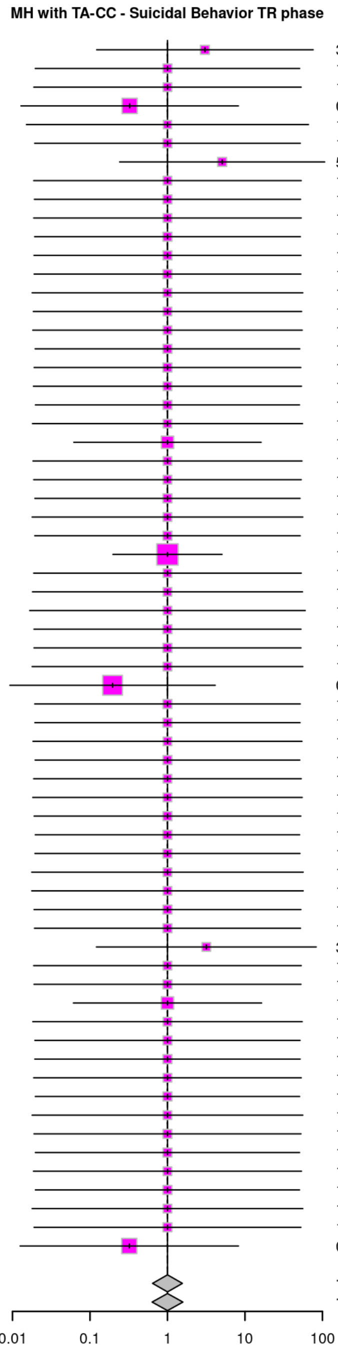

#### Maentel Mantel-Haenszel Risk-Difference

MH RD - Suicidal Behavior TR phase

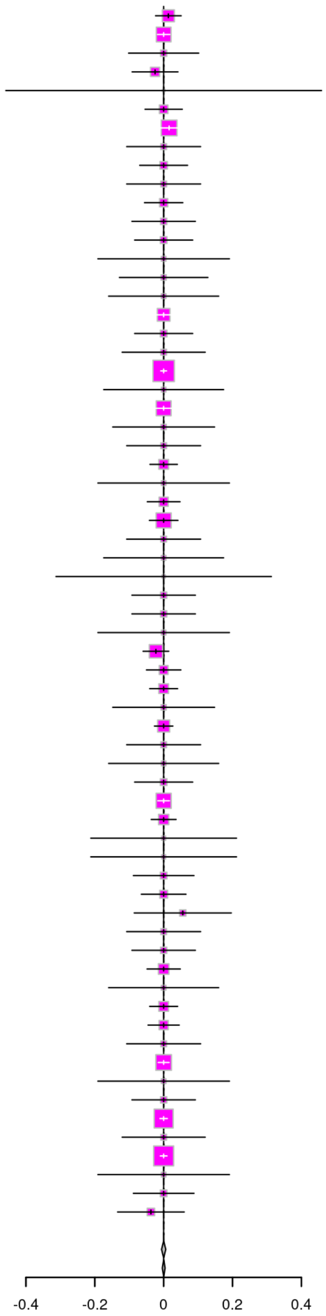

| Study | Experimental Events | Experimental Total | Control Events | Control Total | RD | 95%-CI | Weight (common) | Weight (random) |
|---|---|---|---|---|---|---|---|---|
| Abbar 2022 | 1 | 73 | 0 | 73 | 0.01 | [-0.02; 0.05] | 2.6% | 1.2% |
| Abdallah 2022 | 0 | 108 | 0 | 108 | 0.00 | [-0.02; 0.02] | 3.9% | 5.1% |
| Ahmed 2023 | 0 | 18 | 0 | 18 | 0.00 | [-0.10; 0.10] | 0.7% | 0.2% |
| Anderson 2017 | 0 | 40 | 1 | 40 | -0.03 | [-0.09; 0.04] | 1.5% | 0.4% |
| Burger 2016 | 0 | 3 | 0 | 3 | 0.00 | [-0.46; 0.46] | 0.1% | 0.0% |
| Canuso 2018 | 0 | 35 | 0 | 35 | 0.00 | [-0.05; 0.05] | 1.3% | 0.6% |
| Chen 2023 | 2 | 126 | 0 | 126 | 0.02 | [-0.01; 0.04] | 4.5% | 2.3% |
| Colla 2024 | 0 | 17 | 0 | 17 | 0.00 | [-0.11; 0.11] | 0.6% | 0.1% |
| Dakwar 2019 | 0 | 27 | 0 | 27 | 0.00 | [-0.07; 0.07] | 1.0% | 0.3% |
| Dakwar 2020 | 0 | 17 | 0 | 17 | 0.00 | [-0.11; 0.11] | 0.6% | 0.1% |
| Daly 2018 PA | 0 | 34 | 0 | 34 | 0.00 | [-0.06; 0.06] | 1.2% | 0.5% |
| Daly 2018 PB | 0 | 20 | 0 | 20 | 0.00 | [-0.09; 0.09] | 0.7% | 0.2% |
| Domany 2019 | 0 | 22 | 0 | 22 | 0.00 | [-0.08; 0.08] | 0.8% | 0.2% |
| Domany 2020 | 0 | 9 | 0 | 9 | 0.00 | [-0.19; 0.19] | 0.4% | 0.0% |
| Duek 2023 | 0 | 14 | 0 | 14 | 0.00 | [-0.13; 0.13] | 0.5% | 0.1% |
| Dwyer 2021 | 0 | 11 | 0 | 11 | 0.00 | [-0.16; 0.16] | 0.4% | 0.1% |
| Fava 2020 | 0 | 80 | 0 | 80 | 0.00 | [-0.02; 0.02] | 2.9% | 2.8% |
| Feder 2014 | 0 | 22 | 0 | 22 | 0.00 | [-0.08; 0.08] | 0.8% | 0.2% |
| Feder 2021 | 0 | 15 | 0 | 15 | 0.00 | [-0.12; 0.12] | 0.6% | 0.1% |
| Fedgchin 2019 | 0 | 231 | 0 | 231 | 0.00 | [-0.01; 0.01] | 8.3% | 23.2% |
| Fineberg 2023 | 0 | 10 | 0 | 10 | 0.00 | [-0.17; 0.17] | 0.4% | 0.1% |
| Fu 2020 | 1 | 113 | 1 | 113 | 0.00 | [-0.02; 0.02] | 4.0% | 2.8% |
| George 2017 | 0 | 12 | 0 | 12 | 0.00 | [-0.15; 0.15] | 0.5% | 0.1% |
| Glue 2024b | 0 | 17 | 0 | 17 | 0.00 | [-0.11; 0.11] | 0.6% | 0.1% |
| Grabski 2022 | 0 | 48 | 0 | 48 | 0.00 | [-0.04; 0.04] | 1.7% | 1.0% |
| Grunebaum 2017 | 0 | 9 | 0 | 9 | 0.00 | [-0.19; 0.19] | 0.4% | 0.0% |
| Grunebaum 2018 | 0 | 40 | 0 | 40 | 0.00 | [-0.05; 0.05] | 1.5% | 0.7% |
| Ionescu 2020 | 3 | 114 | 3 | 114 | 0.00 | [-0.04; 0.04] | 4.1% | 1.0% |
| Jones 2024 | 0 | 17 | 0 | 17 | 0.00 | [-0.11; 0.11] | 0.6% | 0.1% |
| Lapidus 2014 | 0 | 10 | 0 | 10 | 0.00 | [-0.17; 0.17] | 0.4% | 0.1% |
| Leal 2023 | 0 | 5 | 0 | 5 | 0.00 | [-0.31; 0.31] | 0.2% | 0.0% |
| Lii 2023 | 0 | 20 | 0 | 20 | 0.00 | [-0.09; 0.09] | 0.7% | 0.2% |
| Lijffijt 2021 | 0 | 20 | 0 | 20 | 0.00 | [-0.09; 0.09] | 0.7% | 0.2% |
| Loo 2016 | 0 | 9 | 0 | 9 | 0.00 | [-0.19; 0.19] | 0.4% | 0.0% |
| Loo 2023 | 0 | 89 | 2 | 89 | -0.02 | [-0.06; 0.01] | 3.2% | 1.2% |
| Milak 2020 | 0 | 38 | 0 | 38 | 0.00 | [-0.05; 0.05] | 1.4% | 0.7% |
| Murrough 2013 | 0 | 47 | 0 | 47 | 0.00 | [-0.04; 0.04] | 1.7% | 1.0% |
| Murrough 2015 II | 0 | 12 | 0 | 12 | 0.00 | [-0.15; 0.15] | 0.5% | 0.1% |
| Ochs-Ross 2020 | 0 | 72 | 0 | 72 | 0.00 | [-0.03; 0.03] | 2.6% | 2.3% |
| Ohtani 2024 | 0 | 17 | 0 | 17 | 0.00 | [-0.11; 0.11] | 0.6% | 0.1% |
| Pattanaseri 2024 | 0 | 11 | 0 | 11 | 0.00 | [-0.16; 0.16] | 0.4% | 0.1% |
| Phillips 2019 | 0 | 22 | 0 | 22 | 0.00 | [-0.08; 0.08] | 0.8% | 0.2% |
| Popova 2019 | 0 | 115 | 0 | 115 | 0.00 | [-0.02; 0.02] | 4.1% | 5.8% |
| Price 2022 | 0 | 53 | 0 | 53 | 0.00 | [-0.04; 0.04] | 1.9% | 1.3% |
| Ray-Griffith 2017 | 0 | 8 | 0 | 8 | 0.00 | [-0.21; 0.21] | 0.3% | 0.0% |
| Rodriguez 2013 | 0 | 8 | 0 | 8 | 0.00 | [-0.21; 0.21] | 0.3% | 0.0% |
| Schiweck 2021 | 0 | 21 | 0 | 21 | 0.00 | [-0.09; 0.09] | 0.8% | 0.2% |
| Shiroma 2020 | 0 | 29 | 0 | 29 | 0.00 | [-0.06; 0.06] | 1.1% | 0.4% |
| Singh 2016 2x doses | 1 | 18 | 0 | 18 | 0.06 | [-0.09; 0.20] | 0.7% | 0.1% |
| Singh 2016 3x doses | 0 | 17 | 0 | 17 | 0.00 | [-0.11; 0.11] | 0.6% | 0.1% |
| Singh 2016 II | 0 | 20 | 0 | 20 | 0.00 | [-0.09; 0.09] | 0.7% | 0.2% |
| Smith-Apeldoorn 2024 | 1 | 57 | 1 | 57 | 0.00 | [-0.05; 0.05] | 2.0% | 0.7% |
| Sos 2013 | 0 | 11 | 0 | 11 | 0.00 | [-0.16; 0.16] | 0.4% | 0.1% |
| Su 2017 | 0 | 47 | 0 | 47 | 0.00 | [-0.04; 0.04] | 1.7% | 1.0% |
| Su 2023 | 0 | 42 | 0 | 42 | 0.00 | [-0.05; 0.05] | 1.5% | 0.8% |
| Sumner 2019 | 0 | 17 | 0 | 17 | 0.00 | [-0.11; 0.11] | 0.6% | 0.1% |
| Takahashi 2021 | 0 | 122 | 0 | 122 | 0.00 | [-0.02; 0.02] | 4.4% | 6.5% |
| Taylor 2018 | 0 | 9 | 0 | 9 | 0.00 | [-0.19; 0.19] | 0.4% | 0.0% |
| Tiger 2020 | 0 | 20 | 0 | 20 | 0.00 | [-0.09; 0.09] | 0.7% | 0.2% |
| Wang 2024 | 0 | 182 | 0 | 182 | 0.00 | [-0.01; 0.01] | 6.5% | 14.4% |
| Xiao 2023 | 0 | 15 | 0 | 15 | 0.00 | [-0.12; 0.12] | 0.6% | 0.1% |
| Yang 2023 | 0 | 200 | 0 | 200 | 0.00 | [-0.01; 0.01] | 7.2% | 17.4% |
| Zarate 2006 | 0 | 9 | 0 | 9 | 0.00 | [-0.19; 0.19] | 0.4% | 0.0% |
| Zarate 2012 | 0 | 21 | 0 | 21 | 0.00 | [-0.09; 0.09] | 0.8% | 0.2% |
| Zhou 2024 | 0 | 27 | 1 | 27 | -0.04 | [-0.13; 0.06] | 1.0% | 0.2% |
| **Common effect model** | | **2742** | | **2742** | **0.00** | **[-0.01; 0.01]** | **100.0%** | **.** |
| **Random effects model** | | | | | **0.00** | **[-0.00; 0.00]** | **.** | **100.0%** |

Heterogeneity: $I^2$ = 0.0%, $\tau^2$ = 0, $p$ = 1.0000

#### Arcsine Risk-Difference

##### Arcsine RD, DB, Suicidal Behavior

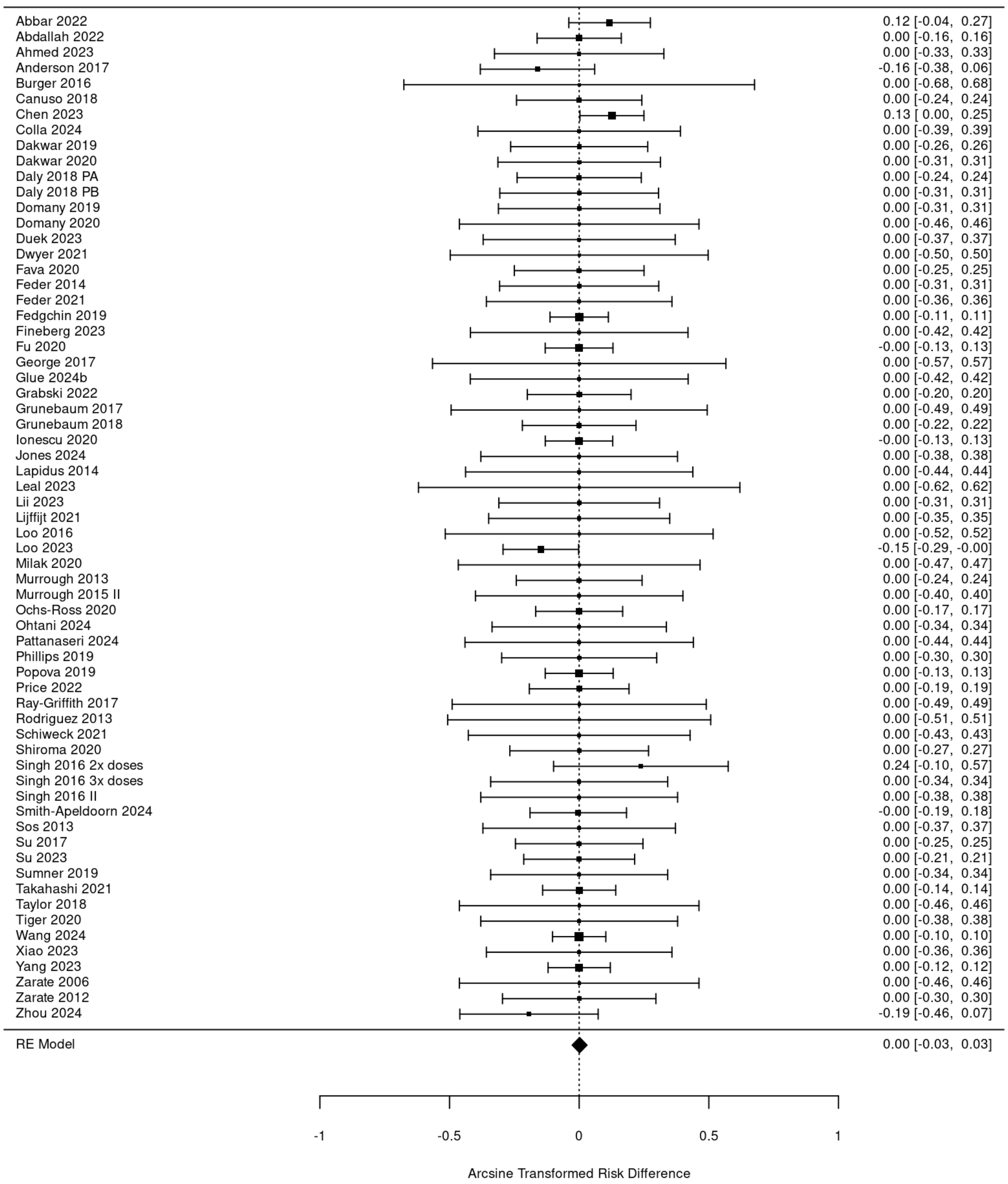

#### Bayesian Meta-Analysis, wider prior

##### Bayesian MA, Delta = 250 - Suicidal Behavior TR

| Study | Estimate | 95% CI |
|---|---|---|
| Abbar 2022 | 1.24 | [-1.98, 4.46] |
| Abdallah 2022 | -0.67 | [-4.60, 3.26] |
| Ahmed 2023 | 0.00 | [-3.97, 3.97] |
| Anderson 2017 | -1.15 | [-4.38, 2.08] |
| Burger 2016 | 0.76 | [-3.36, 4.88] |
| Canuso 2018 | -0.12 | [-4.07, 3.83] |
| Chen 2023 | 1.63 | [-1.42, 4.67] |
| Colla 2024 | -0.51 | [-4.50, 3.48] |
| Dakwar 2019 | 0.04 | [-3.92, 3.99] |
| Dakwar 2020 | 0.29 | [-3.67, 4.26] |
| Daly 2018 PA | -0.03 | [-3.98, 3.92] |
| Daly 2018 PB | 0.05 | [-3.92, 4.01] |
| Domany 2019 | -0.20 | [-4.16, 3.77] |
| Domany 2020 | 0.00 | [-4.02, 4.02] |
| Duek 2023 | 0.00 | [-3.99, 3.99] |
| Dwyer 2021 | -0.57 | [-4.61, 3.47] |
| Fava 2020 | -1.42 | [-5.37, 2.53] |
| Feder 2014 | -0.14 | [-4.11, 3.82] |
| Feder 2021 | 0.00 | [-3.98, 3.98] |
| Fedgchin 2019 | -0.71 | [-4.64, 3.21] |
| Fineberg 2023 | 0.17 | [-3.83, 4.18] |
| Fu 2020 | -0.01 | [-2.79, 2.78] |
| George 2017 | -1.02 | [-5.09, 3.04] |
| Glue 2024b | -0.72 | [-4.73, 3.28] |
| Grabski 2022 | 0.00 | [-3.94, 3.94] |
| Grunebaum 2017 | -0.24 | [-4.27, 3.80] |
| Grunebaum 2018 | 0.00 | [-3.94, 3.94] |
| Ionescu 2020 | -0.01 | [-1.63, 1.61] |
| Jones 2024 | -0.42 | [-4.41, 3.57] |
| Lapidus 2014 | 0.00 | [-4.01, 4.01] |
| Leal 2023 | 0.00 | [-4.09, 4.09] |
| Lii 2023 | 0.00 | [-3.97, 3.97] |
| Lijffijt 2021 | -0.42 | [-4.40, 3.56] |
| Loo 2016 | -0.38 | [-4.42, 3.67] |
| Loo 2023 | -1.60 | [-4.65, 1.45] |
| Milak 2020 | -1.95 | [-5.97, 2.07] |
| Murrough 2013 | -0.62 | [-4.57, 3.33] |
| Murrough 2015 II | 0.00 | [-4.00, 4.00] |
| Ochs-Ross 2020 | -0.10 | [-4.04, 3.83] |
| Ohtani 2024 | 0.00 | [-3.98, 3.98] |
| Pattanaseri 2024 | -0.19 | [-4.20, 3.82] |
| Phillips 2019 | -0.05 | [-4.01, 3.92] |
| Popova 2019 | -0.05 | [-3.98, 3.88] |
| Price 2022 | -0.04 | [-3.98, 3.90] |
| Ray-Griffith 2017 | 0.00 | [-4.03, 4.03] |
| Rodriguez 2013 | -0.13 | [-4.17, 3.92] |
| Schiweck 2021 | -1.05 | [-5.06, 2.95] |
| Shiroma 2020 | -0.15 | [-4.10, 3.81] |
| Singh 2016 2x doses | 1.04 | [-2.23, 4.31] |
| Singh 2016 3x doses | -0.06 | [-4.04, 3.92] |
| Singh 2016 II | -0.67 | [-4.66, 3.32] |
| Smith-Apeldoorn 2024 | -0.06 | [-2.85, 2.74] |
| Sos 2013 | 0.53 | [-3.46, 4.52] |
| Su 2017 | -0.66 | [-4.61, 3.29] |
| Su 2023 | 0.00 | [-3.94, 3.94] |
| Sumner 2019 | -0.06 | [-4.04, 3.92] |
| Takahashi 2021 | -0.42 | [-4.35, 3.51] |
| Taylor 2018 | 0.00 | [-4.02, 4.02] |
| Tiger 2020 | -0.67 | [-4.66, 3.32] |
| Wang 2024 | 0.00 | [-3.93, 3.93] |
| Xiao 2023 | 0.00 | [-3.98, 3.98] |
| Yang 2023 | -0.69 | [-4.62, 3.24] |
| Zarate 2006 | 0.00 | [-4.02, 4.02] |
| Zarate 2012 | 0.09 | [-3.87, 4.05] |
| Zhou 2024 | -1.14 | [-4.38, 2.11] |
| **Summary** | **-0.07** | **[-0.93, 0.79]** |
| **Prediction** | **-0.07** | **[-1.27, 1.12]** |

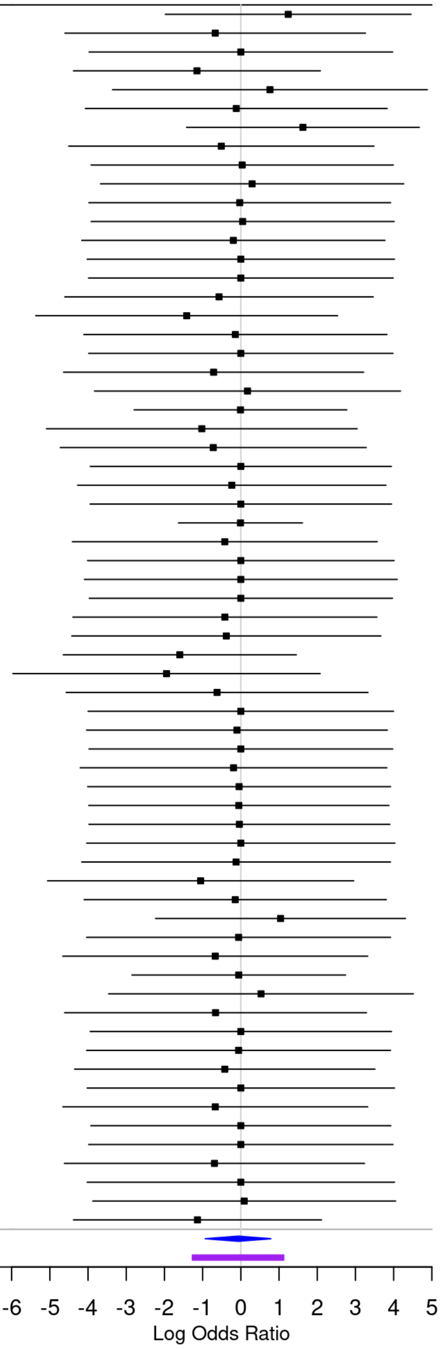

*Heterogeneity (tau): 0.27 [0.00, 0.82]*

Bayesian Meta-Analysis, smaller prior

**Bayesian MA, Delta = 15 - Suicidal Behavior TR**

| Study | Estimate | 95% CI |
|---|---|---|
| Abbar 2022 | 1.24 | [-1.98, 4.46] |
| Abdallah 2022 | -0.67 | [-4.60, 3.26] |
| Ahmed 2023 | 0.00 | [-3.97, 3.97] |
| Anderson 2017 | -1.15 | [-4.38, 2.08] |
| Burger 2016 | 0.76 | [-3.36, 4.88] |
| Canuso 2018 | -0.12 | [-4.07, 3.83] |
| Chen 2023 | 1.63 | [-1.42, 4.67] |
| Colla 2024 | -0.51 | [-4.50, 3.48] |
| Dakwar 2019 | 0.04 | [-3.92, 3.99] |
| Dakwar 2020 | 0.29 | [-3.67, 4.26] |
| Daly 2018 PA | -0.03 | [-3.98, 3.92] |
| Daly 2018 PB | 0.05 | [-3.92, 4.01] |
| Domany 2019 | -0.20 | [-4.16, 3.77] |
| Domany 2020 | 0.00 | [-4.02, 4.02] |
| Duek 2023 | 0.00 | [-3.99, 3.99] |
| Dwyer 2021 | -0.57 | [-4.61, 3.47] |
| Fava 2020 | -1.42 | [-5.37, 2.53] |
| Feder 2014 | -0.14 | [-4.11, 3.82] |
| Feder 2021 | 0.00 | [-3.98, 3.98] |
| Fedgchin 2019 | -0.71 | [-4.64, 3.21] |
| Fineberg 2023 | 0.17 | [-3.83, 4.18] |
| Fu 2020 | -0.01 | [-2.79, 2.78] |
| George 2017 | -1.02 | [-5.09, 3.04] |
| Glue 2024b | -0.72 | [-4.73, 3.28] |
| Grabski 2022 | 0.00 | [-3.94, 3.94] |
| Grunebaum 2017 | -0.24 | [-4.27, 3.80] |
| Grunebaum 2018 | 0.00 | [-3.94, 3.94] |
| Ionescu 2020 | -0.01 | [-1.63, 1.61] |
| Jones 2024 | -0.42 | [-4.41, 3.57] |
| Lapidus 2014 | 0.00 | [-4.01, 4.01] |
| Leal 2023 | 0.00 | [-4.09, 4.09] |
| Lii 2023 | 0.00 | [-3.97, 3.97] |
| Lijffijt 2021 | -0.42 | [-4.40, 3.56] |
| Loo 2016 | -0.38 | [-4.42, 3.67] |
| Loo 2023 | -1.60 | [-4.65, 1.45] |
| Milak 2020 | -1.95 | [-5.97, 2.07] |
| Murrough 2013 | -0.62 | [-4.57, 3.33] |
| Murrough 2015 II | 0.00 | [-4.00, 4.00] |
| Ochs-Ross 2020 | -0.10 | [-4.04, 3.83] |
| Ohtani 2024 | 0.00 | [-3.98, 3.98] |
| Pattanaseri 2024 | -0.19 | [-4.20, 3.82] |
| Phillips 2019 | -0.05 | [-4.01, 3.92] |
| Popova 2019 | -0.05 | [-3.98, 3.88] |
| Price 2022 | -0.04 | [-3.98, 3.90] |
| Ray-Griffith 2017 | 0.00 | [-4.03, 4.03] |
| Rodriguez 2013 | -0.13 | [-4.17, 3.92] |
| Schiweck 2021 | -1.05 | [-5.06, 2.95] |
| Shiroma 2020 | -0.15 | [-4.10, 3.81] |
| Singh 2016 2x doses | 1.04 | [-2.23, 4.31] |
| Singh 2016 3x doses | -0.06 | [-4.04, 3.92] |
| Singh 2016 II | -0.67 | [-4.66, 3.32] |
| Smith-Apeldoorn 2024 | -0.06 | [-2.85, 2.74] |
| Sos 2013 | 0.53 | [-3.46, 4.52] |
| Su 2017 | -0.66 | [-4.61, 3.29] |
| Su 2023 | 0.00 | [-3.94, 3.94] |
| Sumner 2019 | -0.06 | [-4.04, 3.92] |
| Takahashi 2021 | -0.42 | [-4.35, 3.51] |
| Taylor 2018 | 0.00 | [-4.02, 4.02] |
| Tiger 2020 | -0.67 | [-4.66, 3.32] |
| Wang 2024 | 0.00 | [-3.93, 3.93] |
| Xiao 2023 | 0.00 | [-3.98, 3.98] |
| Yang 2023 | -0.69 | [-4.62, 3.24] |
| Zarate 2006 | 0.00 | [-4.02, 4.02] |
| Zarate 2012 | 0.09 | [-3.87, 4.05] |
| Zhou 2024 | -1.14 | [-4.38, 2.11] |
| **Summary** | **-0.06** | **[-0.89, 0.77]** |
| **Prediction** | **-0.06** | **[-1.22, 1.10]** |

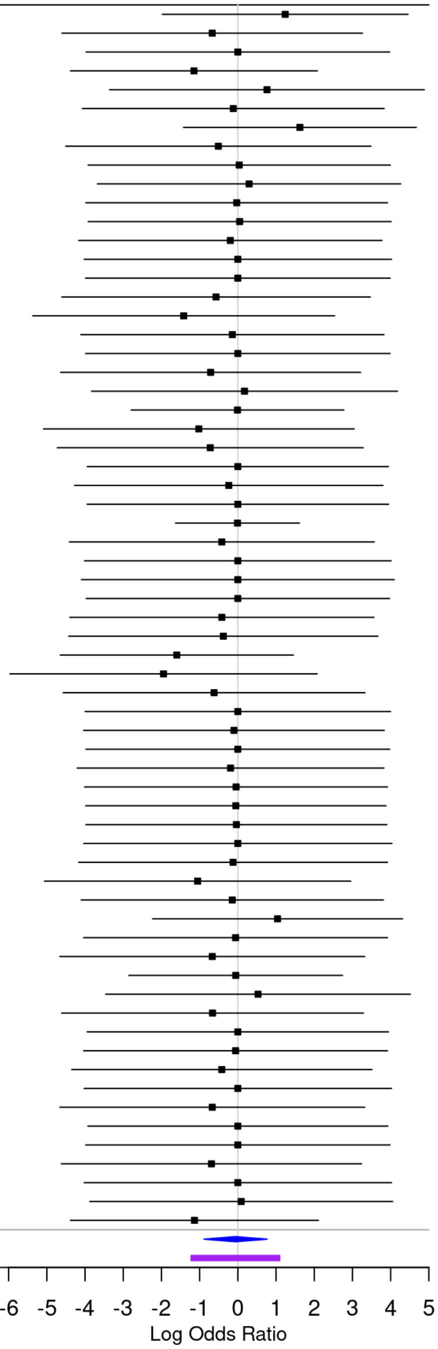

*Heterogeneity (tau): 0.28 [0.00, 0.81]*

**Exact Method**

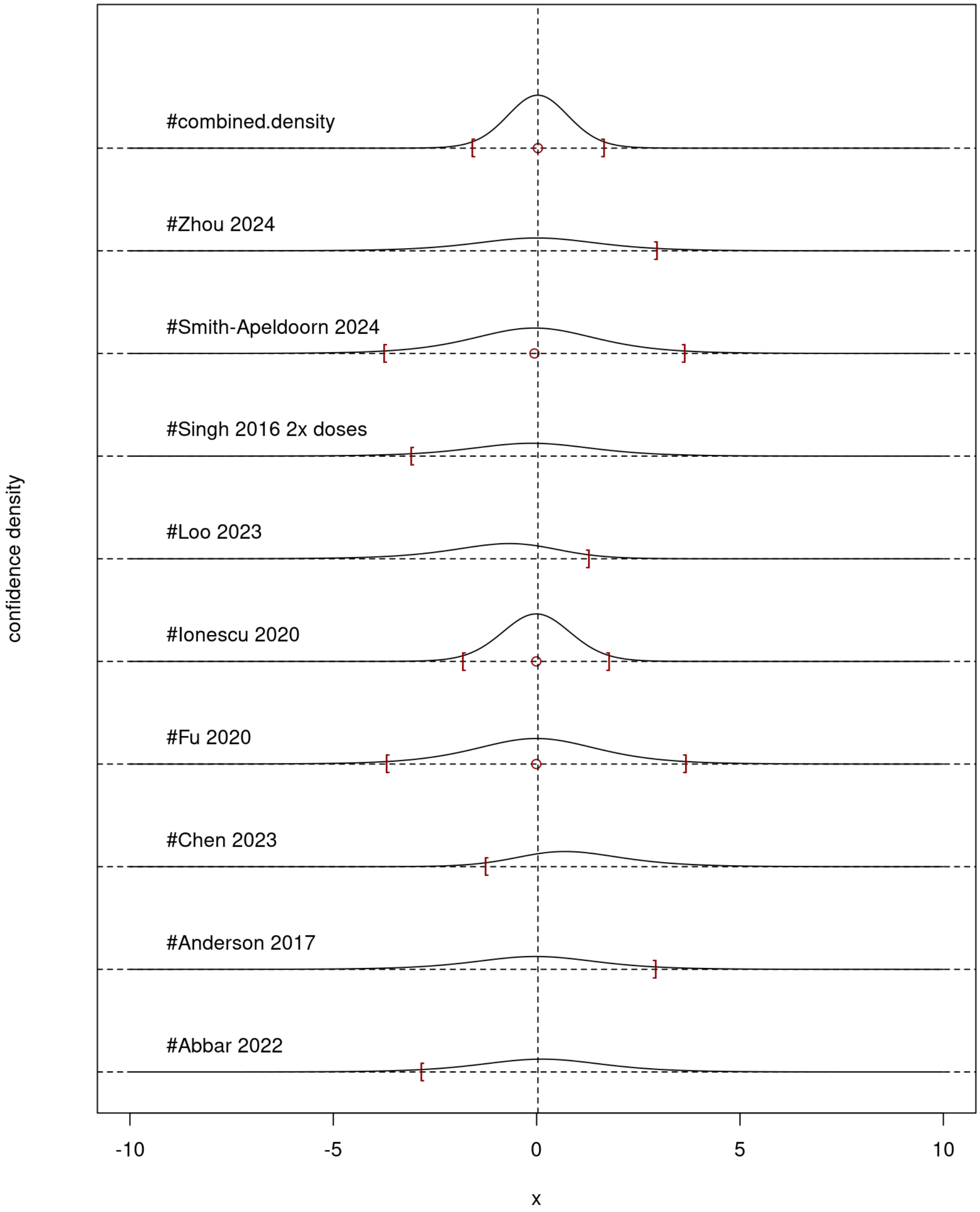

#### Follow-Up Phase

#### Peto's Method

| Study | Experimental Events | Experimental Total | Control Events | Control Total | OR | 95%-CI | Weight (common) | Weight (random) |
|---|---|---|---|---|---|---|---|---|
| Abbar 2022 | 6 | 73 | 8 | 83 | 0.84 | [0.28; 2.52] | 40.1% | 28.6% |
| Ahmed 2023 | 0 | 18 | 0 | 18 | | | 0.0% | 0.0% |
| Anderson 2017 | 0 | 25 | 1 | 23 | 0.12 | [0.00; 6.27] | 3.1% | 4.7% |
| Burger 2016 | 0 | 3 | 0 | 7 | | | 0.0% | 0.0% |
| Canuso 2018 | 0 | 27 | 3 | 22 | 0.10 | [0.01; 1.00] | 8.9% | 11.5% |
| Dakwar 2019 | 0 | 27 | 0 | 28 | | | 0.0% | 0.0% |
| Dakwar 2020 | 0 | 17 | 0 | 23 | | | 0.0% | 0.0% |
| Domany 2019 | 0 | 22 | 0 | 18 | | | 0.0% | 0.0% |
| Domany 2020 | 0 | 9 | 0 | 9 | | | 0.0% | 0.0% |
| Duek 2023 | 0 | 14 | 0 | 14 | | | 0.0% | 0.0% |
| Dwyer 2021 | 0 | 11 | 0 | 6 | | | 0.0% | 0.0% |
| Fava 2020 | 1 | 80 | 0 | 19 | 3.45 | [0.02; 499.91] | 1.9% | 3.0% |
| Feder 2014 | 0 | 22 | 0 | 19 | | | 0.0% | 0.0% |
| Fineberg 2023 | 0 | 10 | 0 | 12 | | | 0.0% | 0.0% |
| Fu 2020 | 4 | 101 | 2 | 91 | 1.78 | [0.35; 9.05] | 18.3% | 18.9% |
| Glue 2024b | 0 | 17 | 0 | 8 | | | 0.0% | 0.0% |
| Grabski 2022 | 0 | 48 | 0 | 48 | | | 0.0% | 0.0% |
| Ionescu 2020 | 4 | 89 | 1 | 94 | 3.61 | [0.61; 21.27] | 15.4% | 16.9% |
| Jones 2024 | 0 | 17 | 0 | 11 | | | 0.0% | 0.0% |
| Lapidus 2014 | 0 | 10 | 0 | 10 | | | 0.0% | 0.0% |
| Leal 2023 | 0 | 5 | 0 | 5 | | | 0.0% | 0.0% |
| Lii 2023 | 0 | 20 | 0 | 20 | | | 0.0% | 0.0% |
| Lijffijt 2021 | 0 | 20 | 1 | 13 | 0.08 | [0.00; 4.36] | 3.0% | 4.5% |
| Murrough 2013 | 0 | 47 | 0 | 25 | | | 0.0% | 0.0% |
| Murrough 2015 II | 0 | 12 | 0 | 12 | | | 0.0% | 0.0% |
| Ohtani 2024 | 0 | 17 | 0 | 17 | | | 0.0% | 0.0% |
| Pattanaseri 2024 | 0 | 11 | 0 | 9 | | | 0.0% | 0.0% |
| Phillips 2019 | 0 | 22 | 0 | 21 | | | 0.0% | 0.0% |
| Price 2022 | 0 | 53 | 0 | 51 | | | 0.0% | 0.0% |
| Ray-Griffith 2017 | 0 | 8 | 0 | 8 | | | 0.0% | 0.0% |
| Rodriguez 2013 | 0 | 8 | 0 | 7 | | | 0.0% | 0.0% |
| Schiweck 2021 | 0 | 21 | 0 | 7 | | | 0.0% | 0.0% |
| Smith-Apeldoorn 2024 | 0 | 49 | 0 | 48 | | | 0.0% | 0.0% |
| Sos 2013 | 0 | 11 | 0 | 19 | | | 0.0% | 0.0% |
| Su 2017 | 0 | 47 | 0 | 24 | | | 0.0% | 0.0% |
| Su 2023 | 1 | 42 | 2 | 42 | 0.51 | [0.05; 4.99] | 9.2% | 11.7% |
| Sumner 2019 | 0 | 17 | 0 | 16 | | | 0.0% | 0.0% |
| Taylor 2018 | 0 | 9 | 0 | 9 | | | 0.0% | 0.0% |
| Wang 2024 | 0 | 182 | 0 | 182 | | | 0.0% | 0.0% |
| Xiao 2023 | 0 | 15 | 0 | 15 | | | 0.0% | 0.0% |
| Yang 2023 | 0 | 200 | 0 | 100 | | | 0.0% | 0.0% |
| Zarate 2006 | 0 | 9 | 0 | 9 | | | 0.0% | 0.0% |
| Zarate 2012 | 0 | 21 | 0 | 23 | | | 0.0% | 0.0% |
| Zhou 2024 | 0 | 27 | 0 | 25 | | | 0.0% | 0.0% |
| **Common effect model** | | **1513** | | **1270** | **0.86** | **[0.43; 1.72]** | **100.0%** | **.** |
| **Random effects model** | | | | | **0.78** | **[0.32; 1.92]** | **.** | **100.0%** |

Heterogeneity: $I^2 = 26.0\%$, $\tau^2 = 0.4166$, $p = 0.2217$

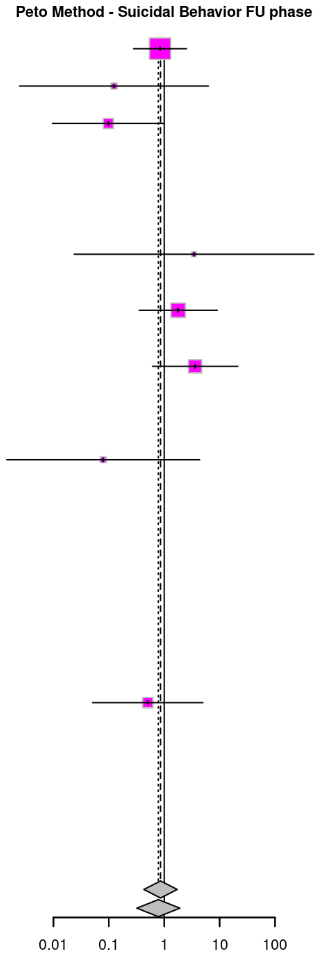

#### Maentel Mantel-Haenszel with CC

| Study | Experimental Events | Experimental Total | Control Events | Control Total | MH with CC - Suicidal Behavior FU phase | OR | 95%-CI | Weight (common) | Weight (random) |
|---|---|---|---|---|---|---|---|---|---|
| Abbar 2022 | 6 | 73 | 8 | 83 | | 0.84 | [0.28; 2.54] | 18.1% | 19.5% |
| Ahmed 2023 | 0 | 18 | 0 | 18 | | 1.00 | [0.02; 53.12] | 1.3% | 1.5% |
| Anderson 2017 | 0 | 25 | 1 | 23 | | 0.29 | [0.01; 7.59] | 4.0% | 2.3% |
| Burger 2016 | 0 | 3 | 0 | 7 | | 2.14 | [0.03; 131.93] | 0.8% | 1.4% |
| Canuso 2018 | 0 | 27 | 3 | 22 | | 0.10 | [0.00; 2.07] | 10.0% | 2.6% |
| Dakwar 2019 | 0 | 27 | 0 | 28 | | 1.04 | [0.02; 54.08] | 1.3% | 1.5% |
| Dakwar 2020 | 0 | 17 | 0 | 23 | | 1.34 | [0.03; 71.04] | 1.1% | 1.5% |
| Domany 2019 | 0 | 22 | 0 | 18 | | 0.82 | [0.02; 43.47] | 1.4% | 1.5% |
| Domany 2020 | 0 | 9 | 0 | 9 | | 1.00 | [0.02; 55.80] | 1.3% | 1.5% |
| Duek 2023 | 0 | 14 | 0 | 14 | | 1.00 | [0.02; 53.89] | 1.3% | 1.5% |
| Dwyer 2021 | 0 | 11 | 0 | 6 | | 0.57 | [0.01; 32.00] | 1.6% | 1.5% |
| Fava 2020 | 1 | 80 | 0 | 19 | | 0.74 | [0.03; 18.76] | 2.1% | 2.3% |
| Feder 2014 | 0 | 22 | 0 | 19 | | 0.87 | [0.02; 45.76] | 1.4% | 1.5% |
| Fineberg 2023 | 0 | 10 | 0 | 12 | | 1.19 | [0.02; 65.32] | 1.2% | 1.5% |
| Fu 2020 | 4 | 101 | 2 | 91 | | 1.84 | [0.33; 10.26] | 5.3% | 8.1% |
| Glue 2024b | 0 | 17 | 0 | 8 | | 0.49 | [0.01; 26.64] | 1.7% | 1.5% |
| Grabski 2022 | 0 | 48 | 0 | 48 | | 1.00 | [0.02; 51.42] | 1.3% | 1.5% |
| Ionescu 2020 | 4 | 89 | 1 | 94 | | 4.38 | [0.48; 39.93] | 2.4% | 4.9% |
| Jones 2024 | 0 | 17 | 0 | 11 | | 0.66 | [0.01; 35.52] | 1.5% | 1.5% |
| Lapidus 2014 | 0 | 10 | 0 | 10 | | 1.00 | [0.02; 55.27] | 1.3% | 1.5% |
| Leal 2023 | 0 | 5 | 0 | 5 | | 1.00 | [0.02; 59.99] | 1.2% | 1.4% |
| Lii 2023 | 0 | 20 | 0 | 20 | | 1.00 | [0.02; 52.85] | 1.3% | 1.5% |
| Lijffijt 2021 | 0 | 20 | 1 | 13 | | 0.20 | [0.01; 5.39] | 4.6% | 2.2% |
| Murrough 2013 | 0 | 47 | 0 | 25 | | 0.54 | [0.01; 27.86] | 1.7% | 1.5% |
| Murrough 2015 II | 0 | 12 | 0 | 12 | | 1.00 | [0.02; 54.46] | 1.3% | 1.5% |
| Ohtani 2024 | 0 | 17 | 0 | 17 | | 1.00 | [0.02; 53.28] | 1.3% | 1.5% |
| Pattanaseri 2024 | 0 | 11 | 0 | 9 | | 0.83 | [0.01; 45.69] | 1.4% | 1.5% |
| Phillips 2019 | 0 | 22 | 0 | 21 | | 0.96 | [0.02; 50.34] | 1.3% | 1.5% |
| Price 2022 | 0 | 53 | 0 | 51 | | 0.96 | [0.02; 49.43] | 1.3% | 1.5% |
| Ray-Griffith 2017 | 0 | 8 | 0 | 8 | | 1.00 | [0.02; 56.46] | 1.2% | 1.5% |
| Rodriguez 2013 | 0 | 8 | 0 | 7 | | 0.88 | [0.02; 50.19] | 1.3% | 1.5% |
| Schiweck 2021 | 0 | 21 | 0 | 7 | | 0.35 | [0.01; 19.18] | 1.9% | 1.5% |
| Smith-Apeldoorn 2024 | 0 | 49 | 0 | 48 | | 0.98 | [0.02; 50.37] | 1.3% | 1.5% |
| Sos 2013 | 0 | 11 | 0 | 19 | | 1.70 | [0.03; 91.39] | 0.9% | 1.5% |
| Su 2017 | 0 | 47 | 0 | 24 | | 0.52 | [0.01; 26.79] | 1.7% | 1.5% |
| Su 2023 | 1 | 42 | 2 | 42 | | 0.49 | [0.04; 5.59] | 5.1% | 4.0% |
| Sumner 2019 | 0 | 17 | 0 | 16 | | 0.94 | [0.02; 50.32] | 1.3% | 1.5% |
| Taylor 2018 | 0 | 9 | 0 | 9 | | 1.00 | [0.02; 55.80] | 1.3% | 1.5% |
| Wang 2024 | 0 | 182 | 0 | 182 | | 1.00 | [0.02; 50.67] | 1.3% | 1.6% |
| Xiao 2023 | 0 | 15 | 0 | 15 | | 1.00 | [0.02; 53.66] | 1.3% | 1.5% |
| Yang 2023 | 0 | 200 | 0 | 100 | | 0.50 | [0.01; 25.45] | 1.8% | 1.6% |
| Zarate 2006 | 0 | 9 | 0 | 9 | | 1.00 | [0.02; 55.80] | 1.3% | 1.5% |
| Zarate 2012 | 0 | 21 | 0 | 23 | | 1.09 | [0.02; 57.53] | 1.2% | 1.5% |
| Zhou 2024 | 0 | 27 | 0 | 25 | | 0.93 | [0.02; 48.49] | 1.3% | 1.5% |
| **Common effect model** | | **1513** | | **1270** | | **0.86** | **[0.54; 1.38]** | **100.0%** | **.** |
| **Random effects model** | | | | | | **0.88** | **[0.54; 1.43]** | **.** | **100.0%** |

Heterogeneity: $I^2 = 0.0\%$, $\tau^2 = 0$, $p = 1.0000$

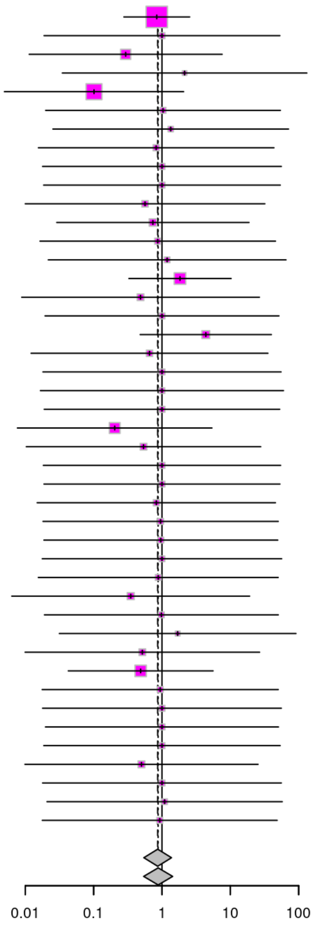

#### Maentel Mantel-Haenszel with Treatment-Arm CC

| Study | Experimental Events | Experimental Total | Control Events | Control Total | OR | 95%-CI | Weight (common) | Weight (random) |
|---|---|---|---|---|---|---|---|---|
| Abbar 2022 | 6 | 73 | 8 | 83 | 0.84 | [0.28; 2.54] | 19.2% | 19.9% |
| Ahmed 2023 | 0 | 18 | 0 | 18 | 1.00 | [0.02; 53.12] | 1.4% | 1.6% |
| Anderson 2017 | 0 | 25 | 1 | 23 | 0.31 | [0.01; 7.67] | 4.2% | 2.4% |
| Burger 2016 | 0 | 3 | 0 | 7 | 1.00 | [0.01; 87.11] | 1.1% | 1.2% |
| Canuso 2018 | 0 | 27 | 3 | 22 | 0.11 | [0.01; 2.05] | 10.4% | 2.9% |
| Dakwar 2019 | 0 | 27 | 0 | 28 | 1.00 | [0.02; 52.22] | 1.4% | 1.6% |
| Dakwar 2020 | 0 | 17 | 0 | 23 | 1.00 | [0.02; 55.30] | 1.3% | 1.5% |
| Domany 2019 | 0 | 22 | 0 | 18 | 1.00 | [0.02; 53.92] | 1.3% | 1.5% |
| Domany 2020 | 0 | 9 | 0 | 9 | 1.00 | [0.02; 55.80] | 1.3% | 1.5% |
| Duek 2023 | 0 | 14 | 0 | 14 | 1.00 | [0.02; 53.89] | 1.3% | 1.5% |
| Dwyer 2021 | 0 | 11 | 0 | 6 | 1.00 | [0.01; 67.61] | 1.2% | 1.4% |
| Fava 2020 | 1 | 80 | 0 | 19 | 2.27 | [0.02; 257.11] | 0.8% | 1.1% |
| Feder 2014 | 0 | 22 | 0 | 19 | 1.00 | [0.02; 53.36] | 1.4% | 1.5% |
| Fineberg 2023 | 0 | 10 | 0 | 12 | 1.00 | [0.02; 55.75] | 1.3% | 1.5% |
| Fu 2020 | 4 | 101 | 2 | 91 | 1.84 | [0.33; 10.26] | 5.6% | 8.3% |
| Glue 2024b | 0 | 17 | 0 | 8 | 1.00 | [0.01; 72.36] | 1.2% | 1.3% |
| Grabski 2022 | 0 | 48 | 0 | 48 | 1.00 | [0.02; 51.42] | 1.4% | 1.6% |
| Ionescu 2020 | 4 | 89 | 1 | 94 | 4.38 | [0.48; 39.93] | 2.6% | 5.0% |
| Jones 2024 | 0 | 17 | 0 | 11 | 1.00 | [0.02; 59.25] | 1.3% | 1.5% |
| Lapidus 2014 | 0 | 10 | 0 | 10 | 1.00 | [0.02; 55.27] | 1.3% | 1.5% |
| Leal 2023 | 0 | 5 | 0 | 5 | 1.00 | [0.02; 59.99] | 1.3% | 1.5% |
| Lii 2023 | 0 | 20 | 0 | 20 | 1.00 | [0.02; 52.85] | 1.4% | 1.6% |
| Lijffijt 2021 | 0 | 20 | 1 | 13 | 0.26 | [0.01; 5.79] | 4.6% | 2.6% |
| Murrough 2013 | 0 | 47 | 0 | 25 | 1.00 | [0.02; 63.11] | 1.2% | 1.4% |
| Murrough 2015 II | 0 | 12 | 0 | 12 | 1.00 | [0.02; 54.46] | 1.3% | 1.5% |
| Ohtani 2024 | 0 | 17 | 0 | 17 | 1.00 | [0.02; 53.28] | 1.4% | 1.5% |
| Pattanaseri 2024 | 0 | 11 | 0 | 9 | 1.00 | [0.02; 56.40] | 1.3% | 1.5% |
| Phillips 2019 | 0 | 22 | 0 | 21 | 1.00 | [0.02; 52.74] | 1.4% | 1.6% |
| Price 2022 | 0 | 53 | 0 | 51 | 1.00 | [0.02; 51.38] | 1.4% | 1.6% |
| Ray-Griffith 2017 | 0 | 8 | 0 | 8 | 1.00 | [0.02; 56.46] | 1.3% | 1.5% |
| Rodriguez 2013 | 0 | 8 | 0 | 7 | 1.00 | [0.02; 57.37] | 1.3% | 1.5% |
| Schiweck 2021 | 0 | 21 | 0 | 7 | 1.00 | [0.01; 99.86] | 1.0% | 1.2% |
| Smith-Apeldoorn 2024 | 0 | 49 | 0 | 48 | 1.00 | [0.02; 51.42] | 1.4% | 1.6% |
| Sos 2013 | 0 | 11 | 0 | 19 | 1.00 | [0.02; 62.32] | 1.3% | 1.4% |
| Su 2017 | 0 | 47 | 0 | 24 | 1.00 | [0.02; 64.85] | 1.2% | 1.4% |
| Su 2023 | 1 | 42 | 2 | 42 | 0.49 | [0.04; 5.59] | 5.4% | 4.1% |
| Sumner 2019 | 0 | 17 | 0 | 16 | 1.00 | [0.02; 53.46] | 1.4% | 1.5% |
| Taylor 2018 | 0 | 9 | 0 | 9 | 1.00 | [0.02; 55.80] | 1.3% | 1.5% |
| Wang 2024 | 0 | 182 | 0 | 182 | 1.00 | [0.02; 50.67] | 1.4% | 1.6% |
| Xiao 2023 | 0 | 15 | 0 | 15 | 1.00 | [0.02; 53.66] | 1.4% | 1.5% |
| Yang 2023 | 0 | 200 | 0 | 100 | 1.00 | [0.02; 64.37] | 1.2% | 1.4% |
| Zarate 2006 | 0 | 9 | 0 | 9 | 1.00 | [0.02; 55.80] | 1.3% | 1.5% |
| Zarate 2012 | 0 | 21 | 0 | 23 | 1.00 | [0.02; 52.84] | 1.4% | 1.6% |
| Zhou 2024 | 0 | 27 | 0 | 25 | 1.00 | [0.02; 52.44] | 1.4% | 1.6% |
| **Common effect model** | | **1513** | | **1270** | **0.93** | **[0.58; 1.49]** | **100.0%** | **.** |
| **Random effects model** | | | | | **0.94** | **[0.58; 1.55]** | **.** | **100.0%** |

Heterogeneity: $I^2 = 0.0\%$, $\tau^2 = 0$, $p = 1.0000$

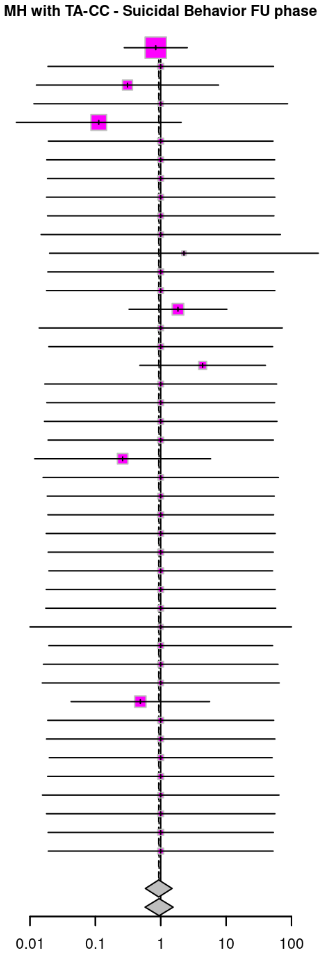

#### Maentel Mantel-Haenszel Risk-Difference

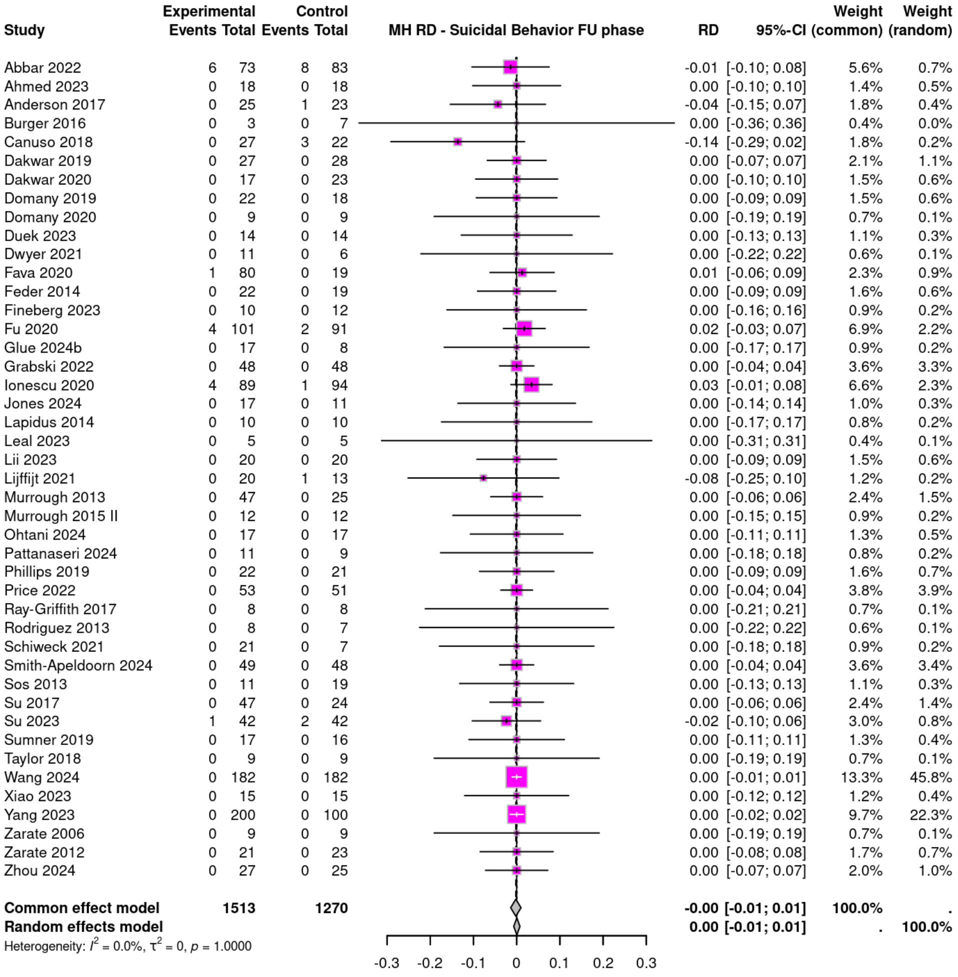

#### Arcsine Risk-Difference

##### Arcsine RD, FU, Suicidal Behavior

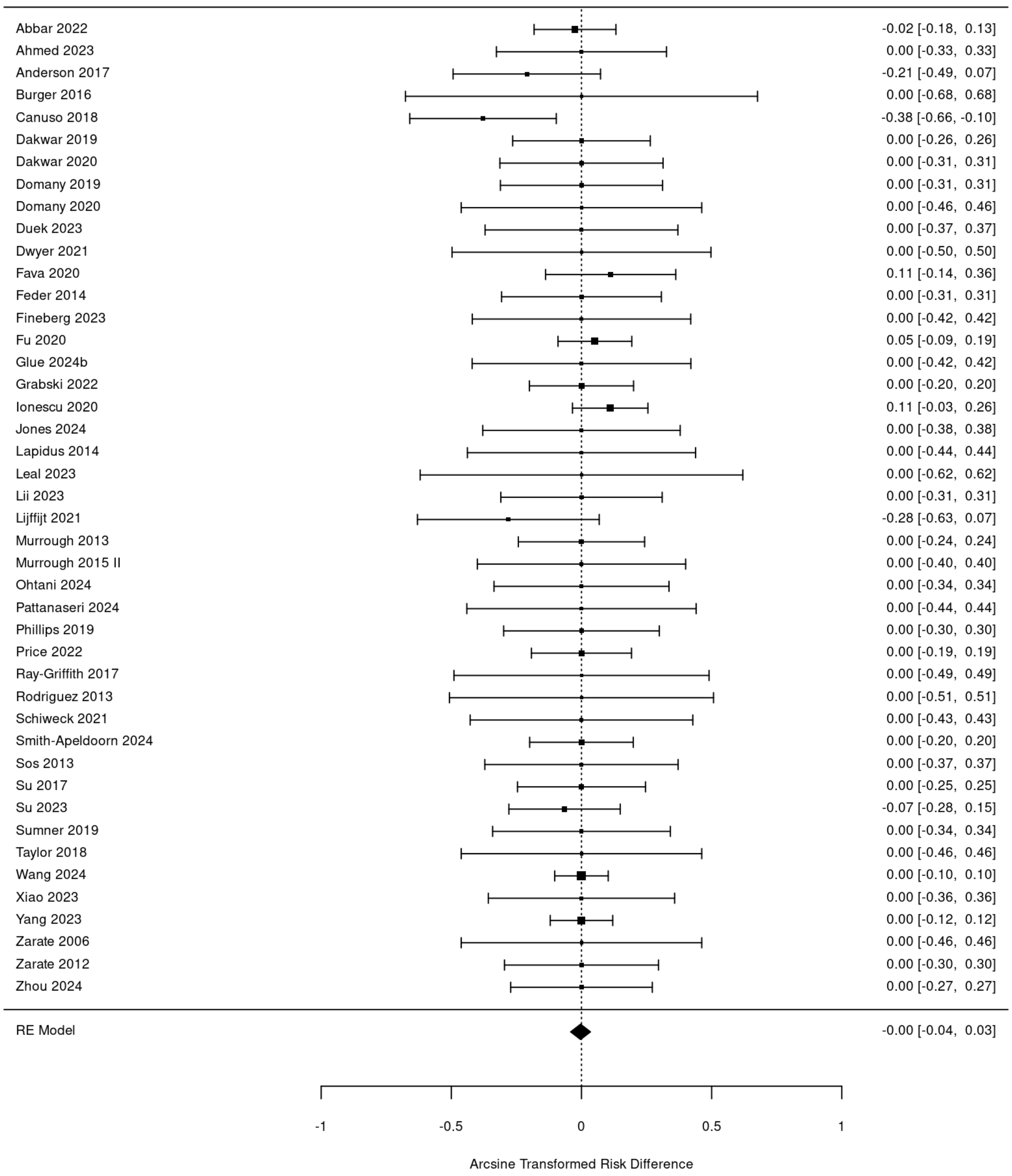

**Bayesian Meta-Analysis, wider prior**

**Bayesian MA, Delta = 250 - Suicidal Behavior FU**

| Study | Estimate | 95% CI |
|---|---|---|
| Abbar 2022 | -0.17 | [-1.28, 0.93] |
| Ahmed 2023 | 0.00 | [-3.97, 3.97] |
| Anderson 2017 | -1.22 | [-4.47, 2.03] |
| Burger 2016 | 0.76 | [-3.36, 4.88] |
| Canuso 2018 | -2.29 | [-5.31, 0.73] |
| Dakwar 2019 | 0.04 | [-3.92, 3.99] |
| Dakwar 2020 | 0.29 | [-3.67, 4.26] |
| Domany 2019 | -0.20 | [-4.16, 3.77] |
| Domany 2020 | 0.00 | [-4.02, 4.02] |
| Duek 2023 | 0.00 | [-3.99, 3.99] |
| Dwyer 2021 | -0.57 | [-4.61, 3.47] |
| Fava 2020 | -0.31 | [-3.55, 2.93] |
| Feder 2014 | -0.14 | [-4.11, 3.82] |
| Fineberg 2023 | 0.17 | [-3.83, 4.18] |
| Fu 2020 | 0.61 | [-1.11, 2.33] |
| Glue 2024b | -0.72 | [-4.73, 3.28] |
| Grabski 2022 | 0.00 | [-3.94, 3.94] |
| Ionescu 2020 | 1.48 | [-0.73, 3.69] |
| Jones 2024 | -0.42 | [-4.41, 3.57] |
| Lapidus 2014 | 0.00 | [-4.01, 4.01] |
| Leal 2023 | 0.00 | [-4.09, 4.09] |
| Lii 2023 | 0.00 | [-3.97, 3.97] |
| Lijffijt 2021 | -1.59 | [-4.87, 1.68] |
| Murrough 2013 | -0.62 | [-4.57, 3.33] |
| Murrough 2015 II | 0.00 | [-4.00, 4.00] |
| Ohtani 2024 | 0.00 | [-3.98, 3.98] |
| Pattanaseri 2024 | -0.19 | [-4.20, 3.82] |
| Phillips 2019 | -0.05 | [-4.01, 3.92] |
| Price 2022 | -0.04 | [-3.98, 3.90] |
| Ray-Griffith 2017 | 0.00 | [-4.03, 4.03] |
| Rodriguez 2013 | -0.13 | [-4.17, 3.92] |
| Schiweck 2021 | -1.05 | [-5.06, 2.95] |
| Smith-Apeldoorn 2024 | -0.02 | [-3.96, 3.92] |
| Sos 2013 | 0.53 | [-3.46, 4.52] |
| Su 2017 | -0.66 | [-4.61, 3.29] |
| Su 2023 | -0.72 | [-3.16, 1.72] |
| Sumner 2019 | -0.06 | [-4.04, 3.92] |
| Taylor 2018 | 0.00 | [-4.02, 4.02] |
| Wang 2024 | 0.00 | [-3.93, 3.93] |
| Xiao 2023 | 0.00 | [-3.98, 3.98] |
| Yang 2023 | -0.69 | [-4.62, 3.24] |
| Zarate 2006 | 0.00 | [-4.02, 4.02] |
| Zarate 2012 | 0.09 | [-3.87, 4.05] |
| Zhou 2024 | -0.08 | [-4.03, 3.88] |
| **Summary** | **-0.19** | **[-0.96, 0.56]** |
| **Prediction** | **-0.18** | **[-1.47, 1.01]** |

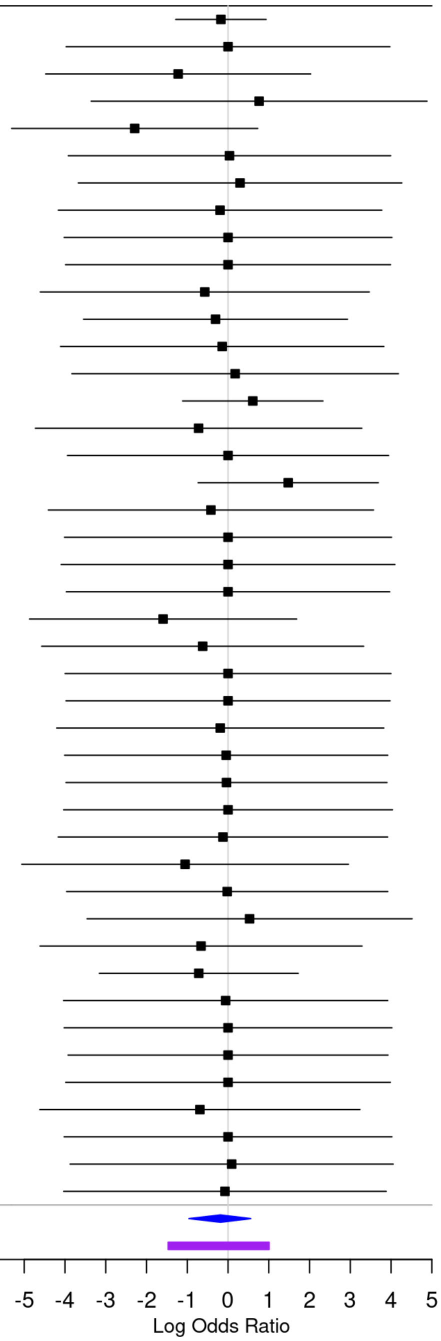

*Heterogeneity (tau): 0.32 [0.00, 0.90]*

Bayesian Meta-Analysis, smaller prior

**Bayesian MA, Delta = 15 - Suicidal Behavior FU**

| Study | Estimate | 95% CI |
|---|---|---|
| Abbar 2022 | -0.17 | [-1.28, 0.93] |
| Ahmed 2023 | 0.00 | [-3.97, 3.97] |
| Anderson 2017 | -1.22 | [-4.47, 2.03] |
| Burger 2016 | 0.76 | [-3.36, 4.88] |
| Canuso 2018 | -2.29 | [-5.31, 0.73] |
| Dakwar 2019 | 0.04 | [-3.92, 3.99] |
| Dakwar 2020 | 0.29 | [-3.67, 4.26] |
| Domany 2019 | -0.20 | [-4.16, 3.77] |
| Domany 2020 | 0.00 | [-4.02, 4.02] |
| Duek 2023 | 0.00 | [-3.99, 3.99] |
| Dwyer 2021 | -0.57 | [-4.61, 3.47] |
| Fava 2020 | -0.31 | [-3.55, 2.93] |
| Feder 2014 | -0.14 | [-4.11, 3.82] |
| Fineberg 2023 | 0.17 | [-3.83, 4.18] |
| Fu 2020 | 0.61 | [-1.11, 2.33] |
| Glue 2024b | -0.72 | [-4.73, 3.28] |
| Grabski 2022 | 0.00 | [-3.94, 3.94] |
| Ionescu 2020 | 1.48 | [-0.73, 3.69] |
| Jones 2024 | -0.42 | [-4.41, 3.57] |
| Lapidus 2014 | 0.00 | [-4.01, 4.01] |
| Leal 2023 | 0.00 | [-4.09, 4.09] |
| Lii 2023 | 0.00 | [-3.97, 3.97] |
| Lijffijt 2021 | -1.59 | [-4.87, 1.68] |
| Murrough 2013 | -0.62 | [-4.57, 3.33] |
| Murrough 2015 II | 0.00 | [-4.00, 4.00] |
| Ohtani 2024 | 0.00 | [-3.98, 3.98] |
| Pattanaseri 2024 | -0.19 | [-4.20, 3.82] |
| Phillips 2019 | -0.05 | [-4.01, 3.92] |
| Price 2022 | -0.04 | [-3.98, 3.90] |
| Ray-Griffith 2017 | 0.00 | [-4.03, 4.03] |
| Rodriguez 2013 | -0.13 | [-4.17, 3.92] |
| Schiweck 2021 | -1.05 | [-5.06, 2.95] |
| Smith-Apeldoorn 2024 | -0.02 | [-3.96, 3.92] |
| Sos 2013 | 0.53 | [-3.46, 4.52] |
| Su 2017 | -0.66 | [-4.61, 3.29] |
| Su 2023 | -0.72 | [-3.16, 1.72] |
| Sumner 2019 | -0.06 | [-4.04, 3.92] |
| Taylor 2018 | 0.00 | [-4.02, 4.02] |
| Wang 2024 | 0.00 | [-3.93, 3.93] |
| Xiao 2023 | 0.00 | [-3.98, 3.98] |
| Yang 2023 | -0.69 | [-4.62, 3.24] |
| Zarate 2006 | 0.00 | [-4.02, 4.02] |
| Zarate 2012 | 0.09 | [-3.87, 4.05] |
| Zhou 2024 | -0.08 | [-4.03, 3.88] |
| **Summary** | **-0.17** | **[-0.93, 0.55]** |
| **Prediction** | **-0.17** | **[-1.45, 1.00]** |

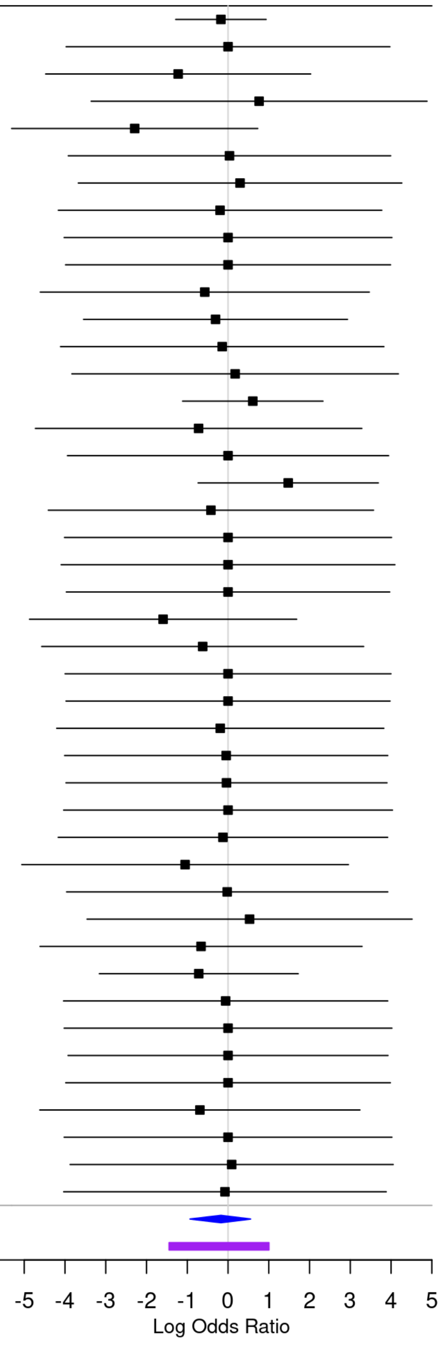

*Heterogeneity (tau): 0.32 [0.00, 0.89]*

**Exact Method**

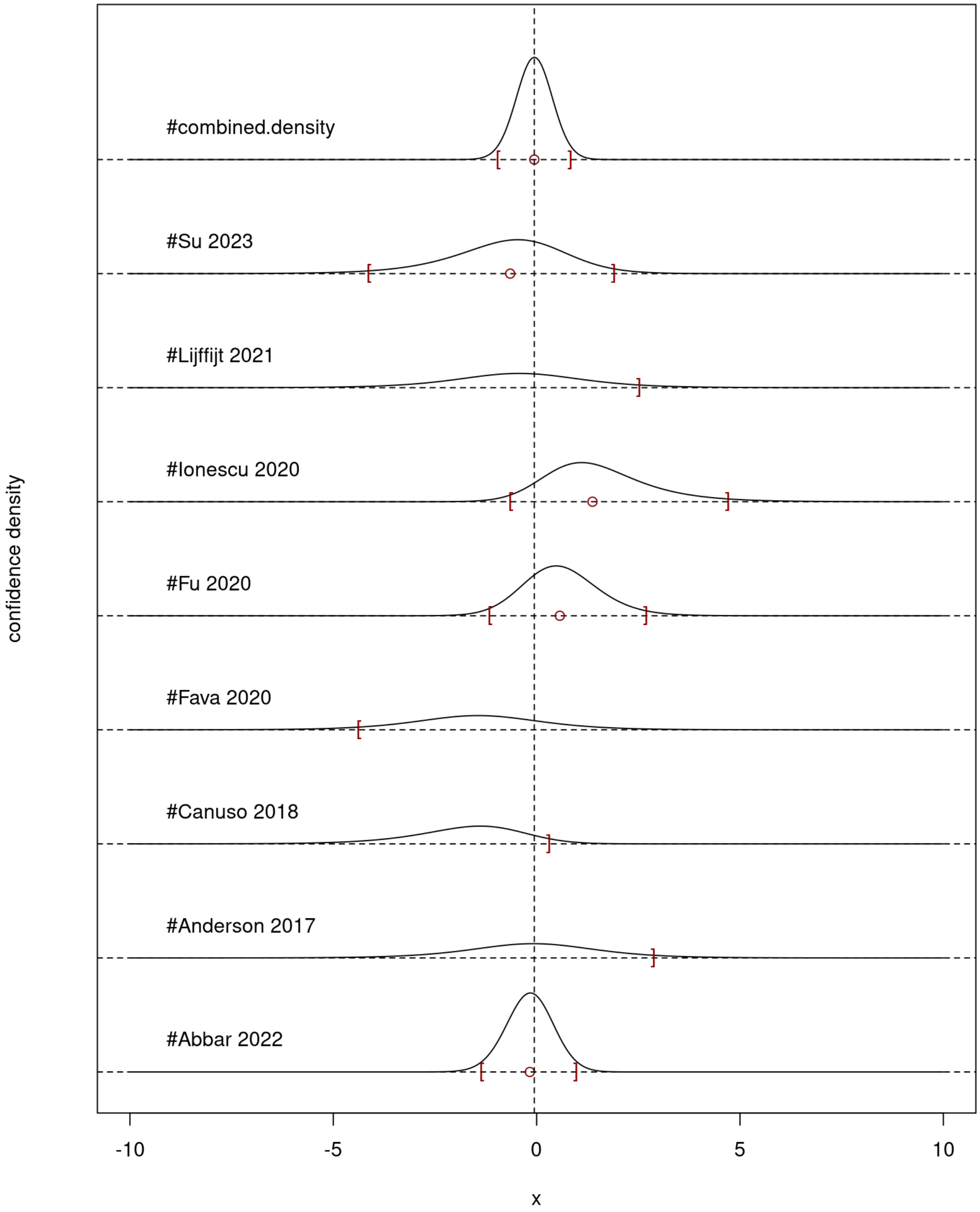

#### Treatment + Follow-Up Phase Combined

#### Peto's Method

| Study | Experimental Events | Experimental Total | Control Events | Control Total | OR | 95%-CI | Weight (common) | Weight (random) |
|---|---|---|---|---|---|---|---|---|
| Abbar 2022 | 7 | 73 | 8 | 83 | 0.99 | [0.34; 2.88] | 27.6% | 25.2% |
| Abdallah 2022 | 0 | 108 | 0 | 55 | | | 0.0% | 0.0% |
| Ahmed 2023 | 0 | 18 | 0 | 18 | | | 0.0% | 0.0% |
| Anderson 2017 | 0 | 40 | 2 | 39 | 0.13 | [0.01; 2.09] | 4.0% | 4.4% |
| Burger 2016 | 0 | 3 | 0 | 7 | | | 0.0% | 0.0% |
| Canuso 2018 | 0 | 35 | 3 | 31 | 0.11 | [0.01; 1.11] | 5.9% | 6.4% |
| Chen 2023 | 2 | 126 | 0 | 126 | 7.45 | [0.46; 119.74] | 4.1% | 4.5% |
| Colla 2024 | 0 | 17 | 0 | 10 | | | 0.0% | 0.0% |
| Dakwar 2019 | 0 | 27 | 0 | 28 | | | 0.0% | 0.0% |
| Dakwar 2020 | 0 | 17 | 0 | 23 | | | 0.0% | 0.0% |
| Daly 2018 PA | 0 | 34 | 0 | 33 | | | 0.0% | 0.0% |
| Daly 2018 PB | 0 | 20 | 0 | 21 | | | 0.0% | 0.0% |
| Domany 2019 | 0 | 22 | 0 | 18 | | | 0.0% | 0.0% |
| Domany 2020 | 0 | 9 | 0 | 9 | | | 0.0% | 0.0% |
| Duek 2023 | 0 | 14 | 0 | 14 | | | 0.0% | 0.0% |
| Dwyer 2021 | 0 | 11 | 0 | 6 | | | 0.0% | 0.0% |
| Fava 2020 | 1 | 80 | 0 | 19 | 3.45 | [0.02; 499.91] | 1.3% | 1.4% |
| Feder 2014 | 0 | 22 | 0 | 19 | | | 0.0% | 0.0% |
| Feder 2021 | 0 | 15 | 0 | 15 | | | 0.0% | 0.0% |
| Fedgchin 2019 | 0 | 231 | 0 | 113 | | | 0.0% | 0.0% |
| Fineberg 2023 | 0 | 10 | 0 | 12 | | | 0.0% | 0.0% |
| Fu 2020 | 5 | 113 | 3 | 112 | 1.66 | [0.41; 6.79] | 15.8% | 15.7% |
| George 2017 | 0 | 12 | 0 | 4 | | | 0.0% | 0.0% |
| Glue 2024b | 0 | 17 | 0 | 8 | | | 0.0% | 0.0% |
| Grabski 2022 | 0 | 48 | 0 | 48 | | | 0.0% | 0.0% |
| Grunebaum 2017 | 0 | 9 | 0 | 7 | | | 0.0% | 0.0% |
| Grunebaum 2018 | 0 | 40 | 0 | 40 | | | 0.0% | 0.0% |
| Ionescu 2020 | 7 | 114 | 4 | 113 | 1.75 | [0.52; 5.87] | 21.4% | 20.4% |
| Jones 2024 | 0 | 17 | 0 | 11 | | | 0.0% | 0.0% |
| Lapidus 2014 | 0 | 10 | 0 | 10 | | | 0.0% | 0.0% |
| Leal 2023 | 0 | 5 | 0 | 5 | | | 0.0% | 0.0% |
| Lii 2023 | 0 | 20 | 0 | 20 | | | 0.0% | 0.0% |
| Lijffijt 2021 | 0 | 20 | 1 | 13 | 0.08 | [0.00; 4.36] | 1.9% | 2.2% |
| Loo 2016 | 0 | 9 | 0 | 6 | | | 0.0% | 0.0% |
| Loo 2023 | 0 | 89 | 2 | 92 | 0.14 | [0.01; 2.23] | 4.0% | 4.5% |
| Milak 2020 | 0 | 38 | 0 | 5 | | | 0.0% | 0.0% |
| Murrough 2013 | 0 | 47 | 0 | 25 | | | 0.0% | 0.0% |
| Murrough 2015 II | 0 | 12 | 0 | 12 | | | 0.0% | 0.0% |
| Ochs-Ross 2020 | 0 | 72 | 0 | 65 | | | 0.0% | 0.0% |
| Ohtani 2024 | 0 | 17 | 0 | 17 | | | 0.0% | 0.0% |
| Pattanaseri 2024 | 0 | 11 | 0 | 9 | | | 0.0% | 0.0% |
| Phillips 2019 | 0 | 22 | 0 | 21 | | | 0.0% | 0.0% |
| Popova 2019 | 0 | 115 | 0 | 109 | | | 0.0% | 0.0% |
| Price 2022 | 0 | 53 | 0 | 51 | | | 0.0% | 0.0% |
| Ray-Griffith 2017 | 0 | 8 | 0 | 8 | | | 0.0% | 0.0% |
| Rodriguez 2013 | 0 | 8 | 0 | 7 | | | 0.0% | 0.0% |
| Schiweck 2021 | 0 | 21 | 0 | 7 | | | 0.0% | 0.0% |
| Shiroma 2020 | 0 | 29 | 0 | 25 | | | 0.0% | 0.0% |
| Singh 2016 2x doses | 1 | 18 | 0 | 16 | 6.61 | [0.13; 335.50] | 2.0% | 2.3% |
| Singh 2016 3x doses | 0 | 17 | 0 | 16 | | | 0.0% | 0.0% |
| Singh 2016 II | 0 | 20 | 0 | 10 | | | 0.0% | 0.0% |
| Smith-Apeldoorn 2024 | 1 | 57 | 1 | 54 | 0.95 | [0.06; 15.35] | 4.0% | 4.4% |
| Sos 2013 | 0 | 11 | 0 | 19 | | | 0.0% | 0.0% |
| Su 2017 | 0 | 47 | 0 | 24 | | | 0.0% | 0.0% |
| Su 2023 | 1 | 42 | 2 | 42 | 0.51 | [0.05; 4.99] | 6.0% | 6.4% |
| Sumner 2019 | 0 | 17 | 0 | 16 | | | 0.0% | 0.0% |
| Takahashi 2021 | 0 | 122 | 0 | 80 | | | 0.0% | 0.0% |
| Taylor 2018 | 0 | 9 | 0 | 9 | | | 0.0% | 0.0% |
| Tiger 2020 | 0 | 20 | 0 | 10 | | | 0.0% | 0.0% |
| Wang 2024 | 0 | 182 | 0 | 182 | | | 0.0% | 0.0% |
| Xiao 2023 | 0 | 15 | 0 | 15 | | | 0.0% | 0.0% |
| Yang 2023 | 0 | 200 | 0 | 100 | | | 0.0% | 0.0% |
| Zarate 2006 | 0 | 9 | 0 | 9 | | | 0.0% | 0.0% |
| Zarate 2012 | 0 | 21 | 0 | 23 | | | 0.0% | 0.0% |
| Zhou 2024 | 0 | 27 | 1 | 27 | 0.14 | [0.00; 6.82] | 2.0% | 2.3% |
| **Common effect model** | | **2742** | | **2201** | **0.91** | **[0.52; 1.60]** | **100.0%** | **.** |
| **Random effects model** | | | | | **0.88** | **[0.49; 1.61]** | **.** | **100.0%** |

Heterogeneity: $I^2 = 18.7\%$, $\tau^2 = 0.0749$, $p = 0.2549$

Peto Method - Suicidal Behavior TR+FU phase

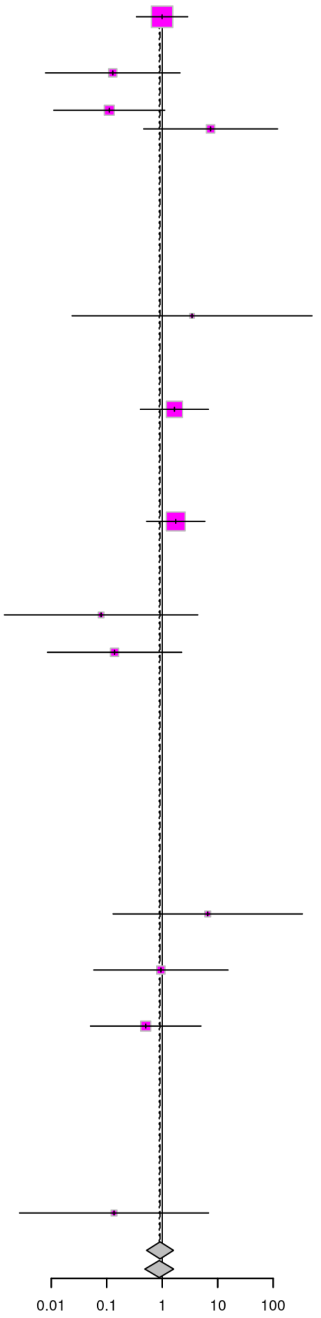

### Maentel Mantel-Haenszel with CC

| Study | Experimental Events | Experimental Total | Control Events | Control Total | OR | 95%-CI | Weight (common) | Weight (random) |
|---|---|---|---|---|---|---|---|---|
| Abbar 2022 | 7 | 73 | 8 | 83 | 0.99 | [0.34; 2.89] | 11.7% | 13.8% |
| Abdallah 2022 | 0 | 108 | 0 | 55 | 0.51 | [0.01; 26.12] | 1.1% | 1.0% |
| Ahmed 2023 | 0 | 18 | 0 | 18 | 1.00 | [0.02; 53.12] | 0.8% | 1.0% |
| Anderson 2017 | 0 | 40 | 2 | 39 | 0.19 | [0.01; 3.98] | 4.3% | 1.7% |
| Burger 2016 | 0 | 3 | 0 | 7 | 2.14 | [0.03; 131.93] | 0.5% | 0.9% |
| Canuso 2018 | 0 | 35 | 3 | 31 | 0.11 | [0.01; 2.31] | 6.3% | 1.7% |
| Chen 2023 | 2 | 126 | 0 | 126 | 5.08 | [0.24; 106.89] | 0.8% | 1.7% |
| Colla 2024 | 0 | 17 | 0 | 10 | 0.60 | [0.01; 32.56] | 1.0% | 1.0% |
| Dakwar 2019 | 0 | 27 | 0 | 28 | 1.04 | [0.02; 54.08] | 0.8% | 1.0% |
| Dakwar 2020 | 0 | 17 | 0 | 23 | 1.34 | [0.03; 71.04] | 0.7% | 1.0% |
| Daly 2018 PA | 0 | 34 | 0 | 33 | 0.97 | [0.02; 50.36] | 0.9% | 1.0% |
| Daly 2018 PB | 0 | 20 | 0 | 21 | 1.05 | [0.02; 55.37] | 0.8% | 1.0% |
| Domany 2019 | 0 | 22 | 0 | 18 | 0.82 | [0.02; 43.47] | 0.9% | 1.0% |
| Domany 2020 | 0 | 9 | 0 | 9 | 1.00 | [0.02; 55.80] | 0.8% | 1.0% |
| Duek 2023 | 0 | 14 | 0 | 14 | 1.00 | [0.02; 53.89] | 0.8% | 1.0% |
| Dwyer 2021 | 0 | 11 | 0 | 6 | 0.57 | [0.01; 32.00] | 1.0% | 1.0% |
| Fava 2020 | 1 | 80 | 0 | 19 | 0.74 | [0.03; 18.76] | 1.4% | 1.5% |
| Feder 2014 | 0 | 22 | 0 | 19 | 0.87 | [0.02; 45.76] | 0.9% | 1.0% |
| Feder 2021 | 0 | 15 | 0 | 15 | 1.00 | [0.02; 53.66] | 0.8% | 1.0% |
| Fedgchin 2019 | 0 | 231 | 0 | 113 | 0.49 | [0.01; 24.87] | 1.2% | 1.0% |
| Fineberg 2023 | 0 | 10 | 0 | 12 | 1.19 | [0.02; 65.32] | 0.8% | 1.0% |
| Fu 2020 | 5 | 113 | 3 | 112 | 1.68 | [0.39; 7.21] | 5.0% | 7.4% |
| George 2017 | 0 | 12 | 0 | 4 | 0.36 | [0.01; 20.98] | 1.2% | 1.0% |
| Glue 2024b | 0 | 17 | 0 | 8 | 0.49 | [0.01; 26.64] | 1.1% | 1.0% |
| Grabski 2022 | 0 | 48 | 0 | 48 | 1.00 | [0.02; 51.42] | 0.9% | 1.0% |
| Grunebaum 2017 | 0 | 9 | 0 | 7 | 0.79 | [0.01; 44.65] | 0.9% | 1.0% |
| Grunebaum 2018 | 0 | 40 | 0 | 40 | 1.00 | [0.02; 51.63] | 0.9% | 1.0% |
| Ionescu 2020 | 7 | 114 | 4 | 113 | 1.78 | [0.51; 6.27] | 6.5% | 9.9% |
| Jones 2024 | 0 | 17 | 0 | 11 | 0.66 | [0.01; 35.52] | 1.0% | 1.0% |
| Lapidus 2014 | 0 | 10 | 0 | 10 | 1.00 | [0.02; 55.27] | 0.8% | 1.0% |
| Leal 2023 | 0 | 5 | 0 | 5 | 1.00 | [0.02; 59.99] | 0.8% | 0.9% |
| Lii 2023 | 0 | 20 | 0 | 20 | 1.00 | [0.02; 52.85] | 0.8% | 1.0% |
| Lijffijt 2021 | 0 | 20 | 1 | 13 | 0.20 | [0.01; 5.39] | 3.0% | 1.5% |
| Loo 2016 | 0 | 9 | 0 | 6 | 0.68 | [0.01; 39.07] | 1.0% | 1.0% |
| Loo 2023 | 0 | 89 | 2 | 92 | 0.20 | [0.01; 4.27] | 4.2% | 1.7% |
| Milak 2020 | 0 | 38 | 0 | 5 | 0.14 | [0.00; 7.96] | 1.5% | 1.0% |
| Murrough 2013 | 0 | 47 | 0 | 25 | 0.54 | [0.01; 27.86] | 1.1% | 1.0% |
| Murrough 2015 II | 0 | 12 | 0 | 12 | 1.00 | [0.02; 54.46] | 0.8% | 1.0% |
| Ochs-Ross 2020 | 0 | 72 | 0 | 65 | 0.90 | [0.02; 46.18] | 0.9% | 1.0% |
| Ohtani 2024 | 0 | 17 | 0 | 17 | 1.00 | [0.02; 53.28] | 0.8% | 1.0% |
| Pattanaseri 2024 | 0 | 11 | 0 | 9 | 0.83 | [0.01; 45.69] | 0.9% | 1.0% |
| Phillips 2019 | 0 | 22 | 0 | 21 | 0.96 | [0.02; 50.34] | 0.9% | 1.0% |
| Popova 2019 | 0 | 115 | 0 | 109 | 0.95 | [0.02; 48.20] | 0.9% | 1.0% |
| Price 2022 | 0 | 53 | 0 | 51 | 0.96 | [0.02; 49.43] | 0.9% | 1.0% |
| Ray-Griffith 2017 | 0 | 8 | 0 | 8 | 1.00 | [0.02; 56.46] | 0.8% | 1.0% |
| Rodriguez 2013 | 0 | 8 | 0 | 7 | 0.88 | [0.02; 50.19] | 0.9% | 1.0% |
| Schiweck 2021 | 0 | 21 | 0 | 7 | 0.35 | [0.01; 19.18] | 1.2% | 1.0% |
| Shiroma 2020 | 0 | 29 | 0 | 25 | 0.86 | [0.02; 45.14] | 0.9% | 1.0% |
| Singh 2016 2x doses | 1 | 18 | 0 | 16 | 2.83 | [0.11; 74.46] | 0.8% | 1.5% |
| Singh 2016 3x doses | 0 | 17 | 0 | 16 | 0.94 | [0.02; 50.32] | 0.9% | 1.0% |
| Singh 2016 II | 0 | 20 | 0 | 10 | 0.51 | [0.01; 27.68] | 1.1% | 1.0% |
| Smith-Apeldoorn 2024 | 1 | 57 | 1 | 54 | 0.95 | [0.06; 15.52] | 1.7% | 2.0% |
| Sos 2013 | 0 | 11 | 0 | 19 | 1.70 | [0.03; 91.39] | 0.6% | 1.0% |
| Su 2017 | 0 | 47 | 0 | 24 | 0.52 | [0.01; 26.79] | 1.1% | 1.0% |
| Su 2023 | 1 | 42 | 2 | 42 | 0.49 | [0.04; 5.59] | 3.4% | 2.6% |
| Sumner 2019 | 0 | 17 | 0 | 16 | 0.94 | [0.02; 50.32] | 0.9% | 1.0% |
| Takahashi 2021 | 0 | 122 | 0 | 80 | 0.66 | [0.01; 33.45] | 1.0% | 1.0% |
| Taylor 2018 | 0 | 9 | 0 | 9 | 1.00 | [0.02; 55.80] | 0.8% | 1.0% |
| Tiger 2020 | 0 | 20 | 0 | 10 | 0.51 | [0.01; 27.68] | 1.1% | 1.0% |
| Wang 2024 | 0 | 182 | 0 | 182 | 1.00 | [0.02; 50.67] | 0.9% | 1.0% |
| Xiao 2023 | 0 | 15 | 0 | 15 | 1.00 | [0.02; 53.66] | 0.8% | 1.0% |
| Yang 2023 | 0 | 200 | 0 | 100 | 0.50 | [0.01; 25.45] | 1.2% | 1.0% |
| Zarate 2006 | 0 | 9 | 0 | 9 | 1.00 | [0.02; 55.80] | 0.8% | 1.0% |
| Zarate 2012 | 0 | 21 | 0 | 23 | 1.09 | [0.02; 57.53] | 0.8% | 1.0% |
| Zhou 2024 | 0 | 27 | 1 | 27 | 0.32 | [0.01; 8.24] | 2.6% | 1.5% |
| **Common effect model** | | **2742** | | **2201** | **0.86** | **[0.59; 1.25]** | **100.0%** | **.** |
| **Random effects model** | | | | | **0.88** | **[0.59; 1.30]** | **.** | **100.0%** |

Heterogeneity: $I^2 = 0.0\%$, $\tau^2 = 0$, $p = 1.0000$

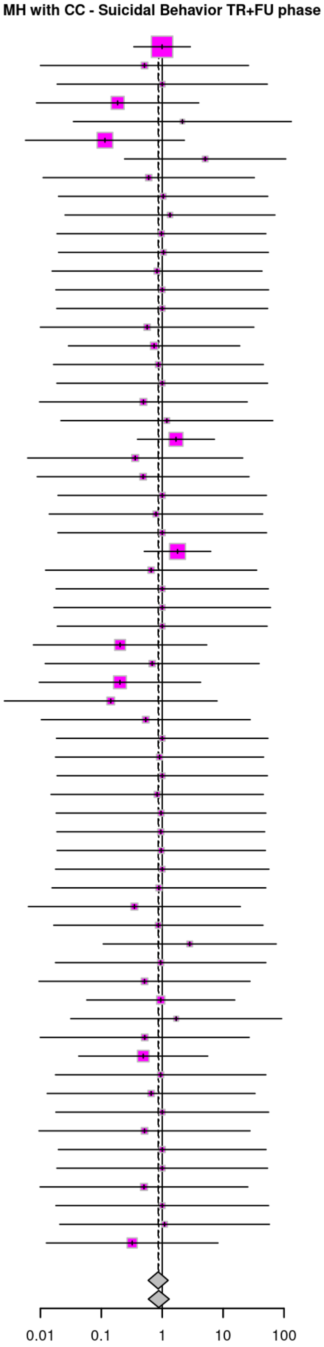

#### Maentel Mantel-Haenszel with Treatment-Arm CC

| Study | Experimental Events | Experimental Total | Control Events | Control Total | MH with TA-CC - Suicidal Behavior TR+FU phase OR | 95%-CI | Weight (common) | Weight (random) |
|---|---|---|---|---|---|---|---|---|
| Abbar 2022 | 7 | 73 | 8 | 83 | 0.99 | [0.34; 2.89] | 12.7% | 14.2% |
| Abdallah 2022 | 0 | 108 | 0 | 55 | 1.00 | [0.02; 63.93] | 0.8% | 0.9% |
| Ahmed 2023 | 0 | 18 | 0 | 18 | 1.00 | [0.02; 53.12] | 0.9% | 1.0% |
| Anderson 2017 | 0 | 40 | 2 | 39 | 0.19 | [0.01; 3.98] | 4.7% | 1.7% |
| Burger 2016 | 0 | 3 | 0 | 7 | 1.00 | [0.01; 87.11] | 0.7% | 0.8% |
| Canuso 2018 | 0 | 35 | 3 | 31 | 0.12 | [0.01; 2.30] | 6.8% | 1.9% |
| Chen 2023 | 2 | 126 | 0 | 126 | 5.08 | [0.24; 106.89] | 0.9% | 1.7% |
| Colla 2024 | 0 | 17 | 0 | 10 | 1.00 | [0.02; 62.21] | 0.8% | 1.0% |
| Dakwar 2019 | 0 | 27 | 0 | 28 | 1.00 | [0.02; 52.22] | 0.9% | 1.0% |
| Dakwar 2020 | 0 | 17 | 0 | 23 | 1.00 | [0.02; 55.30] | 0.9% | 1.0% |
| Daly 2018 PA | 0 | 34 | 0 | 33 | 1.00 | [0.02; 51.89] | 0.9% | 1.0% |
| Daly 2018 PB | 0 | 20 | 0 | 21 | 1.00 | [0.02; 52.85] | 0.9% | 1.0% |
| Domany 2019 | 0 | 22 | 0 | 18 | 1.00 | [0.02; 53.92] | 0.9% | 1.0% |
| Domany 2020 | 0 | 9 | 0 | 9 | 1.00 | [0.02; 55.80] | 0.9% | 1.0% |
| Duek 2023 | 0 | 14 | 0 | 14 | 1.00 | [0.02; 53.89] | 0.9% | 1.0% |
| Dwyer 2021 | 0 | 11 | 0 | 6 | 1.00 | [0.01; 67.61] | 0.8% | 0.9% |
| Fava 2020 | 1 | 80 | 0 | 19 | 2.27 | [0.02; 257.11] | 0.6% | 0.7% |
| Feder 2014 | 0 | 22 | 0 | 19 | 1.00 | [0.02; 53.36] | 0.9% | 1.0% |
| Feder 2021 | 0 | 15 | 0 | 15 | 1.00 | [0.02; 53.66] | 0.9% | 1.0% |
| Fedgchin 2019 | 0 | 231 | 0 | 113 | 1.00 | [0.02; 65.31] | 0.8% | 0.9% |
| Fineberg 2023 | 0 | 10 | 0 | 12 | 1.00 | [0.02; 55.75] | 0.9% | 1.0% |
| Fu 2020 | 5 | 113 | 3 | 112 | 1.68 | [0.39; 7.21] | 5.4% | 7.6% |
| George 2017 | 0 | 12 | 0 | 4 | 1.00 | [0.01; 105.38] | 0.7% | 0.7% |
| Glue 2024b | 0 | 17 | 0 | 8 | 1.00 | [0.01; 72.36] | 0.8% | 0.9% |
| Grabski 2022 | 0 | 48 | 0 | 48 | 1.00 | [0.02; 51.42] | 0.9% | 1.0% |
| Grunebaum 2017 | 0 | 9 | 0 | 7 | 1.00 | [0.02; 58.29] | 0.9% | 1.0% |
| Grunebaum 2018 | 0 | 40 | 0 | 40 | 1.00 | [0.02; 51.63] | 0.9% | 1.0% |
| Ionescu 2020 | 7 | 114 | 4 | 113 | 1.78 | [0.51; 6.27] | 7.1% | 10.3% |
| Jones 2024 | 0 | 17 | 0 | 11 | 1.00 | [0.02; 59.25] | 0.9% | 1.0% |
| Lapidus 2014 | 0 | 10 | 0 | 10 | 1.00 | [0.02; 55.27] | 0.9% | 1.0% |
| Leal 2023 | 0 | 5 | 0 | 5 | 1.00 | [0.02; 59.99] | 0.9% | 1.0% |
| Lii 2023 | 0 | 20 | 0 | 20 | 1.00 | [0.02; 52.85] | 0.9% | 1.0% |
| Lijffijt 2021 | 0 | 20 | 1 | 13 | 0.26 | [0.01; 5.79] | 3.1% | 1.7% |
| Loo 2016 | 0 | 9 | 0 | 6 | 1.00 | [0.02; 61.80] | 0.8% | 1.0% |
| Loo 2023 | 0 | 89 | 2 | 92 | 0.20 | [0.01; 4.27] | 4.6% | 1.7% |
| Milak 2020 | 0 | 38 | 0 | 5 | 1.00 | [0.00; 484.58] | 0.4% | 0.4% |
| Murrough 2013 | 0 | 47 | 0 | 25 | 1.00 | [0.02; 63.11] | 0.8% | 0.9% |
| Murrough 2015 II | 0 | 12 | 0 | 12 | 1.00 | [0.02; 54.46] | 0.9% | 1.0% |
| Ochs-Ross 2020 | 0 | 72 | 0 | 65 | 1.00 | [0.02; 51.38] | 0.9% | 1.0% |
| Ohtani 2024 | 0 | 17 | 0 | 17 | 1.00 | [0.02; 53.28] | 0.9% | 1.0% |
| Pattanaseri 2024 | 0 | 11 | 0 | 9 | 1.00 | [0.02; 56.40] | 0.9% | 1.0% |
| Phillips 2019 | 0 | 22 | 0 | 21 | 1.00 | [0.02; 52.74] | 0.9% | 1.0% |
| Popova 2019 | 0 | 115 | 0 | 109 | 1.00 | [0.02; 50.91] | 0.9% | 1.0% |
| Price 2022 | 0 | 53 | 0 | 51 | 1.00 | [0.02; 51.38] | 0.9% | 1.0% |
| Ray-Griffith 2017 | 0 | 8 | 0 | 8 | 1.00 | [0.02; 56.46] | 0.9% | 1.0% |
| Rodriguez 2013 | 0 | 8 | 0 | 7 | 1.00 | [0.02; 57.37] | 0.9% | 1.0% |
| Schiweck 2021 | 0 | 21 | 0 | 7 | 1.00 | [0.01; 99.86] | 0.7% | 0.8% |
| Shiroma 2020 | 0 | 29 | 0 | 25 | 1.00 | [0.02; 52.79] | 0.9% | 1.0% |
| Singh 2016 2x doses | 1 | 18 | 0 | 16 | 3.05 | [0.11; 85.81] | 0.9% | 1.5% |
| Singh 2016 3x doses | 0 | 17 | 0 | 16 | 1.00 | [0.02; 53.46] | 0.9% | 1.0% |
| Singh 2016 II | 0 | 20 | 0 | 10 | 1.00 | [0.01; 68.32] | 0.8% | 0.9% |
| Smith-Apeldoorn 2024 | 1 | 57 | 1 | 54 | 0.95 | [0.06; 15.52] | 1.9% | 2.1% |
| Sos 2013 | 0 | 11 | 0 | 19 | 1.00 | [0.02; 62.32] | 0.8% | 0.9% |
| Su 2017 | 0 | 47 | 0 | 24 | 1.00 | [0.02; 64.85] | 0.8% | 0.9% |
| Su 2023 | 1 | 42 | 2 | 42 | 0.49 | [0.04; 5.59] | 3.7% | 2.7% |
| Sumner 2019 | 0 | 17 | 0 | 16 | 1.00 | [0.02; 53.46] | 0.9% | 1.0% |
| Takahashi 2021 | 0 | 122 | 0 | 80 | 1.00 | [0.02; 55.55] | 0.9% | 1.0% |
| Taylor 2018 | 0 | 9 | 0 | 9 | 1.00 | [0.02; 55.80] | 0.9% | 1.0% |
| Tiger 2020 | 0 | 20 | 0 | 10 | 1.00 | [0.01; 68.32] | 0.8% | 0.9% |
| Wang 2024 | 0 | 182 | 0 | 182 | 1.00 | [0.02; 50.67] | 0.9% | 1.1% |
| Xiao 2023 | 0 | 15 | 0 | 15 | 1.00 | [0.02; 53.66] | 0.9% | 1.0% |
| Yang 2023 | 0 | 200 | 0 | 100 | 1.00 | [0.02; 64.37] | 0.8% | 0.9% |
| Zarate 2006 | 0 | 9 | 0 | 9 | 1.00 | [0.02; 55.80] | 0.9% | 1.0% |
| Zarate 2012 | 0 | 21 | 0 | 23 | 1.00 | [0.02; 52.84] | 0.9% | 1.0% |
| Zhou 2024 | 0 | 27 | 1 | 27 | 0.32 | [0.01; 8.24] | 2.8% | 1.5% |
| **Common effect model** | | **2742** | | **2201** | **0.96** | **[0.65; 1.41]** | **100.0%** | **.** |
| **Random effects model** | | | | | **0.99** | **[0.66; 1.48]** | **.** | **100.0%** |

Heterogeneity: $I^2 = 0.0\%$, $\tau^2 = 0$, $p = 1.0000$

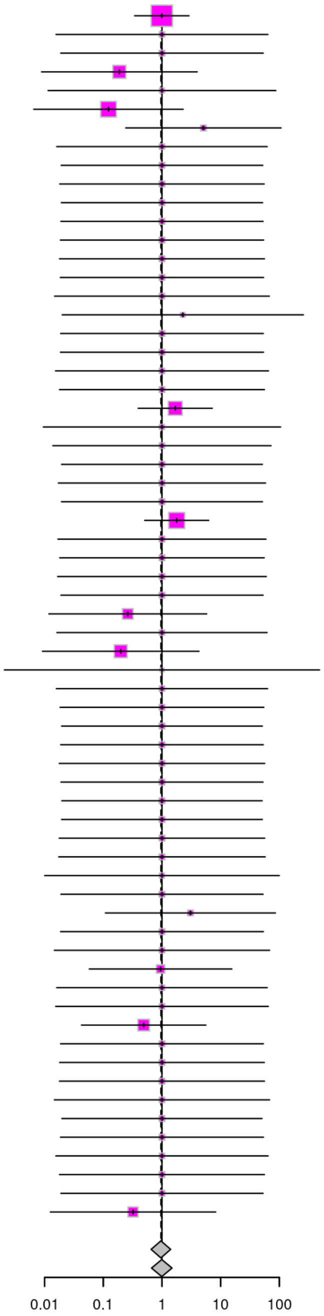

### Maentel Mantel-Haenszel Risk-Difference

| Study | Experimental Events | Experimental Total | Control Events | Control Total | MH RD - Suicidal Behavior TR+FU phase | RD | 95%-CI | Weight (common) | Weight (random) |
|---|---|---|---|---|---|---|---|---|---|
| Abbar 2022 | 7 | 73 | 8 | 83 | | -0.00 | [-0.09; 0.09] | 3.2% | 0.3% |
| Abdallah 2022 | 0 | 108 | 0 | 55 | | 0.00 | [-0.03; 0.03] | 3.0% | 3.6% |
| Ahmed 2023 | 0 | 18 | 0 | 18 | | 0.00 | [-0.10; 0.10] | 0.8% | 0.3% |
| Anderson 2017 | 0 | 40 | 2 | 39 | | -0.05 | [-0.13; 0.03] | 1.7% | 0.4% |
| Burger 2016 | 0 | 3 | 0 | 7 | | 0.00 | [-0.36; 0.36] | 0.2% | 0.0% |
| Canuso 2018 | 0 | 35 | 3 | 31 | | -0.10 | [-0.21; 0.02] | 1.4% | 0.2% |
| Chen 2023 | 2 | 126 | 0 | 126 | | 0.02 | [-0.01; 0.04] | 5.2% | 3.9% |
| Colla 2024 | 0 | 17 | 0 | 10 | | 0.00 | [-0.14; 0.14] | 0.6% | 0.1% |
| Dakwar 2019 | 0 | 27 | 0 | 28 | | 0.00 | [-0.07; 0.07] | 1.2% | 0.6% |
| Dakwar 2020 | 0 | 17 | 0 | 23 | | 0.00 | [-0.10; 0.10] | 0.8% | 0.3% |
| Daly 2018 PA | 0 | 34 | 0 | 33 | | 0.00 | [-0.06; 0.06] | 1.4% | 0.9% |
| Daly 2018 PB | 0 | 20 | 0 | 21 | | 0.00 | [-0.09; 0.09] | 0.9% | 0.3% |
| Domany 2019 | 0 | 22 | 0 | 18 | | 0.00 | [-0.09; 0.09] | 0.9% | 0.3% |
| Domany 2020 | 0 | 9 | 0 | 9 | | 0.00 | [-0.19; 0.19] | 0.4% | 0.1% |
| Duek 2023 | 0 | 14 | 0 | 14 | | 0.00 | [-0.13; 0.13] | 0.6% | 0.2% |
| Dwyer 2021 | 0 | 11 | 0 | 6 | | 0.00 | [-0.22; 0.22] | 0.4% | 0.1% |
| Fava 2020 | 1 | 80 | 0 | 19 | | 0.01 | [-0.06; 0.09] | 1.3% | 0.5% |
| Feder 2014 | 0 | 22 | 0 | 19 | | 0.00 | [-0.09; 0.09] | 0.9% | 0.3% |
| Feder 2021 | 0 | 15 | 0 | 15 | | 0.00 | [-0.12; 0.12] | 0.7% | 0.2% |
| Fedgchin 2019 | 0 | 231 | 0 | 113 | | 0.00 | [-0.01; 0.01] | 6.3% | 15.1% |
| Fineberg 2023 | 0 | 10 | 0 | 12 | | 0.00 | [-0.16; 0.16] | 0.5% | 0.1% |
| Fu 2020 | 5 | 113 | 3 | 112 | | 0.02 | [-0.03; 0.07] | 4.6% | 1.2% |
| George 2017 | 0 | 12 | 0 | 4 | | 0.00 | [-0.28; 0.28] | 0.3% | 0.0% |
| Glue 2024b | 0 | 17 | 0 | 8 | | 0.00 | [-0.17; 0.17] | 0.5% | 0.1% |
| Grabski 2022 | 0 | 48 | 0 | 48 | | 0.00 | [-0.04; 0.04] | 2.0% | 1.7% |
| Grunebaum 2017 | 0 | 9 | 0 | 7 | | 0.00 | [-0.22; 0.22] | 0.4% | 0.1% |
| Grunebaum 2018 | 0 | 40 | 0 | 40 | | 0.00 | [-0.05; 0.05] | 1.7% | 1.2% |
| Ionescu 2020 | 7 | 114 | 4 | 113 | | 0.03 | [-0.03; 0.08] | 4.7% | 0.9% |
| Jones 2024 | 0 | 17 | 0 | 11 | | 0.00 | [-0.14; 0.14] | 0.6% | 0.1% |
| Lapidus 2014 | 0 | 10 | 0 | 10 | | 0.00 | [-0.17; 0.17] | 0.5% | 0.1% |
| Leal 2023 | 0 | 5 | 0 | 5 | | 0.00 | [-0.31; 0.31] | 0.2% | 0.0% |
| Lii 2023 | 0 | 20 | 0 | 20 | | 0.00 | [-0.09; 0.09] | 0.9% | 0.3% |
| Lijffijt 2021 | 0 | 20 | 1 | 13 | | -0.08 | [-0.25; 0.10] | 0.7% | 0.1% |
| Loo 2016 | 0 | 9 | 0 | 6 | | 0.00 | [-0.23; 0.23] | 0.3% | 0.1% |
| Loo 2023 | 0 | 89 | 2 | 92 | | -0.02 | [-0.06; 0.01] | 3.8% | 2.1% |
| Milak 2020 | 0 | 38 | 0 | 5 | | 0.00 | [-0.22; 0.22] | 0.4% | 0.1% |
| Murrough 2013 | 0 | 47 | 0 | 25 | | 0.00 | [-0.06; 0.06] | 1.4% | 0.8% |
| Murrough 2015 II | 0 | 12 | 0 | 12 | | 0.00 | [-0.15; 0.15] | 0.5% | 0.1% |
| Ochs-Ross 2020 | 0 | 72 | 0 | 65 | | 0.00 | [-0.03; 0.03] | 2.9% | 3.5% |
| Ohtani 2024 | 0 | 17 | 0 | 17 | | 0.00 | [-0.11; 0.11] | 0.7% | 0.2% |
| Pattanaseri 2024 | 0 | 11 | 0 | 9 | | 0.00 | [-0.18; 0.18] | 0.4% | 0.1% |
| Phillips 2019 | 0 | 22 | 0 | 21 | | 0.00 | [-0.09; 0.09] | 0.9% | 0.4% |
| Popova 2019 | 0 | 115 | 0 | 109 | | 0.00 | [-0.02; 0.02] | 4.7% | 9.2% |
| Price 2022 | 0 | 53 | 0 | 51 | | 0.00 | [-0.04; 0.04] | 2.2% | 2.0% |
| Ray-Griffith 2017 | 0 | 8 | 0 | 8 | | 0.00 | [-0.21; 0.21] | 0.4% | 0.1% |
| Rodriguez 2013 | 0 | 8 | 0 | 7 | | 0.00 | [-0.22; 0.22] | 0.3% | 0.1% |
| Schiweck 2021 | 0 | 21 | 0 | 7 | | 0.00 | [-0.18; 0.18] | 0.5% | 0.1% |
| Shiroma 2020 | 0 | 29 | 0 | 25 | | 0.00 | [-0.07; 0.07] | 1.1% | 0.6% |
| Singh 2016 2x doses | 1 | 18 | 0 | 16 | | 0.06 | [-0.09; 0.20] | 0.7% | 0.1% |
| Singh 2016 3x doses | 0 | 17 | 0 | 16 | | 0.00 | [-0.11; 0.11] | 0.7% | 0.2% |
| Singh 2016 II | 0 | 20 | 0 | 10 | | 0.00 | [-0.14; 0.14] | 0.6% | 0.1% |
| Smith-Apeldoorn 2024 | 1 | 57 | 1 | 54 | | -0.00 | [-0.05; 0.05] | 2.3% | 1.1% |
| Sos 2013 | 0 | 11 | 0 | 19 | | 0.00 | [-0.13; 0.13] | 0.6% | 0.2% |
| Su 2017 | 0 | 47 | 0 | 24 | | 0.00 | [-0.06; 0.06] | 1.4% | 0.7% |
| Su 2023 | 1 | 42 | 2 | 42 | | -0.02 | [-0.10; 0.06] | 1.7% | 0.4% |
| Sumner 2019 | 0 | 17 | 0 | 16 | | 0.00 | [-0.11; 0.11] | 0.7% | 0.2% |
| Takahashi 2021 | 0 | 122 | 0 | 80 | | 0.00 | [-0.02; 0.02] | 4.0% | 6.6% |
| Taylor 2018 | 0 | 9 | 0 | 9 | | 0.00 | [-0.19; 0.19] | 0.4% | 0.1% |
| Tiger 2020 | 0 | 20 | 0 | 10 | | 0.00 | [-0.14; 0.14] | 0.6% | 0.1% |
| Wang 2024 | 0 | 182 | 0 | 182 | | 0.00 | [-0.01; 0.01] | 7.5% | 24.2% |
| Xiao 2023 | 0 | 15 | 0 | 15 | | 0.00 | [-0.12; 0.12] | 0.7% | 0.2% |
| Yang 2023 | 0 | 200 | 0 | 100 | | 0.00 | [-0.02; 0.02] | 5.5% | 11.8% |
| Zarate 2006 | 0 | 9 | 0 | 9 | | 0.00 | [-0.19; 0.19] | 0.4% | 0.1% |
| Zarate 2012 | 0 | 21 | 0 | 23 | | 0.00 | [-0.08; 0.08] | 0.9% | 0.4% |
| Zhou 2024 | 0 | 27 | 1 | 27 | | -0.04 | [-0.13; 0.06] | 1.2% | 0.3% |
| **Common effect model** | | **2742** | | **2201** | | **-0.00** | **[-0.01; 0.01]** | **100.0%** | **.** |
| **Random effects model** | | | | | | **0.00** | **[-0.01; 0.01]** | **.** | **100.0%** |

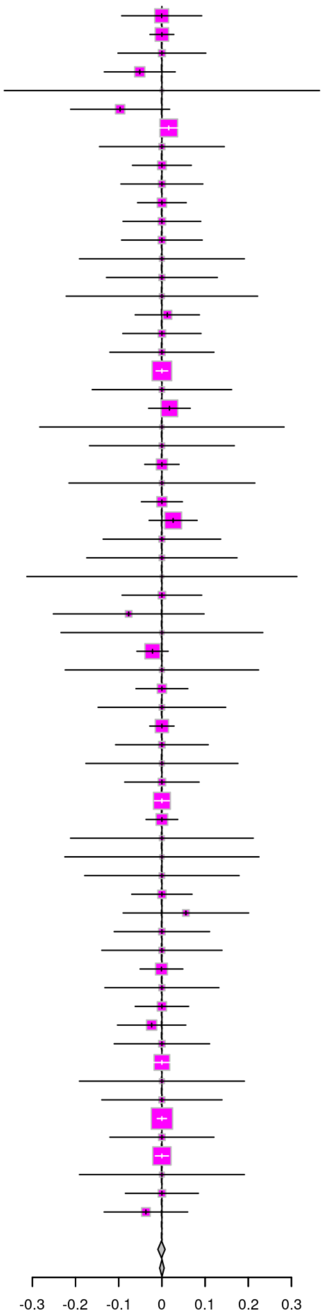

Heterogeneity: $I^2 = 0.0\%$, $\tau^2 = 0$, $p = 1.0000$

#### Arcsine Risk-Difference

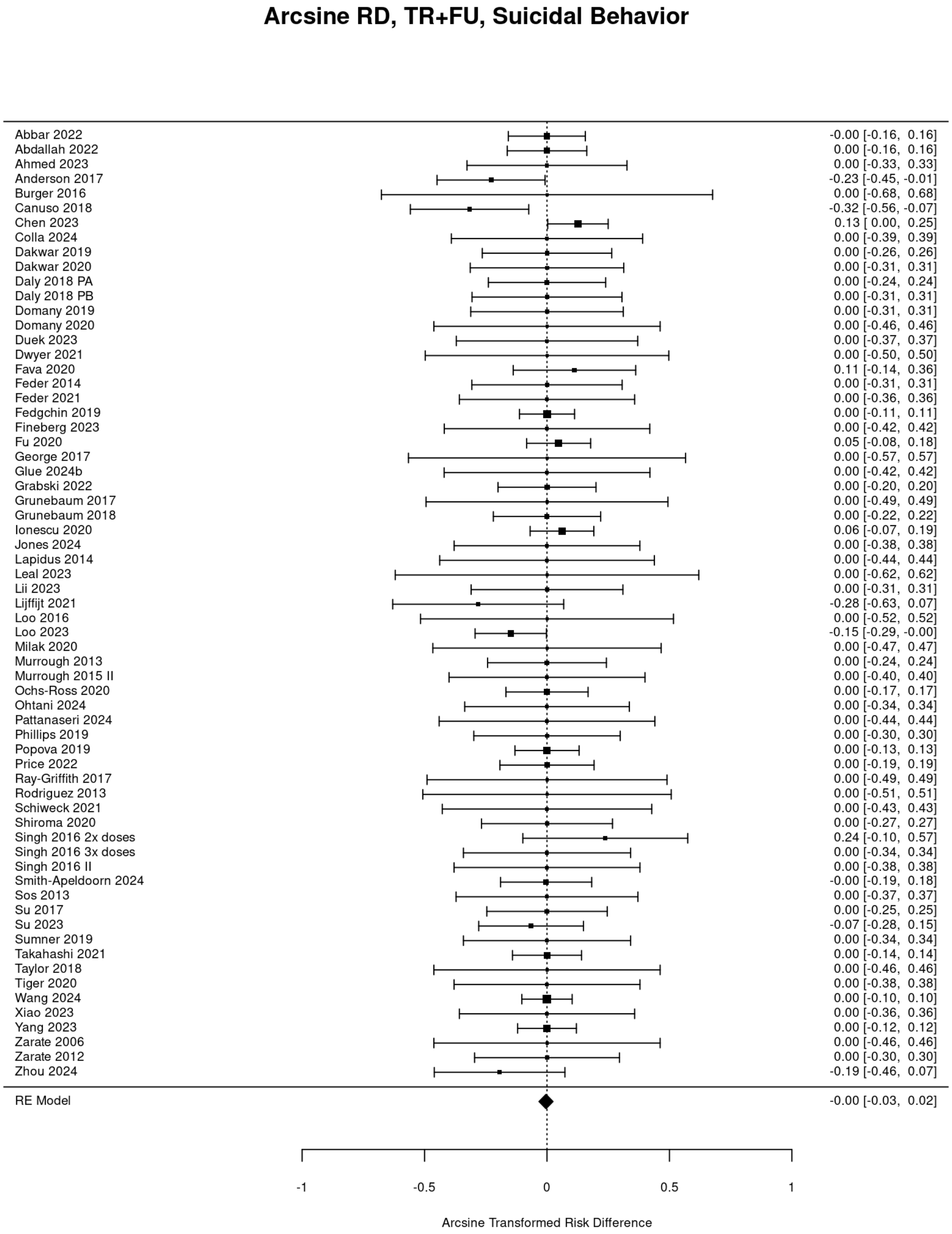

##### Bayesian Meta-Analysis, wider prior

**Bayesian MA, Delta = 250 - Suicidal Behavior TR+FU**

| Study | Estimate | 95% CI |
|---|---|---|
| Abbar 2022 | -0.01 | [-1.07, 1.06] |
| Abdallah 2022 | -0.67 | [-4.60, 3.26] |
| Ahmed 2023 | 0.00 | [-3.97, 3.97] |
| Anderson 2017 | -1.69 | [-4.76, 1.38] |
| Burger 2016 | 0.76 | [-3.36, 4.88] |
| Canuso 2018 | -2.17 | [-5.17, 0.84] |
| Chen 2023 | 1.63 | [-1.42, 4.67] |
| Colla 2024 | -0.51 | [-4.50, 3.48] |
| Dakwar 2019 | 0.04 | [-3.92, 3.99] |
| Dakwar 2020 | 0.29 | [-3.67, 4.26] |
| Daly 2018 PA | -0.03 | [-3.98, 3.92] |
| Daly 2018 PB | 0.05 | [-3.92, 4.01] |
| Domany 2019 | -0.20 | [-4.16, 3.77] |
| Domany 2020 | 0.00 | [-4.02, 4.02] |
| Duek 2023 | 0.00 | [-3.99, 3.99] |
| Dwyer 2021 | -0.57 | [-4.61, 3.47] |
| Fava 2020 | -0.31 | [-3.55, 2.93] |
| Feder 2014 | -0.14 | [-4.11, 3.82] |
| Feder 2021 | 0.00 | [-3.98, 3.98] |
| Fedgchin 2019 | -0.71 | [-4.64, 3.21] |
| Fineberg 2023 | 0.17 | [-3.83, 4.18] |
| Fu 2020 | 0.52 | [-0.94, 1.98] |
| George 2017 | -1.02 | [-5.09, 3.04] |
| Glue 2024b | -0.72 | [-4.73, 3.28] |
| Grabski 2022 | 0.00 | [-3.94, 3.94] |
| Grunebaum 2017 | -0.24 | [-4.27, 3.80] |
| Grunebaum 2018 | 0.00 | [-3.94, 3.94] |
| Ionescu 2020 | 0.58 | [-0.68, 1.84] |
| Jones 2024 | -0.42 | [-4.41, 3.57] |
| Lapidus 2014 | 0.00 | [-4.01, 4.01] |
| Leal 2023 | 0.00 | [-4.09, 4.09] |
| Lii 2023 | 0.00 | [-3.97, 3.97] |
| Lijffijt 2021 | -1.59 | [-4.87, 1.68] |
| Loo 2016 | -0.38 | [-4.42, 3.67] |
| Loo 2023 | -1.60 | [-4.65, 1.45] |
| Milak 2020 | -1.95 | [-5.97, 2.07] |
| Murrough 2013 | -0.62 | [-4.57, 3.33] |
| Murrough 2015 II | 0.00 | [-4.00, 4.00] |
| Ochs-Ross 2020 | -0.10 | [-4.04, 3.83] |
| Ohtani 2024 | 0.00 | [-3.98, 3.98] |
| Pattanaseri 2024 | -0.19 | [-4.20, 3.82] |
| Phillips 2019 | -0.05 | [-4.01, 3.92] |
| Popova 2019 | -0.05 | [-3.98, 3.88] |
| Price 2022 | -0.04 | [-3.98, 3.90] |
| Ray-Griffith 2017 | 0.00 | [-4.03, 4.03] |
| Rodriguez 2013 | -0.13 | [-4.17, 3.92] |
| Schiweck 2021 | -1.05 | [-5.06, 2.95] |
| Shiroma 2020 | -0.15 | [-4.10, 3.81] |
| Singh 2016 2x doses | 1.04 | [-2.23, 4.31] |
| Singh 2016 3x doses | -0.06 | [-4.04, 3.92] |
| Singh 2016 II | -0.67 | [-4.66, 3.32] |
| Smith-Apeldoorn 2024 | -0.06 | [-2.85, 2.74] |
| Sos 2013 | 0.53 | [-3.46, 4.52] |
| Su 2017 | -0.66 | [-4.61, 3.29] |
| Su 2023 | -0.72 | [-3.16, 1.72] |
| Sumner 2019 | -0.06 | [-4.04, 3.92] |
| Takahashi 2021 | -0.42 | [-4.35, 3.51] |
| Taylor 2018 | 0.00 | [-4.02, 4.02] |
| Tiger 2020 | -0.67 | [-4.66, 3.32] |
| Wang 2024 | 0.00 | [-3.93, 3.93] |
| Xiao 2023 | 0.00 | [-3.98, 3.98] |
| Yang 2023 | -0.69 | [-4.62, 3.24] |
| Zarate 2006 | 0.00 | [-4.02, 4.02] |
| Zarate 2012 | 0.09 | [-3.87, 4.05] |
| Zhou 2024 | -1.14 | [-4.38, 2.11] |
| **Summary** | **-0.16** | **[-0.80, 0.44]** |
| **Prediction** | **-0.14** | **[-1.29, 0.84]** |

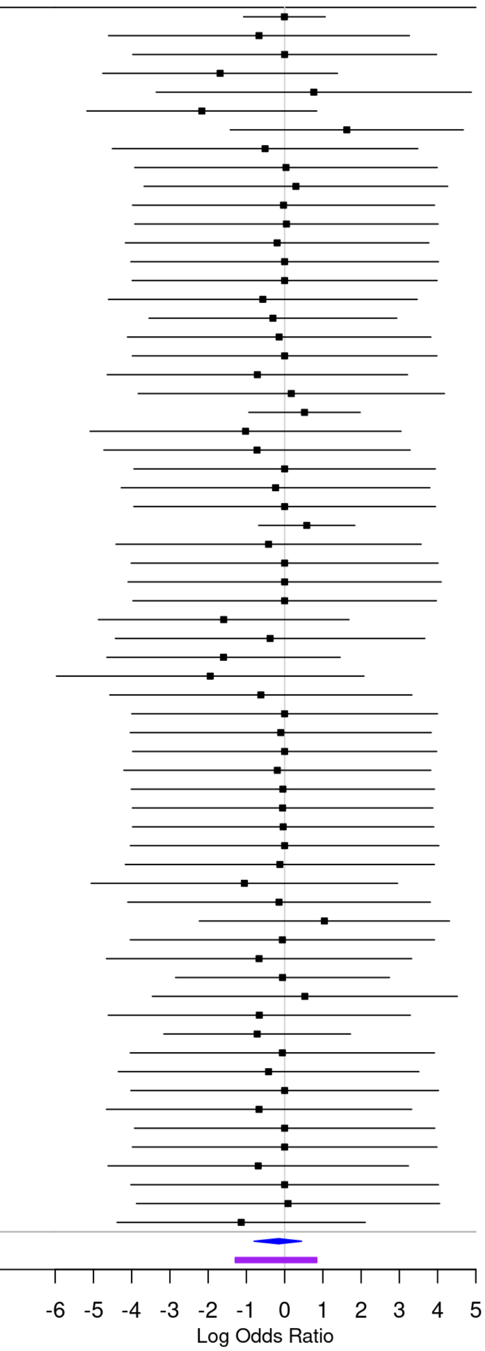

*Heterogeneity (tau): 0.29 [0.00, 0.83]*

**Bayesian Meta-Analysis, smaller prior**

**Bayesian MA, Delta = 15 - Suicidal Behavior TR+FU**

| Study | Estimate | 95% CI |
|---|---|---|
| Abbar 2022 | -0.01 | [-1.07, 1.06] |
| Abdallah 2022 | -0.67 | [-4.60, 3.26] |
| Ahmed 2023 | 0.00 | [-3.97, 3.97] |
| Anderson 2017 | -1.69 | [-4.76, 1.38] |
| Burger 2016 | 0.76 | [-3.36, 4.88] |
| Canuso 2018 | -2.17 | [-5.17, 0.84] |
| Chen 2023 | 1.63 | [-1.42, 4.67] |
| Colla 2024 | -0.51 | [-4.50, 3.48] |
| Dakwar 2019 | 0.04 | [-3.92, 3.99] |
| Dakwar 2020 | 0.29 | [-3.67, 4.26] |
| Daly 2018 PA | -0.03 | [-3.98, 3.92] |
| Daly 2018 PB | 0.05 | [-3.92, 4.01] |
| Domany 2019 | -0.20 | [-4.16, 3.77] |
| Domany 2020 | 0.00 | [-4.02, 4.02] |
| Duek 2023 | 0.00 | [-3.99, 3.99] |
| Dwyer 2021 | -0.57 | [-4.61, 3.47] |
| Fava 2020 | -0.31 | [-3.55, 2.93] |
| Feder 2014 | -0.14 | [-4.11, 3.82] |
| Feder 2021 | 0.00 | [-3.98, 3.98] |
| Fedgchin 2019 | -0.71 | [-4.64, 3.21] |
| Fineberg 2023 | 0.17 | [-3.83, 4.18] |
| Fu 2020 | 0.52 | [-0.94, 1.98] |
| George 2017 | -1.02 | [-5.09, 3.04] |
| Glue 2024b | -0.72 | [-4.73, 3.28] |
| Grabski 2022 | 0.00 | [-3.94, 3.94] |
| Grunebaum 2017 | -0.24 | [-4.27, 3.80] |
| Grunebaum 2018 | 0.00 | [-3.94, 3.94] |
| Ionescu 2020 | 0.58 | [-0.68, 1.84] |
| Jones 2024 | -0.42 | [-4.41, 3.57] |
| Lapidus 2014 | 0.00 | [-4.01, 4.01] |
| Leal 2023 | 0.00 | [-4.09, 4.09] |
| Lii 2023 | 0.00 | [-3.97, 3.97] |
| Lijffijt 2021 | -1.59 | [-4.87, 1.68] |
| Loo 2016 | -0.38 | [-4.42, 3.67] |
| Loo 2023 | -1.60 | [-4.65, 1.45] |
| Milak 2020 | -1.95 | [-5.97, 2.07] |
| Murrough 2013 | -0.62 | [-4.57, 3.33] |
| Murrough 2015 II | 0.00 | [-4.00, 4.00] |
| Ochs-Ross 2020 | -0.10 | [-4.04, 3.83] |
| Ohtani 2024 | 0.00 | [-3.98, 3.98] |
| Pattanaseri 2024 | -0.19 | [-4.20, 3.82] |
| Phillips 2019 | -0.05 | [-4.01, 3.92] |
| Popova 2019 | -0.05 | [-3.98, 3.88] |
| Price 2022 | -0.04 | [-3.98, 3.90] |
| Ray-Griffith 2017 | 0.00 | [-4.03, 4.03] |
| Rodriguez 2013 | -0.13 | [-4.17, 3.92] |
| Schiweck 2021 | -1.05 | [-5.06, 2.95] |
| Shiroma 2020 | -0.15 | [-4.10, 3.81] |
| Singh 2016 2x doses | 1.04 | [-2.23, 4.31] |
| Singh 2016 3x doses | -0.06 | [-4.04, 3.92] |
| Singh 2016 II | -0.67 | [-4.66, 3.32] |
| Smith-Apeldoorn 2024 | -0.06 | [-2.85, 2.74] |
| Sos 2013 | 0.53 | [-3.46, 4.52] |
| Su 2017 | -0.66 | [-4.61, 3.29] |
| Su 2023 | -0.72 | [-3.16, 1.72] |
| Sumner 2019 | -0.06 | [-4.04, 3.92] |
| Takahashi 2021 | -0.42 | [-4.35, 3.51] |
| Taylor 2018 | 0.00 | [-4.02, 4.02] |
| Tiger 2020 | -0.67 | [-4.66, 3.32] |
| Wang 2024 | 0.00 | [-3.93, 3.93] |
| Xiao 2023 | 0.00 | [-3.98, 3.98] |
| Yang 2023 | -0.69 | [-4.62, 3.24] |
| Zarate 2006 | 0.00 | [-4.02, 4.02] |
| Zarate 2012 | 0.09 | [-3.87, 4.05] |
| Zhou 2024 | -1.14 | [-4.38, 2.11] |
| **Summary** | **-0.15** | **[-0.78, 0.43]** |
| **Prediction** | **-0.13** | **[-1.29, 0.88]** |

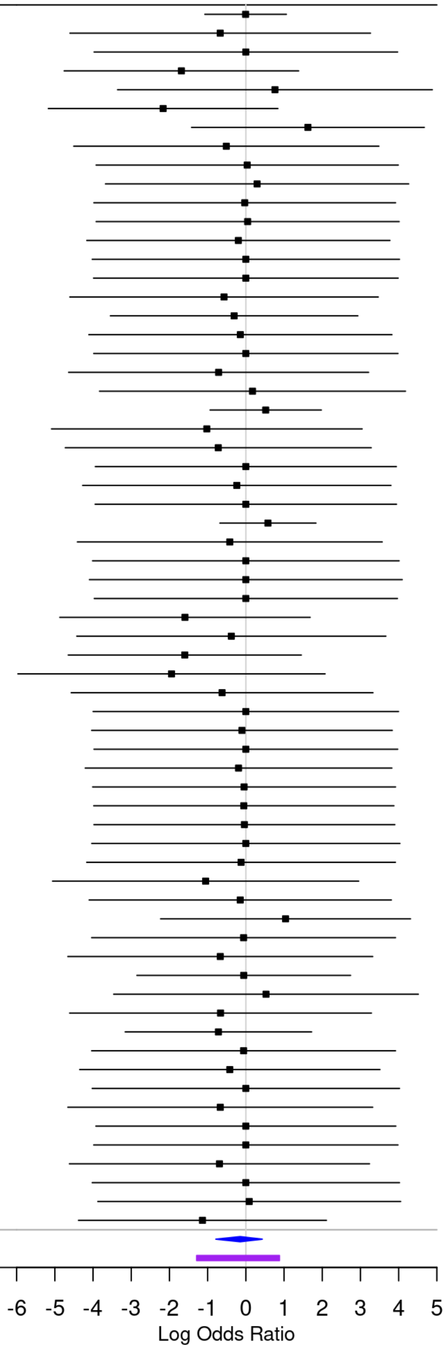

*Heterogeneity (tau): 0.29 [0.00, 0.83]*

**Exact Method**

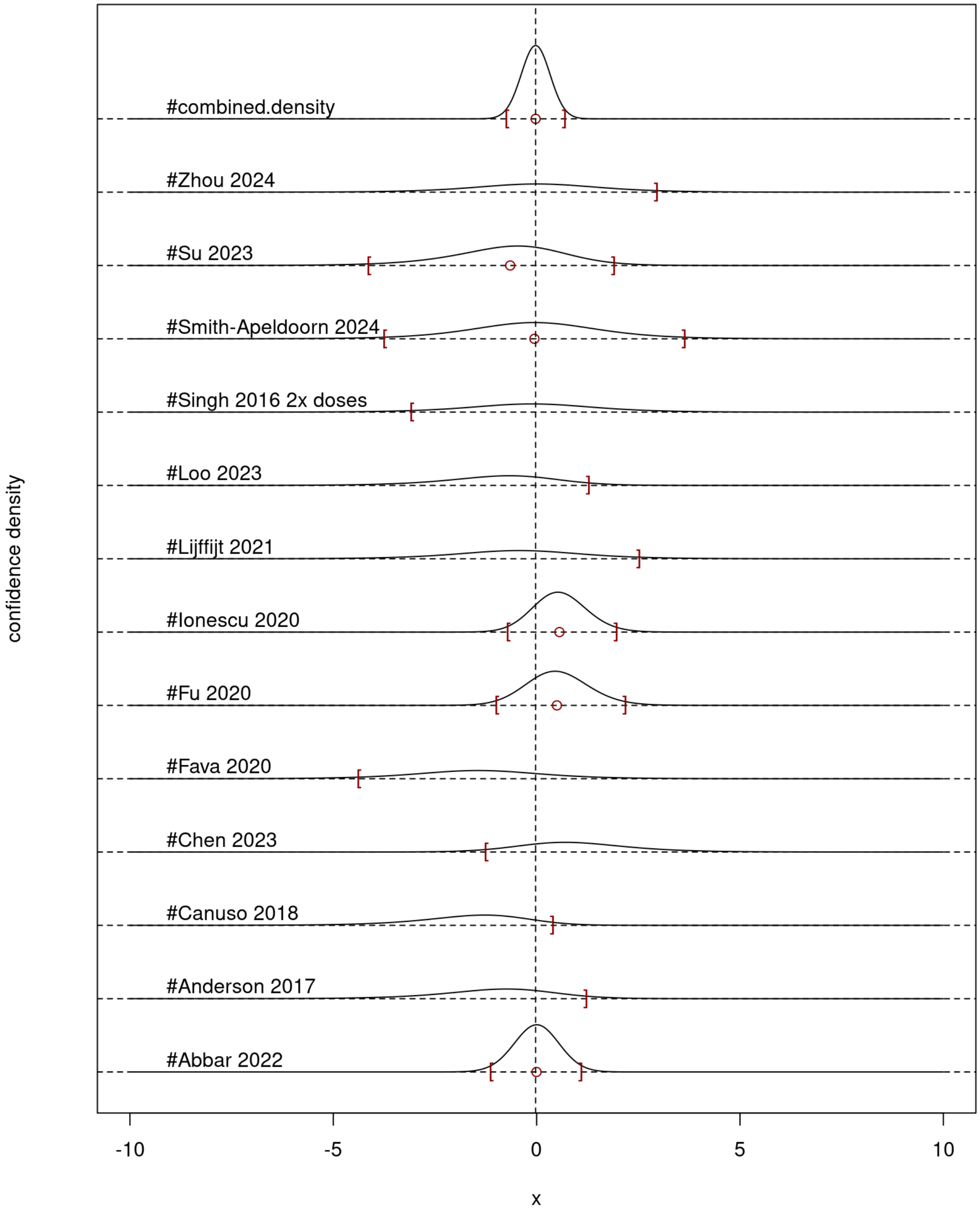

### Suicide Attempts

#### Treatment Phase

#### Peto's Method

| Study | Experimental Events | Experimental Total | Control Events | Control Total | OR | 95%-CI | Weight (common) | Weight (random) |
|---|---|---|---|---|---|---|---|---|
| Abbar 2022 | 1 | 73 | 0 | 73 | 7.39 | [0.15; 372.38] | 6.7% | 6.7% |
| Abdallah 2022 | 0 | 108 | 0 | 108 | | | 0.0% | 0.0% |
| Ahmed 2023 | 0 | 18 | 0 | 18 | | | 0.0% | 0.0% |
| Anderson 2017 | 0 | 40 | 1 | 40 | 0.14 | [0.00; 6.82] | 6.7% | 6.7% |
| Burger 2016 | 0 | 3 | 0 | 3 | | | 0.0% | 0.0% |
| Canuso 2018 | 0 | 35 | 0 | 35 | | | 0.0% | 0.0% |
| Chen 2023 | 1 | 126 | 0 | 126 | 7.39 | [0.15; 372.38] | 6.7% | 6.7% |
| Colla 2024 | 0 | 17 | 0 | 17 | | | 0.0% | 0.0% |
| Dakwar 2019 | 0 | 27 | 0 | 27 | | | 0.0% | 0.0% |
| Dakwar 2020 | 0 | 17 | 0 | 17 | | | 0.0% | 0.0% |
| Daly 2018 PA | 0 | 34 | 0 | 34 | | | 0.0% | 0.0% |
| Daly 2018 PB | 0 | 20 | 0 | 20 | | | 0.0% | 0.0% |
| Domany 2019 | 0 | 22 | 0 | 22 | | | 0.0% | 0.0% |
| Domany 2020 | 0 | 9 | 0 | 9 | | | 0.0% | 0.0% |
| Duek 2023 | 0 | 14 | 0 | 14 | | | 0.0% | 0.0% |
| Dwyer 2021 | 0 | 11 | 0 | 11 | | | 0.0% | 0.0% |
| Fava 2020 | 0 | 80 | 0 | 80 | | | 0.0% | 0.0% |
| Feder 2014 | 0 | 22 | 0 | 22 | | | 0.0% | 0.0% |
| Feder 2021 | 0 | 15 | 0 | 15 | | | 0.0% | 0.0% |
| Fedgchin 2019 | 0 | 231 | 0 | 231 | | | 0.0% | 0.0% |
| Fineberg 2023 | 0 | 10 | 0 | 10 | | | 0.0% | 0.0% |
| Fu 2020 | 1 | 113 | 1 | 113 | 1.00 | [0.06; 16.09] | 13.4% | 13.4% |
| George 2017 | 0 | 12 | 0 | 12 | | | 0.0% | 0.0% |
| Glue 2024b | 0 | 17 | 0 | 17 | | | 0.0% | 0.0% |
| Grabski 2022 | 0 | 48 | 0 | 48 | | | 0.0% | 0.0% |
| Grunebaum 2017 | 0 | 9 | 0 | 9 | | | 0.0% | 0.0% |
| Grunebaum 2018 | 0 | 40 | 0 | 40 | | | 0.0% | 0.0% |
| Ionescu 2020 | 3 | 114 | 3 | 114 | 1.00 | [0.20; 5.04] | 39.5% | 39.5% |
| Jones 2024 | 0 | 17 | 0 | 17 | | | 0.0% | 0.0% |
| Lapidus 2014 | 0 | 10 | 0 | 10 | | | 0.0% | 0.0% |
| Leal 2023 | 0 | 5 | 0 | 5 | | | 0.0% | 0.0% |
| Lii 2023 | 0 | 20 | 0 | 20 | | | 0.0% | 0.0% |
| Lijffijt 2021 | 0 | 20 | 0 | 20 | | | 0.0% | 0.0% |
| Loo 2016 | 0 | 9 | 0 | 9 | | | 0.0% | 0.0% |
| Loo 2023 | 0 | 89 | 2 | 89 | 0.13 | [0.01; 2.16] | 13.4% | 13.4% |
| Milak 2020 | 0 | 38 | 0 | 38 | | | 0.0% | 0.0% |
| Murrough 2013 | 0 | 47 | 0 | 47 | | | 0.0% | 0.0% |
| Murrough 2015 II | 0 | 12 | 0 | 12 | | | 0.0% | 0.0% |
| Ochs-Ross 2020 | 0 | 72 | 0 | 72 | | | 0.0% | 0.0% |
| Ohtani 2024 | 0 | 17 | 0 | 17 | | | 0.0% | 0.0% |
| Pattanaseri 2024 | 0 | 11 | 0 | 11 | | | 0.0% | 0.0% |
| Phillips 2019 | 0 | 22 | 0 | 22 | | | 0.0% | 0.0% |
| Popova 2019 | 0 | 115 | 0 | 115 | | | 0.0% | 0.0% |
| Price 2022 | 0 | 53 | 0 | 53 | | | 0.0% | 0.0% |
| Ray-Griffith 2017 | 0 | 8 | 0 | 8 | | | 0.0% | 0.0% |
| Rodriguez 2013 | 0 | 8 | 0 | 8 | | | 0.0% | 0.0% |
| Schiweck 2021 | 0 | 21 | 0 | 21 | | | 0.0% | 0.0% |
| Shiroma 2020 | 0 | 29 | 0 | 29 | | | 0.0% | 0.0% |
| Singh 2016 2x doses | 1 | 18 | 0 | 18 | 7.39 | [0.15; 372.38] | 6.7% | 6.7% |
| Singh 2016 3x doses | 0 | 17 | 0 | 17 | | | 0.0% | 0.0% |
| Singh 2016 II | 0 | 20 | 0 | 20 | | | 0.0% | 0.0% |
| Smith-Apeldoorn 2024 | 0 | 57 | 0 | 57 | | | 0.0% | 0.0% |
| Sos 2013 | 0 | 11 | 0 | 11 | | | 0.0% | 0.0% |
| Su 2017 | 0 | 47 | 0 | 47 | | | 0.0% | 0.0% |
| Su 2023 | 0 | 42 | 0 | 42 | | | 0.0% | 0.0% |
| Sumner 2019 | 0 | 17 | 0 | 17 | | | 0.0% | 0.0% |
| Takahashi 2021 | 0 | 122 | 0 | 122 | | | 0.0% | 0.0% |
| Taylor 2018 | 0 | 9 | 0 | 9 | | | 0.0% | 0.0% |
| Tiger 2020 | 0 | 20 | 0 | 20 | | | 0.0% | 0.0% |
| Wang 2024 | 0 | 182 | 0 | 182 | | | 0.0% | 0.0% |
| Xiao 2023 | 0 | 15 | 0 | 15 | | | 0.0% | 0.0% |
| Yang 2023 | 0 | 200 | 0 | 200 | | | 0.0% | 0.0% |
| Zarate 2006 | 0 | 9 | 0 | 9 | | | 0.0% | 0.0% |
| Zarate 2012 | 0 | 21 | 0 | 21 | | | 0.0% | 0.0% |
| Zhou 2024 | 0 | 27 | 1 | 27 | 0.14 | [0.00; 6.82] | 6.7% | 6.7% |
| **Common effect model** | | **2742** | | **2742** | **0.87** | **[0.32; 2.42]** | **100.0%** | **.** |
| **Random effects model** | | | | | **0.87** | **[0.32; 2.42]** | **.** | **100.0%** |

Heterogeneity: $I^2 = 0.0\%$, $\tau^2 < 0.0001$, $p = 0.4347$

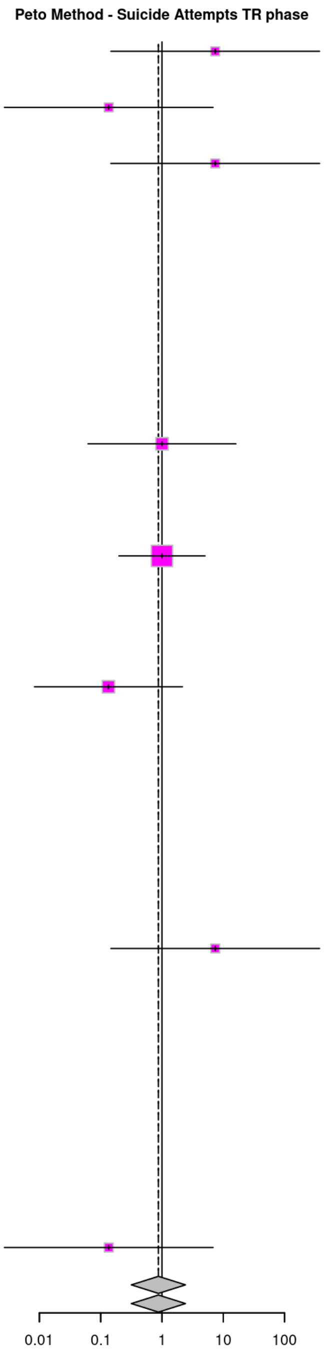

#### Maentel Mantel-Haenszel with CC

| Study | Experimental Events | Experimental Total | Control Events | Control Total | OR | 95%-CI | Weight (common) | Weight (random) |
|---|---|---|---|---|---|---|---|---|
| Abbar 2022 | 1 | 73 | 0 | 73 | 3.04 | [0.12; 75.89] | 1.3% | 2.1% |
| Abdallah 2022 | 0 | 108 | 0 | 108 | 1.00 | [0.02; 50.85] | 1.3% | 1.4% |
| Ahmed 2023 | 0 | 18 | 0 | 18 | 1.00 | [0.02; 53.12] | 1.3% | 1.3% |
| Anderson 2017 | 0 | 40 | 1 | 40 | 0.33 | [0.01; 8.22] | 3.8% | 2.0% |
| Burger 2016 | 0 | 3 | 0 | 3 | 1.00 | [0.02; 66.06] | 1.1% | 1.2% |
| Canuso 2018 | 0 | 35 | 0 | 35 | 1.00 | [0.02; 51.80] | 1.3% | 1.4% |
| Chen 2023 | 1 | 126 | 0 | 126 | 3.02 | [0.12; 74.94] | 1.3% | 2.1% |
| Colla 2024 | 0 | 17 | 0 | 17 | 1.00 | [0.02; 53.28] | 1.3% | 1.3% |
| Dakwar 2019 | 0 | 27 | 0 | 27 | 1.00 | [0.02; 52.22] | 1.3% | 1.4% |
| Dakwar 2020 | 0 | 17 | 0 | 17 | 1.00 | [0.02; 53.28] | 1.3% | 1.3% |
| Daly 2018 PA | 0 | 34 | 0 | 34 | 1.00 | [0.02; 51.84] | 1.3% | 1.4% |
| Daly 2018 PB | 0 | 20 | 0 | 20 | 1.00 | [0.02; 52.85] | 1.3% | 1.4% |
| Domany 2019 | 0 | 22 | 0 | 22 | 1.00 | [0.02; 52.63] | 1.3% | 1.4% |
| Domany 2020 | 0 | 9 | 0 | 9 | 1.00 | [0.02; 55.80] | 1.2% | 1.3% |
| Duek 2023 | 0 | 14 | 0 | 14 | 1.00 | [0.02; 53.89] | 1.3% | 1.3% |
| Dwyer 2021 | 0 | 11 | 0 | 11 | 1.00 | [0.02; 54.83] | 1.2% | 1.3% |
| Fava 2020 | 0 | 80 | 0 | 80 | 1.00 | [0.02; 51.01] | 1.3% | 1.4% |
| Feder 2014 | 0 | 22 | 0 | 22 | 1.00 | [0.02; 52.63] | 1.3% | 1.4% |
| Feder 2021 | 0 | 15 | 0 | 15 | 1.00 | [0.02; 53.66] | 1.3% | 1.3% |
| Fedgchin 2019 | 0 | 231 | 0 | 231 | 1.00 | [0.02; 50.61] | 1.3% | 1.4% |
| Fineberg 2023 | 0 | 10 | 0 | 10 | 1.00 | [0.02; 55.27] | 1.2% | 1.3% |
| Fu 2020 | 1 | 113 | 1 | 113 | 1.00 | [0.06; 16.19] | 2.6% | 2.7% |
| George 2017 | 0 | 12 | 0 | 12 | 1.00 | [0.02; 54.46] | 1.2% | 1.3% |
| Glue 2024b | 0 | 17 | 0 | 17 | 1.00 | [0.02; 53.28] | 1.3% | 1.3% |
| Grabski 2022 | 0 | 48 | 0 | 48 | 1.00 | [0.02; 51.42] | 1.3% | 1.4% |
| Grunebaum 2017 | 0 | 9 | 0 | 9 | 1.00 | [0.02; 55.80] | 1.2% | 1.3% |
| Grunebaum 2018 | 0 | 40 | 0 | 40 | 1.00 | [0.02; 51.63] | 1.3% | 1.4% |
| Ionescu 2020 | 3 | 114 | 3 | 114 | 1.00 | [0.20; 5.06] | 7.6% | 8.1% |
| Jones 2024 | 0 | 17 | 0 | 17 | 1.00 | [0.02; 53.28] | 1.3% | 1.3% |
| Lapidus 2014 | 0 | 10 | 0 | 10 | 1.00 | [0.02; 55.27] | 1.2% | 1.3% |
| Leal 2023 | 0 | 5 | 0 | 5 | 1.00 | [0.02; 59.99] | 1.2% | 1.3% |
| Lii 2023 | 0 | 20 | 0 | 20 | 1.00 | [0.02; 52.85] | 1.3% | 1.4% |
| Lijffijt 2021 | 0 | 20 | 0 | 20 | 1.00 | [0.02; 52.85] | 1.3% | 1.4% |
| Loo 2016 | 0 | 9 | 0 | 9 | 1.00 | [0.02; 55.80] | 1.2% | 1.3% |
| Loo 2023 | 0 | 89 | 2 | 89 | 0.20 | [0.01; 4.13] | 6.5% | 2.3% |
| Milak 2020 | 0 | 38 | 0 | 38 | 1.00 | [0.02; 51.69] | 1.3% | 1.4% |
| Murrough 2013 | 0 | 47 | 0 | 47 | 1.00 | [0.02; 51.44] | 1.3% | 1.4% |
| Murrough 2015 II | 0 | 12 | 0 | 12 | 1.00 | [0.02; 54.46] | 1.2% | 1.3% |
| Ochs-Ross 2020 | 0 | 72 | 0 | 72 | 1.00 | [0.02; 51.08] | 1.3% | 1.4% |
| Ohtani 2024 | 0 | 17 | 0 | 17 | 1.00 | [0.02; 53.28] | 1.3% | 1.3% |
| Pattanaseri 2024 | 0 | 11 | 0 | 11 | 1.00 | [0.02; 54.83] | 1.2% | 1.3% |
| Phillips 2019 | 0 | 22 | 0 | 22 | 1.00 | [0.02; 52.63] | 1.3% | 1.4% |
| Popova 2019 | 0 | 115 | 0 | 115 | 1.00 | [0.02; 50.83] | 1.3% | 1.4% |
| Price 2022 | 0 | 53 | 0 | 53 | 1.00 | [0.02; 51.33] | 1.3% | 1.4% |
| Ray-Griffith 2017 | 0 | 8 | 0 | 8 | 1.00 | [0.02; 56.46] | 1.2% | 1.3% |
| Rodriguez 2013 | 0 | 8 | 0 | 8 | 1.00 | [0.02; 56.46] | 1.2% | 1.3% |
| Schiweck 2021 | 0 | 21 | 0 | 21 | 1.00 | [0.02; 52.73] | 1.3% | 1.4% |
| Shiroma 2020 | 0 | 29 | 0 | 29 | 1.00 | [0.02; 52.09] | 1.3% | 1.4% |
| Singh 2016 2x doses | 1 | 18 | 0 | 18 | 3.17 | [0.12; 83.17] | 1.2% | 2.0% |
| Singh 2016 3x doses | 0 | 17 | 0 | 17 | 1.00 | [0.02; 53.28] | 1.3% | 1.3% |
| Singh 2016 II | 0 | 20 | 0 | 20 | 1.00 | [0.02; 52.85] | 1.3% | 1.4% |
| Smith-Apeldoorn 2024 | 0 | 57 | 0 | 57 | 1.00 | [0.02; 51.26] | 1.3% | 1.4% |
| Sos 2013 | 0 | 11 | 0 | 11 | 1.00 | [0.02; 54.83] | 1.2% | 1.3% |
| Su 2017 | 0 | 47 | 0 | 47 | 1.00 | [0.02; 51.44] | 1.3% | 1.4% |
| Su 2023 | 0 | 42 | 0 | 42 | 1.00 | [0.02; 51.57] | 1.3% | 1.4% |
| Sumner 2019 | 0 | 17 | 0 | 17 | 1.00 | [0.02; 53.28] | 1.3% | 1.3% |
| Takahashi 2021 | 0 | 122 | 0 | 122 | 1.00 | [0.02; 50.80] | 1.3% | 1.4% |
| Taylor 2018 | 0 | 9 | 0 | 9 | 1.00 | [0.02; 55.80] | 1.2% | 1.3% |
| Tiger 2020 | 0 | 20 | 0 | 20 | 1.00 | [0.02; 52.85] | 1.3% | 1.4% |
| Wang 2024 | 0 | 182 | 0 | 182 | 1.00 | [0.02; 50.67] | 1.3% | 1.4% |
| Xiao 2023 | 0 | 15 | 0 | 15 | 1.00 | [0.02; 53.66] | 1.3% | 1.3% |
| Yang 2023 | 0 | 200 | 0 | 200 | 1.00 | [0.02; 50.64] | 1.3% | 1.4% |
| Zarate 2006 | 0 | 9 | 0 | 9 | 1.00 | [0.02; 55.80] | 1.2% | 1.3% |
| Zarate 2012 | 0 | 21 | 0 | 21 | 1.00 | [0.02; 52.73] | 1.3% | 1.4% |
| Zhou 2024 | 0 | 27 | 1 | 27 | 0.32 | [0.01; 8.24] | 3.8% | 2.0% |
| **Common effect model** | | **2742** | | **2742** | **0.97** | **[0.62; 1.53]** | **100.0%** | **.** |
| **Random effects model** | | | | | **0.99** | **[0.62; 1.56]** | **.** | **100.0%** |

Heterogeneity: $I^2 = 0.0\%$, $\tau^2 = 0$, $p = 1.0000$

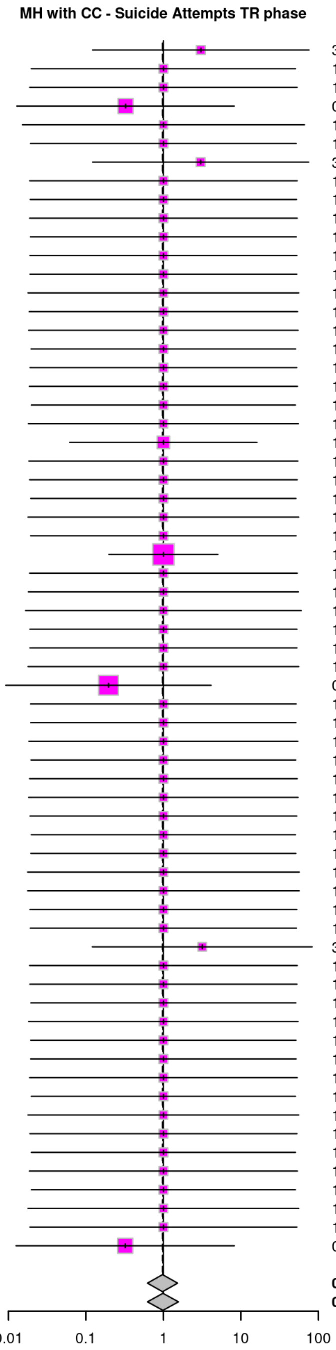

#### Maentel Mantel-Haenszel with Treatment-Arm CC

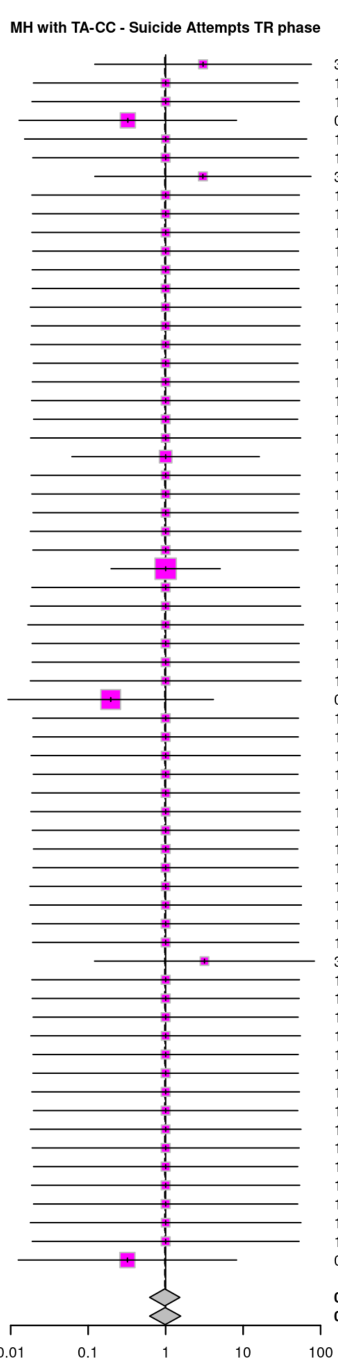

| Study | Experimental Events | Experimental Total | Control Events | Control Total | OR | 95%-CI | Weight (common) | Weight (random) |
|---|---|---|---|---|---|---|---|---|
| Abbar 2022 | 1 | 73 | 0 | 73 | 3.04 | [0.12; 75.89] | 1.3% | 2.1% |
| Abdallah 2022 | 0 | 108 | 0 | 108 | 1.00 | [0.02; 50.85] | 1.3% | 1.4% |
| Ahmed 2023 | 0 | 18 | 0 | 18 | 1.00 | [0.02; 53.12] | 1.3% | 1.3% |
| Anderson 2017 | 0 | 40 | 1 | 40 | 0.33 | [0.01; 8.22] | 3.8% | 2.0% |
| Burger 2016 | 0 | 3 | 0 | 3 | 1.00 | [0.02; 66.06] | 1.1% | 1.2% |
| Canuso 2018 | 0 | 35 | 0 | 35 | 1.00 | [0.02; 51.80] | 1.3% | 1.4% |
| Chen 2023 | 1 | 126 | 0 | 126 | 3.02 | [0.12; 74.94] | 1.3% | 2.1% |
| Colla 2024 | 0 | 17 | 0 | 17 | 1.00 | [0.02; 53.28] | 1.3% | 1.3% |
| Dakwar 2019 | 0 | 27 | 0 | 27 | 1.00 | [0.02; 52.22] | 1.3% | 1.4% |
| Dakwar 2020 | 0 | 17 | 0 | 17 | 1.00 | [0.02; 53.28] | 1.3% | 1.3% |
| Daly 2018 PA | 0 | 34 | 0 | 34 | 1.00 | [0.02; 51.84] | 1.3% | 1.4% |
| Daly 2018 PB | 0 | 20 | 0 | 20 | 1.00 | [0.02; 52.85] | 1.3% | 1.4% |
| Domany 2019 | 0 | 22 | 0 | 22 | 1.00 | [0.02; 52.63] | 1.3% | 1.4% |
| Domany 2020 | 0 | 9 | 0 | 9 | 1.00 | [0.02; 55.80] | 1.2% | 1.3% |
| Duek 2023 | 0 | 14 | 0 | 14 | 1.00 | [0.02; 53.89] | 1.3% | 1.3% |
| Dwyer 2021 | 0 | 11 | 0 | 11 | 1.00 | [0.02; 54.83] | 1.2% | 1.3% |
| Fava 2020 | 0 | 80 | 0 | 80 | 1.00 | [0.02; 51.01] | 1.3% | 1.4% |
| Feder 2014 | 0 | 22 | 0 | 22 | 1.00 | [0.02; 52.63] | 1.3% | 1.4% |
| Feder 2021 | 0 | 15 | 0 | 15 | 1.00 | [0.02; 53.66] | 1.3% | 1.3% |
| Fedgchin 2019 | 0 | 231 | 0 | 231 | 1.00 | [0.02; 50.61] | 1.3% | 1.4% |
| Fineberg 2023 | 0 | 10 | 0 | 10 | 1.00 | [0.02; 55.27] | 1.2% | 1.3% |
| Fu 2020 | 1 | 113 | 1 | 113 | 1.00 | [0.06; 16.19] | 2.6% | 2.7% |
| George 2017 | 0 | 12 | 0 | 12 | 1.00 | [0.02; 54.46] | 1.2% | 1.3% |
| Glue 2024b | 0 | 17 | 0 | 17 | 1.00 | [0.02; 53.28] | 1.3% | 1.3% |
| Grabski 2022 | 0 | 48 | 0 | 48 | 1.00 | [0.02; 51.42] | 1.3% | 1.4% |
| Grunebaum 2017 | 0 | 9 | 0 | 9 | 1.00 | [0.02; 55.80] | 1.2% | 1.3% |
| Grunebaum 2018 | 0 | 40 | 0 | 40 | 1.00 | [0.02; 51.63] | 1.3% | 1.4% |
| Ionescu 2020 | 3 | 114 | 3 | 114 | 1.00 | [0.20; 5.06] | 7.6% | 8.1% |
| Jones 2024 | 0 | 17 | 0 | 17 | 1.00 | [0.02; 53.28] | 1.3% | 1.3% |
| Lapidus 2014 | 0 | 10 | 0 | 10 | 1.00 | [0.02; 55.27] | 1.2% | 1.3% |
| Leal 2023 | 0 | 5 | 0 | 5 | 1.00 | [0.02; 59.99] | 1.2% | 1.3% |
| Lii 2023 | 0 | 20 | 0 | 20 | 1.00 | [0.02; 52.85] | 1.3% | 1.4% |
| Lijffijt 2021 | 0 | 20 | 0 | 20 | 1.00 | [0.02; 52.85] | 1.3% | 1.4% |
| Loo 2016 | 0 | 9 | 0 | 9 | 1.00 | [0.02; 55.80] | 1.2% | 1.3% |
| Loo 2023 | 0 | 89 | 2 | 89 | 0.20 | [0.01; 4.13] | 6.5% | 2.3% |
| Milak 2020 | 0 | 38 | 0 | 38 | 1.00 | [0.02; 51.69] | 1.3% | 1.4% |
| Murrough 2013 | 0 | 47 | 0 | 47 | 1.00 | [0.02; 51.44] | 1.3% | 1.4% |
| Murrough 2015 II | 0 | 12 | 0 | 12 | 1.00 | [0.02; 54.46] | 1.2% | 1.3% |
| Ochs-Ross 2020 | 0 | 72 | 0 | 72 | 1.00 | [0.02; 51.08] | 1.3% | 1.4% |
| Ohtani 2024 | 0 | 17 | 0 | 17 | 1.00 | [0.02; 53.28] | 1.3% | 1.3% |
| Pattanaseri 2024 | 0 | 11 | 0 | 11 | 1.00 | [0.02; 54.83] | 1.2% | 1.3% |
| Phillips 2019 | 0 | 22 | 0 | 22 | 1.00 | [0.02; 52.63] | 1.3% | 1.4% |
| Popova 2019 | 0 | 115 | 0 | 115 | 1.00 | [0.02; 50.83] | 1.3% | 1.4% |
| Price 2022 | 0 | 53 | 0 | 53 | 1.00 | [0.02; 51.33] | 1.3% | 1.4% |
| Ray-Griffith 2017 | 0 | 8 | 0 | 8 | 1.00 | [0.02; 56.46] | 1.2% | 1.3% |
| Rodriguez 2013 | 0 | 8 | 0 | 8 | 1.00 | [0.02; 56.46] | 1.2% | 1.3% |
| Schiweck 2021 | 0 | 21 | 0 | 21 | 1.00 | [0.02; 52.73] | 1.3% | 1.4% |
| Shiroma 2020 | 0 | 29 | 0 | 29 | 1.00 | [0.02; 52.09] | 1.3% | 1.4% |
| Singh 2016 2x doses | 1 | 18 | 0 | 18 | 3.17 | [0.12; 83.17] | 1.2% | 2.0% |
| Singh 2016 3x doses | 0 | 17 | 0 | 17 | 1.00 | [0.02; 53.28] | 1.3% | 1.3% |
| Singh 2016 II | 0 | 20 | 0 | 20 | 1.00 | [0.02; 52.85] | 1.3% | 1.4% |
| Smith-Apeldoorn 2024 | 0 | 57 | 0 | 57 | 1.00 | [0.02; 51.26] | 1.3% | 1.4% |
| Sos 2013 | 0 | 11 | 0 | 11 | 1.00 | [0.02; 54.83] | 1.2% | 1.3% |
| Su 2017 | 0 | 47 | 0 | 47 | 1.00 | [0.02; 51.44] | 1.3% | 1.4% |
| Su 2023 | 0 | 42 | 0 | 42 | 1.00 | [0.02; 51.57] | 1.3% | 1.4% |
| Sumner 2019 | 0 | 17 | 0 | 17 | 1.00 | [0.02; 53.28] | 1.3% | 1.3% |
| Takahashi 2021 | 0 | 122 | 0 | 122 | 1.00 | [0.02; 50.80] | 1.3% | 1.4% |
| Taylor 2018 | 0 | 9 | 0 | 9 | 1.00 | [0.02; 55.80] | 1.2% | 1.3% |
| Tiger 2020 | 0 | 20 | 0 | 20 | 1.00 | [0.02; 52.85] | 1.3% | 1.4% |
| Wang 2024 | 0 | 182 | 0 | 182 | 1.00 | [0.02; 50.67] | 1.3% | 1.4% |
| Xiao 2023 | 0 | 15 | 0 | 15 | 1.00 | [0.02; 53.66] | 1.3% | 1.3% |
| Yang 2023 | 0 | 200 | 0 | 200 | 1.00 | [0.02; 50.64] | 1.3% | 1.4% |
| Zarate 2006 | 0 | 9 | 0 | 9 | 1.00 | [0.02; 55.80] | 1.2% | 1.3% |
| Zarate 2012 | 0 | 21 | 0 | 21 | 1.00 | [0.02; 52.73] | 1.3% | 1.4% |
| Zhou 2024 | 0 | 27 | 1 | 27 | 0.32 | [0.01; 8.24] | 3.8% | 2.0% |
| **Common effect model** | | **2742** | | **2742** | **0.97** | **[0.62; 1.53]** | **100.0%** | **.** |
| **Random effects model** | | | | | **0.99** | **[0.62; 1.56]** | **.** | **100.0%** |

Heterogeneity: $I^2 = 0.0\%$, $\tau^2 = 0$, $p = 1.0000$

#### Maentel Mantel-Haenszel Risk-Difference

| Study | Experimental Events | Experimental Total | Control Events | Control Total | RD | 95%-CI | Weight (common) | Weight (random) |
|---|---|---|---|---|---|---|---|---|
| Abbar 2022 | 1 | 73 | 0 | 73 | 0.01 | [-0.02; 0.05] | 2.6% | 1.2% |
| Abdallah 2022 | 0 | 108 | 0 | 108 | 0.00 | [-0.02; 0.02] | 3.9% | 5.0% |
| Ahmed 2023 | 0 | 18 | 0 | 18 | 0.00 | [-0.10; 0.10] | 0.7% | 0.2% |
| Anderson 2017 | 0 | 40 | 1 | 40 | -0.03 | [-0.09; 0.04] | 1.5% | 0.4% |
| Burger 2016 | 0 | 3 | 0 | 3 | 0.00 | [-0.46; 0.46] | 0.1% | 0.0% |
| Canuso 2018 | 0 | 35 | 0 | 35 | 0.00 | [-0.05; 0.05] | 1.3% | 0.6% |
| Chen 2023 | 1 | 126 | 0 | 126 | 0.01 | [-0.01; 0.03] | 4.5% | 3.4% |
| Colla 2024 | 0 | 17 | 0 | 17 | 0.00 | [-0.11; 0.11] | 0.6% | 0.1% |
| Dakwar 2019 | 0 | 27 | 0 | 27 | 0.00 | [-0.07; 0.07] | 1.0% | 0.3% |
| Dakwar 2020 | 0 | 17 | 0 | 17 | 0.00 | [-0.11; 0.11] | 0.6% | 0.1% |
| Daly 2018 PA | 0 | 34 | 0 | 34 | 0.00 | [-0.06; 0.06] | 1.2% | 0.5% |
| Daly 2018 PB | 0 | 20 | 0 | 20 | 0.00 | [-0.09; 0.09] | 0.7% | 0.2% |
| Domany 2019 | 0 | 22 | 0 | 22 | 0.00 | [-0.08; 0.08] | 0.8% | 0.2% |
| Domany 2020 | 0 | 9 | 0 | 9 | 0.00 | [-0.19; 0.19] | 0.4% | 0.0% |
| Duek 2023 | 0 | 14 | 0 | 14 | 0.00 | [-0.13; 0.13] | 0.5% | 0.1% |
| Dwyer 2021 | 0 | 11 | 0 | 11 | 0.00 | [-0.16; 0.16] | 0.4% | 0.1% |
| Fava 2020 | 0 | 80 | 0 | 80 | 0.00 | [-0.02; 0.02] | 2.9% | 2.8% |
| Feder 2014 | 0 | 22 | 0 | 22 | 0.00 | [-0.08; 0.08] | 0.8% | 0.2% |
| Feder 2021 | 0 | 15 | 0 | 15 | 0.00 | [-0.12; 0.12] | 0.6% | 0.1% |
| Fedgchin 2019 | 0 | 231 | 0 | 231 | 0.00 | [-0.01; 0.01] | 8.3% | 22.7% |
| Fineberg 2023 | 0 | 10 | 0 | 10 | 0.00 | [-0.17; 0.17] | 0.4% | 0.1% |
| Fu 2020 | 1 | 113 | 1 | 113 | 0.00 | [-0.02; 0.02] | 4.0% | 2.7% |
| George 2017 | 0 | 12 | 0 | 12 | 0.00 | [-0.15; 0.15] | 0.5% | 0.1% |
| Glue 2024b | 0 | 17 | 0 | 17 | 0.00 | [-0.11; 0.11] | 0.6% | 0.1% |
| Grabski 2022 | 0 | 48 | 0 | 48 | 0.00 | [-0.04; 0.04] | 1.7% | 1.0% |
| Grunebaum 2017 | 0 | 9 | 0 | 9 | 0.00 | [-0.19; 0.19] | 0.4% | 0.0% |
| Grunebaum 2018 | 0 | 40 | 0 | 40 | 0.00 | [-0.05; 0.05] | 1.5% | 0.7% |
| Ionescu 2020 | 3 | 114 | 3 | 114 | 0.00 | [-0.04; 0.04] | 4.1% | 0.9% |
| Jones 2024 | 0 | 17 | 0 | 17 | 0.00 | [-0.11; 0.11] | 0.6% | 0.1% |
| Lapidus 2014 | 0 | 10 | 0 | 10 | 0.00 | [-0.17; 0.17] | 0.4% | 0.1% |
| Leal 2023 | 0 | 5 | 0 | 5 | 0.00 | [-0.31; 0.31] | 0.2% | 0.0% |
| Lii 2023 | 0 | 20 | 0 | 20 | 0.00 | [-0.09; 0.09] | 0.7% | 0.2% |
| Lijffijt 2021 | 0 | 20 | 0 | 20 | 0.00 | [-0.09; 0.09] | 0.7% | 0.2% |
| Loo 2016 | 0 | 9 | 0 | 9 | 0.00 | [-0.19; 0.19] | 0.4% | 0.0% |
| Loo 2023 | 0 | 89 | 2 | 89 | -0.02 | [-0.06; 0.01] | 3.2% | 1.2% |
| Milak 2020 | 0 | 38 | 0 | 38 | 0.00 | [-0.05; 0.05] | 1.4% | 0.6% |
| Murrough 2013 | 0 | 47 | 0 | 47 | 0.00 | [-0.04; 0.04] | 1.7% | 1.0% |
| Murrough 2015 II | 0 | 12 | 0 | 12 | 0.00 | [-0.15; 0.15] | 0.5% | 0.1% |
| Ochs-Ross 2020 | 0 | 72 | 0 | 72 | 0.00 | [-0.03; 0.03] | 2.6% | 2.3% |
| Ohtani 2024 | 0 | 17 | 0 | 17 | 0.00 | [-0.11; 0.11] | 0.6% | 0.1% |
| Pattanaseri 2024 | 0 | 11 | 0 | 11 | 0.00 | [-0.16; 0.16] | 0.4% | 0.1% |
| Phillips 2019 | 0 | 22 | 0 | 22 | 0.00 | [-0.08; 0.08] | 0.8% | 0.2% |
| Popova 2019 | 0 | 115 | 0 | 115 | 0.00 | [-0.02; 0.02] | 4.1% | 5.7% |
| Price 2022 | 0 | 53 | 0 | 53 | 0.00 | [-0.04; 0.04] | 1.9% | 1.2% |
| Ray-Griffith 2017 | 0 | 8 | 0 | 8 | 0.00 | [-0.21; 0.21] | 0.3% | 0.0% |
| Rodriguez 2013 | 0 | 8 | 0 | 8 | 0.00 | [-0.21; 0.21] | 0.3% | 0.0% |
| Schiweck 2021 | 0 | 21 | 0 | 21 | 0.00 | [-0.09; 0.09] | 0.8% | 0.2% |
| Shiroma 2020 | 0 | 29 | 0 | 29 | 0.00 | [-0.06; 0.06] | 1.1% | 0.4% |
| Singh 2016 2x doses | 1 | 18 | 0 | 18 | 0.06 | [-0.09; 0.20] | 0.7% | 0.1% |
| Singh 2016 3x doses | 0 | 17 | 0 | 17 | 0.00 | [-0.11; 0.11] | 0.6% | 0.1% |
| Singh 2016 II | 0 | 20 | 0 | 20 | 0.00 | [-0.09; 0.09] | 0.7% | 0.2% |
| Smith-Apeldoorn 2024 | 0 | 57 | 0 | 57 | 0.00 | [-0.03; 0.03] | 2.1% | 1.4% |
| Sos 2013 | 0 | 11 | 0 | 11 | 0.00 | [-0.16; 0.16] | 0.4% | 0.1% |
| Su 2017 | 0 | 47 | 0 | 47 | 0.00 | [-0.04; 0.04] | 1.7% | 1.0% |
| Su 2023 | 0 | 42 | 0 | 42 | 0.00 | [-0.05; 0.05] | 1.5% | 0.8% |
| Sumner 2019 | 0 | 17 | 0 | 17 | 0.00 | [-0.11; 0.11] | 0.6% | 0.1% |
| Takahashi 2021 | 0 | 122 | 0 | 122 | 0.00 | [-0.02; 0.02] | 4.4% | 6.4% |
| Taylor 2018 | 0 | 9 | 0 | 9 | 0.00 | [-0.19; 0.19] | 0.4% | 0.0% |
| Tiger 2020 | 0 | 20 | 0 | 20 | 0.00 | [-0.09; 0.09] | 0.7% | 0.2% |
| Wang 2024 | 0 | 182 | 0 | 182 | 0.00 | [-0.01; 0.01] | 6.5% | 14.1% |
| Xiao 2023 | 0 | 15 | 0 | 15 | 0.00 | [-0.12; 0.12] | 0.6% | 0.1% |
| Yang 2023 | 0 | 200 | 0 | 200 | 0.00 | [-0.01; 0.01] | 7.2% | 17.1% |
| Zarate 2006 | 0 | 9 | 0 | 9 | 0.00 | [-0.19; 0.19] | 0.4% | 0.0% |
| Zarate 2012 | 0 | 21 | 0 | 21 | 0.00 | [-0.09; 0.09] | 0.8% | 0.2% |
| Zhou 2024 | 0 | 27 | 1 | 27 | -0.04 | [-0.13; 0.06] | 1.0% | 0.2% |
| **Common effect model** | | **2742** | | **2742** | **-0.00** | **[-0.01; 0.01]** | **100.0%** | **.** |
| **Random effects model** | | | | | **0.00** | **[-0.00; 0.00]** | **.** | **100.0%** |

Heterogeneity: $I^2 = 0.0\%$, $\tau^2 = 0$, $p = 1.0000$

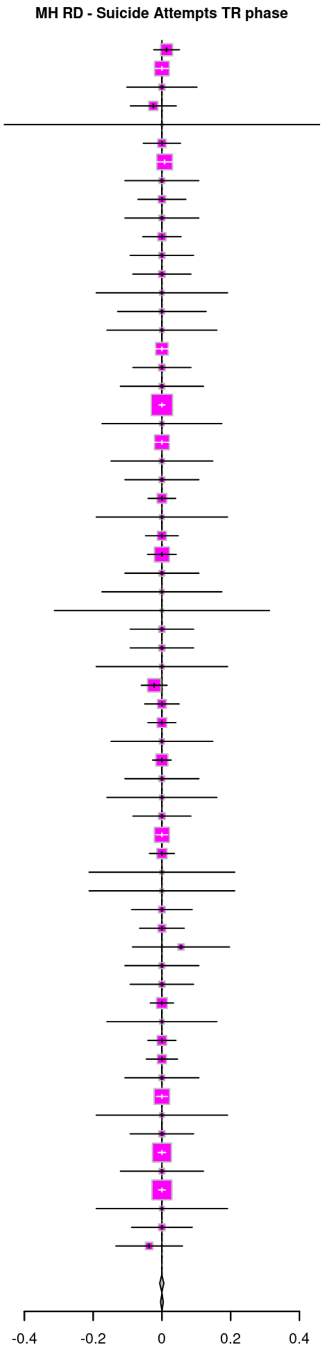

#### Arcsine Risk-Difference

##### Arcsine RD, DB, Suicide Attempts

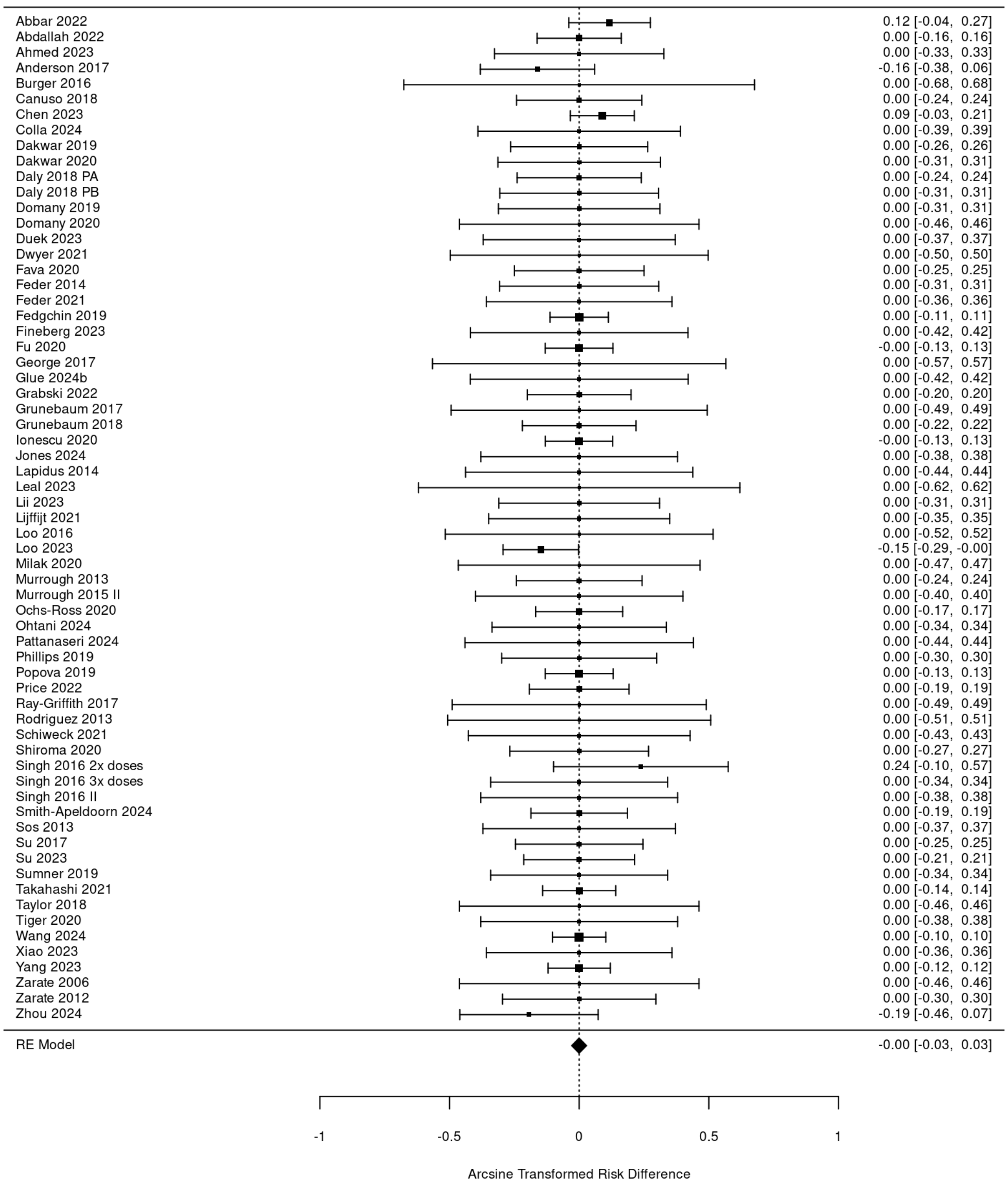

**Bayesian Meta-Analysis, wider prior**

**Bayesian MA, Delta = 250 - Suicide Attempts TR**

| Study | Estimate | 95% CI |
|---|---|---|
| Abbar 2022 | 1.24 | [-1.98, 4.46] |
| Abdallah 2022 | -0.67 | [-4.60, 3.26] |
| Ahmed 2023 | 0.00 | [-3.97, 3.97] |
| Anderson 2017 | -1.15 | [-4.38, 2.08] |
| Burger 2016 | 0.76 | [-3.36, 4.88] |
| Canuso 2018 | -0.12 | [-4.07, 3.83] |
| Chen 2023 | 1.11 | [-2.10, 4.32] |
| Colla 2024 | -0.51 | [-4.50, 3.48] |
| Dakwar 2019 | 0.04 | [-3.92, 3.99] |
| Dakwar 2020 | 0.29 | [-3.67, 4.26] |
| Daly 2018 PA | -0.03 | [-3.98, 3.92] |
| Daly 2018 PB | 0.05 | [-3.92, 4.01] |
| Domany 2019 | -0.20 | [-4.16, 3.77] |
| Domany 2020 | 0.00 | [-4.02, 4.02] |
| Duek 2023 | 0.00 | [-3.99, 3.99] |
| Dwyer 2021 | -0.57 | [-4.61, 3.47] |
| Fava 2020 | -1.42 | [-5.37, 2.53] |
| Feder 2014 | -0.14 | [-4.11, 3.82] |
| Feder 2021 | 0.00 | [-3.98, 3.98] |
| Fedgchin 2019 | -0.71 | [-4.64, 3.21] |
| Fineberg 2023 | 0.17 | [-3.83, 4.18] |
| Fu 2020 | -0.01 | [-2.79, 2.78] |
| George 2017 | -1.02 | [-5.09, 3.04] |
| Glue 2024b | -0.72 | [-4.73, 3.28] |
| Grabski 2022 | 0.00 | [-3.94, 3.94] |
| Grunebaum 2017 | -0.24 | [-4.27, 3.80] |
| Grunebaum 2018 | 0.00 | [-3.94, 3.94] |
| Ionescu 2020 | -0.01 | [-1.63, 1.61] |
| Jones 2024 | -0.42 | [-4.41, 3.57] |
| Lapidus 2014 | 0.00 | [-4.01, 4.01] |
| Leal 2023 | 0.00 | [-4.09, 4.09] |
| Lii 2023 | 0.00 | [-3.97, 3.97] |
| Lijffijt 2021 | -0.42 | [-4.40, 3.56] |
| Loo 2016 | -0.38 | [-4.42, 3.67] |
| Loo 2023 | -1.60 | [-4.65, 1.45] |
| Milak 2020 | -1.95 | [-5.97, 2.07] |
| Murrough 2013 | -0.62 | [-4.57, 3.33] |
| Murrough 2015 II | 0.00 | [-4.00, 4.00] |
| Ochs-Ross 2020 | -0.10 | [-4.04, 3.83] |
| Ohtani 2024 | 0.00 | [-3.98, 3.98] |
| Pattanaseri 2024 | -0.19 | [-4.20, 3.82] |
| Phillips 2019 | -0.05 | [-4.01, 3.92] |
| Popova 2019 | -0.05 | [-3.98, 3.88] |
| Price 2022 | -0.04 | [-3.98, 3.90] |
| Ray-Griffith 2017 | 0.00 | [-4.03, 4.03] |
| Rodriguez 2013 | -0.13 | [-4.17, 3.92] |
| Schiweck 2021 | -1.05 | [-5.06, 2.95] |
| Shiroma 2020 | -0.15 | [-4.10, 3.81] |
| Singh 2016 2x doses | 1.04 | [-2.23, 4.31] |
| Singh 2016 3x doses | -0.06 | [-4.04, 3.92] |
| Singh 2016 II | -0.67 | [-4.66, 3.32] |
| Smith-Apeldoorn 2024 | -0.05 | [-3.99, 3.88] |
| Sos 2013 | 0.53 | [-3.46, 4.52] |
| Su 2017 | -0.66 | [-4.61, 3.29] |
| Su 2023 | 0.00 | [-3.94, 3.94] |
| Sumner 2019 | -0.06 | [-4.04, 3.92] |
| Takahashi 2021 | -0.42 | [-4.35, 3.51] |
| Taylor 2018 | 0.00 | [-4.02, 4.02] |
| Tiger 2020 | -0.67 | [-4.66, 3.32] |
| Wang 2024 | 0.00 | [-3.93, 3.93] |
| Xiao 2023 | 0.00 | [-3.98, 3.98] |
| Yang 2023 | -0.69 | [-4.62, 3.24] |
| Zarate 2006 | 0.00 | [-4.02, 4.02] |
| Zarate 2012 | 0.09 | [-3.87, 4.05] |
| Zhou 2024 | -1.14 | [-4.38, 2.11] |
| **Summary** | **-0.17** | **[-1.08, 0.75]** |
| **Prediction** | **-0.16** | **[-1.41, 1.07]** |

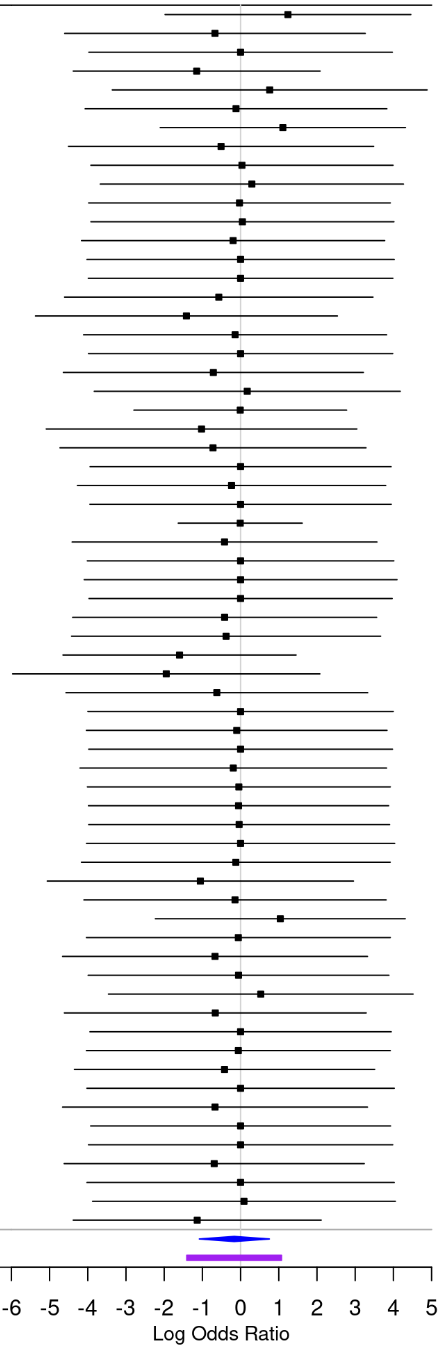

*Heterogeneity (tau): 0.27 [0.00, 0.81]*

**Bayesian Meta-Analysis, smaller prior**

**Bayesian MA, Delta = 15 - Suicide Attempts TR**

| Study | Estimate | 95% CI |
|---|---|---|
| Abbar 2022 | 1.24 | [-1.98, 4.46] |
| Abdallah 2022 | -0.67 | [-4.60, 3.26] |
| Ahmed 2023 | 0.00 | [-3.97, 3.97] |
| Anderson 2017 | -1.15 | [-4.38, 2.08] |
| Burger 2016 | 0.76 | [-3.36, 4.88] |
| Canuso 2018 | -0.12 | [-4.07, 3.83] |
| Chen 2023 | 1.11 | [-2.10, 4.32] |
| Colla 2024 | -0.51 | [-4.50, 3.48] |
| Dakwar 2019 | 0.04 | [-3.92, 3.99] |
| Dakwar 2020 | 0.29 | [-3.67, 4.26] |
| Daly 2018 PA | -0.03 | [-3.98, 3.92] |
| Daly 2018 PB | 0.05 | [-3.92, 4.01] |
| Domany 2019 | -0.20 | [-4.16, 3.77] |
| Domany 2020 | 0.00 | [-4.02, 4.02] |
| Duek 2023 | 0.00 | [-3.99, 3.99] |
| Dwyer 2021 | -0.57 | [-4.61, 3.47] |
| Fava 2020 | -1.42 | [-5.37, 2.53] |
| Feder 2014 | -0.14 | [-4.11, 3.82] |
| Feder 2021 | 0.00 | [-3.98, 3.98] |
| Fedgchin 2019 | -0.71 | [-4.64, 3.21] |
| Fineberg 2023 | 0.17 | [-3.83, 4.18] |
| Fu 2020 | -0.01 | [-2.79, 2.78] |
| George 2017 | -1.02 | [-5.09, 3.04] |
| Glue 2024b | -0.72 | [-4.73, 3.28] |
| Grabski 2022 | 0.00 | [-3.94, 3.94] |
| Grunebaum 2017 | -0.24 | [-4.27, 3.80] |
| Grunebaum 2018 | 0.00 | [-3.94, 3.94] |
| Ionescu 2020 | -0.01 | [-1.63, 1.61] |
| Jones 2024 | -0.42 | [-4.41, 3.57] |
| Lapidus 2014 | 0.00 | [-4.01, 4.01] |
| Leal 2023 | 0.00 | [-4.09, 4.09] |
| Lii 2023 | 0.00 | [-3.97, 3.97] |
| Lijffijt 2021 | -0.42 | [-4.40, 3.56] |
| Loo 2016 | -0.38 | [-4.42, 3.67] |
| Loo 2023 | -1.60 | [-4.65, 1.45] |
| Milak 2020 | -1.95 | [-5.97, 2.07] |
| Murrough 2013 | -0.62 | [-4.57, 3.33] |
| Murrough 2015 II | 0.00 | [-4.00, 4.00] |
| Ochs-Ross 2020 | -0.10 | [-4.04, 3.83] |
| Ohtani 2024 | 0.00 | [-3.98, 3.98] |
| Pattanaseri 2024 | -0.19 | [-4.20, 3.82] |
| Phillips 2019 | -0.05 | [-4.01, 3.92] |
| Popova 2019 | -0.05 | [-3.98, 3.88] |
| Price 2022 | -0.04 | [-3.98, 3.90] |
| Ray-Griffith 2017 | 0.00 | [-4.03, 4.03] |
| Rodriguez 2013 | -0.13 | [-4.17, 3.92] |
| Schiweck 2021 | -1.05 | [-5.06, 2.95] |
| Shiroma 2020 | -0.15 | [-4.10, 3.81] |
| Singh 2016 2x doses | 1.04 | [-2.23, 4.31] |
| Singh 2016 3x doses | -0.06 | [-4.04, 3.92] |
| Singh 2016 II | -0.67 | [-4.66, 3.32] |
| Smith-Apeldoorn 2024 | -0.05 | [-3.99, 3.88] |
| Sos 2013 | 0.53 | [-3.46, 4.52] |
| Su 2017 | -0.66 | [-4.61, 3.29] |
| Su 2023 | 0.00 | [-3.94, 3.94] |
| Sumner 2019 | -0.06 | [-4.04, 3.92] |
| Takahashi 2021 | -0.42 | [-4.35, 3.51] |
| Taylor 2018 | 0.00 | [-4.02, 4.02] |
| Tiger 2020 | -0.67 | [-4.66, 3.32] |
| Wang 2024 | 0.00 | [-3.93, 3.93] |
| Xiao 2023 | 0.00 | [-3.98, 3.98] |
| Yang 2023 | -0.69 | [-4.62, 3.24] |
| Zarate 2006 | 0.00 | [-4.02, 4.02] |
| Zarate 2012 | 0.09 | [-3.87, 4.05] |
| Zhou 2024 | -1.14 | [-4.38, 2.11] |
| **Summary** | **-0.15** | **[-1.02, 0.72]** |
| **Prediction** | **-0.15** | **[-1.36, 1.05]** |

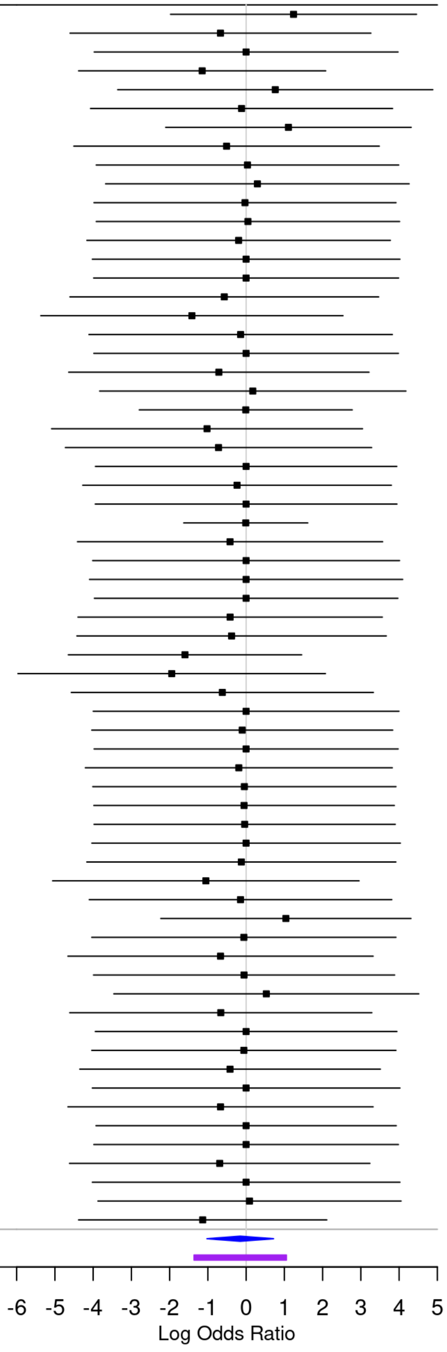

*Heterogeneity (tau): 0.28 [0.00, 0.81]*

**Exact Method**

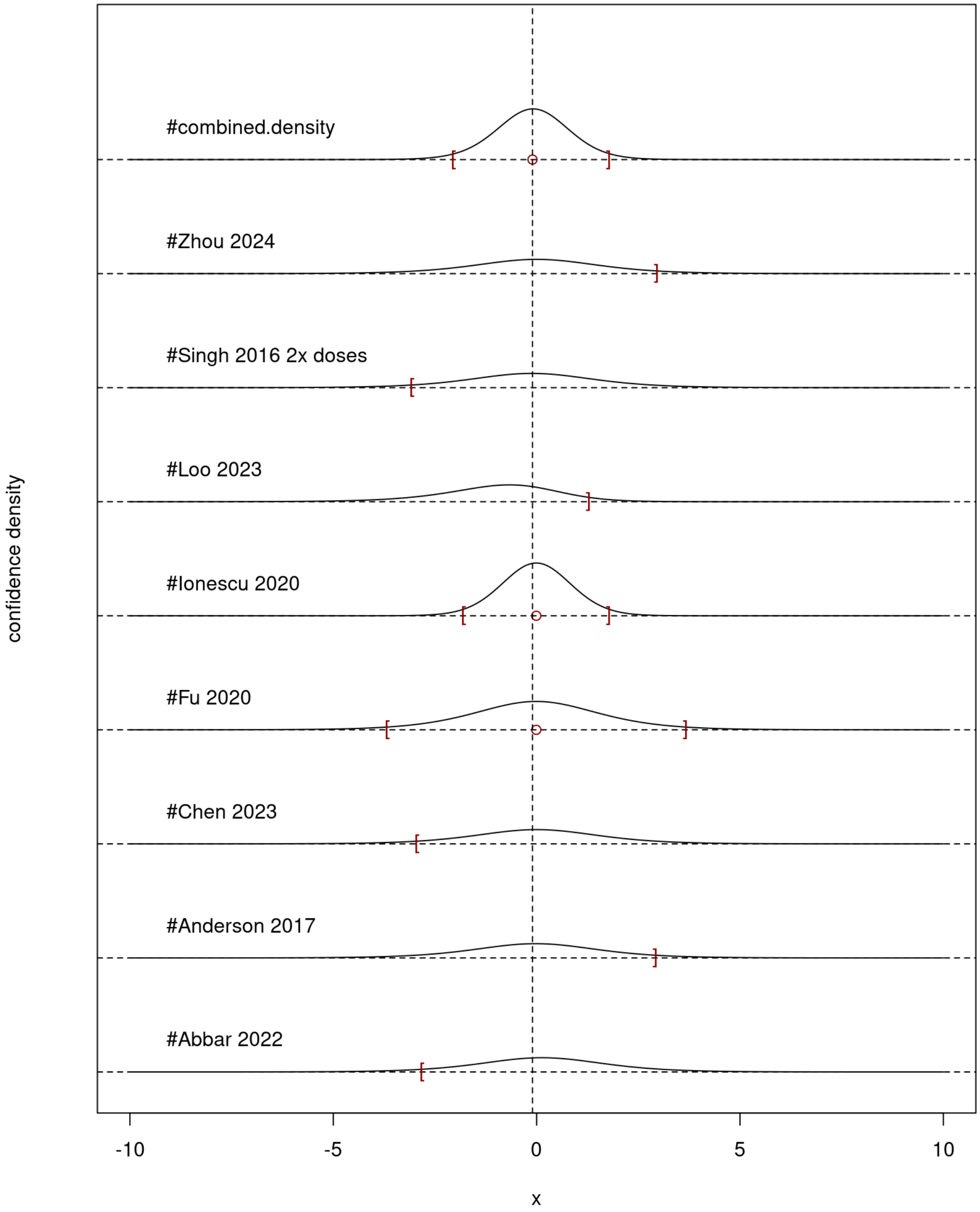

#### Follow-Up Phase

#### Peto's Method

| Study | Experimental Events | Experimental Total | Control Events | Control Total | OR | 95%-CI | Weight (common) | Weight (random) |
|---|---|---|---|---|---|---|---|---|
| Abbar 2022 | 5 | 73 | 8 | 83 | 0.70 | [0.22; 2.16] | 39.7% | 29.7% |
| Ahmed 2023 | 0 | 18 | 0 | 18 | | | 0.0% | 0.0% |
| Anderson 2017 | 0 | 25 | 1 | 23 | 0.12 | [0.00; 6.27] | 3.3% | 4.7% |
| Burger 2016 | 0 | 3 | 0 | 7 | | | 0.0% | 0.0% |
| Canuso 2018 | 0 | 27 | 3 | 22 | 0.10 | [0.01; 1.00] | 9.5% | 11.6% |
| Dakwar 2019 | 0 | 27 | 0 | 28 | | | 0.0% | 0.0% |
| Dakwar 2020 | 0 | 17 | 0 | 23 | | | 0.0% | 0.0% |
| Domany 2019 | 0 | 22 | 0 | 18 | | | 0.0% | 0.0% |
| Domany 2020 | 0 | 9 | 0 | 9 | | | 0.0% | 0.0% |
| Duek 2023 | 0 | 14 | 0 | 14 | | | 0.0% | 0.0% |
| Dwyer 2021 | 0 | 11 | 0 | 6 | | | 0.0% | 0.0% |
| Fava 2020 | 1 | 80 | 0 | 19 | 3.45 | [0.02; 499.91] | 2.1% | 3.0% |
| Feder 2014 | 0 | 22 | 0 | 19 | | | 0.0% | 0.0% |
| Fineberg 2023 | 0 | 10 | 0 | 12 | | | 0.0% | 0.0% |
| Fu 2020 | 3 | 101 | 2 | 91 | 1.35 | [0.23; 7.98] | 16.2% | 17.4% |
| Glue 2024b | 0 | 17 | 0 | 8 | | | 0.0% | 0.0% |
| Grabski 2022 | 0 | 48 | 0 | 48 | | | 0.0% | 0.0% |
| Ionescu 2020 | 4 | 89 | 1 | 94 | 3.61 | [0.61; 21.27] | 16.3% | 17.4% |
| Jones 2024 | 0 | 17 | 0 | 11 | | | 0.0% | 0.0% |
| Lapidus 2014 | 0 | 10 | 0 | 10 | | | 0.0% | 0.0% |
| Leal 2023 | 0 | 5 | 0 | 5 | | | 0.0% | 0.0% |
| Lii 2023 | 0 | 20 | 0 | 20 | | | 0.0% | 0.0% |
| Lijffijt 2021 | 0 | 20 | 1 | 13 | 0.08 | [0.00; 4.36] | 3.2% | 4.5% |
| Murrough 2013 | 0 | 47 | 0 | 25 | | | 0.0% | 0.0% |
| Murrough 2015 II | 0 | 12 | 0 | 12 | | | 0.0% | 0.0% |
| Ohtani 2024 | 0 | 17 | 0 | 17 | | | 0.0% | 0.0% |
| Pattanaseri 2024 | 0 | 11 | 0 | 9 | | | 0.0% | 0.0% |
| Phillips 2019 | 0 | 22 | 0 | 21 | | | 0.0% | 0.0% |
| Price 2022 | 0 | 53 | 0 | 51 | | | 0.0% | 0.0% |
| Ray-Griffith 2017 | 0 | 8 | 0 | 8 | | | 0.0% | 0.0% |
| Rodriguez 2013 | 0 | 8 | 0 | 7 | | | 0.0% | 0.0% |
| Schiweck 2021 | 0 | 21 | 0 | 7 | | | 0.0% | 0.0% |
| Smith-Apeldoorn 2024 | 0 | 49 | 0 | 48 | | | 0.0% | 0.0% |
| Sos 2013 | 0 | 11 | 0 | 19 | | | 0.0% | 0.0% |
| Su 2017 | 0 | 47 | 0 | 24 | | | 0.0% | 0.0% |
| Su 2023 | 1 | 42 | 2 | 42 | 0.51 | [0.05; 4.99] | 9.7% | 11.8% |
| Sumner 2019 | 0 | 17 | 0 | 16 | | | 0.0% | 0.0% |
| Taylor 2018 | 0 | 9 | 0 | 9 | | | 0.0% | 0.0% |
| Wang 2024 | 0 | 182 | 0 | 182 | | | 0.0% | 0.0% |
| Xiao 2023 | 0 | 15 | 0 | 15 | | | 0.0% | 0.0% |
| Yang 2023 | 0 | 200 | 0 | 100 | | | 0.0% | 0.0% |
| Zarate 2006 | 0 | 9 | 0 | 9 | | | 0.0% | 0.0% |
| Zarate 2012 | 0 | 21 | 0 | 23 | | | 0.0% | 0.0% |
| Zhou 2024 | 0 | 27 | 0 | 25 | | | 0.0% | 0.0% |
| **Common effect model** | | **1513** | | **1270** | **0.74** | **[0.36; 1.52]** | **100.0%** | **.** |
| **Random effects model** | | | | | **0.70** | **[0.29; 1.69]** | **.** | **100.0%** |

Heterogeneity: $I^2 = 21.4\%$, $\tau^2 = 0.3469$, $p = 0.2598$

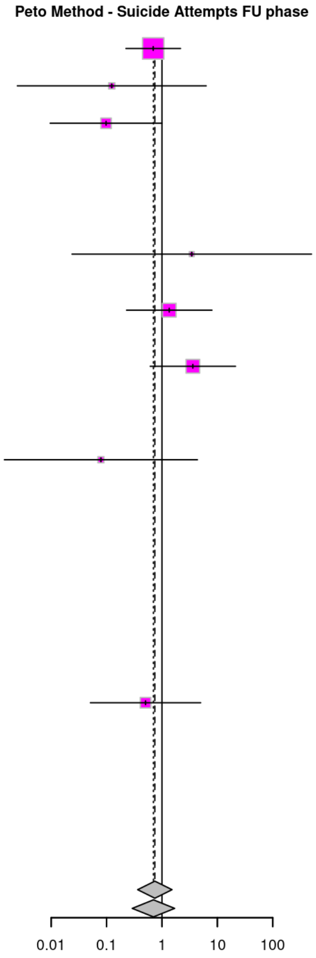

#### Maentel Mantel-Haenszel with CC

| Study | Experimental Events | Experimental Total | Control Events | Control Total | MH with CC - Suicide Attempts FU phase | OR | 95%-CI | Weight (common) | Weight (random) |
|---|---|---|---|---|---|---|---|---|---|
| Abbar 2022 | 5 | 73 | 8 | 83 | | 0.69 | [0.22; 2.21] | 18.3% | 18.1% |
| Ahmed 2023 | 0 | 18 | 0 | 18 | | 1.00 | [0.02; 53.12] | 1.3% | 1.6% |
| Anderson 2017 | 0 | 25 | 1 | 23 | | 0.29 | [0.01; 7.59] | 4.0% | 2.3% |
| Burger 2016 | 0 | 3 | 0 | 7 | | 2.14 | [0.03; 131.93] | 0.8% | 1.4% |
| Canuso 2018 | 0 | 27 | 3 | 22 | | 0.10 | [0.00; 2.07] | 9.9% | 2.7% |
| Dakwar 2019 | 0 | 27 | 0 | 28 | | 1.04 | [0.02; 54.08] | 1.3% | 1.6% |
| Dakwar 2020 | 0 | 17 | 0 | 23 | | 1.34 | [0.03; 71.04] | 1.1% | 1.6% |
| Domany 2019 | 0 | 22 | 0 | 18 | | 0.82 | [0.02; 43.47] | 1.4% | 1.6% |
| Domany 2020 | 0 | 9 | 0 | 9 | | 1.00 | [0.02; 55.80] | 1.2% | 1.5% |
| Duek 2023 | 0 | 14 | 0 | 14 | | 1.00 | [0.02; 53.89] | 1.3% | 1.5% |
| Dwyer 2021 | 0 | 11 | 0 | 6 | | 0.57 | [0.01; 32.00] | 1.6% | 1.5% |
| Fava 2020 | 1 | 80 | 0 | 19 | | 0.74 | [0.03; 18.76] | 2.1% | 2.3% |
| Feder 2014 | 0 | 22 | 0 | 19 | | 0.87 | [0.02; 45.76] | 1.4% | 1.6% |
| Fineberg 2023 | 0 | 10 | 0 | 12 | | 1.19 | [0.02; 65.32] | 1.2% | 1.5% |
| Fu 2020 | 3 | 101 | 2 | 91 | | 1.36 | [0.22; 8.34] | 5.4% | 7.5% |
| Glue 2024b | 0 | 17 | 0 | 8 | | 0.49 | [0.01; 26.64] | 1.7% | 1.5% |
| Grabski 2022 | 0 | 48 | 0 | 48 | | 1.00 | [0.02; 51.42] | 1.3% | 1.6% |
| Ionescu 2020 | 4 | 89 | 1 | 94 | | 4.38 | [0.48; 39.93] | 2.4% | 5.0% |
| Jones 2024 | 0 | 17 | 0 | 11 | | 0.66 | [0.01; 35.52] | 1.5% | 1.5% |
| Lapidus 2014 | 0 | 10 | 0 | 10 | | 1.00 | [0.02; 55.27] | 1.3% | 1.5% |
| Leal 2023 | 0 | 5 | 0 | 5 | | 1.00 | [0.02; 59.99] | 1.2% | 1.5% |
| Lii 2023 | 0 | 20 | 0 | 20 | | 1.00 | [0.02; 52.85] | 1.3% | 1.6% |
| Lijffijt 2021 | 0 | 20 | 1 | 13 | | 0.20 | [0.01; 5.39] | 4.6% | 2.3% |
| Murrough 2013 | 0 | 47 | 0 | 25 | | 0.54 | [0.01; 27.86] | 1.7% | 1.6% |
| Murrough 2015 II | 0 | 12 | 0 | 12 | | 1.00 | [0.02; 54.46] | 1.3% | 1.5% |
| Ohtani 2024 | 0 | 17 | 0 | 17 | | 1.00 | [0.02; 53.28] | 1.3% | 1.6% |
| Pattanaseri 2024 | 0 | 11 | 0 | 9 | | 0.83 | [0.01; 45.69] | 1.4% | 1.5% |
| Phillips 2019 | 0 | 22 | 0 | 21 | | 0.96 | [0.02; 50.34] | 1.3% | 1.6% |
| Price 2022 | 0 | 53 | 0 | 51 | | 0.96 | [0.02; 49.43] | 1.3% | 1.6% |
| Ray-Griffith 2017 | 0 | 8 | 0 | 8 | | 1.00 | [0.02; 56.46] | 1.2% | 1.5% |
| Rodriguez 2013 | 0 | 8 | 0 | 7 | | 0.88 | [0.02; 50.19] | 1.3% | 1.5% |
| Schiweck 2021 | 0 | 21 | 0 | 7 | | 0.35 | [0.01; 19.18] | 1.9% | 1.5% |
| Smith-Apeldoorn 2024 | 0 | 49 | 0 | 48 | | 0.98 | [0.02; 50.37] | 1.3% | 1.6% |
| Sos 2013 | 0 | 11 | 0 | 19 | | 1.70 | [0.03; 91.39] | 0.9% | 1.5% |
| Su 2017 | 0 | 47 | 0 | 24 | | 0.52 | [0.01; 26.79] | 1.7% | 1.6% |
| Su 2023 | 1 | 42 | 2 | 42 | | 0.49 | [0.04; 5.59] | 5.1% | 4.1% |
| Sumner 2019 | 0 | 17 | 0 | 16 | | 0.94 | [0.02; 50.32] | 1.3% | 1.6% |
| Taylor 2018 | 0 | 9 | 0 | 9 | | 1.00 | [0.02; 55.80] | 1.2% | 1.5% |
| Wang 2024 | 0 | 182 | 0 | 182 | | 1.00 | [0.02; 50.67] | 1.3% | 1.6% |
| Xiao 2023 | 0 | 15 | 0 | 15 | | 1.00 | [0.02; 53.66] | 1.3% | 1.5% |
| Yang 2023 | 0 | 200 | 0 | 100 | | 0.50 | [0.01; 25.45] | 1.7% | 1.6% |
| Zarate 2006 | 0 | 9 | 0 | 9 | | 1.00 | [0.02; 55.80] | 1.2% | 1.5% |
| Zarate 2012 | 0 | 21 | 0 | 23 | | 1.09 | [0.02; 57.53] | 1.2% | 1.6% |
| Zhou 2024 | 0 | 27 | 0 | 25 | | 0.93 | [0.02; 48.49] | 1.3% | 1.6% |
| **Common effect model** | | **1513** | | **1270** | | **0.81** | **[0.51; 1.30]** | **100.0%** | **.** |
| **Random effects model** | | | | | | **0.83** | **[0.50; 1.35]** | **.** | **100.0%** |

Heterogeneity: $I^2 = 0.0\%$, $\tau^2 = 0$, $p = 1.0000$

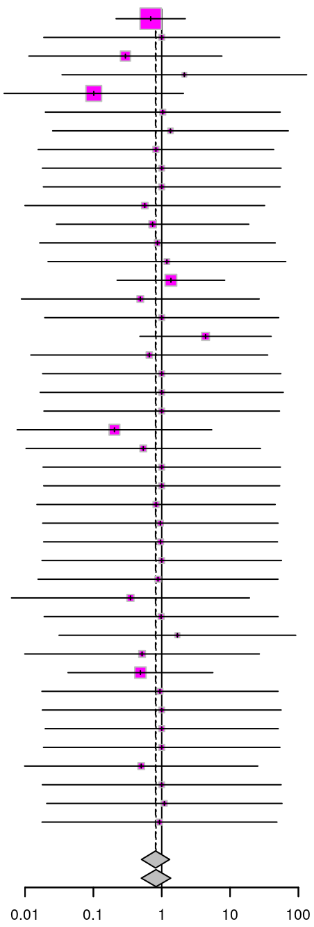

#### Maentel Mantel-Haenszel with Treatment-Arm CC

| Study | Experimental Events | Experimental Total | Control Events | Control Total | OR | 95%-CI | Weight (common) | Weight (random) |
|---|---|---|---|---|---|---|---|---|
| Abbar 2022 | 5 | 73 | 8 | 83 | 0.69 | [0.22; 2.21] | 19.4% | 18.5% |
| Ahmed 2023 | 0 | 18 | 0 | 18 | 1.00 | [0.02; 53.12] | 1.4% | 1.6% |
| Anderson 2017 | 0 | 25 | 1 | 23 | 0.31 | [0.01; 7.67] | 4.2% | 2.4% |
| Burger 2016 | 0 | 3 | 0 | 7 | 1.00 | [0.01; 87.11] | 1.1% | 1.3% |
| Canuso 2018 | 0 | 27 | 3 | 22 | 0.11 | [0.01; 2.05] | 10.4% | 3.0% |
| Dakwar 2019 | 0 | 27 | 0 | 28 | 1.00 | [0.02; 52.22] | 1.4% | 1.6% |
| Dakwar 2020 | 0 | 17 | 0 | 23 | 1.00 | [0.02; 55.30] | 1.3% | 1.6% |
| Domany 2019 | 0 | 22 | 0 | 18 | 1.00 | [0.02; 53.92] | 1.3% | 1.6% |
| Domany 2020 | 0 | 9 | 0 | 9 | 1.00 | [0.02; 55.80] | 1.3% | 1.6% |
| Duek 2023 | 0 | 14 | 0 | 14 | 1.00 | [0.02; 53.89] | 1.3% | 1.6% |
| Dwyer 2021 | 0 | 11 | 0 | 6 | 1.00 | [0.01; 67.61] | 1.2% | 1.4% |
| Fava 2020 | 1 | 80 | 0 | 19 | 2.27 | [0.02; 257.11] | 0.8% | 1.1% |
| Feder 2014 | 0 | 22 | 0 | 19 | 1.00 | [0.02; 53.36] | 1.4% | 1.6% |
| Fineberg 2023 | 0 | 10 | 0 | 12 | 1.00 | [0.02; 55.75] | 1.3% | 1.6% |
| Fu 2020 | 3 | 101 | 2 | 91 | 1.36 | [0.22; 8.34] | 5.7% | 7.7% |
| Glue 2024b | 0 | 17 | 0 | 8 | 1.00 | [0.01; 72.36] | 1.2% | 1.4% |
| Grabski 2022 | 0 | 48 | 0 | 48 | 1.00 | [0.02; 51.42] | 1.4% | 1.6% |
| Ionescu 2020 | 4 | 89 | 1 | 94 | 4.38 | [0.48; 39.93] | 2.6% | 5.1% |
| Jones 2024 | 0 | 17 | 0 | 11 | 1.00 | [0.02; 59.25] | 1.3% | 1.5% |
| Lapidus 2014 | 0 | 10 | 0 | 10 | 1.00 | [0.02; 55.27] | 1.3% | 1.6% |
| Leal 2023 | 0 | 5 | 0 | 5 | 1.00 | [0.02; 59.99] | 1.3% | 1.5% |
| Lii 2023 | 0 | 20 | 0 | 20 | 1.00 | [0.02; 52.85] | 1.4% | 1.6% |
| Lijffijt 2021 | 0 | 20 | 1 | 13 | 0.26 | [0.01; 5.79] | 4.6% | 2.6% |
| Murrough 2013 | 0 | 47 | 0 | 25 | 1.00 | [0.02; 63.11] | 1.2% | 1.5% |
| Murrough 2015 II | 0 | 12 | 0 | 12 | 1.00 | [0.02; 54.46] | 1.3% | 1.6% |
| Ohtani 2024 | 0 | 17 | 0 | 17 | 1.00 | [0.02; 53.28] | 1.4% | 1.6% |
| Pattanaseri 2024 | 0 | 11 | 0 | 9 | 1.00 | [0.02; 56.40] | 1.3% | 1.5% |
| Phillips 2019 | 0 | 22 | 0 | 21 | 1.00 | [0.02; 52.74] | 1.4% | 1.6% |
| Price 2022 | 0 | 53 | 0 | 51 | 1.00 | [0.02; 51.38] | 1.4% | 1.6% |
| Ray-Griffith 2017 | 0 | 8 | 0 | 8 | 1.00 | [0.02; 56.46] | 1.3% | 1.5% |
| Rodriguez 2013 | 0 | 8 | 0 | 7 | 1.00 | [0.02; 57.37] | 1.3% | 1.5% |
| Schiweck 2021 | 0 | 21 | 0 | 7 | 1.00 | [0.01; 99.86] | 1.0% | 1.2% |
| Smith-Apeldoorn 2024 | 0 | 49 | 0 | 48 | 1.00 | [0.02; 51.42] | 1.4% | 1.6% |
| Sos 2013 | 0 | 11 | 0 | 19 | 1.00 | [0.02; 62.32] | 1.3% | 1.5% |
| Su 2017 | 0 | 47 | 0 | 24 | 1.00 | [0.02; 64.85] | 1.2% | 1.4% |
| Su 2023 | 1 | 42 | 2 | 42 | 0.49 | [0.04; 5.59] | 5.4% | 4.2% |
| Sumner 2019 | 0 | 17 | 0 | 16 | 1.00 | [0.02; 53.46] | 1.3% | 1.6% |
| Taylor 2018 | 0 | 9 | 0 | 9 | 1.00 | [0.02; 55.80] | 1.3% | 1.6% |
| Wang 2024 | 0 | 182 | 0 | 182 | 1.00 | [0.02; 50.67] | 1.4% | 1.6% |
| Xiao 2023 | 0 | 15 | 0 | 15 | 1.00 | [0.02; 53.66] | 1.3% | 1.6% |
| Yang 2023 | 0 | 200 | 0 | 100 | 1.00 | [0.02; 64.37] | 1.2% | 1.5% |
| Zarate 2006 | 0 | 9 | 0 | 9 | 1.00 | [0.02; 55.80] | 1.3% | 1.6% |
| Zarate 2012 | 0 | 21 | 0 | 23 | 1.00 | [0.02; 52.84] | 1.4% | 1.6% |
| Zhou 2024 | 0 | 27 | 0 | 25 | 1.00 | [0.02; 52.44] | 1.4% | 1.6% |
| **Common effect model** | | **1513** | | **1270** | **0.88** | **[0.54; 1.41]** | **100.0%** | . |
| **Random effects model** | | | | | **0.89** | **[0.54; 1.47]** | . | **100.0%** |

Heterogeneity: $I^2 = 0.0\%$, $\tau^2 = 0$, $p = 1.0000$

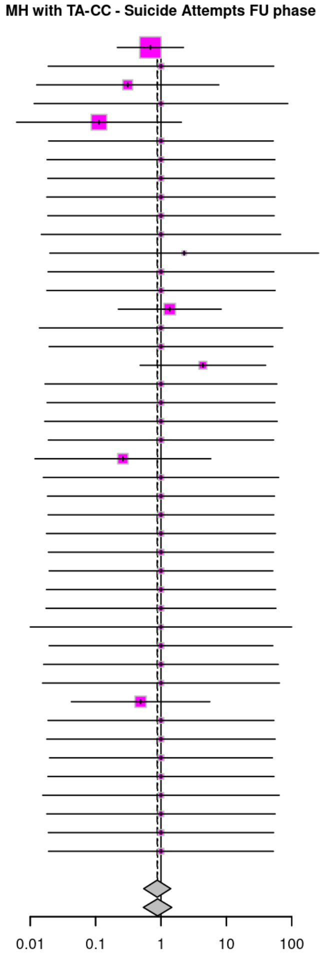

#### Maentel Mantel-Haenszel Risk-Difference

| Study | Experimental Events | Experimental Total | Control Events | Control Total | RD | 95%-CI | Weight (common) | Weight (random) |
|---|---|---|---|---|---|---|---|---|
| Abbar 2022 | 5 | 73 | 8 | 83 | -0.03 | [-0.11; 0.06] | 5.6% | 0.7% |
| Ahmed 2023 | 0 | 18 | 0 | 18 | 0.00 | [-0.10; 0.10] | 1.4% | 0.5% |
| Anderson 2017 | 0 | 25 | 1 | 23 | -0.04 | [-0.15; 0.07] | 1.8% | 0.4% |
| Burger 2016 | 0 | 3 | 0 | 7 | 0.00 | [-0.36; 0.36] | 0.4% | 0.0% |
| Canuso 2018 | 0 | 27 | 3 | 22 | -0.14 | [-0.29; 0.02] | 1.8% | 0.2% |
| Dakwar 2019 | 0 | 27 | 0 | 28 | 0.00 | [-0.07; 0.07] | 2.1% | 1.1% |
| Dakwar 2020 | 0 | 17 | 0 | 23 | 0.00 | [-0.10; 0.10] | 1.5% | 0.6% |
| Domany 2019 | 0 | 22 | 0 | 18 | 0.00 | [-0.09; 0.09] | 1.5% | 0.6% |
| Domany 2020 | 0 | 9 | 0 | 9 | 0.00 | [-0.19; 0.19] | 0.7% | 0.1% |
| Duek 2023 | 0 | 14 | 0 | 14 | 0.00 | [-0.13; 0.13] | 1.1% | 0.3% |
| Dwyer 2021 | 0 | 11 | 0 | 6 | 0.00 | [-0.22; 0.22] | 0.6% | 0.1% |
| Fava 2020 | 1 | 80 | 0 | 19 | 0.01 | [-0.06; 0.09] | 2.3% | 0.9% |
| Feder 2014 | 0 | 22 | 0 | 19 | 0.00 | [-0.09; 0.09] | 1.6% | 0.6% |
| Fineberg 2023 | 0 | 10 | 0 | 12 | 0.00 | [-0.16; 0.16] | 0.9% | 0.2% |
| Fu 2020 | 3 | 101 | 2 | 91 | 0.01 | [-0.04; 0.05] | 6.9% | 2.6% |
| Glue 2024b | 0 | 17 | 0 | 8 | 0.00 | [-0.17; 0.17] | 0.9% | 0.2% |
| Grabski 2022 | 0 | 48 | 0 | 48 | 0.00 | [-0.04; 0.04] | 3.6% | 3.3% |
| Ionescu 2020 | 4 | 89 | 1 | 94 | 0.03 | [-0.01; 0.08] | 6.6% | 2.3% |
| Jones 2024 | 0 | 17 | 0 | 11 | 0.00 | [-0.14; 0.14] | 1.0% | 0.3% |
| Lapidus 2014 | 0 | 10 | 0 | 10 | 0.00 | [-0.17; 0.17] | 0.8% | 0.2% |
| Leal 2023 | 0 | 5 | 0 | 5 | 0.00 | [-0.31; 0.31] | 0.4% | 0.1% |
| Lii 2023 | 0 | 20 | 0 | 20 | 0.00 | [-0.09; 0.09] | 1.5% | 0.6% |
| Lijffijt 2021 | 0 | 20 | 1 | 13 | -0.08 | [-0.25; 0.10] | 1.2% | 0.2% |
| Murrough 2013 | 0 | 47 | 0 | 25 | 0.00 | [-0.06; 0.06] | 2.4% | 1.4% |
| Murrough 2015 II | 0 | 12 | 0 | 12 | 0.00 | [-0.15; 0.15] | 0.9% | 0.2% |
| Ohtani 2024 | 0 | 17 | 0 | 17 | 0.00 | [-0.11; 0.11] | 1.3% | 0.5% |
| Pattanaseri 2024 | 0 | 11 | 0 | 9 | 0.00 | [-0.18; 0.18] | 0.8% | 0.2% |
| Phillips 2019 | 0 | 22 | 0 | 21 | 0.00 | [-0.09; 0.09] | 1.6% | 0.7% |
| Price 2022 | 0 | 53 | 0 | 51 | 0.00 | [-0.04; 0.04] | 3.8% | 3.9% |
| Ray-Griffith 2017 | 0 | 8 | 0 | 8 | 0.00 | [-0.21; 0.21] | 0.7% | 0.1% |
| Rodriguez 2013 | 0 | 8 | 0 | 7 | 0.00 | [-0.22; 0.22] | 0.6% | 0.1% |
| Schiweck 2021 | 0 | 21 | 0 | 7 | 0.00 | [-0.18; 0.18] | 0.9% | 0.2% |
| Smith-Apeldoorn 2024 | 0 | 49 | 0 | 48 | 0.00 | [-0.04; 0.04] | 3.6% | 3.4% |
| Sos 2013 | 0 | 11 | 0 | 19 | 0.00 | [-0.13; 0.13] | 1.1% | 0.3% |
| Su 2017 | 0 | 47 | 0 | 24 | 0.00 | [-0.06; 0.06] | 2.4% | 1.4% |
| Su 2023 | 1 | 42 | 2 | 42 | -0.02 | [-0.10; 0.06] | 3.0% | 0.8% |
| Sumner 2019 | 0 | 17 | 0 | 16 | 0.00 | [-0.11; 0.11] | 1.3% | 0.4% |
| Taylor 2018 | 0 | 9 | 0 | 9 | 0.00 | [-0.19; 0.19] | 0.7% | 0.1% |
| Wang 2024 | 0 | 182 | 0 | 182 | 0.00 | [-0.01; 0.01] | 13.3% | 45.6% |
| Xiao 2023 | 0 | 15 | 0 | 15 | 0.00 | [-0.12; 0.12] | 1.2% | 0.4% |
| Yang 2023 | 0 | 200 | 0 | 100 | 0.00 | [-0.02; 0.02] | 9.7% | 22.2% |
| Zarate 2006 | 0 | 9 | 0 | 9 | 0.00 | [-0.19; 0.19] | 0.7% | 0.1% |
| Zarate 2012 | 0 | 21 | 0 | 23 | 0.00 | [-0.08; 0.08] | 1.7% | 0.7% |
| Zhou 2024 | 0 | 27 | 0 | 25 | 0.00 | [-0.07; 0.07] | 2.0% | 1.0% |
| **Common effect model** | | **1513** | | **1270** | **-0.00** | **[-0.02; 0.01]** | **100.0%** | **.** |
| **Random effects model** | | | | | **0.00** | **[-0.01; 0.01]** | **.** | **100.0%** |

Heterogeneity: $I^2 = 0.0\%$, $\tau^2 = 0$, $p = 1.0000$

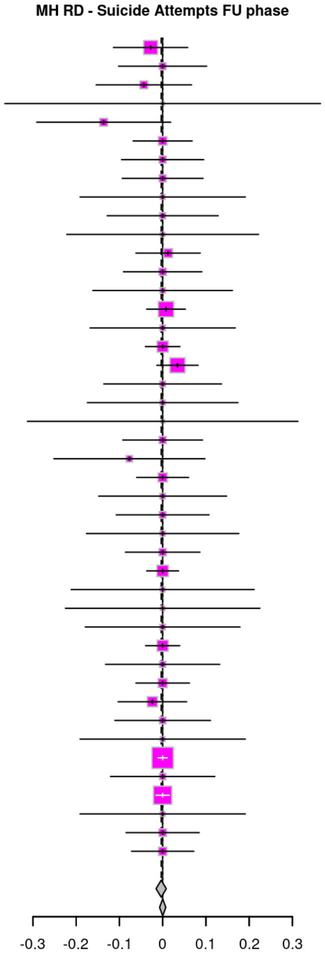

#### Arcsine Risk-Difference

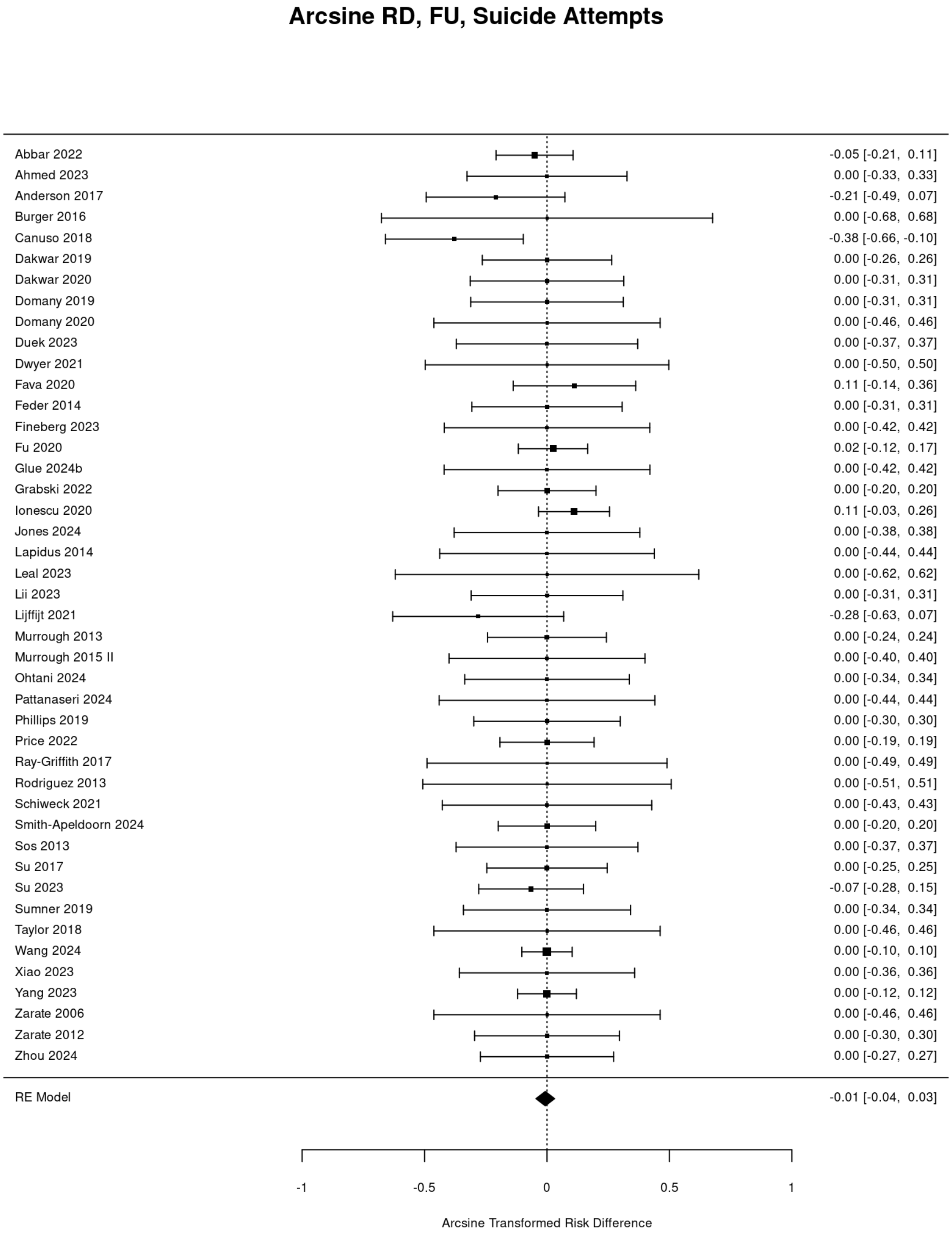

#### Bayesian Meta-Analysis, wider prior

**Bayesian MA, Delta = 250 - Suicide Attempts FU**

| Study | Estimate | 95% CI |
|---|---|---|
| Abbar 2022 | -0.37 | [-1.54, 0.79] |
| Ahmed 2023 | 0.00 | [-3.97, 3.97] |
| Anderson 2017 | -1.22 | [-4.47, 2.03] |
| Burger 2016 | 0.76 | [-3.36, 4.88] |
| Canuso 2018 | -2.29 | [-5.31, 0.73] |
| Dakwar 2019 | 0.04 | [-3.92, 3.99] |
| Dakwar 2020 | 0.29 | [-3.67, 4.26] |
| Domany 2019 | -0.20 | [-4.16, 3.77] |
| Domany 2020 | 0.00 | [-4.02, 4.02] |
| Duek 2023 | 0.00 | [-3.99, 3.99] |
| Dwyer 2021 | -0.57 | [-4.61, 3.47] |
| Fava 2020 | -0.31 | [-3.55, 2.93] |
| Feder 2014 | -0.14 | [-4.11, 3.82] |
| Fineberg 2023 | 0.17 | [-3.83, 4.18] |
| Fu 2020 | 0.31 | [-1.50, 2.12] |
| Glue 2024b | -0.72 | [-4.73, 3.28] |
| Grabski 2022 | 0.00 | [-3.94, 3.94] |
| Ionescu 2020 | 1.48 | [-0.73, 3.69] |
| Jones 2024 | -0.42 | [-4.41, 3.57] |
| Lapidus 2014 | 0.00 | [-4.01, 4.01] |
| Leal 2023 | 0.00 | [-4.09, 4.09] |
| Lii 2023 | 0.00 | [-3.97, 3.97] |
| Lijffijt 2021 | -1.59 | [-4.87, 1.68] |
| Murrough 2013 | -0.62 | [-4.57, 3.33] |
| Murrough 2015 II | 0.00 | [-4.00, 4.00] |
| Ohtani 2024 | 0.00 | [-3.98, 3.98] |
| Pattanaseri 2024 | -0.19 | [-4.20, 3.82] |
| Phillips 2019 | -0.05 | [-4.01, 3.92] |
| Price 2022 | -0.04 | [-3.98, 3.90] |
| Ray-Griffith 2017 | 0.00 | [-4.03, 4.03] |
| Rodriguez 2013 | -0.13 | [-4.17, 3.92] |
| Schiweck 2021 | -1.05 | [-5.06, 2.95] |
| Smith-Apeldoorn 2024 | -0.02 | [-3.96, 3.92] |
| Sos 2013 | 0.53 | [-3.46, 4.52] |
| Su 2017 | -0.66 | [-4.61, 3.29] |
| Su 2023 | -0.72 | [-3.16, 1.72] |
| Sumner 2019 | -0.06 | [-4.04, 3.92] |
| Taylor 2018 | 0.00 | [-4.02, 4.02] |
| Wang 2024 | 0.00 | [-3.93, 3.93] |
| Xiao 2023 | 0.00 | [-3.98, 3.98] |
| Yang 2023 | -0.69 | [-4.62, 3.24] |
| Zarate 2006 | 0.00 | [-4.02, 4.02] |
| Zarate 2012 | 0.09 | [-3.87, 4.05] |
| Zhou 2024 | -0.08 | [-4.03, 3.88] |
| **Summary** | **-0.30** | **[-1.10, 0.45]** |
| **Prediction** | **-0.30** | **[-1.61, 0.90]** |

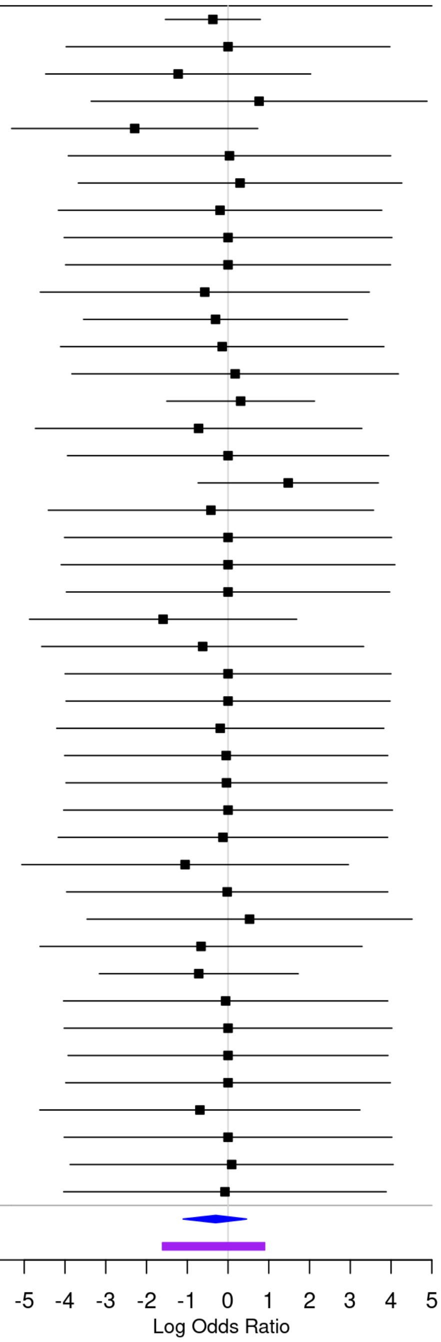

*Heterogeneity (tau): 0.32 [0.00, 0.90]*

#### Bayesian Meta-Analysis, smaller prior

##### Bayesian MA, Delta = 15 - Suicide Attempts FU

| Study | Estimate | 95% CI |
|---|---|---|
| Abbar 2022 | -0.37 | [-1.54, 0.79] |
| Ahmed 2023 | 0.00 | [-3.97, 3.97] |
| Anderson 2017 | -1.22 | [-4.47, 2.03] |
| Burger 2016 | 0.76 | [-3.36, 4.88] |
| Canuso 2018 | -2.29 | [-5.31, 0.73] |
| Dakwar 2019 | 0.04 | [-3.92, 3.99] |
| Dakwar 2020 | 0.29 | [-3.67, 4.26] |
| Domany 2019 | -0.20 | [-4.16, 3.77] |
| Domany 2020 | 0.00 | [-4.02, 4.02] |
| Duek 2023 | 0.00 | [-3.99, 3.99] |
| Dwyer 2021 | -0.57 | [-4.61, 3.47] |
| Fava 2020 | -0.31 | [-3.55, 2.93] |
| Feder 2014 | -0.14 | [-4.11, 3.82] |
| Fineberg 2023 | 0.17 | [-3.83, 4.18] |
| Fu 2020 | 0.31 | [-1.50, 2.12] |
| Glue 2024b | -0.72 | [-4.73, 3.28] |
| Grabski 2022 | 0.00 | [-3.94, 3.94] |
| Ionescu 2020 | 1.48 | [-0.73, 3.69] |
| Jones 2024 | -0.42 | [-4.41, 3.57] |
| Lapidus 2014 | 0.00 | [-4.01, 4.01] |
| Leal 2023 | 0.00 | [-4.09, 4.09] |
| Lii 2023 | 0.00 | [-3.97, 3.97] |
| Lijffijt 2021 | -1.59 | [-4.87, 1.68] |
| Murrough 2013 | -0.62 | [-4.57, 3.33] |
| Murrough 2015 II | 0.00 | [-4.00, 4.00] |
| Ohtani 2024 | 0.00 | [-3.98, 3.98] |
| Pattanaseri 2024 | -0.19 | [-4.20, 3.82] |
| Phillips 2019 | -0.05 | [-4.01, 3.92] |
| Price 2022 | -0.04 | [-3.98, 3.90] |
| Ray-Griffith 2017 | 0.00 | [-4.03, 4.03] |
| Rodriguez 2013 | -0.13 | [-4.17, 3.92] |
| Schiweck 2021 | -1.05 | [-5.06, 2.95] |
| Smith-Apeldoorn 2024 | -0.02 | [-3.96, 3.92] |
| Sos 2013 | 0.53 | [-3.46, 4.52] |
| Su 2017 | -0.66 | [-4.61, 3.29] |
| Su 2023 | -0.72 | [-3.16, 1.72] |
| Sumner 2019 | -0.06 | [-4.04, 3.92] |
| Taylor 2018 | 0.00 | [-4.02, 4.02] |
| Wang 2024 | 0.00 | [-3.93, 3.93] |
| Xiao 2023 | 0.00 | [-3.98, 3.98] |
| Yang 2023 | -0.69 | [-4.62, 3.24] |
| Zarate 2006 | 0.00 | [-4.02, 4.02] |
| Zarate 2012 | 0.09 | [-3.87, 4.05] |
| Zhou 2024 | -0.08 | [-4.03, 3.88] |
| **Summary** | **-0.29** | **[-1.06, 0.46]** |
| **Prediction** | **-0.29** | **[-1.55, 0.90]** |

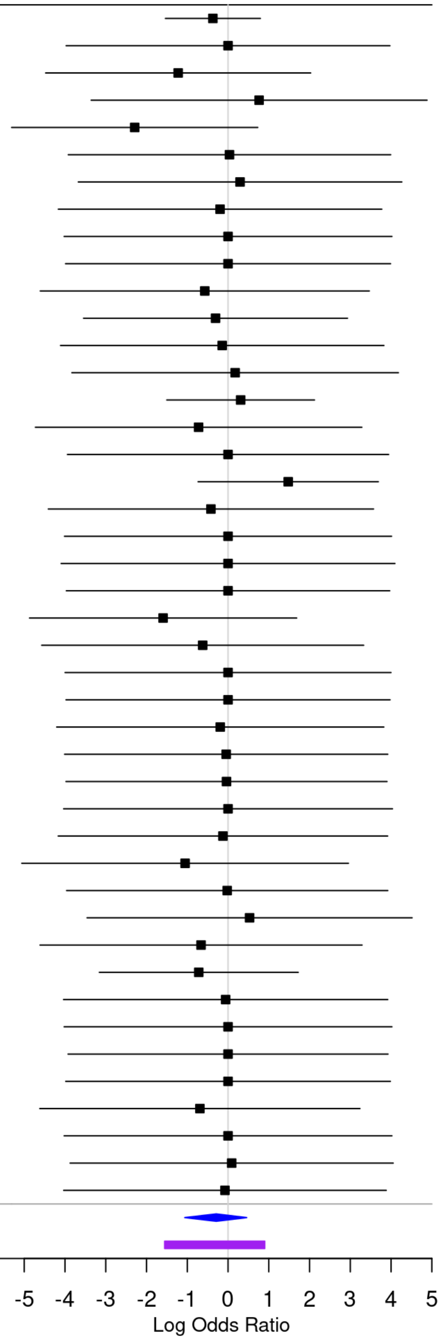

*Heterogeneity (tau): 0.32 [0.00, 0.89]*

**Exact Method**

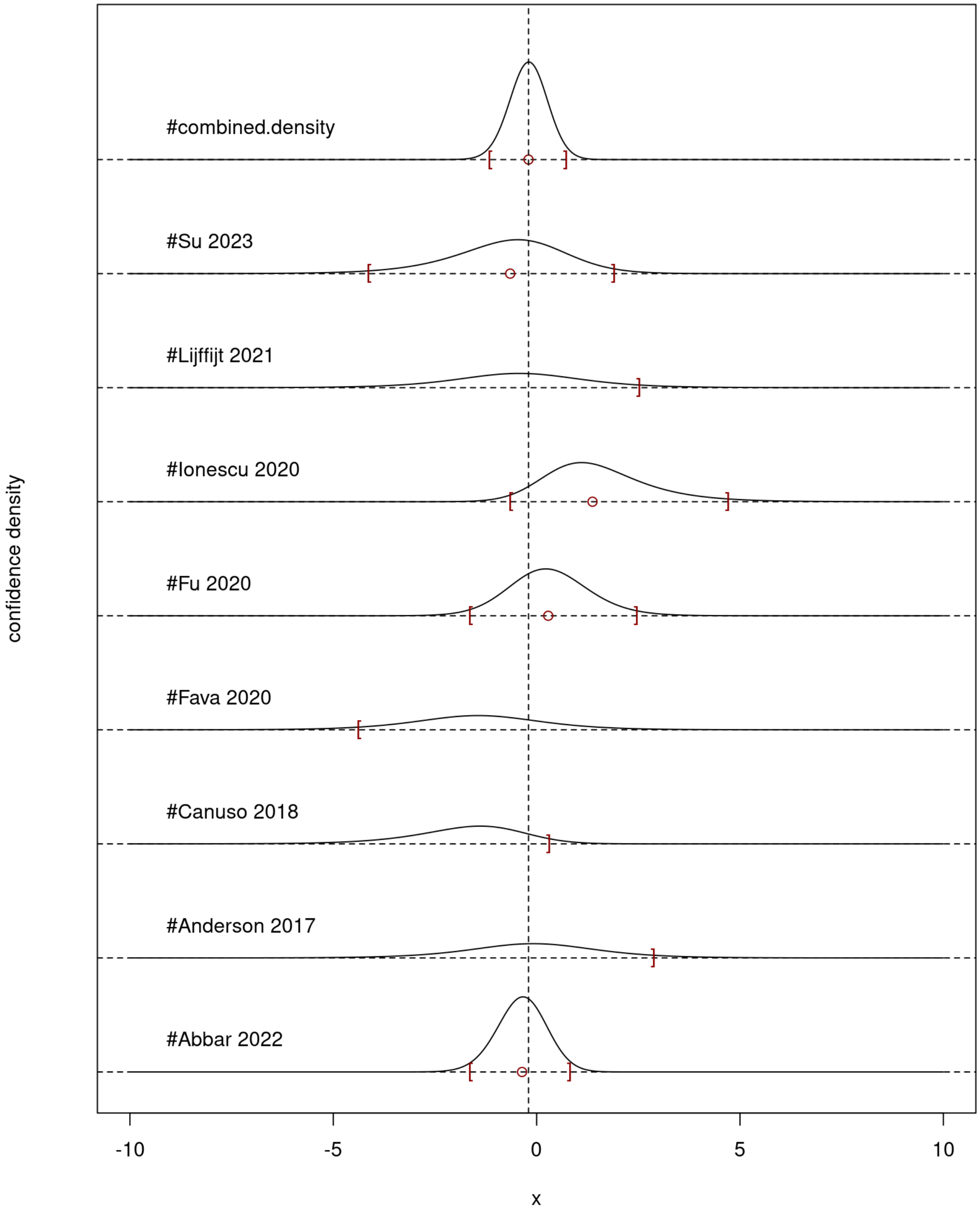

#### Treatment + Follow-Up Phase Combined

#### Peto's Method

| Study | Experimental Events | Experimental Total | Control Events | Control Total | OR | 95%-CI | Weight (common) | Weight (random) |
|---|---|---|---|---|---|---|---|---|
| Abbar 2022 | 6 | 73 | 8 | 83 | 0.84 | [0.28; 2.52] | 28.7% | 25.4% |
| Abdallah 2022 | 0 | 108 | 0 | 55 | | | 0.0% | 0.0% |
| Ahmed 2023 | 0 | 18 | 0 | 18 | | | 0.0% | 0.0% |
| Anderson 2017 | 0 | 40 | 2 | 39 | 0.13 | [0.01; 2.09] | 4.4% | 5.1% |
| Burger 2016 | 0 | 3 | 0 | 7 | | | 0.0% | 0.0% |
| Canuso 2018 | 0 | 35 | 3 | 31 | 0.11 | [0.01; 1.11] | 6.5% | 7.3% |
| Chen 2023 | 1 | 126 | 0 | 126 | 7.39 | [0.15; 372.38] | 2.2% | 2.7% |
| Colla 2024 | 0 | 17 | 0 | 10 | | | 0.0% | 0.0% |
| Dakwar 2019 | 0 | 27 | 0 | 28 | | | 0.0% | 0.0% |
| Dakwar 2020 | 0 | 17 | 0 | 23 | | | 0.0% | 0.0% |
| Daly 2018 PA | 0 | 34 | 0 | 33 | | | 0.0% | 0.0% |
| Daly 2018 PB | 0 | 20 | 0 | 21 | | | 0.0% | 0.0% |
| Domany 2019 | 0 | 22 | 0 | 18 | | | 0.0% | 0.0% |
| Domany 2020 | 0 | 9 | 0 | 9 | | | 0.0% | 0.0% |
| Duek 2023 | 0 | 14 | 0 | 14 | | | 0.0% | 0.0% |
| Dwyer 2021 | 0 | 11 | 0 | 6 | | | 0.0% | 0.0% |
| Fava 2020 | 1 | 80 | 0 | 19 | 3.45 | [0.02; 499.91] | 1.4% | 1.7% |
| Feder 2014 | 0 | 22 | 0 | 19 | | | 0.0% | 0.0% |
| Feder 2021 | 0 | 15 | 0 | 15 | | | 0.0% | 0.0% |
| Fedgchin 2019 | 0 | 231 | 0 | 113 | | | 0.0% | 0.0% |
| Fineberg 2023 | 0 | 10 | 0 | 12 | | | 0.0% | 0.0% |
| Fu 2020 | 4 | 113 | 3 | 112 | 1.33 | [0.30; 5.97] | 15.3% | 15.5% |
| George 2017 | 0 | 12 | 0 | 4 | | | 0.0% | 0.0% |
| Glue 2024b | 0 | 17 | 0 | 8 | | | 0.0% | 0.0% |
| Grabski 2022 | 0 | 48 | 0 | 48 | | | 0.0% | 0.0% |
| Grunebaum 2017 | 0 | 9 | 0 | 7 | | | 0.0% | 0.0% |
| Grunebaum 2018 | 0 | 40 | 0 | 40 | | | 0.0% | 0.0% |
| Ionescu 2020 | 7 | 114 | 4 | 113 | 1.75 | [0.52; 5.87] | 23.6% | 21.9% |
| Jones 2024 | 0 | 17 | 0 | 11 | | | 0.0% | 0.0% |
| Lapidus 2014 | 0 | 10 | 0 | 10 | | | 0.0% | 0.0% |
| Leal 2023 | 0 | 5 | 0 | 5 | | | 0.0% | 0.0% |
| Lii 2023 | 0 | 20 | 0 | 20 | | | 0.0% | 0.0% |
| Lijffijt 2021 | 0 | 20 | 1 | 13 | 0.08 | [0.00; 4.36] | 2.1% | 2.5% |
| Loo 2016 | 0 | 9 | 0 | 6 | | | 0.0% | 0.0% |
| Loo 2023 | 0 | 89 | 2 | 92 | 0.14 | [0.01; 2.23] | 4.5% | 5.1% |
| Milak 2020 | 0 | 38 | 0 | 5 | | | 0.0% | 0.0% |
| Murrough 2013 | 0 | 47 | 0 | 25 | | | 0.0% | 0.0% |
| Murrough 2015 II | 0 | 12 | 0 | 12 | | | 0.0% | 0.0% |
| Ochs-Ross 2020 | 0 | 72 | 0 | 65 | | | 0.0% | 0.0% |
| Ohtani 2024 | 0 | 17 | 0 | 17 | | | 0.0% | 0.0% |
| Pattanaseri 2024 | 0 | 11 | 0 | 9 | | | 0.0% | 0.0% |
| Phillips 2019 | 0 | 22 | 0 | 21 | | | 0.0% | 0.0% |
| Popova 2019 | 0 | 115 | 0 | 109 | | | 0.0% | 0.0% |
| Price 2022 | 0 | 53 | 0 | 51 | | | 0.0% | 0.0% |
| Ray-Griffith 2017 | 0 | 8 | 0 | 8 | | | 0.0% | 0.0% |
| Rodriguez 2013 | 0 | 8 | 0 | 7 | | | 0.0% | 0.0% |
| Schiweck 2021 | 0 | 21 | 0 | 7 | | | 0.0% | 0.0% |
| Shiroma 2020 | 0 | 29 | 0 | 25 | | | 0.0% | 0.0% |
| Singh 2016 2x doses | 1 | 18 | 0 | 16 | 6.61 | [0.13; 335.50] | 2.2% | 2.7% |
| Singh 2016 3x doses | 0 | 17 | 0 | 16 | | | 0.0% | 0.0% |
| Singh 2016 II | 0 | 20 | 0 | 10 | | | 0.0% | 0.0% |
| Smith-Apeldoorn 2024 | 0 | 57 | 0 | 54 | | | 0.0% | 0.0% |
| Sos 2013 | 0 | 11 | 0 | 19 | | | 0.0% | 0.0% |
| Su 2017 | 0 | 47 | 0 | 24 | | | 0.0% | 0.0% |
| Su 2023 | 1 | 42 | 2 | 42 | 0.51 | [0.05; 4.99] | 6.6% | 7.4% |
| Sumner 2019 | 0 | 17 | 0 | 16 | | | 0.0% | 0.0% |
| Takahashi 2021 | 0 | 122 | 0 | 80 | | | 0.0% | 0.0% |
| Taylor 2018 | 0 | 9 | 0 | 9 | | | 0.0% | 0.0% |
| Tiger 2020 | 0 | 20 | 0 | 10 | | | 0.0% | 0.0% |
| Wang 2024 | 0 | 182 | 0 | 182 | | | 0.0% | 0.0% |
| Xiao 2023 | 0 | 15 | 0 | 15 | | | 0.0% | 0.0% |
| Yang 2023 | 0 | 200 | 0 | 100 | | | 0.0% | 0.0% |
| Zarate 2006 | 0 | 9 | 0 | 9 | | | 0.0% | 0.0% |
| Zarate 2012 | 0 | 21 | 0 | 23 | | | 0.0% | 0.0% |
| Zhou 2024 | 0 | 27 | 1 | 27 | 0.14 | [0.00; 6.82] | 2.2% | 2.7% |
| **Common effect model** | | **2742** | | **2201** | **0.79** | **[0.44; 1.42]** | **100.0%** | . |
| **Random effects model** | | | | | **0.75** | **[0.39; 1.43]** | . | **100.0%** |

Heterogeneity: $I^2 = 15.2\%$, $\tau^2 = 0.1196$, $p = 0.2954$

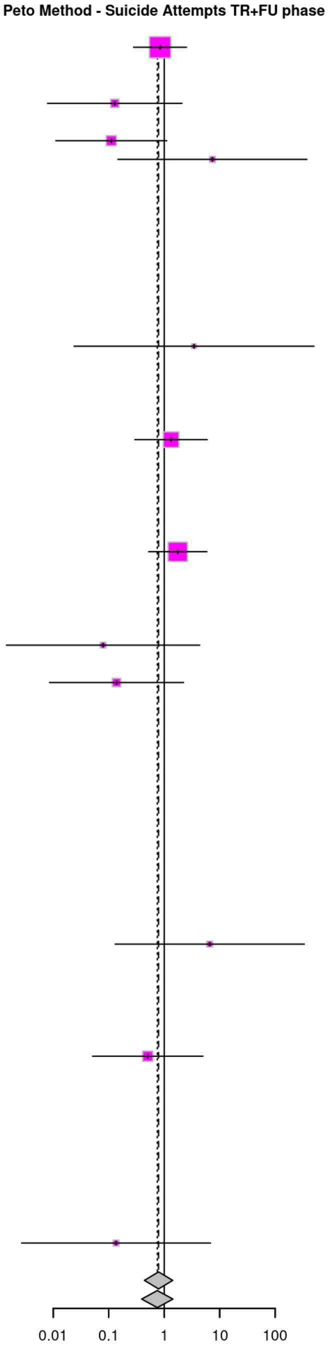

#### Maentel Mantel-Haenszel with CC

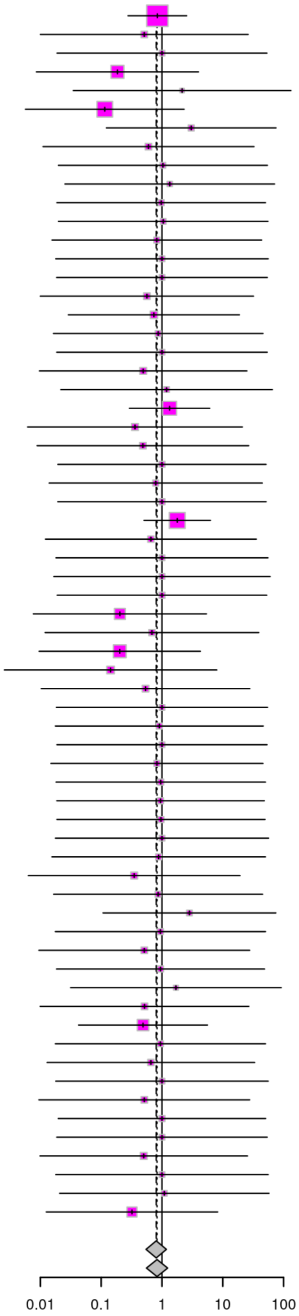

| Study | Experimental Events | Experimental Total | Control Events | Control Total | OR | 95%-CI | Weight (common) | Weight (random) |
|---|---|---|---|---|---|---|---|---|
| Abbar 2022 | 6 | 73 | 8 | 83 | 0.84 | [0.28; 2.54] | 12.0% | 13.2% |
| Abdallah 2022 | 0 | 108 | 0 | 55 | 0.51 | [0.01; 26.12] | 1.1% | 1.0% |
| Ahmed 2023 | 0 | 18 | 0 | 18 | 1.00 | [0.02; 53.12] | 0.8% | 1.0% |
| Anderson 2017 | 0 | 40 | 2 | 39 | 0.19 | [0.01; 3.98] | 4.4% | 1.7% |
| Burger 2016 | 0 | 3 | 0 | 7 | 2.14 | [0.03; 131.93] | 0.5% | 1.0% |
| Canuso 2018 | 0 | 35 | 3 | 31 | 0.11 | [0.01; 2.31] | 6.4% | 1.8% |
| Chen 2023 | 1 | 126 | 0 | 126 | 3.02 | [0.12; 74.94] | 0.9% | 1.6% |
| Colla 2024 | 0 | 17 | 0 | 10 | 0.60 | [0.01; 32.56] | 1.1% | 1.0% |
| Dakwar 2019 | 0 | 27 | 0 | 28 | 1.04 | [0.02; 54.08] | 0.8% | 1.0% |
| Dakwar 2020 | 0 | 17 | 0 | 23 | 1.34 | [0.03; 71.04] | 0.7% | 1.0% |
| Daly 2018 PA | 0 | 34 | 0 | 33 | 0.97 | [0.02; 50.36] | 0.9% | 1.0% |
| Daly 2018 PB | 0 | 20 | 0 | 21 | 1.05 | [0.02; 55.37] | 0.8% | 1.0% |
| Domany 2019 | 0 | 22 | 0 | 18 | 0.82 | [0.02; 43.47] | 0.9% | 1.0% |
| Domany 2020 | 0 | 9 | 0 | 9 | 1.00 | [0.02; 55.80] | 0.8% | 1.0% |
| Duek 2023 | 0 | 14 | 0 | 14 | 1.00 | [0.02; 53.89] | 0.8% | 1.0% |
| Dwyer 2021 | 0 | 11 | 0 | 6 | 0.57 | [0.01; 32.00] | 1.1% | 1.0% |
| Fava 2020 | 1 | 80 | 0 | 19 | 0.74 | [0.03; 18.76] | 1.4% | 1.5% |
| Feder 2014 | 0 | 22 | 0 | 19 | 0.87 | [0.02; 45.76] | 0.9% | 1.0% |
| Feder 2021 | 0 | 15 | 0 | 15 | 1.00 | [0.02; 53.66] | 0.8% | 1.0% |
| Fedgchin 2019 | 0 | 231 | 0 | 113 | 0.49 | [0.01; 24.87] | 1.2% | 1.0% |
| Fineberg 2023 | 0 | 10 | 0 | 12 | 1.19 | [0.02; 65.32] | 0.8% | 1.0% |
| Fu 2020 | 4 | 113 | 3 | 112 | 1.33 | [0.29; 6.10] | 5.1% | 7.0% |
| George 2017 | 0 | 12 | 0 | 4 | 0.36 | [0.01; 20.98] | 1.2% | 1.0% |
| Glue 2024b | 0 | 17 | 0 | 8 | 0.49 | [0.01; 26.64] | 1.1% | 1.0% |
| Grabski 2022 | 0 | 48 | 0 | 48 | 1.00 | [0.02; 51.42] | 0.9% | 1.0% |
| Grunebaum 2017 | 0 | 9 | 0 | 7 | 0.79 | [0.01; 44.65] | 0.9% | 1.0% |
| Grunebaum 2018 | 0 | 40 | 0 | 40 | 1.00 | [0.02; 51.63] | 0.9% | 1.0% |
| Ionescu 2020 | 7 | 114 | 4 | 113 | 1.78 | [0.51; 6.27] | 6.6% | 10.2% |
| Jones 2024 | 0 | 17 | 0 | 11 | 0.66 | [0.01; 35.52] | 1.0% | 1.0% |
| Lapidus 2014 | 0 | 10 | 0 | 10 | 1.00 | [0.02; 55.27] | 0.8% | 1.0% |
| Leal 2023 | 0 | 5 | 0 | 5 | 1.00 | [0.02; 59.99] | 0.8% | 1.0% |
| Lii 2023 | 0 | 20 | 0 | 20 | 1.00 | [0.02; 52.85] | 0.9% | 1.0% |
| Lijffijt 2021 | 0 | 20 | 1 | 13 | 0.20 | [0.01; 5.39] | 3.1% | 1.5% |
| Loo 2016 | 0 | 9 | 0 | 6 | 0.68 | [0.01; 39.07] | 1.0% | 1.0% |
| Loo 2023 | 0 | 89 | 2 | 92 | 0.20 | [0.01; 4.27] | 4.3% | 1.7% |
| Milak 2020 | 0 | 38 | 0 | 5 | 0.14 | [0.00; 7.96] | 1.5% | 1.0% |
| Murrough 2013 | 0 | 47 | 0 | 25 | 0.54 | [0.01; 27.86] | 1.1% | 1.0% |
| Murrough 2015 II | 0 | 12 | 0 | 12 | 1.00 | [0.02; 54.46] | 0.8% | 1.0% |
| Ochs-Ross 2020 | 0 | 72 | 0 | 65 | 0.90 | [0.02; 46.18] | 0.9% | 1.0% |
| Ohtani 2024 | 0 | 17 | 0 | 17 | 1.00 | [0.02; 53.28] | 0.8% | 1.0% |
| Pattanaseri 2024 | 0 | 11 | 0 | 9 | 0.83 | [0.01; 45.69] | 0.9% | 1.0% |
| Phillips 2019 | 0 | 22 | 0 | 21 | 0.96 | [0.02; 50.34] | 0.9% | 1.0% |
| Popova 2019 | 0 | 115 | 0 | 109 | 0.95 | [0.02; 48.20] | 0.9% | 1.0% |
| Price 2022 | 0 | 53 | 0 | 51 | 0.96 | [0.02; 49.43] | 0.9% | 1.0% |
| Ray-Griffith 2017 | 0 | 8 | 0 | 8 | 1.00 | [0.02; 56.46] | 0.8% | 1.0% |
| Rodriguez 2013 | 0 | 8 | 0 | 7 | 0.88 | [0.02; 50.19] | 0.9% | 1.0% |
| Schiweck 2021 | 0 | 21 | 0 | 7 | 0.35 | [0.01; 19.18] | 1.2% | 1.0% |
| Shiroma 2020 | 0 | 29 | 0 | 25 | 0.86 | [0.02; 45.14] | 0.9% | 1.0% |
| Singh 2016 2x doses | 1 | 18 | 0 | 16 | 2.83 | [0.11; 74.46] | 0.8% | 1.5% |
| Singh 2016 3x doses | 0 | 17 | 0 | 16 | 0.94 | [0.02; 50.32] | 0.9% | 1.0% |
| Singh 2016 II | 0 | 20 | 0 | 10 | 0.51 | [0.01; 27.68] | 1.1% | 1.0% |
| Smith-Apeldoorn 2024 | 0 | 57 | 0 | 54 | 0.95 | [0.02; 48.61] | 0.9% | 1.0% |
| Sos 2013 | 0 | 11 | 0 | 19 | 1.70 | [0.03; 91.39] | 0.6% | 1.0% |
| Su 2017 | 0 | 47 | 0 | 24 | 0.52 | [0.01; 26.79] | 1.1% | 1.0% |
| Su 2023 | 1 | 42 | 2 | 42 | 0.49 | [0.04; 5.59] | 3.4% | 2.7% |
| Sumner 2019 | 0 | 17 | 0 | 16 | 0.94 | [0.02; 50.32] | 0.9% | 1.0% |
| Takahashi 2021 | 0 | 122 | 0 | 80 | 0.66 | [0.01; 33.45] | 1.0% | 1.0% |
| Taylor 2018 | 0 | 9 | 0 | 9 | 1.00 | [0.02; 55.80] | 0.8% | 1.0% |
| Tiger 2020 | 0 | 20 | 0 | 10 | 0.51 | [0.01; 27.68] | 1.1% | 1.0% |
| Wang 2024 | 0 | 182 | 0 | 182 | 1.00 | [0.02; 50.67] | 0.9% | 1.0% |
| Xiao 2023 | 0 | 15 | 0 | 15 | 1.00 | [0.02; 53.66] | 0.8% | 1.0% |
| Yang 2023 | 0 | 200 | 0 | 100 | 0.50 | [0.01; 25.45] | 1.2% | 1.0% |
| Zarate 2006 | 0 | 9 | 0 | 9 | 1.00 | [0.02; 55.80] | 0.8% | 1.0% |
| Zarate 2012 | 0 | 21 | 0 | 23 | 1.09 | [0.02; 57.53] | 0.8% | 1.0% |
| Zhou 2024 | 0 | 27 | 1 | 27 | 0.32 | [0.01; 8.24] | 2.6% | 1.5% |
| **Common effect model** | | **2742** | | **2201** | **0.80** | **[0.55; 1.18]** | **100.0%** | **.** |
| **Random effects model** | | | | | **0.83** | **[0.55; 1.24]** | **.** | **100.0%** |

Heterogeneity: $I^2 = 0.0\%$, $\tau^2 = 0$, $p = 1.0000$

#### Maentel Mantel-Haenszel with Treatment-Arm CC

| Study | Experimental Events | Experimental Total | Control Events | Control Total | OR | 95%-CI | Weight (common) | Weight (random) |
|---|---|---|---|---|---|---|---|---|
| Abbar 2022 | 6 | 73 | 8 | 83 | 0.84 | [0.28; 2.54] | 13.0% | 13.6% |
| Abdallah 2022 | 0 | 108 | 0 | 55 | 1.00 | [0.02; 63.93] | 0.8% | 1.0% |
| Ahmed 2023 | 0 | 18 | 0 | 18 | 1.00 | [0.02; 53.12] | 0.9% | 1.1% |
| Anderson 2017 | 0 | 40 | 2 | 39 | 0.19 | [0.01; 3.98] | 4.7% | 1.8% |
| Burger 2016 | 0 | 3 | 0 | 7 | 1.00 | [0.01; 87.11] | 0.7% | 0.8% |
| Canuso 2018 | 0 | 35 | 3 | 31 | 0.12 | [0.01; 2.30] | 6.9% | 1.9% |
| Chen 2023 | 1 | 126 | 0 | 126 | 3.02 | [0.12; 74.94] | 0.9% | 1.6% |
| Colla 2024 | 0 | 17 | 0 | 10 | 1.00 | [0.02; 62.21] | 0.9% | 1.0% |
| Dakwar 2019 | 0 | 27 | 0 | 28 | 1.00 | [0.02; 52.22] | 0.9% | 1.1% |
| Dakwar 2020 | 0 | 17 | 0 | 23 | 1.00 | [0.02; 55.30] | 0.9% | 1.0% |
| Daly 2018 PA | 0 | 34 | 0 | 33 | 1.00 | [0.02; 51.89] | 0.9% | 1.1% |
| Daly 2018 PB | 0 | 20 | 0 | 21 | 1.00 | [0.02; 52.85] | 0.9% | 1.1% |
| Domany 2019 | 0 | 22 | 0 | 18 | 1.00 | [0.02; 53.92] | 0.9% | 1.0% |
| Domany 2020 | 0 | 9 | 0 | 9 | 1.00 | [0.02; 55.80] | 0.9% | 1.0% |
| Duek 2023 | 0 | 14 | 0 | 14 | 1.00 | [0.02; 53.89] | 0.9% | 1.1% |
| Dwyer 2021 | 0 | 11 | 0 | 6 | 1.00 | [0.01; 67.61] | 0.8% | 0.9% |
| Fava 2020 | 1 | 80 | 0 | 19 | 2.27 | [0.02; 257.11] | 0.6% | 0.7% |
| Feder 2014 | 0 | 22 | 0 | 19 | 1.00 | [0.02; 53.36] | 0.9% | 1.1% |
| Feder 2021 | 0 | 15 | 0 | 15 | 1.00 | [0.02; 53.66] | 0.9% | 1.1% |
| Fedgchin 2019 | 0 | 231 | 0 | 113 | 1.00 | [0.02; 65.31] | 0.8% | 1.0% |
| Fineberg 2023 | 0 | 10 | 0 | 12 | 1.00 | [0.02; 55.75] | 0.9% | 1.0% |
| Fu 2020 | 4 | 113 | 3 | 112 | 1.33 | [0.29; 6.10] | 5.5% | 7.2% |
| George 2017 | 0 | 12 | 0 | 4 | 1.00 | [0.01; 105.38] | 0.7% | 0.8% |
| Glue 2024b | 0 | 17 | 0 | 8 | 1.00 | [0.01; 72.36] | 0.8% | 0.9% |
| Grabski 2022 | 0 | 48 | 0 | 48 | 1.00 | [0.02; 51.42] | 0.9% | 1.1% |
| Grunebaum 2017 | 0 | 9 | 0 | 7 | 1.00 | [0.02; 58.29] | 0.9% | 1.0% |
| Grunebaum 2018 | 0 | 40 | 0 | 40 | 1.00 | [0.02; 51.63] | 0.9% | 1.1% |
| Ionescu 2020 | 7 | 114 | 4 | 113 | 1.78 | [0.51; 6.27] | 7.1% | 10.6% |
| Jones 2024 | 0 | 17 | 0 | 11 | 1.00 | [0.02; 59.25] | 0.9% | 1.0% |
| Lapidus 2014 | 0 | 10 | 0 | 10 | 1.00 | [0.02; 55.27] | 0.9% | 1.0% |
| Leal 2023 | 0 | 5 | 0 | 5 | 1.00 | [0.02; 59.99] | 0.9% | 1.0% |
| Lii 2023 | 0 | 20 | 0 | 20 | 1.00 | [0.02; 52.85] | 0.9% | 1.1% |
| Lijffijt 2021 | 0 | 20 | 1 | 13 | 0.26 | [0.01; 5.79] | 3.1% | 1.7% |
| Loo 2016 | 0 | 9 | 0 | 6 | 1.00 | [0.02; 61.80] | 0.9% | 1.0% |
| Loo 2023 | 0 | 89 | 2 | 92 | 0.20 | [0.01; 4.27] | 4.6% | 1.8% |
| Milak 2020 | 0 | 38 | 0 | 5 | 1.00 | [0.00; 484.58] | 0.4% | 0.4% |
| Murrough 2013 | 0 | 47 | 0 | 25 | 1.00 | [0.02; 63.11] | 0.8% | 1.0% |
| Murrough 2015 II | 0 | 12 | 0 | 12 | 1.00 | [0.02; 54.46] | 0.9% | 1.0% |
| Ochs-Ross 2020 | 0 | 72 | 0 | 65 | 1.00 | [0.02; 51.38] | 0.9% | 1.1% |
| Ohtani 2024 | 0 | 17 | 0 | 17 | 1.00 | [0.02; 53.28] | 0.9% | 1.1% |
| Pattanaseri 2024 | 0 | 11 | 0 | 9 | 1.00 | [0.02; 56.40] | 0.9% | 1.0% |
| Phillips 2019 | 0 | 22 | 0 | 21 | 1.00 | [0.02; 52.74] | 0.9% | 1.1% |
| Popova 2019 | 0 | 115 | 0 | 109 | 1.00 | [0.02; 50.91] | 0.9% | 1.1% |
| Price 2022 | 0 | 53 | 0 | 51 | 1.00 | [0.02; 51.38] | 0.9% | 1.1% |
| Ray-Griffith 2017 | 0 | 8 | 0 | 8 | 1.00 | [0.02; 56.46] | 0.9% | 1.0% |
| Rodriguez 2013 | 0 | 8 | 0 | 7 | 1.00 | [0.02; 57.37] | 0.9% | 1.0% |
| Schiweck 2021 | 0 | 21 | 0 | 7 | 1.00 | [0.01; 99.86] | 0.7% | 0.8% |
| Shiroma 2020 | 0 | 29 | 0 | 25 | 1.00 | [0.02; 52.79] | 0.9% | 1.1% |
| Singh 2016 2x doses | 1 | 18 | 0 | 16 | 3.05 | [0.11; 85.81] | 0.9% | 1.5% |
| Singh 2016 3x doses | 0 | 17 | 0 | 16 | 1.00 | [0.02; 53.46] | 0.9% | 1.1% |
| Singh 2016 II | 0 | 20 | 0 | 10 | 1.00 | [0.01; 68.32] | 0.8% | 0.9% |
| Smith-Apeldoorn 2024 | 0 | 57 | 0 | 54 | 1.00 | [0.02; 51.36] | 0.9% | 1.1% |
| Sos 2013 | 0 | 11 | 0 | 19 | 1.00 | [0.02; 62.32] | 0.9% | 1.0% |
| Su 2017 | 0 | 47 | 0 | 24 | 1.00 | [0.02; 64.85] | 0.8% | 1.0% |
| Su 2023 | 1 | 42 | 2 | 42 | 0.49 | [0.04; 5.59] | 3.7% | 2.8% |
| Sumner 2019 | 0 | 17 | 0 | 16 | 1.00 | [0.02; 53.46] | 0.9% | 1.1% |
| Takahashi 2021 | 0 | 122 | 0 | 80 | 1.00 | [0.02; 55.55] | 0.9% | 1.0% |
| Taylor 2018 | 0 | 9 | 0 | 9 | 1.00 | [0.02; 55.80] | 0.9% | 1.0% |
| Tiger 2020 | 0 | 20 | 0 | 10 | 1.00 | [0.01; 68.32] | 0.8% | 0.9% |
| Wang 2024 | 0 | 182 | 0 | 182 | 1.00 | [0.02; 50.67] | 0.9% | 1.1% |
| Xiao 2023 | 0 | 15 | 0 | 15 | 1.00 | [0.02; 53.66] | 0.9% | 1.1% |
| Yang 2023 | 0 | 200 | 0 | 100 | 1.00 | [0.02; 64.37] | 0.8% | 1.0% |
| Zarate 2006 | 0 | 9 | 0 | 9 | 1.00 | [0.02; 55.80] | 0.9% | 1.0% |
| Zarate 2012 | 0 | 21 | 0 | 23 | 1.00 | [0.02; 52.84] | 0.9% | 1.1% |
| Zhou 2024 | 0 | 27 | 1 | 27 | 0.32 | [0.01; 8.24] | 2.8% | 1.6% |
| **Common effect model** | | **2742** | | **2201** | **0.90** | **[0.61; 1.33]** | **100.0%** | **.** |
| **Random effects model** | | | | | **0.94** | **[0.62; 1.41]** | **.** | **100.0%** |

Heterogeneity: $I^2 = 0.0\%$, $\tau^2 = 0$, $p = 1.0000$

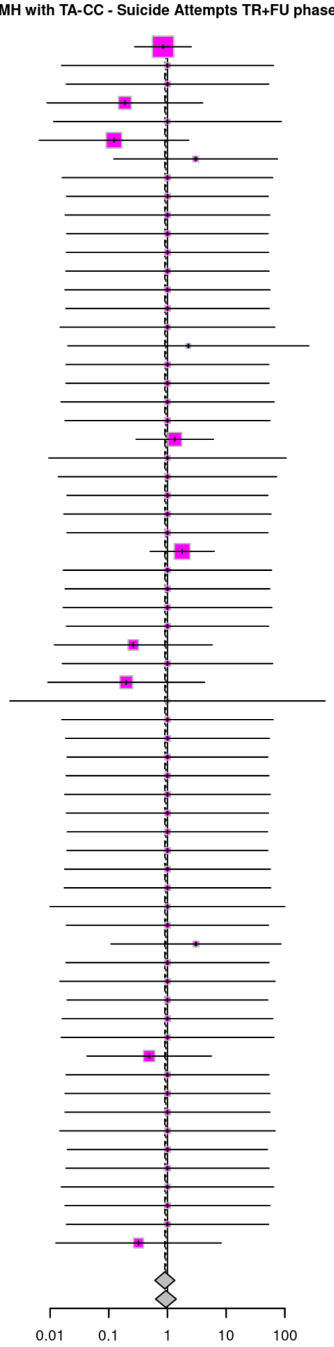

#### Maentel Mantel-Haenszel Risk-Difference

| Study | Experimental Events | Experimental Total | Control Events | Control Total | RD | 95%-CI | Weight (common) | Weight (random) |
|---|---|---|---|---|---|---|---|---|
| Abbar 2022 | 6 | 73 | 8 | 83 | -0.01 | [-0.10; 0.08] | 3.2% | 0.3% |
| Abdallah 2022 | 0 | 108 | 0 | 55 | 0.00 | [-0.03; 0.03] | 3.0% | 3.5% |
| Ahmed 2023 | 0 | 18 | 0 | 18 | 0.00 | [-0.10; 0.10] | 0.8% | 0.3% |
| Anderson 2017 | 0 | 40 | 2 | 39 | -0.05 | [-0.13; 0.03] | 1.7% | 0.4% |
| Burger 2016 | 0 | 3 | 0 | 7 | 0.00 | [-0.36; 0.36] | 0.2% | 0.0% |
| Canuso 2018 | 0 | 35 | 3 | 31 | -0.10 | [-0.21; 0.02] | 1.4% | 0.2% |
| Chen 2023 | 1 | 126 | 0 | 126 | 0.01 | [-0.01; 0.03] | 5.2% | 5.7% |
| Colla 2024 | 0 | 17 | 0 | 10 | 0.00 | [-0.14; 0.14] | 0.6% | 0.1% |
| Dakwar 2019 | 0 | 27 | 0 | 28 | 0.00 | [-0.07; 0.07] | 1.2% | 0.6% |
| Dakwar 2020 | 0 | 17 | 0 | 23 | 0.00 | [-0.10; 0.10] | 0.8% | 0.3% |
| Daly 2018 PA | 0 | 34 | 0 | 33 | 0.00 | [-0.06; 0.06] | 1.4% | 0.8% |
| Daly 2018 PB | 0 | 20 | 0 | 21 | 0.00 | [-0.09; 0.09] | 0.9% | 0.3% |
| Domany 2019 | 0 | 22 | 0 | 18 | 0.00 | [-0.09; 0.09] | 0.9% | 0.3% |
| Domany 2020 | 0 | 9 | 0 | 9 | 0.00 | [-0.19; 0.19] | 0.4% | 0.1% |
| Duek 2023 | 0 | 14 | 0 | 14 | 0.00 | [-0.13; 0.13] | 0.6% | 0.2% |
| Dwyer 2021 | 0 | 11 | 0 | 6 | 0.00 | [-0.22; 0.22] | 0.4% | 0.1% |
| Fava 2020 | 1 | 80 | 0 | 19 | 0.01 | [-0.06; 0.09] | 1.3% | 0.5% |
| Feder 2014 | 0 | 22 | 0 | 19 | 0.00 | [-0.09; 0.09] | 0.9% | 0.3% |
| Feder 2021 | 0 | 15 | 0 | 15 | 0.00 | [-0.12; 0.12] | 0.7% | 0.2% |
| Fedgchin 2019 | 0 | 231 | 0 | 113 | 0.00 | [-0.01; 0.01] | 6.3% | 14.6% |
| Fineberg 2023 | 0 | 10 | 0 | 12 | 0.00 | [-0.16; 0.16] | 0.5% | 0.1% |
| Fu 2020 | 4 | 113 | 3 | 112 | 0.01 | [-0.04; 0.05] | 4.6% | 1.3% |
| George 2017 | 0 | 12 | 0 | 4 | 0.00 | [-0.28; 0.28] | 0.3% | 0.0% |
| Glue 2024b | 0 | 17 | 0 | 8 | 0.00 | [-0.17; 0.17] | 0.5% | 0.1% |
| Grabski 2022 | 0 | 48 | 0 | 48 | 0.00 | [-0.04; 0.04] | 2.0% | 1.7% |
| Grunebaum 2017 | 0 | 9 | 0 | 7 | 0.00 | [-0.22; 0.22] | 0.4% | 0.1% |
| Grunebaum 2018 | 0 | 40 | 0 | 40 | 0.00 | [-0.05; 0.05] | 1.7% | 1.2% |
| Ionescu 2020 | 7 | 114 | 4 | 113 | 0.03 | [-0.03; 0.08] | 4.7% | 0.9% |
| Jones 2024 | 0 | 17 | 0 | 11 | 0.00 | [-0.14; 0.14] | 0.6% | 0.1% |
| Lapidus 2014 | 0 | 10 | 0 | 10 | 0.00 | [-0.17; 0.17] | 0.5% | 0.1% |
| Leal 2023 | 0 | 5 | 0 | 5 | 0.00 | [-0.31; 0.31] | 0.2% | 0.0% |
| Lii 2023 | 0 | 20 | 0 | 20 | 0.00 | [-0.09; 0.09] | 0.9% | 0.3% |
| Lijffijt 2021 | 0 | 20 | 1 | 13 | -0.08 | [-0.25; 0.10] | 0.7% | 0.1% |
| Loo 2016 | 0 | 9 | 0 | 6 | 0.00 | [-0.23; 0.23] | 0.3% | 0.0% |
| Loo 2023 | 0 | 89 | 2 | 92 | -0.02 | [-0.06; 0.01] | 3.8% | 2.0% |
| Milak 2020 | 0 | 38 | 0 | 5 | 0.00 | [-0.22; 0.22] | 0.4% | 0.1% |
| Murrough 2013 | 0 | 47 | 0 | 25 | 0.00 | [-0.06; 0.06] | 1.4% | 0.7% |
| Murrough 2015 II | 0 | 12 | 0 | 12 | 0.00 | [-0.15; 0.15] | 0.5% | 0.1% |
| Ochs-Ross 2020 | 0 | 72 | 0 | 65 | 0.00 | [-0.03; 0.03] | 2.9% | 3.4% |
| Ohtani 2024 | 0 | 17 | 0 | 17 | 0.00 | [-0.11; 0.11] | 0.7% | 0.2% |
| Pattanaseri 2024 | 0 | 11 | 0 | 9 | 0.00 | [-0.18; 0.18] | 0.4% | 0.1% |
| Phillips 2019 | 0 | 22 | 0 | 21 | 0.00 | [-0.09; 0.09] | 0.9% | 0.4% |
| Popova 2019 | 0 | 115 | 0 | 109 | 0.00 | [-0.02; 0.02] | 4.6% | 8.9% |
| Price 2022 | 0 | 53 | 0 | 51 | 0.00 | [-0.04; 0.04] | 2.2% | 2.0% |
| Ray-Griffith 2017 | 0 | 8 | 0 | 8 | 0.00 | [-0.21; 0.21] | 0.4% | 0.1% |
| Rodriguez 2013 | 0 | 8 | 0 | 7 | 0.00 | [-0.22; 0.22] | 0.3% | 0.1% |
| Schiweck 2021 | 0 | 21 | 0 | 7 | 0.00 | [-0.18; 0.18] | 0.5% | 0.1% |
| Shiroma 2020 | 0 | 29 | 0 | 25 | 0.00 | [-0.07; 0.07] | 1.1% | 0.5% |
| Singh 2016 2x doses | 1 | 18 | 0 | 16 | 0.06 | [-0.09; 0.20] | 0.7% | 0.1% |
| Singh 2016 3x doses | 0 | 17 | 0 | 16 | 0.00 | [-0.11; 0.11] | 0.7% | 0.2% |
| Singh 2016 II | 0 | 20 | 0 | 10 | 0.00 | [-0.14; 0.14] | 0.6% | 0.1% |
| Smith-Apeldoorn 2024 | 0 | 57 | 0 | 54 | 0.00 | [-0.03; 0.03] | 2.3% | 2.2% |
| Sos 2013 | 0 | 11 | 0 | 19 | 0.00 | [-0.13; 0.13] | 0.6% | 0.2% |
| Su 2017 | 0 | 47 | 0 | 24 | 0.00 | [-0.06; 0.06] | 1.4% | 0.7% |
| Su 2023 | 1 | 42 | 2 | 42 | -0.02 | [-0.10; 0.06] | 1.7% | 0.4% |
| Sumner 2019 | 0 | 17 | 0 | 16 | 0.00 | [-0.11; 0.11] | 0.7% | 0.2% |
| Takahashi 2021 | 0 | 122 | 0 | 80 | 0.00 | [-0.02; 0.02] | 4.0% | 6.4% |
| Taylor 2018 | 0 | 9 | 0 | 9 | 0.00 | [-0.19; 0.19] | 0.4% | 0.1% |
| Tiger 2020 | 0 | 20 | 0 | 10 | 0.00 | [-0.14; 0.14] | 0.6% | 0.1% |
| Wang 2024 | 0 | 182 | 0 | 182 | 0.00 | [-0.01; 0.01] | 7.5% | 23.4% |
| Xiao 2023 | 0 | 15 | 0 | 15 | 0.00 | [-0.12; 0.12] | 0.7% | 0.2% |
| Yang 2023 | 0 | 200 | 0 | 100 | 0.00 | [-0.02; 0.02] | 5.5% | 11.4% |
| Zarate 2006 | 0 | 9 | 0 | 9 | 0.00 | [-0.19; 0.19] | 0.4% | 0.1% |
| Zarate 2012 | 0 | 21 | 0 | 23 | 0.00 | [-0.08; 0.08] | 0.9% | 0.4% |
| Zhou 2024 | 0 | 27 | 1 | 27 | -0.04 | [-0.13; 0.06] | 1.2% | 0.3% |
| **Common effect model** | | **2742** | | **2201** | **-0.00** | **[-0.01; 0.01]** | **100.0%** | **.** |
| **Random effects model** | | | | | **-0.00** | **[-0.01; 0.00]** | **.** | **100.0%** |

Heterogeneity: $I^2 = 0.0\%$, $\tau^2 = 0$, $p = 1.0000$

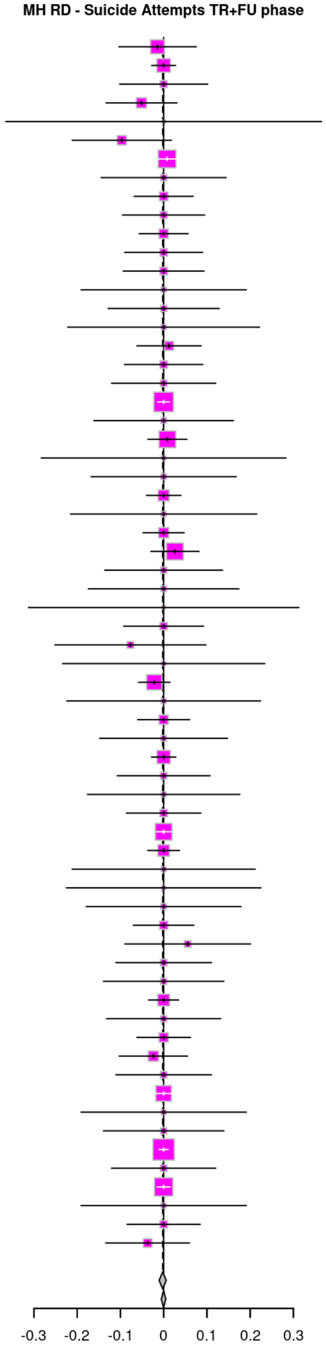

#### Arcsine Risk-Difference

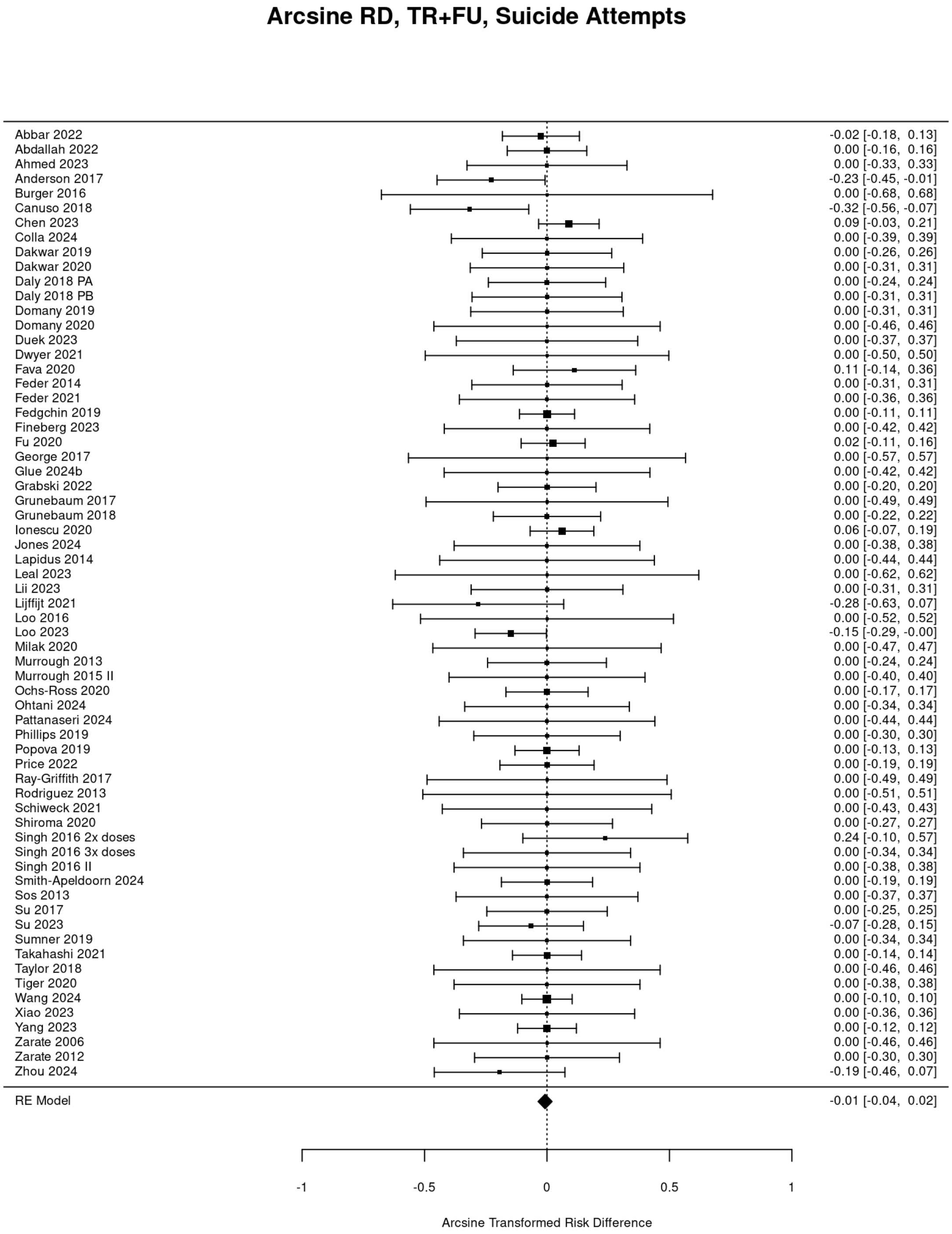

Bayesian Meta-Analysis, wider prior

**Bayesian MA, Delta = 250 - Suicide Attempts TR-FU**

| Study | Estimate | 95% CI |
|---|---|---|
| Abbar 2022 | -0.17 | [-1.28, 0.93] |
| Abdallah 2022 | -0.67 | [-4.60, 3.26] |
| Ahmed 2023 | 0.00 | [-3.97, 3.97] |
| Anderson 2017 | -1.69 | [-4.76, 1.38] |
| Burger 2016 | 0.76 | [-3.36, 4.88] |
| Canuso 2018 | -2.17 | [-5.17, 0.84] |
| Chen 2023 | 1.11 | [-2.10, 4.32] |
| Colla 2024 | -0.51 | [-4.50, 3.48] |
| Dakwar 2019 | 0.04 | [-3.92, 3.99] |
| Dakwar 2020 | 0.29 | [-3.67, 4.26] |
| Daly 2018 PA | -0.03 | [-3.98, 3.92] |
| Daly 2018 PB | 0.05 | [-3.92, 4.01] |
| Domany 2019 | -0.20 | [-4.16, 3.77] |
| Domany 2020 | 0.00 | [-4.02, 4.02] |
| Duek 2023 | 0.00 | [-3.99, 3.99] |
| Dwyer 2021 | -0.57 | [-4.61, 3.47] |
| Fava 2020 | -0.31 | [-3.55, 2.93] |
| Feder 2014 | -0.14 | [-4.11, 3.82] |
| Feder 2021 | 0.00 | [-3.98, 3.98] |
| Fedgchin 2019 | -0.71 | [-4.64, 3.21] |
| Fineberg 2023 | 0.17 | [-3.83, 4.18] |
| Fu 2020 | 0.29 | [-1.23, 1.81] |
| George 2017 | -1.02 | [-5.09, 3.04] |
| Glue 2024b | -0.72 | [-4.73, 3.28] |
| Grabski 2022 | 0.00 | [-3.94, 3.94] |
| Grunebaum 2017 | -0.24 | [-4.27, 3.80] |
| Grunebaum 2018 | 0.00 | [-3.94, 3.94] |
| Ionescu 2020 | 0.58 | [-0.68, 1.84] |
| Jones 2024 | -0.42 | [-4.41, 3.57] |
| Lapidus 2014 | 0.00 | [-4.01, 4.01] |
| Leal 2023 | 0.00 | [-4.09, 4.09] |
| Lii 2023 | 0.00 | [-3.97, 3.97] |
| Lijffijt 2021 | -1.59 | [-4.87, 1.68] |
| Loo 2016 | -0.38 | [-4.42, 3.67] |
| Loo 2023 | -1.60 | [-4.65, 1.45] |
| Milak 2020 | -1.95 | [-5.97, 2.07] |
| Murrough 2013 | -0.62 | [-4.57, 3.33] |
| Murrough 2015 II | 0.00 | [-4.00, 4.00] |
| Ochs-Ross 2020 | -0.10 | [-4.04, 3.83] |
| Ohtani 2024 | 0.00 | [-3.98, 3.98] |
| Pattanaseri 2024 | -0.19 | [-4.20, 3.82] |
| Phillips 2019 | -0.05 | [-4.01, 3.92] |
| Popova 2019 | -0.05 | [-3.98, 3.88] |
| Price 2022 | -0.04 | [-3.98, 3.90] |
| Ray-Griffith 2017 | 0.00 | [-4.03, 4.03] |
| Rodriguez 2013 | -0.13 | [-4.17, 3.92] |
| Schiweck 2021 | -1.05 | [-5.06, 2.95] |
| Shiroma 2020 | -0.15 | [-4.10, 3.81] |
| Singh 2016 2x doses | 1.04 | [-2.23, 4.31] |
| Singh 2016 3x doses | -0.06 | [-4.04, 3.92] |
| Singh 2016 II | -0.67 | [-4.66, 3.32] |
| Smith-Apeldoorn 2024 | -0.05 | [-3.99, 3.88] |
| Sos 2013 | 0.53 | [-3.46, 4.52] |
| Su 2017 | -0.66 | [-4.61, 3.29] |
| Su 2023 | -0.72 | [-3.16, 1.72] |
| Sumner 2019 | -0.06 | [-4.04, 3.92] |
| Takahashi 2021 | -0.42 | [-4.35, 3.51] |
| Taylor 2018 | 0.00 | [-4.02, 4.02] |
| Tiger 2020 | -0.67 | [-4.66, 3.32] |
| Wang 2024 | 0.00 | [-3.93, 3.93] |
| Xiao 2023 | 0.00 | [-3.98, 3.98] |
| Yang 2023 | -0.69 | [-4.62, 3.24] |
| Zarate 2006 | 0.00 | [-4.02, 4.02] |
| Zarate 2012 | 0.09 | [-3.87, 4.05] |
| Zhou 2024 | -1.14 | [-4.38, 2.11] |
| **Summary** | **-0.29** | **[-0.95, 0.32]** |
| **Prediction** | **-0.28** | **[-1.44, 0.74]** |

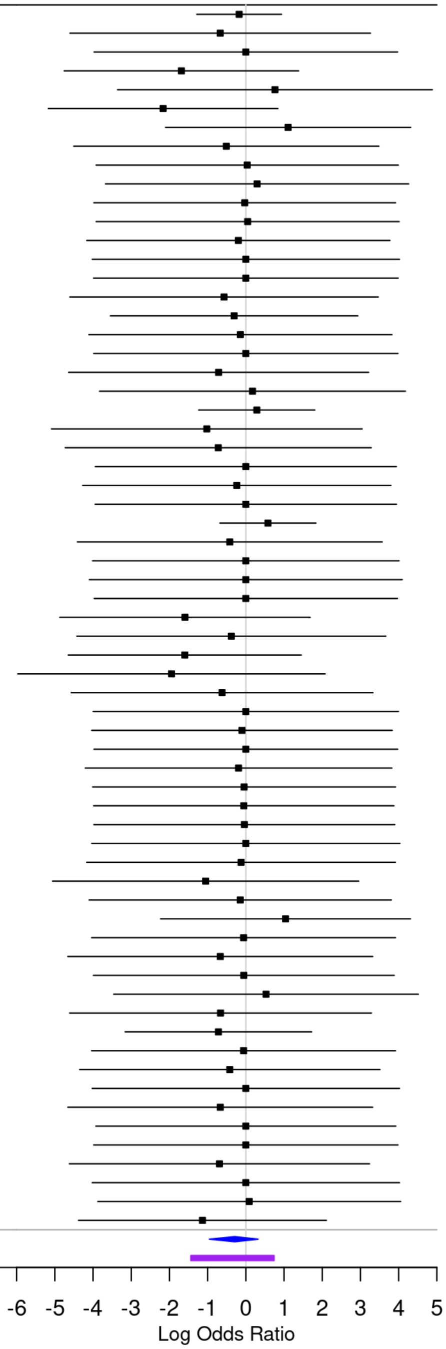

*Heterogeneity (tau): 0.29 [0.00, 0.83]*

Bayesian Meta-Analysis, smaller prior

**Bayesian MA, Delta = 15 - Suicide Attempts TR+FU**

| Study | Estimate | 95% CI |
|---|---|---|
| Abbar 2022 | -0.17 | [-1.28, 0.93] |
| Abdallah 2022 | -0.67 | [-4.60, 3.26] |
| Ahmed 2023 | 0.00 | [-3.97, 3.97] |
| Anderson 2017 | -1.69 | [-4.76, 1.38] |
| Burger 2016 | 0.76 | [-3.36, 4.88] |
| Canuso 2018 | -2.17 | [-5.17, 0.84] |
| Chen 2023 | 1.11 | [-2.10, 4.32] |
| Colla 2024 | -0.51 | [-4.50, 3.48] |
| Dakwar 2019 | 0.04 | [-3.92, 3.99] |
| Dakwar 2020 | 0.29 | [-3.67, 4.26] |
| Daly 2018 PA | -0.03 | [-3.98, 3.92] |
| Daly 2018 PB | 0.05 | [-3.92, 4.01] |
| Domany 2019 | -0.20 | [-4.16, 3.77] |
| Domany 2020 | 0.00 | [-4.02, 4.02] |
| Duek 2023 | 0.00 | [-3.99, 3.99] |
| Dwyer 2021 | -0.57 | [-4.61, 3.47] |
| Fava 2020 | -0.31 | [-3.55, 2.93] |
| Feder 2014 | -0.14 | [-4.11, 3.82] |
| Feder 2021 | 0.00 | [-3.98, 3.98] |
| Fedgchin 2019 | -0.71 | [-4.64, 3.21] |
| Fineberg 2023 | 0.17 | [-3.83, 4.18] |
| Fu 2020 | 0.29 | [-1.23, 1.81] |
| George 2017 | -1.02 | [-5.09, 3.04] |
| Glue 2024b | -0.72 | [-4.73, 3.28] |
| Grabski 2022 | 0.00 | [-3.94, 3.94] |
| Grunebaum 2017 | -0.24 | [-4.27, 3.80] |
| Grunebaum 2018 | 0.00 | [-3.94, 3.94] |
| Ionescu 2020 | 0.58 | [-0.68, 1.84] |
| Jones 2024 | -0.42 | [-4.41, 3.57] |
| Lapidus 2014 | 0.00 | [-4.01, 4.01] |
| Leal 2023 | 0.00 | [-4.09, 4.09] |
| Lii 2023 | 0.00 | [-3.97, 3.97] |
| Lijffijt 2021 | -1.59 | [-4.87, 1.68] |
| Loo 2016 | -0.38 | [-4.42, 3.67] |
| Loo 2023 | -1.60 | [-4.65, 1.45] |
| Milak 2020 | -1.95 | [-5.97, 2.07] |
| Murrough 2013 | -0.62 | [-4.57, 3.33] |
| Murrough 2015 II | 0.00 | [-4.00, 4.00] |
| Ochs-Ross 2020 | -0.10 | [-4.04, 3.83] |
| Ohtani 2024 | 0.00 | [-3.98, 3.98] |
| Pattanaseri 2024 | -0.19 | [-4.20, 3.82] |
| Phillips 2019 | -0.05 | [-4.01, 3.92] |
| Popova 2019 | -0.05 | [-3.98, 3.88] |
| Price 2022 | -0.04 | [-3.98, 3.90] |
| Ray-Griffith 2017 | 0.00 | [-4.03, 4.03] |
| Rodriguez 2013 | -0.13 | [-4.17, 3.92] |
| Schiweck 2021 | -1.05 | [-5.06, 2.95] |
| Shiroma 2020 | -0.15 | [-4.10, 3.81] |
| Singh 2016 2x doses | 1.04 | [-2.23, 4.31] |
| Singh 2016 3x doses | -0.06 | [-4.04, 3.92] |
| Singh 2016 II | -0.67 | [-4.66, 3.32] |
| Smith-Apeldoorn 2024 | -0.05 | [-3.99, 3.88] |
| Sos 2013 | 0.53 | [-3.46, 4.52] |
| Su 2017 | -0.66 | [-4.61, 3.29] |
| Su 2023 | -0.72 | [-3.16, 1.72] |
| Sumner 2019 | -0.06 | [-4.04, 3.92] |
| Takahashi 2021 | -0.42 | [-4.35, 3.51] |
| Taylor 2018 | 0.00 | [-4.02, 4.02] |
| Tiger 2020 | -0.67 | [-4.66, 3.32] |
| Wang 2024 | 0.00 | [-3.93, 3.93] |
| Xiao 2023 | 0.00 | [-3.98, 3.98] |
| Yang 2023 | -0.69 | [-4.62, 3.24] |
| Zarate 2006 | 0.00 | [-4.02, 4.02] |
| Zarate 2012 | 0.09 | [-3.87, 4.05] |
| Zhou 2024 | -1.14 | [-4.38, 2.11] |
| **Summary** | **-0.28** | **[-0.93, 0.33]** |
| **Prediction** | **-0.26** | **[-1.41, 0.76]** |

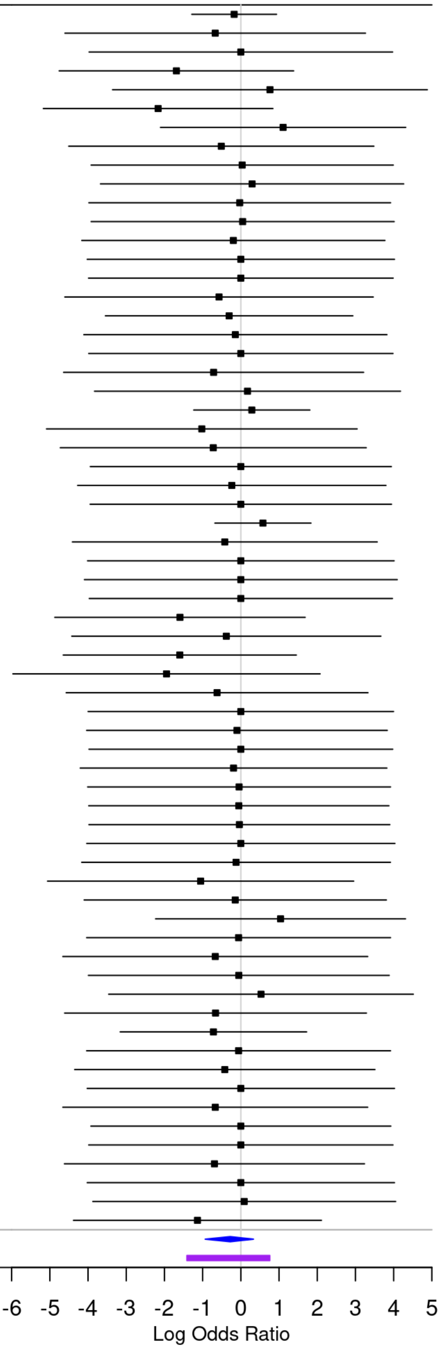

*Heterogeneity (tau): 0.29 [0.00, 0.82]*

##### Exact Method

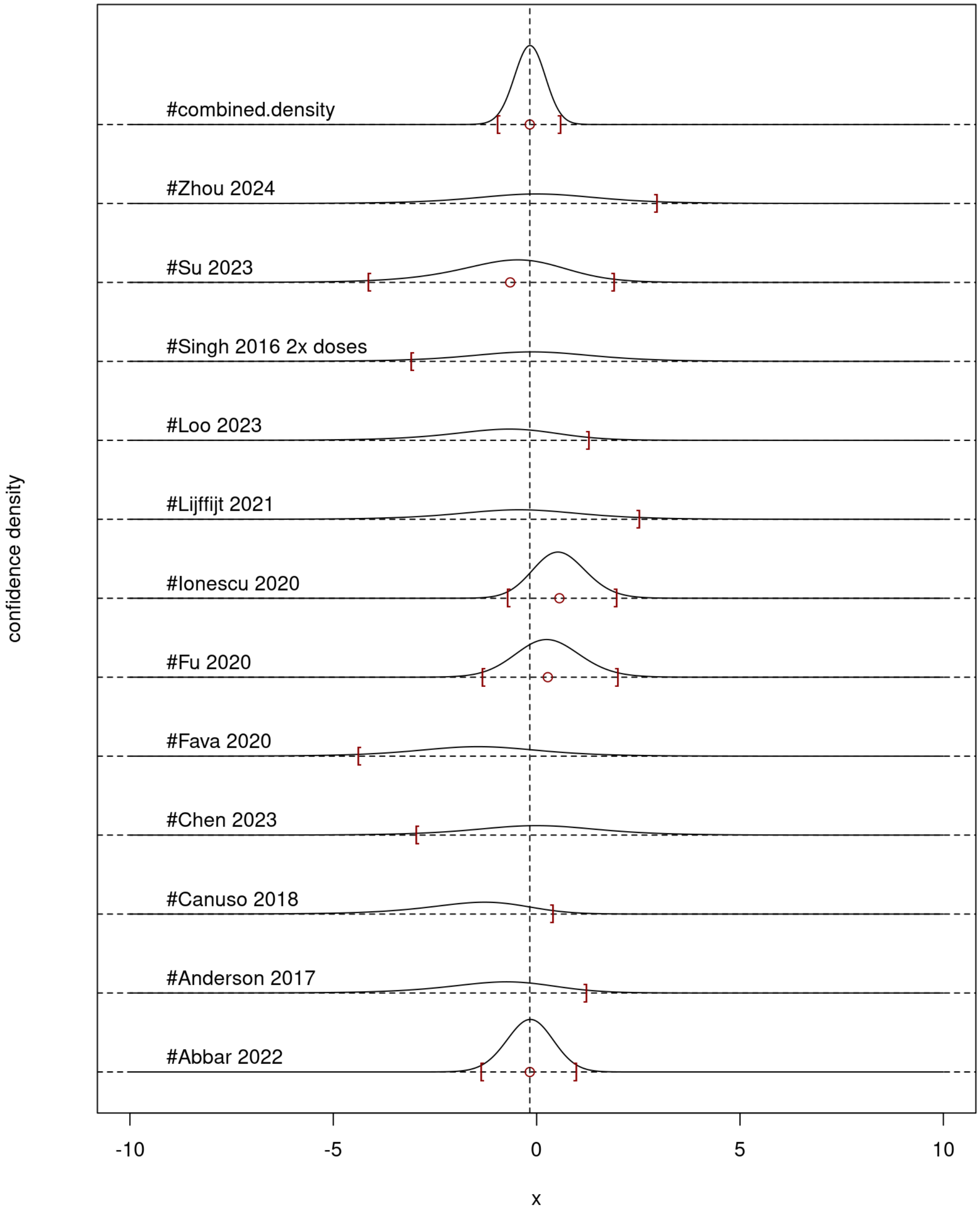

### Suicides

#### Treatment Phase

#### Peto's Method

| Study | Experimental Events | Experimental Total | Control Events | Control Total | OR | 95%-CI | Weight (common) | Weight (random) |
|---|---|---|---|---|---|---|---|---|
| Abbar 2022 | 0 | 73 | 0 | 73 | | | 0.0% | 0.0% |
| Abdallah 2022 | 0 | 108 | 0 | 108 | | | 0.0% | 0.0% |
| Ahmed 2023 | 0 | 18 | 0 | 18 | | | 0.0% | 0.0% |
| Anderson 2017 | 0 | 40 | 0 | 40 | | | 0.0% | 0.0% |
| Burger 2016 | 0 | 3 | 0 | 3 | | | 0.0% | 0.0% |
| Canuso 2018 | 0 | 35 | 0 | 35 | | | 0.0% | 0.0% |
| Chen 2023 | 1 | 126 | 0 | 126 | 7.39 | [0.15; 372.38] | 33.5% | 33.5% |
| Colla 2024 | 0 | 17 | 0 | 17 | | | 0.0% | 0.0% |
| Dakwar 2019 | 0 | 27 | 0 | 27 | | | 0.0% | 0.0% |
| Dakwar 2020 | 0 | 17 | 0 | 17 | | | 0.0% | 0.0% |
| Daly 2018 PA | 0 | 34 | 0 | 34 | | | 0.0% | 0.0% |
| Daly 2018 PB | 0 | 20 | 0 | 20 | | | 0.0% | 0.0% |
| Domany 2019 | 0 | 22 | 0 | 22 | | | 0.0% | 0.0% |
| Domany 2020 | 0 | 9 | 0 | 9 | | | 0.0% | 0.0% |
| Duek 2023 | 0 | 14 | 0 | 14 | | | 0.0% | 0.0% |
| Dwyer 2021 | 0 | 11 | 0 | 11 | | | 0.0% | 0.0% |
| Fava 2020 | 0 | 80 | 0 | 80 | | | 0.0% | 0.0% |
| Feder 2014 | 0 | 22 | 0 | 22 | | | 0.0% | 0.0% |
| Feder 2021 | 0 | 15 | 0 | 15 | | | 0.0% | 0.0% |
| Fedgchin 2019 | 0 | 231 | 0 | 231 | | | 0.0% | 0.0% |
| Fineberg 2023 | 0 | 10 | 0 | 10 | | | 0.0% | 0.0% |
| Fu 2020 | 0 | 113 | 0 | 113 | | | 0.0% | 0.0% |
| George 2017 | 0 | 12 | 0 | 12 | | | 0.0% | 0.0% |
| Glue 2024b | 0 | 17 | 0 | 17 | | | 0.0% | 0.0% |
| Grabski 2022 | 0 | 48 | 0 | 48 | | | 0.0% | 0.0% |
| Grunebaum 2017 | 0 | 9 | 0 | 9 | | | 0.0% | 0.0% |
| Grunebaum 2018 | 0 | 40 | 0 | 40 | | | 0.0% | 0.0% |
| Ionescu 2020 | 0 | 114 | 0 | 114 | | | 0.0% | 0.0% |
| Jones 2024 | 0 | 17 | 0 | 17 | | | 0.0% | 0.0% |
| Lapidus 2014 | 0 | 10 | 0 | 10 | | | 0.0% | 0.0% |
| Leal 2023 | 0 | 5 | 0 | 5 | | | 0.0% | 0.0% |
| Lii 2023 | 0 | 20 | 0 | 20 | | | 0.0% | 0.0% |
| Lijffijt 2021 | 0 | 20 | 0 | 20 | | | 0.0% | 0.0% |
| Loo 2016 | 0 | 9 | 0 | 9 | | | 0.0% | 0.0% |
| Loo 2023 | 0 | 89 | 0 | 89 | | | 0.0% | 0.0% |
| Milak 2020 | 0 | 38 | 0 | 38 | | | 0.0% | 0.0% |
| Murrough 2013 | 0 | 47 | 0 | 47 | | | 0.0% | 0.0% |
| Murrough 2015 II | 0 | 12 | 0 | 12 | | | 0.0% | 0.0% |
| Ochs-Ross 2020 | 0 | 72 | 0 | 72 | | | 0.0% | 0.0% |
| Ohtani 2024 | 0 | 17 | 0 | 17 | | | 0.0% | 0.0% |
| Pattanaseri 2024 | 0 | 11 | 0 | 11 | | | 0.0% | 0.0% |
| Phillips 2019 | 0 | 22 | 0 | 22 | | | 0.0% | 0.0% |
| Popova 2019 | 0 | 115 | 0 | 115 | | | 0.0% | 0.0% |
| Price 2022 | 0 | 103 | 0 | 103 | | | 0.0% | 0.0% |
| Ray-Griffith 2017 | 0 | 8 | 0 | 8 | | | 0.0% | 0.0% |
| Rodriguez 2013 | 0 | 8 | 0 | 8 | | | 0.0% | 0.0% |
| Schiweck 2021 | 0 | 21 | 0 | 21 | | | 0.0% | 0.0% |
| Shiroma 2020 | 0 | 29 | 0 | 29 | | | 0.0% | 0.0% |
| Singh 2016 2x doses | 0 | 18 | 0 | 18 | | | 0.0% | 0.0% |
| Singh 2016 3x doses | 0 | 17 | 0 | 17 | | | 0.0% | 0.0% |
| Singh 2016 II | 0 | 20 | 0 | 20 | | | 0.0% | 0.0% |
| Smith-Apeldoorn 2024 | 1 | 57 | 1 | 57 | 1.00 | [0.06; 16.19] | 66.5% | 66.5% |
| Sos 2013 | 0 | 11 | 0 | 11 | | | 0.0% | 0.0% |
| Su 2017 | 0 | 47 | 0 | 47 | | | 0.0% | 0.0% |
| Su 2023 | 0 | 42 | 0 | 42 | | | 0.0% | 0.0% |
| Sumner 2019 | 0 | 17 | 0 | 17 | | | 0.0% | 0.0% |
| Takahashi 2021 | 0 | 122 | 0 | 122 | | | 0.0% | 0.0% |
| Taylor 2018 | 0 | 9 | 0 | 9 | | | 0.0% | 0.0% |
| Tiger 2020 | 0 | 20 | 0 | 20 | | | 0.0% | 0.0% |
| Wang 2024 | 0 | 182 | 0 | 182 | | | 0.0% | 0.0% |
| Xiao 2023 | 0 | 15 | 0 | 15 | | | 0.0% | 0.0% |
| Yang 2023 | 0 | 200 | 0 | 200 | | | 0.0% | 0.0% |
| Zarate 2006 | 0 | 9 | 0 | 9 | | | 0.0% | 0.0% |
| Zarate 2012 | 0 | 21 | 0 | 21 | | | 0.0% | 0.0% |
| Zhou 2024 | 0 | 27 | 0 | 27 | | | 0.0% | 0.0% |
| **Common effect model** | | **2792** | | **2792** | **1.96** | **[0.20; 18.93]** | **100.0%** | **.** |
| **Random effects model** | | | | | **1.96** | **[0.20; 18.93]** | **.** | **100.0%** |

Heterogeneity: $I^2 = 0.0\%$, $\tau^2 = 0$, $p = 0.4149$

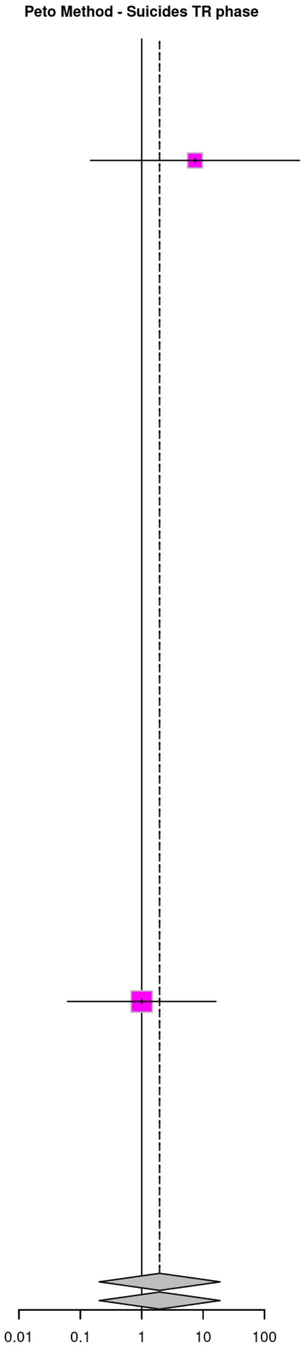

### Maentel Mantel-Haenszel with CC

| Study | Experimental Events | Experimental Total | Control Events | Control Total | OR | 95%-CI | Weight (common) | Weight (random) |
|---|---|---|---|---|---|---|---|---|
| Abbar 2022 | 0 | 73 | 0 | 73 | 1.00 | [0.02; 51.07] | 1.5% | 1.5% |
| Abdallah 2022 | 0 | 108 | 0 | 108 | 1.00 | [0.02; 50.85] | 1.5% | 1.5% |
| Ahmed 2023 | 0 | 18 | 0 | 18 | 1.00 | [0.02; 53.12] | 1.5% | 1.5% |
| Anderson 2017 | 0 | 40 | 0 | 40 | 1.00 | [0.02; 51.63] | 1.5% | 1.5% |
| Burger 2016 | 0 | 3 | 0 | 3 | 1.00 | [0.02; 66.06] | 1.4% | 1.4% |
| Canuso 2018 | 0 | 35 | 0 | 35 | 1.00 | [0.02; 51.80] | 1.5% | 1.5% |
| Chen 2023 | 1 | 126 | 0 | 126 | 3.02 | [0.12; 74.94] | 1.5% | 2.3% |
| Colla 2024 | 0 | 17 | 0 | 17 | 1.00 | [0.02; 53.28] | 1.5% | 1.5% |
| Dakwar 2019 | 0 | 27 | 0 | 27 | 1.00 | [0.02; 52.22] | 1.5% | 1.5% |
| Dakwar 2020 | 0 | 17 | 0 | 17 | 1.00 | [0.02; 53.28] | 1.5% | 1.5% |
| Daly 2018 PA | 0 | 34 | 0 | 34 | 1.00 | [0.02; 51.84] | 1.5% | 1.5% |
| Daly 2018 PB | 0 | 20 | 0 | 20 | 1.00 | [0.02; 52.85] | 1.5% | 1.5% |
| Domany 2019 | 0 | 22 | 0 | 22 | 1.00 | [0.02; 52.63] | 1.5% | 1.5% |
| Domany 2020 | 0 | 9 | 0 | 9 | 1.00 | [0.02; 55.80] | 1.5% | 1.5% |
| Duek 2023 | 0 | 14 | 0 | 14 | 1.00 | [0.02; 53.89] | 1.5% | 1.5% |
| Dwyer 2021 | 0 | 11 | 0 | 11 | 1.00 | [0.02; 54.83] | 1.5% | 1.5% |
| Fava 2020 | 0 | 80 | 0 | 80 | 1.00 | [0.02; 51.01] | 1.5% | 1.5% |
| Feder 2014 | 0 | 22 | 0 | 22 | 1.00 | [0.02; 52.63] | 1.5% | 1.5% |
| Feder 2021 | 0 | 15 | 0 | 15 | 1.00 | [0.02; 53.66] | 1.5% | 1.5% |
| Fedgchin 2019 | 0 | 231 | 0 | 231 | 1.00 | [0.02; 50.61] | 1.6% | 1.5% |
| Fineberg 2023 | 0 | 10 | 0 | 10 | 1.00 | [0.02; 55.27] | 1.5% | 1.5% |
| Fu 2020 | 0 | 113 | 0 | 113 | 1.00 | [0.02; 50.83] | 1.5% | 1.5% |
| George 2017 | 0 | 12 | 0 | 12 | 1.00 | [0.02; 54.46] | 1.5% | 1.5% |
| Glue 2024b | 0 | 17 | 0 | 17 | 1.00 | [0.02; 53.28] | 1.5% | 1.5% |
| Grabski 2022 | 0 | 48 | 0 | 48 | 1.00 | [0.02; 51.42] | 1.5% | 1.5% |
| Grunebaum 2017 | 0 | 9 | 0 | 9 | 1.00 | [0.02; 55.80] | 1.5% | 1.5% |
| Grunebaum 2018 | 0 | 40 | 0 | 40 | 1.00 | [0.02; 51.63] | 1.5% | 1.5% |
| Ionescu 2020 | 0 | 114 | 0 | 114 | 1.00 | [0.02; 50.83] | 1.5% | 1.5% |
| Jones 2024 | 0 | 17 | 0 | 17 | 1.00 | [0.02; 53.28] | 1.5% | 1.5% |
| Lapidus 2014 | 0 | 10 | 0 | 10 | 1.00 | [0.02; 55.27] | 1.5% | 1.5% |
| Leal 2023 | 0 | 5 | 0 | 5 | 1.00 | [0.02; 59.99] | 1.4% | 1.4% |
| Lii 2023 | 0 | 20 | 0 | 20 | 1.00 | [0.02; 52.85] | 1.5% | 1.5% |
| Lijffijt 2021 | 0 | 20 | 0 | 20 | 1.00 | [0.02; 52.85] | 1.5% | 1.5% |
| Loo 2016 | 0 | 9 | 0 | 9 | 1.00 | [0.02; 55.80] | 1.5% | 1.5% |
| Loo 2023 | 0 | 89 | 0 | 89 | 1.00 | [0.02; 50.95] | 1.5% | 1.5% |
| Milak 2020 | 0 | 38 | 0 | 38 | 1.00 | [0.02; 51.69] | 1.5% | 1.5% |
| Murrough 2013 | 0 | 47 | 0 | 47 | 1.00 | [0.02; 51.44] | 1.5% | 1.5% |
| Murrough 2015 II | 0 | 12 | 0 | 12 | 1.00 | [0.02; 54.46] | 1.5% | 1.5% |
| Ochs-Ross 2020 | 0 | 72 | 0 | 72 | 1.00 | [0.02; 51.08] | 1.5% | 1.5% |
| Ohtani 2024 | 0 | 17 | 0 | 17 | 1.00 | [0.02; 53.28] | 1.5% | 1.5% |
| Pattanaseri 2024 | 0 | 11 | 0 | 11 | 1.00 | [0.02; 54.83] | 1.5% | 1.5% |
| Phillips 2019 | 0 | 22 | 0 | 22 | 1.00 | [0.02; 52.63] | 1.5% | 1.5% |
| Popova 2019 | 0 | 115 | 0 | 115 | 1.00 | [0.02; 50.83] | 1.5% | 1.5% |
| Price 2022 | 0 | 103 | 0 | 103 | 1.00 | [0.02; 50.88] | 1.5% | 1.5% |
| Ray-Griffith 2017 | 0 | 8 | 0 | 8 | 1.00 | [0.02; 56.46] | 1.5% | 1.5% |
| Rodriguez 2013 | 0 | 8 | 0 | 8 | 1.00 | [0.02; 56.46] | 1.5% | 1.5% |
| Schiweck 2021 | 0 | 21 | 0 | 21 | 1.00 | [0.02; 52.73] | 1.5% | 1.5% |
| Shiroma 2020 | 0 | 29 | 0 | 29 | 1.00 | [0.02; 52.09] | 1.5% | 1.5% |
| Singh 2016 2x doses | 0 | 18 | 0 | 18 | 1.00 | [0.02; 53.12] | 1.5% | 1.5% |
| Singh 2016 3x doses | 0 | 17 | 0 | 17 | 1.00 | [0.02; 53.28] | 1.5% | 1.5% |
| Singh 2016 II | 0 | 20 | 0 | 20 | 1.00 | [0.02; 52.85] | 1.5% | 1.5% |
| Smith-Apeldoorn 2024 | 1 | 57 | 1 | 57 | 1.00 | [0.06; 16.39] | 3.1% | 3.0% |
| Sos 2013 | 0 | 11 | 0 | 11 | 1.00 | [0.02; 54.83] | 1.5% | 1.5% |
| Su 2017 | 0 | 47 | 0 | 47 | 1.00 | [0.02; 51.44] | 1.5% | 1.5% |
| Su 2023 | 0 | 42 | 0 | 42 | 1.00 | [0.02; 51.57] | 1.5% | 1.5% |
| Sumner 2019 | 0 | 17 | 0 | 17 | 1.00 | [0.02; 53.28] | 1.5% | 1.5% |
| Takahashi 2021 | 0 | 122 | 0 | 122 | 1.00 | [0.02; 50.80] | 1.5% | 1.5% |
| Taylor 2018 | 0 | 9 | 0 | 9 | 1.00 | [0.02; 55.80] | 1.5% | 1.5% |
| Tiger 2020 | 0 | 20 | 0 | 20 | 1.00 | [0.02; 52.85] | 1.5% | 1.5% |
| Wang 2024 | 0 | 182 | 0 | 182 | 1.00 | [0.02; 50.67] | 1.6% | 1.5% |
| Xiao 2023 | 0 | 15 | 0 | 15 | 1.00 | [0.02; 53.66] | 1.5% | 1.5% |
| Yang 2023 | 0 | 200 | 0 | 200 | 1.00 | [0.02; 50.64] | 1.6% | 1.5% |
| Zarate 2006 | 0 | 9 | 0 | 9 | 1.00 | [0.02; 55.80] | 1.5% | 1.5% |
| Zarate 2012 | 0 | 21 | 0 | 21 | 1.00 | [0.02; 52.73] | 1.5% | 1.5% |
| Zhou 2024 | 0 | 27 | 0 | 27 | 1.00 | [0.02; 52.22] | 1.5% | 1.5% |
| **Common effect model** | | **2792** | | **2792** | **1.03** | **[0.63; 1.67]** | **100.0%** | **.** |
| **Random effects model** | | | | | **1.03** | **[0.63; 1.67]** | **.** | **100.0%** |

Heterogeneity: $I^2 = 0.0\%$, $\tau^2 = 0$, $p = 1.0000$

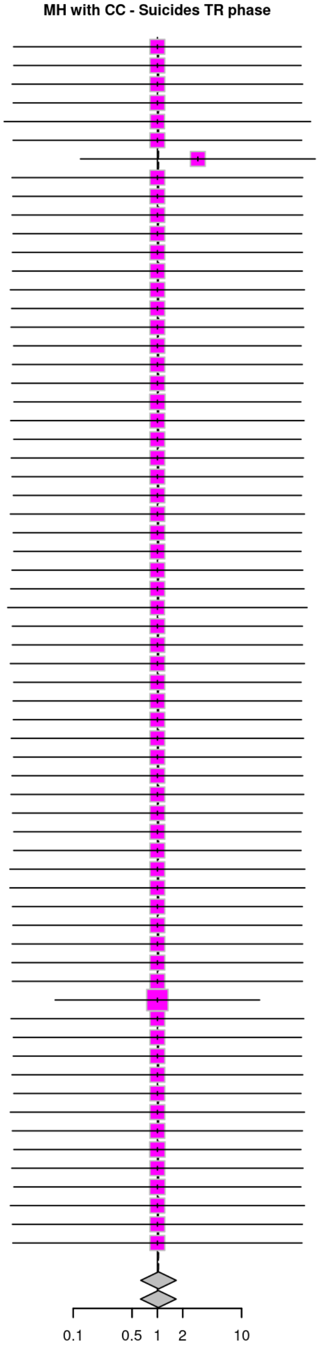

#### Maentel Mantel-Haenszel with Treatment-Arm CC

| Study | Experimental Events | Experimental Total | Control Events | Control Total | MH with TA-CC - Suicides TR phase | OR | 95%-CI | Weight (common) | Weight (random) |
|---|---|---|---|---|---|---|---|---|---|
| Abbar 2022 | 0 | 73 | 0 | 73 | | 1.00 | [0.02; 51.07] | 1.5% | 1.5% |
| Abdallah 2022 | 0 | 108 | 0 | 108 | | 1.00 | [0.02; 50.85] | 1.5% | 1.5% |
| Ahmed 2023 | 0 | 18 | 0 | 18 | | 1.00 | [0.02; 53.12] | 1.5% | 1.5% |
| Anderson 2017 | 0 | 40 | 0 | 40 | | 1.00 | [0.02; 51.63] | 1.5% | 1.5% |
| Burger 2016 | 0 | 3 | 0 | 3 | | 1.00 | [0.02; 66.06] | 1.4% | 1.4% |
| Canuso 2018 | 0 | 35 | 0 | 35 | | 1.00 | [0.02; 51.80] | 1.5% | 1.5% |
| Chen 2023 | 1 | 126 | 0 | 126 | | 3.02 | [0.12; 74.94] | 1.5% | 2.3% |
| Colla 2024 | 0 | 17 | 0 | 17 | | 1.00 | [0.02; 53.28] | 1.5% | 1.5% |
| Dakwar 2019 | 0 | 27 | 0 | 27 | | 1.00 | [0.02; 52.22] | 1.5% | 1.5% |
| Dakwar 2020 | 0 | 17 | 0 | 17 | | 1.00 | [0.02; 53.28] | 1.5% | 1.5% |
| Daly 2018 PA | 0 | 34 | 0 | 34 | | 1.00 | [0.02; 51.84] | 1.5% | 1.5% |
| Daly 2018 PB | 0 | 20 | 0 | 20 | | 1.00 | [0.02; 52.85] | 1.5% | 1.5% |
| Domany 2019 | 0 | 22 | 0 | 22 | | 1.00 | [0.02; 52.63] | 1.5% | 1.5% |
| Domany 2020 | 0 | 9 | 0 | 9 | | 1.00 | [0.02; 55.80] | 1.5% | 1.5% |
| Duek 2023 | 0 | 14 | 0 | 14 | | 1.00 | [0.02; 53.89] | 1.5% | 1.5% |
| Dwyer 2021 | 0 | 11 | 0 | 11 | | 1.00 | [0.02; 54.83] | 1.5% | 1.5% |
| Fava 2020 | 0 | 80 | 0 | 80 | | 1.00 | [0.02; 51.01] | 1.5% | 1.5% |
| Feder 2014 | 0 | 22 | 0 | 22 | | 1.00 | [0.02; 52.63] | 1.5% | 1.5% |
| Feder 2021 | 0 | 15 | 0 | 15 | | 1.00 | [0.02; 53.66] | 1.5% | 1.5% |
| Fedgchin 2019 | 0 | 231 | 0 | 231 | | 1.00 | [0.02; 50.61] | 1.6% | 1.5% |
| Fineberg 2023 | 0 | 10 | 0 | 10 | | 1.00 | [0.02; 55.27] | 1.5% | 1.5% |
| Fu 2020 | 0 | 113 | 0 | 113 | | 1.00 | [0.02; 50.83] | 1.5% | 1.5% |
| George 2017 | 0 | 12 | 0 | 12 | | 1.00 | [0.02; 54.46] | 1.5% | 1.5% |
| Glue 2024b | 0 | 17 | 0 | 17 | | 1.00 | [0.02; 53.28] | 1.5% | 1.5% |
| Grabski 2022 | 0 | 48 | 0 | 48 | | 1.00 | [0.02; 51.42] | 1.5% | 1.5% |
| Grunebaum 2017 | 0 | 9 | 0 | 9 | | 1.00 | [0.02; 55.80] | 1.5% | 1.5% |
| Grunebaum 2018 | 0 | 40 | 0 | 40 | | 1.00 | [0.02; 51.63] | 1.5% | 1.5% |
| Ionescu 2020 | 0 | 114 | 0 | 114 | | 1.00 | [0.02; 50.83] | 1.5% | 1.5% |
| Jones 2024 | 0 | 17 | 0 | 17 | | 1.00 | [0.02; 53.28] | 1.5% | 1.5% |
| Lapidus 2014 | 0 | 10 | 0 | 10 | | 1.00 | [0.02; 55.27] | 1.5% | 1.5% |
| Leal 2023 | 0 | 5 | 0 | 5 | | 1.00 | [0.02; 59.99] | 1.4% | 1.4% |
| Lii 2023 | 0 | 20 | 0 | 20 | | 1.00 | [0.02; 52.85] | 1.5% | 1.5% |
| Lijffijt 2021 | 0 | 20 | 0 | 20 | | 1.00 | [0.02; 52.85] | 1.5% | 1.5% |
| Loo 2016 | 0 | 9 | 0 | 9 | | 1.00 | [0.02; 55.80] | 1.5% | 1.5% |
| Loo 2023 | 0 | 89 | 0 | 89 | | 1.00 | [0.02; 50.95] | 1.5% | 1.5% |
| Milak 2020 | 0 | 38 | 0 | 38 | | 1.00 | [0.02; 51.69] | 1.5% | 1.5% |
| Murrough 2013 | 0 | 47 | 0 | 47 | | 1.00 | [0.02; 51.44] | 1.5% | 1.5% |
| Murrough 2015 II | 0 | 12 | 0 | 12 | | 1.00 | [0.02; 54.46] | 1.5% | 1.5% |
| Ochs-Ross 2020 | 0 | 72 | 0 | 72 | | 1.00 | [0.02; 51.08] | 1.5% | 1.5% |
| Ohtani 2024 | 0 | 17 | 0 | 17 | | 1.00 | [0.02; 53.28] | 1.5% | 1.5% |
| Pattanaseri 2024 | 0 | 11 | 0 | 11 | | 1.00 | [0.02; 54.83] | 1.5% | 1.5% |
| Phillips 2019 | 0 | 22 | 0 | 22 | | 1.00 | [0.02; 52.63] | 1.5% | 1.5% |
| Popova 2019 | 0 | 115 | 0 | 115 | | 1.00 | [0.02; 50.83] | 1.5% | 1.5% |
| Price 2022 | 0 | 103 | 0 | 103 | | 1.00 | [0.02; 50.88] | 1.5% | 1.5% |
| Ray-Griffith 2017 | 0 | 8 | 0 | 8 | | 1.00 | [0.02; 56.46] | 1.5% | 1.5% |
| Rodriguez 2013 | 0 | 8 | 0 | 8 | | 1.00 | [0.02; 56.46] | 1.5% | 1.5% |
| Schiweck 2021 | 0 | 21 | 0 | 21 | | 1.00 | [0.02; 52.73] | 1.5% | 1.5% |
| Shiroma 2020 | 0 | 29 | 0 | 29 | | 1.00 | [0.02; 52.09] | 1.5% | 1.5% |
| Singh 2016 2x doses | 0 | 18 | 0 | 18 | | 1.00 | [0.02; 53.12] | 1.5% | 1.5% |
| Singh 2016 3x doses | 0 | 17 | 0 | 17 | | 1.00 | [0.02; 53.28] | 1.5% | 1.5% |
| Singh 2016 II | 0 | 20 | 0 | 20 | | 1.00 | [0.02; 52.85] | 1.5% | 1.5% |
| Smith-Apeldoorn 2024 | 1 | 57 | 1 | 57 | | 1.00 | [0.06; 16.39] | 3.1% | 3.0% |
| Sos 2013 | 0 | 11 | 0 | 11 | | 1.00 | [0.02; 54.83] | 1.5% | 1.5% |
| Su 2017 | 0 | 47 | 0 | 47 | | 1.00 | [0.02; 51.44] | 1.5% | 1.5% |
| Su 2023 | 0 | 42 | 0 | 42 | | 1.00 | [0.02; 51.57] | 1.5% | 1.5% |
| Sumner 2019 | 0 | 17 | 0 | 17 | | 1.00 | [0.02; 53.28] | 1.5% | 1.5% |
| Takahashi 2021 | 0 | 122 | 0 | 122 | | 1.00 | [0.02; 50.80] | 1.5% | 1.5% |
| Taylor 2018 | 0 | 9 | 0 | 9 | | 1.00 | [0.02; 55.80] | 1.5% | 1.5% |
| Tiger 2020 | 0 | 20 | 0 | 20 | | 1.00 | [0.02; 52.85] | 1.5% | 1.5% |
| Wang 2024 | 0 | 182 | 0 | 182 | | 1.00 | [0.02; 50.67] | 1.6% | 1.5% |
| Xiao 2023 | 0 | 15 | 0 | 15 | | 1.00 | [0.02; 53.66] | 1.5% | 1.5% |
| Yang 2023 | 0 | 200 | 0 | 200 | | 1.00 | [0.02; 50.64] | 1.6% | 1.5% |
| Zarate 2006 | 0 | 9 | 0 | 9 | | 1.00 | [0.02; 55.80] | 1.5% | 1.5% |
| Zarate 2012 | 0 | 21 | 0 | 21 | | 1.00 | [0.02; 52.73] | 1.5% | 1.5% |
| Zhou 2024 | 0 | 27 | 0 | 27 | | 1.00 | [0.02; 52.22] | 1.5% | 1.5% |
| **Common effect model** | | **2792** | | **2792** | | **1.03** | **[0.63; 1.67]** | **100.0%** | **.** |
| **Random effects model** | | | | | | **1.03** | **[0.63; 1.67]** | **.** | **100.0%** |

Heterogeneity: $I^2 = 0.0\%$, $\tau^2 = 0$, $p = 1.0000$

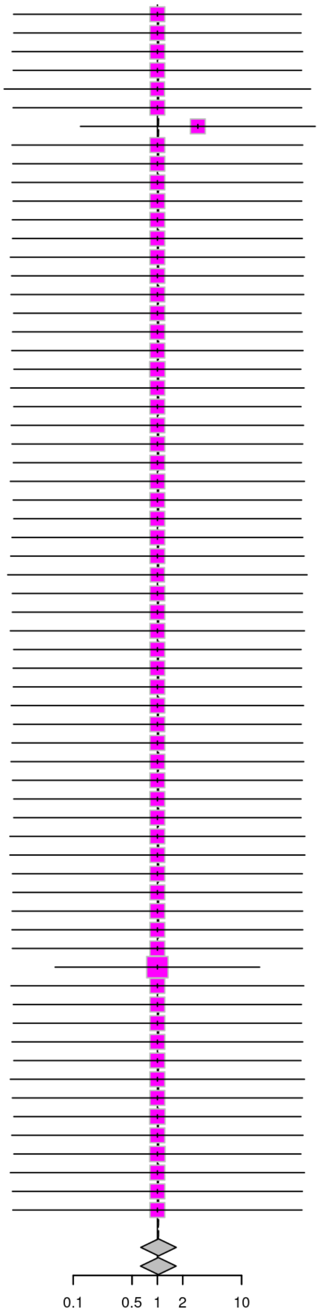

#### Maentel Mantel-Haenszel Risk-Difference

| Study | Experimental Events | Experimental Total | Control Events | Control Total | RD | 95%-CI | Weight (common) | Weight (random) |
|---|---|---|---|---|---|---|---|---|
| Abbar 2022 | 0 | 73 | 0 | 73 | 0.00 | [-0.03; 0.03] | 2.6% | 2.0% |
| Abdallah 2022 | 0 | 108 | 0 | 108 | 0.00 | [-0.02; 0.02] | 3.8% | 4.4% |
| Ahmed 2023 | 0 | 18 | 0 | 18 | 0.00 | [-0.10; 0.10] | 0.7% | 0.1% |
| Anderson 2017 | 0 | 40 | 0 | 40 | 0.00 | [-0.05; 0.05] | 1.4% | 0.6% |
| Burger 2016 | 0 | 3 | 0 | 3 | 0.00 | [-0.46; 0.46] | 0.1% | 0.0% |
| Canuso 2018 | 0 | 35 | 0 | 35 | 0.00 | [-0.05; 0.05] | 1.3% | 0.5% |
| Chen 2023 | 1 | 126 | 0 | 126 | 0.01 | [-0.01; 0.03] | 4.4% | 3.0% |
| Colla 2024 | 0 | 17 | 0 | 17 | 0.00 | [-0.11; 0.11] | 0.6% | 0.1% |
| Dakwar 2019 | 0 | 27 | 0 | 27 | 0.00 | [-0.07; 0.07] | 1.0% | 0.3% |
| Dakwar 2020 | 0 | 17 | 0 | 17 | 0.00 | [-0.11; 0.11] | 0.6% | 0.1% |
| Daly 2018 PA | 0 | 34 | 0 | 34 | 0.00 | [-0.06; 0.06] | 1.2% | 0.5% |
| Daly 2018 PB | 0 | 20 | 0 | 20 | 0.00 | [-0.09; 0.09] | 0.7% | 0.2% |
| Domany 2019 | 0 | 22 | 0 | 22 | 0.00 | [-0.08; 0.08] | 0.8% | 0.2% |
| Domany 2020 | 0 | 9 | 0 | 9 | 0.00 | [-0.19; 0.19] | 0.4% | 0.0% |
| Duek 2023 | 0 | 14 | 0 | 14 | 0.00 | [-0.13; 0.13] | 0.5% | 0.1% |
| Dwyer 2021 | 0 | 11 | 0 | 11 | 0.00 | [-0.16; 0.16] | 0.4% | 0.1% |
| Fava 2020 | 0 | 80 | 0 | 80 | 0.00 | [-0.02; 0.02] | 2.8% | 2.4% |
| Feder 2014 | 0 | 22 | 0 | 22 | 0.00 | [-0.08; 0.08] | 0.8% | 0.2% |
| Feder 2021 | 0 | 15 | 0 | 15 | 0.00 | [-0.12; 0.12] | 0.6% | 0.1% |
| Fedgchin 2019 | 0 | 231 | 0 | 231 | 0.00 | [-0.01; 0.01] | 8.1% | 19.9% |
| Fineberg 2023 | 0 | 10 | 0 | 10 | 0.00 | [-0.17; 0.17] | 0.4% | 0.0% |
| Fu 2020 | 0 | 113 | 0 | 113 | 0.00 | [-0.02; 0.02] | 4.0% | 4.8% |
| George 2017 | 0 | 12 | 0 | 12 | 0.00 | [-0.15; 0.15] | 0.5% | 0.1% |
| Glue 2024b | 0 | 17 | 0 | 17 | 0.00 | [-0.11; 0.11] | 0.6% | 0.1% |
| Grabski 2022 | 0 | 48 | 0 | 48 | 0.00 | [-0.04; 0.04] | 1.7% | 0.9% |
| Grunebaum 2017 | 0 | 9 | 0 | 9 | 0.00 | [-0.19; 0.19] | 0.4% | 0.0% |
| Grunebaum 2018 | 0 | 40 | 0 | 40 | 0.00 | [-0.05; 0.05] | 1.4% | 0.6% |
| Ionescu 2020 | 0 | 114 | 0 | 114 | 0.00 | [-0.02; 0.02] | 4.0% | 4.9% |
| Jones 2024 | 0 | 17 | 0 | 17 | 0.00 | [-0.11; 0.11] | 0.6% | 0.1% |
| Lapidus 2014 | 0 | 10 | 0 | 10 | 0.00 | [-0.17; 0.17] | 0.4% | 0.0% |
| Leal 2023 | 0 | 5 | 0 | 5 | 0.00 | [-0.31; 0.31] | 0.2% | 0.0% |
| Lii 2023 | 0 | 20 | 0 | 20 | 0.00 | [-0.09; 0.09] | 0.7% | 0.2% |
| Lijffijt 2021 | 0 | 20 | 0 | 20 | 0.00 | [-0.09; 0.09] | 0.7% | 0.2% |
| Loo 2016 | 0 | 9 | 0 | 9 | 0.00 | [-0.19; 0.19] | 0.4% | 0.0% |
| Loo 2023 | 0 | 89 | 0 | 89 | 0.00 | [-0.02; 0.02] | 3.2% | 3.0% |
| Milak 2020 | 0 | 38 | 0 | 38 | 0.00 | [-0.05; 0.05] | 1.4% | 0.6% |
| Murrough 2013 | 0 | 47 | 0 | 47 | 0.00 | [-0.04; 0.04] | 1.7% | 0.9% |
| Murrough 2015 II | 0 | 12 | 0 | 12 | 0.00 | [-0.15; 0.15] | 0.5% | 0.1% |
| Ochs-Ross 2020 | 0 | 72 | 0 | 72 | 0.00 | [-0.03; 0.03] | 2.6% | 2.0% |
| Ohtani 2024 | 0 | 17 | 0 | 17 | 0.00 | [-0.11; 0.11] | 0.6% | 0.1% |
| Pattanaseri 2024 | 0 | 11 | 0 | 11 | 0.00 | [-0.16; 0.16] | 0.4% | 0.1% |
| Phillips 2019 | 0 | 22 | 0 | 22 | 0.00 | [-0.08; 0.08] | 0.8% | 0.2% |
| Popova 2019 | 0 | 115 | 0 | 115 | 0.00 | [-0.02; 0.02] | 4.1% | 5.0% |
| Price 2022 | 0 | 103 | 0 | 103 | 0.00 | [-0.02; 0.02] | 3.6% | 4.0% |
| Ray-Griffith 2017 | 0 | 8 | 0 | 8 | 0.00 | [-0.21; 0.21] | 0.3% | 0.0% |
| Rodriguez 2013 | 0 | 8 | 0 | 8 | 0.00 | [-0.21; 0.21] | 0.3% | 0.0% |
| Schiweck 2021 | 0 | 21 | 0 | 21 | 0.00 | [-0.09; 0.09] | 0.8% | 0.2% |
| Shiroma 2020 | 0 | 29 | 0 | 29 | 0.00 | [-0.06; 0.06] | 1.1% | 0.3% |
| Singh 2016 2x doses | 0 | 18 | 0 | 18 | 0.00 | [-0.10; 0.10] | 0.7% | 0.1% |
| Singh 2016 3x doses | 0 | 17 | 0 | 17 | 0.00 | [-0.11; 0.11] | 0.6% | 0.1% |
| Singh 2016 II | 0 | 20 | 0 | 20 | 0.00 | [-0.09; 0.09] | 0.7% | 0.2% |
| Smith-Apeldoorn 2024 | 1 | 57 | 1 | 57 | 0.00 | [-0.05; 0.05] | 2.0% | 0.6% |
| Sos 2013 | 0 | 11 | 0 | 11 | 0.00 | [-0.16; 0.16] | 0.4% | 0.1% |
| Su 2017 | 0 | 47 | 0 | 47 | 0.00 | [-0.04; 0.04] | 1.7% | 0.9% |
| Su 2023 | 0 | 42 | 0 | 42 | 0.00 | [-0.05; 0.05] | 1.5% | 0.7% |
| Sumner 2019 | 0 | 17 | 0 | 17 | 0.00 | [-0.11; 0.11] | 0.6% | 0.1% |
| Takahashi 2021 | 0 | 122 | 0 | 122 | 0.00 | [-0.02; 0.02] | 4.3% | 5.6% |
| Taylor 2018 | 0 | 9 | 0 | 9 | 0.00 | [-0.19; 0.19] | 0.4% | 0.0% |
| Tiger 2020 | 0 | 20 | 0 | 20 | 0.00 | [-0.09; 0.09] | 0.7% | 0.2% |
| Wang 2024 | 0 | 182 | 0 | 182 | 0.00 | [-0.01; 0.01] | 6.4% | 12.4% |
| Xiao 2023 | 0 | 15 | 0 | 15 | 0.00 | [-0.12; 0.12] | 0.6% | 0.1% |
| Yang 2023 | 0 | 200 | 0 | 200 | 0.00 | [-0.01; 0.01] | 7.0% | 15.0% |
| Zarate 2006 | 0 | 9 | 0 | 9 | 0.00 | [-0.19; 0.19] | 0.4% | 0.0% |
| Zarate 2012 | 0 | 21 | 0 | 21 | 0.00 | [-0.09; 0.09] | 0.8% | 0.2% |
| Zhou 2024 | 0 | 27 | 0 | 27 | 0.00 | [-0.07; 0.07] | 1.0% | 0.3% |
| **Common effect model** | | **2792** | | **2792** | **0.00** | **[-0.01; 0.01]** | **100.0%** | **.** |
| **Random effects model** | | | | | **0.00** | **[-0.00; 0.00]** | **.** | **100.0%** |

Heterogeneity: $I^2 = 0.0\%$, $\tau^2 = 0$, $p = 1.0000$

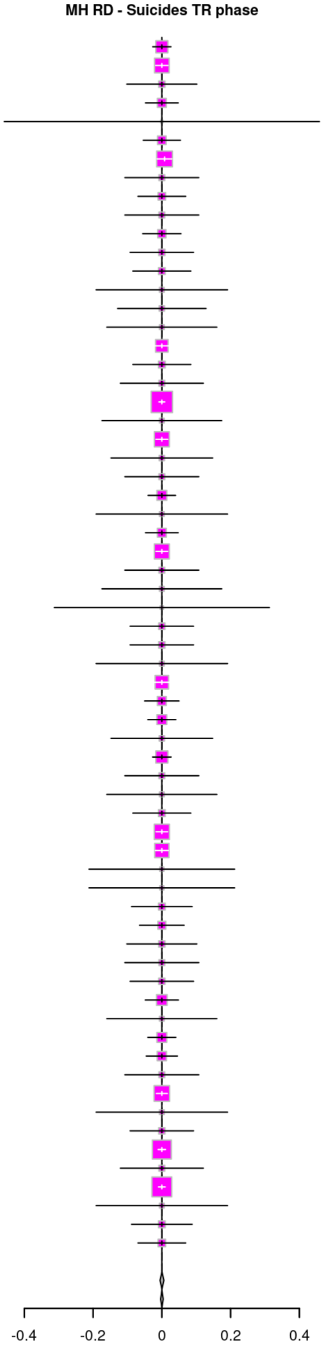

#### Arcsine Risk-Difference

##### Arcsine RD, DB, Suicides

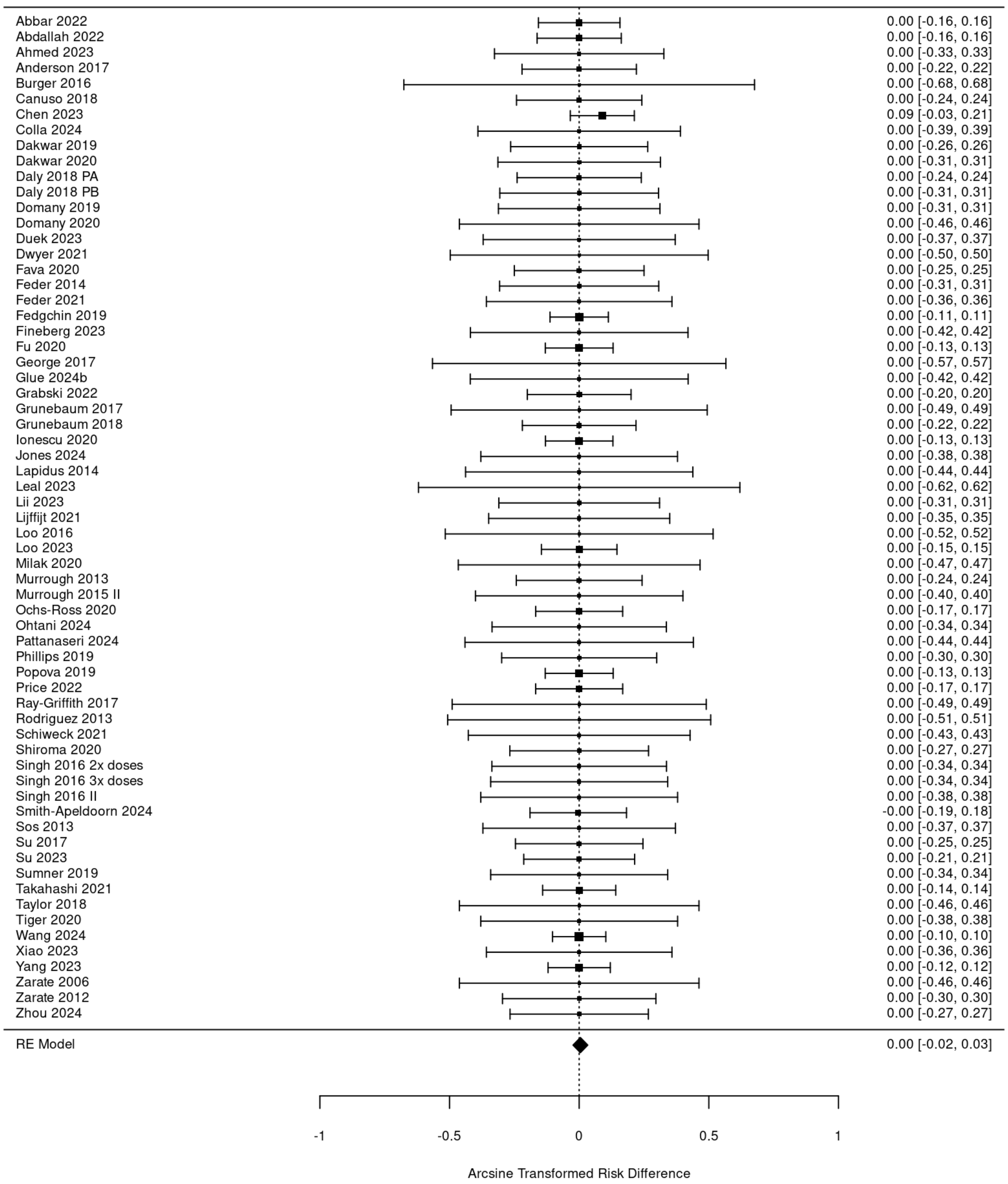

**Bayesian Meta-Analysis, wider prior**

**Bayesian MA, Delta = 250 - Suicides TR**

| Study | Estimate | 95% CI |
|---|---|---|
| Abbar 2022 | 0.13 | [-3.80, 4.06] |
| Abdallah 2022 | -0.67 | [-4.60, 3.26] |
| Ahmed 2023 | 0.00 | [-3.97, 3.97] |
| Anderson 2017 | -0.03 | [-3.97, 3.92] |
| Burger 2016 | 0.76 | [-3.36, 4.88] |
| Canuso 2018 | -0.12 | [-4.07, 3.83] |
| Chen 2023 | 1.11 | [-2.10, 4.32] |
| Colla 2024 | -0.51 | [-4.50, 3.48] |
| Dakwar 2019 | 0.04 | [-3.92, 3.99] |
| Dakwar 2020 | 0.29 | [-3.67, 4.26] |
| Daly 2018 PA | -0.03 | [-3.98, 3.92] |
| Daly 2018 PB | 0.05 | [-3.92, 4.01] |
| Domany 2019 | -0.20 | [-4.16, 3.77] |
| Domany 2020 | 0.00 | [-4.02, 4.02] |
| Duek 2023 | 0.00 | [-3.99, 3.99] |
| Dwyer 2021 | -0.57 | [-4.61, 3.47] |
| Fava 2020 | -1.42 | [-5.37, 2.53] |
| Feder 2014 | -0.14 | [-4.11, 3.82] |
| Feder 2021 | 0.00 | [-3.98, 3.98] |
| Fedgchin 2019 | -0.71 | [-4.64, 3.21] |
| Fineberg 2023 | 0.17 | [-3.83, 4.18] |
| Fu 2020 | -0.01 | [-3.94, 3.92] |
| George 2017 | -1.02 | [-5.09, 3.04] |
| Glue 2024b | -0.72 | [-4.73, 3.28] |
| Grabski 2022 | 0.00 | [-3.94, 3.94] |
| Grunebaum 2017 | -0.24 | [-4.27, 3.80] |
| Grunebaum 2018 | 0.00 | [-3.94, 3.94] |
| Ionescu 2020 | -0.01 | [-3.94, 3.92] |
| Jones 2024 | -0.42 | [-4.41, 3.57] |
| Lapidus 2014 | 0.00 | [-4.01, 4.01] |
| Leal 2023 | 0.00 | [-4.09, 4.09] |
| Lii 2023 | 0.00 | [-3.97, 3.97] |
| Lijffijt 2021 | -0.42 | [-4.40, 3.56] |
| Loo 2016 | -0.38 | [-4.42, 3.67] |
| Loo 2023 | 0.03 | [-3.90, 3.96] |
| Milak 2020 | -1.95 | [-5.97, 2.07] |
| Murrough 2013 | -0.62 | [-4.57, 3.33] |
| Murrough 2015 II | 0.00 | [-4.00, 4.00] |
| Ochs-Ross 2020 | -0.10 | [-4.04, 3.83] |
| Ohtani 2024 | 0.00 | [-3.98, 3.98] |
| Pattanaseri 2024 | -0.19 | [-4.20, 3.82] |
| Phillips 2019 | -0.05 | [-4.01, 3.92] |
| Popova 2019 | -0.05 | [-3.98, 3.88] |
| Price 2022 | -0.70 | [-4.63, 3.24] |
| Ray-Griffith 2017 | 0.00 | [-4.03, 4.03] |
| Rodriguez 2013 | -0.13 | [-4.17, 3.92] |
| Schiweck 2021 | -1.05 | [-5.06, 2.95] |
| Shiroma 2020 | -0.15 | [-4.10, 3.81] |
| Singh 2016 2x doses | -0.11 | [-4.09, 3.86] |
| Singh 2016 3x doses | -0.06 | [-4.04, 3.92] |
| Singh 2016 II | -0.67 | [-4.66, 3.32] |
| Smith-Apeldoorn 2024 | -0.06 | [-2.85, 2.74] |
| Sos 2013 | 0.53 | [-3.46, 4.52] |
| Su 2017 | -0.66 | [-4.61, 3.29] |
| Su 2023 | 0.00 | [-3.94, 3.94] |
| Sumner 2019 | -0.06 | [-4.04, 3.92] |
| Takahashi 2021 | -0.42 | [-4.35, 3.51] |
| Taylor 2018 | 0.00 | [-4.02, 4.02] |
| Tiger 2020 | -0.67 | [-4.66, 3.32] |
| Wang 2024 | 0.00 | [-3.93, 3.93] |
| Xiao 2023 | 0.00 | [-3.98, 3.98] |
| Yang 2023 | -0.69 | [-4.62, 3.24] |
| Zarate 2006 | 0.00 | [-4.02, 4.02] |
| Zarate 2012 | 0.09 | [-3.87, 4.05] |
| Zhou 2024 | 0.00 | [-3.96, 3.96] |
| **Summary** | **0.05** | **[-1.26, 1.42]** |
| **Prediction** | **0.06** | **[-1.51, 1.64]** |

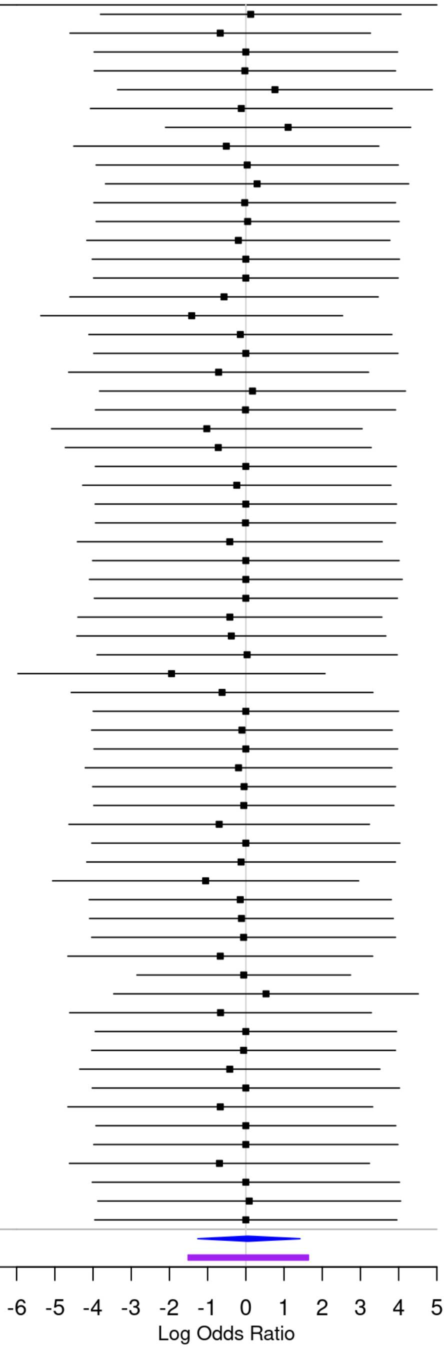

*Heterogeneity (tau): 0.28 [0.00, 0.83]*

**Bayesian Meta-Analysis, smaller prior**

##### Bayesian MA, Delta = 15 - Suicides TR

| Study | Estimate | 95% CI |
|---|---|---|
| Abbar 2022 | 0.13 | [-3.80, 4.06] |
| Abdallah 2022 | -0.67 | [-4.60, 3.26] |
| Ahmed 2023 | 0.00 | [-3.97, 3.97] |
| Anderson 2017 | -0.03 | [-3.97, 3.92] |
| Burger 2016 | 0.76 | [-3.36, 4.88] |
| Canuso 2018 | -0.12 | [-4.07, 3.83] |
| Chen 2023 | 1.11 | [-2.10, 4.32] |
| Colla 2024 | -0.51 | [-4.50, 3.48] |
| Dakwar 2019 | 0.04 | [-3.92, 3.99] |
| Dakwar 2020 | 0.29 | [-3.67, 4.26] |
| Daly 2018 PA | -0.03 | [-3.98, 3.92] |
| Daly 2018 PB | 0.05 | [-3.92, 4.01] |
| Domany 2019 | -0.20 | [-4.16, 3.77] |
| Domany 2020 | 0.00 | [-4.02, 4.02] |
| Duek 2023 | 0.00 | [-3.99, 3.99] |
| Dwyer 2021 | -0.57 | [-4.61, 3.47] |
| Fava 2020 | -1.42 | [-5.37, 2.53] |
| Feder 2014 | -0.14 | [-4.11, 3.82] |
| Feder 2021 | 0.00 | [-3.98, 3.98] |
| Fedgchin 2019 | -0.71 | [-4.64, 3.21] |
| Fineberg 2023 | 0.17 | [-3.83, 4.18] |
| Fu 2020 | -0.01 | [-3.94, 3.92] |
| George 2017 | -1.02 | [-5.09, 3.04] |
| Glue 2024b | -0.72 | [-4.73, 3.28] |
| Grabski 2022 | 0.00 | [-3.94, 3.94] |
| Grunebaum 2017 | -0.24 | [-4.27, 3.80] |
| Grunebaum 2018 | 0.00 | [-3.94, 3.94] |
| Ionescu 2020 | -0.01 | [-3.94, 3.92] |
| Jones 2024 | -0.42 | [-4.41, 3.57] |
| Lapidus 2014 | 0.00 | [-4.01, 4.01] |
| Leal 2023 | 0.00 | [-4.09, 4.09] |
| Lii 2023 | 0.00 | [-3.97, 3.97] |
| Lijffijt 2021 | -0.42 | [-4.40, 3.56] |
| Loo 2016 | -0.38 | [-4.42, 3.67] |
| Loo 2023 | 0.03 | [-3.90, 3.96] |
| Milak 2020 | -1.95 | [-5.97, 2.07] |
| Murrough 2013 | -0.62 | [-4.57, 3.33] |
| Murrough 2015 II | 0.00 | [-4.00, 4.00] |
| Ochs-Ross 2020 | -0.10 | [-4.04, 3.83] |
| Ohtani 2024 | 0.00 | [-3.98, 3.98] |
| Pattanaseri 2024 | -0.19 | [-4.20, 3.82] |
| Phillips 2019 | -0.05 | [-4.01, 3.92] |
| Popova 2019 | -0.05 | [-3.98, 3.88] |
| Price 2022 | -0.70 | [-4.63, 3.24] |
| Ray-Griffith 2017 | 0.00 | [-4.03, 4.03] |
| Rodriguez 2013 | -0.13 | [-4.17, 3.92] |
| Schiweck 2021 | -1.05 | [-5.06, 2.95] |
| Shiroma 2020 | -0.15 | [-4.10, 3.81] |
| Singh 2016 2x doses | -0.11 | [-4.09, 3.86] |
| Singh 2016 3x doses | -0.06 | [-4.04, 3.92] |
| Singh 2016 II | -0.67 | [-4.66, 3.32] |
| Smith-Apeldoorn 2024 | -0.06 | [-2.85, 2.74] |
| Sos 2013 | 0.53 | [-3.46, 4.52] |
| Su 2017 | -0.66 | [-4.61, 3.29] |
| Su 2023 | 0.00 | [-3.94, 3.94] |
| Sumner 2019 | -0.06 | [-4.04, 3.92] |
| Takahashi 2021 | -0.42 | [-4.35, 3.51] |
| Taylor 2018 | 0.00 | [-4.02, 4.02] |
| Tiger 2020 | -0.67 | [-4.66, 3.32] |
| Wang 2024 | 0.00 | [-3.93, 3.93] |
| Xiao 2023 | 0.00 | [-3.98, 3.98] |
| Yang 2023 | -0.69 | [-4.62, 3.24] |
| Zarate 2006 | 0.00 | [-4.02, 4.02] |
| Zarate 2012 | 0.09 | [-3.87, 4.05] |
| Zhou 2024 | 0.00 | [-3.96, 3.96] |
| **Summary** | **0.04** | **[-1.16, 1.27]** |
| **Prediction** | **0.04** | **[-1.42, 1.54]** |

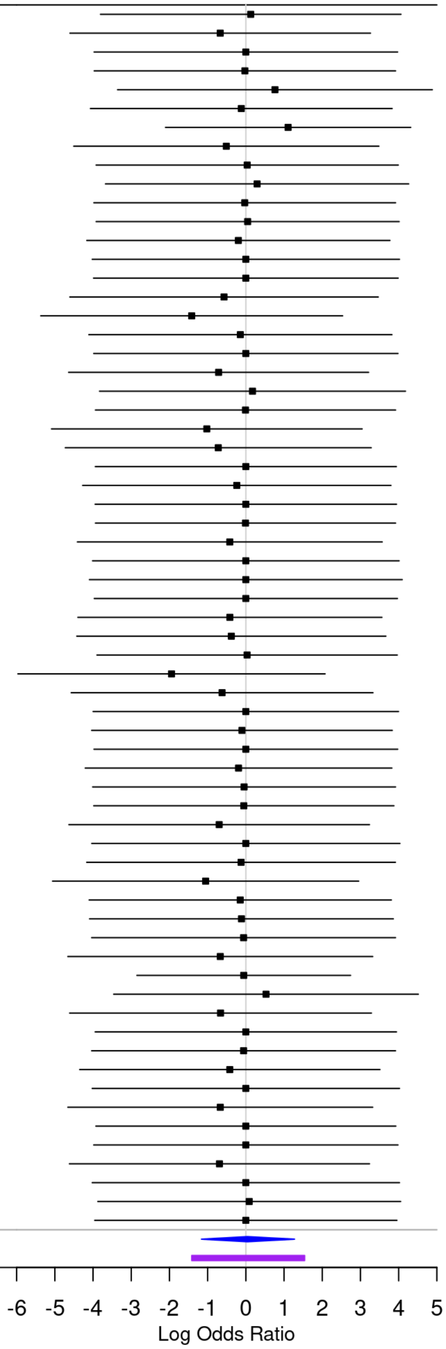

*Heterogeneity (tau): 0.28 [0.00, 0.82]*

**Exact Method**

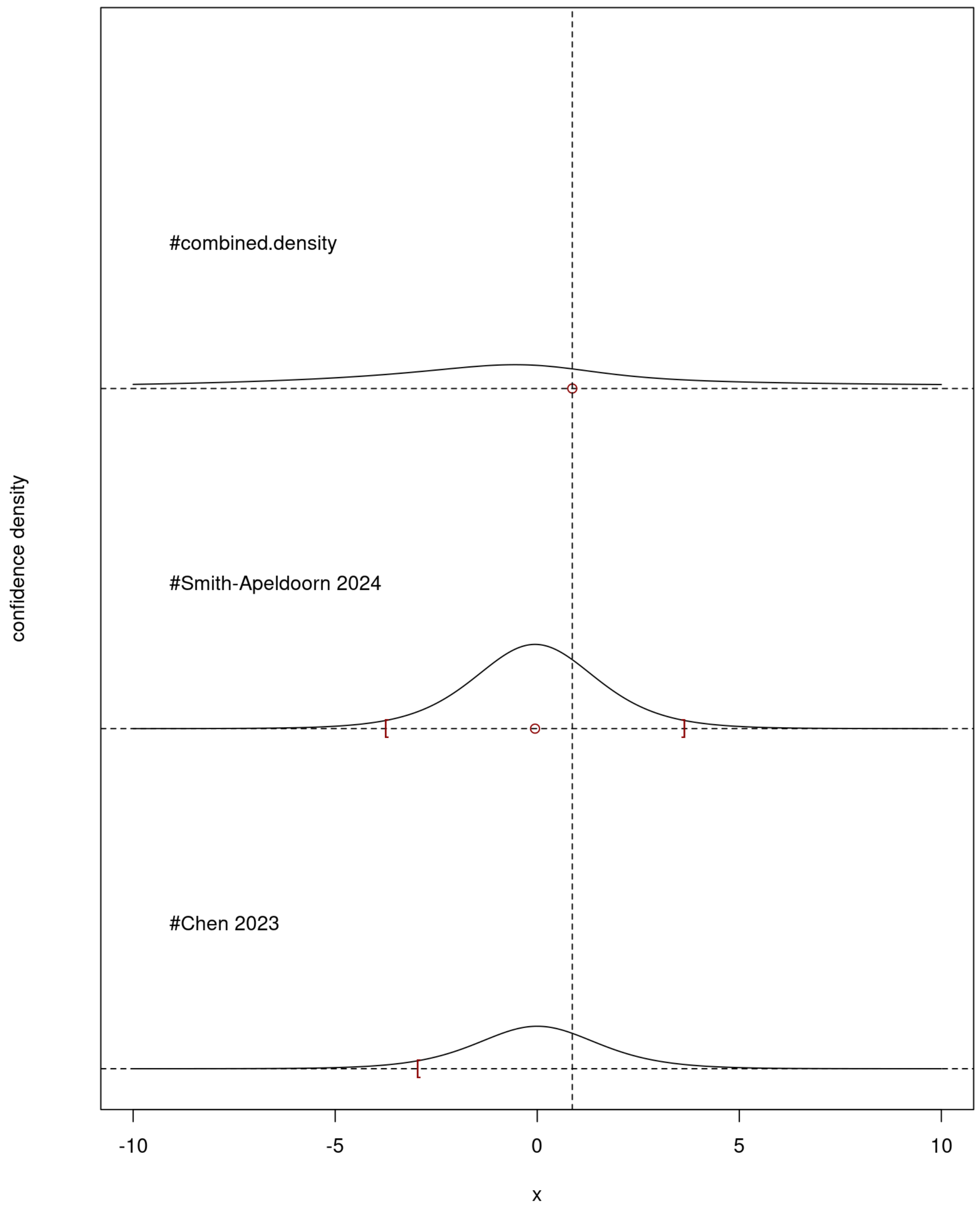

#### Follow-Up Phase

#### Peto's Method

| Study | Experimental Events | Experimental Total | Control Events | Control Total | OR | 95%-CI | Weight (common) | Weight (random) |
|---|---|---|---|---|---|---|---|---|
| Abbar 2022 | 1 | 73 | 0 | 83 | 8.47 | [0.17; 430.52] | 50.0% | 50.0% |
| Ahmed 2023 | 0 | 18 | 0 | 18 | | | 0.0% | 0.0% |
| Anderson 2017 | 0 | 25 | 0 | 23 | | | 0.0% | 0.0% |
| Burger 2016 | 0 | 3 | 0 | 7 | | | 0.0% | 0.0% |
| Canuso 2018 | 0 | 27 | 0 | 22 | | | 0.0% | 0.0% |
| Dakwar 2019 | 0 | 27 | 0 | 28 | | | 0.0% | 0.0% |
| Dakwar 2020 | 0 | 17 | 0 | 23 | | | 0.0% | 0.0% |
| Domany 2019 | 0 | 22 | 0 | 18 | | | 0.0% | 0.0% |
| Domany 2020 | 0 | 9 | 0 | 9 | | | 0.0% | 0.0% |
| Duek 2023 | 0 | 14 | 0 | 14 | | | 0.0% | 0.0% |
| Dwyer 2021 | 0 | 11 | 0 | 6 | | | 0.0% | 0.0% |
| Fava 2020 | 0 | 80 | 0 | 19 | | | 0.0% | 0.0% |
| Feder 2014 | 0 | 22 | 0 | 19 | | | 0.0% | 0.0% |
| Fineberg 2023 | 0 | 10 | 0 | 12 | | | 0.0% | 0.0% |
| Fu 2020 | 1 | 101 | 0 | 91 | 6.69 | [0.13; 339.08] | 50.0% | 50.0% |
| Glue 2024b | 0 | 17 | 0 | 8 | | | 0.0% | 0.0% |
| Grabski 2022 | 0 | 48 | 0 | 48 | | | 0.0% | 0.0% |
| Ionescu 2020 | 0 | 89 | 0 | 94 | | | 0.0% | 0.0% |
| Jones 2024 | 0 | 17 | 0 | 11 | | | 0.0% | 0.0% |
| Lapidus 2014 | 0 | 10 | 0 | 10 | | | 0.0% | 0.0% |
| Leal 2023 | 0 | 5 | 0 | 5 | | | 0.0% | 0.0% |
| Lii 2023 | 0 | 20 | 0 | 20 | | | 0.0% | 0.0% |
| Lijffijt 2021 | 0 | 20 | 0 | 13 | | | 0.0% | 0.0% |
| Murrough 2013 | 0 | 47 | 0 | 25 | | | 0.0% | 0.0% |
| Murrough 2015 II | 0 | 12 | 0 | 12 | | | 0.0% | 0.0% |
| Ohtani 2024 | 0 | 17 | 0 | 17 | | | 0.0% | 0.0% |
| Pattanaseri 2024 | 0 | 11 | 0 | 9 | | | 0.0% | 0.0% |
| Phillips 2019 | 0 | 22 | 0 | 21 | | | 0.0% | 0.0% |
| Price 2022 | 0 | 103 | 0 | 51 | | | 0.0% | 0.0% |
| Ray-Griffith 2017 | 0 | 8 | 0 | 8 | | | 0.0% | 0.0% |
| Rodriguez 2013 | 0 | 8 | 0 | 7 | | | 0.0% | 0.0% |
| Schiweck 2021 | 0 | 21 | 0 | 7 | | | 0.0% | 0.0% |
| Smith-Apeldoorn 2024 | 0 | 49 | 0 | 48 | | | 0.0% | 0.0% |
| Sos 2013 | 0 | 11 | 0 | 19 | | | 0.0% | 0.0% |
| Su 2017 | 0 | 47 | 0 | 24 | | | 0.0% | 0.0% |
| Su 2023 | 0 | 42 | 0 | 42 | | | 0.0% | 0.0% |
| Sumner 2019 | 0 | 17 | 0 | 16 | | | 0.0% | 0.0% |
| Taylor 2018 | 0 | 9 | 0 | 8 | | | 0.0% | 0.0% |
| Wang 2024 | 0 | 182 | 0 | 182 | | | 0.0% | 0.0% |
| Xiao 2023 | 0 | 15 | 0 | 15 | | | 0.0% | 0.0% |
| Yang 2023 | 0 | 200 | 0 | 100 | | | 0.0% | 0.0% |
| Zarate 2006 | 0 | 9 | 0 | 9 | | | 0.0% | 0.0% |
| Zarate 2012 | 0 | 21 | 0 | 23 | | | 0.0% | 0.0% |
| Zhou 2024 | 0 | 27 | 0 | 25 | | | 0.0% | 0.0% |
| **Common effect model** | | **1563** | | **1269** | **7.53** | **[0.47; 120.96]** | **100.0%** | **.** |
| **Random effects model** | | | | | **7.53** | **[0.47; 120.96]** | **.** | **100.0%** |

Heterogeneity: $I^2 = 0.0\%$, $\tau^2 = 0$, $p = 0.9336$

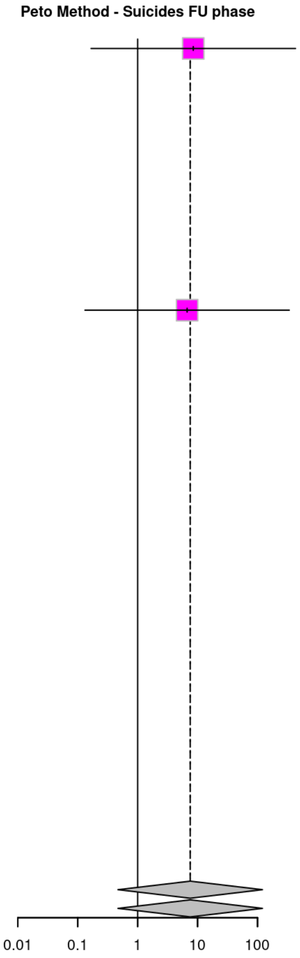

#### Maentel Mantel-Haenszel with CC

| Study | Experimental Events | Experimental Total | Control Events | Control Total | MH with CC - Suicides FU phase | OR | 95%-CI | Weight (common) | Weight (random) |
|---|---|---|---|---|---|---|---|---|---|
| Abbar 2022 | 1 | 73 | 0 | 83 | | 3.46 | [0.14; 86.13] | 2.0% | 3.4% |
| Ahmed 2023 | 0 | 18 | 0 | 18 | | 1.00 | [0.02; 53.12] | 2.1% | 2.2% |
| Anderson 2017 | 0 | 25 | 0 | 23 | | 0.92 | [0.02; 48.33] | 2.2% | 2.2% |
| Burger 2016 | 0 | 3 | 0 | 7 | | 2.14 | [0.03; 131.93] | 1.3% | 2.1% |
| Canuso 2018 | 0 | 27 | 0 | 22 | | 0.82 | [0.02; 42.89] | 2.4% | 2.2% |
| Dakwar 2019 | 0 | 27 | 0 | 28 | | 1.04 | [0.02; 54.08] | 2.1% | 2.3% |
| Dakwar 2020 | 0 | 17 | 0 | 23 | | 1.34 | [0.03; 71.04] | 1.8% | 2.2% |
| Domany 2019 | 0 | 22 | 0 | 18 | | 0.82 | [0.02; 43.47] | 2.3% | 2.2% |
| Domany 2020 | 0 | 9 | 0 | 9 | | 1.00 | [0.02; 55.80] | 2.1% | 2.2% |
| Duek 2023 | 0 | 14 | 0 | 14 | | 1.00 | [0.02; 53.89] | 2.1% | 2.2% |
| Dwyer 2021 | 0 | 11 | 0 | 6 | | 0.57 | [0.01; 32.00] | 2.6% | 2.2% |
| Fava 2020 | 0 | 80 | 0 | 19 | | 0.24 | [0.00; 12.59] | 3.5% | 2.3% |
| Feder 2014 | 0 | 22 | 0 | 19 | | 0.87 | [0.02; 45.76] | 2.3% | 2.2% |
| Fineberg 2023 | 0 | 10 | 0 | 12 | | 1.19 | [0.02; 65.32] | 1.9% | 2.2% |
| Fu 2020 | 1 | 101 | 0 | 91 | | 2.73 | [0.11; 67.89] | 2.3% | 3.4% |
| Glue 2024b | 0 | 17 | 0 | 8 | | 0.49 | [0.01; 26.64] | 2.8% | 2.2% |
| Grabski 2022 | 0 | 48 | 0 | 48 | | 1.00 | [0.02; 51.42] | 2.2% | 2.3% |
| Ionescu 2020 | 0 | 89 | 0 | 94 | | 1.06 | [0.02; 53.78] | 2.1% | 2.3% |
| Jones 2024 | 0 | 17 | 0 | 11 | | 0.66 | [0.01; 35.52] | 2.6% | 2.2% |
| Lapidus 2014 | 0 | 10 | 0 | 10 | | 1.00 | [0.02; 55.27] | 2.1% | 2.2% |
| Leal 2023 | 0 | 5 | 0 | 5 | | 1.00 | [0.02; 59.99] | 2.0% | 2.1% |
| Lii 2023 | 0 | 20 | 0 | 20 | | 1.00 | [0.02; 52.85] | 2.1% | 2.2% |
| Lijffijt 2021 | 0 | 20 | 0 | 13 | | 0.66 | [0.01; 35.23] | 2.6% | 2.2% |
| Murrough 2013 | 0 | 47 | 0 | 25 | | 0.54 | [0.01; 27.86] | 2.8% | 2.3% |
| Murrough 2015 II | 0 | 12 | 0 | 12 | | 1.00 | [0.02; 54.46] | 2.1% | 2.2% |
| Ohtani 2024 | 0 | 17 | 0 | 17 | | 1.00 | [0.02; 53.28] | 2.1% | 2.2% |
| Pattanaseri 2024 | 0 | 11 | 0 | 9 | | 0.83 | [0.01; 45.69] | 2.3% | 2.2% |
| Phillips 2019 | 0 | 22 | 0 | 21 | | 0.96 | [0.02; 50.34] | 2.2% | 2.2% |
| Price 2022 | 0 | 103 | 0 | 51 | | 0.50 | [0.01; 25.44] | 2.9% | 2.3% |
| Ray-Griffith 2017 | 0 | 8 | 0 | 8 | | 1.00 | [0.02; 56.46] | 2.1% | 2.2% |
| Rodriguez 2013 | 0 | 8 | 0 | 7 | | 0.88 | [0.02; 50.19] | 2.2% | 2.2% |
| Schiweck 2021 | 0 | 21 | 0 | 7 | | 0.35 | [0.01; 19.18] | 3.1% | 2.2% |
| Smith-Apeldoorn 2024 | 0 | 49 | 0 | 48 | | 0.98 | [0.02; 50.37] | 2.2% | 2.3% |
| Sos 2013 | 0 | 11 | 0 | 19 | | 1.70 | [0.03; 91.39] | 1.6% | 2.2% |
| Su 2017 | 0 | 47 | 0 | 24 | | 0.52 | [0.01; 26.79] | 2.8% | 2.3% |
| Su 2023 | 0 | 42 | 0 | 42 | | 1.00 | [0.02; 51.57] | 2.2% | 2.3% |
| Sumner 2019 | 0 | 17 | 0 | 16 | | 0.94 | [0.02; 50.32] | 2.2% | 2.2% |
| Taylor 2018 | 0 | 9 | 0 | 8 | | 0.89 | [0.02; 50.22] | 2.2% | 2.2% |
| Wang 2024 | 0 | 182 | 0 | 182 | | 1.00 | [0.02; 50.67] | 2.2% | 2.3% |
| Xiao 2023 | 0 | 15 | 0 | 15 | | 1.00 | [0.02; 53.66] | 2.1% | 2.2% |
| Yang 2023 | 0 | 200 | 0 | 100 | | 0.50 | [0.01; 25.45] | 2.9% | 2.3% |
| Zarate 2006 | 0 | 9 | 0 | 9 | | 1.00 | [0.02; 55.80] | 2.1% | 2.2% |
| Zarate 2012 | 0 | 21 | 0 | 23 | | 1.09 | [0.02; 57.53] | 2.0% | 2.2% |
| Zhou 2024 | 0 | 27 | 0 | 25 | | 0.93 | [0.02; 48.49] | 2.2% | 2.2% |
| **Common effect model** | | **1563** | | **1269** | | **0.96** | **[0.53; 1.72]** | **100.0%** | **.** |
| **Random effects model** | | | | | | **0.93** | **[0.51; 1.68]** | **.** | **100.0%** |

Heterogeneity: $I^2 = 0.0\%$, $\tau^2 = 0$, $p = 1.0000$

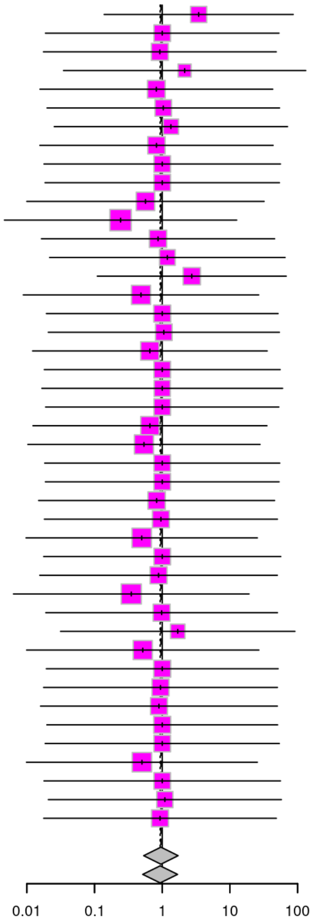

#### Maentel Mantel-Haenszel with Treatment-Arm CC

| Study | Experimental Events | Experimental Total | Control Events | Control Total | OR | 95%-CI | Weight (common) | Weight (random) |
|---|---|---|---|---|---|---|---|---|
| Abbar 2022 | 1 | 73 | 0 | 83 | 3.18 | [0.14; 74.37] | 2.4% | 3.7% |
| Ahmed 2023 | 0 | 18 | 0 | 18 | 1.00 | [0.02; 53.12] | 2.4% | 2.3% |
| Anderson 2017 | 0 | 25 | 0 | 23 | 1.00 | [0.02; 52.62] | 2.4% | 2.3% |
| Burger 2016 | 0 | 3 | 0 | 7 | 1.00 | [0.01; 87.11] | 1.9% | 1.8% |
| Canuso 2018 | 0 | 27 | 0 | 22 | 1.00 | [0.02; 53.50] | 2.4% | 2.3% |
| Dakwar 2019 | 0 | 27 | 0 | 28 | 1.00 | [0.02; 52.22] | 2.4% | 2.3% |
| Dakwar 2020 | 0 | 17 | 0 | 23 | 1.00 | [0.02; 55.30] | 2.3% | 2.3% |
| Domany 2019 | 0 | 22 | 0 | 18 | 1.00 | [0.02; 53.92] | 2.4% | 2.3% |
| Domany 2020 | 0 | 9 | 0 | 9 | 1.00 | [0.02; 55.80] | 2.3% | 2.3% |
| Duek 2023 | 0 | 14 | 0 | 14 | 1.00 | [0.02; 53.89] | 2.4% | 2.3% |
| Dwyer 2021 | 0 | 11 | 0 | 6 | 1.00 | [0.01; 67.61] | 2.1% | 2.1% |
| Fava 2020 | 0 | 80 | 0 | 19 | 1.00 | [0.01; 148.67] | 1.5% | 1.5% |
| Feder 2014 | 0 | 22 | 0 | 19 | 1.00 | [0.02; 53.36] | 2.4% | 2.3% |
| Fineberg 2023 | 0 | 10 | 0 | 12 | 1.00 | [0.02; 55.75] | 2.3% | 2.3% |
| Fu 2020 | 1 | 101 | 0 | 91 | 2.93 | [0.11; 77.20] | 2.4% | 3.4% |
| Glue 2024b | 0 | 17 | 0 | 8 | 1.00 | [0.01; 72.36] | 2.0% | 2.0% |
| Grabski 2022 | 0 | 48 | 0 | 48 | 1.00 | [0.02; 51.42] | 2.4% | 2.4% |
| Ionescu 2020 | 0 | 89 | 0 | 94 | 1.00 | [0.02; 51.01] | 2.4% | 2.4% |
| Jones 2024 | 0 | 17 | 0 | 11 | 1.00 | [0.02; 59.25] | 2.2% | 2.2% |
| Lapidus 2014 | 0 | 10 | 0 | 10 | 1.00 | [0.02; 55.27] | 2.3% | 2.3% |
| Leal 2023 | 0 | 5 | 0 | 5 | 1.00 | [0.02; 59.99] | 2.2% | 2.2% |
| Lii 2023 | 0 | 20 | 0 | 20 | 1.00 | [0.02; 52.85] | 2.4% | 2.3% |
| Lijffijt 2021 | 0 | 20 | 0 | 13 | 1.00 | [0.02; 58.54] | 2.3% | 2.2% |
| Murrough 2013 | 0 | 47 | 0 | 25 | 1.00 | [0.02; 63.11] | 2.2% | 2.1% |
| Murrough 2015 II | 0 | 12 | 0 | 12 | 1.00 | [0.02; 54.46] | 2.3% | 2.3% |
| Ohtani 2024 | 0 | 17 | 0 | 17 | 1.00 | [0.02; 53.28] | 2.4% | 2.3% |
| Pattanaseri 2024 | 0 | 11 | 0 | 9 | 1.00 | [0.02; 56.40] | 2.3% | 2.2% |
| Phillips 2019 | 0 | 22 | 0 | 21 | 1.00 | [0.02; 52.74] | 2.4% | 2.3% |
| Price 2022 | 0 | 103 | 0 | 51 | 1.00 | [0.02; 65.23] | 2.1% | 2.1% |
| Ray-Griffith 2017 | 0 | 8 | 0 | 8 | 1.00 | [0.02; 56.46] | 2.3% | 2.2% |
| Rodriguez 2013 | 0 | 8 | 0 | 7 | 1.00 | [0.02; 57.37] | 2.3% | 2.2% |
| Schiweck 2021 | 0 | 21 | 0 | 7 | 1.00 | [0.01; 99.86] | 1.8% | 1.7% |
| Smith-Apeldoorn 2024 | 0 | 49 | 0 | 48 | 1.00 | [0.02; 51.42] | 2.4% | 2.4% |
| Sos 2013 | 0 | 11 | 0 | 19 | 1.00 | [0.02; 62.32] | 2.2% | 2.1% |
| Su 2017 | 0 | 47 | 0 | 24 | 1.00 | [0.02; 64.85] | 2.1% | 2.1% |
| Su 2023 | 0 | 42 | 0 | 42 | 1.00 | [0.02; 51.57] | 2.4% | 2.3% |
| Sumner 2019 | 0 | 17 | 0 | 16 | 1.00 | [0.02; 53.46] | 2.4% | 2.3% |
| Taylor 2018 | 0 | 9 | 0 | 8 | 1.00 | [0.02; 56.50] | 2.3% | 2.2% |
| Wang 2024 | 0 | 182 | 0 | 182 | 1.00 | [0.02; 50.67] | 2.4% | 2.4% |
| Xiao 2023 | 0 | 15 | 0 | 15 | 1.00 | [0.02; 53.66] | 2.4% | 2.3% |
| Yang 2023 | 0 | 200 | 0 | 100 | 1.00 | [0.02; 64.37] | 2.2% | 2.1% |
| Zarate 2006 | 0 | 9 | 0 | 9 | 1.00 | [0.02; 55.80] | 2.3% | 2.3% |
| Zarate 2012 | 0 | 21 | 0 | 23 | 1.00 | [0.02; 52.84] | 2.4% | 2.3% |
| Zhou 2024 | 0 | 27 | 0 | 25 | 1.00 | [0.02; 52.44] | 2.4% | 2.3% |
| **Common effect model** | | **1563** | | **1269** | **1.10** | **[0.60; 2.00]** | **100.0%** | **.** |
| **Random effects model** | | | | | **1.08** | **[0.59; 1.98]** | **.** | **100.0%** |

Heterogeneity: $I^2 = 0.0\%$, $\tau^2 = 0$, $p = 1.0000$

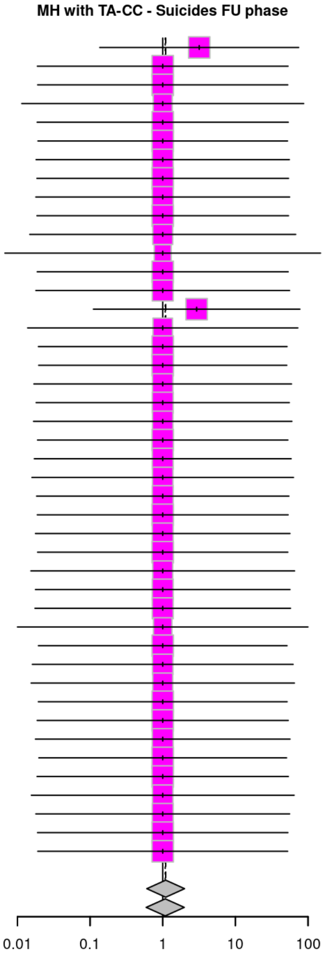

#### Maentel Mantel-Haenszel Risk-Difference

| Study | Experimental Events | Experimental Total | Control Events | Control Total | MH RD - Suicides FU phase | RD | 95%-CI | Weight (common) | Weight (random) |
|---|---|---|---|---|---|---|---|---|---|
| Abbar 2022 | 1 | 73 | 0 | 83 | | 0.01 | [-0.02; 0.05] | 5.6% | 3.3% |
| Ahmed 2023 | 0 | 18 | 0 | 18 | | 0.00 | [-0.10; 0.10] | 1.4% | 0.4% |
| Anderson 2017 | 0 | 25 | 0 | 23 | | 0.00 | [-0.08; 0.08] | 1.8% | 0.7% |
| Burger 2016 | 0 | 3 | 0 | 7 | | 0.00 | [-0.36; 0.36] | 0.4% | 0.0% |
| Canuso 2018 | 0 | 27 | 0 | 22 | | 0.00 | [-0.08; 0.08] | 1.8% | 0.7% |
| Dakwar 2019 | 0 | 27 | 0 | 28 | | 0.00 | [-0.07; 0.07] | 2.0% | 0.9% |
| Dakwar 2020 | 0 | 17 | 0 | 23 | | 0.00 | [-0.10; 0.10] | 1.5% | 0.5% |
| Domany 2019 | 0 | 22 | 0 | 18 | | 0.00 | [-0.09; 0.09] | 1.5% | 0.5% |
| Domany 2020 | 0 | 9 | 0 | 9 | | 0.00 | [-0.19; 0.19] | 0.7% | 0.1% |
| Duek 2023 | 0 | 14 | 0 | 14 | | 0.00 | [-0.13; 0.13] | 1.1% | 0.3% |
| Dwyer 2021 | 0 | 11 | 0 | 6 | | 0.00 | [-0.22; 0.22] | 0.6% | 0.1% |
| Fava 2020 | 0 | 80 | 0 | 19 | | 0.00 | [-0.07; 0.07] | 2.3% | 0.9% |
| Feder 2014 | 0 | 22 | 0 | 19 | | 0.00 | [-0.09; 0.09] | 1.5% | 0.5% |
| Fineberg 2023 | 0 | 10 | 0 | 12 | | 0.00 | [-0.16; 0.16] | 0.9% | 0.2% |
| Fu 2020 | 1 | 101 | 0 | 91 | | 0.01 | [-0.02; 0.04] | 6.9% | 5.5% |
| Glue 2024b | 0 | 17 | 0 | 8 | | 0.00 | [-0.17; 0.17] | 0.9% | 0.2% |
| Grabski 2022 | 0 | 48 | 0 | 48 | | 0.00 | [-0.04; 0.04] | 3.5% | 2.7% |
| Ionescu 2020 | 0 | 89 | 0 | 94 | | 0.00 | [-0.02; 0.02] | 6.6% | 9.6% |
| Jones 2024 | 0 | 17 | 0 | 11 | | 0.00 | [-0.14; 0.14] | 1.0% | 0.2% |
| Lapidus 2014 | 0 | 10 | 0 | 10 | | 0.00 | [-0.17; 0.17] | 0.8% | 0.1% |
| Leal 2023 | 0 | 5 | 0 | 5 | | 0.00 | [-0.31; 0.31] | 0.4% | 0.0% |
| Lii 2023 | 0 | 20 | 0 | 20 | | 0.00 | [-0.09; 0.09] | 1.5% | 0.5% |
| Lijffijt 2021 | 0 | 20 | 0 | 13 | | 0.00 | [-0.12; 0.12] | 1.2% | 0.3% |
| Murrough 2013 | 0 | 47 | 0 | 25 | | 0.00 | [-0.06; 0.06] | 2.4% | 1.2% |
| Murrough 2015 II | 0 | 12 | 0 | 12 | | 0.00 | [-0.15; 0.15] | 0.9% | 0.2% |
| Ohtani 2024 | 0 | 17 | 0 | 17 | | 0.00 | [-0.11; 0.11] | 1.3% | 0.4% |
| Pattanaseri 2024 | 0 | 11 | 0 | 9 | | 0.00 | [-0.18; 0.18] | 0.8% | 0.1% |
| Phillips 2019 | 0 | 22 | 0 | 21 | | 0.00 | [-0.09; 0.09] | 1.6% | 0.6% |
| Price 2022 | 0 | 103 | 0 | 51 | | 0.00 | [-0.03; 0.03] | 5.0% | 4.9% |
| Ray-Griffith 2017 | 0 | 8 | 0 | 8 | | 0.00 | [-0.21; 0.21] | 0.6% | 0.1% |
| Rodriguez 2013 | 0 | 8 | 0 | 7 | | 0.00 | [-0.22; 0.22] | 0.6% | 0.1% |
| Schiweck 2021 | 0 | 21 | 0 | 7 | | 0.00 | [-0.18; 0.18] | 0.8% | 0.1% |
| Smith-Apeldoorn 2024 | 0 | 49 | 0 | 48 | | 0.00 | [-0.04; 0.04] | 3.5% | 2.8% |
| Sos 2013 | 0 | 11 | 0 | 19 | | 0.00 | [-0.13; 0.13] | 1.1% | 0.2% |
| Su 2017 | 0 | 47 | 0 | 24 | | 0.00 | [-0.06; 0.06] | 2.3% | 1.1% |
| Su 2023 | 0 | 42 | 0 | 42 | | 0.00 | [-0.05; 0.05] | 3.1% | 2.1% |
| Sumner 2019 | 0 | 17 | 0 | 16 | | 0.00 | [-0.11; 0.11] | 1.2% | 0.4% |
| Taylor 2018 | 0 | 9 | 0 | 8 | | 0.00 | [-0.20; 0.20] | 0.7% | 0.1% |
| Wang 2024 | 0 | 182 | 0 | 182 | | 0.00 | [-0.01; 0.01] | 13.1% | 37.4% |
| Xiao 2023 | 0 | 15 | 0 | 15 | | 0.00 | [-0.12; 0.12] | 1.1% | 0.3% |
| Yang 2023 | 0 | 200 | 0 | 100 | | 0.00 | [-0.02; 0.02] | 9.6% | 18.2% |
| Zarate 2006 | 0 | 9 | 0 | 9 | | 0.00 | [-0.19; 0.19] | 0.7% | 0.1% |
| Zarate 2012 | 0 | 21 | 0 | 23 | | 0.00 | [-0.08; 0.08] | 1.6% | 0.6% |
| Zhou 2024 | 0 | 27 | 0 | 25 | | 0.00 | [-0.07; 0.07] | 1.9% | 0.8% |
| **Common effect model** | | **1563** | | **1269** | | **0.00** | **[-0.01; 0.01]** | **100.0%** | **.** |
| **Random effects model** | | | | | | **0.00** | **[-0.01; 0.01]** | **.** | **100.0%** |

Heterogeneity: $I^2 = 0.0\%$, $\tau^2 = 0$, $p = 1.0000$

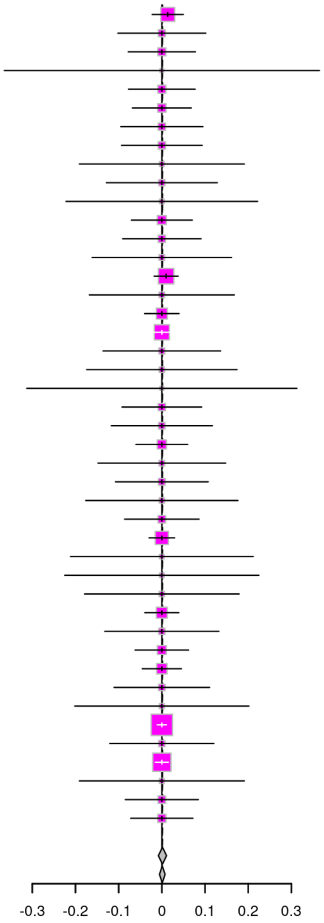

#### Arcsine Risk-Difference

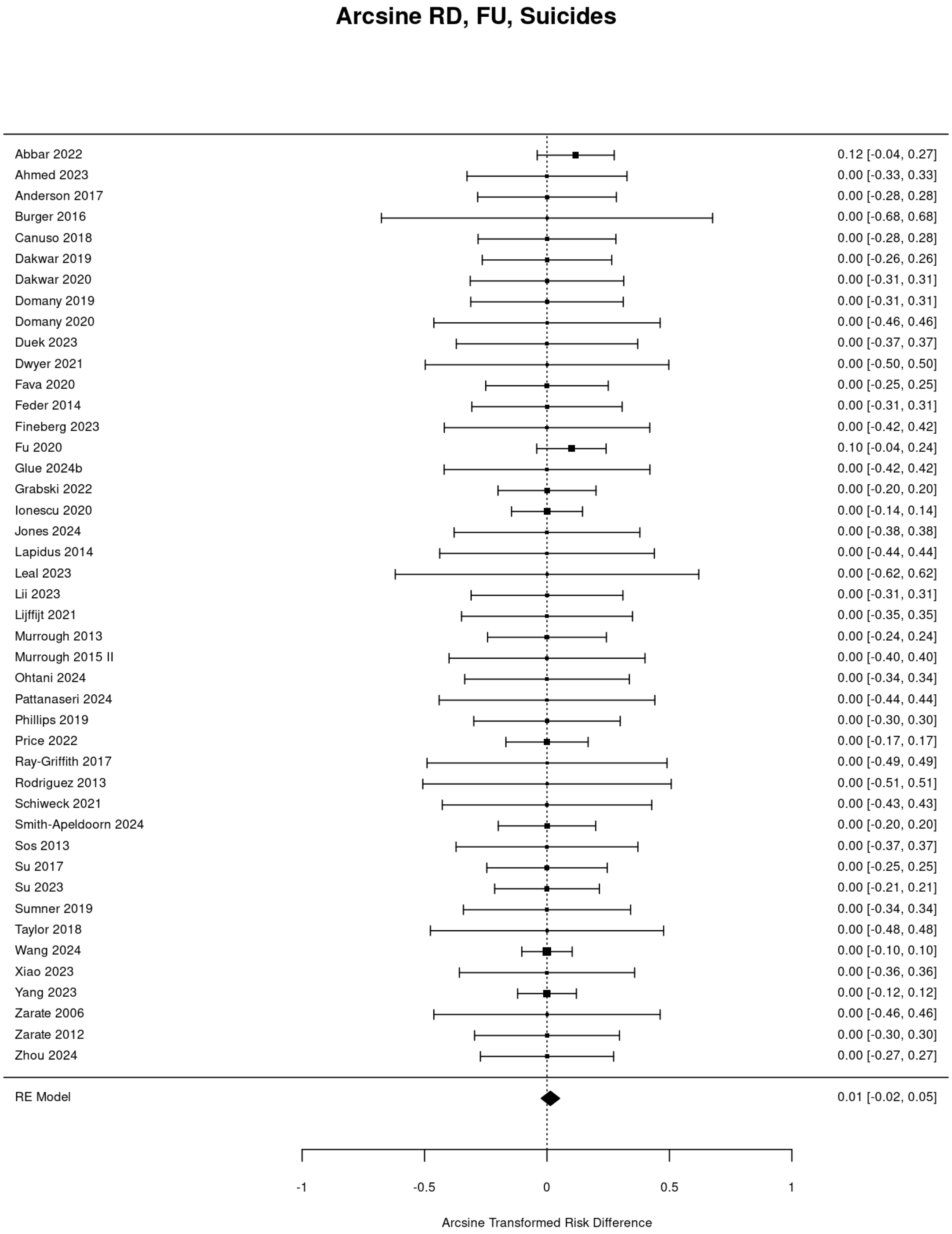

Bayesian Meta-Analysis, wider prior

**Bayesian MA, Delta = 250 - Suicides FU**

| Study | Estimate | 95% CI |
|---|---|---|
| Abbar 2022 | 1.24 | [-1.98, 4.46] |
| Ahmed 2023 | 0.00 | [-3.97, 3.97] |
| Anderson 2017 | -0.08 | [-4.04, 3.88] |
| Burger 2016 | 0.76 | [-3.36, 4.88] |
| Canuso 2018 | -0.20 | [-4.16, 3.76] |
| Dakwar 2019 | 0.04 | [-3.92, 3.99] |
| Dakwar 2020 | 0.29 | [-3.67, 4.26] |
| Domany 2019 | -0.20 | [-4.16, 3.77] |
| Domany 2020 | 0.00 | [-4.02, 4.02] |
| Duek 2023 | 0.00 | [-3.99, 3.99] |
| Dwyer 2021 | -0.57 | [-4.61, 3.47] |
| Fava 2020 | -1.42 | [-5.37, 2.53] |
| Feder 2014 | -0.14 | [-4.11, 3.82] |
| Fineberg 2023 | 0.17 | [-3.83, 4.18] |
| Fu 2020 | 1.00 | [-2.21, 4.22] |
| Glue 2024b | -0.72 | [-4.73, 3.28] |
| Grabski 2022 | 0.00 | [-3.94, 3.94] |
| Ionescu 2020 | 0.05 | [-3.88, 3.98] |
| Jones 2024 | -0.42 | [-4.41, 3.57] |
| Lapidus 2014 | 0.00 | [-4.01, 4.01] |
| Leal 2023 | 0.00 | [-4.09, 4.09] |
| Lii 2023 | 0.00 | [-3.97, 3.97] |
| Lijffijt 2021 | -0.42 | [-4.40, 3.56] |
| Murrough 2013 | -0.62 | [-4.57, 3.33] |
| Murrough 2015 II | 0.00 | [-4.00, 4.00] |
| Ohtani 2024 | 0.00 | [-3.98, 3.98] |
| Pattanaseri 2024 | -0.19 | [-4.20, 3.82] |
| Phillips 2019 | -0.05 | [-4.01, 3.92] |
| Price 2022 | -0.70 | [-4.63, 3.24] |
| Ray-Griffith 2017 | 0.00 | [-4.03, 4.03] |
| Rodriguez 2013 | -0.13 | [-4.17, 3.92] |
| Schiweck 2021 | -1.05 | [-5.06, 2.95] |
| Smith-Apeldoorn 2024 | -0.02 | [-3.96, 3.92] |
| Sos 2013 | 0.53 | [-3.46, 4.52] |
| Su 2017 | -0.66 | [-4.61, 3.29] |
| Su 2023 | 0.00 | [-3.94, 3.94] |
| Sumner 2019 | -0.06 | [-4.04, 3.92] |
| Taylor 2018 | -0.11 | [-4.14, 3.92] |
| Wang 2024 | 0.00 | [-3.93, 3.93] |
| Xiao 2023 | 0.00 | [-3.98, 3.98] |
| Yang 2023 | -0.69 | [-4.62, 3.24] |
| Zarate 2006 | 0.00 | [-4.02, 4.02] |
| Zarate 2012 | 0.09 | [-3.87, 4.05] |
| Zhou 2024 | -0.08 | [-4.03, 3.88] |
| **Summary** | **0.55** | **[-1.05, 2.33]** |
| **Prediction** | **0.55** | **[-1.31, 2.54]** |

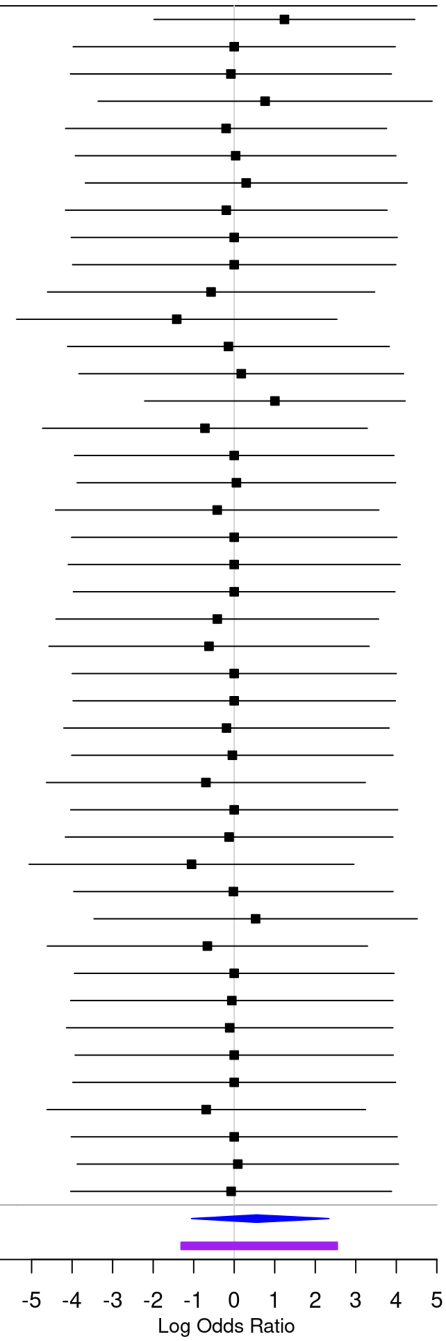

*Heterogeneity (tau): 0.30 [0.00, 0.88]*

**Bayesian Meta-Analysis, smaller prior**

**Bayesian MA, Delta = 15 - Suicides FU**

| Study | Estimate | 95% CI |
|---|---|---|
| Abbar 2022 | 1.24 | [-1.98, 4.46] |
| Ahmed 2023 | 0.00 | [-3.97, 3.97] |
| Anderson 2017 | -0.08 | [-4.04, 3.88] |
| Burger 2016 | 0.76 | [-3.36, 4.88] |
| Canuso 2018 | -0.20 | [-4.16, 3.76] |
| Dakwar 2019 | 0.04 | [-3.92, 3.99] |
| Dakwar 2020 | 0.29 | [-3.67, 4.26] |
| Domany 2019 | -0.20 | [-4.16, 3.77] |
| Domany 2020 | 0.00 | [-4.02, 4.02] |
| Duek 2023 | 0.00 | [-3.99, 3.99] |
| Dwyer 2021 | -0.57 | [-4.61, 3.47] |
| Fava 2020 | -1.42 | [-5.37, 2.53] |
| Feder 2014 | -0.14 | [-4.11, 3.82] |
| Fineberg 2023 | 0.17 | [-3.83, 4.18] |
| Fu 2020 | 1.00 | [-2.21, 4.22] |
| Glue 2024b | -0.72 | [-4.73, 3.28] |
| Grabski 2022 | 0.00 | [-3.94, 3.94] |
| Ionescu 2020 | 0.05 | [-3.88, 3.98] |
| Jones 2024 | -0.42 | [-4.41, 3.57] |
| Lapidus 2014 | 0.00 | [-4.01, 4.01] |
| Leal 2023 | 0.00 | [-4.09, 4.09] |
| Lii 2023 | 0.00 | [-3.97, 3.97] |
| Lijffijt 2021 | -0.42 | [-4.40, 3.56] |
| Murrough 2013 | -0.62 | [-4.57, 3.33] |
| Murrough 2015 II | 0.00 | [-4.00, 4.00] |
| Ohtani 2024 | 0.00 | [-3.98, 3.98] |
| Pattanaseri 2024 | -0.19 | [-4.20, 3.82] |
| Phillips 2019 | -0.05 | [-4.01, 3.92] |
| Price 2022 | -0.70 | [-4.63, 3.24] |
| Ray-Griffith 2017 | 0.00 | [-4.03, 4.03] |
| Rodriguez 2013 | -0.13 | [-4.17, 3.92] |
| Schiweck 2021 | -1.05 | [-5.06, 2.95] |
| Smith-Apeldoorn 2024 | -0.02 | [-3.96, 3.92] |
| Sos 2013 | 0.53 | [-3.46, 4.52] |
| Su 2017 | -0.66 | [-4.61, 3.29] |
| Su 2023 | 0.00 | [-3.94, 3.94] |
| Sumner 2019 | -0.06 | [-4.04, 3.92] |
| Taylor 2018 | -0.11 | [-4.14, 3.92] |
| Wang 2024 | 0.00 | [-3.93, 3.93] |
| Xiao 2023 | 0.00 | [-3.98, 3.98] |
| Yang 2023 | -0.69 | [-4.62, 3.24] |
| Zarate 2006 | 0.00 | [-4.02, 4.02] |
| Zarate 2012 | 0.09 | [-3.87, 4.05] |
| Zhou 2024 | -0.08 | [-4.03, 3.88] |
| **Summary** | **0.42** | **[-0.98, 1.92]** |
| **Prediction** | **0.42** | **[-1.26, 2.15]** |

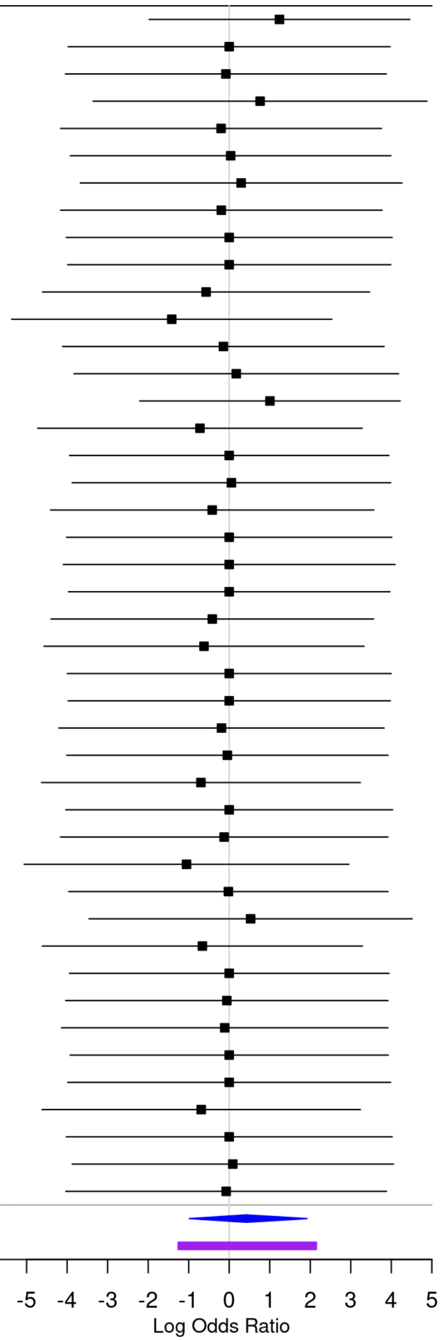

*Heterogeneity (tau): 0.31 [0.00, 0.89]*

**Exact Method** Not possible to calculate because zero events in control arms

#### Treatment + Follow-Up Phase Combined

#### Peto's Method

| Study | Experimental Events | Experimental Total | Control Events | Control Total | OR | 95%-CI | Weight (common) | Weight (random) |
|---|---|---|---|---|---|---|---|---|
| Abbar 2022 | 1 | 73 | 0 | 83 | 8.47 | [0.17; 430.52] | 20.0% | 20.0% |
| Abdallah 2022 | 0 | 108 | 0 | 55 | | | 0.0% | 0.0% |
| Ahmed 2023 | 0 | 18 | 0 | 18 | | | 0.0% | 0.0% |
| Anderson 2017 | 0 | 40 | 0 | 39 | | | 0.0% | 0.0% |
| Burger 2016 | 0 | 3 | 0 | 7 | | | 0.0% | 0.0% |
| Canuso 2018 | 0 | 35 | 0 | 31 | | | 0.0% | 0.0% |
| Chen 2023 | 1 | 126 | 0 | 126 | 7.39 | [0.15; 372.38] | 20.1% | 20.1% |
| Colla 2024 | 0 | 17 | 0 | 10 | | | 0.0% | 0.0% |
| Dakwar 2019 | 0 | 27 | 0 | 28 | | | 0.0% | 0.0% |
| Dakwar 2020 | 0 | 17 | 0 | 23 | | | 0.0% | 0.0% |
| Daly 2018 PA | 0 | 34 | 0 | 33 | | | 0.0% | 0.0% |
| Daly 2018 PB | 0 | 20 | 0 | 21 | | | 0.0% | 0.0% |
| Domany 2019 | 0 | 22 | 0 | 18 | | | 0.0% | 0.0% |
| Domany 2020 | 0 | 9 | 0 | 9 | | | 0.0% | 0.0% |
| Duek 2023 | 0 | 14 | 0 | 14 | | | 0.0% | 0.0% |
| Dwyer 2021 | 0 | 11 | 0 | 6 | | | 0.0% | 0.0% |
| Fava 2020 | 0 | 80 | 0 | 19 | | | 0.0% | 0.0% |
| Feder 2014 | 0 | 22 | 0 | 19 | | | 0.0% | 0.0% |
| Feder 2021 | 0 | 15 | 0 | 15 | | | 0.0% | 0.0% |
| Fedgchin 2019 | 0 | 231 | 0 | 113 | | | 0.0% | 0.0% |
| Fineberg 2023 | 0 | 10 | 0 | 12 | | | 0.0% | 0.0% |
| Fu 2020 | 1 | 113 | 0 | 112 | 7.32 | [0.15; 369.12] | 20.1% | 20.1% |
| George 2017 | 0 | 12 | 0 | 4 | | | 0.0% | 0.0% |
| Glue 2024b | 0 | 17 | 0 | 8 | | | 0.0% | 0.0% |
| Grabski 2022 | 0 | 48 | 0 | 48 | | | 0.0% | 0.0% |
| Grunebaum 2017 | 0 | 9 | 0 | 7 | | | 0.0% | 0.0% |
| Grunebaum 2018 | 0 | 40 | 0 | 40 | | | 0.0% | 0.0% |
| Ionescu 2020 | 0 | 114 | 0 | 113 | | | 0.0% | 0.0% |
| Jones 2024 | 0 | 17 | 0 | 11 | | | 0.0% | 0.0% |
| Lapidus 2014 | 0 | 10 | 0 | 10 | | | 0.0% | 0.0% |
| Leal 2023 | 0 | 5 | 0 | 5 | | | 0.0% | 0.0% |
| Lii 2023 | 0 | 20 | 0 | 20 | | | 0.0% | 0.0% |
| Lijffijt 2021 | 0 | 20 | 0 | 13 | | | 0.0% | 0.0% |
| Loo 2016 | 0 | 9 | 0 | 6 | | | 0.0% | 0.0% |
| Loo 2023 | 0 | 89 | 0 | 92 | | | 0.0% | 0.0% |
| Milak 2020 | 0 | 38 | 0 | 5 | | | 0.0% | 0.0% |
| Murrough 2013 | 0 | 47 | 0 | 25 | | | 0.0% | 0.0% |
| Murrough 2015 II | 0 | 12 | 0 | 12 | | | 0.0% | 0.0% |
| Ochs-Ross 2020 | 0 | 72 | 0 | 65 | | | 0.0% | 0.0% |
| Ohtani 2024 | 0 | 17 | 0 | 17 | | | 0.0% | 0.0% |
| Pattanaseri 2024 | 0 | 11 | 0 | 9 | | | 0.0% | 0.0% |
| Phillips 2019 | 0 | 22 | 0 | 21 | | | 0.0% | 0.0% |
| Popova 2019 | 0 | 115 | 0 | 109 | | | 0.0% | 0.0% |
| Price 2022 | 0 | 103 | 0 | 51 | | | 0.0% | 0.0% |
| Ray-Griffith 2017 | 0 | 8 | 0 | 8 | | | 0.0% | 0.0% |
| Rodriguez 2013 | 0 | 8 | 0 | 7 | | | 0.0% | 0.0% |
| Schiweck 2021 | 0 | 21 | 0 | 7 | | | 0.0% | 0.0% |
| Shiroma 2020 | 0 | 29 | 0 | 25 | | | 0.0% | 0.0% |
| Singh 2016 2x doses | 0 | 18 | 0 | 16 | | | 0.0% | 0.0% |
| Singh 2016 3x doses | 0 | 17 | 0 | 16 | | | 0.0% | 0.0% |
| Singh 2016 II | 0 | 20 | 0 | 10 | | | 0.0% | 0.0% |
| Smith-Apeldoorn 2024 | 1 | 57 | 1 | 54 | 0.95 | [0.06; 15.35] | 39.8% | 39.8% |
| Sos 2013 | 0 | 11 | 0 | 19 | | | 0.0% | 0.0% |
| Su 2017 | 0 | 47 | 0 | 24 | | | 0.0% | 0.0% |
| Su 2023 | 0 | 42 | 0 | 42 | | | 0.0% | 0.0% |
| Sumner 2019 | 0 | 17 | 0 | 16 | | | 0.0% | 0.0% |
| Takahashi 2021 | 0 | 122 | 0 | 80 | | | 0.0% | 0.0% |
| Taylor 2018 | 0 | 9 | 0 | 9 | | | 0.0% | 0.0% |
| Tiger 2020 | 0 | 20 | 0 | 10 | | | 0.0% | 0.0% |
| Wang 2024 | 0 | 182 | 0 | 182 | | | 0.0% | 0.0% |
| Xiao 2023 | 0 | 15 | 0 | 15 | | | 0.0% | 0.0% |
| Yang 2023 | 0 | 200 | 0 | 100 | | | 0.0% | 0.0% |
| Zarate 2006 | 0 | 9 | 0 | 9 | | | 0.0% | 0.0% |
| Zarate 2012 | 0 | 21 | 0 | 23 | | | 0.0% | 0.0% |
| Zhou 2024 | 0 | 27 | 0 | 27 | | | 0.0% | 0.0% |
| **Common effect model** | | **2792** | | **2201** | **3.35** | **[0.58; 19.40]** | **100.0%** | **.** |
| **Random effects model** | | | | | **3.35** | **[0.58; 19.40]** | **.** | **100.0%** |

Heterogeneity: $I^2 = 0.0\%$, $\tau^2 = 0$, $p = 0.7258$

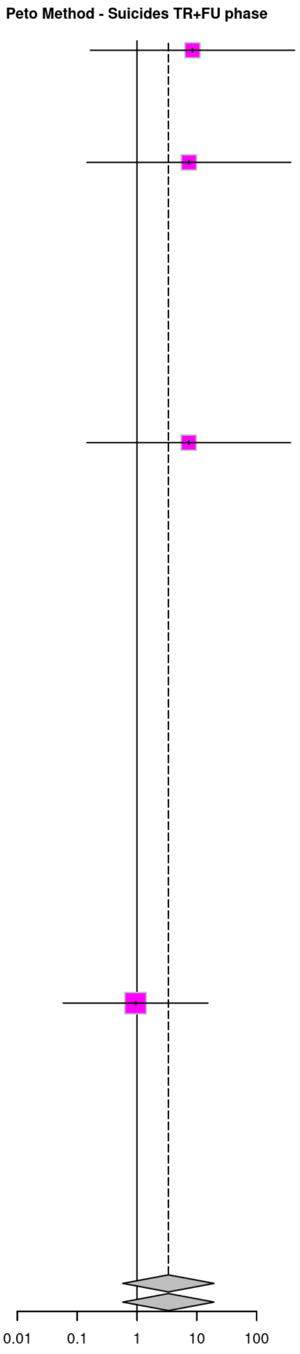

#### Maentel Mantel-Haenszel with CC

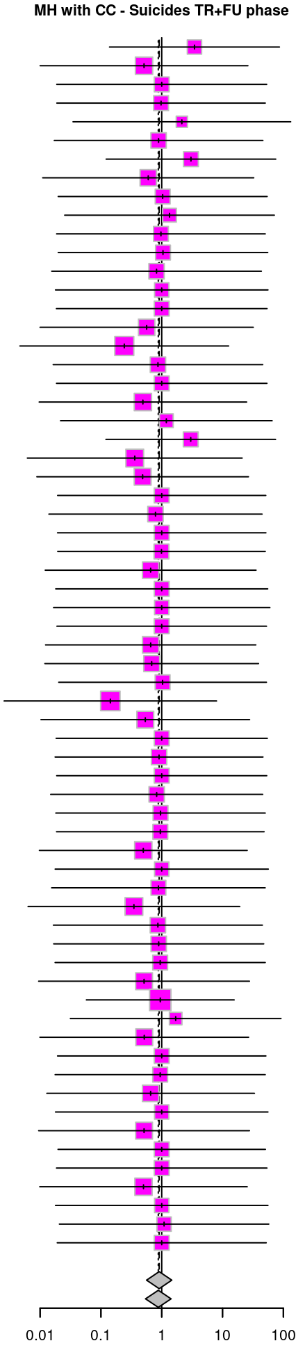

| Study | Experimental Events | Experimental Total | Control Events | Control Total | OR | 95%-CI | Weight (common) | Weight (random) |
|---|---|---|---|---|---|---|---|---|
| Abbar 2022 | 1 | 73 | 0 | 83 | 3.46 | [0.14; 86.13] | 1.3% | 2.3% |
| Abdallah 2022 | 0 | 108 | 0 | 55 | 0.51 | [0.01; 26.12] | 1.9% | 1.5% |
| Ahmed 2023 | 0 | 18 | 0 | 18 | 1.00 | [0.02; 53.12] | 1.4% | 1.5% |
| Anderson 2017 | 0 | 40 | 0 | 39 | 0.98 | [0.02; 50.37] | 1.4% | 1.5% |
| Burger 2016 | 0 | 3 | 0 | 7 | 2.14 | [0.03; 131.93] | 0.8% | 1.4% |
| Canuso 2018 | 0 | 35 | 0 | 31 | 0.89 | [0.02; 46.05] | 1.5% | 1.5% |
| Chen 2023 | 1 | 126 | 0 | 126 | 3.02 | [0.12; 74.94] | 1.4% | 2.3% |
| Colla 2024 | 0 | 17 | 0 | 10 | 0.60 | [0.01; 32.56] | 1.7% | 1.5% |
| Dakwar 2019 | 0 | 27 | 0 | 28 | 1.04 | [0.02; 54.08] | 1.4% | 1.5% |
| Dakwar 2020 | 0 | 17 | 0 | 23 | 1.34 | [0.03; 71.04] | 1.2% | 1.5% |
| Daly 2018 PA | 0 | 34 | 0 | 33 | 0.97 | [0.02; 50.36] | 1.4% | 1.5% |
| Daly 2018 PB | 0 | 20 | 0 | 21 | 1.05 | [0.02; 55.37] | 1.4% | 1.5% |
| Domany 2019 | 0 | 22 | 0 | 18 | 0.82 | [0.02; 43.47] | 1.5% | 1.5% |
| Domany 2020 | 0 | 9 | 0 | 9 | 1.00 | [0.02; 55.80] | 1.3% | 1.4% |
| Duek 2023 | 0 | 14 | 0 | 14 | 1.00 | [0.02; 53.89] | 1.4% | 1.5% |
| Dwyer 2021 | 0 | 11 | 0 | 6 | 0.57 | [0.01; 32.00] | 1.7% | 1.4% |
| Fava 2020 | 0 | 80 | 0 | 19 | 0.24 | [0.00; 12.59] | 2.3% | 1.5% |
| Feder 2014 | 0 | 22 | 0 | 19 | 0.87 | [0.02; 45.76] | 1.5% | 1.5% |
| Feder 2021 | 0 | 15 | 0 | 15 | 1.00 | [0.02; 53.66] | 1.4% | 1.5% |
| Fedgchin 2019 | 0 | 231 | 0 | 113 | 0.49 | [0.01; 24.87] | 1.9% | 1.5% |
| Fineberg 2023 | 0 | 10 | 0 | 12 | 1.19 | [0.02; 65.32] | 1.2% | 1.5% |
| Fu 2020 | 1 | 113 | 0 | 112 | 3.00 | [0.12; 74.43] | 1.4% | 2.3% |
| George 2017 | 0 | 12 | 0 | 4 | 0.36 | [0.01; 20.98] | 2.0% | 1.4% |
| Glue 2024b | 0 | 17 | 0 | 8 | 0.49 | [0.01; 26.64] | 1.8% | 1.5% |
| Grabski 2022 | 0 | 48 | 0 | 48 | 1.00 | [0.02; 51.42] | 1.4% | 1.5% |
| Grunebaum 2017 | 0 | 9 | 0 | 7 | 0.79 | [0.01; 44.65] | 1.5% | 1.4% |
| Grunebaum 2018 | 0 | 40 | 0 | 40 | 1.00 | [0.02; 51.63] | 1.4% | 1.5% |
| Ionescu 2020 | 0 | 114 | 0 | 113 | 0.99 | [0.02; 50.39] | 1.4% | 1.5% |
| Jones 2024 | 0 | 17 | 0 | 11 | 0.66 | [0.01; 35.52] | 1.7% | 1.5% |
| Lapidus 2014 | 0 | 10 | 0 | 10 | 1.00 | [0.02; 55.27] | 1.4% | 1.5% |
| Leal 2023 | 0 | 5 | 0 | 5 | 1.00 | [0.02; 59.99] | 1.3% | 1.4% |
| Lii 2023 | 0 | 20 | 0 | 20 | 1.00 | [0.02; 52.85] | 1.4% | 1.5% |
| Lijffijt 2021 | 0 | 20 | 0 | 13 | 0.66 | [0.01; 35.23] | 1.7% | 1.5% |
| Loo 2016 | 0 | 9 | 0 | 6 | 0.68 | [0.01; 39.07] | 1.6% | 1.4% |
| Loo 2023 | 0 | 89 | 0 | 92 | 1.03 | [0.02; 52.65] | 1.4% | 1.5% |
| Milak 2020 | 0 | 38 | 0 | 5 | 0.14 | [0.00; 7.96] | 2.4% | 1.4% |
| Murrough 2013 | 0 | 47 | 0 | 25 | 0.54 | [0.01; 27.86] | 1.8% | 1.5% |
| Murrough 2015 II | 0 | 12 | 0 | 12 | 1.00 | [0.02; 54.46] | 1.4% | 1.5% |
| Ochs-Ross 2020 | 0 | 72 | 0 | 65 | 0.90 | [0.02; 46.18] | 1.5% | 1.5% |
| Ohtani 2024 | 0 | 17 | 0 | 17 | 1.00 | [0.02; 53.28] | 1.4% | 1.5% |
| Pattanaseri 2024 | 0 | 11 | 0 | 9 | 0.83 | [0.01; 45.69] | 1.5% | 1.5% |
| Phillips 2019 | 0 | 22 | 0 | 21 | 0.96 | [0.02; 50.34] | 1.4% | 1.5% |
| Popova 2019 | 0 | 115 | 0 | 109 | 0.95 | [0.02; 48.20] | 1.4% | 1.5% |
| Price 2022 | 0 | 103 | 0 | 51 | 0.50 | [0.01; 25.44] | 1.9% | 1.5% |
| Ray-Griffith 2017 | 0 | 8 | 0 | 8 | 1.00 | [0.02; 56.46] | 1.3% | 1.4% |
| Rodriguez 2013 | 0 | 8 | 0 | 7 | 0.88 | [0.02; 50.19] | 1.4% | 1.4% |
| Schiweck 2021 | 0 | 21 | 0 | 7 | 0.35 | [0.01; 19.18] | 2.0% | 1.5% |
| Shiroma 2020 | 0 | 29 | 0 | 25 | 0.86 | [0.02; 45.14] | 1.5% | 1.5% |
| Singh 2016 2x doses | 0 | 18 | 0 | 16 | 0.89 | [0.02; 47.53] | 1.5% | 1.5% |
| Singh 2016 3x doses | 0 | 17 | 0 | 16 | 0.94 | [0.02; 50.32] | 1.4% | 1.5% |
| Singh 2016 II | 0 | 20 | 0 | 10 | 0.51 | [0.01; 27.68] | 1.8% | 1.5% |
| Smith-Apeldoorn 2024 | 1 | 57 | 1 | 54 | 0.95 | [0.06; 15.52] | 2.9% | 3.0% |
| Sos 2013 | 0 | 11 | 0 | 19 | 1.70 | [0.03; 91.39] | 1.0% | 1.5% |
| Su 2017 | 0 | 47 | 0 | 24 | 0.52 | [0.01; 26.79] | 1.8% | 1.5% |
| Su 2023 | 0 | 42 | 0 | 42 | 1.00 | [0.02; 51.57] | 1.4% | 1.5% |
| Sumner 2019 | 0 | 17 | 0 | 16 | 0.94 | [0.02; 50.32] | 1.4% | 1.5% |
| Takahashi 2021 | 0 | 122 | 0 | 80 | 0.66 | [0.01; 33.45] | 1.7% | 1.5% |
| Taylor 2018 | 0 | 9 | 0 | 9 | 1.00 | [0.02; 55.80] | 1.3% | 1.4% |
| Tiger 2020 | 0 | 20 | 0 | 10 | 0.51 | [0.01; 27.68] | 1.8% | 1.5% |
| Wang 2024 | 0 | 182 | 0 | 182 | 1.00 | [0.02; 50.67] | 1.4% | 1.5% |
| Xiao 2023 | 0 | 15 | 0 | 15 | 1.00 | [0.02; 53.66] | 1.4% | 1.5% |
| Yang 2023 | 0 | 200 | 0 | 100 | 0.50 | [0.01; 25.45] | 1.9% | 1.5% |
| Zarate 2006 | 0 | 9 | 0 | 9 | 1.00 | [0.02; 55.80] | 1.3% | 1.4% |
| Zarate 2012 | 0 | 21 | 0 | 23 | 1.09 | [0.02; 57.53] | 1.3% | 1.5% |
| Zhou 2024 | 0 | 27 | 0 | 27 | 1.00 | [0.02; 52.22] | 1.4% | 1.5% |
| **Common effect model** | | **2792** | | **2201** | **0.91** | **[0.56; 1.46]** | **100.0%** | **.** |
| **Random effects model** | | | | | **0.88** | **[0.54; 1.42]** | **.** | **100.0%** |

Heterogeneity: $I^2 = 0.0\%$, $\tau^2 = 0$, $p = 1.0000$

#### Maentel Mantel-Haenszel with Treatment-Arm CC

| Study | Experimental Events | Experimental Total | Control Events | Control Total | OR | 95%-CI | Weight (common) | Weight (random) |
|---|---|---|---|---|---|---|---|---|
| Abbar 2022 | 1 | 73 | 0 | 83 | 3.18 | [0.14; 74.37] | 1.6% | 2.5% |
| Abdallah 2022 | 0 | 108 | 0 | 55 | 1.00 | [0.02; 63.93] | 1.5% | 1.4% |
| Ahmed 2023 | 0 | 18 | 0 | 18 | 1.00 | [0.02; 53.12] | 1.6% | 1.6% |
| Anderson 2017 | 0 | 40 | 0 | 39 | 1.00 | [0.02; 51.66] | 1.6% | 1.6% |
| Burger 2016 | 0 | 3 | 0 | 7 | 1.00 | [0.01; 87.11] | 1.3% | 1.2% |
| Canuso 2018 | 0 | 35 | 0 | 31 | 1.00 | [0.02; 52.27] | 1.6% | 1.6% |
| Chen 2023 | 1 | 126 | 0 | 126 | 3.02 | [0.12; 74.94] | 1.6% | 2.4% |
| Colla 2024 | 0 | 17 | 0 | 10 | 1.00 | [0.02; 62.21] | 1.5% | 1.4% |
| Dakwar 2019 | 0 | 27 | 0 | 28 | 1.00 | [0.02; 52.22] | 1.6% | 1.6% |
| Dakwar 2020 | 0 | 17 | 0 | 23 | 1.00 | [0.02; 55.30] | 1.6% | 1.5% |
| Daly 2018 PA | 0 | 34 | 0 | 33 | 1.00 | [0.02; 51.89] | 1.6% | 1.6% |
| Daly 2018 PB | 0 | 20 | 0 | 21 | 1.00 | [0.02; 52.85] | 1.6% | 1.6% |
| Domany 2019 | 0 | 22 | 0 | 18 | 1.00 | [0.02; 53.92] | 1.6% | 1.5% |
| Domany 2020 | 0 | 9 | 0 | 9 | 1.00 | [0.02; 55.80] | 1.6% | 1.5% |
| Duek 2023 | 0 | 14 | 0 | 14 | 1.00 | [0.02; 53.89] | 1.6% | 1.5% |
| Dwyer 2021 | 0 | 11 | 0 | 6 | 1.00 | [0.01; 67.61] | 1.4% | 1.4% |
| Fava 2020 | 0 | 80 | 0 | 19 | 1.00 | [0.01; 148.67] | 1.0% | 1.0% |
| Feder 2014 | 0 | 22 | 0 | 19 | 1.00 | [0.02; 53.36] | 1.6% | 1.5% |
| Feder 2021 | 0 | 15 | 0 | 15 | 1.00 | [0.02; 53.66] | 1.6% | 1.5% |
| Fedgchin 2019 | 0 | 231 | 0 | 113 | 1.00 | [0.02; 65.31] | 1.4% | 1.4% |
| Fineberg 2023 | 0 | 10 | 0 | 12 | 1.00 | [0.02; 55.75] | 1.6% | 1.5% |
| Fu 2020 | 1 | 113 | 0 | 112 | 3.02 | [0.12; 75.23] | 1.6% | 2.4% |
| George 2017 | 0 | 12 | 0 | 4 | 1.00 | [0.01; 105.38] | 1.2% | 1.1% |
| Glue 2024b | 0 | 17 | 0 | 8 | 1.00 | [0.01; 72.36] | 1.4% | 1.3% |
| Grabski 2022 | 0 | 48 | 0 | 48 | 1.00 | [0.02; 51.42] | 1.6% | 1.6% |
| Grunebaum 2017 | 0 | 9 | 0 | 7 | 1.00 | [0.02; 58.29] | 1.5% | 1.5% |
| Grunebaum 2018 | 0 | 40 | 0 | 40 | 1.00 | [0.02; 51.63] | 1.6% | 1.6% |
| Ionescu 2020 | 0 | 114 | 0 | 113 | 1.00 | [0.02; 50.83] | 1.6% | 1.6% |
| Jones 2024 | 0 | 17 | 0 | 11 | 1.00 | [0.02; 59.25] | 1.5% | 1.5% |
| Lapidus 2014 | 0 | 10 | 0 | 10 | 1.00 | [0.02; 55.27] | 1.6% | 1.5% |
| Leal 2023 | 0 | 5 | 0 | 5 | 1.00 | [0.02; 59.99] | 1.5% | 1.5% |
| Lii 2023 | 0 | 20 | 0 | 20 | 1.00 | [0.02; 52.85] | 1.6% | 1.6% |
| Lijffijt 2021 | 0 | 20 | 0 | 13 | 1.00 | [0.02; 58.54] | 1.5% | 1.5% |
| Loo 2016 | 0 | 9 | 0 | 6 | 1.00 | [0.02; 61.80] | 1.5% | 1.4% |
| Loo 2023 | 0 | 89 | 0 | 92 | 1.00 | [0.02; 50.97] | 1.6% | 1.6% |
| Milak 2020 | 0 | 38 | 0 | 5 | 1.00 | [0.00; 484.58] | 0.7% | 0.6% |
| Murrough 2013 | 0 | 47 | 0 | 25 | 1.00 | [0.02; 63.11] | 1.5% | 1.4% |
| Murrough 2015 II | 0 | 12 | 0 | 12 | 1.00 | [0.02; 54.46] | 1.6% | 1.5% |
| Ochs-Ross 2020 | 0 | 72 | 0 | 65 | 1.00 | [0.02; 51.38] | 1.6% | 1.6% |
| Ohtani 2024 | 0 | 17 | 0 | 17 | 1.00 | [0.02; 53.28] | 1.6% | 1.5% |
| Pattanaseri 2024 | 0 | 11 | 0 | 9 | 1.00 | [0.02; 56.40] | 1.5% | 1.5% |
| Phillips 2019 | 0 | 22 | 0 | 21 | 1.00 | [0.02; 52.74] | 1.6% | 1.6% |
| Popova 2019 | 0 | 115 | 0 | 109 | 1.00 | [0.02; 50.91] | 1.6% | 1.6% |
| Price 2022 | 0 | 103 | 0 | 51 | 1.00 | [0.02; 65.23] | 1.4% | 1.4% |
| Ray-Griffith 2017 | 0 | 8 | 0 | 8 | 1.00 | [0.02; 56.46] | 1.5% | 1.5% |
| Rodriguez 2013 | 0 | 8 | 0 | 7 | 1.00 | [0.02; 57.37] | 1.5% | 1.5% |
| Schiweck 2021 | 0 | 21 | 0 | 7 | 1.00 | [0.01; 99.86] | 1.2% | 1.2% |
| Shiroma 2020 | 0 | 29 | 0 | 25 | 1.00 | [0.02; 52.79] | 1.6% | 1.6% |
| Singh 2016 2x doses | 0 | 18 | 0 | 16 | 1.00 | [0.02; 53.65] | 1.6% | 1.5% |
| Singh 2016 3x doses | 0 | 17 | 0 | 16 | 1.00 | [0.02; 53.46] | 1.6% | 1.5% |
| Singh 2016 II | 0 | 20 | 0 | 10 | 1.00 | [0.01; 68.32] | 1.4% | 1.4% |
| Smith-Apeldoorn 2024 | 1 | 57 | 1 | 54 | 0.95 | [0.06; 15.52] | 3.3% | 3.1% |
| Sos 2013 | 0 | 11 | 0 | 19 | 1.00 | [0.02; 62.32] | 1.5% | 1.4% |
| Su 2017 | 0 | 47 | 0 | 24 | 1.00 | [0.02; 64.85] | 1.4% | 1.4% |
| Su 2023 | 0 | 42 | 0 | 42 | 1.00 | [0.02; 51.57] | 1.6% | 1.6% |
| Sumner 2019 | 0 | 17 | 0 | 16 | 1.00 | [0.02; 53.46] | 1.6% | 1.5% |
| Takahashi 2021 | 0 | 122 | 0 | 80 | 1.00 | [0.02; 55.55] | 1.6% | 1.5% |
| Taylor 2018 | 0 | 9 | 0 | 9 | 1.00 | [0.02; 55.80] | 1.6% | 1.5% |
| Tiger 2020 | 0 | 20 | 0 | 10 | 1.00 | [0.01; 68.32] | 1.4% | 1.4% |
| Wang 2024 | 0 | 182 | 0 | 182 | 1.00 | [0.02; 50.67] | 1.6% | 1.6% |
| Xiao 2023 | 0 | 15 | 0 | 15 | 1.00 | [0.02; 53.66] | 1.6% | 1.5% |
| Yang 2023 | 0 | 200 | 0 | 100 | 1.00 | [0.02; 64.37] | 1.4% | 1.4% |
| Zarate 2006 | 0 | 9 | 0 | 9 | 1.00 | [0.02; 55.80] | 1.6% | 1.5% |
| Zarate 2012 | 0 | 21 | 0 | 23 | 1.00 | [0.02; 52.84] | 1.6% | 1.6% |
| Zhou 2024 | 0 | 27 | 0 | 27 | 1.00 | [0.02; 52.22] | 1.6% | 1.6% |
| **Common effect model** | | **2792** | | **2201** | **1.10** | **[0.67; 1.79]** | **100.0%** | **.** |
| **Random effects model** | | | | | **1.08** | **[0.66; 1.78]** | **.** | **100.0%** |

Heterogeneity: $I^2 = 0.0\%$, $\tau^2 = 0$, $p = 1.0000$

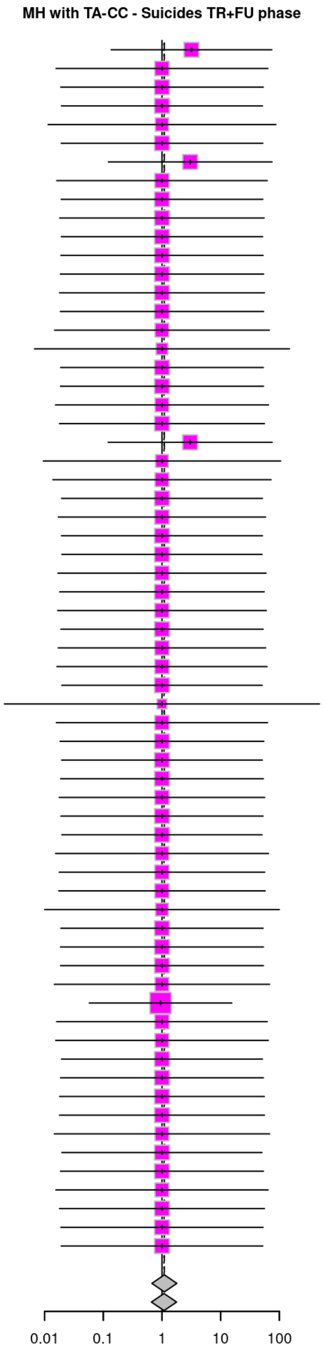

#### Maentel Mantel-Haenszel Risk-Difference

| Study | Experimental Events | Experimental Total | Control Events | Control Total | RD | 95%-CI | Weight (common) | Weight (random) |
|---|---|---|---|---|---|---|---|---|
| Abbar 2022 | 1 | 73 | 0 | 83 | 0.01 | [-0.02; 0.05] | 3.2% | 1.7% |
| Abdallah 2022 | 0 | 108 | 0 | 55 | 0.00 | [-0.03; 0.03] | 3.0% | 2.9% |
| Ahmed 2023 | 0 | 18 | 0 | 18 | 0.00 | [-0.10; 0.10] | 0.8% | 0.2% |
| Anderson 2017 | 0 | 40 | 0 | 39 | 0.00 | [-0.05; 0.05] | 1.7% | 1.0% |
| Burger 2016 | 0 | 3 | 0 | 7 | 0.00 | [-0.36; 0.36] | 0.2% | 0.0% |
| Canuso 2018 | 0 | 35 | 0 | 31 | 0.00 | [-0.06; 0.06] | 1.4% | 0.7% |
| Chen 2023 | 1 | 126 | 0 | 126 | 0.01 | [-0.01; 0.03] | 5.2% | 4.7% |
| Colla 2024 | 0 | 17 | 0 | 10 | 0.00 | [-0.14; 0.14] | 0.6% | 0.1% |
| Dakwar 2019 | 0 | 27 | 0 | 28 | 0.00 | [-0.07; 0.07] | 1.2% | 0.5% |
| Dakwar 2020 | 0 | 17 | 0 | 23 | 0.00 | [-0.10; 0.10] | 0.8% | 0.2% |
| Daly 2018 PA | 0 | 34 | 0 | 33 | 0.00 | [-0.06; 0.06] | 1.4% | 0.7% |
| Daly 2018 PB | 0 | 20 | 0 | 21 | 0.00 | [-0.09; 0.09] | 0.9% | 0.3% |
| Domany 2019 | 0 | 22 | 0 | 18 | 0.00 | [-0.09; 0.09] | 0.8% | 0.3% |
| Domany 2020 | 0 | 9 | 0 | 9 | 0.00 | [-0.19; 0.19] | 0.4% | 0.1% |
| Duek 2023 | 0 | 14 | 0 | 14 | 0.00 | [-0.13; 0.13] | 0.6% | 0.1% |
| Dwyer 2021 | 0 | 11 | 0 | 6 | 0.00 | [-0.22; 0.22] | 0.4% | 0.0% |
| Fava 2020 | 0 | 80 | 0 | 19 | 0.00 | [-0.07; 0.07] | 1.3% | 0.4% |
| Feder 2014 | 0 | 22 | 0 | 19 | 0.00 | [-0.09; 0.09] | 0.9% | 0.3% |
| Feder 2021 | 0 | 15 | 0 | 15 | 0.00 | [-0.12; 0.12] | 0.7% | 0.2% |
| Fedgchin 2019 | 0 | 231 | 0 | 113 | 0.00 | [-0.01; 0.01] | 6.2% | 12.2% |
| Fineberg 2023 | 0 | 10 | 0 | 12 | 0.00 | [-0.16; 0.16] | 0.5% | 0.1% |
| Fu 2020 | 1 | 113 | 0 | 112 | 0.01 | [-0.02; 0.03] | 4.6% | 3.8% |
| George 2017 | 0 | 12 | 0 | 4 | 0.00 | [-0.28; 0.28] | 0.3% | 0.0% |
| Glue 2024b | 0 | 17 | 0 | 8 | 0.00 | [-0.17; 0.17] | 0.5% | 0.1% |
| Grabski 2022 | 0 | 48 | 0 | 48 | 0.00 | [-0.04; 0.04] | 2.0% | 1.4% |
| Grunebaum 2017 | 0 | 9 | 0 | 7 | 0.00 | [-0.22; 0.22] | 0.4% | 0.0% |
| Grunebaum 2018 | 0 | 40 | 0 | 40 | 0.00 | [-0.05; 0.05] | 1.7% | 1.0% |
| Ionescu 2020 | 0 | 114 | 0 | 113 | 0.00 | [-0.02; 0.02] | 4.7% | 7.7% |
| Jones 2024 | 0 | 17 | 0 | 11 | 0.00 | [-0.14; 0.14] | 0.6% | 0.1% |
| Lapidus 2014 | 0 | 10 | 0 | 10 | 0.00 | [-0.17; 0.17] | 0.4% | 0.1% |
| Leal 2023 | 0 | 5 | 0 | 5 | 0.00 | [-0.31; 0.31] | 0.2% | 0.0% |
| Lii 2023 | 0 | 20 | 0 | 20 | 0.00 | [-0.09; 0.09] | 0.9% | 0.3% |
| Lijffijt 2021 | 0 | 20 | 0 | 13 | 0.00 | [-0.12; 0.12] | 0.7% | 0.2% |
| Loo 2016 | 0 | 9 | 0 | 6 | 0.00 | [-0.23; 0.23] | 0.3% | 0.0% |
| Loo 2023 | 0 | 89 | 0 | 92 | 0.00 | [-0.02; 0.02] | 3.7% | 4.9% |
| Milak 2020 | 0 | 38 | 0 | 5 | 0.00 | [-0.22; 0.22] | 0.4% | 0.0% |
| Murrough 2013 | 0 | 47 | 0 | 25 | 0.00 | [-0.06; 0.06] | 1.4% | 0.6% |
| Murrough 2015 II | 0 | 12 | 0 | 12 | 0.00 | [-0.15; 0.15] | 0.5% | 0.1% |
| Ochs-Ross 2020 | 0 | 72 | 0 | 65 | 0.00 | [-0.03; 0.03] | 2.8% | 2.8% |
| Ohtani 2024 | 0 | 17 | 0 | 17 | 0.00 | [-0.11; 0.11] | 0.7% | 0.2% |
| Pattanaseri 2024 | 0 | 11 | 0 | 9 | 0.00 | [-0.18; 0.18] | 0.4% | 0.1% |
| Phillips 2019 | 0 | 22 | 0 | 21 | 0.00 | [-0.09; 0.09] | 0.9% | 0.3% |
| Popova 2019 | 0 | 115 | 0 | 109 | 0.00 | [-0.02; 0.02] | 4.6% | 7.4% |
| Price 2022 | 0 | 103 | 0 | 51 | 0.00 | [-0.03; 0.03] | 2.8% | 2.5% |
| Ray-Griffith 2017 | 0 | 8 | 0 | 8 | 0.00 | [-0.21; 0.21] | 0.4% | 0.0% |
| Rodriguez 2013 | 0 | 8 | 0 | 7 | 0.00 | [-0.22; 0.22] | 0.3% | 0.0% |
| Schiweck 2021 | 0 | 21 | 0 | 7 | 0.00 | [-0.18; 0.18] | 0.5% | 0.1% |
| Shiroma 2020 | 0 | 29 | 0 | 25 | 0.00 | [-0.07; 0.07] | 1.1% | 0.5% |
| Singh 2016 2x doses | 0 | 18 | 0 | 16 | 0.00 | [-0.11; 0.11] | 0.7% | 0.2% |
| Singh 2016 3x doses | 0 | 17 | 0 | 16 | 0.00 | [-0.11; 0.11] | 0.7% | 0.2% |
| Singh 2016 II | 0 | 20 | 0 | 10 | 0.00 | [-0.14; 0.14] | 0.6% | 0.1% |
| Smith-Apeldoorn 2024 | 1 | 57 | 1 | 54 | -0.00 | [-0.05; 0.05] | 2.3% | 0.9% |
| Sos 2013 | 0 | 11 | 0 | 19 | 0.00 | [-0.13; 0.13] | 0.6% | 0.1% |
| Su 2017 | 0 | 47 | 0 | 24 | 0.00 | [-0.06; 0.06] | 1.3% | 0.6% |
| Su 2023 | 0 | 42 | 0 | 42 | 0.00 | [-0.05; 0.05] | 1.8% | 1.1% |
| Sumner 2019 | 0 | 17 | 0 | 16 | 0.00 | [-0.11; 0.11] | 0.7% | 0.2% |
| Takahashi 2021 | 0 | 122 | 0 | 80 | 0.00 | [-0.02; 0.02] | 4.0% | 5.4% |
| Taylor 2018 | 0 | 9 | 0 | 9 | 0.00 | [-0.19; 0.19] | 0.4% | 0.1% |
| Tiger 2020 | 0 | 20 | 0 | 10 | 0.00 | [-0.14; 0.14] | 0.6% | 0.1% |
| Wang 2024 | 0 | 182 | 0 | 182 | 0.00 | [-0.01; 0.01] | 7.5% | 19.5% |
| Xiao 2023 | 0 | 15 | 0 | 15 | 0.00 | [-0.12; 0.12] | 0.7% | 0.2% |
| Yang 2023 | 0 | 200 | 0 | 100 | 0.00 | [-0.02; 0.02] | 5.5% | 9.5% |
| Zarate 2006 | 0 | 9 | 0 | 9 | 0.00 | [-0.19; 0.19] | 0.4% | 0.1% |
| Zarate 2012 | 0 | 21 | 0 | 23 | 0.00 | [-0.08; 0.08] | 0.9% | 0.3% |
| Zhou 2024 | 0 | 27 | 0 | 27 | 0.00 | [-0.07; 0.07] | 1.1% | 0.5% |
| **Common effect model** | | **2792** | | **2201** | **0.00** | **[-0.01; 0.01]** | **100.0%** | **.** |
| **Random effects model** | | | | | **0.00** | **[-0.00; 0.01]** | **.** | **100.0%** |

Heterogeneity: $I^2 = 0.0\%$, $\tau^2 = 0$, $p = 1.0000$

**MH RD - Suicides TR+FU phase**

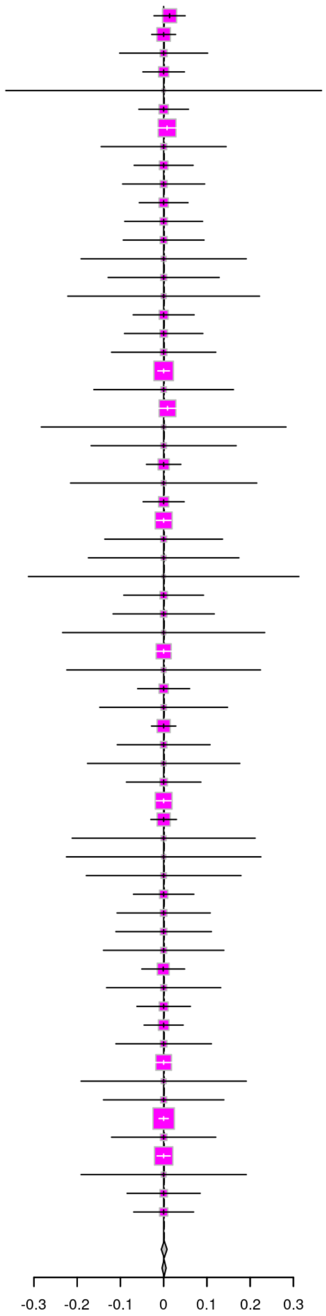

#### Arcsine Risk-Difference

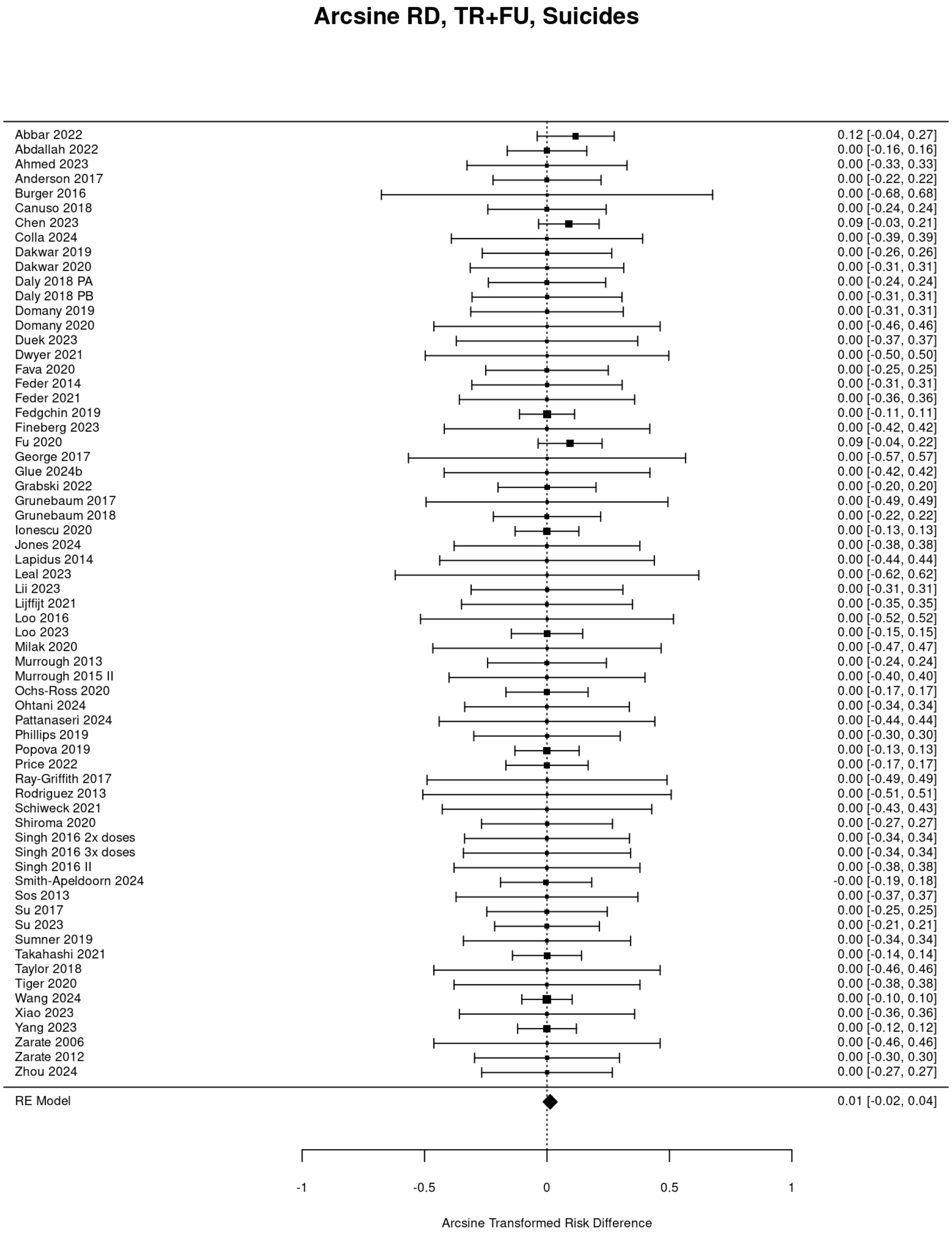

**Bayesian Meta-Analysis, wider prior**

**Bayesian MA, Delta = 250 - Suicides TR+FU**

| Study | Estimate | 95% CI |
|---|---|---|
| Abbar 2022 | 1.24 | [-1.98, 4.46] |
| Abdallah 2022 | -0.67 | [-4.60, 3.26] |
| Ahmed 2023 | 0.00 | [-3.97, 3.97] |
| Anderson 2017 | -0.03 | [-3.97, 3.92] |
| Burger 2016 | 0.76 | [-3.36, 4.88] |
| Canuso 2018 | -0.12 | [-4.07, 3.83] |
| Chen 2023 | 1.11 | [-2.10, 4.32] |
| Colla 2024 | -0.51 | [-4.50, 3.48] |
| Dakwar 2019 | 0.04 | [-3.92, 3.99] |
| Dakwar 2020 | 0.29 | [-3.67, 4.26] |
| Daly 2018 PA | -0.03 | [-3.98, 3.92] |
| Daly 2018 PB | 0.05 | [-3.92, 4.01] |
| Domany 2019 | -0.20 | [-4.16, 3.77] |
| Domany 2020 | 0.00 | [-4.02, 4.02] |
| Duek 2023 | 0.00 | [-3.99, 3.99] |
| Dwyer 2021 | -0.57 | [-4.61, 3.47] |
| Fava 2020 | -1.42 | [-5.37, 2.53] |
| Feder 2014 | -0.14 | [-4.11, 3.82] |
| Feder 2021 | 0.00 | [-3.98, 3.98] |
| Fedgchin 2019 | -0.71 | [-4.64, 3.21] |
| Fineberg 2023 | 0.17 | [-3.83, 4.18] |
| Fu 2020 | 1.10 | [-2.11, 4.31] |
| George 2017 | -1.02 | [-5.09, 3.04] |
| Glue 2024b | -0.72 | [-4.73, 3.28] |
| Grabski 2022 | 0.00 | [-3.94, 3.94] |
| Grunebaum 2017 | -0.24 | [-4.27, 3.80] |
| Grunebaum 2018 | 0.00 | [-3.94, 3.94] |
| Ionescu 2020 | -0.01 | [-3.94, 3.92] |
| Jones 2024 | -0.42 | [-4.41, 3.57] |
| Lapidus 2014 | 0.00 | [-4.01, 4.01] |
| Leal 2023 | 0.00 | [-4.09, 4.09] |
| Lii 2023 | 0.00 | [-3.97, 3.97] |
| Lijffijt 2021 | -0.42 | [-4.40, 3.56] |
| Loo 2016 | -0.38 | [-4.42, 3.67] |
| Loo 2023 | 0.03 | [-3.90, 3.96] |
| Milak 2020 | -1.95 | [-5.97, 2.07] |
| Murrough 2013 | -0.62 | [-4.57, 3.33] |
| Murrough 2015 II | 0.00 | [-4.00, 4.00] |
| Ochs-Ross 2020 | -0.10 | [-4.04, 3.83] |
| Ohtani 2024 | 0.00 | [-3.98, 3.98] |
| Pattanaseri 2024 | -0.19 | [-4.20, 3.82] |
| Phillips 2019 | -0.05 | [-4.01, 3.92] |
| Popova 2019 | -0.05 | [-3.98, 3.88] |
| Price 2022 | -0.70 | [-4.63, 3.24] |
| Ray-Griffith 2017 | 0.00 | [-4.03, 4.03] |
| Rodriguez 2013 | -0.13 | [-4.17, 3.92] |
| Schiweck 2021 | -1.05 | [-5.06, 2.95] |
| Shiroma 2020 | -0.15 | [-4.10, 3.81] |
| Singh 2016 2x doses | -0.11 | [-4.09, 3.86] |
| Singh 2016 3x doses | -0.06 | [-4.04, 3.92] |
| Singh 2016 II | -0.67 | [-4.66, 3.32] |
| Smith-Apeldoorn 2024 | -0.06 | [-2.85, 2.74] |
| Sos 2013 | 0.53 | [-3.46, 4.52] |
| Su 2017 | -0.66 | [-4.61, 3.29] |
| Su 2023 | 0.00 | [-3.94, 3.94] |
| Sumner 2019 | -0.06 | [-4.04, 3.92] |
| Takahashi 2021 | -0.42 | [-4.35, 3.51] |
| Taylor 2018 | 0.00 | [-4.02, 4.02] |
| Tiger 2020 | -0.67 | [-4.66, 3.32] |
| Wang 2024 | 0.00 | [-3.93, 3.93] |
| Xiao 2023 | 0.00 | [-3.98, 3.98] |
| Yang 2023 | -0.69 | [-4.62, 3.24] |
| Zarate 2006 | 0.00 | [-4.02, 4.02] |
| Zarate 2012 | 0.09 | [-3.87, 4.05] |
| Zhou 2024 | 0.00 | [-3.96, 3.96] |
| **Summary** | **0.42** | **[-0.76, 1.71]** |
| **Prediction** | **0.43** | **[-1.02, 1.98]** |

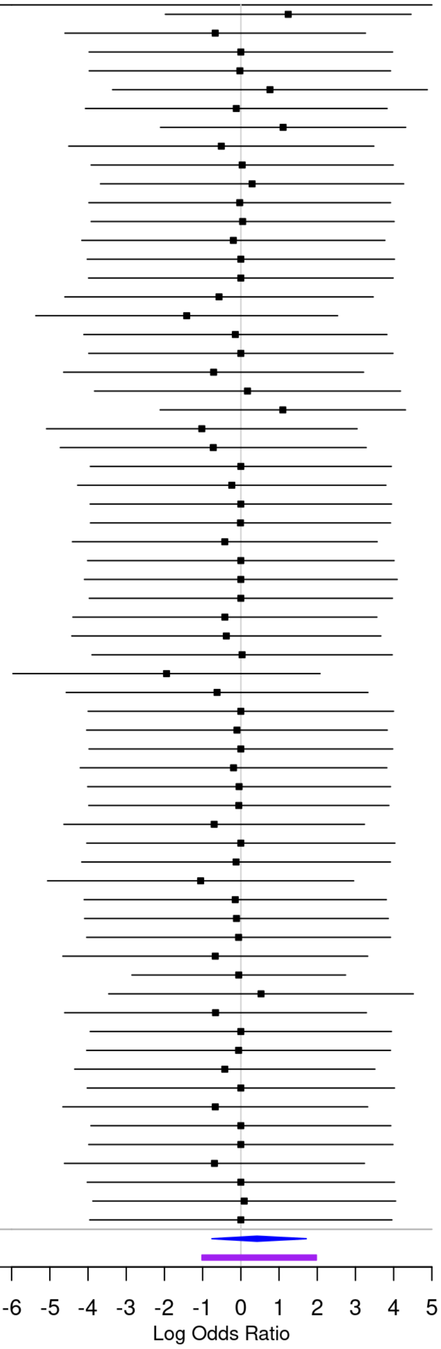

*Heterogeneity (tau): 0.28 [0.00, 0.83]*

**Bayesian Meta-Analysis, smaller prior**

**Bayesian MA, Delta = 15 - Suicides TR+FU**

| Study | Estimate | 95% CI |
|---|---|---|
| Abbar 2022 | 1.24 | [-1.98, 4.46] |
| Abdallah 2022 | -0.67 | [-4.60, 3.26] |
| Ahmed 2023 | 0.00 | [-3.97, 3.97] |
| Anderson 2017 | -0.03 | [-3.97, 3.92] |
| Burger 2016 | 0.76 | [-3.36, 4.88] |
| Canuso 2018 | -0.12 | [-4.07, 3.83] |
| Chen 2023 | 1.11 | [-2.10, 4.32] |
| Colla 2024 | -0.51 | [-4.50, 3.48] |
| Dakwar 2019 | 0.04 | [-3.92, 3.99] |
| Dakwar 2020 | 0.29 | [-3.67, 4.26] |
| Daly 2018 PA | -0.03 | [-3.98, 3.92] |
| Daly 2018 PB | 0.05 | [-3.92, 4.01] |
| Domany 2019 | -0.20 | [-4.16, 3.77] |
| Domany 2020 | 0.00 | [-4.02, 4.02] |
| Duek 2023 | 0.00 | [-3.99, 3.99] |
| Dwyer 2021 | -0.57 | [-4.61, 3.47] |
| Fava 2020 | -1.42 | [-5.37, 2.53] |
| Feder 2014 | -0.14 | [-4.11, 3.82] |
| Feder 2021 | 0.00 | [-3.98, 3.98] |
| Fedgchin 2019 | -0.71 | [-4.64, 3.21] |
| Fineberg 2023 | 0.17 | [-3.83, 4.18] |
| Fu 2020 | 1.10 | [-2.11, 4.31] |
| George 2017 | -1.02 | [-5.09, 3.04] |
| Glue 2024b | -0.72 | [-4.73, 3.28] |
| Grabski 2022 | 0.00 | [-3.94, 3.94] |
| Grunebaum 2017 | -0.24 | [-4.27, 3.80] |
| Grunebaum 2018 | 0.00 | [-3.94, 3.94] |
| Ionescu 2020 | -0.01 | [-3.94, 3.92] |
| Jones 2024 | -0.42 | [-4.41, 3.57] |
| Lapidus 2014 | 0.00 | [-4.01, 4.01] |
| Leal 2023 | 0.00 | [-4.09, 4.09] |
| Lii 2023 | 0.00 | [-3.97, 3.97] |
| Lijffijt 2021 | -0.42 | [-4.40, 3.56] |
| Loo 2016 | -0.38 | [-4.42, 3.67] |
| Loo 2023 | 0.03 | [-3.90, 3.96] |
| Milak 2020 | -1.95 | [-5.97, 2.07] |
| Murrough 2013 | -0.62 | [-4.57, 3.33] |
| Murrough 2015 II | 0.00 | [-4.00, 4.00] |
| Ochs-Ross 2020 | -0.10 | [-4.04, 3.83] |
| Ohtani 2024 | 0.00 | [-3.98, 3.98] |
| Pattanaseri 2024 | -0.19 | [-4.20, 3.82] |
| Phillips 2019 | -0.05 | [-4.01, 3.92] |
| Popova 2019 | -0.05 | [-3.98, 3.88] |
| Price 2022 | -0.70 | [-4.63, 3.24] |
| Ray-Griffith 2017 | 0.00 | [-4.03, 4.03] |
| Rodriguez 2013 | -0.13 | [-4.17, 3.92] |
| Schiweck 2021 | -1.05 | [-5.06, 2.95] |
| Shiroma 2020 | -0.15 | [-4.10, 3.81] |
| Singh 2016 2x doses | -0.11 | [-4.09, 3.86] |
| Singh 2016 3x doses | -0.06 | [-4.04, 3.92] |
| Singh 2016 II | -0.67 | [-4.66, 3.32] |
| Smith-Apeldoorn 2024 | -0.06 | [-2.85, 2.74] |
| Sos 2013 | 0.53 | [-3.46, 4.52] |
| Su 2017 | -0.66 | [-4.61, 3.29] |
| Su 2023 | 0.00 | [-3.94, 3.94] |
| Sumner 2019 | -0.06 | [-4.04, 3.92] |
| Takahashi 2021 | -0.42 | [-4.35, 3.51] |
| Taylor 2018 | 0.00 | [-4.02, 4.02] |
| Tiger 2020 | -0.67 | [-4.66, 3.32] |
| Wang 2024 | 0.00 | [-3.93, 3.93] |
| Xiao 2023 | 0.00 | [-3.98, 3.98] |
| Yang 2023 | -0.69 | [-4.62, 3.24] |
| Zarate 2006 | 0.00 | [-4.02, 4.02] |
| Zarate 2012 | 0.09 | [-3.87, 4.05] |
| Zhou 2024 | 0.00 | [-3.96, 3.96] |
| **Summary** | **0.36** | **[-0.76, 1.54]** |
| **Prediction** | **0.37** | **[-1.05, 1.82]** |

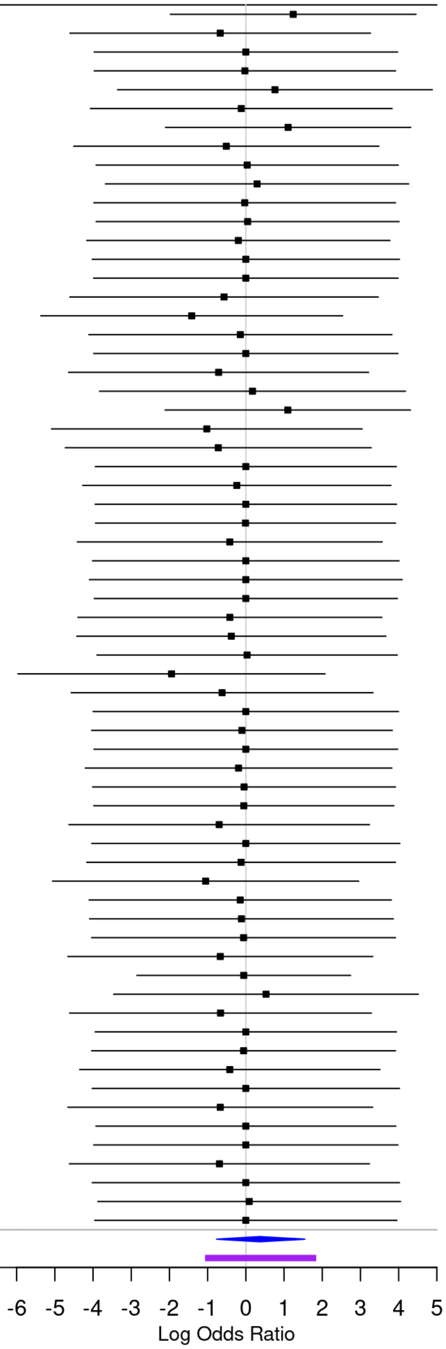

*Heterogeneity (tau): 0.28 [0.00, 0.83]*

**Exact Method**

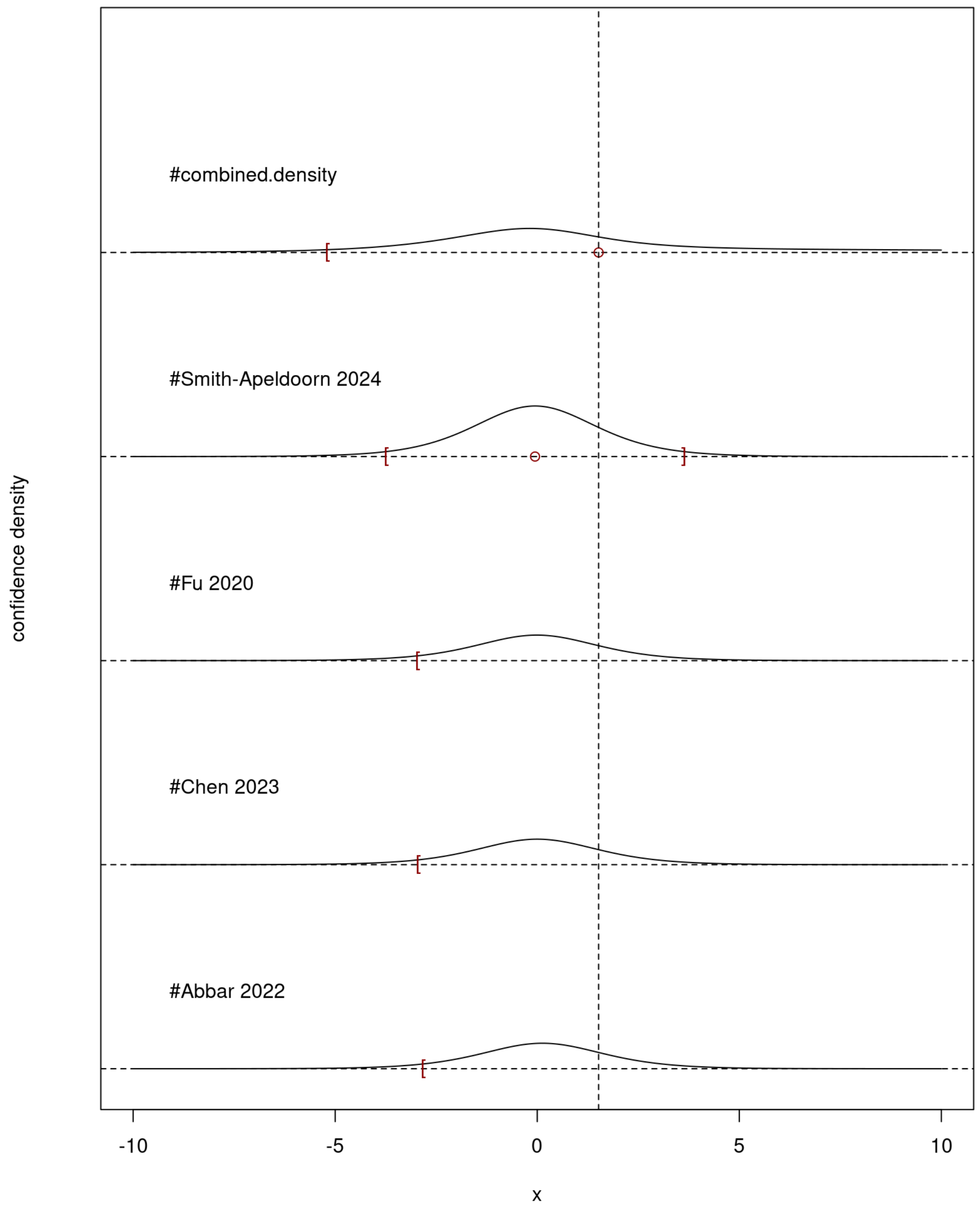

### Deaths

#### Treatment Phase

#### Peto's Method

| Study | Experimental Events | Experimental Total | Control Events | Control Total | OR | 95%-CI | Weight (common) | Weight (random) |
|---|---|---|---|---|---|---|---|---|
| Abbar 2022 | 0 | 73 | 0 | 73 | | | 0.0% | 0.0% |
| Abdallah 2022 | 0 | 108 | 0 | 108 | | | 0.0% | 0.0% |
| Ahmed 2023 | 0 | 18 | 0 | 18 | | | 0.0% | 0.0% |
| Anderson 2017 | 0 | 40 | 0 | 40 | | | 0.0% | 0.0% |
| Burger 2016 | 0 | 3 | 0 | 3 | | | 0.0% | 0.0% |
| Canuso 2018 | 0 | 35 | 0 | 35 | | | 0.0% | 0.0% |
| Chen 2023 | 1 | 126 | 0 | 126 | 7.39 | [0.15; 372.38] | 25.1% | 25.1% |
| Colla 2024 | 0 | 17 | 0 | 17 | | | 0.0% | 0.0% |
| Dakwar 2019 | 0 | 27 | 0 | 27 | | | 0.0% | 0.0% |
| Dakwar 2020 | 0 | 17 | 0 | 17 | | | 0.0% | 0.0% |
| Daly 2018 PA | 0 | 34 | 0 | 34 | | | 0.0% | 0.0% |
| Daly 2018 PB | 0 | 20 | 0 | 20 | | | 0.0% | 0.0% |
| Domany 2019 | 0 | 22 | 0 | 22 | | | 0.0% | 0.0% |
| Domany 2020 | 0 | 9 | 0 | 9 | | | 0.0% | 0.0% |
| Duek 2023 | 0 | 14 | 0 | 14 | | | 0.0% | 0.0% |
| Dwyer 2021 | 0 | 11 | 0 | 11 | | | 0.0% | 0.0% |
| Fava 2020 | 0 | 80 | 0 | 80 | | | 0.0% | 0.0% |
| Feder 2014 | 0 | 22 | 0 | 22 | | | 0.0% | 0.0% |
| Feder 2021 | 0 | 15 | 0 | 15 | | | 0.0% | 0.0% |
| Fedgchin 2019 | 0 | 231 | 0 | 231 | | | 0.0% | 0.0% |
| Fineberg 2023 | 0 | 10 | 0 | 10 | | | 0.0% | 0.0% |
| Fu 2020 | 0 | 113 | 0 | 113 | | | 0.0% | 0.0% |
| George 2017 | 0 | 12 | 0 | 12 | | | 0.0% | 0.0% |
| Glue 2024b | 0 | 17 | 0 | 17 | | | 0.0% | 0.0% |
| Grabski 2022 | 0 | 48 | 0 | 48 | | | 0.0% | 0.0% |
| Grunebaum 2017 | 0 | 9 | 0 | 9 | | | 0.0% | 0.0% |
| Grunebaum 2018 | 0 | 40 | 0 | 40 | | | 0.0% | 0.0% |
| Ionescu 2020 | 0 | 114 | 0 | 114 | | | 0.0% | 0.0% |
| Jones 2024 | 0 | 17 | 0 | 17 | | | 0.0% | 0.0% |
| Lapidus 2014 | 0 | 10 | 0 | 10 | | | 0.0% | 0.0% |
| Leal 2023 | 0 | 5 | 0 | 5 | | | 0.0% | 0.0% |
| Lii 2023 | 0 | 20 | 0 | 20 | | | 0.0% | 0.0% |
| Lijffijt 2021 | 0 | 20 | 0 | 20 | | | 0.0% | 0.0% |
| Loo 2016 | 0 | 9 | 0 | 9 | | | 0.0% | 0.0% |
| Loo 2023 | 0 | 89 | 0 | 89 | | | 0.0% | 0.0% |
| Milak 2020 | 0 | 38 | 0 | 38 | | | 0.0% | 0.0% |
| Murrough 2013 | 0 | 47 | 0 | 47 | | | 0.0% | 0.0% |
| Murrough 2015 II | 0 | 12 | 0 | 12 | | | 0.0% | 0.0% |
| Ochs-Ross 2020 | 0 | 72 | 0 | 72 | | | 0.0% | 0.0% |
| Ohtani 2024 | 0 | 17 | 0 | 17 | | | 0.0% | 0.0% |
| Pattanaseri 2024 | 0 | 11 | 0 | 11 | | | 0.0% | 0.0% |
| Phillips 2019 | 0 | 22 | 0 | 22 | | | 0.0% | 0.0% |
| Popova 2019 | 0 | 115 | 0 | 115 | | | 0.0% | 0.0% |
| Price 2022 | 0 | 103 | 0 | 103 | | | 0.0% | 0.0% |
| Ray-Griffith 2017 | 0 | 8 | 0 | 8 | | | 0.0% | 0.0% |
| Rodriguez 2013 | 0 | 8 | 0 | 8 | | | 0.0% | 0.0% |
| Schiweck 2021 | 0 | 21 | 0 | 21 | | | 0.0% | 0.0% |
| Shiroma 2020 | 0 | 29 | 0 | 29 | | | 0.0% | 0.0% |
| Singh 2016 2x doses | 0 | 18 | 0 | 18 | | | 0.0% | 0.0% |
| Singh 2016 3x doses | 0 | 17 | 0 | 17 | | | 0.0% | 0.0% |
| Singh 2016 II | 0 | 20 | 0 | 20 | | | 0.0% | 0.0% |
| Smith-Apeldoorn 2024 | 1 | 57 | 1 | 57 | 1.00 | [0.06; 16.19] | 49.8% | 49.8% |
| Sos 2013 | 0 | 11 | 0 | 11 | | | 0.0% | 0.0% |
| Su 2017 | 0 | 47 | 0 | 47 | | | 0.0% | 0.0% |
| Su 2023 | 0 | 42 | 0 | 42 | | | 0.0% | 0.0% |
| Sumner 2019 | 0 | 17 | 0 | 17 | | | 0.0% | 0.0% |
| Takahashi 2021 | 0 | 122 | 0 | 122 | | | 0.0% | 0.0% |
| Taylor 2018 | 0 | 9 | 0 | 9 | | | 0.0% | 0.0% |
| Tiger 2020 | 0 | 20 | 0 | 20 | | | 0.0% | 0.0% |
| Wang 2024 | 0 | 182 | 0 | 182 | | | 0.0% | 0.0% |
| Xiao 2023 | 0 | 15 | 0 | 15 | | | 0.0% | 0.0% |
| Yang 2023 | 0 | 200 | 0 | 200 | | | 0.0% | 0.0% |
| Zarate 2006 | 0 | 9 | 0 | 9 | | | 0.0% | 0.0% |
| Zarate 2012 | 0 | 21 | 0 | 21 | | | 0.0% | 0.0% |
| Zhou 2024 | 0 | 27 | 1 | 27 | 0.14 | [0.00; 6.82] | 25.1% | 25.1% |
| **Common effect model** | | **2792** | | **2792** | **1.00** | **[0.14; 7.13]** | **100.0%** | **.** |
| **Random effects model** | | | | | **1.00** | **[0.14; 7.13]** | **.** | **100.0%** |

Heterogeneity: $I^2 = 0.0\%$, $\tau^2 < 0.0001$, $p = 0.3679$

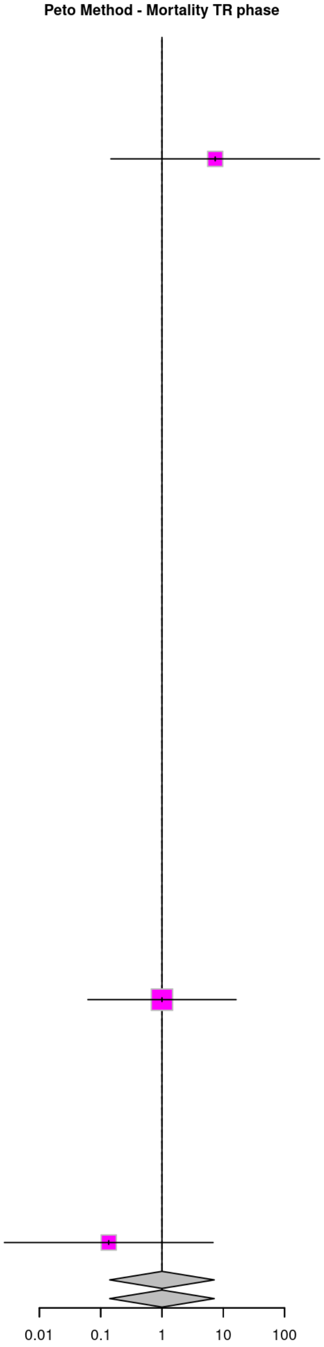

#### Maentel Mantel-Haenszel with CC

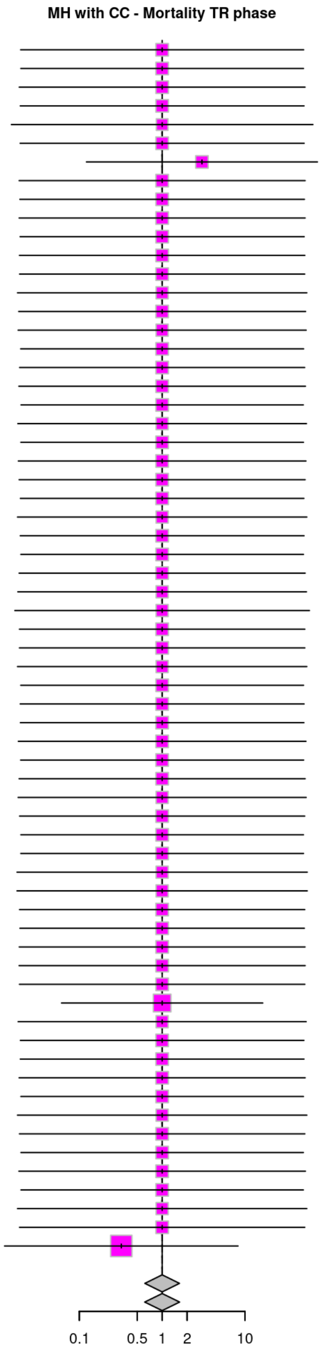

| Study | Experimental Events | Experimental Total | Control Events | Control Total | OR | 95%-CI | Weight (common) | Weight (random) |
|---|---|---|---|---|---|---|---|---|
| Abbar 2022 | 0 | 73 | 0 | 73 | 1.00 | [0.02; 51.07] | 1.5% | 1.5% |
| Abdallah 2022 | 0 | 108 | 0 | 108 | 1.00 | [0.02; 50.85] | 1.5% | 1.5% |
| Ahmed 2023 | 0 | 18 | 0 | 18 | 1.00 | [0.02; 53.12] | 1.5% | 1.5% |
| Anderson 2017 | 0 | 40 | 0 | 40 | 1.00 | [0.02; 51.63] | 1.5% | 1.5% |
| Burger 2016 | 0 | 3 | 0 | 3 | 1.00 | [0.02; 66.06] | 1.3% | 1.3% |
| Canuso 2018 | 0 | 35 | 0 | 35 | 1.00 | [0.02; 51.80] | 1.5% | 1.5% |
| Chen 2023 | 1 | 126 | 0 | 126 | 3.02 | [0.12; 74.94] | 1.5% | 2.3% |
| Colla 2024 | 0 | 17 | 0 | 17 | 1.00 | [0.02; 53.28] | 1.5% | 1.5% |
| Dakwar 2019 | 0 | 27 | 0 | 27 | 1.00 | [0.02; 52.22] | 1.5% | 1.5% |
| Dakwar 2020 | 0 | 17 | 0 | 17 | 1.00 | [0.02; 53.28] | 1.5% | 1.5% |
| Daly 2018 PA | 0 | 34 | 0 | 34 | 1.00 | [0.02; 51.84] | 1.5% | 1.5% |
| Daly 2018 PB | 0 | 20 | 0 | 20 | 1.00 | [0.02; 52.85] | 1.5% | 1.5% |
| Domany 2019 | 0 | 22 | 0 | 22 | 1.00 | [0.02; 52.63] | 1.5% | 1.5% |
| Domany 2020 | 0 | 9 | 0 | 9 | 1.00 | [0.02; 55.80] | 1.4% | 1.5% |
| Duek 2023 | 0 | 14 | 0 | 14 | 1.00 | [0.02; 53.89] | 1.5% | 1.5% |
| Dwyer 2021 | 0 | 11 | 0 | 11 | 1.00 | [0.02; 54.83] | 1.4% | 1.5% |
| Fava 2020 | 0 | 80 | 0 | 80 | 1.00 | [0.02; 51.01] | 1.5% | 1.5% |
| Feder 2014 | 0 | 22 | 0 | 22 | 1.00 | [0.02; 52.63] | 1.5% | 1.5% |
| Feder 2021 | 0 | 15 | 0 | 15 | 1.00 | [0.02; 53.66] | 1.5% | 1.5% |
| Fedgchin 2019 | 0 | 231 | 0 | 231 | 1.00 | [0.02; 50.61] | 1.5% | 1.5% |
| Fineberg 2023 | 0 | 10 | 0 | 10 | 1.00 | [0.02; 55.27] | 1.4% | 1.5% |
| Fu 2020 | 0 | 113 | 0 | 113 | 1.00 | [0.02; 50.83] | 1.5% | 1.5% |
| George 2017 | 0 | 12 | 0 | 12 | 1.00 | [0.02; 54.46] | 1.5% | 1.5% |
| Glue 2024b | 0 | 17 | 0 | 17 | 1.00 | [0.02; 53.28] | 1.5% | 1.5% |
| Grabski 2022 | 0 | 48 | 0 | 48 | 1.00 | [0.02; 51.42] | 1.5% | 1.5% |
| Grunebaum 2017 | 0 | 9 | 0 | 9 | 1.00 | [0.02; 55.80] | 1.4% | 1.5% |
| Grunebaum 2018 | 0 | 40 | 0 | 40 | 1.00 | [0.02; 51.63] | 1.5% | 1.5% |
| Ionescu 2020 | 0 | 114 | 0 | 114 | 1.00 | [0.02; 50.83] | 1.5% | 1.5% |
| Jones 2024 | 0 | 17 | 0 | 17 | 1.00 | [0.02; 53.28] | 1.5% | 1.5% |
| Lapidus 2014 | 0 | 10 | 0 | 10 | 1.00 | [0.02; 55.27] | 1.4% | 1.5% |
| Leal 2023 | 0 | 5 | 0 | 5 | 1.00 | [0.02; 59.99] | 1.4% | 1.4% |
| Lii 2023 | 0 | 20 | 0 | 20 | 1.00 | [0.02; 52.85] | 1.5% | 1.5% |
| Lijffijt 2021 | 0 | 20 | 0 | 20 | 1.00 | [0.02; 52.85] | 1.5% | 1.5% |
| Loo 2016 | 0 | 9 | 0 | 9 | 1.00 | [0.02; 55.80] | 1.4% | 1.5% |
| Loo 2023 | 0 | 89 | 0 | 89 | 1.00 | [0.02; 50.95] | 1.5% | 1.5% |
| Milak 2020 | 0 | 38 | 0 | 38 | 1.00 | [0.02; 51.69] | 1.5% | 1.5% |
| Murrough 2013 | 0 | 47 | 0 | 47 | 1.00 | [0.02; 51.44] | 1.5% | 1.5% |
| Murrough 2015 II | 0 | 12 | 0 | 12 | 1.00 | [0.02; 54.46] | 1.5% | 1.5% |
| Ochs-Ross 2020 | 0 | 72 | 0 | 72 | 1.00 | [0.02; 51.08] | 1.5% | 1.5% |
| Ohtani 2024 | 0 | 17 | 0 | 17 | 1.00 | [0.02; 53.28] | 1.5% | 1.5% |
| Pattanaseri 2024 | 0 | 11 | 0 | 11 | 1.00 | [0.02; 54.83] | 1.4% | 1.5% |
| Phillips 2019 | 0 | 22 | 0 | 22 | 1.00 | [0.02; 52.63] | 1.5% | 1.5% |
| Popova 2019 | 0 | 115 | 0 | 115 | 1.00 | [0.02; 50.83] | 1.5% | 1.5% |
| Price 2022 | 0 | 103 | 0 | 103 | 1.00 | [0.02; 50.88] | 1.5% | 1.5% |
| Ray-Griffith 2017 | 0 | 8 | 0 | 8 | 1.00 | [0.02; 56.46] | 1.4% | 1.4% |
| Rodriguez 2013 | 0 | 8 | 0 | 8 | 1.00 | [0.02; 56.46] | 1.4% | 1.4% |
| Schiweck 2021 | 0 | 21 | 0 | 21 | 1.00 | [0.02; 52.73] | 1.5% | 1.5% |
| Shiroma 2020 | 0 | 29 | 0 | 29 | 1.00 | [0.02; 52.09] | 1.5% | 1.5% |
| Singh 2016 2x doses | 0 | 18 | 0 | 18 | 1.00 | [0.02; 53.12] | 1.5% | 1.5% |
| Singh 2016 3x doses | 0 | 17 | 0 | 17 | 1.00 | [0.02; 53.28] | 1.5% | 1.5% |
| Singh 2016 II | 0 | 20 | 0 | 20 | 1.00 | [0.02; 52.85] | 1.5% | 1.5% |
| Smith-Apeldoorn 2024 | 1 | 57 | 1 | 57 | 1.00 | [0.06; 16.39] | 3.0% | 3.0% |
| Sos 2013 | 0 | 11 | 0 | 11 | 1.00 | [0.02; 54.83] | 1.4% | 1.5% |
| Su 2017 | 0 | 47 | 0 | 47 | 1.00 | [0.02; 51.44] | 1.5% | 1.5% |
| Su 2023 | 0 | 42 | 0 | 42 | 1.00 | [0.02; 51.57] | 1.5% | 1.5% |
| Sumner 2019 | 0 | 17 | 0 | 17 | 1.00 | [0.02; 53.28] | 1.5% | 1.5% |
| Takahashi 2021 | 0 | 122 | 0 | 122 | 1.00 | [0.02; 50.80] | 1.5% | 1.5% |
| Taylor 2018 | 0 | 9 | 0 | 9 | 1.00 | [0.02; 55.80] | 1.4% | 1.5% |
| Tiger 2020 | 0 | 20 | 0 | 20 | 1.00 | [0.02; 52.85] | 1.5% | 1.5% |
| Wang 2024 | 0 | 182 | 0 | 182 | 1.00 | [0.02; 50.67] | 1.5% | 1.5% |
| Xiao 2023 | 0 | 15 | 0 | 15 | 1.00 | [0.02; 53.66] | 1.5% | 1.5% |
| Yang 2023 | 0 | 200 | 0 | 200 | 1.00 | [0.02; 50.64] | 1.5% | 1.5% |
| Zarate 2006 | 0 | 9 | 0 | 9 | 1.00 | [0.02; 55.80] | 1.4% | 1.5% |
| Zarate 2012 | 0 | 21 | 0 | 21 | 1.00 | [0.02; 52.73] | 1.5% | 1.5% |
| Zhou 2024 | 0 | 27 | 1 | 27 | 0.32 | [0.01; 8.24] | 4.4% | 2.2% |
| **Common effect model** | | **2792** | | **2792** | **1.00** | **[0.62; 1.62]** | **100.0%** | **.** |
| **Random effects model** | | | | | **1.00** | **[0.62; 1.62]** | **.** | **100.0%** |

Heterogeneity: $I^2 = 0.0\%$, $\tau^2 = 0$, $p = 1.0000$

#### Maentel Mantel-Haenszel with Treatment-Arm CC

| Study | Experimental Events | Experimental Total | Control Events | Control Total | OR | 95%-CI | Weight (common) | Weight (random) |
|---|---|---|---|---|---|---|---|---|
| Abbar 2022 | 0 | 73 | 0 | 73 | 1.00 | [0.02; 51.07] | 1.5% | 1.5% |
| Abdallah 2022 | 0 | 108 | 0 | 108 | 1.00 | [0.02; 50.85] | 1.5% | 1.5% |
| Ahmed 2023 | 0 | 18 | 0 | 18 | 1.00 | [0.02; 53.12] | 1.5% | 1.5% |
| Anderson 2017 | 0 | 40 | 0 | 40 | 1.00 | [0.02; 51.63] | 1.5% | 1.5% |
| Burger 2016 | 0 | 3 | 0 | 3 | 1.00 | [0.02; 66.06] | 1.3% | 1.3% |
| Canuso 2018 | 0 | 35 | 0 | 35 | 1.00 | [0.02; 51.80] | 1.5% | 1.5% |
| Chen 2023 | 1 | 126 | 0 | 126 | 3.02 | [0.12; 74.94] | 1.5% | 2.3% |
| Colla 2024 | 0 | 17 | 0 | 17 | 1.00 | [0.02; 53.28] | 1.5% | 1.5% |
| Dakwar 2019 | 0 | 27 | 0 | 27 | 1.00 | [0.02; 52.22] | 1.5% | 1.5% |
| Dakwar 2020 | 0 | 17 | 0 | 17 | 1.00 | [0.02; 53.28] | 1.5% | 1.5% |
| Daly 2018 PA | 0 | 34 | 0 | 34 | 1.00 | [0.02; 51.84] | 1.5% | 1.5% |
| Daly 2018 PB | 0 | 20 | 0 | 20 | 1.00 | [0.02; 52.85] | 1.5% | 1.5% |
| Domany 2019 | 0 | 22 | 0 | 22 | 1.00 | [0.02; 52.63] | 1.5% | 1.5% |
| Domany 2020 | 0 | 9 | 0 | 9 | 1.00 | [0.02; 55.80] | 1.4% | 1.5% |
| Duek 2023 | 0 | 14 | 0 | 14 | 1.00 | [0.02; 53.89] | 1.5% | 1.5% |
| Dwyer 2021 | 0 | 11 | 0 | 11 | 1.00 | [0.02; 54.83] | 1.4% | 1.5% |
| Fava 2020 | 0 | 80 | 0 | 80 | 1.00 | [0.02; 51.01] | 1.5% | 1.5% |
| Feder 2014 | 0 | 22 | 0 | 22 | 1.00 | [0.02; 52.63] | 1.5% | 1.5% |
| Feder 2021 | 0 | 15 | 0 | 15 | 1.00 | [0.02; 53.66] | 1.5% | 1.5% |
| Fedgchin 2019 | 0 | 231 | 0 | 231 | 1.00 | [0.02; 50.61] | 1.5% | 1.5% |
| Fineberg 2023 | 0 | 10 | 0 | 10 | 1.00 | [0.02; 55.27] | 1.4% | 1.5% |
| Fu 2020 | 0 | 113 | 0 | 113 | 1.00 | [0.02; 50.83] | 1.5% | 1.5% |
| George 2017 | 0 | 12 | 0 | 12 | 1.00 | [0.02; 54.46] | 1.5% | 1.5% |
| Glue 2024b | 0 | 17 | 0 | 17 | 1.00 | [0.02; 53.28] | 1.5% | 1.5% |
| Grabski 2022 | 0 | 48 | 0 | 48 | 1.00 | [0.02; 51.42] | 1.5% | 1.5% |
| Grunebaum 2017 | 0 | 9 | 0 | 9 | 1.00 | [0.02; 55.80] | 1.4% | 1.5% |
| Grunebaum 2018 | 0 | 40 | 0 | 40 | 1.00 | [0.02; 51.63] | 1.5% | 1.5% |
| Ionescu 2020 | 0 | 114 | 0 | 114 | 1.00 | [0.02; 50.83] | 1.5% | 1.5% |
| Jones 2024 | 0 | 17 | 0 | 17 | 1.00 | [0.02; 53.28] | 1.5% | 1.5% |
| Lapidus 2014 | 0 | 10 | 0 | 10 | 1.00 | [0.02; 55.27] | 1.4% | 1.5% |
| Leal 2023 | 0 | 5 | 0 | 5 | 1.00 | [0.02; 59.99] | 1.4% | 1.4% |
| Lii 2023 | 0 | 20 | 0 | 20 | 1.00 | [0.02; 52.85] | 1.5% | 1.5% |
| Lijffijt 2021 | 0 | 20 | 0 | 20 | 1.00 | [0.02; 52.85] | 1.5% | 1.5% |
| Loo 2016 | 0 | 9 | 0 | 9 | 1.00 | [0.02; 55.80] | 1.4% | 1.5% |
| Loo 2023 | 0 | 89 | 0 | 89 | 1.00 | [0.02; 50.95] | 1.5% | 1.5% |
| Milak 2020 | 0 | 38 | 0 | 38 | 1.00 | [0.02; 51.69] | 1.5% | 1.5% |
| Murrough 2013 | 0 | 47 | 0 | 47 | 1.00 | [0.02; 51.44] | 1.5% | 1.5% |
| Murrough 2015 II | 0 | 12 | 0 | 12 | 1.00 | [0.02; 54.46] | 1.5% | 1.5% |
| Ochs-Ross 2020 | 0 | 72 | 0 | 72 | 1.00 | [0.02; 51.08] | 1.5% | 1.5% |
| Ohtani 2024 | 0 | 17 | 0 | 17 | 1.00 | [0.02; 53.28] | 1.5% | 1.5% |
| Pattanaseri 2024 | 0 | 11 | 0 | 11 | 1.00 | [0.02; 54.83] | 1.4% | 1.5% |
| Phillips 2019 | 0 | 22 | 0 | 22 | 1.00 | [0.02; 52.63] | 1.5% | 1.5% |
| Popova 2019 | 0 | 115 | 0 | 115 | 1.00 | [0.02; 50.83] | 1.5% | 1.5% |
| Price 2022 | 0 | 103 | 0 | 103 | 1.00 | [0.02; 50.88] | 1.5% | 1.5% |
| Ray-Griffith 2017 | 0 | 8 | 0 | 8 | 1.00 | [0.02; 56.46] | 1.4% | 1.4% |
| Rodriguez 2013 | 0 | 8 | 0 | 8 | 1.00 | [0.02; 56.46] | 1.4% | 1.4% |
| Schiweck 2021 | 0 | 21 | 0 | 21 | 1.00 | [0.02; 52.73] | 1.5% | 1.5% |
| Shiroma 2020 | 0 | 29 | 0 | 29 | 1.00 | [0.02; 52.09] | 1.5% | 1.5% |
| Singh 2016 2x doses | 0 | 18 | 0 | 18 | 1.00 | [0.02; 53.12] | 1.5% | 1.5% |
| Singh 2016 3x doses | 0 | 17 | 0 | 17 | 1.00 | [0.02; 53.28] | 1.5% | 1.5% |
| Singh 2016 II | 0 | 20 | 0 | 20 | 1.00 | [0.02; 52.85] | 1.5% | 1.5% |
| Smith-Apeldoorn 2024 | 1 | 57 | 1 | 57 | 1.00 | [0.06; 16.39] | 3.0% | 3.0% |
| Sos 2013 | 0 | 11 | 0 | 11 | 1.00 | [0.02; 54.83] | 1.4% | 1.5% |
| Su 2017 | 0 | 47 | 0 | 47 | 1.00 | [0.02; 51.44] | 1.5% | 1.5% |
| Su 2023 | 0 | 42 | 0 | 42 | 1.00 | [0.02; 51.57] | 1.5% | 1.5% |
| Sumner 2019 | 0 | 17 | 0 | 17 | 1.00 | [0.02; 53.28] | 1.5% | 1.5% |
| Takahashi 2021 | 0 | 122 | 0 | 122 | 1.00 | [0.02; 50.80] | 1.5% | 1.5% |
| Taylor 2018 | 0 | 9 | 0 | 9 | 1.00 | [0.02; 55.80] | 1.4% | 1.5% |
| Tiger 2020 | 0 | 20 | 0 | 20 | 1.00 | [0.02; 52.85] | 1.5% | 1.5% |
| Wang 2024 | 0 | 182 | 0 | 182 | 1.00 | [0.02; 50.67] | 1.5% | 1.5% |
| Xiao 2023 | 0 | 15 | 0 | 15 | 1.00 | [0.02; 53.66] | 1.5% | 1.5% |
| Yang 2023 | 0 | 200 | 0 | 200 | 1.00 | [0.02; 50.64] | 1.5% | 1.5% |
| Zarate 2006 | 0 | 9 | 0 | 9 | 1.00 | [0.02; 55.80] | 1.4% | 1.5% |
| Zarate 2012 | 0 | 21 | 0 | 21 | 1.00 | [0.02; 52.73] | 1.5% | 1.5% |
| Zhou 2024 | 0 | 27 | 1 | 27 | 0.32 | [0.01; 8.24] | 4.4% | 2.2% |
| **Common effect model** | | **2792** | | **2792** | **1.00** | **[0.62; 1.62]** | **100.0%** | **.** |
| **Random effects model** | | | | | **1.00** | **[0.62; 1.62]** | **.** | **100.0%** |

Heterogeneity: $I^2 = 0.0\%$, $\tau^2 = 0$, $p = 1.0000$

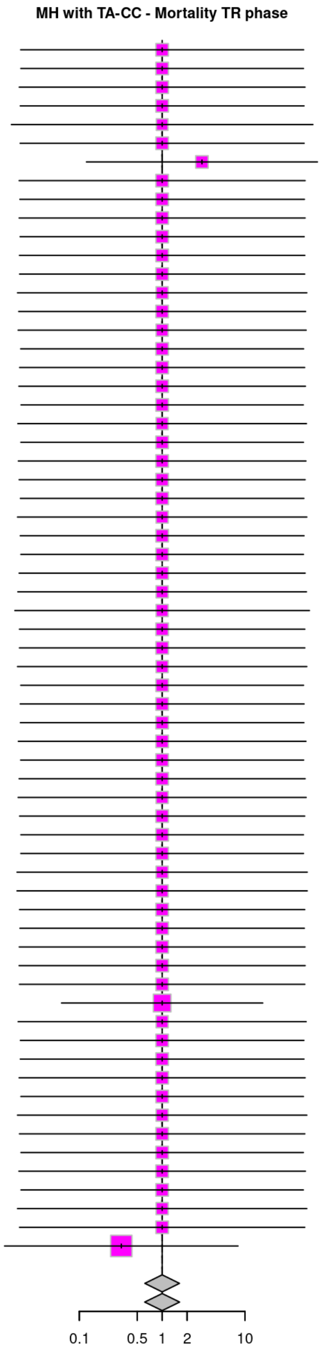

#### Maentel Mantel-Haenszel Risk-Difference

| Study | Experimental Events | Experimental Total | Control Events | Control Total | RD | 95%-CI | Weight (common) | Weight (random) |
|---|---|---|---|---|---|---|---|---|
| Abbar 2022 | 0 | 73 | 0 | 73 | 0.00 | [-0.03; 0.03] | 2.6% | 2.0% |
| Abdallah 2022 | 0 | 108 | 0 | 108 | 0.00 | [-0.02; 0.02] | 3.8% | 4.4% |
| Ahmed 2023 | 0 | 18 | 0 | 18 | 0.00 | [-0.10; 0.10] | 0.7% | 0.1% |
| Anderson 2017 | 0 | 40 | 0 | 40 | 0.00 | [-0.05; 0.05] | 1.4% | 0.6% |
| Burger 2016 | 0 | 3 | 0 | 3 | 0.00 | [-0.46; 0.46] | 0.1% | 0.0% |
| Canuso 2018 | 0 | 35 | 0 | 35 | 0.00 | [-0.05; 0.05] | 1.3% | 0.5% |
| Chen 2023 | 1 | 126 | 0 | 126 | 0.01 | [-0.01; 0.03] | 4.4% | 3.0% |
| Colla 2024 | 0 | 17 | 0 | 17 | 0.00 | [-0.11; 0.11] | 0.6% | 0.1% |
| Dakwar 2019 | 0 | 27 | 0 | 27 | 0.00 | [-0.07; 0.07] | 1.0% | 0.3% |
| Dakwar 2020 | 0 | 17 | 0 | 17 | 0.00 | [-0.11; 0.11] | 0.6% | 0.1% |
| Daly 2018 PA | 0 | 34 | 0 | 34 | 0.00 | [-0.06; 0.06] | 1.2% | 0.5% |
| Daly 2018 PB | 0 | 20 | 0 | 20 | 0.00 | [-0.09; 0.09] | 0.7% | 0.2% |
| Domany 2019 | 0 | 22 | 0 | 22 | 0.00 | [-0.08; 0.08] | 0.8% | 0.2% |
| Domany 2020 | 0 | 9 | 0 | 9 | 0.00 | [-0.19; 0.19] | 0.4% | 0.0% |
| Duek 2023 | 0 | 14 | 0 | 14 | 0.00 | [-0.13; 0.13] | 0.5% | 0.1% |
| Dwyer 2021 | 0 | 11 | 0 | 11 | 0.00 | [-0.16; 0.16] | 0.4% | 0.1% |
| Fava 2020 | 0 | 80 | 0 | 80 | 0.00 | [-0.02; 0.02] | 2.8% | 2.4% |
| Feder 2014 | 0 | 22 | 0 | 22 | 0.00 | [-0.08; 0.08] | 0.8% | 0.2% |
| Feder 2021 | 0 | 15 | 0 | 15 | 0.00 | [-0.12; 0.12] | 0.6% | 0.1% |
| Fedgchin 2019 | 0 | 231 | 0 | 231 | 0.00 | [-0.01; 0.01] | 8.1% | 19.9% |
| Fineberg 2023 | 0 | 10 | 0 | 10 | 0.00 | [-0.17; 0.17] | 0.4% | 0.0% |
| Fu 2020 | 0 | 113 | 0 | 113 | 0.00 | [-0.02; 0.02] | 4.0% | 4.8% |
| George 2017 | 0 | 12 | 0 | 12 | 0.00 | [-0.15; 0.15] | 0.5% | 0.1% |
| Glue 2024b | 0 | 17 | 0 | 17 | 0.00 | [-0.11; 0.11] | 0.6% | 0.1% |
| Grabski 2022 | 0 | 48 | 0 | 48 | 0.00 | [-0.04; 0.04] | 1.7% | 0.9% |
| Grunebaum 2017 | 0 | 9 | 0 | 9 | 0.00 | [-0.19; 0.19] | 0.4% | 0.0% |
| Grunebaum 2018 | 0 | 40 | 0 | 40 | 0.00 | [-0.05; 0.05] | 1.4% | 0.6% |
| Ionescu 2020 | 0 | 114 | 0 | 114 | 0.00 | [-0.02; 0.02] | 4.0% | 4.9% |
| Jones 2024 | 0 | 17 | 0 | 17 | 0.00 | [-0.11; 0.11] | 0.6% | 0.1% |
| Lapidus 2014 | 0 | 10 | 0 | 10 | 0.00 | [-0.17; 0.17] | 0.4% | 0.0% |
| Leal 2023 | 0 | 5 | 0 | 5 | 0.00 | [-0.31; 0.31] | 0.2% | 0.0% |
| Lii 2023 | 0 | 20 | 0 | 20 | 0.00 | [-0.09; 0.09] | 0.7% | 0.2% |
| Lijffijt 2021 | 0 | 20 | 0 | 20 | 0.00 | [-0.09; 0.09] | 0.7% | 0.2% |
| Loo 2016 | 0 | 9 | 0 | 9 | 0.00 | [-0.19; 0.19] | 0.4% | 0.0% |
| Loo 2023 | 0 | 89 | 0 | 89 | 0.00 | [-0.02; 0.02] | 3.2% | 3.0% |
| Milak 2020 | 0 | 38 | 0 | 38 | 0.00 | [-0.05; 0.05] | 1.4% | 0.6% |
| Murrough 2013 | 0 | 47 | 0 | 47 | 0.00 | [-0.04; 0.04] | 1.7% | 0.9% |
| Murrough 2015 II | 0 | 12 | 0 | 12 | 0.00 | [-0.15; 0.15] | 0.5% | 0.1% |
| Ochs-Ross 2020 | 0 | 72 | 0 | 72 | 0.00 | [-0.03; 0.03] | 2.6% | 2.0% |
| Ohtani 2024 | 0 | 17 | 0 | 17 | 0.00 | [-0.11; 0.11] | 0.6% | 0.1% |
| Pattanaseri 2024 | 0 | 11 | 0 | 11 | 0.00 | [-0.16; 0.16] | 0.4% | 0.1% |
| Phillips 2019 | 0 | 22 | 0 | 22 | 0.00 | [-0.08; 0.08] | 0.8% | 0.2% |
| Popova 2019 | 0 | 115 | 0 | 115 | 0.00 | [-0.02; 0.02] | 4.1% | 5.0% |
| Price 2022 | 0 | 103 | 0 | 103 | 0.00 | [-0.02; 0.02] | 3.6% | 4.0% |
| Ray-Griffith 2017 | 0 | 8 | 0 | 8 | 0.00 | [-0.21; 0.21] | 0.3% | 0.0% |
| Rodriguez 2013 | 0 | 8 | 0 | 8 | 0.00 | [-0.21; 0.21] | 0.3% | 0.0% |
| Schiweck 2021 | 0 | 21 | 0 | 21 | 0.00 | [-0.09; 0.09] | 0.8% | 0.2% |
| Shiroma 2020 | 0 | 29 | 0 | 29 | 0.00 | [-0.06; 0.06] | 1.1% | 0.3% |
| Singh 2016 2x doses | 0 | 18 | 0 | 18 | 0.00 | [-0.10; 0.10] | 0.7% | 0.1% |
| Singh 2016 3x doses | 0 | 17 | 0 | 17 | 0.00 | [-0.11; 0.11] | 0.6% | 0.1% |
| Singh 2016 II | 0 | 20 | 0 | 20 | 0.00 | [-0.09; 0.09] | 0.7% | 0.2% |
| Smith-Apeldoorn 2024 | 1 | 57 | 1 | 57 | 0.00 | [-0.05; 0.05] | 2.0% | 0.6% |
| Sos 2013 | 0 | 11 | 0 | 11 | 0.00 | [-0.16; 0.16] | 0.4% | 0.1% |
| Su 2017 | 0 | 47 | 0 | 47 | 0.00 | [-0.04; 0.04] | 1.7% | 0.9% |
| Su 2023 | 0 | 42 | 0 | 42 | 0.00 | [-0.05; 0.05] | 1.5% | 0.7% |
| Sumner 2019 | 0 | 17 | 0 | 17 | 0.00 | [-0.11; 0.11] | 0.6% | 0.1% |
| Takahashi 2021 | 0 | 122 | 0 | 122 | 0.00 | [-0.02; 0.02] | 4.3% | 5.6% |
| Taylor 2018 | 0 | 9 | 0 | 9 | 0.00 | [-0.19; 0.19] | 0.4% | 0.0% |
| Tiger 2020 | 0 | 20 | 0 | 20 | 0.00 | [-0.09; 0.09] | 0.7% | 0.2% |
| Wang 2024 | 0 | 182 | 0 | 182 | 0.00 | [-0.01; 0.01] | 6.4% | 12.4% |
| Xiao 2023 | 0 | 15 | 0 | 15 | 0.00 | [-0.12; 0.12] | 0.6% | 0.1% |
| Yang 2023 | 0 | 200 | 0 | 200 | 0.00 | [-0.01; 0.01] | 7.0% | 15.0% |
| Zarate 2006 | 0 | 9 | 0 | 9 | 0.00 | [-0.19; 0.19] | 0.4% | 0.0% |
| Zarate 2012 | 0 | 21 | 0 | 21 | 0.00 | [-0.09; 0.09] | 0.8% | 0.2% |
| Zhou 2024 | 0 | 27 | 1 | 27 | -0.04 | [-0.13; 0.06] | 1.0% | 0.2% |
| **Common effect model** | | **2792** | | **2792** | **-0.00** | **[-0.01; 0.01]** | **100.0%** | **.** |
| **Random effects model** | | | | | **0.00** | **[-0.00; 0.00]** | **.** | **100.0%** |

Heterogeneity: $I^2 = 0.0\%$, $\tau^2 = 0$, $p = 1.0000$

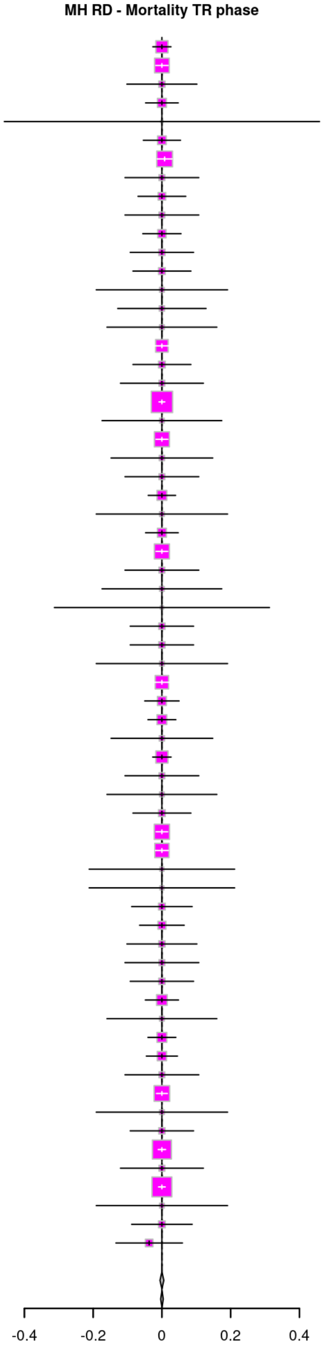

#### Arcsine Risk-Difference

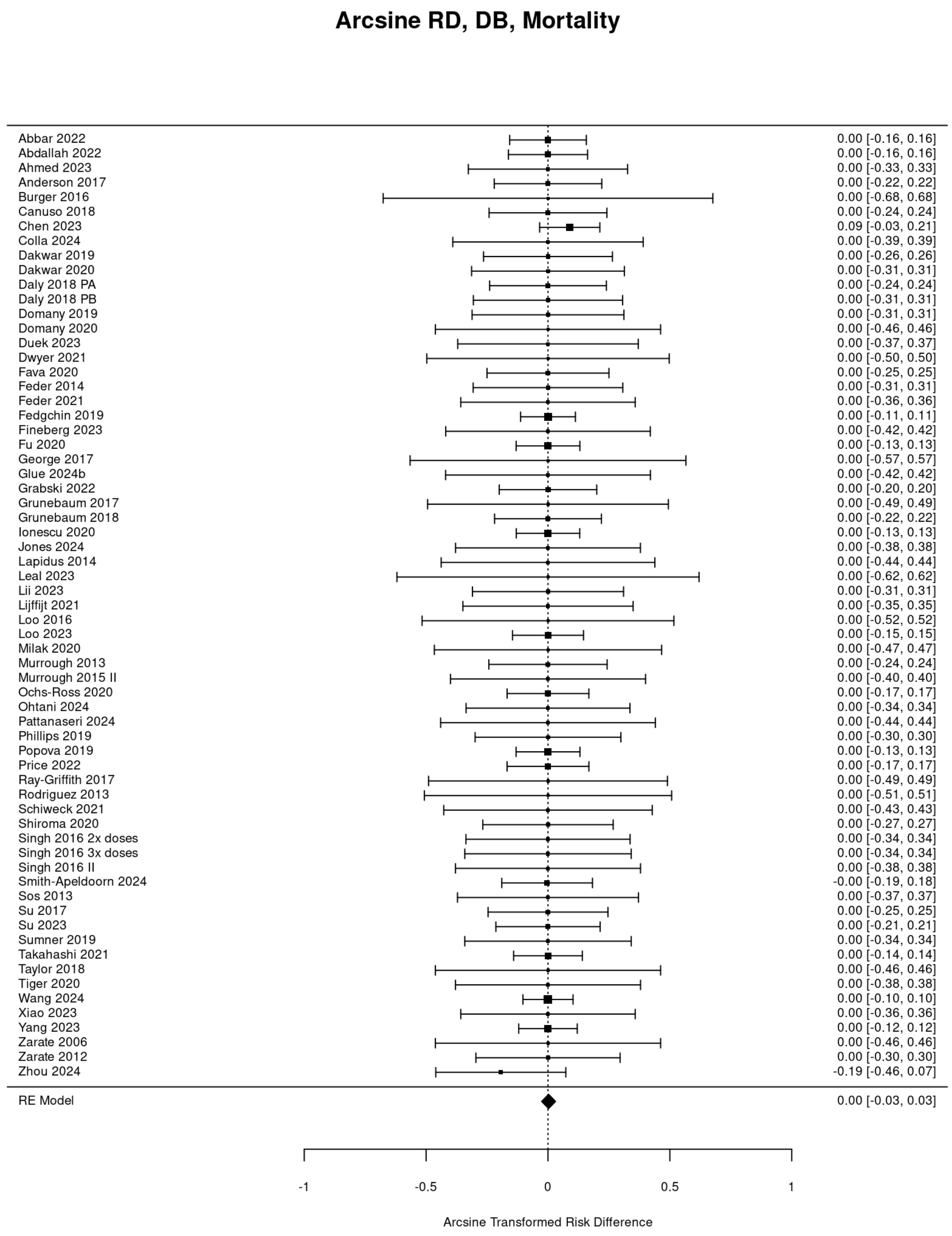

**Bayesian Meta-Analysis, wider prior**

**Bayesian MA, Delta = 250 - Deaths TR**

| Study | Estimate | 95% CI |
|---|---|---|
| Abbar 2022 | 0.13 | [-3.80, 4.06] |
| Abdallah 2022 | -0.67 | [-4.60, 3.26] |
| Ahmed 2023 | 0.00 | [-3.97, 3.97] |
| Anderson 2017 | -0.03 | [-3.97, 3.92] |
| Burger 2016 | 0.76 | [-3.36, 4.88] |
| Canuso 2018 | -0.12 | [-4.07, 3.83] |
| Chen 2023 | 1.11 | [-2.10, 4.32] |
| Colla 2024 | -0.51 | [-4.50, 3.48] |
| Dakwar 2019 | 0.04 | [-3.92, 3.99] |
| Dakwar 2020 | 0.29 | [-3.67, 4.26] |
| Daly 2018 PA | -0.03 | [-3.98, 3.92] |
| Daly 2018 PB | 0.05 | [-3.92, 4.01] |
| Domany 2019 | -0.20 | [-4.16, 3.77] |
| Domany 2020 | 0.00 | [-4.02, 4.02] |
| Duek 2023 | 0.00 | [-3.99, 3.99] |
| Dwyer 2021 | -0.57 | [-4.61, 3.47] |
| Fava 2020 | -1.42 | [-5.37, 2.53] |
| Feder 2014 | -0.14 | [-4.11, 3.82] |
| Feder 2021 | 0.00 | [-3.98, 3.98] |
| Fedgchin 2019 | -0.71 | [-4.64, 3.21] |
| Fineberg 2023 | 0.17 | [-3.83, 4.18] |
| Fu 2020 | -0.01 | [-3.94, 3.92] |
| George 2017 | -1.02 | [-5.09, 3.04] |
| Glue 2024b | -0.72 | [-4.73, 3.28] |
| Grabski 2022 | 0.00 | [-3.94, 3.94] |
| Grunebaum 2017 | -0.24 | [-4.27, 3.80] |
| Grunebaum 2018 | 0.00 | [-3.94, 3.94] |
| Ionescu 2020 | -0.01 | [-3.94, 3.92] |
| Jones 2024 | -0.42 | [-4.41, 3.57] |
| Lapidus 2014 | 0.00 | [-4.01, 4.01] |
| Leal 2023 | 0.00 | [-4.09, 4.09] |
| Lii 2023 | 0.00 | [-3.97, 3.97] |
| Lijffijt 2021 | -0.42 | [-4.40, 3.56] |
| Loo 2016 | -0.38 | [-4.42, 3.67] |
| Loo 2023 | 0.03 | [-3.90, 3.96] |
| Milak 2020 | -1.95 | [-5.97, 2.07] |
| Murrough 2013 | -0.62 | [-4.57, 3.33] |
| Murrough 2015 II | 0.00 | [-4.00, 4.00] |
| Ochs-Ross 2020 | -0.10 | [-4.04, 3.83] |
| Ohtani 2024 | 0.00 | [-3.98, 3.98] |
| Pattanaseri 2024 | -0.19 | [-4.20, 3.82] |
| Phillips 2019 | -0.05 | [-4.01, 3.92] |
| Popova 2019 | -0.05 | [-3.98, 3.88] |
| Price 2022 | -0.70 | [-4.63, 3.24] |
| Ray-Griffith 2017 | 0.00 | [-4.03, 4.03] |
| Rodriguez 2013 | -0.13 | [-4.17, 3.92] |
| Schiweck 2021 | -1.05 | [-5.06, 2.95] |
| Shiroma 2020 | -0.15 | [-4.10, 3.81] |
| Singh 2016 2x doses | -0.11 | [-4.09, 3.86] |
| Singh 2016 3x doses | -0.06 | [-4.04, 3.92] |
| Singh 2016 II | -0.67 | [-4.66, 3.32] |
| Smith-Apeldoorn 2024 | -0.06 | [-2.85, 2.74] |
| Sos 2013 | 0.53 | [-3.46, 4.52] |
| Su 2017 | -0.66 | [-4.61, 3.29] |
| Su 2023 | 0.00 | [-3.94, 3.94] |
| Sumner 2019 | -0.06 | [-4.04, 3.92] |
| Takahashi 2021 | -0.42 | [-4.35, 3.51] |
| Taylor 2018 | 0.00 | [-4.02, 4.02] |
| Tiger 2020 | -0.67 | [-4.66, 3.32] |
| Wang 2024 | 0.00 | [-3.93, 3.93] |
| Xiao 2023 | 0.00 | [-3.98, 3.98] |
| Yang 2023 | -0.69 | [-4.62, 3.24] |
| Zarate 2006 | 0.00 | [-4.02, 4.02] |
| Zarate 2012 | 0.09 | [-3.87, 4.05] |
| Zhou 2024 | -1.14 | [-4.38, 2.11] |
| **Summary** | **-0.14** | **[-1.43, 1.12]** |
| **Prediction** | **-0.14** | **[-1.70, 1.38]** |

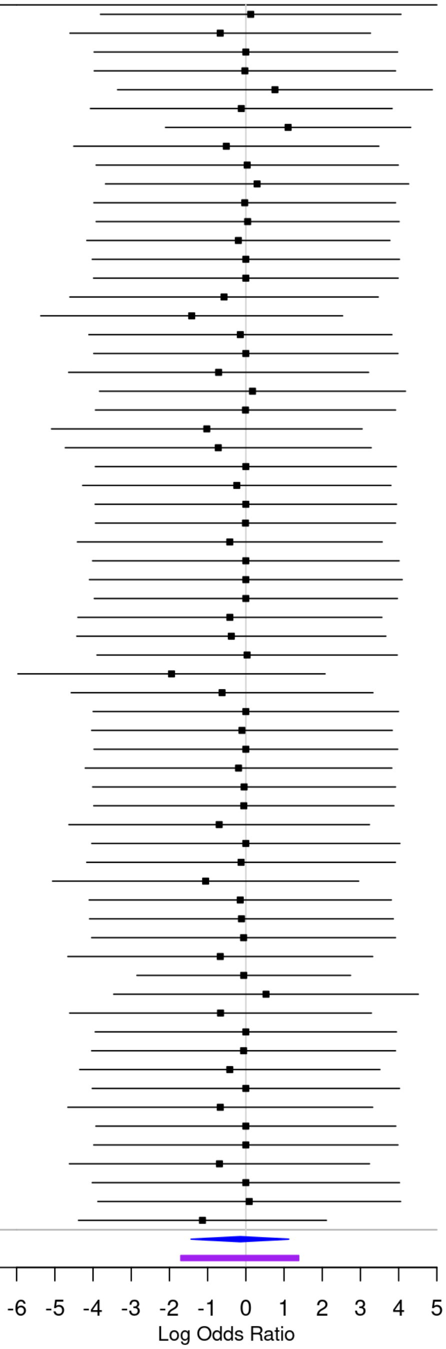

*Heterogeneity (tau): 0.28 [0.00, 0.83]*

**Bayesian Meta-Analysis, smaller prior**

**Bayesian MA, Delta = 15 - Deaths TR**

| Study | Estimate | 95% CI |
|---|---|---|
| Abbar 2022 | 0.13 | [-3.80, 4.06] |
| Abdallah 2022 | -0.67 | [-4.60, 3.26] |
| Ahmed 2023 | 0.00 | [-3.97, 3.97] |
| Anderson 2017 | -0.03 | [-3.97, 3.92] |
| Burger 2016 | 0.76 | [-3.36, 4.88] |
| Canuso 2018 | -0.12 | [-4.07, 3.83] |
| Chen 2023 | 1.11 | [-2.10, 4.32] |
| Colla 2024 | -0.51 | [-4.50, 3.48] |
| Dakwar 2019 | 0.04 | [-3.92, 3.99] |
| Dakwar 2020 | 0.29 | [-3.67, 4.26] |
| Daly 2018 PA | -0.03 | [-3.98, 3.92] |
| Daly 2018 PB | 0.05 | [-3.92, 4.01] |
| Domany 2019 | -0.20 | [-4.16, 3.77] |
| Domany 2020 | 0.00 | [-4.02, 4.02] |
| Duek 2023 | 0.00 | [-3.99, 3.99] |
| Dwyer 2021 | -0.57 | [-4.61, 3.47] |
| Fava 2020 | -1.42 | [-5.37, 2.53] |
| Feder 2014 | -0.14 | [-4.11, 3.82] |
| Feder 2021 | 0.00 | [-3.98, 3.98] |
| Fedgchin 2019 | -0.71 | [-4.64, 3.21] |
| Fineberg 2023 | 0.17 | [-3.83, 4.18] |
| Fu 2020 | -0.01 | [-3.94, 3.92] |
| George 2017 | -1.02 | [-5.09, 3.04] |
| Glue 2024b | -0.72 | [-4.73, 3.28] |
| Grabski 2022 | 0.00 | [-3.94, 3.94] |
| Grunebaum 2017 | -0.24 | [-4.27, 3.80] |
| Grunebaum 2018 | 0.00 | [-3.94, 3.94] |
| Ionescu 2020 | -0.01 | [-3.94, 3.92] |
| Jones 2024 | -0.42 | [-4.41, 3.57] |
| Lapidus 2014 | 0.00 | [-4.01, 4.01] |
| Leal 2023 | 0.00 | [-4.09, 4.09] |
| Lii 2023 | 0.00 | [-3.97, 3.97] |
| Lijffijt 2021 | -0.42 | [-4.40, 3.56] |
| Loo 2016 | -0.38 | [-4.42, 3.67] |
| Loo 2023 | 0.03 | [-3.90, 3.96] |
| Milak 2020 | -1.95 | [-5.97, 2.07] |
| Murrough 2013 | -0.62 | [-4.57, 3.33] |
| Murrough 2015 II | 0.00 | [-4.00, 4.00] |
| Ochs-Ross 2020 | -0.10 | [-4.04, 3.83] |
| Ohtani 2024 | 0.00 | [-3.98, 3.98] |
| Pattanaseri 2024 | -0.19 | [-4.20, 3.82] |
| Phillips 2019 | -0.05 | [-4.01, 3.92] |
| Popova 2019 | -0.05 | [-3.98, 3.88] |
| Price 2022 | -0.70 | [-4.63, 3.24] |
| Ray-Griffith 2017 | 0.00 | [-4.03, 4.03] |
| Rodriguez 2013 | -0.13 | [-4.17, 3.92] |
| Schiweck 2021 | -1.05 | [-5.06, 2.95] |
| Shiroma 2020 | -0.15 | [-4.10, 3.81] |
| Singh 2016 2x doses | -0.11 | [-4.09, 3.86] |
| Singh 2016 3x doses | -0.06 | [-4.04, 3.92] |
| Singh 2016 II | -0.67 | [-4.66, 3.32] |
| Smith-Apeldoorn 2024 | -0.06 | [-2.85, 2.74] |
| Sos 2013 | 0.53 | [-3.46, 4.52] |
| Su 2017 | -0.66 | [-4.61, 3.29] |
| Su 2023 | 0.00 | [-3.94, 3.94] |
| Sumner 2019 | -0.06 | [-4.04, 3.92] |
| Takahashi 2021 | -0.42 | [-4.35, 3.51] |
| Taylor 2018 | 0.00 | [-4.02, 4.02] |
| Tiger 2020 | -0.67 | [-4.66, 3.32] |
| Wang 2024 | 0.00 | [-3.93, 3.93] |
| Xiao 2023 | 0.00 | [-3.98, 3.98] |
| Yang 2023 | -0.69 | [-4.62, 3.24] |
| Zarate 2006 | 0.00 | [-4.02, 4.02] |
| Zarate 2012 | 0.09 | [-3.87, 4.05] |
| Zhou 2024 | -1.14 | [-4.38, 2.11] |
| **Summary** | **-0.13** | **[-1.30, 1.05]** |
| **Prediction** | **-0.12** | **[-1.59, 1.32]** |

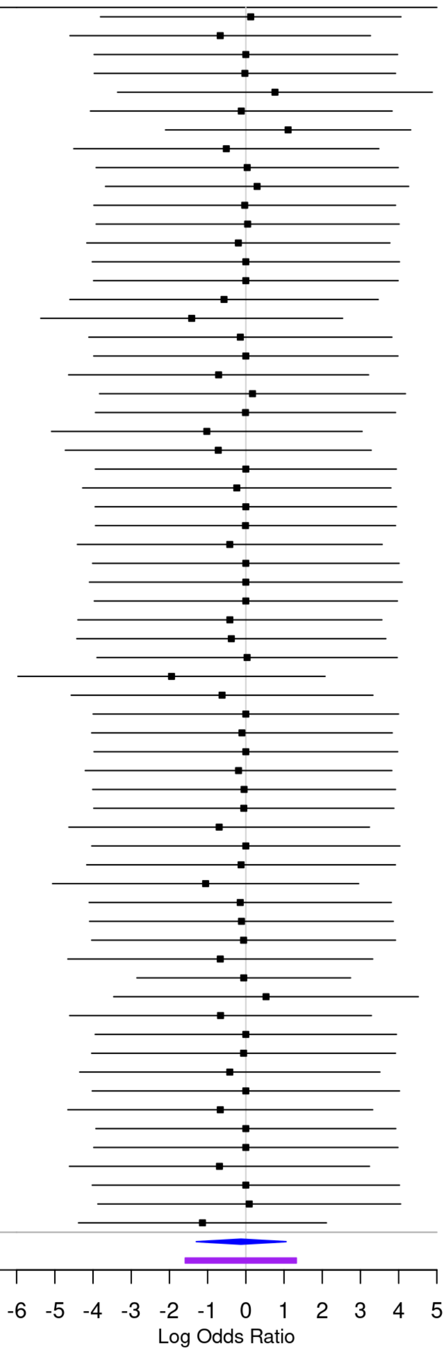

*Heterogeneity (tau): 0.29 [0.00, 0.83]*

**Exact Method**

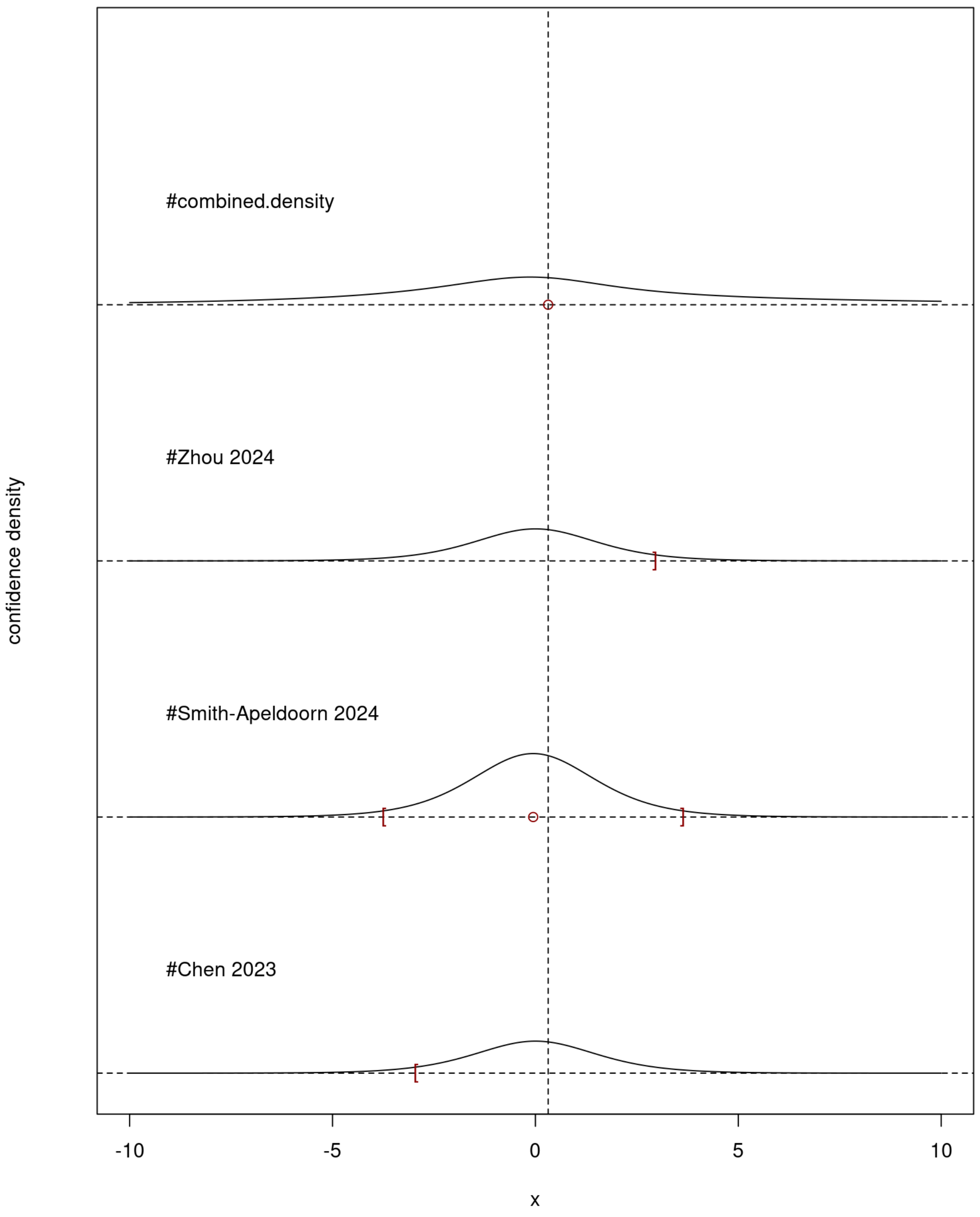

#### Follow-Up Phase

#### Peto's Method

| Study | Experimental Events | Experimental Total | Control Events | Control Total | OR | 95%-CI | Weight (common) | Weight (random) |
|---|---|---|---|---|---|---|---|---|
| Abbar 2022 | 1 | 73 | 0 | 83 | 8.47 | [0.17; 430.52] | 24.9% | 24.9% |
| Ahmed 2023 | 0 | 18 | 0 | 18 | | | 0.0% | 0.0% |
| Anderson 2017 | 0 | 25 | 0 | 23 | | | 0.0% | 0.0% |
| Burger 2016 | 0 | 3 | 0 | 7 | | | 0.0% | 0.0% |
| Canuso 2018 | 0 | 27 | 0 | 22 | | | 0.0% | 0.0% |
| Dakwar 2019 | 0 | 27 | 0 | 28 | | | 0.0% | 0.0% |
| Dakwar 2020 | 0 | 17 | 0 | 23 | | | 0.0% | 0.0% |
| Domany 2019 | 0 | 22 | 0 | 18 | | | 0.0% | 0.0% |
| Domany 2020 | 0 | 9 | 0 | 9 | | | 0.0% | 0.0% |
| Duek 2023 | 0 | 14 | 0 | 14 | | | 0.0% | 0.0% |
| Dwyer 2021 | 0 | 11 | 0 | 6 | | | 0.0% | 0.0% |
| Fava 2020 | 0 | 80 | 0 | 19 | | | 0.0% | 0.0% |
| Feder 2014 | 0 | 22 | 0 | 19 | | | 0.0% | 0.0% |
| Fineberg 2023 | 0 | 10 | 0 | 12 | | | 0.0% | 0.0% |
| Fu 2020 | 1 | 101 | 0 | 91 | 6.69 | [0.13; 339.08] | 25.0% | 25.0% |
| Glue 2024b | 0 | 17 | 0 | 8 | | | 0.0% | 0.0% |
| Grabski 2022 | 0 | 48 | 0 | 48 | | | 0.0% | 0.0% |
| Ionescu 2020 | 0 | 89 | 0 | 94 | | | 0.0% | 0.0% |
| Jones 2024 | 0 | 17 | 0 | 11 | | | 0.0% | 0.0% |
| Lapidus 2014 | 0 | 10 | 0 | 10 | | | 0.0% | 0.0% |
| Leal 2023 | 0 | 5 | 0 | 5 | | | 0.0% | 0.0% |
| Lii 2023 | 1 | 20 | 0 | 20 | 7.39 | [0.15; 372.38] | 25.0% | 25.0% |
| Lijffijt 2021 | 0 | 20 | 0 | 13 | | | 0.0% | 0.0% |
| Murrough 2013 | 0 | 47 | 0 | 25 | | | 0.0% | 0.0% |
| Murrough 2015 II | 1 | 12 | 0 | 12 | 7.39 | [0.15; 372.38] | 25.0% | 25.0% |
| Ohtani 2024 | 0 | 17 | 0 | 17 | | | 0.0% | 0.0% |
| Pattanaseri 2024 | 0 | 11 | 0 | 9 | | | 0.0% | 0.0% |
| Phillips 2019 | 0 | 22 | 0 | 21 | | | 0.0% | 0.0% |
| Price 2022 | 0 | 103 | 0 | 51 | | | 0.0% | 0.0% |
| Ray-Griffith 2017 | 0 | 8 | 0 | 8 | | | 0.0% | 0.0% |
| Rodriguez 2013 | 0 | 8 | 0 | 7 | | | 0.0% | 0.0% |
| Schiweck 2021 | 0 | 21 | 0 | 7 | | | 0.0% | 0.0% |
| Smith-Apeldoorn 2024 | 0 | 49 | 0 | 48 | | | 0.0% | 0.0% |
| Sos 2013 | 0 | 11 | 0 | 19 | | | 0.0% | 0.0% |
| Su 2017 | 0 | 47 | 0 | 24 | | | 0.0% | 0.0% |
| Su 2023 | 0 | 42 | 0 | 42 | | | 0.0% | 0.0% |
| Sumner 2019 | 0 | 17 | 0 | 16 | | | 0.0% | 0.0% |
| Taylor 2018 | 0 | 9 | 0 | 8 | | | 0.0% | 0.0% |
| Wang 2024 | 0 | 182 | 0 | 182 | | | 0.0% | 0.0% |
| Xiao 2023 | 0 | 15 | 0 | 15 | | | 0.0% | 0.0% |
| Yang 2023 | 0 | 200 | 0 | 100 | | | 0.0% | 0.0% |
| Zarate 2006 | 0 | 9 | 0 | 9 | | | 0.0% | 0.0% |
| Zarate 2012 | 0 | 21 | 0 | 23 | | | 0.0% | 0.0% |
| Zhou 2024 | 0 | 27 | 0 | 25 | | | 0.0% | 0.0% |
| **Common effect model** | | **1563** | | **1269** | **7.46** | **[1.05; 53.04]** | **100.0%** | **.** |
| **Random effects model** | | | | | **7.46** | **[1.05; 53.04]** | **.** | **100.0%** |

Heterogeneity: $I^2 = 0.0\%$, $\tau^2 = 0$, $p = 0.9998$

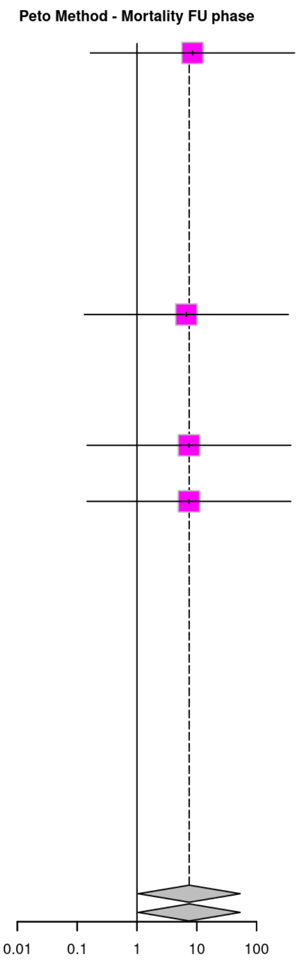

#### Maentel Mantel-Haenszel with CC

| Study | Experimental Events | Experimental Total | Control Events | Control Total | OR | 95%-CI | Weight (common) | Weight (random) |
|---|---|---|---|---|---|---|---|---|
| Abbar 2022 | 1 | 73 | 0 | 83 | 3.46 | [0.14; 86.13] | 2.0% | 3.3% |
| Ahmed 2023 | 0 | 18 | 0 | 18 | 1.00 | [0.02; 53.12] | 2.1% | 2.2% |
| Anderson 2017 | 0 | 25 | 0 | 23 | 0.92 | [0.02; 48.33] | 2.2% | 2.2% |
| Burger 2016 | 0 | 3 | 0 | 7 | 2.14 | [0.03; 131.93] | 1.3% | 2.0% |
| Canuso 2018 | 0 | 27 | 0 | 22 | 0.82 | [0.02; 42.89] | 2.4% | 2.2% |
| Dakwar 2019 | 0 | 27 | 0 | 28 | 1.04 | [0.02; 54.08] | 2.1% | 2.2% |
| Dakwar 2020 | 0 | 17 | 0 | 23 | 1.34 | [0.03; 71.04] | 1.8% | 2.2% |
| Domany 2019 | 0 | 22 | 0 | 18 | 0.82 | [0.02; 43.47] | 2.3% | 2.2% |
| Domany 2020 | 0 | 9 | 0 | 9 | 1.00 | [0.02; 55.80] | 2.1% | 2.1% |
| Duek 2023 | 0 | 14 | 0 | 14 | 1.00 | [0.02; 53.89] | 2.1% | 2.2% |
| Dwyer 2021 | 0 | 11 | 0 | 6 | 0.57 | [0.01; 32.00] | 2.7% | 2.1% |
| Fava 2020 | 0 | 80 | 0 | 19 | 0.24 | [0.00; 12.59] | 3.5% | 2.2% |
| Feder 2014 | 0 | 22 | 0 | 19 | 0.87 | [0.02; 45.76] | 2.3% | 2.2% |
| Fineberg 2023 | 0 | 10 | 0 | 12 | 1.19 | [0.02; 65.32] | 1.9% | 2.1% |
| Fu 2020 | 1 | 101 | 0 | 91 | 2.73 | [0.11; 67.89] | 2.3% | 3.3% |
| Glue 2024b | 0 | 17 | 0 | 8 | 0.49 | [0.01; 26.64] | 2.8% | 2.1% |
| Grabski 2022 | 0 | 48 | 0 | 48 | 1.00 | [0.02; 51.42] | 2.2% | 2.2% |
| Ionescu 2020 | 0 | 89 | 0 | 94 | 1.06 | [0.02; 53.78] | 2.1% | 2.2% |
| Jones 2024 | 0 | 17 | 0 | 11 | 0.66 | [0.01; 35.52] | 2.6% | 2.2% |
| Lapidus 2014 | 0 | 10 | 0 | 10 | 1.00 | [0.02; 55.27] | 2.1% | 2.1% |
| Leal 2023 | 0 | 5 | 0 | 5 | 1.00 | [0.02; 59.99] | 2.0% | 2.1% |
| Lii 2023 | 1 | 20 | 0 | 20 | 3.15 | [0.12; 82.16] | 2.0% | 3.2% |
| Lijffijt 2021 | 0 | 20 | 0 | 13 | 0.66 | [0.01; 35.23] | 2.6% | 2.2% |
| Murrough 2013 | 0 | 47 | 0 | 25 | 0.54 | [0.01; 27.86] | 2.8% | 2.2% |
| Murrough 2015 II | 1 | 12 | 0 | 12 | 3.26 | [0.12; 88.35] | 1.9% | 3.2% |
| Ohtani 2024 | 0 | 17 | 0 | 17 | 1.00 | [0.02; 53.28] | 2.1% | 2.2% |
| Pattanaseri 2024 | 0 | 11 | 0 | 9 | 0.83 | [0.01; 45.69] | 2.3% | 2.1% |
| Phillips 2019 | 0 | 22 | 0 | 21 | 0.96 | [0.02; 50.34] | 2.2% | 2.2% |
| Price 2022 | 0 | 103 | 0 | 51 | 0.50 | [0.01; 25.44] | 2.9% | 2.2% |
| Ray-Griffith 2017 | 0 | 8 | 0 | 8 | 1.00 | [0.02; 56.46] | 2.1% | 2.1% |
| Rodriguez 2013 | 0 | 8 | 0 | 7 | 0.88 | [0.02; 50.19] | 2.2% | 2.1% |
| Schiweck 2021 | 0 | 21 | 0 | 7 | 0.35 | [0.01; 19.18] | 3.1% | 2.1% |
| Smith-Apeldoorn 2024 | 0 | 49 | 0 | 48 | 0.98 | [0.02; 50.37] | 2.2% | 2.2% |
| Sos 2013 | 0 | 11 | 0 | 19 | 1.70 | [0.03; 91.39] | 1.6% | 2.2% |
| Su 2017 | 0 | 47 | 0 | 24 | 0.52 | [0.01; 26.79] | 2.9% | 2.2% |
| Su 2023 | 0 | 42 | 0 | 42 | 1.00 | [0.02; 51.57] | 2.2% | 2.2% |
| Sumner 2019 | 0 | 17 | 0 | 16 | 0.94 | [0.02; 50.32] | 2.2% | 2.2% |
| Taylor 2018 | 0 | 9 | 0 | 8 | 0.89 | [0.02; 50.22] | 2.2% | 2.1% |
| Wang 2024 | 0 | 182 | 0 | 182 | 1.00 | [0.02; 50.67] | 2.2% | 2.2% |
| Xiao 2023 | 0 | 15 | 0 | 15 | 1.00 | [0.02; 53.66] | 2.1% | 2.2% |
| Yang 2023 | 0 | 200 | 0 | 100 | 0.50 | [0.01; 25.45] | 2.9% | 2.2% |
| Zarate 2006 | 0 | 9 | 0 | 9 | 1.00 | [0.02; 55.80] | 2.1% | 2.1% |
| Zarate 2012 | 0 | 21 | 0 | 23 | 1.09 | [0.02; 57.53] | 2.0% | 2.2% |
| Zhou 2024 | 0 | 27 | 0 | 25 | 0.93 | [0.02; 48.49] | 2.2% | 2.2% |
| **Common effect model** | | **1563** | | **1269** | **1.04** | **[0.59; 1.85]** | **100.0%** | **.** |
| **Random effects model** | | | | | **1.00** | **[0.56; 1.81]** | **.** | **100.0%** |

Heterogeneity: $I^2 = 0.0\%$, $\tau^2 = 0$, $p = 1.0000$

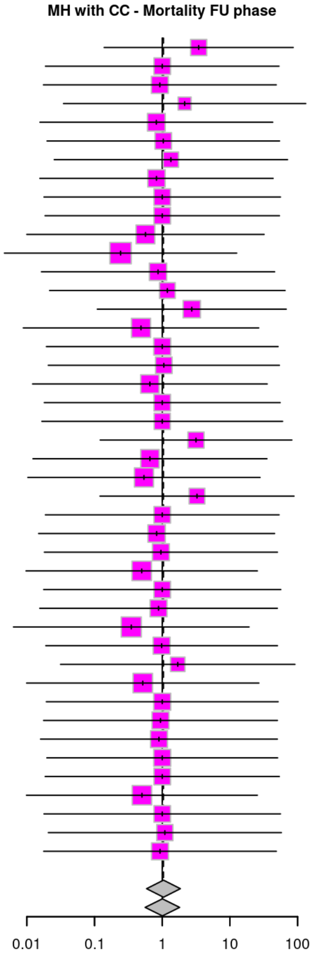

#### Maentel Mantel-Haenszel with Treatment-Arm CC

| Study | Experimental Events | Experimental Total | Control Events | Control Total | MH with TA-CC - Mortality FU phase | OR | 95%-CI | Weight (common) | Weight (random) |
|---|---|---|---|---|---|---|---|---|---|
| Abbar 2022 | 1 | 73 | 0 | 83 | | 3.18 | [0.14; 74.37] | 2.4% | 3.6% |
| Ahmed 2023 | 0 | 18 | 0 | 18 | | 1.00 | [0.02; 53.12] | 2.4% | 2.3% |
| Anderson 2017 | 0 | 25 | 0 | 23 | | 1.00 | [0.02; 52.62] | 2.4% | 2.3% |
| Burger 2016 | 0 | 3 | 0 | 7 | | 1.00 | [0.01; 87.11] | 1.9% | 1.8% |
| Canuso 2018 | 0 | 27 | 0 | 22 | | 1.00 | [0.02; 53.50] | 2.4% | 2.3% |
| Dakwar 2019 | 0 | 27 | 0 | 28 | | 1.00 | [0.02; 52.22] | 2.4% | 2.3% |
| Dakwar 2020 | 0 | 17 | 0 | 23 | | 1.00 | [0.02; 55.30] | 2.3% | 2.2% |
| Domany 2019 | 0 | 22 | 0 | 18 | | 1.00 | [0.02; 53.92] | 2.4% | 2.2% |
| Domany 2020 | 0 | 9 | 0 | 9 | | 1.00 | [0.02; 55.80] | 2.3% | 2.2% |
| Duek 2023 | 0 | 14 | 0 | 14 | | 1.00 | [0.02; 53.89] | 2.4% | 2.2% |
| Dwyer 2021 | 0 | 11 | 0 | 6 | | 1.00 | [0.01; 67.61] | 2.1% | 2.0% |
| Fava 2020 | 0 | 80 | 0 | 19 | | 1.00 | [0.01; 148.67] | 1.5% | 1.4% |
| Feder 2014 | 0 | 22 | 0 | 19 | | 1.00 | [0.02; 53.36] | 2.4% | 2.3% |
| Fineberg 2023 | 0 | 10 | 0 | 12 | | 1.00 | [0.02; 55.75] | 2.3% | 2.2% |
| Fu 2020 | 1 | 101 | 0 | 91 | | 2.93 | [0.11; 77.20] | 2.4% | 3.3% |
| Glue 2024b | 0 | 17 | 0 | 8 | | 1.00 | [0.01; 72.36] | 2.0% | 1.9% |
| Grabski 2022 | 0 | 48 | 0 | 48 | | 1.00 | [0.02; 51.42] | 2.4% | 2.3% |
| Ionescu 2020 | 0 | 89 | 0 | 94 | | 1.00 | [0.02; 51.01] | 2.4% | 2.3% |
| Jones 2024 | 0 | 17 | 0 | 11 | | 1.00 | [0.02; 59.25] | 2.3% | 2.1% |
| Lapidus 2014 | 0 | 10 | 0 | 10 | | 1.00 | [0.02; 55.27] | 2.3% | 2.2% |
| Leal 2023 | 0 | 5 | 0 | 5 | | 1.00 | [0.02; 59.99] | 2.2% | 2.1% |
| Lii 2023 | 1 | 20 | 0 | 20 | | 3.15 | [0.12; 82.16] | 2.3% | 3.4% |
| Lijffijt 2021 | 0 | 20 | 0 | 13 | | 1.00 | [0.02; 58.54] | 2.3% | 2.2% |
| Murrough 2013 | 0 | 47 | 0 | 25 | | 1.00 | [0.02; 63.11] | 2.2% | 2.1% |
| Murrough 2015 II | 1 | 12 | 0 | 12 | | 3.26 | [0.12; 88.35] | 2.2% | 3.3% |
| Ohtani 2024 | 0 | 17 | 0 | 17 | | 1.00 | [0.02; 53.28] | 2.4% | 2.3% |
| Pattanaseri 2024 | 0 | 11 | 0 | 9 | | 1.00 | [0.02; 56.40] | 2.3% | 2.2% |
| Phillips 2019 | 0 | 22 | 0 | 21 | | 1.00 | [0.02; 52.74] | 2.4% | 2.3% |
| Price 2022 | 0 | 103 | 0 | 51 | | 1.00 | [0.02; 65.23] | 2.2% | 2.0% |
| Ray-Griffith 2017 | 0 | 8 | 0 | 8 | | 1.00 | [0.02; 56.46] | 2.3% | 2.2% |
| Rodriguez 2013 | 0 | 8 | 0 | 7 | | 1.00 | [0.02; 57.37] | 2.3% | 2.2% |
| Schiweck 2021 | 0 | 21 | 0 | 7 | | 1.00 | [0.01; 99.86] | 1.8% | 1.7% |
| Smith-Apeldoorn 2024 | 0 | 49 | 0 | 48 | | 1.00 | [0.02; 51.42] | 2.4% | 2.3% |
| Sos 2013 | 0 | 11 | 0 | 19 | | 1.00 | [0.02; 62.32] | 2.2% | 2.1% |
| Su 2017 | 0 | 47 | 0 | 24 | | 1.00 | [0.02; 64.85] | 2.2% | 2.1% |
| Su 2023 | 0 | 42 | 0 | 42 | | 1.00 | [0.02; 51.57] | 2.4% | 2.3% |
| Sumner 2019 | 0 | 17 | 0 | 16 | | 1.00 | [0.02; 53.46] | 2.4% | 2.3% |
| Taylor 2018 | 0 | 9 | 0 | 8 | | 1.00 | [0.02; 56.50] | 2.3% | 2.2% |
| Wang 2024 | 0 | 182 | 0 | 182 | | 1.00 | [0.02; 50.67] | 2.4% | 2.3% |
| Xiao 2023 | 0 | 15 | 0 | 15 | | 1.00 | [0.02; 53.66] | 2.4% | 2.3% |
| Yang 2023 | 0 | 200 | 0 | 100 | | 1.00 | [0.02; 64.37] | 2.2% | 2.1% |
| Zarate 2006 | 0 | 9 | 0 | 9 | | 1.00 | [0.02; 55.80] | 2.3% | 2.2% |
| Zarate 2012 | 0 | 21 | 0 | 23 | | 1.00 | [0.02; 52.84] | 2.4% | 2.3% |
| Zhou 2024 | 0 | 27 | 0 | 25 | | 1.00 | [0.02; 52.44] | 2.4% | 2.3% |
| **Common effect model** | | **1563** | | **1269** | | **1.20** | **[0.66; 2.15]** | **100.0%** | **.** |
| **Random effects model** | | | | | | **1.17** | **[0.64; 2.12]** | **.** | **100.0%** |

Heterogeneity: $I^2 = 0.0\%$, $\tau^2 = 0$, $p = 1.0000$

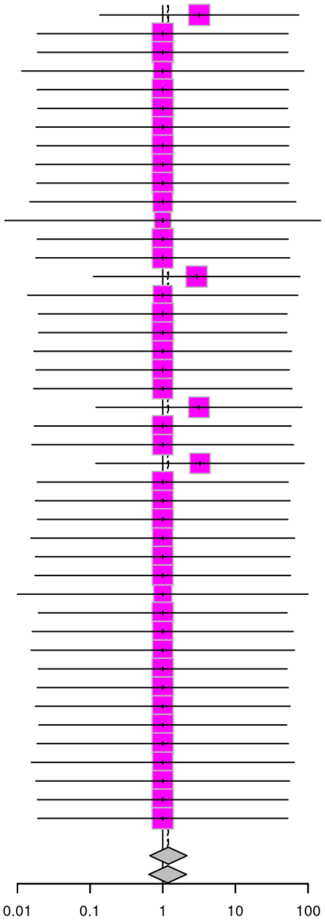

#### Maentel Mantel-Haenszel Risk-Difference

| Study | Experimental Events | Experimental Total | Control Events | Control Total | RD | 95%-CI | Weight (common) | Weight (random) |
|---|---|---|---|---|---|---|---|---|
| Abbar 2022 | 1 | 73 | 0 | 83 | 0.01 | [-0.02; 0.05] | 5.6% | 3.3% |
| Ahmed 2023 | 0 | 18 | 0 | 18 | 0.00 | [-0.10; 0.10] | 1.4% | 0.4% |
| Anderson 2017 | 0 | 25 | 0 | 23 | 0.00 | [-0.08; 0.08] | 1.8% | 0.7% |
| Burger 2016 | 0 | 3 | 0 | 7 | 0.00 | [-0.36; 0.36] | 0.4% | 0.0% |
| Canuso 2018 | 0 | 27 | 0 | 22 | 0.00 | [-0.08; 0.08] | 1.8% | 0.7% |
| Dakwar 2019 | 0 | 27 | 0 | 28 | 0.00 | [-0.07; 0.07] | 2.0% | 0.9% |
| Dakwar 2020 | 0 | 17 | 0 | 23 | 0.00 | [-0.10; 0.10] | 1.5% | 0.5% |
| Domany 2019 | 0 | 22 | 0 | 18 | 0.00 | [-0.09; 0.09] | 1.5% | 0.5% |
| Domany 2020 | 0 | 9 | 0 | 9 | 0.00 | [-0.19; 0.19] | 0.7% | 0.1% |
| Duek 2023 | 0 | 14 | 0 | 14 | 0.00 | [-0.13; 0.13] | 1.1% | 0.3% |
| Dwyer 2021 | 0 | 11 | 0 | 6 | 0.00 | [-0.22; 0.22] | 0.6% | 0.1% |
| Fava 2020 | 0 | 80 | 0 | 19 | 0.00 | [-0.07; 0.07] | 2.3% | 0.9% |
| Feder 2014 | 0 | 22 | 0 | 19 | 0.00 | [-0.09; 0.09] | 1.5% | 0.5% |
| Fineberg 2023 | 0 | 10 | 0 | 12 | 0.00 | [-0.16; 0.16] | 0.9% | 0.2% |
| Fu 2020 | 1 | 101 | 0 | 91 | 0.01 | [-0.02; 0.04] | 6.9% | 5.6% |
| Glue 2024b | 0 | 17 | 0 | 8 | 0.00 | [-0.17; 0.17] | 0.9% | 0.2% |
| Grabski 2022 | 0 | 48 | 0 | 48 | 0.00 | [-0.04; 0.04] | 3.5% | 2.7% |
| Ionescu 2020 | 0 | 89 | 0 | 94 | 0.00 | [-0.02; 0.02] | 6.6% | 9.6% |
| Jones 2024 | 0 | 17 | 0 | 11 | 0.00 | [-0.14; 0.14] | 1.0% | 0.2% |
| Lapidus 2014 | 0 | 10 | 0 | 10 | 0.00 | [-0.17; 0.17] | 0.8% | 0.1% |
| Leal 2023 | 0 | 5 | 0 | 5 | 0.00 | [-0.31; 0.31] | 0.4% | 0.0% |
| Lii 2023 | 1 | 20 | 0 | 20 | 0.05 | [-0.08; 0.18] | 1.5% | 0.3% |
| Lijffijt 2021 | 0 | 20 | 0 | 13 | 0.00 | [-0.12; 0.12] | 1.2% | 0.3% |
| Murrough 2013 | 0 | 47 | 0 | 25 | 0.00 | [-0.06; 0.06] | 2.4% | 1.2% |
| Murrough 2015 II | 1 | 12 | 0 | 12 | 0.08 | [-0.12; 0.29] | 0.9% | 0.1% |
| Ohtani 2024 | 0 | 17 | 0 | 17 | 0.00 | [-0.11; 0.11] | 1.3% | 0.4% |
| Pattanaseri 2024 | 0 | 11 | 0 | 9 | 0.00 | [-0.18; 0.18] | 0.8% | 0.1% |
| Phillips 2019 | 0 | 22 | 0 | 21 | 0.00 | [-0.09; 0.09] | 1.6% | 0.6% |
| Price 2022 | 0 | 103 | 0 | 51 | 0.00 | [-0.03; 0.03] | 5.0% | 4.9% |
| Ray-Griffith 2017 | 0 | 8 | 0 | 8 | 0.00 | [-0.21; 0.21] | 0.6% | 0.1% |
| Rodriguez 2013 | 0 | 8 | 0 | 7 | 0.00 | [-0.22; 0.22] | 0.6% | 0.1% |
| Schiweck 2021 | 0 | 21 | 0 | 7 | 0.00 | [-0.18; 0.18] | 0.8% | 0.1% |
| Smith-Apeldoorn 2024 | 0 | 49 | 0 | 48 | 0.00 | [-0.04; 0.04] | 3.5% | 2.8% |
| Sos 2013 | 0 | 11 | 0 | 19 | 0.00 | [-0.13; 0.13] | 1.1% | 0.2% |
| Su 2017 | 0 | 47 | 0 | 24 | 0.00 | [-0.06; 0.06] | 2.3% | 1.1% |
| Su 2023 | 0 | 42 | 0 | 42 | 0.00 | [-0.05; 0.05] | 3.1% | 2.1% |
| Sumner 2019 | 0 | 17 | 0 | 16 | 0.00 | [-0.11; 0.11] | 1.2% | 0.4% |
| Taylor 2018 | 0 | 9 | 0 | 8 | 0.00 | [-0.20; 0.20] | 0.7% | 0.1% |
| Wang 2024 | 0 | 182 | 0 | 182 | 0.00 | [-0.01; 0.01] | 13.1% | 37.5% |
| Xiao 2023 | 0 | 15 | 0 | 15 | 0.00 | [-0.12; 0.12] | 1.1% | 0.3% |
| Yang 2023 | 0 | 200 | 0 | 100 | 0.00 | [-0.02; 0.02] | 9.6% | 18.3% |
| Zarate 2006 | 0 | 9 | 0 | 9 | 0.00 | [-0.19; 0.19] | 0.7% | 0.1% |
| Zarate 2012 | 0 | 21 | 0 | 23 | 0.00 | [-0.08; 0.08] | 1.6% | 0.6% |
| Zhou 2024 | 0 | 27 | 0 | 25 | 0.00 | [-0.07; 0.07] | 1.9% | 0.8% |
| **Common effect model** | | **1563** | | **1269** | **0.00** | **[-0.01; 0.01]** | **100.0%** | **.** |
| **Random effects model** | | | | | **0.00** | **[-0.01; 0.01]** | **.** | **100.0%** |

Heterogeneity: $I^2 = 0.0\%$, $\tau^2 = 0$, $p = 1.0000$

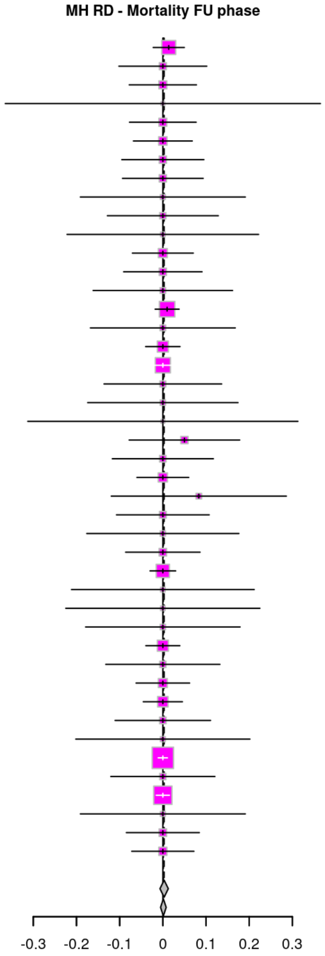

#### Arcsine Risk-Difference

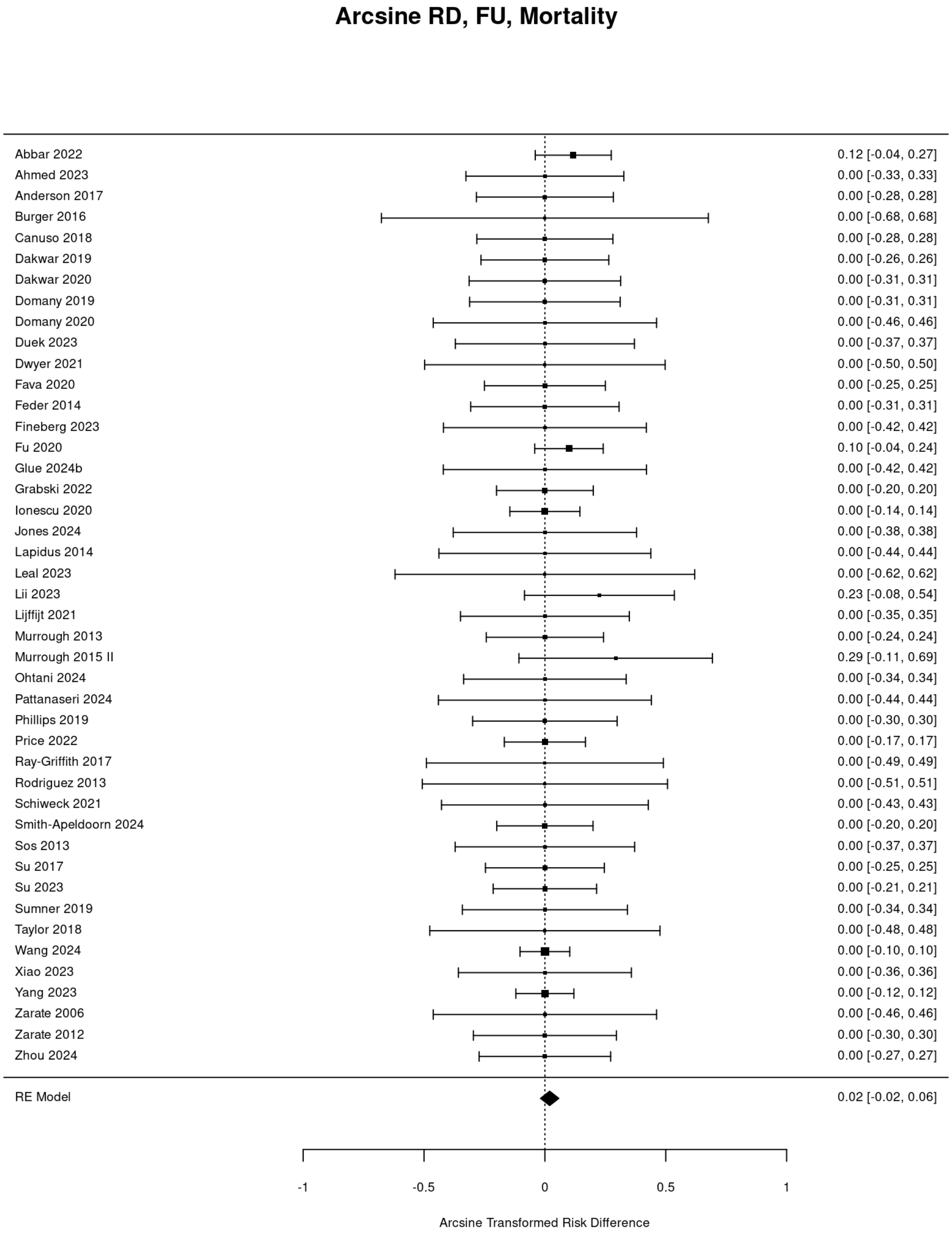

#### Bayesian Meta-Analysis, wider prior

| Study | Experimental Events | Experimental Total | Control Events | Control Total | OR | 95%-CI | Weight (common) | Weight (random) |
|---|---|---|---|---|---|---|---|---|
| Abbar 2022 | 0 | 73 | 0 | 73 | 1.00 | [0.02; 51.07] | 1.5% | 1.5% |
| Abdallah 2022 | 0 | 108 | 0 | 108 | 1.00 | [0.02; 50.85] | 1.5% | 1.5% |
| Ahmed 2023 | 0 | 18 | 0 | 18 | 1.00 | [0.02; 53.12] | 1.5% | 1.5% |
| Anderson 2017 | 0 | 40 | 0 | 40 | 1.00 | [0.02; 51.63] | 1.5% | 1.5% |
| Burger 2016 | 0 | 3 | 0 | 3 | 1.00 | [0.02; 66.06] | 1.3% | 1.3% |
| Canuso 2018 | 0 | 35 | 0 | 35 | 1.00 | [0.02; 51.80] | 1.5% | 1.5% |
| Chen 2023 | 1 | 126 | 0 | 126 | 3.02 | [0.12; 74.94] | 1.5% | 2.3% |
| Colla 2024 | 0 | 17 | 0 | 17 | 1.00 | [0.02; 53.28] | 1.5% | 1.5% |
| Dakwar 2019 | 0 | 27 | 0 | 27 | 1.00 | [0.02; 52.22] | 1.5% | 1.5% |
| Dakwar 2020 | 0 | 17 | 0 | 17 | 1.00 | [0.02; 53.28] | 1.5% | 1.5% |
| Daly 2018 PA | 0 | 34 | 0 | 34 | 1.00 | [0.02; 51.84] | 1.5% | 1.5% |
| Daly 2018 PB | 0 | 20 | 0 | 20 | 1.00 | [0.02; 52.85] | 1.5% | 1.5% |
| Domany 2019 | 0 | 22 | 0 | 22 | 1.00 | [0.02; 52.63] | 1.5% | 1.5% |
| Domany 2020 | 0 | 9 | 0 | 9 | 1.00 | [0.02; 55.80] | 1.4% | 1.5% |
| Duek 2023 | 0 | 14 | 0 | 14 | 1.00 | [0.02; 53.89] | 1.5% | 1.5% |
| Dwyer 2021 | 0 | 11 | 0 | 11 | 1.00 | [0.02; 54.83] | 1.4% | 1.5% |
| Fava 2020 | 0 | 80 | 0 | 80 | 1.00 | [0.02; 51.01] | 1.5% | 1.5% |
| Feder 2014 | 0 | 22 | 0 | 22 | 1.00 | [0.02; 52.63] | 1.5% | 1.5% |
| Feder 2021 | 0 | 15 | 0 | 15 | 1.00 | [0.02; 53.66] | 1.5% | 1.5% |
| Fedgchin 2019 | 0 | 231 | 0 | 231 | 1.00 | [0.02; 50.61] | 1.5% | 1.5% |
| Fineberg 2023 | 0 | 10 | 0 | 10 | 1.00 | [0.02; 55.27] | 1.4% | 1.5% |
| Fu 2020 | 0 | 113 | 0 | 113 | 1.00 | [0.02; 50.83] | 1.5% | 1.5% |
| George 2017 | 0 | 12 | 0 | 12 | 1.00 | [0.02; 54.46] | 1.5% | 1.5% |
| Glue 2024b | 0 | 17 | 0 | 17 | 1.00 | [0.02; 53.28] | 1.5% | 1.5% |
| Grabski 2022 | 0 | 48 | 0 | 48 | 1.00 | [0.02; 51.42] | 1.5% | 1.5% |
| Grunebaum 2017 | 0 | 9 | 0 | 9 | 1.00 | [0.02; 55.80] | 1.4% | 1.5% |
| Grunebaum 2018 | 0 | 40 | 0 | 40 | 1.00 | [0.02; 51.63] | 1.5% | 1.5% |
| Ionescu 2020 | 0 | 114 | 0 | 114 | 1.00 | [0.02; 50.83] | 1.5% | 1.5% |
| Jones 2024 | 0 | 17 | 0 | 17 | 1.00 | [0.02; 53.28] | 1.5% | 1.5% |
| Lapidus 2014 | 0 | 10 | 0 | 10 | 1.00 | [0.02; 55.27] | 1.4% | 1.5% |
| Leal 2023 | 0 | 5 | 0 | 5 | 1.00 | [0.02; 59.99] | 1.4% | 1.4% |
| Lii 2023 | 0 | 20 | 0 | 20 | 1.00 | [0.02; 52.85] | 1.5% | 1.5% |
| Lijffijt 2021 | 0 | 20 | 0 | 20 | 1.00 | [0.02; 52.85] | 1.5% | 1.5% |
| Loo 2016 | 0 | 9 | 0 | 9 | 1.00 | [0.02; 55.80] | 1.4% | 1.5% |
| Loo 2023 | 0 | 89 | 0 | 89 | 1.00 | [0.02; 50.95] | 1.5% | 1.5% |
| Milak 2020 | 0 | 38 | 0 | 38 | 1.00 | [0.02; 51.69] | 1.5% | 1.5% |
| Murrough 2013 | 0 | 47 | 0 | 47 | 1.00 | [0.02; 51.44] | 1.5% | 1.5% |
| Murrough 2015 II | 0 | 12 | 0 | 12 | 1.00 | [0.02; 54.46] | 1.5% | 1.5% |
| Ochs-Ross 2020 | 0 | 72 | 0 | 72 | 1.00 | [0.02; 51.08] | 1.5% | 1.5% |
| Ohtani 2024 | 0 | 17 | 0 | 17 | 1.00 | [0.02; 53.28] | 1.5% | 1.5% |
| Pattanaseri 2024 | 0 | 11 | 0 | 11 | 1.00 | [0.02; 54.83] | 1.4% | 1.5% |
| Phillips 2019 | 0 | 22 | 0 | 22 | 1.00 | [0.02; 52.63] | 1.5% | 1.5% |
| Popova 2019 | 0 | 115 | 0 | 115 | 1.00 | [0.02; 50.83] | 1.5% | 1.5% |
| Price 2022 | 0 | 103 | 0 | 103 | 1.00 | [0.02; 50.88] | 1.5% | 1.5% |
| Ray-Griffith 2017 | 0 | 8 | 0 | 8 | 1.00 | [0.02; 56.46] | 1.4% | 1.4% |
| Rodriguez 2013 | 0 | 8 | 0 | 8 | 1.00 | [0.02; 56.46] | 1.4% | 1.4% |
| Schiweck 2021 | 0 | 21 | 0 | 21 | 1.00 | [0.02; 52.73] | 1.5% | 1.5% |
| Shiroma 2020 | 0 | 29 | 0 | 29 | 1.00 | [0.02; 52.09] | 1.5% | 1.5% |
| Singh 2016 2x doses | 0 | 18 | 0 | 18 | 1.00 | [0.02; 53.12] | 1.5% | 1.5% |
| Singh 2016 3x doses | 0 | 17 | 0 | 17 | 1.00 | [0.02; 53.28] | 1.5% | 1.5% |
| Singh 2016 II | 0 | 20 | 0 | 20 | 1.00 | [0.02; 52.85] | 1.5% | 1.5% |
| Smith-Apeldoorn 2024 | 1 | 57 | 1 | 57 | 1.00 | [0.06; 16.39] | 3.0% | 3.0% |
| Sos 2013 | 0 | 11 | 0 | 11 | 1.00 | [0.02; 54.83] | 1.4% | 1.5% |
| Su 2017 | 0 | 47 | 0 | 47 | 1.00 | [0.02; 51.44] | 1.5% | 1.5% |
| Su 2023 | 0 | 42 | 0 | 42 | 1.00 | [0.02; 51.57] | 1.5% | 1.5% |
| Sumner 2019 | 0 | 17 | 0 | 17 | 1.00 | [0.02; 53.28] | 1.5% | 1.5% |
| Takahashi 2021 | 0 | 122 | 0 | 122 | 1.00 | [0.02; 50.80] | 1.5% | 1.5% |
| Taylor 2018 | 0 | 9 | 0 | 9 | 1.00 | [0.02; 55.80] | 1.4% | 1.5% |
| Tiger 2020 | 0 | 20 | 0 | 20 | 1.00 | [0.02; 52.85] | 1.5% | 1.5% |
| Wang 2024 | 0 | 182 | 0 | 182 | 1.00 | [0.02; 50.67] | 1.5% | 1.5% |
| Xiao 2023 | 0 | 15 | 0 | 15 | 1.00 | [0.02; 53.66] | 1.5% | 1.5% |
| Yang 2023 | 0 | 200 | 0 | 200 | 1.00 | [0.02; 50.64] | 1.5% | 1.5% |
| Zarate 2006 | 0 | 9 | 0 | 9 | 1.00 | [0.02; 55.80] | 1.4% | 1.5% |
| Zarate 2012 | 0 | 21 | 0 | 21 | 1.00 | [0.02; 52.73] | 1.5% | 1.5% |
| Zhou 2024 | 0 | 27 | 1 | 27 | 0.32 | [0.01; 8.24] | 4.4% | 2.2% |
| **Common effect model** | | **2792** | | **2792** | **1.00** | **[0.62; 1.62]** | **100.0%** | **.** |
| **Random effects model** | | | | | **1.00** | **[0.62; 1.62]** | **.** | **100.0%** |

Heterogeneity: $I^2 = 0.0\%$, $\tau^2 = 0$, $p = 1.0000$

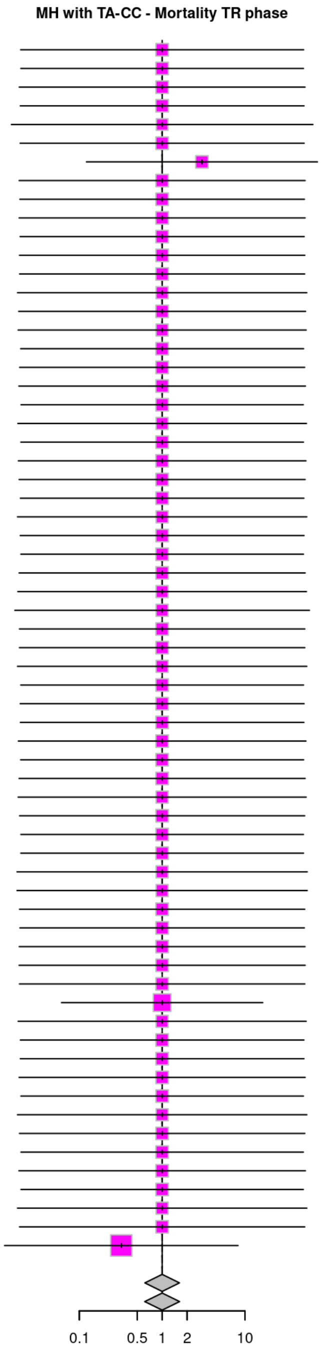

**Bayesian Meta-Analysis, smaller prior**

**Bayesian MA, Delta = 15 - Deaths FU**

| Study | Estimate | 95% CI |
|---|---|---|
| Abbar 2022 | 1.24 | [-1.98, 4.46] |
| Ahmed 2023 | 0.00 | [-3.97, 3.97] |
| Anderson 2017 | -0.08 | [-4.04, 3.88] |
| Burger 2016 | 0.76 | [-3.36, 4.88] |
| Canuso 2018 | -0.20 | [-4.16, 3.76] |
| Dakwar 2019 | 0.04 | [-3.92, 3.99] |
| Dakwar 2020 | 0.29 | [-3.67, 4.26] |
| Domany 2019 | -0.20 | [-4.16, 3.77] |
| Domany 2020 | 0.00 | [-4.02, 4.02] |
| Duek 2023 | 0.00 | [-3.99, 3.99] |
| Dwyer 2021 | -0.57 | [-4.61, 3.47] |
| Fava 2020 | -1.42 | [-5.37, 2.53] |
| Feder 2014 | -0.14 | [-4.11, 3.82] |
| Fineberg 2023 | 0.17 | [-3.83, 4.18] |
| Fu 2020 | 1.00 | [-2.21, 4.22] |
| Glue 2024b | -0.72 | [-4.73, 3.28] |
| Grabski 2022 | 0.00 | [-3.94, 3.94] |
| Ionescu 2020 | 0.05 | [-3.88, 3.98] |
| Jones 2024 | -0.42 | [-4.41, 3.57] |
| Lapidus 2014 | 0.00 | [-4.01, 4.01] |
| Leal 2023 | 0.00 | [-4.09, 4.09] |
| Lii 2023 | 1.15 | [-2.11, 4.41] |
| Lijffijt 2021 | -0.42 | [-4.40, 3.56] |
| Murrough 2013 | -0.62 | [-4.57, 3.33] |
| Murrough 2015 II | 1.18 | [-2.12, 4.48] |
| Ohtani 2024 | 0.00 | [-3.98, 3.98] |
| Pattanaseri 2024 | -0.19 | [-4.20, 3.82] |
| Phillips 2019 | -0.05 | [-4.01, 3.92] |
| Price 2022 | -0.70 | [-4.63, 3.24] |
| Ray-Griffith 2017 | 0.00 | [-4.03, 4.03] |
| Rodriguez 2013 | -0.13 | [-4.17, 3.92] |
| Schiweck 2021 | -1.05 | [-5.06, 2.95] |
| Smith-Apeldoorn 2024 | -0.02 | [-3.96, 3.92] |
| Sos 2013 | 0.53 | [-3.46, 4.52] |
| Su 2017 | -0.66 | [-4.61, 3.29] |
| Su 2023 | 0.00 | [-3.94, 3.94] |
| Sumner 2019 | -0.06 | [-4.04, 3.92] |
| Taylor 2018 | -0.11 | [-4.14, 3.92] |
| Wang 2024 | 0.00 | [-3.93, 3.93] |
| Xiao 2023 | 0.00 | [-3.98, 3.98] |
| Yang 2023 | -0.69 | [-4.62, 3.24] |
| Zarate 2006 | 0.00 | [-4.02, 4.02] |
| Zarate 2012 | 0.09 | [-3.87, 4.05] |
| Zhou 2024 | -0.08 | [-4.03, 3.88] |
| **Summary** | **0.79** | **[-0.54, 2.24]** |
| **Prediction** | **0.79** | **[-0.79, 2.49]** |

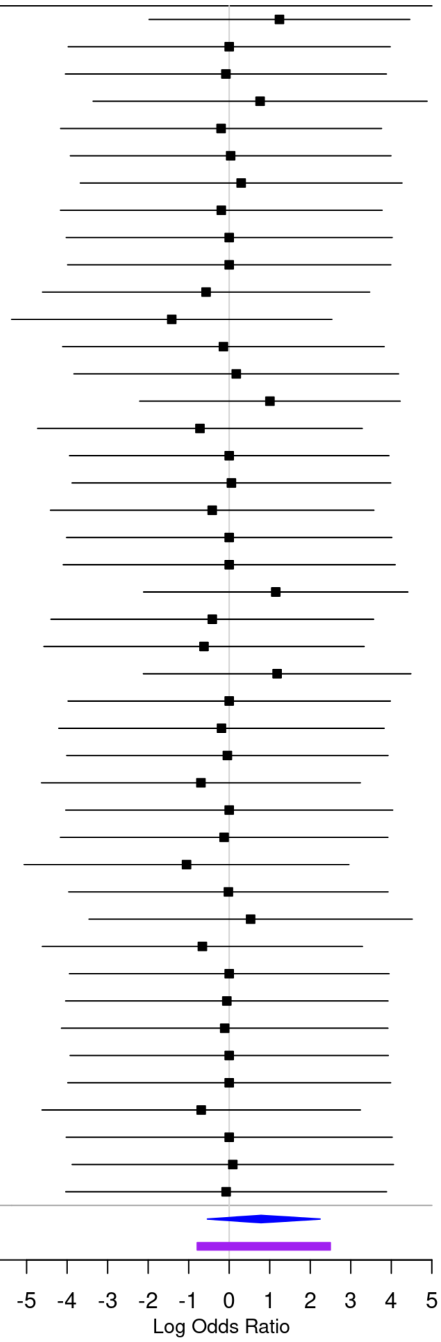

*Heterogeneity (tau): 0.30 [0.00, 0.88]*

**Exact Method** Not possible to run because no events in the control arms

#### Treatment + Follow-Up Phase Combined

#### Peto's Method

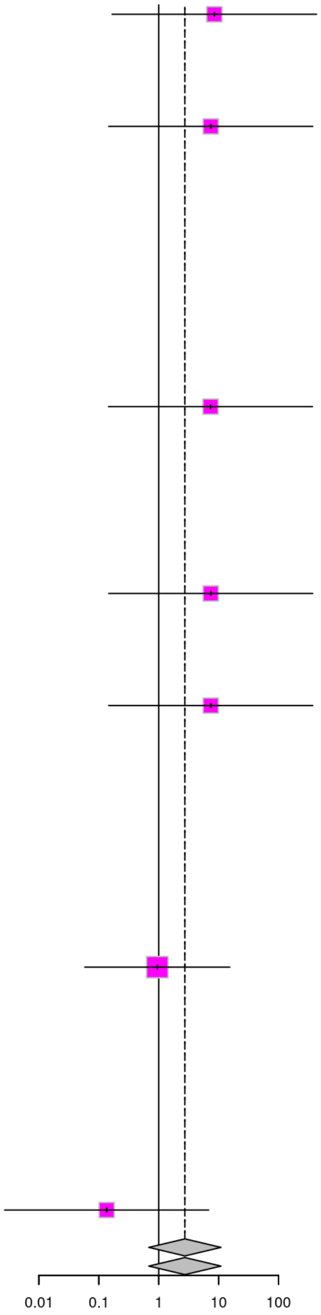

| Study | Experimental Events | Experimental Total | Control Events | Control Total | OR | 95%-CI | Weight (common) | Weight (random) |
|---|---|---|---|---|---|---|---|---|
| Abbar 2022 | 1 | 73 | 0 | 83 | 8.47 | [0.17; 430.52] | 12.5% | 12.5% |
| Abdallah 2022 | 0 | 108 | 0 | 55 | | | 0.0% | 0.0% |
| Ahmed 2023 | 0 | 18 | 0 | 18 | | | 0.0% | 0.0% |
| Anderson 2017 | 0 | 40 | 0 | 39 | | | 0.0% | 0.0% |
| Burger 2016 | 0 | 3 | 0 | 7 | | | 0.0% | 0.0% |
| Canuso 2018 | 0 | 35 | 0 | 31 | | | 0.0% | 0.0% |
| Chen 2023 | 1 | 126 | 0 | 126 | 7.39 | [0.15; 372.38] | 12.5% | 12.5% |
| Colla 2024 | 0 | 17 | 0 | 10 | | | 0.0% | 0.0% |
| Dakwar 2019 | 0 | 27 | 0 | 28 | | | 0.0% | 0.0% |
| Dakwar 2020 | 0 | 17 | 0 | 23 | | | 0.0% | 0.0% |
| Daly 2018 PA | 0 | 34 | 0 | 33 | | | 0.0% | 0.0% |
| Daly 2018 PB | 0 | 20 | 0 | 21 | | | 0.0% | 0.0% |
| Domany 2019 | 0 | 22 | 0 | 18 | | | 0.0% | 0.0% |
| Domany 2020 | 0 | 9 | 0 | 9 | | | 0.0% | 0.0% |
| Duek 2023 | 0 | 14 | 0 | 14 | | | 0.0% | 0.0% |
| Dwyer 2021 | 0 | 11 | 0 | 6 | | | 0.0% | 0.0% |
| Fava 2020 | 0 | 80 | 0 | 19 | | | 0.0% | 0.0% |
| Feder 2014 | 0 | 22 | 0 | 19 | | | 0.0% | 0.0% |
| Feder 2021 | 0 | 15 | 0 | 15 | | | 0.0% | 0.0% |
| Fedgchin 2019 | 0 | 231 | 0 | 113 | | | 0.0% | 0.0% |
| Fineberg 2023 | 0 | 10 | 0 | 12 | | | 0.0% | 0.0% |
| Fu 2020 | 1 | 113 | 0 | 112 | 7.32 | [0.15; 369.12] | 12.5% | 12.5% |
| George 2017 | 0 | 12 | 0 | 4 | | | 0.0% | 0.0% |
| Glue 2024b | 0 | 17 | 0 | 8 | | | 0.0% | 0.0% |
| Grabski 2022 | 0 | 48 | 0 | 48 | | | 0.0% | 0.0% |
| Grunebaum 2017 | 0 | 9 | 0 | 7 | | | 0.0% | 0.0% |
| Grunebaum 2018 | 0 | 40 | 0 | 40 | | | 0.0% | 0.0% |
| Ionescu 2020 | 0 | 114 | 0 | 113 | | | 0.0% | 0.0% |
| Jones 2024 | 0 | 17 | 0 | 11 | | | 0.0% | 0.0% |
| Lapidus 2014 | 0 | 10 | 0 | 10 | | | 0.0% | 0.0% |
| Leal 2023 | 0 | 5 | 0 | 5 | | | 0.0% | 0.0% |
| Lii 2023 | 1 | 20 | 0 | 20 | 7.39 | [0.15; 372.38] | 12.5% | 12.5% |
| Lijffijt 2021 | 0 | 20 | 0 | 13 | | | 0.0% | 0.0% |
| Loo 2016 | 0 | 9 | 0 | 6 | | | 0.0% | 0.0% |
| Loo 2023 | 0 | 89 | 0 | 92 | | | 0.0% | 0.0% |
| Milak 2020 | 0 | 38 | 0 | 5 | | | 0.0% | 0.0% |
| Murrough 2013 | 0 | 47 | 0 | 25 | | | 0.0% | 0.0% |
| Murrough 2015 II | 1 | 12 | 0 | 12 | 7.39 | [0.15; 372.38] | 12.5% | 12.5% |
| Ochs-Ross 2020 | 0 | 72 | 0 | 65 | | | 0.0% | 0.0% |
| Ohtani 2024 | 0 | 17 | 0 | 17 | | | 0.0% | 0.0% |
| Pattanaseri 2024 | 0 | 11 | 0 | 9 | | | 0.0% | 0.0% |
| Phillips 2019 | 0 | 22 | 0 | 21 | | | 0.0% | 0.0% |
| Popova 2019 | 0 | 115 | 0 | 109 | | | 0.0% | 0.0% |
| Price 2022 | 0 | 103 | 0 | 51 | | | 0.0% | 0.0% |
| Ray-Griffith 2017 | 0 | 8 | 0 | 8 | | | 0.0% | 0.0% |
| Rodriguez 2013 | 0 | 8 | 0 | 7 | | | 0.0% | 0.0% |
| Schiweck 2021 | 0 | 21 | 0 | 7 | | | 0.0% | 0.0% |
| Shiroma 2020 | 0 | 29 | 0 | 25 | | | 0.0% | 0.0% |
| Singh 2016 2x doses | 0 | 18 | 0 | 16 | | | 0.0% | 0.0% |
| Singh 2016 3x doses | 0 | 17 | 0 | 16 | | | 0.0% | 0.0% |
| Singh 2016 II | 0 | 20 | 0 | 10 | | | 0.0% | 0.0% |
| Smith-Apeldoorn 2024 | 1 | 57 | 1 | 54 | 0.95 | [0.06; 15.35] | 24.8% | 24.8% |
| Sos 2013 | 0 | 11 | 0 | 19 | | | 0.0% | 0.0% |
| Su 2017 | 0 | 47 | 0 | 24 | | | 0.0% | 0.0% |
| Su 2023 | 0 | 42 | 0 | 42 | | | 0.0% | 0.0% |
| Sumner 2019 | 0 | 17 | 0 | 16 | | | 0.0% | 0.0% |
| Takahashi 2021 | 0 | 122 | 0 | 80 | | | 0.0% | 0.0% |
| Taylor 2018 | 0 | 9 | 0 | 9 | | | 0.0% | 0.0% |
| Tiger 2020 | 0 | 20 | 0 | 10 | | | 0.0% | 0.0% |
| Wang 2024 | 0 | 182 | 0 | 182 | | | 0.0% | 0.0% |
| Xiao 2023 | 0 | 15 | 0 | 15 | | | 0.0% | 0.0% |
| Yang 2023 | 0 | 200 | 0 | 100 | | | 0.0% | 0.0% |
| Zarate 2006 | 0 | 9 | 0 | 9 | | | 0.0% | 0.0% |
| Zarate 2012 | 0 | 21 | 0 | 23 | | | 0.0% | 0.0% |
| Zhou 2024 | 0 | 27 | 1 | 27 | 0.14 | [0.00; 6.82] | 12.5% | 12.5% |
| **Common effect model** | | **2792** | | **2201** | **2.73** | **[0.68; 10.94]** | **100.0%** | **.** |
| **Random effects model** | | | | | **2.73** | **[0.68; 10.94]** | **.** | **100.0%** |

Heterogeneity: $I^2 = 0.0\%$, $\tau^2 = 0$, $p = 0.6607$

#### Maentel Mantel-Haenszel with CC

MH with CC - Mortality TR+FU phase

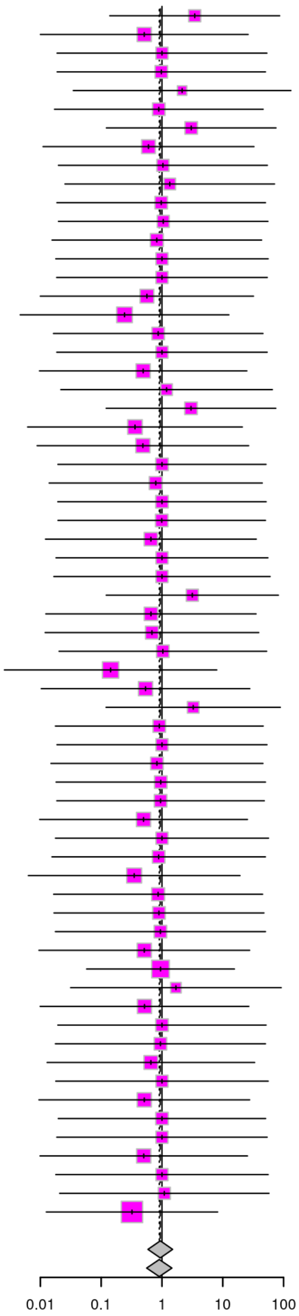

| Study | Experimental Events | Experimental Total | Control Events | Control Total | OR | 95%-CI | Weight (common) | Weight (random) |
|---|---|---|---|---|---|---|---|---|
| Abbar 2022 | 1 | 73 | 0 | 83 | 3.46 | [0.14; 86.13] | 1.3% | 2.2% |
| Abdallah 2022 | 0 | 108 | 0 | 55 | 0.51 | [0.01; 26.12] | 1.8% | 1.5% |
| Ahmed 2023 | 0 | 18 | 0 | 18 | 1.00 | [0.02; 53.12] | 1.3% | 1.5% |
| Anderson 2017 | 0 | 40 | 0 | 39 | 0.98 | [0.02; 50.37] | 1.4% | 1.5% |
| Burger 2016 | 0 | 3 | 0 | 7 | 2.14 | [0.03; 131.93] | 0.8% | 1.4% |
| Canuso 2018 | 0 | 35 | 0 | 31 | 0.89 | [0.02; 46.05] | 1.4% | 1.5% |
| Chen 2023 | 1 | 126 | 0 | 126 | 3.02 | [0.12; 74.94] | 1.4% | 2.2% |
| Colla 2024 | 0 | 17 | 0 | 10 | 0.60 | [0.01; 32.56] | 1.7% | 1.4% |
| Dakwar 2019 | 0 | 27 | 0 | 28 | 1.04 | [0.02; 54.08] | 1.3% | 1.5% |
| Dakwar 2020 | 0 | 17 | 0 | 23 | 1.34 | [0.03; 71.04] | 1.2% | 1.5% |
| Daly 2018 PA | 0 | 34 | 0 | 33 | 0.97 | [0.02; 50.36] | 1.4% | 1.5% |
| Daly 2018 PB | 0 | 20 | 0 | 21 | 1.05 | [0.02; 55.37] | 1.3% | 1.5% |
| Domany 2019 | 0 | 22 | 0 | 18 | 0.82 | [0.02; 43.47] | 1.5% | 1.5% |
| Domany 2020 | 0 | 9 | 0 | 9 | 1.00 | [0.02; 55.80] | 1.3% | 1.4% |
| Duek 2023 | 0 | 14 | 0 | 14 | 1.00 | [0.02; 53.89] | 1.3% | 1.4% |
| Dwyer 2021 | 0 | 11 | 0 | 6 | 0.57 | [0.01; 32.00] | 1.7% | 1.4% |
| Fava 2020 | 0 | 80 | 0 | 19 | 0.24 | [0.00; 12.59] | 2.2% | 1.5% |
| Feder 2014 | 0 | 22 | 0 | 19 | 0.87 | [0.02; 45.76] | 1.4% | 1.5% |
| Feder 2021 | 0 | 15 | 0 | 15 | 1.00 | [0.02; 53.66] | 1.3% | 1.4% |
| Fedgchin 2019 | 0 | 231 | 0 | 113 | 0.49 | [0.01; 24.87] | 1.8% | 1.5% |
| Fineberg 2023 | 0 | 10 | 0 | 12 | 1.19 | [0.02; 65.32] | 1.2% | 1.4% |
| Fu 2020 | 1 | 113 | 0 | 112 | 3.00 | [0.12; 74.43] | 1.4% | 2.2% |
| George 2017 | 0 | 12 | 0 | 4 | 0.36 | [0.01; 20.98] | 1.9% | 1.4% |
| Glue 2024b | 0 | 17 | 0 | 8 | 0.49 | [0.01; 26.64] | 1.8% | 1.4% |
| Grabski 2022 | 0 | 48 | 0 | 48 | 1.00 | [0.02; 51.42] | 1.4% | 1.5% |
| Grunebaum 2017 | 0 | 9 | 0 | 7 | 0.79 | [0.01; 44.65] | 1.5% | 1.4% |
| Grunebaum 2018 | 0 | 40 | 0 | 40 | 1.00 | [0.02; 51.63] | 1.4% | 1.5% |
| Ionescu 2020 | 0 | 114 | 0 | 113 | 0.99 | [0.02; 50.39] | 1.4% | 1.5% |
| Jones 2024 | 0 | 17 | 0 | 11 | 0.66 | [0.01; 35.52] | 1.6% | 1.4% |
| Lapidus 2014 | 0 | 10 | 0 | 10 | 1.00 | [0.02; 55.27] | 1.3% | 1.4% |
| Leal 2023 | 0 | 5 | 0 | 5 | 1.00 | [0.02; 59.99] | 1.3% | 1.4% |
| Lii 2023 | 1 | 20 | 0 | 20 | 3.15 | [0.12; 82.16] | 1.3% | 2.2% |
| Lijffijt 2021 | 0 | 20 | 0 | 13 | 0.66 | [0.01; 35.23] | 1.6% | 1.4% |
| Loo 2016 | 0 | 9 | 0 | 6 | 0.68 | [0.01; 39.07] | 1.5% | 1.4% |
| Loo 2023 | 0 | 89 | 0 | 92 | 1.03 | [0.02; 52.65] | 1.4% | 1.5% |
| Milak 2020 | 0 | 38 | 0 | 5 | 0.14 | [0.00; 7.96] | 2.4% | 1.4% |
| Murrough 2013 | 0 | 47 | 0 | 25 | 0.54 | [0.01; 27.86] | 1.8% | 1.5% |
| Murrough 2015 II | 1 | 12 | 0 | 12 | 3.26 | [0.12; 88.35] | 1.2% | 2.1% |
| Ochs-Ross 2020 | 0 | 72 | 0 | 65 | 0.90 | [0.02; 46.18] | 1.4% | 1.5% |
| Ohtani 2024 | 0 | 17 | 0 | 17 | 1.00 | [0.02; 53.28] | 1.3% | 1.5% |
| Pattanaseri 2024 | 0 | 11 | 0 | 9 | 0.83 | [0.01; 45.69] | 1.4% | 1.4% |
| Phillips 2019 | 0 | 22 | 0 | 21 | 0.96 | [0.02; 50.34] | 1.4% | 1.5% |
| Popova 2019 | 0 | 115 | 0 | 109 | 0.95 | [0.02; 48.20] | 1.4% | 1.5% |
| Price 2022 | 0 | 103 | 0 | 51 | 0.50 | [0.01; 25.44] | 1.8% | 1.5% |
| Ray-Griffith 2017 | 0 | 8 | 0 | 8 | 1.00 | [0.02; 56.46] | 1.3% | 1.4% |
| Rodriguez 2013 | 0 | 8 | 0 | 7 | 0.88 | [0.02; 50.19] | 1.4% | 1.4% |
| Schiweck 2021 | 0 | 21 | 0 | 7 | 0.35 | [0.01; 19.18] | 2.0% | 1.4% |
| Shiroma 2020 | 0 | 29 | 0 | 25 | 0.86 | [0.02; 45.14] | 1.5% | 1.5% |
| Singh 2016 2x doses | 0 | 18 | 0 | 16 | 0.89 | [0.02; 47.53] | 1.4% | 1.5% |
| Singh 2016 3x doses | 0 | 17 | 0 | 16 | 0.94 | [0.02; 50.32] | 1.4% | 1.4% |
| Singh 2016 II | 0 | 20 | 0 | 10 | 0.51 | [0.01; 27.68] | 1.8% | 1.4% |
| Smith-Apeldoorn 2024 | 1 | 57 | 1 | 54 | 0.95 | [0.06; 15.52] | 2.8% | 2.9% |
| Sos 2013 | 0 | 11 | 0 | 19 | 1.70 | [0.03; 91.39] | 1.0% | 1.4% |
| Su 2017 | 0 | 47 | 0 | 24 | 0.52 | [0.01; 26.79] | 1.8% | 1.5% |
| Su 2023 | 0 | 42 | 0 | 42 | 1.00 | [0.02; 51.57] | 1.4% | 1.5% |
| Sumner 2019 | 0 | 17 | 0 | 16 | 0.94 | [0.02; 50.32] | 1.4% | 1.4% |
| Takahashi 2021 | 0 | 122 | 0 | 80 | 0.66 | [0.01; 33.45] | 1.7% | 1.5% |
| Taylor 2018 | 0 | 9 | 0 | 9 | 1.00 | [0.02; 55.80] | 1.3% | 1.4% |
| Tiger 2020 | 0 | 20 | 0 | 10 | 0.51 | [0.01; 27.68] | 1.8% | 1.4% |
| Wang 2024 | 0 | 182 | 0 | 182 | 1.00 | [0.02; 50.67] | 1.4% | 1.5% |
| Xiao 2023 | 0 | 15 | 0 | 15 | 1.00 | [0.02; 53.66] | 1.3% | 1.4% |
| Yang 2023 | 0 | 200 | 0 | 100 | 0.50 | [0.01; 25.45] | 1.8% | 1.5% |
| Zarate 2006 | 0 | 9 | 0 | 9 | 1.00 | [0.02; 55.80] | 1.3% | 1.4% |
| Zarate 2012 | 0 | 21 | 0 | 23 | 1.09 | [0.02; 57.53] | 1.3% | 1.5% |
| Zhou 2024 | 0 | 27 | 1 | 27 | 0.32 | [0.01; 8.24] | 4.1% | 2.2% |
| **Common effect model** | | **2792** | | **2201** | **0.94** | **[0.59; 1.50]** | **100.0%** | **.** |
| **Random effects model** | | | | | **0.90** | **[0.56; 1.46]** | **.** | **100.0%** |

Heterogeneity: $I^2 = 0.0\%$, $\tau^2 = 0$, $p = 1.0000$

#### Maentel Mantel-Haenszel with Treatment-Arm CC

| Study | Experimental Events | Experimental Total | Control Events | Control Total | OR | 95%-CI | Weight (common) | Weight (random) |
|---|---|---|---|---|---|---|---|---|
| Abbar 2022 | 1 | 73 | 0 | 83 | 3.18 | [0.14; 74.37] | 1.5% | 2.4% |
| Abdallah 2022 | 0 | 108 | 0 | 55 | 1.00 | [0.02; 63.93] | 1.4% | 1.4% |
| Ahmed 2023 | 0 | 18 | 0 | 18 | 1.00 | [0.02; 53.12] | 1.5% | 1.5% |
| Anderson 2017 | 0 | 40 | 0 | 39 | 1.00 | [0.02; 51.66] | 1.6% | 1.5% |
| Burger 2016 | 0 | 3 | 0 | 7 | 1.00 | [0.01; 87.11] | 1.2% | 1.2% |
| Canuso 2018 | 0 | 35 | 0 | 31 | 1.00 | [0.02; 52.27] | 1.6% | 1.5% |
| Chen 2023 | 1 | 126 | 0 | 126 | 3.02 | [0.12; 74.94] | 1.6% | 2.3% |
| Colla 2024 | 0 | 17 | 0 | 10 | 1.00 | [0.02; 62.21] | 1.4% | 1.4% |
| Dakwar 2019 | 0 | 27 | 0 | 28 | 1.00 | [0.02; 52.22] | 1.6% | 1.5% |
| Dakwar 2020 | 0 | 17 | 0 | 23 | 1.00 | [0.02; 55.30] | 1.5% | 1.5% |
| Daly 2018 PA | 0 | 34 | 0 | 33 | 1.00 | [0.02; 51.89] | 1.6% | 1.5% |
| Daly 2018 PB | 0 | 20 | 0 | 21 | 1.00 | [0.02; 52.85] | 1.5% | 1.5% |
| Domany 2019 | 0 | 22 | 0 | 18 | 1.00 | [0.02; 53.92] | 1.5% | 1.5% |
| Domany 2020 | 0 | 9 | 0 | 9 | 1.00 | [0.02; 55.80] | 1.5% | 1.5% |
| Duek 2023 | 0 | 14 | 0 | 14 | 1.00 | [0.02; 53.89] | 1.5% | 1.5% |
| Dwyer 2021 | 0 | 11 | 0 | 6 | 1.00 | [0.01; 67.61] | 1.4% | 1.3% |
| Fava 2020 | 0 | 80 | 0 | 19 | 1.00 | [0.01; 148.67] | 1.0% | 1.0% |
| Feder 2014 | 0 | 22 | 0 | 19 | 1.00 | [0.02; 53.36] | 1.5% | 1.5% |
| Feder 2021 | 0 | 15 | 0 | 15 | 1.00 | [0.02; 53.66] | 1.5% | 1.5% |
| Fedgchin 2019 | 0 | 231 | 0 | 113 | 1.00 | [0.02; 65.31] | 1.4% | 1.4% |
| Fineberg 2023 | 0 | 10 | 0 | 12 | 1.00 | [0.02; 55.75] | 1.5% | 1.5% |
| Fu 2020 | 1 | 113 | 0 | 112 | 3.02 | [0.12; 75.23] | 1.6% | 2.3% |
| George 2017 | 0 | 12 | 0 | 4 | 1.00 | [0.01; 105.38] | 1.1% | 1.1% |
| Glue 2024b | 0 | 17 | 0 | 8 | 1.00 | [0.01; 72.36] | 1.3% | 1.3% |
| Grabski 2022 | 0 | 48 | 0 | 48 | 1.00 | [0.02; 51.42] | 1.6% | 1.5% |
| Grunebaum 2017 | 0 | 9 | 0 | 7 | 1.00 | [0.02; 58.29] | 1.5% | 1.4% |
| Grunebaum 2018 | 0 | 40 | 0 | 40 | 1.00 | [0.02; 51.63] | 1.6% | 1.5% |
| Ionescu 2020 | 0 | 114 | 0 | 113 | 1.00 | [0.02; 50.83] | 1.6% | 1.6% |
| Jones 2024 | 0 | 17 | 0 | 11 | 1.00 | [0.02; 59.25] | 1.5% | 1.4% |
| Lapidus 2014 | 0 | 10 | 0 | 10 | 1.00 | [0.02; 55.27] | 1.5% | 1.5% |
| Leal 2023 | 0 | 5 | 0 | 5 | 1.00 | [0.02; 59.99] | 1.5% | 1.4% |
| Lii 2023 | 1 | 20 | 0 | 20 | 3.15 | [0.12; 82.16] | 1.5% | 2.3% |
| Lijffijt 2021 | 0 | 20 | 0 | 13 | 1.00 | [0.02; 58.54] | 1.5% | 1.4% |
| Loo 2016 | 0 | 9 | 0 | 6 | 1.00 | [0.02; 61.80] | 1.4% | 1.4% |
| Loo 2023 | 0 | 89 | 0 | 92 | 1.00 | [0.02; 50.97] | 1.6% | 1.6% |
| Milak 2020 | 0 | 38 | 0 | 5 | 1.00 | [0.00; 484.58] | 0.6% | 0.6% |
| Murrough 2013 | 0 | 47 | 0 | 25 | 1.00 | [0.02; 63.11] | 1.4% | 1.4% |
| Murrough 2015 II | 1 | 12 | 0 | 12 | 3.26 | [0.12; 88.35] | 1.4% | 2.2% |
| Ochs-Ross 2020 | 0 | 72 | 0 | 65 | 1.00 | [0.02; 51.38] | 1.6% | 1.5% |
| Ohtani 2024 | 0 | 17 | 0 | 17 | 1.00 | [0.02; 53.28] | 1.5% | 1.5% |
| Pattanaseri 2024 | 0 | 11 | 0 | 9 | 1.00 | [0.02; 56.40] | 1.5% | 1.5% |
| Phillips 2019 | 0 | 22 | 0 | 21 | 1.00 | [0.02; 52.74] | 1.5% | 1.5% |
| Popova 2019 | 0 | 115 | 0 | 109 | 1.00 | [0.02; 50.91] | 1.6% | 1.6% |
| Price 2022 | 0 | 103 | 0 | 51 | 1.00 | [0.02; 65.23] | 1.4% | 1.4% |
| Ray-Griffith 2017 | 0 | 8 | 0 | 8 | 1.00 | [0.02; 56.46] | 1.5% | 1.5% |
| Rodriguez 2013 | 0 | 8 | 0 | 7 | 1.00 | [0.02; 57.37] | 1.5% | 1.5% |
| Schiweck 2021 | 0 | 21 | 0 | 7 | 1.00 | [0.01; 99.86] | 1.1% | 1.1% |
| Shiroma 2020 | 0 | 29 | 0 | 25 | 1.00 | [0.02; 52.79] | 1.5% | 1.5% |
| Singh 2016 2x doses | 0 | 18 | 0 | 16 | 1.00 | [0.02; 53.65] | 1.5% | 1.5% |
| Singh 2016 3x doses | 0 | 17 | 0 | 16 | 1.00 | [0.02; 53.46] | 1.5% | 1.5% |
| Singh 2016 II | 0 | 20 | 0 | 10 | 1.00 | [0.01; 68.32] | 1.4% | 1.3% |
| Smith-Apeldoorn 2024 | 1 | 57 | 1 | 54 | 0.95 | [0.06; 15.52] | 3.2% | 3.1% |
| Sos 2013 | 0 | 11 | 0 | 19 | 1.00 | [0.02; 62.32] | 1.4% | 1.4% |
| Su 2017 | 0 | 47 | 0 | 24 | 1.00 | [0.02; 64.85] | 1.4% | 1.4% |
| Su 2023 | 0 | 42 | 0 | 42 | 1.00 | [0.02; 51.57] | 1.6% | 1.5% |
| Sumner 2019 | 0 | 17 | 0 | 16 | 1.00 | [0.02; 53.46] | 1.5% | 1.5% |
| Takahashi 2021 | 0 | 122 | 0 | 80 | 1.00 | [0.02; 55.55] | 1.5% | 1.5% |
| Taylor 2018 | 0 | 9 | 0 | 9 | 1.00 | [0.02; 55.80] | 1.5% | 1.5% |
| Tiger 2020 | 0 | 20 | 0 | 10 | 1.00 | [0.01; 68.32] | 1.4% | 1.3% |
| Wang 2024 | 0 | 182 | 0 | 182 | 1.00 | [0.02; 50.67] | 1.6% | 1.6% |
| Xiao 2023 | 0 | 15 | 0 | 15 | 1.00 | [0.02; 53.66] | 1.5% | 1.5% |
| Yang 2023 | 0 | 200 | 0 | 100 | 1.00 | [0.02; 64.37] | 1.4% | 1.4% |
| Zarate 2006 | 0 | 9 | 0 | 9 | 1.00 | [0.02; 55.80] | 1.5% | 1.5% |
| Zarate 2012 | 0 | 21 | 0 | 23 | 1.00 | [0.02; 52.84] | 1.5% | 1.5% |
| Zhou 2024 | 0 | 27 | 1 | 27 | 0.32 | [0.01; 8.24] | 4.7% | 2.3% |
| **Common effect model** | | **2792** | | **2201** | **1.13** | **[0.70; 1.82]** | **100.0%** | **.** |
| **Random effects model** | | | | | **1.11** | **[0.68; 1.81]** | **.** | **100.0%** |

Heterogeneity: $I^2 = 0.0\%$, $\tau^2 = 0$, $p = 1.0000$

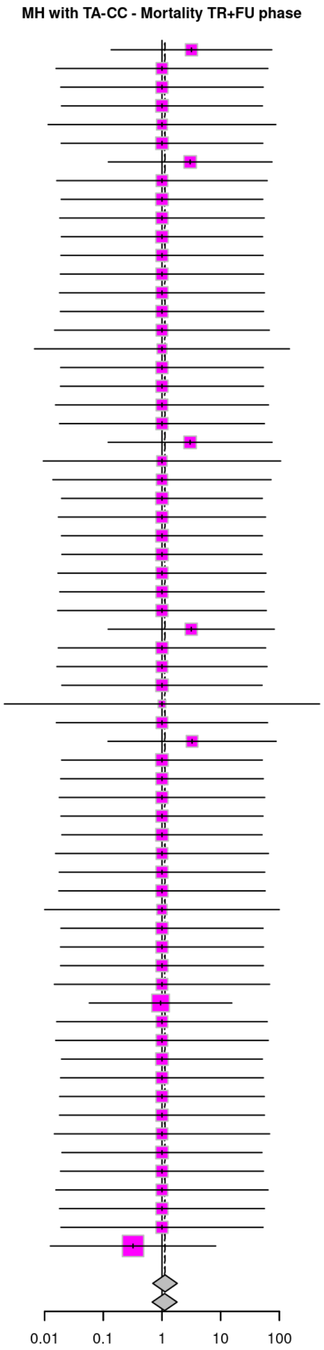

#### Maentel Mantel-Haenszel Risk-Difference

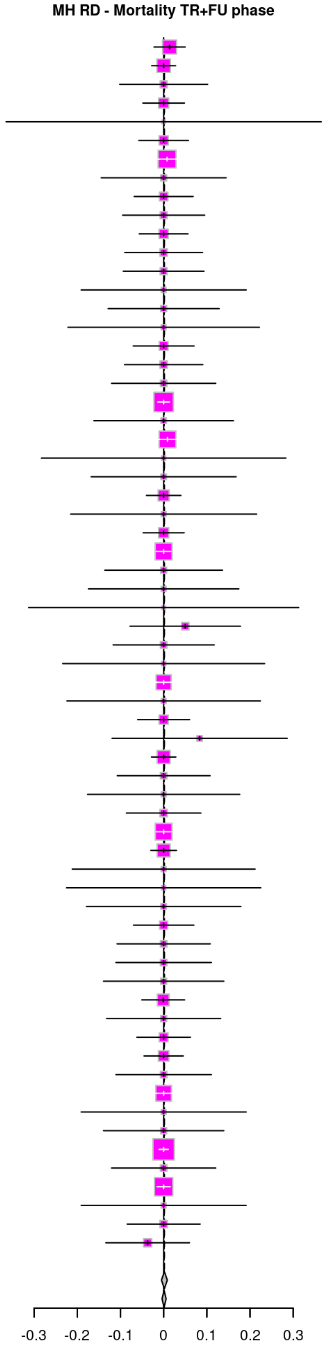

| Study | Experimental Events | Experimental Total | Control Events | Control Total | RD | 95%-CI | Weight (common) | Weight (random) |
|---|---|---|---|---|---|---|---|---|
| Abbar 2022 | 1 | 73 | 0 | 83 | 0.01 | [-0.02; 0.05] | 3.2% | 1.7% |
| Abdallah 2022 | 0 | 108 | 0 | 55 | 0.00 | [-0.03; 0.03] | 3.0% | 2.9% |
| Ahmed 2023 | 0 | 18 | 0 | 18 | 0.00 | [-0.10; 0.10] | 0.8% | 0.2% |
| Anderson 2017 | 0 | 40 | 0 | 39 | 0.00 | [-0.05; 0.05] | 1.7% | 1.0% |
| Burger 2016 | 0 | 3 | 0 | 7 | 0.00 | [-0.36; 0.36] | 0.2% | 0.0% |
| Canuso 2018 | 0 | 35 | 0 | 31 | 0.00 | [-0.06; 0.06] | 1.4% | 0.7% |
| Chen 2023 | 1 | 126 | 0 | 126 | 0.01 | [-0.01; 0.03] | 5.2% | 4.8% |
| Colla 2024 | 0 | 17 | 0 | 10 | 0.00 | [-0.14; 0.14] | 0.6% | 0.1% |
| Dakwar 2019 | 0 | 27 | 0 | 28 | 0.00 | [-0.07; 0.07] | 1.2% | 0.5% |
| Dakwar 2020 | 0 | 17 | 0 | 23 | 0.00 | [-0.10; 0.10] | 0.8% | 0.2% |
| Daly 2018 PA | 0 | 34 | 0 | 33 | 0.00 | [-0.06; 0.06] | 1.4% | 0.7% |
| Daly 2018 PB | 0 | 20 | 0 | 21 | 0.00 | [-0.09; 0.09] | 0.9% | 0.3% |
| Domany 2019 | 0 | 22 | 0 | 18 | 0.00 | [-0.09; 0.09] | 0.8% | 0.3% |
| Domany 2020 | 0 | 9 | 0 | 9 | 0.00 | [-0.19; 0.19] | 0.4% | 0.1% |
| Duek 2023 | 0 | 14 | 0 | 14 | 0.00 | [-0.13; 0.13] | 0.6% | 0.1% |
| Dwyer 2021 | 0 | 11 | 0 | 6 | 0.00 | [-0.22; 0.22] | 0.4% | 0.0% |
| Fava 2020 | 0 | 80 | 0 | 19 | 0.00 | [-0.07; 0.07] | 1.3% | 0.5% |
| Feder 2014 | 0 | 22 | 0 | 19 | 0.00 | [-0.09; 0.09] | 0.9% | 0.3% |
| Feder 2021 | 0 | 15 | 0 | 15 | 0.00 | [-0.12; 0.12] | 0.7% | 0.2% |
| Fedgchin 2019 | 0 | 231 | 0 | 113 | 0.00 | [-0.01; 0.01] | 6.2% | 12.3% |
| Fineberg 2023 | 0 | 10 | 0 | 12 | 0.00 | [-0.16; 0.16] | 0.5% | 0.1% |
| Fu 2020 | 1 | 113 | 0 | 112 | 0.01 | [-0.02; 0.03] | 4.6% | 3.8% |
| George 2017 | 0 | 12 | 0 | 4 | 0.00 | [-0.28; 0.28] | 0.3% | 0.0% |
| Glue 2024b | 0 | 17 | 0 | 8 | 0.00 | [-0.17; 0.17] | 0.5% | 0.1% |
| Grabski 2022 | 0 | 48 | 0 | 48 | 0.00 | [-0.04; 0.04] | 2.0% | 1.4% |
| Grunebaum 2017 | 0 | 9 | 0 | 7 | 0.00 | [-0.22; 0.22] | 0.4% | 0.0% |
| Grunebaum 2018 | 0 | 40 | 0 | 40 | 0.00 | [-0.05; 0.05] | 1.7% | 1.0% |
| Ionescu 2020 | 0 | 114 | 0 | 113 | 0.00 | [-0.02; 0.02] | 4.7% | 7.7% |
| Jones 2024 | 0 | 17 | 0 | 11 | 0.00 | [-0.14; 0.14] | 0.6% | 0.1% |
| Lapidus 2014 | 0 | 10 | 0 | 10 | 0.00 | [-0.17; 0.17] | 0.4% | 0.1% |
| Leal 2023 | 0 | 5 | 0 | 5 | 0.00 | [-0.31; 0.31] | 0.2% | 0.0% |
| Lii 2023 | 1 | 20 | 0 | 20 | 0.05 | [-0.08; 0.18] | 0.9% | 0.1% |
| Lijffijt 2021 | 0 | 20 | 0 | 13 | 0.00 | [-0.12; 0.12] | 0.7% | 0.2% |
| Loo 2016 | 0 | 9 | 0 | 6 | 0.00 | [-0.23; 0.23] | 0.3% | 0.0% |
| Loo 2023 | 0 | 89 | 0 | 92 | 0.00 | [-0.02; 0.02] | 3.7% | 4.9% |
| Milak 2020 | 0 | 38 | 0 | 5 | 0.00 | [-0.22; 0.22] | 0.4% | 0.0% |
| Murrough 2013 | 0 | 47 | 0 | 25 | 0.00 | [-0.06; 0.06] | 1.4% | 0.6% |
| Murrough 2015 II | 1 | 12 | 0 | 12 | 0.08 | [-0.12; 0.29] | 0.5% | 0.1% |
| Ochs-Ross 2020 | 0 | 72 | 0 | 65 | 0.00 | [-0.03; 0.03] | 2.8% | 2.8% |
| Ohtani 2024 | 0 | 17 | 0 | 17 | 0.00 | [-0.11; 0.11] | 0.7% | 0.2% |
| Pattanaseri 2024 | 0 | 11 | 0 | 9 | 0.00 | [-0.18; 0.18] | 0.4% | 0.1% |
| Phillips 2019 | 0 | 22 | 0 | 21 | 0.00 | [-0.09; 0.09] | 0.9% | 0.3% |
| Popova 2019 | 0 | 115 | 0 | 109 | 0.00 | [-0.02; 0.02] | 4.6% | 7.5% |
| Price 2022 | 0 | 103 | 0 | 51 | 0.00 | [-0.03; 0.03] | 2.8% | 2.5% |
| Ray-Griffith 2017 | 0 | 8 | 0 | 8 | 0.00 | [-0.21; 0.21] | 0.4% | 0.1% |
| Rodriguez 2013 | 0 | 8 | 0 | 7 | 0.00 | [-0.22; 0.22] | 0.3% | 0.0% |
| Schiweck 2021 | 0 | 21 | 0 | 7 | 0.00 | [-0.18; 0.18] | 0.5% | 0.1% |
| Shiroma 2020 | 0 | 29 | 0 | 25 | 0.00 | [-0.07; 0.07] | 1.1% | 0.5% |
| Singh 2016 2x doses | 0 | 18 | 0 | 16 | 0.00 | [-0.11; 0.11] | 0.7% | 0.2% |
| Singh 2016 3x doses | 0 | 17 | 0 | 16 | 0.00 | [-0.11; 0.11] | 0.7% | 0.2% |
| Singh 2016 II | 0 | 20 | 0 | 10 | 0.00 | [-0.14; 0.14] | 0.6% | 0.1% |
| Smith-Apeldoorn 2024 | 1 | 57 | 1 | 54 | -0.00 | [-0.05; 0.05] | 2.3% | 0.9% |
| Sos 2013 | 0 | 11 | 0 | 19 | 0.00 | [-0.13; 0.13] | 0.6% | 0.1% |
| Su 2017 | 0 | 47 | 0 | 24 | 0.00 | [-0.06; 0.06] | 1.3% | 0.6% |
| Su 2023 | 0 | 42 | 0 | 42 | 0.00 | [-0.05; 0.05] | 1.8% | 1.1% |
| Sumner 2019 | 0 | 17 | 0 | 16 | 0.00 | [-0.11; 0.11] | 0.7% | 0.2% |
| Takahashi 2021 | 0 | 122 | 0 | 80 | 0.00 | [-0.02; 0.02] | 4.0% | 5.4% |
| Taylor 2018 | 0 | 9 | 0 | 9 | 0.00 | [-0.19; 0.19] | 0.4% | 0.1% |
| Tiger 2020 | 0 | 20 | 0 | 10 | 0.00 | [-0.14; 0.14] | 0.6% | 0.1% |
| Wang 2024 | 0 | 182 | 0 | 182 | 0.00 | [-0.01; 0.01] | 7.5% | 19.6% |
| Xiao 2023 | 0 | 15 | 0 | 15 | 0.00 | [-0.12; 0.12] | 0.7% | 0.2% |
| Yang 2023 | 0 | 200 | 0 | 100 | 0.00 | [-0.02; 0.02] | 5.5% | 9.6% |
| Zarate 2006 | 0 | 9 | 0 | 9 | 0.00 | [-0.19; 0.19] | 0.4% | 0.1% |
| Zarate 2012 | 0 | 21 | 0 | 23 | 0.00 | [-0.08; 0.08] | 0.9% | 0.3% |
| Zhou 2024 | 0 | 27 | 1 | 27 | -0.04 | [-0.13; 0.06] | 1.1% | 0.2% |
| **Common effect model** | | **2792** | | **2201** | **0.00** | **[-0.01; 0.01]** | **100.0%** | **.** |
| **Random effects model** | | | | | **0.00** | **[-0.00; 0.01]** | **.** | **100.0%** |

Heterogeneity: $I^2 = 0.0\%$, $\tau^2 = 0$, $p = 1.0000$

#### Arcsine Risk-Difference

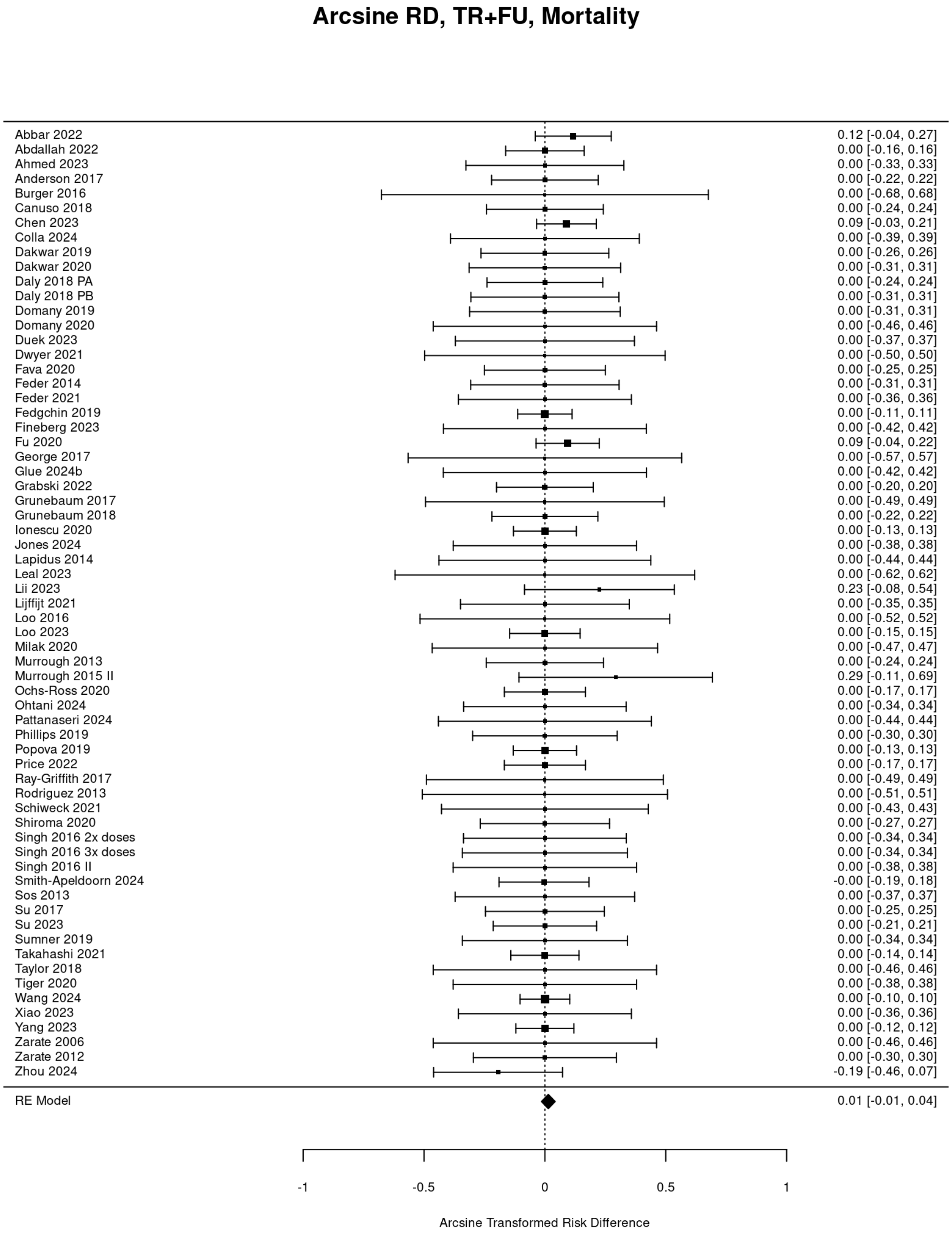

#### Bayesian Meta-Analysis, wider prior

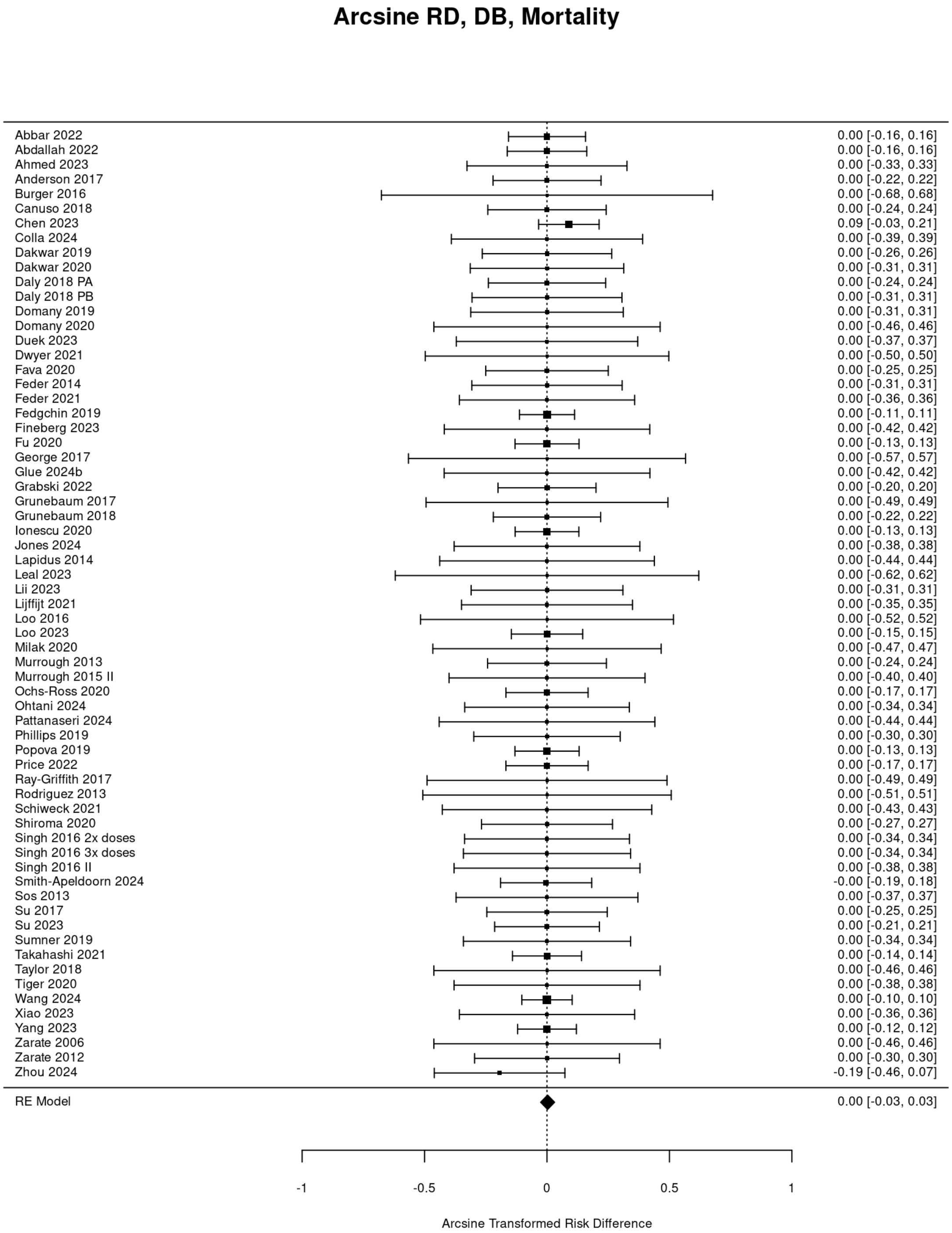

#### Bayesian Meta-Analysis, smaller prior

##### Bayesian MA, Delta = 15 - Deaths TR+FU

| Study | Estimate | 95% CI |
|---|---|---|
| Abbar 2022 | 1.24 | [-1.98, 4.46] |
| Abdallah 2022 | -0.67 | [-4.60, 3.26] |
| Ahmed 2023 | 0.00 | [-3.97, 3.97] |
| Anderson 2017 | -0.03 | [-3.97, 3.92] |
| Burger 2016 | 0.76 | [-3.36, 4.88] |
| Canuso 2018 | -0.12 | [-4.07, 3.83] |
| Chen 2023 | 1.11 | [-2.10, 4.32] |
| Colla 2024 | -0.51 | [-4.50, 3.48] |
| Dakwar 2019 | 0.04 | [-3.92, 3.99] |
| Dakwar 2020 | 0.29 | [-3.67, 4.26] |
| Daly 2018 PA | -0.03 | [-3.98, 3.92] |
| Daly 2018 PB | 0.05 | [-3.92, 4.01] |
| Domany 2019 | -0.20 | [-4.16, 3.77] |
| Domany 2020 | 0.00 | [-4.02, 4.02] |
| Duek 2023 | 0.00 | [-3.99, 3.99] |
| Dwyer 2021 | -0.57 | [-4.61, 3.47] |
| Fava 2020 | -1.42 | [-5.37, 2.53] |
| Feder 2014 | -0.14 | [-4.11, 3.82] |
| Feder 2021 | 0.00 | [-3.98, 3.98] |
| Fedgchin 2019 | -0.71 | [-4.64, 3.21] |
| Fineberg 2023 | 0.17 | [-3.83, 4.18] |
| Fu 2020 | 1.10 | [-2.11, 4.31] |
| George 2017 | -1.02 | [-5.09, 3.04] |
| Glue 2024b | -0.72 | [-4.73, 3.28] |
| Grabski 2022 | 0.00 | [-3.94, 3.94] |
| Grunebaum 2017 | -0.24 | [-4.27, 3.80] |
| Grunebaum 2018 | 0.00 | [-3.94, 3.94] |
| Ionescu 2020 | -0.01 | [-3.94, 3.92] |
| Jones 2024 | -0.42 | [-4.41, 3.57] |
| Lapidus 2014 | 0.00 | [-4.01, 4.01] |
| Leal 2023 | 0.00 | [-4.09, 4.09] |
| Lii 2023 | 1.15 | [-2.11, 4.41] |
| Lijffijt 2021 | -0.42 | [-4.40, 3.56] |
| Loo 2016 | -0.38 | [-4.42, 3.67] |
| Loo 2023 | 0.03 | [-3.90, 3.96] |
| Milak 2020 | -1.95 | [-5.97, 2.07] |
| Murrough 2013 | -0.62 | [-4.57, 3.33] |
| Murrough 2015 II | 1.18 | [-2.12, 4.48] |
| Ochs-Ross 2020 | -0.10 | [-4.04, 3.83] |
| Ohtani 2024 | 0.00 | [-3.98, 3.98] |
| Pattanaseri 2024 | -0.19 | [-4.20, 3.82] |
| Phillips 2019 | -0.05 | [-4.01, 3.92] |
| Popova 2019 | -0.05 | [-3.98, 3.88] |
| Price 2022 | -0.70 | [-4.63, 3.24] |
| Ray-Griffith 2017 | 0.00 | [-4.03, 4.03] |
| Rodriguez 2013 | -0.13 | [-4.17, 3.92] |
| Schiweck 2021 | -1.05 | [-5.06, 2.95] |
| Shiroma 2020 | -0.15 | [-4.10, 3.81] |
| Singh 2016 2x doses | -0.11 | [-4.09, 3.86] |
| Singh 2016 3x doses | -0.06 | [-4.04, 3.92] |
| Singh 2016 II | -0.67 | [-4.66, 3.32] |
| Smith-Apeldoorn 2024 | -0.06 | [-2.85, 2.74] |
| Sos 2013 | 0.53 | [-3.46, 4.52] |
| Su 2017 | -0.66 | [-4.61, 3.29] |
| Su 2023 | 0.00 | [-3.94, 3.94] |
| Sumner 2019 | -0.06 | [-4.04, 3.92] |
| Takahashi 2021 | -0.42 | [-4.35, 3.51] |
| Taylor 2018 | 0.00 | [-4.02, 4.02] |
| Tiger 2020 | -0.67 | [-4.66, 3.32] |
| Wang 2024 | 0.00 | [-3.93, 3.93] |
| Xiao 2023 | 0.00 | [-3.98, 3.98] |
| Yang 2023 | -0.69 | [-4.62, 3.24] |
| Zarate 2006 | 0.00 | [-4.02, 4.02] |
| Zarate 2012 | 0.09 | [-3.87, 4.05] |
| Zhou 2024 | -1.14 | [-4.38, 2.11] |
| **Summary** | **0.44** | **[-0.60, 1.51]** |
| **Prediction** | **0.44** | **[-0.91, 1.81]** |

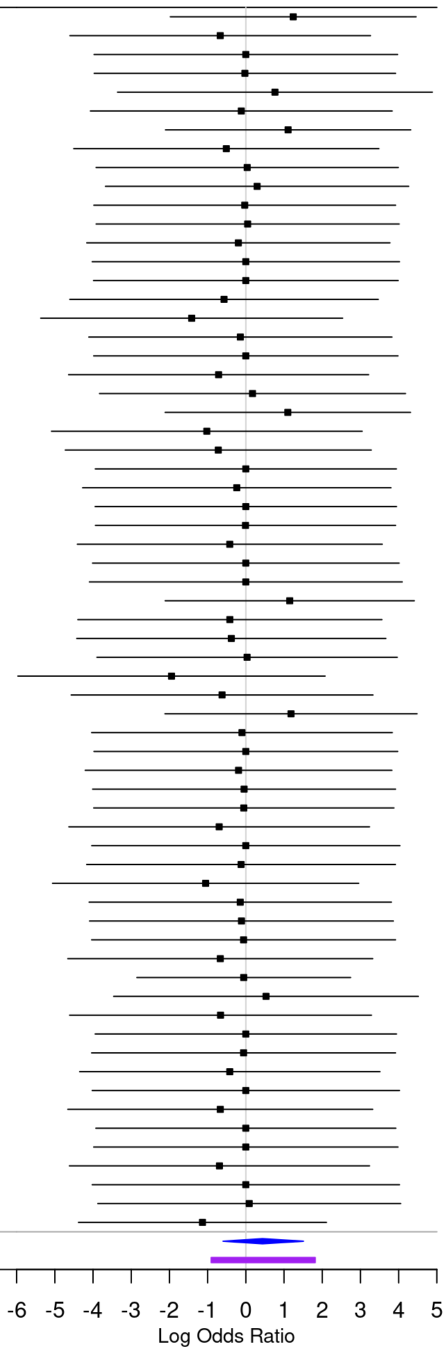

*Heterogeneity (tau): 0.28 [0.00, 0.83]*

**Exact Method**

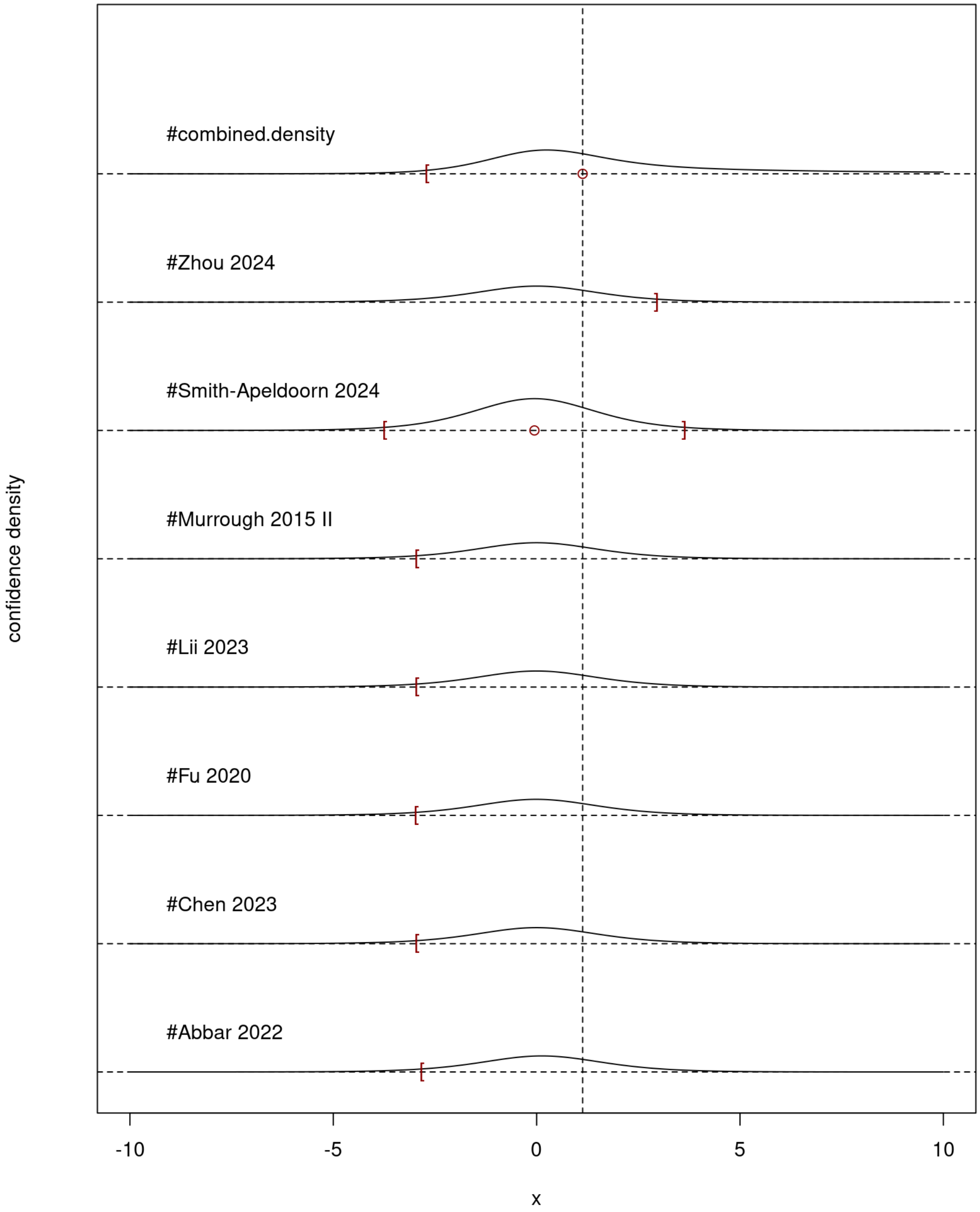

### Suicide Ideation

#### Main Analysis

## Up to 4h

| Study | Experimental Total | Experimental Mean | Experimental SD | Control Total | Control Mean | Control SD | SMD | 95%-CI | Weight (common) | Weight (random) |
|---|---|---|---|---|---|---|---|---|---|---|
| Abbar 2022 | 73 | 6.17 | 8.70 | 83 | 14.33 | 10.11 | -0.86 | [-1.19; -0.53] | 11.9% | 6.3% |
| Ahmed 2023 | 18 | 83.89 | 19.55 | 18 | 80.56 | 13.02 | 0.20 | [-0.46; 0.85] | 3.0% | 4.6% |
| Burger 2016 | 3 | 9.40 | 11.75 | 7 | 22.70 | 11.65 | -1.03 | [-2.50; 0.44] | 0.6% | 1.9% |
| Canuso 2018 | 35 | 4.26 | 2.09 | 31 | 4.81 | 1.70 | -0.28 | [-0.77; 0.20] | 5.4% | 5.5% |
| Domany 2020 | 9 | 13.10 | 14.20 | 9 | 12.30 | 14.70 | 0.05 | [-0.87; 0.98] | 1.5% | 3.4% |
| Domany 2021 | 15 | 8.60 | 9.30 | 15 | 13.60 | 8.60 | -0.54 | [-1.27; 0.19] | 2.4% | 4.3% |
| Fineberg 2023 | 10 | 4.40 | 5.23 | 12 | 3.42 | 5.71 | 0.17 | [-0.67; 1.01] | 1.8% | 3.8% |
| Fu 2020 | 110 | 2.49 | 1.65 | 112 | 2.74 | 1.44 | -0.16 | [-0.42; 0.10] | 18.5% | 6.6% |
| George 2017 | 12 | 2.17 | 1.53 | 4 | 1.75 | 1.50 | 0.26 | [-0.88; 1.40] | 1.0% | 2.7% |
| Glue 2024b | 17 | 0.41 | 0.71 | 8 | 0.25 | 0.46 | 0.24 | [-0.60; 1.08] | 1.8% | 3.8% |
| Grunebaum 2017 | 7 | 5.71 | 6.10 | 9 | 5.89 | 5.58 | -0.03 | [-1.02; 0.96] | 1.3% | 3.2% |
| Grunebaum 2018 | 40 | 5.00 | 5.18 | 40 | 9.85 | 7.21 | -0.77 | [-1.22; -0.31] | 6.2% | 5.7% |
| Hu 2016 | 13 | 0.30 | 0.50 | 14 | 1.30 | 0.40 | -2.15 | [-3.13; -1.18] | 1.4% | 3.2% |
| Ionescu 2019 | 13 | 1.90 | 1.40 | 13 | 1.50 | 1.40 | 0.28 | [-0.50; 1.05] | 2.2% | 4.1% |
| Ionescu 2020 | 113 | 2.25 | 1.47 | 112 | 2.92 | 1.32 | -0.48 | [-0.74; -0.21] | 18.3% | 6.6% |
| Jones 2024 | 17 | 8.50 | 7.99 | 11 | 14.20 | 8.63 | -0.67 | [-1.45; 0.11] | 2.1% | 4.0% |
| Loo 2016 | 9 | 1.44 | 1.31 | 6 | 0.83 | 0.75 | 0.51 | [-0.55; 1.56] | 1.2% | 3.0% |
| Singh 2016 2x doses | 18 | 0.11 | 0.32 | 16 | 0.75 | 1.18 | -0.74 | [-1.44; -0.04] | 2.6% | 4.4% |
| Singh 2016 3x doses | 17 | 0.12 | 0.49 | 16 | 0.50 | 1.03 | -0.46 | [-1.16; 0.23] | 2.7% | 4.5% |
| Su 2023 | 42 | 1.57 | 1.41 | 42 | 2.36 | 1.23 | -0.59 | [-1.03; -0.16] | 6.7% | 5.8% |
| Sumner 2019 | 17 | 0.41 | 0.87 | 16 | 1.00 | 0.89 | -0.65 | [-1.36; 0.05] | 2.6% | 4.4% |
| Zarate 2006 | 9 | 0.12 | 0.35 | 9 | 0.56 | 0.73 | -0.72 | [-1.68; 0.24] | 1.4% | 3.3% |
| Zarate 2012 | 19 | 1.10 | 1.24 | 22 | 1.59 | 1.22 | -0.39 | [-1.01; 0.23] | 3.3% | 4.8% |
| **Common effect model** | **636** | | | **625** | | | **-0.43** | **[-0.54; -0.31]** | **100.0%** | **--** |
| **Random effects model** | | | | | | | **-0.40** | **[-0.63; -0.16]** | **--** | **100.0%** |

Heterogeneity: $I^2 = 52\%$, $\tau^2 = 0.2017$, $p < 0.01$

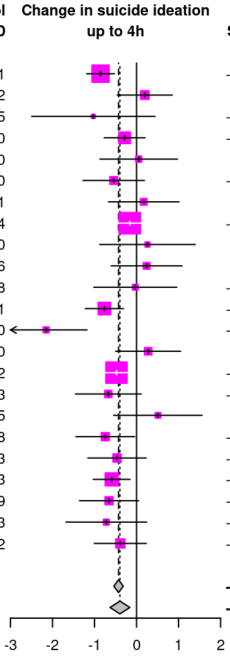

## >12h up to 24h

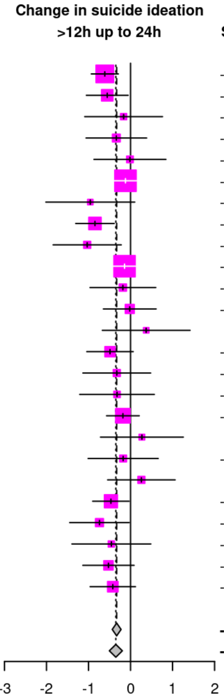

| Study | Experimental Total | Experimental Mean | Experimental SD | Control Total | Control Mean | Control SD | SMD | 95%-CI | Weight (common) | Weight (random) |
|---|---|---|---|---|---|---|---|---|---|---|
| Abbar 2022 | 73 | 5.77 | 9.68 | 83 | 11.97 | 10.41 | -0.61 | [-0.93; -0.29] | 10.7% | 7.4% |
| Canuso 2018 | 35 | 3.29 | 2.47 | 30 | 4.47 | 1.57 | -0.55 | [-1.05; -0.06] | 4.5% | 5.2% |
| Domany 2020 | 9 | 9.20 | 14.80 | 9 | 11.90 | 16.40 | -0.16 | [-1.09; 0.76] | 1.3% | 2.2% |
| Domany 2021 | 15 | 7.80 | 10.50 | 15 | 11.50 | 10.80 | -0.34 | [-1.06; 0.38] | 2.1% | 3.3% |
| Fineberg 2023 | 10 | 3.10 | 5.63 | 11 | 3.18 | 5.67 | -0.01 | [-0.87; 0.84] | 1.5% | 2.5% |
| Fu 2020 | 110 | 2.23 | 1.67 | 112 | 2.43 | 1.63 | -0.12 | [-0.38; 0.14] | 16.0% | 8.3% |
| Grunebaum 2017 | 7 | 4.29 | 5.74 | 9 | 10.11 | 5.75 | -0.96 | [-2.02; 0.10] | 1.0% | 1.8% |
| Grunebaum 2018 | 40 | 6.05 | 5.48 | 40 | 11.63 | 7.43 | -0.85 | [-1.30; -0.39] | 5.3% | 5.6% |
| Hu 2016 | 13 | 0.70 | 0.70 | 14 | 1.30 | 0.40 | -1.03 | [-1.84; -0.22] | 1.7% | 2.7% |
| Ionescu 2020 | 110 | 2.22 | 1.51 | 110 | 2.44 | 1.57 | -0.14 | [-0.41; 0.12] | 15.8% | 8.3% |
| Jones 2024 | 17 | 10.80 | 8.08 | 10 | 12.30 | 7.83 | -0.18 | [-0.96; 0.60] | 1.8% | 2.9% |
| Lii 2023 | 20 | 0.20 | 0.52 | 19 | 0.21 | 0.54 | -0.02 | [-0.65; 0.61] | 2.8% | 3.9% |
| Loo 2016 | 9 | 1.78 | 1.20 | 6 | 1.33 | 1.03 | 0.37 | [-0.67; 1.42] | 1.0% | 1.8% |
| Murrough 2013 | 26 | 0.85 | 1.12 | 26 | 1.46 | 1.33 | -0.49 | [-1.04; 0.06] | 3.6% | 4.6% |
| Murrough 2015 II | 12 | 10.83 | 8.48 | 12 | 14.00 | 10.15 | -0.33 | [-1.13; 0.48] | 1.7% | 2.8% |
| Pattanaseri 2024 | 11 | 1.64 | 1.57 | 9 | 2.11 | 1.16 | -0.32 | [-1.21; 0.57] | 1.4% | 2.4% |
| Price 2022 | 53 | 0.38 | 0.98 | 49 | 0.59 | 1.31 | -0.18 | [-0.57; 0.21] | 7.3% | 6.5% |
| Ray-Griffith 2017 | 8 | 2.87 | 0.99 | 8 | 2.62 | 0.74 | 0.27 | [-0.72; 1.26] | 1.1% | 2.0% |
| Singh 2016 II | 18 | 0.61 | 1.42 | 8 | 0.88 | 1.64 | -0.18 | [-1.01; 0.66] | 1.6% | 2.6% |
| Sos 2013 | 9 | 0.56 | 0.73 | 18 | 0.39 | 0.59 | 0.26 | [-0.55; 1.06] | 1.7% | 2.8% |
| Su 2023 | 42 | 1.48 | 1.42 | 42 | 2.10 | 1.21 | -0.47 | [-0.90; -0.03] | 5.9% | 5.9% |
| Sumner 2019 | 17 | 0.65 | 0.79 | 16 | 1.31 | 0.95 | -0.74 | [-1.45; -0.03] | 2.2% | 3.3% |
| Zarate 2006 | 9 | 0.44 | 0.53 | 9 | 0.78 | 0.83 | -0.46 | [-1.40; 0.48] | 1.3% | 2.2% |
| Zarate 2012 | 20 | 1.30 | 1.38 | 23 | 2.00 | 1.24 | -0.53 | [-1.14; 0.09] | 3.0% | 4.1% |
| Zhou 2024 | 27 | -2.40 | 2.20 | 27 | -1.50 | 2.00 | -0.42 | [-0.96; 0.12] | 3.8% | 4.7% |
| **Common effect model** | **720** | | | **715** | | | **-0.33** | **[-0.44; -0.23]** | **100.0%** | **--** |
| **Random effects model** | | | | | | | **-0.35** | **[-0.51; -0.19]** | **--** | **100.0%** |

Heterogeneity: $I^2 = 12\%$, $\tau^2 = 0.0573$, $p = 0.29$

## >24h up to 72h

| Study | Experimental Total | Experimental Mean | Experimental SD | Control Total | Control Mean | Control SD | SMD | 95%-CI | Weight (common) | Weight (random) |
|---|---|---|---|---|---|---|---|---|---|---|
| Abbar 2022 | 73 | 3.00 | 6.05 | 80 | 8.50 | 10.19 | -0.65 | [-0.97; -0.32] | 6.0% | 5.6% |
| Canuso 2018 | 30 | 2.87 | 2.61 | 28 | 3.32 | 2.33 | -0.18 | [-0.70; 0.34] | 2.4% | 3.5% |
| Chen 2023 | 123 | 0.25 | 0.61 | 126 | 0.23 | 0.54 | 0.03 | [-0.21; 0.28] | 10.3% | 6.8% |
| Daly 2018 PA | 33 | 0.18 | 0.46 | 33 | 0.12 | 0.33 | 0.15 | [-0.34; 0.63] | 2.7% | 3.8% |
| Daly 2018 PB | 20 | 0.25 | 0.72 | 21 | 0.24 | 0.54 | 0.02 | [-0.60; 0.63] | 1.7% | 2.8% |
| Domany 2020 | 9 | 7.00 | 16.30 | 9 | 10.90 | 14.40 | -0.24 | [-1.17; 0.69] | 0.7% | 1.4% |
| Domany 2021 | 15 | 6.30 | 8.90 | 15 | 6.60 | 10.40 | -0.03 | [-0.75; 0.69] | 1.2% | 2.2% |
| Fedgchin 2019 | 218 | 0.18 | 0.55 | 110 | 0.18 | 0.47 | 0.00 | [-0.23; 0.23] | 12.1% | 7.1% |
| Fineberg 2023 | 10 | 3.20 | 4.76 | 11 | 3.00 | 5.27 | 0.04 | [-0.82; 0.89] | 0.9% | 1.6% |
| Fu 2020 | 108 | 1.84 | 1.47 | 109 | 2.13 | 1.55 | -0.19 | [-0.46; 0.08] | 9.0% | 6.5% |
| George 2017 | 12 | 1.42 | 1.38 | 4 | 2.00 | 1.41 | -0.40 | [-1.54; 0.75] | 0.5% | 1.0% |
| Hu 2016 | 12 | 0.50 | 0.70 | 14 | 1.20 | 0.40 | -1.22 | [-2.06; -0.37] | 0.9% | 1.7% |
| Ionescu 2020 | 106 | 1.84 | 1.54 | 109 | 1.95 | 1.60 | -0.07 | [-0.34; 0.20] | 8.9% | 6.5% |
| Jones 2024 | 17 | 7.80 | 7.36 | 11 | 10.50 | 9.65 | -0.32 | [-1.08; 0.45] | 1.1% | 2.0% |
| Lii 2023 | 18 | 0.32 | 0.95 | 20 | 0.35 | 0.59 | -0.04 | [-0.67; 0.60] | 1.6% | 2.6% |
| Loo 2016 | 9 | 1.78 | 1.20 | 6 | 1.33 | 1.03 | 0.37 | [-0.67; 1.42] | 0.6% | 1.2% |
| Murrough 2013 | 26 | 1.15 | 1.29 | 26 | 1.42 | 1.42 | -0.20 | [-0.74; 0.35] | 2.1% | 3.2% |
| Murrough 2015 II | 12 | 9.33 | 9.07 | 12 | 12.92 | 10.20 | -0.36 | [-1.17; 0.45] | 1.0% | 1.8% |
| Ochs-Ross 2020 | 70 | 0.16 | 0.40 | 66 | 0.12 | 0.33 | 0.11 | [-0.23; 0.44] | 5.6% | 5.5% |
| Pattanaseri 2024 | 11 | 1.18 | 1.60 | 9 | 1.22 | 1.30 | -0.03 | [-0.91; 0.86] | 0.8% | 1.6% |
| Popova 2019 | 113 | 0.16 | 0.54 | 109 | 0.12 | 0.35 | 0.09 | [-0.18; 0.35] | 9.2% | 6.5% |
| Singh 2016 2x doses | 18 | 0.00 | 0.00 | 16 | 0.25 | 0.77 | | | 0.0% | 0.0% |
| Singh 2016 3x doses | 17 | 0.12 | 0.49 | 16 | 0.31 | 0.60 | -0.34 | [-1.03; 0.35] | 1.3% | 2.3% |
| Singh 2016 II | 18 | 0.56 | 1.29 | 8 | 0.88 | 1.64 | -0.22 | [-1.06; 0.61] | 0.9% | 1.7% |
| Su 2017 | 47 | 0.72 | 0.74 | 24 | 1.17 | 0.82 | -0.58 | [-1.08; -0.08] | 2.5% | 3.6% |
| Su 2023 | 42 | 1.62 | 1.50 | 42 | 2.12 | 1.21 | -0.36 | [-0.79; 0.07] | 3.4% | 4.3% |
| Takahashi 2021 | 122 | 0.17 | 0.53 | 80 | 0.28 | 0.67 | -0.19 | [-0.47; 0.10] | 8.0% | 6.2% |
| Zarate 2006 | 9 | 0.78 | 1.09 | 9 | 0.67 | 0.87 | 0.11 | [-0.82; 1.03] | 0.7% | 1.4% |
| Zarate 2012 | 15 | 1.47 | 1.46 | 22 | 1.96 | 1.36 | -0.34 | [-1.00; 0.32] | 1.5% | 2.5% |
| Zhou 2024 | 27 | -2.50 | 2.20 | 24 | -1.60 | 2.20 | -0.40 | [-0.96; 0.15] | 2.1% | 3.2% |
| **Common effect model** | **1360** | | | **1169** | | | **-0.13** | **[-0.21; -0.05]** | **100.0%** | **--** |
| **Random effects model** | | | | | | | **-0.16** | **[-0.28; -0.04]** | **--** | **100.0%** |

Heterogeneity: $I^2 = 17\%$, $\tau^2 = 0.0399$, $p = 0.21$

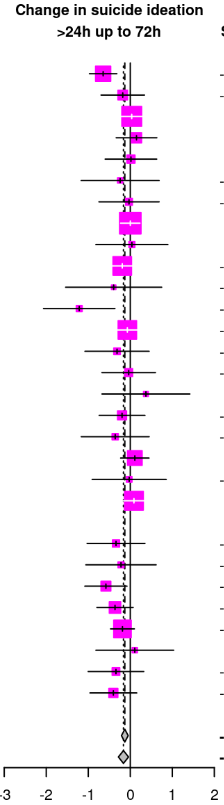

## >72h up to 2w

| Study | Experimental Total | Experimental Mean | Experimental SD | Control Total | Control Mean | Control SD | SMD | 95%-CI | Weight (common) | Weight (random) |
|---|---|---|---|---|---|---|---|---|---|---|
| Abbar 2022 | 73 | 3.50 | 7.18 | 80 | 8.00 | 11.30 | -0.47 | [-0.79; -0.15] | 5.6% | 5.1% |
| Ahmed 2023 | 18 | 75.72 | 20.58 | 18 | 80.67 | 13.15 | -0.28 | [-0.94; 0.38] | 1.3% | 1.9% |
| Anderson 2017 | 35 | 2.94 | 1.76 | 37 | 3.08 | 2.38 | -0.07 | [-0.53; 0.40] | 2.7% | 3.3% |
| Burger 2016 | 3 | 2.40 | 13.61 | 7 | 6.70 | 8.93 | -0.38 | [-1.75; 0.99] | 0.3% | 0.5% |
| Canuso 2018 | 29 | 1.76 | 2.03 | 26 | 1.54 | 1.68 | 0.12 | [-0.41; 0.65] | 2.1% | 2.7% |
| Chen 2023 | 117 | 0.25 | 0.60 | 111 | 0.23 | 0.61 | 0.03 | [-0.23; 0.29] | 8.6% | 6.2% |
| Colla 2024 | 17 | 1.18 | 0.87 | 10 | 1.60 | 0.95 | -0.45 | [-1.24; 0.34] | 0.9% | 1.4% |
| Daly 2018 PA | 40 | 0.20 | 0.55 | 32 | 0.22 | 0.42 | -0.04 | [-0.50; 0.43] | 2.7% | 3.3% |
| Daly 2018 PB | 20 | 0.15 | 0.49 | 21 | 0.38 | 0.59 | -0.41 | [-1.03; 0.20] | 1.5% | 2.1% |
| Domany 2020 | 9 | 7.40 | 18.00 | 9 | 1.40 | 2.20 | 0.45 | [-0.49; 1.38] | 0.7% | 1.0% |
| Domany 2021 | 15 | 3.10 | 14.10 | 15 | 11.40 | 15.20 | -0.55 | [-1.28; 0.18] | 1.1% | 1.6% |
| Duek 2023 | 14 | 0.14 | 0.36 | 13 | 0.08 | 0.28 | 0.20 | [-0.56; 0.95] | 1.0% | 1.5% |
| Fedgchin 2019 | 211 | 0.16 | 0.46 | 110 | 0.18 | 0.39 | -0.05 | [-0.28; 0.18] | 10.9% | 6.7% |
| Fineberg 2023 | 10 | 4.30 | 7.10 | 12 | 5.67 | 7.04 | -0.19 | [-1.03; 0.65] | 0.8% | 1.3% |
| Fu 2020 | 105 | 1.28 | 1.31 | 99 | 1.33 | 1.14 | -0.04 | [-0.32; 0.23] | 7.7% | 5.9% |
| George 2017 | 12 | 1.25 | 1.21 | 4 | 2.50 | 1.73 | -0.88 | [-2.07; 0.30] | 0.4% | 0.7% |
| Grabski 2022 | 41 | 0.10 | 0.30 | 44 | 0.00 | 0.00 | | | 0.0% | 0.0% |
| Hu 2016 | 12 | 0.90 | 0.90 | 14 | 1.00 | 0.50 | -0.14 | [-0.91; 0.64] | 1.0% | 1.5% |
| Ionescu 2019 | 13 | 1.30 | 1.40 | 13 | 1.50 | 1.40 | -0.14 | [-0.91; 0.63] | 1.0% | 1.5% |
| Ionescu 2020 | 92 | 0.97 | 1.19 | 100 | 1.35 | 1.34 | -0.30 | [-0.58; -0.01] | 7.1% | 5.7% |
| Lii 2023 | 19 | 0.32 | 0.58 | 20 | 0.40 | 0.77 | -0.11 | [-0.74; 0.51] | 1.5% | 2.1% |
| Loo 2016 | 9 | 2.00 | 1.66 | 5 | 1.60 | 0.89 | 0.26 | [-0.84; 1.36] | 0.5% | 0.8% |
| Murrough 2015 II | 12 | 7.92 | 8.73 | 12 | 10.08 | 9.39 | -0.23 | [-1.03; 0.57] | 0.9% | 1.4% |
| Ochs-Ross 2020 | 65 | 0.21 | 0.65 | 62 | 0.18 | 0.58 | 0.05 | [-0.30; 0.40] | 4.8% | 4.7% |
| Pattanaseri 2024 | 11 | 1.64 | 1.75 | 9 | 1.44 | 1.33 | 0.12 | [-0.76; 1.00] | 0.7% | 1.2% |
| Popova 2019 | 109 | 0.08 | 0.36 | 107 | 0.18 | 0.53 | -0.22 | [-0.49; 0.05] | 8.1% | 6.0% |
| Price 2022 | 52 | 0.79 | 1.32 | 46 | 1.30 | 1.62 | -0.34 | [-0.74; 0.05] | 3.6% | 4.0% |
| Singh 2016 2x doses | 15 | 0.07 | 0.26 | 12 | 0.33 | 0.65 | -0.53 | [-1.31; 0.24] | 1.0% | 1.5% |
| Singh 2016 3x doses | 15 | 0.00 | 0.00 | 12 | 0.25 | 0.45 | | | 0.0% | 0.0% |
| Smith-Apeldoorn 2024 | 53 | 1.34 | 0.90 | 51 | 1.47 | 0.83 | -0.15 | [-0.53; 0.24] | 3.9% | 4.2% |
| Sos 2013 | 9 | 0.44 | 0.73 | 18 | 0.17 | 0.87 | 0.32 | [-0.49; 1.12] | 0.9% | 1.4% |
| Su 2017 | 47 | 0.98 | 0.87 | 24 | 1.29 | 0.81 | -0.36 | [-0.86; 0.13] | 2.4% | 3.0% |
| Su 2023 | 42 | 1.95 | 1.36 | 42 | 2.12 | 1.33 | -0.13 | [-0.55; 0.30] | 3.2% | 3.6% |
| Sumner 2019 | 17 | 0.94 | 0.97 | 16 | 1.63 | 0.96 | -0.70 | [-1.40; 0.01] | 1.2% | 1.7% |
| Takahashi 2021 | 116 | 0.18 | 0.61 | 74 | 0.27 | 0.76 | -0.13 | [-0.43; 0.16] | 6.8% | 5.6% |
| Zarate 2006 | 9 | 0.89 | 1.27 | 9 | 1.00 | 0.82 | -0.10 | [-1.02; 0.83] | 0.7% | 1.1% |
| Zarate 2012 | 11 | 2.09 | 1.38 | 17 | 2.12 | 1.05 | -0.02 | [-0.78; 0.74] | 1.0% | 1.5% |
| Zhou 2024 | 27 | -2.80 | 2.10 | 23 | -2.20 | 2.50 | -0.26 | [-0.82; 0.30] | 1.9% | 2.5% |
| **Common effect model** | **1524** | | | **1334** | | | **-0.15** | **[-0.23; -0.07]** | **100.0%** | **--** |
| **Random effects model** | | | | | | | **-0.16** | **[-0.26; -0.06]** | **--** | **100.0%** |

Heterogeneity: $I^2 = 0\%$, $\tau^2 = 0.0259$, $p = 0.90$

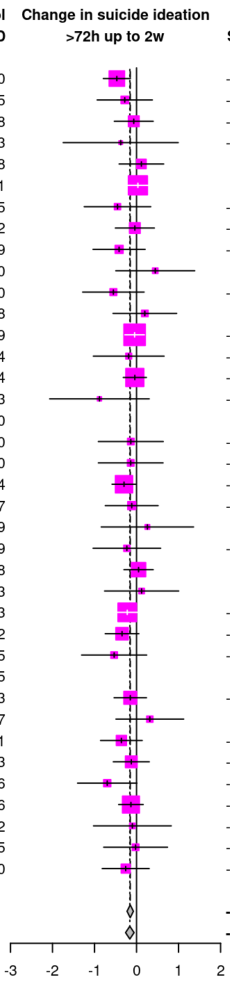

## >2w up to 4w

| Study | Experimental Total | Experimental Mean | Experimental SD | Control Total | Control Mean | Control SD | SMD | 95%-CI | Weight (common) | Weight (random) |
|---|---|---|---|---|---|---|---|---|---|---|
| Abbar 2022 | 73 | 2.83 | 5.67 | 80 | 6.43 | 9.66 | -0.45 | [-0.77; -0.13] | 7.4% | 7.1% |
| Canuso 2018 | 26 | 1.00 | 1.44 | 21 | 0.90 | 1.37 | 0.07 | [-0.51; 0.64] | 2.3% | 3.8% |
| Chen 2023 | 108 | 0.26 | 0.62 | 107 | 0.21 | 0.53 | 0.09 | [-0.18; 0.35] | 10.7% | 8.1% |
| Domany 2021 | 15 | 3.00 | 14.50 | 15 | 8.30 | 16.00 | -0.34 | [-1.06; 0.38] | 1.5% | 2.8% |
| Duek 2023 | 13 | 0.31 | 0.48 | 11 | 0.18 | 0.41 | 0.27 | [-0.54; 1.08] | 1.2% | 2.3% |
| Fedgchin 2019 | 204 | 0.11 | 0.37 | 107 | 0.17 | 0.40 | -0.16 | [-0.39; 0.08] | 13.9% | 8.7% |
| Fineberg 2023 | 10 | 7.10 | 6.01 | 11 | 4.73 | 5.75 | 0.39 | [-0.48; 1.25] | 1.0% | 2.1% |
| Fu 2020 | 100 | 0.97 | 1.27 | 93 | 1.08 | 1.21 | -0.09 | [-0.37; 0.19] | 9.6% | 7.8% |
| Grabski 2022 | 39 | 0.05 | 0.22 | 43 | 0.00 | 0.00 | | | 0.0% | 0.0% |
| Hu 2016 | 12 | 0.40 | 0.90 | 14 | 0.20 | 0.40 | 0.29 | [-0.49; 1.06] | 1.3% | 2.5% |
| Ionescu 2019 | 12 | 1.20 | 1.40 | 12 | 1.40 | 1.40 | -0.14 | [-0.94; 0.66] | 1.2% | 2.3% |
| Ionescu 2020 | 90 | 0.68 | 1.04 | 92 | 0.93 | 1.18 | -0.22 | [-0.52; 0.07] | 9.0% | 7.7% |
| Ochs-Ross 2020 | 62 | 0.18 | 0.51 | 61 | 0.17 | 0.45 | 0.02 | [-0.33; 0.37] | 6.1% | 6.6% |
| Ohtani 2024 | 15 | 1.40 | 1.08 | 16 | 1.94 | 1.14 | -0.47 | [-1.19; 0.25] | 1.5% | 2.8% |
| Popova 2019 | 103 | 0.07 | 0.29 | 102 | 0.08 | 0.31 | -0.03 | [-0.31; 0.24] | 10.2% | 8.0% |
| Price 2022 | 49 | 0.61 | 1.27 | 41 | 1.68 | 1.71 | -0.71 | [-1.14; -0.29] | 4.2% | 5.5% |
| Smith-Apeldoorn 2024 | 50 | 1.42 | 0.83 | 49 | 1.39 | 1.02 | 0.03 | [-0.36; 0.43] | 4.9% | 5.9% |
| Su 2017 | 47 | 1.00 | 0.75 | 24 | 1.33 | 1.01 | -0.39 | [-0.88; 0.11] | 3.1% | 4.6% |
| Takahashi 2021 | 111 | 0.15 | 0.65 | 72 | 0.14 | 0.54 | 0.02 | [-0.28; 0.31] | 8.7% | 7.6% |
| Zhou 2024 | 26 | -2.90 | 2.20 | 23 | -1.90 | 2.30 | -0.44 | [-1.01; 0.13] | 2.4% | 3.9% |
| **Common effect model** | **1165** | | | **994** | | | **-0.13** | **[-0.21; -0.04]** | **100.0%** | **--** |
| **Random effects model** | | | | | | | **-0.14** | **[-0.27; 0.00]** | **--** | **100.0%** |

Heterogeneity: $I^2 = 25\%$, $\tau^2 = 0.0414$, $p = 0.16$

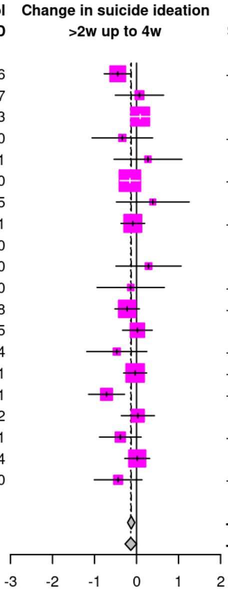

**>4w**

| Study | Experimental Total | Experimental Mean | Experimental SD | Control Total | Control Mean | Control SD | SMD | 95%-CI | Weight (common) | Weight (random) |
|---|---|---|---|---|---|---|---|---|---|---|
| Abbar 2022 | 60 | 2.60 | 5.90 | 66 | 4.00 | 8.30 | -0.19 | [-0.54; 0.16] | 15.9% | 15.1% |
| Duek 2023 | 11 | 0.30 | 0.48 | 11 | 0.20 | 0.42 | 0.21 | [-0.63; 1.05] | 2.8% | 4.1% |
| Fu 2020 | 84 | 1.04 | 1.31 | 79 | 0.84 | 1.20 | 0.16 | [-0.15; 0.47] | 20.7% | 17.3% |
| Grabski 2022 | 38 | 0.05 | 0.23 | 43 | 0.07 | 0.26 | -0.07 | [-0.51; 0.37] | 10.3% | 11.5% |
| Ionescu 2020 | 80 | 0.49 | 1.08 | 86 | 0.66 | 0.98 | -0.16 | [-0.47; 0.14] | 21.0% | 17.5% |
| Pattanaseri 2024 | 11 | 1.64 | 1.50 | 9 | 1.89 | 1.27 | -0.17 | [-1.05; 0.71] | 2.5% | 3.7% |
| Price 2022 | 36 | 0.56 | 0.97 | 35 | 0.60 | 0.85 | -0.04 | [-0.51; 0.42] | 9.0% | 10.5% |
| Smith-Apeldoorn 2024 | 48 | 1.67 | 0.75 | 46 | 1.35 | 0.82 | 0.40 | [-0.01; 0.81] | 11.7% | 12.5% |
| Zhou 2024 | 26 | -2.80 | 2.10 | 23 | -2.10 | 2.30 | -0.31 | [-0.88; 0.25] | 6.1% | 7.9% |
| **Common effect model** | **394** | | | **398** | | | **-0.01** | **[-0.15; 0.13]** | **100.0%** | **--** |
| **Random effects model** | | | | | | | **-0.01** | **[-0.20; 0.17]** | **--** | **100.0%** |

Heterogeneity: $I^2 = 8\%$, $\tau^2 = 0.0242$, $p = 0.37$

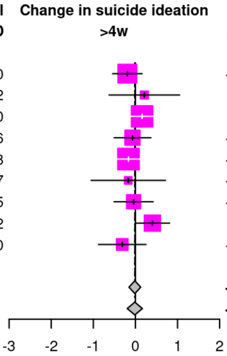

#### End of Treatment

| Study | Experimental Total | Experimental Mean | Experimental SD | Control Total | Control Mean | Control SD | SMD | 95%-CI | Weight (common) | Weight (random) |
|---|---|---|---|---|---|---|---|---|---|---|
| Abbar 2022 (day 2) | 73 | 3.00 | 6.05 | 80 | 8.50 | 10.19 | -0.65 | [-0.97; -0.32] | 5.4% | 4.4% |
| Ahmed 2023 (2w) | 18 | 75.72 | 20.58 | 18 | 80.67 | 13.15 | -0.28 | [-0.94; 0.38] | 1.3% | 2.1% |
| Canuso 2018 (25d) | 26 | 1.00 | 1.44 | 21 | 0.90 | 1.37 | 0.07 | [-0.51; 0.64] | 1.7% | 2.5% |
| Chen 2023 (4w) | 108 | 0.26 | 0.62 | 107 | 0.21 | 0.53 | 0.09 | [-0.18; 0.35] | 8.0% | 5.0% |
| Colla 2024 (2w) | 17 | 1.18 | 0.87 | 10 | 1.60 | 0.95 | -0.45 | [-1.24; 0.34] | 0.9% | 1.6% |
| Daly 2018 PA (7d) | 40 | 0.20 | 0.55 | 32 | 0.22 | 0.42 | -0.04 | [-0.50; 0.43] | 2.6% | 3.2% |
| Daly 2018 PB (7d) | 20 | 0.15 | 0.49 | 21 | 0.38 | 0.59 | -0.41 | [-1.03; 0.20] | 1.5% | 2.3% |
| Domany 2020 (24h) | 9 | 9.20 | 14.80 | 9 | 11.90 | 16.40 | -0.16 | [-1.09; 0.76] | 0.7% | 1.2% |
| Domany 2021 (24h) | 15 | 7.80 | 10.50 | 15 | 11.50 | 10.80 | -0.34 | [-1.06; 0.38] | 1.1% | 1.8% |
| Fedgchin 2019 (4w) | 204 | 0.11 | 0.37 | 107 | 0.17 | 0.40 | -0.16 | [-0.39; 0.08] | 10.4% | 5.3% |
| Fineberg 2023 (24h) | 10 | 3.10 | 5.63 | 11 | 3.18 | 5.67 | -0.01 | [-0.87; 0.84] | 0.8% | 1.4% |
| Fu 2020 (25d) | 100 | 0.97 | 1.27 | 93 | 1.08 | 1.21 | -0.09 | [-0.37; 0.19] | 7.1% | 4.8% |
| George 2017 (1w) | 12 | 1.25 | 1.21 | 4 | 2.50 | 1.73 | -0.88 | [-2.07; 0.30] | 0.4% | 0.8% |
| Grabski 2022 (1-3w) | 38 | 0.05 | 0.23 | 43 | 0.07 | 0.26 | -0.07 | [-0.51; 0.37] | 3.0% | 3.4% |
| Grunebaum 2017 (24h) | 7 | 4.29 | 5.74 | 9 | 10.11 | 5.75 | -0.96 | [-2.02; 0.10] | 0.5% | 1.0% |
| Grunebaum 2018 (24h) | 40 | 6.05 | 5.48 | 40 | 11.63 | 7.43 | -0.85 | [-1.30; -0.39] | 2.7% | 3.3% |
| Hu 2016 (24h) | 13 | 0.70 | 0.70 | 14 | 1.30 | 0.40 | -1.03 | [-1.84; -0.22] | 0.9% | 1.5% |
| Ionescu 2019 (3w) | 12 | 1.20 | 1.40 | 12 | 1.40 | 1.40 | -0.14 | [-0.94; 0.66] | 0.9% | 1.6% |
| Ionescu 2020 (25d) | 90 | 0.68 | 1.04 | 92 | 0.93 | 1.18 | -0.22 | [-0.52; 0.07] | 6.7% | 4.8% |
| Jones 2024 (24h) | 17 | 10.80 | 8.08 | 10 | 12.30 | 7.83 | -0.18 | [-0.96; 0.60] | 0.9% | 1.6% |
| Lii 2023 (24h) | 20 | 0.20 | 0.52 | 19 | 0.21 | 0.54 | -0.02 | [-0.65; 0.61] | 1.4% | 2.2% |
| Loo 2016 (1w-2w) | 9 | 2.00 | 1.66 | 5 | 1.60 | 0.89 | 0.26 | [-0.84; 1.36] | 0.5% | 0.9% |
| Murrough 2013 (24h) | 26 | 0.85 | 1.12 | 26 | 1.46 | 1.33 | -0.49 | [-1.04; 0.06] | 1.9% | 2.6% |
| Murrough 2015 II (24h) | 12 | 10.83 | 8.48 | 12 | 14.00 | 10.15 | -0.33 | [-1.13; 0.48] | 0.9% | 1.5% |
| Ochs-Ross 2020 (4w) | 62 | 0.18 | 0.51 | 61 | 0.17 | 0.45 | 0.02 | [-0.33; 0.37] | 4.6% | 4.1% |
| Ohtani 2024 (2w) | 15 | 1.40 | 1.08 | 16 | 1.94 | 1.14 | -0.47 | [-1.19; 0.25] | 1.1% | 1.8% |
| Pattanaseri 2024 (3d) | 11 | 1.18 | 1.60 | 9 | 1.22 | 1.30 | -0.03 | [-0.91; 0.86] | 0.7% | 1.3% |
| Popova 2019 (4w) | 103 | 0.07 | 0.29 | 102 | 0.08 | 0.31 | -0.03 | [-0.31; 0.24] | 7.6% | 4.9% |
| Price 2022 (24h) | 53 | 0.38 | 0.98 | 49 | 0.59 | 1.31 | -0.18 | [-0.57; 0.21] | 3.8% | 3.8% |
| Singh 2016 2x doses (2w) | 15 | 0.07 | 0.26 | 12 | 0.33 | 0.65 | -0.53 | [-1.31; 0.24] | 1.0% | 1.6% |
| Singh 2016 3x doses (2w) | 15 | 0.00 | 0.00 | 12 | 0.25 | 0.45 | | | 0.0% | 0.0% |
| Singh 2016 II (4d) | 18 | 0.56 | 1.29 | 8 | 0.88 | 1.64 | -0.22 | [-1.06; 0.61] | 0.8% | 1.5% |
| Smith-Apeldoorn 2024 (6w) | 50 | 1.60 | 0.83 | 48 | 1.42 | 0.85 | 0.21 | [-0.18; 0.61] | 3.6% | 3.7% |
| Sos 2013 (24h) | 9 | 0.56 | 0.73 | . | 0.39 | 0.59 | | | 0.0% | 0.0% |
| Su 2023 (24h) | 42 | 1.48 | 1.42 | 42 | 2.10 | 1.21 | -0.47 | [-0.90; -0.03] | 3.0% | 3.4% |
| Sumner 2019 (24h) | 17 | 0.65 | 0.79 | 16 | 1.31 | 0.95 | -0.74 | [-1.45; -0.03] | 1.1% | 1.9% |
| Takahashi 2021 (4w) | 111 | 0.15 | 0.65 | 72 | 0.14 | 0.54 | 0.02 | [-0.28; 0.31] | 6.5% | 4.7% |
| Zarate 2006 (24h) | 9 | 0.44 | 0.53 | 9 | 0.78 | 0.83 | -0.46 | [-1.40; 0.48] | 0.6% | 1.2% |
| Zarate 2012 (24h) | 20 | 1.30 | 1.38 | 23 | 2.00 | 1.24 | -0.53 | [-1.14; 0.09] | 1.5% | 2.3% |
| Zhou 2024 (6d) | 27 | -2.60 | 2.00 | 25 | -1.70 | 2.20 | -0.42 | [-0.97; 0.13] | 1.9% | 2.6% |
| **Common effect model** | **1513** | | | **1314** | | | **-0.19** | **[-0.27; -0.12]** | **100.0%** | **--** |
| **Random effects model** | | | | | | | **-0.24** | **[-0.35; -0.13]** | **--** | **100.0%** |

Heterogeneity: $I^2 = 24\%$, $\tau^2 = 0.0484$, $p = 0.10$

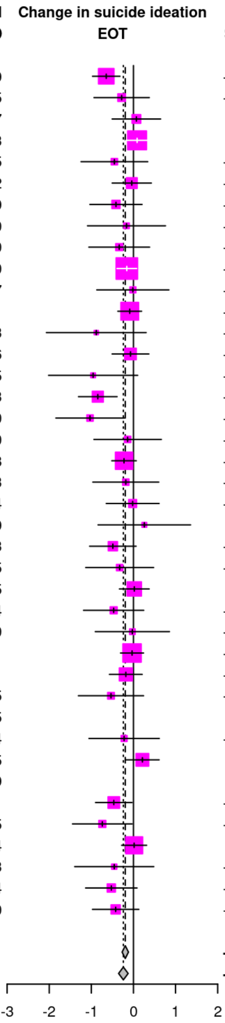

### Subgroup Analysis - Drug

## Up to 4h

| Study | Experimental Total | Experimental Mean | Experimental SD | Control Total | Control Mean | Control SD | SMD | 95%-CI | Weight (common) | Weight (random) |
|---|---|---|---|---|---|---|---|---|---|---|
| **drug = Esketamine** | | | | | | | | | | |
| Canuso 2018 | 35 | 4.26 | 2.09 | 31 | 4.81 | 1.70 | -0.28 | [-0.77; 0.20] | 5.4% | 5.5% |
| Fu 2020 | 110 | 2.49 | 1.65 | 112 | 2.74 | 1.44 | -0.16 | [-0.42; 0.10] | 18.5% | 6.6% |
| Ionescu 2020 | 113 | 2.25 | 1.47 | 112 | 2.92 | 1.32 | -0.48 | [-0.74; -0.21] | 18.3% | 6.6% |
| Common effect model | **258** | | | **255** | | | **-0.31** | **[-0.49; -0.14]** | **42.3%** | **--** |
| Random effects model | | | | | | | **-0.31** | **[-0.53; -0.09]** | **--** | **18.8%** |
| Heterogeneity: $I^2$ = 28%, $\tau^2$ = 0.0122, $p$ = 0.25 | | | | | | | | | | |
| **drug = Ketamine** | | | | | | | | | | |
| Abbar 2022 | 73 | 6.17 | 8.70 | 83 | 14.33 | 10.11 | -0.86 | [-1.19; -0.53] | 11.9% | 6.3% |
| Ahmed 2023 | 18 | 83.89 | 19.55 | 18 | 80.56 | 13.02 | 0.20 | [-0.46; 0.85] | 3.0% | 4.6% |
| Burger 2016 | 3 | 9.40 | 11.75 | 7 | 22.70 | 11.65 | -1.03 | [-2.50; 0.44] | 0.6% | 1.9% |
| Domany 2020 | 9 | 13.10 | 14.20 | 9 | 12.30 | 14.70 | 0.05 | [-0.87; 0.98] | 1.5% | 3.4% |
| Domany 2021 | 15 | 8.60 | 9.30 | 15 | 13.60 | 8.60 | -0.54 | [-1.27; 0.19] | 2.4% | 4.3% |
| Fineberg 2023 | 10 | 4.40 | 5.23 | 12 | 3.42 | 5.71 | 0.17 | [-0.67; 1.01] | 1.8% | 3.8% |
| George 2017 | 12 | 2.17 | 1.53 | 4 | 1.75 | 1.50 | 0.26 | [-0.88; 1.40] | 1.0% | 2.7% |
| Glue 2024b | 17 | 0.41 | 0.71 | 8 | 0.25 | 0.46 | 0.24 | [-0.60; 1.08] | 1.8% | 3.8% |
| Grunebaum 2017 | 7 | 5.71 | 6.10 | 9 | 5.89 | 5.58 | -0.03 | [-1.02; 0.96] | 1.3% | 3.2% |
| Grunebaum 2018 | 40 | 5.00 | 5.18 | 40 | 9.85 | 7.21 | -0.77 | [-1.22; -0.31] | 6.2% | 5.7% |
| Hu 2016 | 13 | 0.30 | 0.50 | 14 | 1.30 | 0.40 | -2.15 | [-3.13; -1.18] | 1.4% | 3.2% |
| Ionescu 2019 | 13 | 1.90 | 1.40 | 13 | 1.50 | 1.40 | 0.28 | [-0.50; 1.05] | 2.2% | 4.1% |
| Jones 2024 | 17 | 8.50 | 7.99 | 11 | 14.20 | 8.63 | -0.67 | [-1.45; 0.11] | 2.1% | 4.0% |
| Loo 2016 | 9 | 1.44 | 1.31 | 6 | 0.83 | 0.75 | 0.51 | [-0.55; 1.56] | 1.2% | 3.0% |
| Singh 2016 2x doses | 18 | 0.11 | 0.32 | 16 | 0.75 | 1.18 | -0.74 | [-1.44; -0.04] | 2.6% | 4.4% |
| Singh 2016 3x doses | 17 | 0.12 | 0.49 | 16 | 0.50 | 1.03 | -0.46 | [-1.16; 0.23] | 2.7% | 4.5% |
| Su 2023 | 42 | 1.57 | 1.41 | 42 | 2.36 | 1.23 | -0.59 | [-1.03; -0.16] | 6.7% | 5.8% |
| Sumner 2019 | 17 | 0.41 | 0.87 | 16 | 1.00 | 0.89 | -0.65 | [-1.36; 0.05] | 2.6% | 4.4% |
| Zarate 2006 | 9 | 0.12 | 0.35 | 9 | 0.56 | 0.73 | -0.72 | [-1.68; 0.24] | 1.4% | 3.3% |
| Zarate 2012 | 19 | 1.10 | 1.24 | 22 | 1.59 | 1.22 | -0.39 | [-1.01; 0.23] | 3.3% | 4.8% |
| Common effect model | **378** | | | **370** | | | **-0.51** | **[-0.66; -0.36]** | **57.7%** | **--** |
| Random effects model | | | | | | | **-0.41** | **[-0.69; -0.13]** | **--** | **81.2%** |
| Heterogeneity: $I^2$ = 52%, $\tau^2$ = 0.2413, $p$ < 0.01 | | | | | | | | | | |
| **Common effect model** | **636** | | | **625** | | | **-0.43** | **[-0.54; -0.31]** | **100.0%** | **--** |
| **Random effects model** | | | | | | | **-0.40** | **[-0.63; -0.16]** | **--** | **100.0%** |

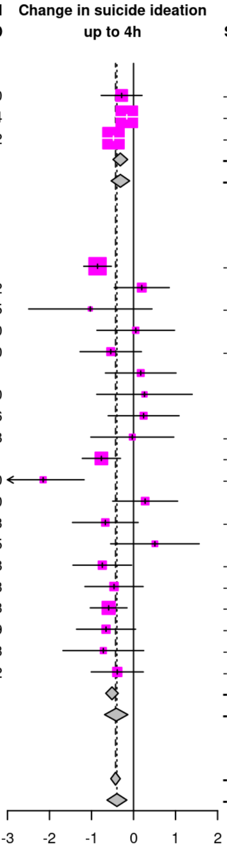

Heterogeneity: $I^2$ = 52%, $\tau^2$ = 0.2017, $p$ < 0.01
Test for subgroup differences (common effect): $\chi^2_1$ = 2.86, df = 1 ($p$ = 0.09)
RE model$\chi^2_1$ = 0.30, df = 1 ($p$ = 0.58)

## >12h up to 24h

| Study | Experimental Total | Experimental Mean | Experimental SD | Control Total | Control Mean | Control SD | SMD | 95%-CI | Weight (common) | Weight (random) |
|---|---|---|---|---|---|---|---|---|---|---|
| **drug = Esketamine** | | | | | | | | | | |
| Canuso 2018 | 35 | 3.29 | 2.47 | 30 | 4.47 | 1.57 | -0.55 | [-1.05; -0.06] | 4.5% | 5.2% |
| Fu 2020 | 110 | 2.23 | 1.67 | 112 | 2.43 | 1.63 | -0.12 | [-0.38; 0.14] | 16.0% | 8.3% |
| Ionescu 2020 | 110 | 2.22 | 1.51 | 110 | 2.44 | 1.57 | -0.14 | [-0.41; 0.12] | 15.8% | 8.3% |
| Singh 2016 II | 18 | 0.61 | 1.42 | 8 | 0.88 | 1.64 | -0.18 | [-1.01; 0.66] | 1.6% | 2.6% |
| Zhou 2024 | 27 | -2.40 | 2.20 | 27 | -1.50 | 2.00 | -0.42 | [-0.96; 0.12] | 3.8% | 4.7% |
| Common effect model | **300** | | | **287** | | | **-0.21** | **[-0.37; -0.04]** | **41.6%** | **--** |
| Random effects model | | | | | | | **-0.23** | **[-0.43; -0.02]** | **--** | **29.2%** |
| Heterogeneity: $I^2 = 0\%$, $\tau^2 = 0.0140$, $p = 0.54$ | | | | | | | | | | |
| **drug = Ketamine** | | | | | | | | | | |
| Abbar 2022 | 73 | 5.77 | 9.68 | 83 | 11.97 | 10.41 | -0.61 | [-0.93; -0.29] | 10.7% | 7.4% |
| Domany 2020 | 9 | 9.20 | 14.80 | 9 | 11.90 | 16.40 | -0.16 | [-1.09; 0.76] | 1.3% | 2.2% |
| Domany 2021 | 15 | 7.80 | 10.50 | 15 | 11.50 | 10.80 | -0.34 | [-1.06; 0.38] | 2.1% | 3.3% |
| Fineberg 2023 | 10 | 3.10 | 5.63 | 11 | 3.18 | 5.67 | -0.01 | [-0.87; 0.84] | 1.5% | 2.5% |
| Grunebaum 2017 | 7 | 4.29 | 5.74 | 9 | 10.11 | 5.75 | -0.96 | [-2.02; 0.10] | 1.0% | 1.8% |
| Grunebaum 2018 | 40 | 6.05 | 5.48 | 40 | 11.63 | 7.43 | -0.85 | [-1.30; -0.39] | 5.3% | 5.6% |
| Hu 2016 | 13 | 0.70 | 0.70 | 14 | 1.30 | 0.40 | -1.03 | [-1.84; -0.22] | 1.7% | 2.7% |
| Jones 2024 | 17 | 10.80 | 8.08 | 10 | 12.30 | 7.83 | -0.18 | [-0.96; 0.60] | 1.8% | 2.9% |
| Lii 2023 | 20 | 0.20 | 0.52 | 19 | 0.21 | 0.54 | -0.02 | [-0.65; 0.61] | 2.8% | 3.9% |
| Loo 2016 | 9 | 1.78 | 1.20 | 6 | 1.33 | 1.03 | 0.37 | [-0.67; 1.42] | 1.0% | 1.8% |
| Murrough 2013 | 26 | 0.85 | 1.12 | 26 | 1.46 | 1.33 | -0.49 | [-1.04; 0.06] | 3.6% | 4.6% |
| Murrough 2015 II | 12 | 10.83 | 8.48 | 12 | 14.00 | 10.15 | -0.33 | [-1.13; 0.48] | 1.7% | 2.8% |
| Pattanaseri 2024 | 11 | 1.64 | 1.57 | 9 | 2.11 | 1.16 | -0.32 | [-1.21; 0.57] | 1.4% | 2.4% |
| Price 2022 | 53 | 0.38 | 0.98 | 49 | 0.59 | 1.31 | -0.18 | [-0.57; 0.21] | 7.3% | 6.5% |
| Ray-Griffith 2017 | 8 | 2.87 | 0.99 | 8 | 2.62 | 0.74 | 0.27 | [-0.72; 1.26] | 1.1% | 2.0% |
| Sos 2013 | 9 | 0.56 | 0.73 | 18 | 0.39 | 0.59 | 0.26 | [-0.55; 1.06] | 1.7% | 2.8% |
| Su 2023 | 42 | 1.48 | 1.42 | 42 | 2.10 | 1.21 | -0.47 | [-0.90; -0.03] | 5.9% | 5.9% |
| Sumner 2019 | 17 | 0.65 | 0.79 | 16 | 1.31 | 0.95 | -0.74 | [-1.45; -0.03] | 2.2% | 3.3% |
| Zarate 2006 | 9 | 0.44 | 0.53 | 9 | 0.78 | 0.83 | -0.46 | [-1.40; 0.48] | 1.3% | 2.2% |
| Zarate 2012 | 20 | 1.30 | 1.38 | 23 | 2.00 | 1.24 | -0.53 | [-1.14; 0.09] | 3.0% | 4.1% |
| Common effect model | **420** | | | **428** | | | **-0.42** | **[-0.56; -0.28]** | **58.4%** | **--** |
| Random effects model | | | | | | | **-0.38** | **[-0.58; -0.19]** | **--** | **70.8%** |
| Heterogeneity: $I^2 = 7\%$, $\tau^2 = 0.0715$, $p = 0.37$ | | | | | | | | | | |
| **Common effect model** | **720** | | | **715** | | | **-0.33** | **[-0.44; -0.23]** | **100.0%** | **--** |
| **Random effects model** | | | | | | | **-0.35** | **[-0.51; -0.19]** | **--** | **100.0%** |

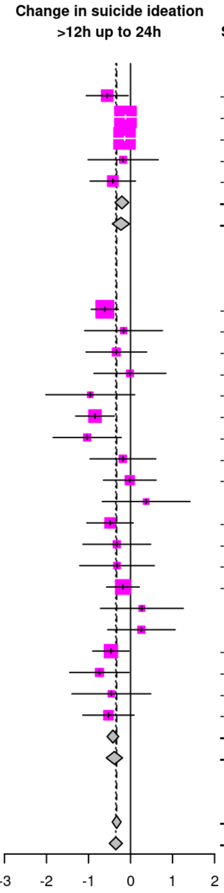

Heterogeneity: $I^2 = 12\%$, $\tau^2 = 0.0573$, $p = 0.29$
Test for subgroup differences (common effect): $\chi^2_1 = 3.88$, df = 1 ($p = 0.05$)
RE model$\chi^2_1 = 1.16$, df = 1 ($p = 0.28$)

## >24h up to 72h

Change in suicide ideation >24h up to 72h

| Study | Experimental Total | Experimental Mean | Experimental SD | Control Total | Control Mean | Control SD | SMD | 95%-CI | Weight (common) | Weight (random) |
|---|---|---|---|---|---|---|---|---|---|---|
| drug = Esketamine | | | | | | | | | | |
| Canuso 2018 | 30 | 2.87 | 2.61 | 28 | 3.32 | 2.33 | -0.18 | [-0.70; 0.34] | 2.4% | 3.5% |
| Chen 2023 | 123 | 0.25 | 0.61 | 126 | 0.23 | 0.54 | 0.03 | [-0.21; 0.28] | 10.3% | 6.8% |
| Daly 2018 PA | 33 | 0.18 | 0.46 | 33 | 0.12 | 0.33 | 0.15 | [-0.34; 0.63] | 2.7% | 3.8% |
| Daly 2018 PB | 20 | 0.25 | 0.72 | 21 | 0.24 | 0.54 | 0.02 | [-0.60; 0.63] | 1.7% | 2.8% |
| Fedgchin 2019 | 218 | 0.18 | 0.55 | 110 | 0.18 | 0.47 | 0.00 | [-0.23; 0.23] | 12.1% | 7.1% |
| Fu 2020 | 108 | 1.84 | 1.47 | 109 | 2.13 | 1.55 | -0.19 | [-0.46; 0.08] | 9.0% | 6.5% |
| Ionescu 2020 | 106 | 1.84 | 1.54 | 109 | 1.95 | 1.60 | -0.07 | [-0.34; 0.20] | 8.9% | 6.5% |
| Ochs-Ross 2020 | 70 | 0.16 | 0.40 | 66 | 0.12 | 0.33 | 0.11 | [-0.23; 0.44] | 5.6% | 5.5% |
| Popova 2019 | 113 | 0.16 | 0.54 | 109 | 0.12 | 0.35 | 0.09 | [-0.18; 0.35] | 9.2% | 6.5% |
| Singh 2016 II | 18 | 0.56 | 1.29 | 8 | 0.88 | 1.64 | -0.22 | [-1.06; 0.61] | 0.9% | 1.7% |
| Takahashi 2021 | 122 | 0.17 | 0.53 | 80 | 0.28 | 0.67 | -0.19 | [-0.47; 0.10] | 8.0% | 6.2% |
| Zhou 2024 | 27 | -2.50 | 2.20 | 24 | -1.60 | 2.20 | -0.40 | [-0.96; 0.15] | 2.1% | 3.2% |
| Common effect model | **988** | | | **823** | | | **-0.04** | **[-0.14; 0.05]** | **73.0%** | **--** |
| Random effects model | | | | | | | **-0.05** | **[-0.16; 0.07]** | **--** | **59.9%** |
| Heterogeneity: $I^2 = 0\%$, $\tau^2 = 0.0091$, $p = 0.79$ | | | | | | | | | | |
| drug = Ketamine | | | | | | | | | | |
| Abbar 2022 | 73 | 3.00 | 6.05 | 80 | 8.50 | 10.19 | -0.65 | [-0.97; -0.32] | 6.0% | 5.6% |
| Domany 2020 | 9 | 7.00 | 16.30 | 9 | 10.90 | 14.40 | -0.24 | [-1.17; 0.69] | 0.7% | 1.4% |
| Domany 2021 | 15 | 6.30 | 8.90 | 15 | 6.60 | 10.40 | -0.03 | [-0.75; 0.69] | 1.2% | 2.2% |
| Fineberg 2023 | 10 | 3.20 | 4.76 | 11 | 3.00 | 5.27 | 0.04 | [-0.82; 0.89] | 0.9% | 1.6% |
| George 2017 | 12 | 1.42 | 1.38 | 4 | 2.00 | 1.41 | -0.40 | [-1.54; 0.75] | 0.5% | 1.0% |
| Hu 2016 | 12 | 0.50 | 0.70 | 14 | 1.20 | 0.40 | -1.22 | [-2.06; -0.37] | 0.9% | 1.7% |
| Jones 2024 | 17 | 7.80 | 7.36 | 11 | 10.50 | 9.65 | -0.32 | [-1.08; 0.45] | 1.1% | 2.0% |
| Lii 2023 | 18 | 0.32 | 0.95 | 20 | 0.35 | 0.59 | -0.04 | [-0.67; 0.60] | 1.6% | 2.6% |
| Loo 2016 | 9 | 1.78 | 1.20 | 6 | 1.33 | 1.03 | 0.37 | [-0.67; 1.42] | 0.6% | 1.2% |
| Murrough 2013 | 26 | 1.15 | 1.29 | 26 | 1.42 | 1.42 | -0.20 | [-0.74; 0.35] | 2.1% | 3.2% |
| Murrough 2015 II | 12 | 9.33 | 9.07 | 12 | 12.92 | 10.20 | -0.36 | [-1.17; 0.45] | 1.0% | 1.8% |
| Pattanaseri 2024 | 11 | 1.18 | 1.60 | 9 | 1.22 | 1.30 | -0.03 | [-0.91; 0.86] | 0.8% | 1.6% |
| Singh 2016 2x doses | 18 | 0.00 | 0.00 | 16 | 0.25 | 0.77 | | | 0.0% | 0.0% |
| Singh 2016 3x doses | 17 | 0.12 | 0.49 | 16 | 0.31 | 0.60 | -0.34 | [-1.03; 0.35] | 1.3% | 2.3% |
| Su 2017 | 47 | 0.72 | 0.74 | 24 | 1.17 | 0.82 | -0.58 | [-1.08; -0.08] | 2.5% | 3.6% |
| Su 2023 | 42 | 1.62 | 1.50 | 42 | 2.12 | 1.21 | -0.36 | [-0.79; 0.07] | 3.4% | 4.3% |
| Zarate 2006 | 9 | 0.78 | 1.09 | 9 | 0.67 | 0.87 | 0.11 | [-0.82; 1.03] | 0.7% | 1.4% |
| Zarate 2012 | 15 | 1.47 | 1.46 | 22 | 1.96 | 1.36 | -0.34 | [-1.00; 0.32] | 1.5% | 2.5% |
| Common effect model | **372** | | | **346** | | | **-0.37** | **[-0.52; -0.21]** | **27.0%** | **--** |
| Random effects model | | | | | | | **-0.32** | **[-0.52; -0.12]** | **--** | **40.1%** |
| Heterogeneity: $I^2 = 0\%$, $\tau^2 = 0.0511$, $p = 0.59$ | | | | | | | | | | |
| **Common effect model** | **1360** | | | **1169** | | | **-0.13** | **[-0.21; -0.05]** | **100.0%** | **--** |
| **Random effects model** | | | | | | | **-0.16** | **[-0.28; -0.04]** | **--** | **100.0%** |

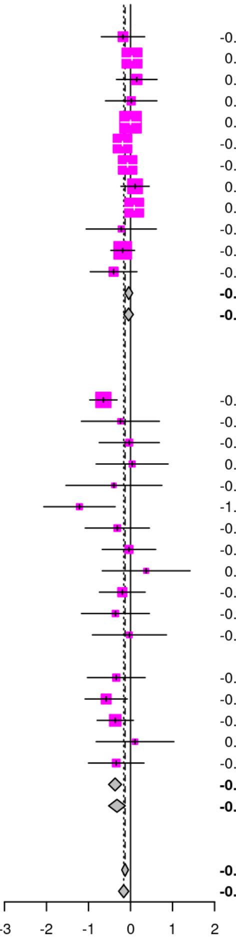

Heterogeneity: $I^2 = 17\%$, $\tau^2 = 0.0399$, $p = 0.21$
Test for subgroup differences (common effect): $\chi^2_1 = 12.54$, df = 1 ($p < 0.01$)
RE model$\chi^2_1 = 5.65$, df = 1 ($p = 0.02$)

## >72h up to 2w

| Study | Experimental Total | Experimental Mean | Experimental SD | Control Total | Control Mean | Control SD | SMD | 95%-CI (common) | Weight (common) | Weight (random) |
|---|---|---|---|---|---|---|---|---|---|---|
| **drug = Esketamine** | | | | | | | | | | |
| Canuso 2018 | 29 | 1.76 | 2.03 | 26 | 1.54 | 1.68 | 0.12 | [-0.41; 0.65] | 2.1% | 2.7% |
| Chen 2023 | 117 | 0.25 | 0.60 | 111 | 0.23 | 0.61 | 0.03 | [-0.23; 0.29] | 8.6% | 6.2% |
| Daly 2018 PA | 40 | 0.20 | 0.55 | 32 | 0.22 | 0.42 | -0.04 | [-0.50; 0.43] | 2.7% | 3.3% |
| Daly 2018 PB | 20 | 0.15 | 0.49 | 21 | 0.38 | 0.59 | -0.41 | [-1.03; 0.20] | 1.5% | 2.1% |
| Fedgchin 2019 | 211 | 0.16 | 0.46 | 110 | 0.18 | 0.39 | -0.05 | [-0.28; 0.18] | 10.9% | 6.7% |
| Fu 2020 | 105 | 1.28 | 1.31 | 99 | 1.33 | 1.14 | -0.04 | [-0.32; 0.23] | 7.7% | 5.9% |
| Ionescu 2020 | 92 | 0.97 | 1.19 | 100 | 1.35 | 1.34 | -0.30 | [-0.58; -0.01] | 7.1% | 5.7% |
| Ochs-Ross 2020 | 65 | 0.21 | 0.65 | 62 | 0.18 | 0.58 | 0.05 | [-0.30; 0.40] | 4.8% | 4.7% |
| Popova 2019 | 109 | 0.08 | 0.36 | 107 | 0.18 | 0.53 | -0.22 | [-0.49; 0.05] | 8.1% | 6.0% |
| Smith-Apeldoorn 2024 | 53 | 1.34 | 0.90 | 51 | 1.47 | 0.83 | -0.15 | [-0.53; 0.24] | 3.9% | 4.2% |
| Takahashi 2021 | 116 | 0.18 | 0.61 | 74 | 0.27 | 0.76 | -0.13 | [-0.43; 0.16] | 6.8% | 5.6% |
| Zhou 2024 | 27 | -2.80 | 2.10 | 23 | -2.20 | 2.50 | -0.26 | [-0.82; 0.30] | 1.9% | 2.5% |
| **Common effect model** | **984** | | | **816** | | | **-0.10** | **[-0.19; -0.01]** | **65.8%** | **--** |
| **Random effects model** | | | | | | | **-0.10** | **[-0.21; 0.01]** | **--** | **55.5%** |
| Heterogeneity: $I^2 = 0\%$, $\tau^2 = 0.0081$, $p = 0.81$ | | | | | | | | | | |
| **drug = Ketamine** | | | | | | | | | | |
| Abbar 2022 | 73 | 3.50 | 7.18 | 80 | 8.00 | 11.30 | -0.47 | [-0.79; -0.15] | 5.6% | 5.1% |
| Ahmed 2023 | 18 | 75.72 | 20.58 | 18 | 80.67 | 13.15 | -0.28 | [-0.94; 0.38] | 1.3% | 1.9% |
| Anderson 2017 | 35 | 2.94 | 1.76 | 37 | 3.08 | 2.38 | -0.07 | [-0.53; 0.40] | 2.7% | 3.3% |
| Burger 2016 | 3 | 2.40 | 13.61 | 7 | 6.70 | 8.93 | -0.38 | [-1.75; 0.99] | 0.3% | 0.5% |
| Colla 2024 | 17 | 1.18 | 0.87 | 10 | 1.60 | 0.95 | -0.45 | [-1.24; 0.34] | 0.9% | 1.4% |
| Domany 2020 | 9 | 7.40 | 18.00 | 9 | 1.40 | 2.20 | 0.45 | [-0.49; 1.38] | 0.7% | 1.0% |
| Domany 2021 | 15 | 3.10 | 14.10 | 15 | 11.40 | 15.20 | -0.55 | [-1.28; 0.18] | 1.1% | 1.6% |
| Duek 2023 | 14 | 0.14 | 0.36 | 13 | 0.08 | 0.28 | 0.20 | [-0.56; 0.95] | 1.0% | 1.5% |
| Fineberg 2023 | 10 | 4.30 | 7.10 | 12 | 5.67 | 7.04 | -0.19 | [-1.03; 0.65] | 0.8% | 1.3% |
| George 2017 | 12 | 1.25 | 1.21 | 4 | 2.50 | 1.73 | -0.88 | [-2.07; 0.30] | 0.4% | 0.7% |
| Grabski 2022 | 41 | 0.10 | 0.30 | 44 | 0.00 | 0.00 | | | 0.0% | 0.0% |
| Hu 2016 | 12 | 0.90 | 0.90 | 14 | 1.00 | 0.50 | -0.14 | [-0.91; 0.64] | 1.0% | 1.5% |
| Ionescu 2019 | 13 | 1.30 | 1.40 | 13 | 1.50 | 1.40 | -0.14 | [-0.91; 0.63] | 1.0% | 1.5% |
| Lii 2023 | 19 | 0.32 | 0.58 | 20 | 0.40 | 0.77 | -0.11 | [-0.74; 0.51] | 1.5% | 2.1% |
| Loo 2016 | 9 | 2.00 | 1.66 | 5 | 1.60 | 0.89 | 0.26 | [-0.84; 1.36] | 0.5% | 0.8% |
| Murrough 2015 II | 12 | 7.92 | 8.73 | 12 | 10.08 | 9.39 | -0.23 | [-1.03; 0.57] | 0.9% | 1.4% |
| Pattanaseri 2024 | 11 | 1.64 | 1.75 | 9 | 1.44 | 1.33 | 0.12 | [-0.76; 1.00] | 0.7% | 1.2% |
| Price 2022 | 52 | 0.79 | 1.32 | 46 | 1.30 | 1.62 | -0.34 | [-0.74; 0.05] | 3.6% | 4.0% |
| Singh 2016 2x doses | 15 | 0.07 | 0.26 | 12 | 0.33 | 0.65 | -0.53 | [-1.31; 0.24] | 1.0% | 1.5% |
| Singh 2016 3x doses | 15 | 0.00 | 0.00 | 12 | 0.25 | 0.45 | | | 0.0% | 0.0% |
| Sos 2013 | 9 | 0.44 | 0.73 | 18 | 0.17 | 0.87 | 0.32 | [-0.49; 1.12] | 0.9% | 1.4% |
| Su 2017 | 47 | 0.98 | 0.87 | 24 | 1.29 | 0.81 | -0.36 | [-0.86; 0.13] | 2.4% | 3.0% |
| Su 2023 | 42 | 1.95 | 1.36 | 42 | 2.12 | 1.33 | -0.13 | [-0.55; 0.30] | 3.2% | 3.6% |
| Sumner 2019 | 17 | 0.94 | 0.97 | 16 | 1.63 | 0.96 | -0.70 | [-1.40; 0.01] | 1.2% | 1.7% |
| Zarate 2006 | 9 | 0.89 | 1.27 | 9 | 1.00 | 0.82 | -0.10 | [-1.02; 0.83] | 0.7% | 1.1% |
| Zarate 2012 | 11 | 2.09 | 1.38 | 17 | 2.12 | 1.05 | -0.02 | [-0.78; 0.74] | 1.0% | 1.5% |
| **Common effect model** | **540** | | | **518** | | | **-0.24** | **[-0.37; -0.11]** | **34.2%** | **--** |
| **Random effects model** | | | | | | | **-0.22** | **[-0.38; -0.07]** | **--** | **44.5%** |
| Heterogeneity: $I^2 = 0\%$, $\tau^2 = 0.0363$, $p = 0.90$ | | | | | | | | | | |
| **Common effect model** | **1524** | | | **1334** | | | **-0.15** | **[-0.23; -0.07]** | **100.0%** | **--** |
| **Random effects model** | | | | | | | **-0.16** | **[-0.26; -0.06]** | **--** | **100.0%** |

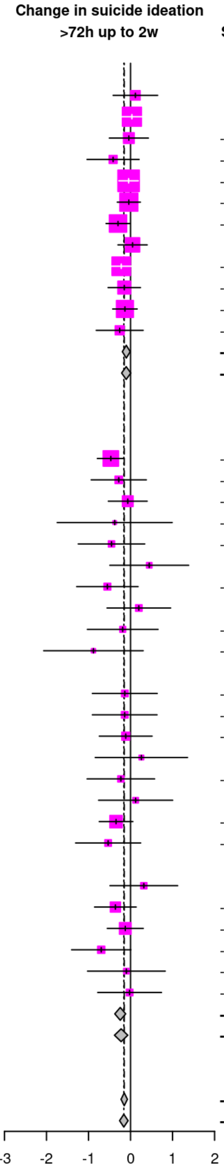

Heterogeneity: $I^2 = 0\%$, $\tau^2 = 0.0259$, $p = 0.90$
Test for subgroup differences (common effect): $\chi^2_1 = 3.10$, df = 1 ($p = 0.08$)
RE model$\chi^2_1 = 1.51$, df = 1 ($p = 0.22$)

## >2w up to 4w

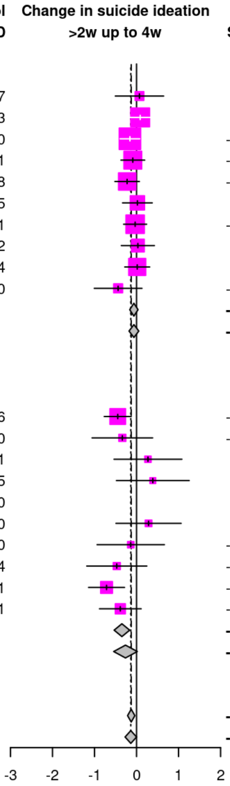

| Study | Experimental Total | Experimental Mean | Experimental SD | Control Total | Control Mean | Control SD | SMD | 95%-CI | Weight (common) | Weight (random) |
|---|---|---|---|---|---|---|---|---|---|---|
| drug = Esketamine | | | | | | | | | | |
| Canuso 2018 | 26 | 1.00 | 1.44 | 21 | 0.90 | 1.37 | 0.07 | [-0.51; 0.64] | 2.3% | 3.8% |
| Chen 2023 | 108 | 0.26 | 0.62 | 107 | 0.21 | 0.53 | 0.09 | [-0.18; 0.35] | 10.7% | 8.1% |
| Fedgchin 2019 | 204 | 0.11 | 0.37 | 107 | 0.17 | 0.40 | -0.16 | [-0.39; 0.08] | 13.9% | 8.7% |
| Fu 2020 | 100 | 0.97 | 1.27 | 93 | 1.08 | 1.21 | -0.09 | [-0.37; 0.19] | 9.6% | 7.8% |
| Ionescu 2020 | 90 | 0.68 | 1.04 | 92 | 0.93 | 1.18 | -0.22 | [-0.52; 0.07] | 9.0% | 7.7% |
| Ochs-Ross 2020 | 62 | 0.18 | 0.51 | 61 | 0.17 | 0.45 | 0.02 | [-0.33; 0.37] | 6.1% | 6.6% |
| Popova 2019 | 103 | 0.07 | 0.29 | 102 | 0.08 | 0.31 | -0.03 | [-0.31; 0.24] | 10.2% | 8.0% |
| Smith-Apeldoorn 2024 | 50 | 1.42 | 0.83 | 49 | 1.39 | 1.02 | 0.03 | [-0.36; 0.43] | 4.9% | 5.9% |
| Takahashi 2021 | 111 | 0.15 | 0.65 | 72 | 0.14 | 0.54 | 0.02 | [-0.28; 0.31] | 8.7% | 7.6% |
| Zhou 2024 | 26 | -2.90 | 2.20 | 23 | -1.90 | 2.30 | -0.44 | [-1.01; 0.13] | 2.4% | 3.9% |
| Common effect model | **880** | | | **727** | | | **-0.06** | **[-0.16; 0.04]** | **77.7%** | **--** |
| Random effects model | | | | | | | **-0.06** | **[-0.18; 0.05]** | **--** | **68.1%** |
| Heterogeneity: $I^2 = 0\%$, $\tau^2 = 0.0085$, $p = 0.77$ | | | | | | | | | | |
| drug = Ketamine | | | | | | | | | | |
| Abbar 2022 | 73 | 2.83 | 5.67 | 80 | 6.43 | 9.66 | -0.45 | [-0.77; -0.13] | 7.4% | 7.1% |
| Domany 2021 | 15 | 3.00 | 14.50 | 15 | 8.30 | 16.00 | -0.34 | [-1.06; 0.38] | 1.5% | 2.8% |
| Duek 2023 | 13 | 0.31 | 0.48 | 11 | 0.18 | 0.41 | 0.27 | [-0.54; 1.08] | 1.2% | 2.3% |
| Fineberg 2023 | 10 | 7.10 | 6.01 | 11 | 4.73 | 5.75 | 0.39 | [-0.48; 1.25] | 1.0% | 2.1% |
| Grabski 2022 | 39 | 0.05 | 0.22 | 43 | 0.00 | 0.00 | | | 0.0% | 0.0% |
| Hu 2016 | 12 | 0.40 | 0.90 | 14 | 0.20 | 0.40 | 0.29 | [-0.49; 1.06] | 1.3% | 2.5% |
| Ionescu 2019 | 12 | 1.20 | 1.40 | 12 | 1.40 | 1.40 | -0.14 | [-0.94; 0.66] | 1.2% | 2.3% |
| Ohtani 2024 | 15 | 1.40 | 1.08 | 16 | 1.94 | 1.14 | -0.47 | [-1.19; 0.25] | 1.5% | 2.8% |
| Price 2022 | 49 | 0.61 | 1.27 | 41 | 1.68 | 1.71 | -0.71 | [-1.14; -0.29] | 4.2% | 5.5% |
| Su 2017 | 47 | 1.00 | 0.75 | 24 | 1.33 | 1.01 | -0.39 | [-0.88; 0.11] | 3.1% | 4.6% |
| Common effect model | **285** | | | **267** | | | **-0.35** | **[-0.53; -0.16]** | **22.3%** | **--** |
| Random effects model | | | | | | | **-0.26** | **[-0.54; 0.02]** | **--** | **31.9%** |
| Heterogeneity: $I^2 = 28\%$, $\tau^2 = 0.0833$, $p = 0.19$ | | | | | | | | | | |
| **Common effect model** | **1165** | | | **994** | | | **-0.13** | **[-0.21; -0.04]** | **100.0%** | **--** |
| **Random effects model** | | | | | | | **-0.14** | **[-0.27; 0.00]** | **--** | **100.0%** |

Heterogeneity: $I^2 = 25\%$, $\tau^2 = 0.0414$, $p = 0.16$

Test for subgroup differences (common effect): $\chi^2_1 = 7.09$, df = 1 ($p < 0.01$)

RE model$\chi^2_1 = 1.66$, df = 1 ($p = 0.20$)

# >4w

| Study | Experimental Total | Experimental Mean | Experimental SD | Control Total | Control Mean | Control SD | Change in suicide ideation >4w | SMD | 95%-CI | Weight (common) | Weight (random) |
|---|---|---|---|---|---|---|---|---|---|---|---|
| **drug = Esketamine** | | | | | | | | | | | |
| Fu 2020 | 84 | 1.04 | 1.31 | 79 | 0.84 | 1.20 | | 0.16 | [-0.15; 0.47] | 20.7% | 17.3% |
| Ionescu 2020 | 80 | 0.49 | 1.08 | 86 | 0.66 | 0.98 | | -0.16 | [-0.47; 0.14] | 21.0% | 17.5% |
| Smith-Apeldoorn 2024 | 48 | 1.67 | 0.75 | 46 | 1.35 | 0.82 | | 0.40 | [-0.01; 0.81] | 11.7% | 12.5% |
| Zhou 2024 | 26 | -2.80 | 2.10 | 23 | -2.10 | 2.30 | | -0.31 | [-0.88; 0.25] | 6.1% | 7.9% |
| **Common effect model** | **238** | | | **234** | | | | **0.04** | **[-0.14; 0.23]** | **59.5%** | **--** |
| **Random effects model** | | | | | | | | **0.04** | **[-0.27; 0.35]** | **--** | **55.2%** |
| Heterogeneity: $I^2$ = 56%, $\tau^2$ = 0.0625, $p$ = 0.08 | | | | | | | | | | | |
| **drug = Ketamine** | | | | | | | | | | | |
| Abbar 2022 | 60 | 2.60 | 5.90 | 66 | 4.00 | 8.30 | | -0.19 | [-0.54; 0.16] | 15.9% | 15.1% |
| Duek 2023 | 11 | 0.30 | 0.48 | 11 | 0.20 | 0.42 | | 0.21 | [-0.63; 1.05] | 2.8% | 4.1% |
| Grabski 2022 | 38 | 0.05 | 0.23 | 43 | 0.07 | 0.26 | | -0.07 | [-0.51; 0.37] | 10.3% | 11.5% |
| Pattanaseri 2024 | 11 | 1.64 | 1.50 | 9 | 1.89 | 1.27 | | -0.17 | [-1.05; 0.71] | 2.5% | 3.7% |
| Price 2022 | 36 | 0.56 | 0.97 | 35 | 0.60 | 0.85 | | -0.04 | [-0.51; 0.42] | 9.0% | 10.5% |
| **Common effect model** | **156** | | | **164** | | | | **-0.10** | **[-0.32; 0.12]** | **40.5%** | **--** |
| **Random effects model** | | | | | | | | **-0.10** | **[-0.32; 0.13]** | **--** | **44.8%** |
| Heterogeneity: $I^2$ = 0%, $\tau^2$ = 0.0037, $p$ = 0.93 | | | | | | | | | | | |
| **Common effect model** | **394** | | | **398** | | | | **-0.01** | **[-0.15; 0.13]** | **100.0%** | **--** |
| **Random effects model** | | | | | | | | **-0.01** | **[-0.20; 0.17]** | **--** | **100.0%** |

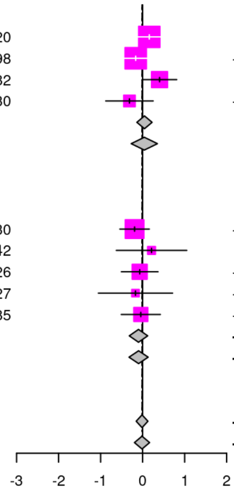

Heterogeneity: $I^2$ = 8%, $\tau^2$ = 0.0242, $p$ = 0.37
Test for subgroup differences (common effect): $\chi^2_1$ = 0.96, df = 1 ($p$ = 0.33)
RE model$\chi^2_1$ = 0.48, df = 1 ($p$ = 0.49)

#### End of Treatment

| Study | Experimental Total | Experimental Mean | Experimental SD | Control Total | Control Mean | Control SD | SMD | 95%-CI | Weight (common) | Weight (random) |
|---|---|---|---|---|---|---|---|---|---|---|
| **drug = Esketamine** | | | | | | | | | | |
| Canuso 2018 (25d) | 26 | 1.00 | 1.44 | 21 | 0.90 | 1.37 | 0.07 | [-0.51; 0.64] | 1.7% | 2.5% |
| Chen 2023 (4w) | 108 | 0.26 | 0.62 | 107 | 0.21 | 0.53 | 0.09 | [-0.18; 0.35] | 8.0% | 5.0% |
| Daly 2018 PA (7d) | 40 | 0.20 | 0.55 | 32 | 0.22 | 0.42 | -0.04 | [-0.50; 0.43] | 2.6% | 3.2% |
| Daly 2018 PB (7d) | 20 | 0.15 | 0.49 | 21 | 0.38 | 0.59 | -0.41 | [-1.03; 0.20] | 1.5% | 2.3% |
| Fedgchin 2019 (4w) | 204 | 0.11 | 0.37 | 107 | 0.17 | 0.40 | -0.16 | [-0.39; 0.08] | 10.4% | 5.3% |
| Fu 2020 (25d) | 100 | 0.97 | 1.27 | 93 | 1.08 | 1.21 | -0.09 | [-0.37; 0.19] | 7.1% | 4.8% |
| Ionescu 2020 (25d) | 90 | 0.68 | 1.04 | 92 | 0.93 | 1.18 | -0.22 | [-0.52; 0.07] | 6.7% | 4.8% |
| Ochs-Ross 2020 (4w) | 62 | 0.18 | 0.51 | 61 | 0.17 | 0.45 | 0.02 | [-0.33; 0.37] | 4.6% | 4.1% |
| Popova 2019 (4w) | 103 | 0.07 | 0.29 | 102 | 0.08 | 0.31 | -0.03 | [-0.31; 0.24] | 7.6% | 4.9% |
| Singh 2016 II (4d) | 18 | 0.56 | 1.29 | 8 | 0.88 | 1.64 | -0.22 | [-1.06; 0.61] | 0.8% | 1.5% |
| Smith-Apeldoorn 2024 (6w) | 50 | 1.60 | 0.83 | 48 | 1.42 | 0.85 | 0.21 | [-0.18; 0.61] | 3.6% | 3.7% |
| Takahashi 2021 (4w) | 111 | 0.15 | 0.65 | 72 | 0.14 | 0.54 | 0.02 | [-0.28; 0.31] | 6.5% | 4.7% |
| Zhou 2024 (6d) | 27 | -2.60 | 2.00 | 25 | -1.70 | 2.20 | -0.42 | [-0.97; 0.13] | 1.9% | 2.6% |
| **Common effect model** | **959** | | | **789** | | | **-0.06** | **[-0.16; 0.03]** | **63.0%** | **--** |
| **Random effects model** | | | | | | | **-0.06** | **[-0.18; 0.05]** | **--** | **49.5%** |
| Heterogeneity: $I^2 = 0\%$, $\tau^2 = 0.0130$, $p = 0.73$ | | | | | | | | | | |
| **drug = Ketamine** | | | | | | | | | | |
| Abbar 2022 (day 2) | 73 | 3.00 | 6.05 | 80 | 8.50 | 10.19 | -0.65 | [-0.97; -0.32] | 5.4% | 4.4% |
| Ahmed 2023 (2w) | 18 | 75.72 | 20.58 | 18 | 80.67 | 13.15 | -0.28 | [-0.94; 0.38] | 1.3% | 2.1% |
| Colla 2024 (2w) | 17 | 1.18 | 0.87 | 10 | 1.60 | 0.95 | -0.45 | [-1.24; 0.34] | 0.9% | 1.6% |
| Domany 2020 (24h) | 9 | 9.20 | 14.80 | 9 | 11.90 | 16.40 | -0.16 | [-1.09; 0.76] | 0.7% | 1.2% |
| Domany 2021 (24h) | 15 | 7.80 | 10.50 | 15 | 11.50 | 10.80 | -0.34 | [-1.06; 0.38] | 1.1% | 1.8% |
| Fineberg 2023 (24h) | 10 | 3.10 | 5.63 | 11 | 3.18 | 5.67 | -0.01 | [-0.87; 0.84] | 0.8% | 1.4% |
| George 2017 (1w) | 12 | 1.25 | 1.21 | 4 | 2.50 | 1.73 | -0.88 | [-2.07; 0.30] | 0.4% | 0.8% |
| Grabski 2022 (1-3w) | 38 | 0.05 | 0.23 | 43 | 0.07 | 0.26 | -0.07 | [-0.51; 0.37] | 3.0% | 3.4% |
| Grunebaum 2017 (24h) | 7 | 4.29 | 5.74 | 9 | 10.11 | 5.75 | -0.96 | [-2.02; 0.10] | 0.5% | 1.0% |
| Grunebaum 2018 (24h) | 40 | 6.05 | 5.48 | 40 | 11.63 | 7.43 | -0.85 | [-1.30; -0.39] | 2.7% | 3.3% |
| Hu 2016 (24h) | 13 | 0.70 | 0.70 | 14 | 1.30 | 0.40 | -1.03 | [-1.84; -0.22] | 0.9% | 1.5% |
| Ionescu 2019 (3w) | 12 | 1.20 | 1.40 | 12 | 1.40 | 1.40 | -0.14 | [-0.94; 0.66] | 0.9% | 1.6% |
| Jones 2024 (24h) | 17 | 10.80 | 8.08 | 10 | 12.30 | 7.83 | -0.18 | [-0.96; 0.60] | 0.9% | 1.6% |
| Lii 2023 (24h) | 20 | 0.20 | 0.52 | 19 | 0.21 | 0.54 | -0.02 | [-0.65; 0.61] | 1.4% | 2.2% |
| Loo 2016 (1w-2w) | 9 | 2.00 | 1.66 | 5 | 1.60 | 0.89 | 0.26 | [-0.84; 1.36] | 0.5% | 0.9% |
| Murrough 2013 (24h) | 26 | 0.85 | 1.12 | 26 | 1.46 | 1.33 | -0.49 | [-1.04; 0.06] | 1.9% | 2.6% |
| Murrough 2015 II (24h) | 12 | 10.83 | 8.48 | 12 | 14.00 | 10.15 | -0.33 | [-1.13; 0.48] | 0.9% | 1.5% |
| Ohtani 2024 (2w) | 15 | 1.40 | 1.08 | 16 | 1.94 | 1.14 | -0.47 | [-1.19; 0.25] | 1.1% | 1.8% |
| Pattanaseri 2024 (3d) | 11 | 1.18 | 1.60 | 9 | 1.22 | 1.30 | -0.03 | [-0.91; 0.86] | 0.7% | 1.3% |
| Price 2022 (24h) | 53 | 0.38 | 0.98 | 49 | 0.59 | 1.31 | -0.18 | [-0.57; 0.21] | 3.8% | 3.8% |
| Singh 2016 2x doses (2w) | 15 | 0.07 | 0.26 | 12 | 0.33 | 0.65 | -0.53 | [-1.31; 0.24] | 1.0% | 1.6% |
| Singh 2016 3x doses (2w) | 15 | 0.00 | 0.00 | 12 | 0.25 | 0.45 | | | 0.0% | 0.0% |
| Sos 2013 (24h) | 9 | 0.56 | 0.73 | . | 0.39 | 0.59 | | | 0.0% | 0.0% |
| Su 2023 (24h) | 42 | 1.48 | 1.42 | 42 | 2.10 | 1.21 | -0.47 | [-0.90; -0.03] | 3.0% | 3.4% |
| Sumner 2019 (24h) | 17 | 0.65 | 0.79 | 16 | 1.31 | 0.95 | -0.74 | [-1.45; -0.03] | 1.1% | 1.9% |
| Zarate 2006 (24h) | 9 | 0.44 | 0.53 | 9 | 0.78 | 0.83 | -0.46 | [-1.40; 0.48] | 0.6% | 1.2% |
| Zarate 2012 (24h) | 20 | 1.30 | 1.38 | 23 | 2.00 | 1.24 | -0.53 | [-1.14; 0.09] | 1.5% | 2.3% |
| **Common effect model** | **554** | | | **525** | | | **-0.42** | **[-0.54; -0.29]** | **37.0%** | **--** |
| **Random effects model** | | | | | | | **-0.41** | **[-0.56; -0.25]** | **--** | **50.5%** |
| Heterogeneity: $I^2 = 0\%$, $\tau^2 = 0.0417$, $p = 0.70$ | | | | | | | | | | |
| **Common effect model** | **1513** | | | **1314** | | | **-0.19** | **[-0.27; -0.12]** | **100.0%** | **--** |
| **Random effects model** | | | | | | | **-0.24** | **[-0.35; -0.13]** | **--** | **100.0%** |

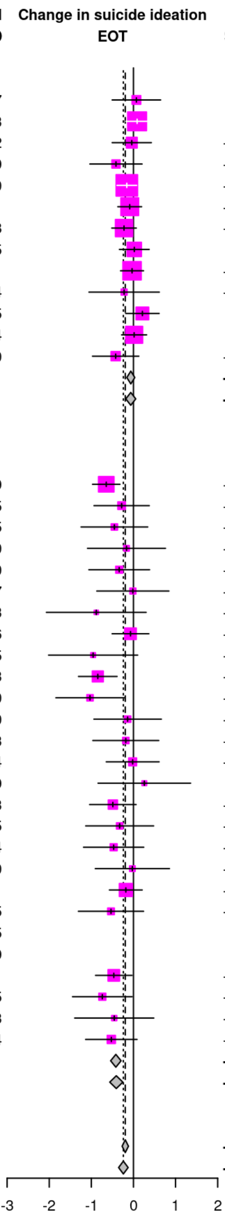

Heterogeneity: $I^2 = 24\%$, $\tau^2 = 0.0484$, $p = 0.10$
Test for subgroup differences (common effect): $\chi^2_1 = 19.85$, df = 1 ($p < 0.01$)
RE model $\chi^2_1 = 11.88$, df = 1 ($p < 0.01$)

#### Subgroup Analysis - Suicide as inclusion/exclusion criterion

## Up to 4h

| Study | Experimental Total | Experimental Mean | Experimental SD | Control Total | Control Mean | Control SD | SMD | 95%-CI | Weight (common) | Weight (random) |
|---|---|---|---|---|---|---|---|---|---|---|
| suic_excl_incl = excl/unknown | | | | | | | | | | |
| George 2017 | 12 | 2.17 | 1.53 | 4 | 1.75 | 1.50 | 0.26 | [-0.88; 1.40] | 1.0% | 2.7% |
| Glue 2024b | 17 | 0.41 | 0.71 | 8 | 0.25 | 0.46 | 0.24 | [-0.60; 1.08] | 1.8% | 3.8% |
| Loo 2016 | 9 | 1.44 | 1.31 | 6 | 0.83 | 0.75 | 0.51 | [-0.55; 1.56] | 1.2% | 3.0% |
| Singh 2016 2x doses | 18 | 0.11 | 0.32 | 16 | 0.75 | 1.18 | -0.74 | [-1.44; -0.04] | 2.6% | 4.4% |
| Singh 2016 3x doses | 17 | 0.12 | 0.49 | 16 | 0.50 | 1.03 | -0.46 | [-1.16; 0.23] | 2.7% | 4.5% |
| Sumner 2019 | 17 | 0.41 | 0.87 | 16 | 1.00 | 0.89 | -0.65 | [-1.36; 0.05] | 2.6% | 4.4% |
| Zarate 2006 | 9 | 0.12 | 0.35 | 9 | 0.56 | 0.73 | -0.72 | [-1.68; 0.24] | 1.4% | 3.3% |
| Zarate 2012 | 19 | 1.10 | 1.24 | 22 | 1.59 | 1.22 | -0.39 | [-1.01; 0.23] | 3.3% | 4.8% |
| Common effect model | **118** | | | **97** | | | **-0.36** | **[-0.63; -0.08]** | **16.6%** | **--** |
| Random effects model | | | | | | | **-0.32** | **[-0.69; 0.06]** | **--** | **30.8%** |
| Heterogeneity: $I^2$ = 14%, $\tau^2$ = 0.1253, $p$ = 0.32 | | | | | | | | | | |
| suic_excl_incl = incl | | | | | | | | | | |
| Abbar 2022 | 73 | 6.17 | 8.70 | 83 | 14.33 | 10.11 | -0.86 | [-1.19; -0.53] | 11.9% | 6.3% |
| Ahmed 2023 | 18 | 83.89 | 19.55 | 18 | 80.56 | 13.02 | 0.20 | [-0.46; 0.85] | 3.0% | 4.6% |
| Burger 2016 | 3 | 9.40 | 11.75 | 7 | 22.70 | 11.65 | -1.03 | [-2.50; 0.44] | 0.6% | 1.9% |
| Canuso 2018 | 35 | 4.26 | 2.09 | 31 | 4.81 | 1.70 | -0.28 | [-0.77; 0.20] | 5.4% | 5.5% |
| Domany 2020 | 9 | 13.10 | 14.20 | 9 | 12.30 | 14.70 | 0.05 | [-0.87; 0.98] | 1.5% | 3.4% |
| Domany 2021 | 15 | 8.60 | 9.30 | 15 | 13.60 | 8.60 | -0.54 | [-1.27; 0.19] | 2.4% | 4.3% |
| Fineberg 2023 | 10 | 4.40 | 5.23 | 12 | 3.42 | 5.71 | 0.17 | [-0.67; 1.01] | 1.8% | 3.8% |
| Fu 2020 | 110 | 2.49 | 1.65 | 112 | 2.74 | 1.44 | -0.16 | [-0.42; 0.10] | 18.5% | 6.6% |
| Grunebaum 2017 | 7 | 5.71 | 6.10 | 9 | 5.89 | 5.58 | -0.03 | [-1.02; 0.96] | 1.3% | 3.2% |
| Grunebaum 2018 | 40 | 5.00 | 5.18 | 40 | 9.85 | 7.21 | -0.77 | [-1.22; -0.31] | 6.2% | 5.7% |
| Hu 2016 | 13 | 0.30 | 0.50 | 14 | 1.30 | 0.40 | -2.15 | [-3.13; -1.18] | 1.4% | 3.2% |
| Ionescu 2019 | 13 | 1.90 | 1.40 | 13 | 1.50 | 1.40 | 0.28 | [-0.50; 1.05] | 2.2% | 4.1% |
| Ionescu 2020 | 113 | 2.25 | 1.47 | 112 | 2.92 | 1.32 | -0.48 | [-0.74; -0.21] | 18.3% | 6.6% |
| Jones 2024 | 17 | 8.50 | 7.99 | 11 | 14.20 | 8.63 | -0.67 | [-1.45; 0.11] | 2.1% | 4.0% |
| Su 2023 | 42 | 1.57 | 1.41 | 42 | 2.36 | 1.23 | -0.59 | [-1.03; -0.16] | 6.7% | 5.8% |
| Common effect model | **518** | | | **528** | | | **-0.44** | **[-0.57; -0.32]** | **83.4%** | **--** |
| Random effects model | | | | | | | **-0.44** | **[-0.74; -0.13]** | **--** | **69.2%** |
| Heterogeneity: $I^2$ = 62%, $\tau^2$ = 0.2470, $p$ < 0.01 | | | | | | | | | | |
| **Common effect model** | **636** | | | **625** | | | **-0.43** | **[-0.54; -0.31]** | **100.0%** | **--** |
| **Random effects model** | | | | | | | **-0.40** | **[-0.63; -0.16]** | **--** | **100.0%** |

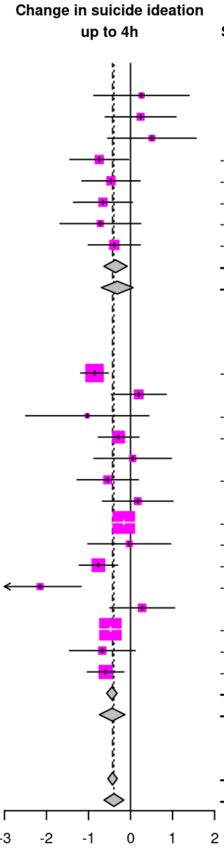

Heterogeneity: $I^2$ = 52%, $\tau^2$ = 0.2017, $p$ < 0.01
Test for subgroup differences (common effect): $\chi^2_1$ = 0.31, df = 1 ($p$ = 0.58)
RE model$\chi^2_1$ = 0.24, df = 1 ($p$ = 0.63)

## >12h up to 24h

| Study | Experimental Total | Experimental Mean | Experimental SD | Control Total | Control Mean | Control SD | SMD | 95%-CI | Weight (common) | Weight (random) |
|---|---|---|---|---|---|---|---|---|---|---|
| **suic_excl_incl = excl/unknown** | | | | | | | | | | |
| Lii 2023 | 20 | 0.20 | 0.52 | 19 | 0.21 | 0.54 | -0.02 | [-0.65; 0.61] | 2.8% | 3.9% |
| Loo 2016 | 9 | 1.78 | 1.20 | 6 | 1.33 | 1.03 | 0.37 | [-0.67; 1.42] | 1.0% | 1.8% |
| Murrough 2013 | 26 | 0.85 | 1.12 | 26 | 1.46 | 1.33 | -0.49 | [-1.04; 0.06] | 3.6% | 4.6% |
| Murrough 2015 II | 12 | 10.83 | 8.48 | 12 | 14.00 | 10.15 | -0.33 | [-1.13; 0.48] | 1.7% | 2.8% |
| Pattanaseri 2024 | 11 | 1.64 | 1.57 | 9 | 2.11 | 1.16 | -0.32 | [-1.21; 0.57] | 1.4% | 2.4% |
| Price 2022 | 53 | 0.38 | 0.98 | 49 | 0.59 | 1.31 | -0.18 | [-0.57; 0.21] | 7.3% | 6.5% |
| Ray-Griffith 2017 | 8 | 2.87 | 0.99 | 8 | 2.62 | 0.74 | 0.27 | [-0.72; 1.26] | 1.1% | 2.0% |
| Singh 2016 II | 18 | 0.61 | 1.42 | 8 | 0.88 | 1.64 | -0.18 | [-1.01; 0.66] | 1.6% | 2.6% |
| Sos 2013 | 9 | 0.56 | 0.73 | 18 | 0.39 | 0.59 | 0.26 | [-0.55; 1.06] | 1.7% | 2.8% |
| Sumner 2019 | 17 | 0.65 | 0.79 | 16 | 1.31 | 0.95 | -0.74 | [-1.45; -0.03] | 2.2% | 3.3% |
| Zarate 2006 | 9 | 0.44 | 0.53 | 9 | 0.78 | 0.83 | -0.46 | [-1.40; 0.48] | 1.3% | 2.2% |
| Zarate 2012 | 20 | 1.30 | 1.38 | 23 | 2.00 | 1.24 | -0.53 | [-1.14; 0.09] | 3.0% | 4.1% |
| Common effect model | **212** | | | **203** | | | **-0.25** | **[-0.44; -0.05]** | **28.7%** | **--** |
| Random effects model | | | | | | | **-0.24** | **[-0.48; 0.00]** | **--** | **39.0%** |
| Heterogeneity: $I^2 = 0\%$, $\tau^2 = 0.0487$, $p = 0.69$ | | | | | | | | | | |
| **suic_excl_incl = incl** | | | | | | | | | | |
| Abbar 2022 | 73 | 5.77 | 9.68 | 83 | 11.97 | 10.41 | -0.61 | [-0.93; -0.29] | 10.7% | 7.4% |
| Canuso 2018 | 35 | 3.29 | 2.47 | 30 | 4.47 | 1.57 | -0.55 | [-1.05; -0.06] | 4.5% | 5.2% |
| Domany 2020 | 9 | 9.20 | 14.80 | 9 | 11.90 | 16.40 | -0.16 | [-1.09; 0.76] | 1.3% | 2.2% |
| Domany 2021 | 15 | 7.80 | 10.50 | 15 | 11.50 | 10.80 | -0.34 | [-1.06; 0.38] | 2.1% | 3.3% |
| Fineberg 2023 | 10 | 3.10 | 5.63 | 11 | 3.18 | 5.67 | -0.01 | [-0.87; 0.84] | 1.5% | 2.5% |
| Fu 2020 | 110 | 2.23 | 1.67 | 112 | 2.43 | 1.63 | -0.12 | [-0.38; 0.14] | 16.0% | 8.3% |
| Grunebaum 2017 | 7 | 4.29 | 5.74 | 9 | 10.11 | 5.75 | -0.96 | [-2.02; 0.10] | 1.0% | 1.8% |
| Grunebaum 2018 | 40 | 6.05 | 5.48 | 40 | 11.63 | 7.43 | -0.85 | [-1.30; -0.39] | 5.3% | 5.6% |
| Hu 2016 | 13 | 0.70 | 0.70 | 14 | 1.30 | 0.40 | -1.03 | [-1.84; -0.22] | 1.7% | 2.7% |
| Ionescu 2020 | 110 | 2.22 | 1.51 | 110 | 2.44 | 1.57 | -0.14 | [-0.41; 0.12] | 15.8% | 8.3% |
| Jones 2024 | 17 | 10.80 | 8.08 | 10 | 12.30 | 7.83 | -0.18 | [-0.96; 0.60] | 1.8% | 2.9% |
| Su 2023 | 42 | 1.48 | 1.42 | 42 | 2.10 | 1.21 | -0.47 | [-0.90; -0.03] | 5.9% | 5.9% |
| Zhou 2024 | 27 | -2.40 | 2.20 | 27 | -1.50 | 2.00 | -0.42 | [-0.96; 0.12] | 3.8% | 4.7% |
| Common effect model | **508** | | | **512** | | | **-0.36** | **[-0.49; -0.24]** | **71.3%** | **--** |
| Random effects model | | | | | | | **-0.42** | **[-0.61; -0.23]** | **--** | **61.0%** |
| Heterogeneity: $I^2 = 34\%$, $\tau^2 = 0.0519$, $p = 0.11$ | | | | | | | | | | |
| **Common effect model** | **720** | | | **715** | | | **-0.33** | **[-0.44; -0.23]** | **100.0%** | **--** |
| **Random effects model** | | | | | | | **-0.35** | **[-0.51; -0.19]** | **--** | **100.0%** |

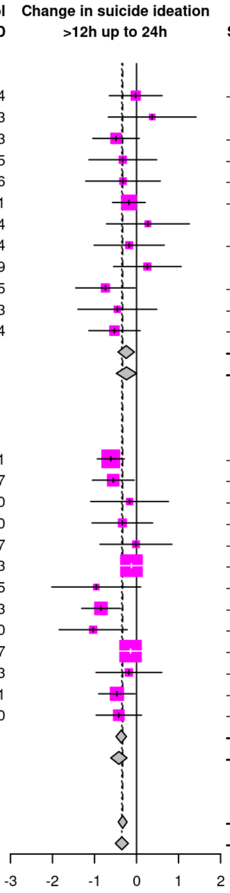

Heterogeneity: $I^2 = 12\%$, $\tau^2 = 0.0573$, $p = 0.29$
Test for subgroup differences (common effect): $\chi^2_1 = 0.99$, df = 1 ($p = 0.32$)
RE model$\chi^2_1 = 1.30$, df = 1 ($p = 0.25$)

## >24h up to 72h

| Study | Experimental Total | Experimental Mean | Experimental SD | Control Total | Control Mean | Control SD | SMD | 95%-CI | Weight (common) | Weight (random) |
|---|---|---|---|---|---|---|---|---|---|---|
| suic_excl_incl = excl/unknown | | | | | | | | | | |
| Chen 2023 | 123 | 0.25 | 0.61 | 126 | 0.23 | 0.54 | 0.03 | [-0.21; 0.28] | 10.3% | 6.8% |
| Daly 2018 PA | 33 | 0.18 | 0.46 | 33 | 0.12 | 0.33 | 0.15 | [-0.34; 0.63] | 2.7% | 3.8% |
| Daly 2018 PB | 20 | 0.25 | 0.72 | 21 | 0.24 | 0.54 | 0.02 | [-0.60; 0.63] | 1.7% | 2.8% |
| Fedgchin 2019 | 218 | 0.18 | 0.55 | 110 | 0.18 | 0.47 | 0.00 | [-0.23; 0.23] | 12.1% | 7.1% |
| George 2017 | 12 | 1.42 | 1.38 | 4 | 2.00 | 1.41 | -0.40 | [-1.54; 0.75] | 0.5% | 1.0% |
| Lii 2023 | 18 | 0.32 | 0.95 | 20 | 0.35 | 0.59 | -0.04 | [-0.67; 0.60] | 1.6% | 2.6% |
| Loo 2016 | 9 | 1.78 | 1.20 | 6 | 1.33 | 1.03 | 0.37 | [-0.67; 1.42] | 0.6% | 1.2% |
| Murrough 2013 | 26 | 1.15 | 1.29 | 26 | 1.42 | 1.42 | -0.20 | [-0.74; 0.35] | 2.1% | 3.2% |
| Murrough 2015 II | 12 | 9.33 | 9.07 | 12 | 12.92 | 10.20 | -0.36 | [-1.17; 0.45] | 1.0% | 1.8% |
| Ochs-Ross 2020 | 70 | 0.16 | 0.40 | 66 | 0.12 | 0.33 | 0.11 | [-0.23; 0.44] | 5.6% | 5.5% |
| Pattanaseri 2024 | 11 | 1.18 | 1.60 | 9 | 1.22 | 1.30 | -0.03 | [-0.91; 0.86] | 0.8% | 1.6% |
| Popova 2019 | 113 | 0.16 | 0.54 | 109 | 0.12 | 0.35 | 0.09 | [-0.18; 0.35] | 9.2% | 6.5% |
| Singh 2016 2x doses | 18 | 0.00 | 0.00 | 16 | 0.25 | 0.77 | | | 0.0% | 0.0% |
| Singh 2016 3x doses | 17 | 0.12 | 0.49 | 16 | 0.31 | 0.60 | -0.34 | [-1.03; 0.35] | 1.3% | 2.3% |
| Singh 2016 II | 18 | 0.56 | 1.29 | 8 | 0.88 | 1.64 | -0.22 | [-1.06; 0.61] | 0.9% | 1.7% |
| Su 2017 | 47 | 0.72 | 0.74 | 24 | 1.17 | 0.82 | -0.58 | [-1.08; -0.08] | 2.5% | 3.6% |
| Takahashi 2021 | 122 | 0.17 | 0.53 | 80 | 0.28 | 0.67 | -0.19 | [-0.47; 0.10] | 8.0% | 6.2% |
| Zarate 2006 | 9 | 0.78 | 1.09 | 9 | 0.67 | 0.87 | 0.11 | [-0.82; 1.03] | 0.7% | 1.4% |
| Zarate 2012 | 15 | 1.47 | 1.46 | 22 | 1.96 | 1.36 | -0.34 | [-1.00; 0.32] | 1.5% | 2.5% |
| Common effect model | **911** | | | **717** | | | **-0.04** | **[-0.14; 0.06]** | **63.4%** | **--** |
| Random effects model | | | | | | | **-0.06** | **[-0.19; 0.07]** | **--** | **61.6%** |
| Heterogeneity: $I^2$ = 0%, $\tau^2$ = 0.0198, $p$ = 0.81 | | | | | | | | | | |
| suic_excl_incl = incl | | | | | | | | | | |
| Abbar 2022 | 73 | 3.00 | 6.05 | 80 | 8.50 | 10.19 | -0.65 | [-0.97; -0.32] | 6.0% | 5.6% |
| Canuso 2018 | 30 | 2.87 | 2.61 | 28 | 3.32 | 2.33 | -0.18 | [-0.70; 0.34] | 2.4% | 3.5% |
| Domany 2020 | 9 | 7.00 | 16.30 | 9 | 10.90 | 14.40 | -0.24 | [-1.17; 0.69] | 0.7% | 1.4% |
| Domany 2021 | 15 | 6.30 | 8.90 | 15 | 6.60 | 10.40 | -0.03 | [-0.75; 0.69] | 1.2% | 2.2% |
| Fineberg 2023 | 10 | 3.20 | 4.76 | 11 | 3.00 | 5.27 | 0.04 | [-0.82; 0.89] | 0.9% | 1.6% |
| Fu 2020 | 108 | 1.84 | 1.47 | 109 | 2.13 | 1.55 | -0.19 | [-0.46; 0.08] | 9.0% | 6.5% |
| Hu 2016 | 12 | 0.50 | 0.70 | 14 | 1.20 | 0.40 | -1.22 | [-2.06; -0.37] | 0.9% | 1.7% |
| Ionescu 2020 | 106 | 1.84 | 1.54 | 109 | 1.95 | 1.60 | -0.07 | [-0.34; 0.20] | 8.9% | 6.5% |
| Jones 2024 | 17 | 7.80 | 7.36 | 11 | 10.50 | 9.65 | -0.32 | [-1.08; 0.45] | 1.1% | 2.0% |
| Su 2023 | 42 | 1.62 | 1.50 | 42 | 2.12 | 1.21 | -0.36 | [-0.79; 0.07] | 3.4% | 4.3% |
| Zhou 2024 | 27 | -2.50 | 2.20 | 24 | -1.60 | 2.20 | -0.40 | [-0.96; 0.15] | 2.1% | 3.2% |
| Common effect model | **449** | | | **452** | | | **-0.28** | **[-0.41; -0.15]** | **36.6%** | **--** |
| Random effects model | | | | | | | **-0.31** | **[-0.52; -0.10]** | **--** | **38.4%** |
| Heterogeneity: $I^2$ = 28%, $\tau^2$ = 0.0558, $p$ = 0.18 | | | | | | | | | | |
| **Common effect model** | **1360** | | | **1169** | | | **-0.13** | **[-0.21; -0.05]** | **100.0%** | **--** |
| **Random effects model** | | | | | | | **-0.16** | **[-0.28; -0.04]** | **--** | **100.0%** |

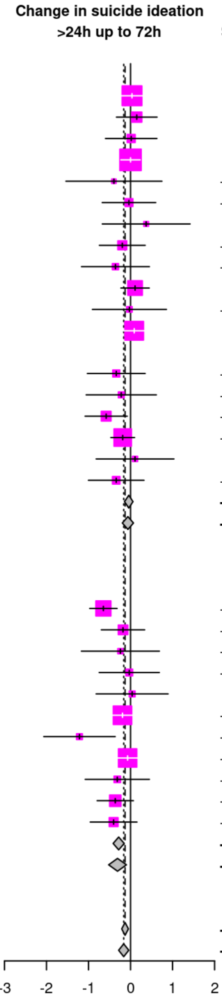

Heterogeneity: $I^2$ = 17%, $\tau^2$ = 0.0399, $p$ = 0.21
Test for subgroup differences (common effect): $\chi^2_1$ = 8.06, df = 1 ($p$ < 0.01)
RE model$\chi^2_1$ = 3.76, df = 1 ($p$ = 0.05)

## >72h up to 2w

| Study | Experimental Total | Experimental Mean | Experimental SD | Control Total | Control Mean | Control SD | SMD | 95%-CI | Weight (common) | Weight (random) |
|---|---|---|---|---|---|---|---|---|---|---|
| suic_excl_incl = excl/unknown | | | | | | | | | | |
| Anderson 2017 | 35 | 2.94 | 1.76 | 37 | 3.08 | 2.38 | -0.07 | [-0.53; 0.40] | 2.7% | 3.3% |
| Chen 2023 | 117 | 0.25 | 0.60 | 111 | 0.23 | 0.61 | 0.03 | [-0.23; 0.29] | 8.6% | 6.2% |
| Colla 2024 | 17 | 1.18 | 0.87 | 10 | 1.60 | 0.95 | -0.45 | [-1.24; 0.34] | 0.9% | 1.4% |
| Daly 2018 PA | 40 | 0.20 | 0.55 | 32 | 0.22 | 0.42 | -0.04 | [-0.50; 0.43] | 2.7% | 3.3% |
| Daly 2018 PB | 20 | 0.15 | 0.49 | 21 | 0.38 | 0.59 | -0.41 | [-1.03; 0.20] | 1.5% | 2.1% |
| Duek 2023 | 14 | 0.14 | 0.36 | 13 | 0.08 | 0.28 | 0.20 | [-0.56; 0.95] | 1.0% | 1.5% |
| Fedgchin 2019 | 211 | 0.16 | 0.46 | 110 | 0.18 | 0.39 | -0.05 | [-0.28; 0.18] | 10.9% | 6.7% |
| George 2017 | 12 | 1.25 | 1.21 | 4 | 2.50 | 1.73 | -0.88 | [-2.07; 0.30] | 0.4% | 0.7% |
| Grabski 2022 | 41 | 0.10 | 0.30 | 44 | 0.00 | 0.00 | | | 0.0% | 0.0% |
| Lii 2023 | 19 | 0.32 | 0.58 | 20 | 0.40 | 0.77 | -0.11 | [-0.74; 0.51] | 1.5% | 2.1% |
| Loo 2016 | 9 | 2.00 | 1.66 | 5 | 1.60 | 0.89 | 0.26 | [-0.84; 1.36] | 0.5% | 0.8% |
| Murrough 2015 II | 12 | 7.92 | 8.73 | 12 | 10.08 | 9.39 | -0.23 | [-1.03; 0.57] | 0.9% | 1.4% |
| Ochs-Ross 2020 | 65 | 0.21 | 0.65 | 62 | 0.18 | 0.58 | 0.05 | [-0.30; 0.40] | 4.8% | 4.7% |
| Pattanaseri 2024 | 11 | 1.64 | 1.75 | 9 | 1.44 | 1.33 | 0.12 | [-0.76; 1.00] | 0.7% | 1.2% |
| Popova 2019 | 109 | 0.08 | 0.36 | 107 | 0.18 | 0.53 | -0.22 | [-0.49; 0.05] | 8.1% | 6.0% |
| Price 2022 | 52 | 0.79 | 1.32 | 46 | 1.30 | 1.62 | -0.34 | [-0.74; 0.05] | 3.6% | 4.0% |
| Singh 2016 2x doses | 15 | 0.07 | 0.26 | 12 | 0.33 | 0.65 | -0.53 | [-1.31; 0.24] | 1.0% | 1.5% |
| Singh 2016 3x doses | 15 | 0.00 | 0.00 | 12 | 0.25 | 0.45 | | | 0.0% | 0.0% |
| Smith-Apeldoorn 2024 | 53 | 1.34 | 0.90 | 51 | 1.47 | 0.83 | -0.15 | [-0.53; 0.24] | 3.9% | 4.2% |
| Sos 2013 | 9 | 0.44 | 0.73 | 18 | 0.17 | 0.87 | 0.32 | [-0.49; 1.12] | 0.9% | 1.4% |
| Su 2017 | 47 | 0.98 | 0.87 | 24 | 1.29 | 0.81 | -0.36 | [-0.86; 0.13] | 2.4% | 3.0% |
| Sumner 2019 | 17 | 0.94 | 0.97 | 16 | 1.63 | 0.96 | -0.70 | [-1.40; 0.01] | 1.2% | 1.7% |
| Takahashi 2021 | 116 | 0.18 | 0.61 | 74 | 0.27 | 0.76 | -0.13 | [-0.43; 0.16] | 6.8% | 5.6% |
| Zarate 2006 | 9 | 0.89 | 1.27 | 9 | 1.00 | 0.82 | -0.10 | [-1.02; 0.83] | 0.7% | 1.1% |
| Zarate 2012 | 11 | 2.09 | 1.38 | 17 | 2.12 | 1.05 | -0.02 | [-0.78; 0.74] | 1.0% | 1.5% |
| Common effect model | **1076** | | | **876** | | | **-0.12** | **[-0.22; -0.03]** | **66.4%** | **--** |
| Random effects model | | | | | | | **-0.14** | **[-0.27; -0.01]** | **--** | **65.1%** |
| Heterogeneity: $I^2 = 0\%$, $\tau^2 = 0.0294$, $p = 0.86$ | | | | | | | | | | |
| suic_excl_incl = incl | | | | | | | | | | |
| Abbar 2022 | 73 | 3.50 | 7.18 | 80 | 8.00 | 11.30 | -0.47 | [-0.79; -0.15] | 5.6% | 5.1% |
| Ahmed 2023 | 18 | 75.72 | 20.58 | 18 | 80.67 | 13.15 | -0.28 | [-0.94; 0.38] | 1.3% | 1.9% |
| Burger 2016 | 3 | 2.40 | 13.61 | 7 | 6.70 | 8.93 | -0.38 | [-1.75; 0.99] | 0.3% | 0.5% |
| Canuso 2018 | 29 | 1.76 | 2.03 | 26 | 1.54 | 1.68 | 0.12 | [-0.41; 0.65] | 2.1% | 2.7% |
| Domany 2020 | 9 | 7.40 | 18.00 | 9 | 1.40 | 2.20 | 0.45 | [-0.49; 1.38] | 0.7% | 1.0% |
| Domany 2021 | 15 | 3.10 | 14.10 | 15 | 11.40 | 15.20 | -0.55 | [-1.28; 0.18] | 1.1% | 1.6% |
| Fineberg 2023 | 10 | 4.30 | 7.10 | 12 | 5.67 | 7.04 | -0.19 | [-1.03; 0.65] | 0.8% | 1.3% |
| Fu 2020 | 105 | 1.28 | 1.31 | 99 | 1.33 | 1.14 | -0.04 | [-0.32; 0.23] | 7.7% | 5.9% |
| Hu 2016 | 12 | 0.90 | 0.90 | 14 | 1.00 | 0.50 | -0.14 | [-0.91; 0.64] | 1.0% | 1.5% |
| Ionescu 2019 | 13 | 1.30 | 1.40 | 13 | 1.50 | 1.40 | -0.14 | [-0.91; 0.63] | 1.0% | 1.5% |
| Ionescu 2020 | 92 | 0.97 | 1.19 | 100 | 1.35 | 1.34 | -0.30 | [-0.58; -0.01] | 7.1% | 5.7% |
| Su 2023 | 42 | 1.95 | 1.36 | 42 | 2.12 | 1.33 | -0.13 | [-0.55; 0.30] | 3.2% | 3.6% |
| Zhou 2024 | 27 | -2.80 | 2.10 | 23 | -2.20 | 2.50 | -0.26 | [-0.82; 0.30] | 1.9% | 2.5% |
| Common effect model | **448** | | | **458** | | | **-0.21** | **[-0.34; -0.07]** | **33.6%** | **--** |
| Random effects model | | | | | | | **-0.20** | **[-0.37; -0.03]** | **--** | **34.9%** |
| Heterogeneity: $I^2 = 0\%$, $\tau^2 = 0.0216$, $p = 0.72$ | | | | | | | | | | |
| **Common effect model** | **1524** | | | **1334** | | | **-0.15** | **[-0.23; -0.07]** | **100.0%** | **--** |
| **Random effects model** | | | | | | | **-0.16** | **[-0.26; -0.06]** | **--** | **100.0%** |

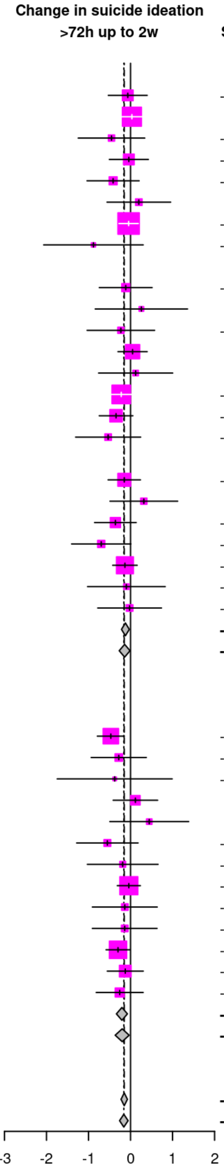

Heterogeneity: $I^2 = 0\%$, $\tau^2 = 0.0259$, $p = 0.90$
Test for subgroup differences (common effect): $\chi^2_1 = 1.02$, df = 1 ($p = 0.31$)
RE model$\chi^2_1 = 0.30$, df = 1 ($p = 0.59$)

## >2w up to 4w

| Study | Experimental Total | Mean | SD | Control Total | Mean | SD | SMD | 95%-CI | Weight (common) | Weight (random) |
|---|---|---|---|---|---|---|---|---|---|---|
| **suic_excl_incl = excl/unknown** | | | | | | | | | | |
| Chen 2023 | 108 | 0.26 | 0.62 | 107 | 0.21 | 0.53 | 0.09 | [-0.18; 0.35] | 10.7% | 8.1% |
| Duek 2023 | 13 | 0.31 | 0.48 | 11 | 0.18 | 0.41 | 0.27 | [-0.54; 1.08] | 1.2% | 2.3% |
| Fedgchin 2019 | 204 | 0.11 | 0.37 | 107 | 0.17 | 0.40 | -0.16 | [-0.39; 0.08] | 13.9% | 8.7% |
| Grabski 2022 | 39 | 0.05 | 0.22 | 43 | 0.00 | 0.00 | | | 0.0% | 0.0% |
| Ochs-Ross 2020 | 62 | 0.18 | 0.51 | 61 | 0.17 | 0.45 | 0.02 | [-0.33; 0.37] | 6.1% | 6.6% |
| Ohtani 2024 | 15 | 1.40 | 1.08 | 16 | 1.94 | 1.14 | -0.47 | [-1.19; 0.25] | 1.5% | 2.8% |
| Popova 2019 | 103 | 0.07 | 0.29 | 102 | 0.08 | 0.31 | -0.03 | [-0.31; 0.24] | 10.2% | 8.0% |
| Price 2022 | 49 | 0.61 | 1.27 | 41 | 1.68 | 1.71 | -0.71 | [-1.14; -0.29] | 4.2% | 5.5% |
| Smith-Apeldoorn 2024 | 50 | 1.42 | 0.83 | 49 | 1.39 | 1.02 | 0.03 | [-0.36; 0.43] | 4.9% | 5.9% |
| Su 2017 | 47 | 1.00 | 0.75 | 24 | 1.33 | 1.01 | -0.39 | [-0.88; 0.11] | 3.1% | 4.6% |
| Takahashi 2021 | 111 | 0.15 | 0.65 | 72 | 0.14 | 0.54 | 0.02 | [-0.28; 0.31] | 8.7% | 7.6% |
| **Common effect model** | **801** | | | **633** | | | **-0.09** | **[-0.20; 0.02]** | **64.4%** | **--** |
| **Random effects model** | | | | | | | **-0.12** | **[-0.30; 0.07]** | **--** | **60.1%** |
| Heterogeneity: $I^2$ = 39%, $\tau^2$ = 0.0492, $p$ = 0.10 | | | | | | | | | | |
| **suic_excl_incl = incl** | | | | | | | | | | |
| Abbar 2022 | 73 | 2.83 | 5.67 | 80 | 6.43 | 9.66 | -0.45 | [-0.77; -0.13] | 7.4% | 7.1% |
| Canuso 2018 | 26 | 1.00 | 1.44 | 21 | 0.90 | 1.37 | 0.07 | [-0.51; 0.64] | 2.3% | 3.8% |
| Domany 2021 | 15 | 3.00 | 14.50 | 15 | 8.30 | 16.00 | -0.34 | [-1.06; 0.38] | 1.5% | 2.8% |
| Fineberg 2023 | 10 | 7.10 | 6.01 | 11 | 4.73 | 5.75 | 0.39 | [-0.48; 1.25] | 1.0% | 2.1% |
| Fu 2020 | 100 | 0.97 | 1.27 | 93 | 1.08 | 1.21 | -0.09 | [-0.37; 0.19] | 9.6% | 7.8% |
| Hu 2016 | 12 | 0.40 | 0.90 | 14 | 0.20 | 0.40 | 0.29 | [-0.49; 1.06] | 1.3% | 2.5% |
| Ionescu 2019 | 12 | 1.20 | 1.40 | 12 | 1.40 | 1.40 | -0.14 | [-0.94; 0.66] | 1.2% | 2.3% |
| Ionescu 2020 | 90 | 0.68 | 1.04 | 92 | 0.93 | 1.18 | -0.22 | [-0.52; 0.07] | 9.0% | 7.7% |
| Zhou 2024 | 26 | -2.90 | 2.20 | 23 | -1.90 | 2.30 | -0.44 | [-1.01; 0.13] | 2.4% | 3.9% |
| **Common effect model** | **364** | | | **361** | | | **-0.20** | **[-0.34; -0.05]** | **35.6%** | **--** |
| **Random effects model** | | | | | | | **-0.17** | **[-0.38; 0.04]** | **--** | **39.9%** |
| Heterogeneity: $I^2$ = 0%, $\tau^2$ = 0.0374, $p$ = 0.45 | | | | | | | | | | |
| **Common effect model** | **1165** | | | **994** | | | **-0.13** | **[-0.21; -0.04]** | **100.0%** | **--** |
| **Random effects model** | | | | | | | **-0.14** | **[-0.27; 0.00]** | **--** | **100.0%** |

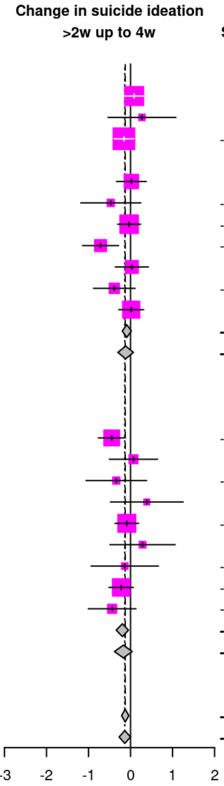

Heterogeneity: $I^2$ = 25%, $\tau^2$ = 0.0414, $p$ = 0.16

Test for subgroup differences (common effect): $\chi^2_1$ = 1.30, df = 1 ($p$ = 0.25)

RE model$\chi^2_1$ = 0.15, df = 1 ($p$ = 0.70)

## >4w

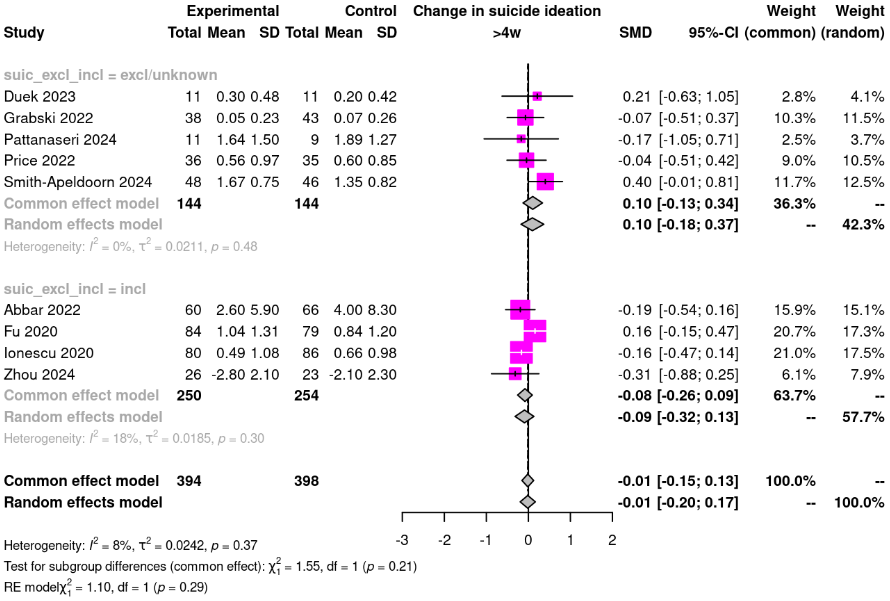

Experimental Control Change in suicide ideation Weight Weight
Study Total Mean SD Total Mean SD >4w SMD 95%-CI (common) (random)
suic_excl_incl = excl/unknown
Duek 2023 11 0.30 0.48 11 0.20 0.42 0.21 [-0.63; 1.05] 2.8% 4.1%
Grabski 2022 38 0.05 0.23 43 0.07 0.26 -0.07 [-0.51; 0.37] 10.3% 11.5%
Pattanaseri 2024 11 1.64 1.50 9 1.89 1.27 -0.17 [-1.05; 0.71] 2.5% 3.7%
Price 2022 36 0.56 0.97 35 0.60 0.85 -0.04 [-0.51; 0.42] 9.0% 10.5%
Smith-Apeldoorn 2024 48 1.67 0.75 46 1.35 0.82 0.40 [-0.01; 0.81] 11.7% 12.5%
Common effect model 144 144 0.10 [-0.13; 0.34] 36.3% --
Random effects model 0.10 [-0.18; 0.37] -- 42.3%
Heterogeneity: $I^2$ = 0%, $\tau^2$ = 0.0211, $p$ = 0.48
suic_excl_incl = incl
Abbar 2022 60 2.60 5.90 66 4.00 8.30 -0.19 [-0.54; 0.16] 15.9% 15.1%
Fu 2020 84 1.04 1.31 79 0.84 1.20 0.16 [-0.15; 0.47] 20.7% 17.3%
Ionescu 2020 80 0.49 1.08 86 0.66 0.98 -0.16 [-0.47; 0.14] 21.0% 17.5%
Zhou 2024 26 -2.80 2.10 23 -2.10 2.30 -0.31 [-0.88; 0.25] 6.1% 7.9%
Common effect model 250 254 -0.08 [-0.26; 0.09] 63.7% --
Random effects model -0.09 [-0.32; 0.13] -- 57.7%
Heterogeneity: $I^2$ = 18%, $\tau^2$ = 0.0185, $p$ = 0.30
Common effect model 394 398 -0.01 [-0.15; 0.13] 100.0% --
Random effects model -0.01 [-0.20; 0.17] -- 100.0%
-3 -2 -1 0 1 2
Heterogeneity: $I^2$ = 8%, $\tau^2$ = 0.0242, $p$ = 0.37
Test for subgroup differences (common effect): $\chi^2_1$ = 1.55, df = 1 ($p$ = 0.21)
RE model$\chi^2_1$ = 1.10, df = 1 ($p$ = 0.29)

#### End of Treatment

| Study | Experimental Total | Experimental Mean | Experimental SD | Control Total | Control Mean | Control SD | SMD | 95%-CI | Weight (common) | Weight (random) |
|---|---|---|---|---|---|---|---|---|---|---|
| **suic_excl_incl = excl/unknown** | | | | | | | | | | |
| Chen 2023 (4w) | 108 | 0.26 | 0.62 | 107 | 0.21 | 0.53 | 0.09 | [-0.18; 0.35] | 8.0% | 5.0% |
| Colla 2024 (2w) | 17 | 1.18 | 0.87 | 10 | 1.60 | 0.95 | -0.45 | [-1.24; 0.34] | 0.9% | 1.6% |
| Daly 2018 PA (7d) | 40 | 0.20 | 0.55 | 32 | 0.22 | 0.42 | -0.04 | [-0.50; 0.43] | 2.6% | 3.2% |
| Daly 2018 PB (7d) | 20 | 0.15 | 0.49 | 21 | 0.38 | 0.59 | -0.41 | [-1.03; 0.20] | 1.5% | 2.3% |
| Fedgchin 2019 (4w) | 204 | 0.11 | 0.37 | 107 | 0.17 | 0.40 | -0.16 | [-0.39; 0.08] | 10.4% | 5.3% |
| George 2017 (1w) | 12 | 1.25 | 1.21 | 4 | 2.50 | 1.73 | -0.88 | [-2.07; 0.30] | 0.4% | 0.8% |
| Grabski 2022 (1-3w) | 38 | 0.05 | 0.23 | 43 | 0.07 | 0.26 | -0.07 | [-0.51; 0.37] | 3.0% | 3.4% |
| Lii 2023 (24h) | 20 | 0.20 | 0.52 | 19 | 0.21 | 0.54 | -0.02 | [-0.65; 0.61] | 1.4% | 2.2% |
| Loo 2016 (1w-2w) | 9 | 2.00 | 1.66 | 5 | 1.60 | 0.89 | 0.26 | [-0.84; 1.36] | 0.5% | 0.9% |
| Murrough 2013 (24h) | 26 | 0.85 | 1.12 | 26 | 1.46 | 1.33 | -0.49 | [-1.04; 0.06] | 1.9% | 2.6% |
| Murrough 2015 II (24h) | 12 | 10.83 | 8.48 | 12 | 14.00 | 10.15 | -0.33 | [-1.13; 0.48] | 0.9% | 1.5% |
| Ochs-Ross 2020 (4w) | 62 | 0.18 | 0.51 | 61 | 0.17 | 0.45 | 0.02 | [-0.33; 0.37] | 4.6% | 4.1% |
| Ohtani 2024 (2w) | 15 | 1.40 | 1.08 | 16 | 1.94 | 1.14 | -0.47 | [-1.19; 0.25] | 1.1% | 1.8% |
| Pattanaseri 2024 (3d) | 11 | 1.18 | 1.60 | 9 | 1.22 | 1.30 | -0.03 | [-0.91; 0.86] | 0.7% | 1.3% |
| Popova 2019 (4w) | 103 | 0.07 | 0.29 | 102 | 0.08 | 0.31 | -0.03 | [-0.31; 0.24] | 7.6% | 4.9% |
| Price 2022 (24h) | 53 | 0.38 | 0.98 | 49 | 0.59 | 1.31 | -0.18 | [-0.57; 0.21] | 3.8% | 3.8% |
| Singh 2016 2x doses (2w) | 15 | 0.07 | 0.26 | 12 | 0.33 | 0.65 | -0.53 | [-1.31; 0.24] | 1.0% | 1.6% |
| Singh 2016 3x doses (2w) | 15 | 0.00 | 0.00 | 12 | 0.25 | 0.45 | | | 0.0% | 0.0% |
| Singh 2016 II (4d) | 18 | 0.56 | 1.29 | 8 | 0.88 | 1.64 | -0.22 | [-1.06; 0.61] | 0.8% | 1.5% |
| Smith-Apeldoorn 2024 (6w) | 50 | 1.60 | 0.83 | 48 | 1.42 | 0.85 | 0.21 | [-0.18; 0.61] | 3.6% | 3.7% |
| Sos 2013 (24h) | 9 | 0.56 | 0.73 | . | 0.39 | 0.59 | | | 0.0% | 0.0% |
| Sumner 2019 (24h) | 17 | 0.65 | 0.79 | 16 | 1.31 | 0.95 | -0.74 | [-1.45; -0.03] | 1.1% | 1.9% |
| Takahashi 2021 (4w) | 111 | 0.15 | 0.65 | 72 | 0.14 | 0.54 | 0.02 | [-0.28; 0.31] | 6.5% | 4.7% |
| Zarate 2006 (24h) | 9 | 0.44 | 0.53 | 9 | 0.78 | 0.83 | -0.46 | [-1.40; 0.48] | 0.6% | 1.2% |
| Zarate 2012 (24h) | 20 | 1.30 | 1.38 | 23 | 2.00 | 1.24 | -0.53 | [-1.14; 0.09] | 1.5% | 2.3% |
| **Common effect model** | **1014** | | | **823** | | | **-0.11** | **[-0.20; -0.01]** | **64.4%** | -- |
| **Random effects model** | | | | | | | **-0.15** | **[-0.29; -0.02]** | -- | **61.9%** |
| Heterogeneity: $I^2 = 0\%$, $\tau^2 = 0.0365$, $p = 0.59$ | | | | | | | | | | |
| **suic_excl_incl = incl** | | | | | | | | | | |
| Abbar 2022 (day 2) | 73 | 3.00 | 6.05 | 80 | 8.50 | 10.19 | -0.65 | [-0.97; -0.32] | 5.4% | 4.4% |
| Ahmed 2023 (2w) | 18 | 75.72 | 20.58 | 18 | 80.67 | 13.15 | -0.28 | [-0.94; 0.38] | 1.3% | 2.1% |
| Canuso 2018 (25d) | 26 | 1.00 | 1.44 | 21 | 0.90 | 1.37 | 0.07 | [-0.51; 0.64] | 1.7% | 2.5% |
| Domany 2020 (24h) | 9 | 9.20 | 14.80 | 9 | 11.90 | 16.40 | -0.16 | [-1.09; 0.76] | 0.7% | 1.2% |
| Domany 2021 (24h) | 15 | 7.80 | 10.50 | 15 | 11.50 | 10.80 | -0.34 | [-1.06; 0.38] | 1.1% | 1.8% |
| Fineberg 2023 (24h) | 10 | 3.10 | 5.63 | 11 | 3.18 | 5.67 | -0.01 | [-0.87; 0.84] | 0.8% | 1.4% |
| Fu 2020 (25d) | 100 | 0.97 | 1.27 | 93 | 1.08 | 1.21 | -0.09 | [-0.37; 0.19] | 7.1% | 4.8% |
| Grunebaum 2017 (24h) | 7 | 4.29 | 5.74 | 9 | 10.11 | 5.75 | -0.96 | [-2.02; 0.10] | 0.5% | 1.0% |
| Grunebaum 2018 (24h) | 40 | 6.05 | 5.48 | 40 | 11.63 | 7.43 | -0.85 | [-1.30; -0.39] | 2.7% | 3.3% |
| Hu 2016 (24h) | 13 | 0.70 | 0.70 | 14 | 1.30 | 0.40 | -1.03 | [-1.84; -0.22] | 0.9% | 1.5% |
| Ionescu 2019 (3w) | 12 | 1.20 | 1.40 | 12 | 1.40 | 1.40 | -0.14 | [-0.94; 0.66] | 0.9% | 1.6% |
| Ionescu 2020 (25d) | 90 | 0.68 | 1.04 | 92 | 0.93 | 1.18 | -0.22 | [-0.52; 0.07] | 6.7% | 4.8% |
| Jones 2024 (24h) | 17 | 10.80 | 8.08 | 10 | 12.30 | 7.83 | -0.18 | [-0.96; 0.60] | 0.9% | 1.6% |
| Su 2023 (24h) | 42 | 1.48 | 1.42 | 42 | 2.10 | 1.21 | -0.47 | [-0.90; -0.03] | 3.0% | 3.4% |
| Zhou 2024 (6d) | 27 | -2.60 | 2.00 | 25 | -1.70 | 2.20 | -0.42 | [-0.97; 0.13] | 1.9% | 2.6% |
| **Common effect model** | **499** | | | **491** | | | **-0.35** | **[-0.48; -0.22]** | **35.6%** | -- |
| **Random effects model** | | | | | | | **-0.37** | **[-0.56; -0.18]** | -- | **38.1%** |
| Heterogeneity: $I^2 = 27\%$, $\tau^2 = 0.0558$, $p = 0.15$ | | | | | | | | | | |
| **Common effect model** | **1513** | | | **1314** | | | **-0.19** | **[-0.27; -0.12]** | **100.0%** | -- |
| **Random effects model** | | | | | | | **-0.24** | **[-0.35; -0.13]** | -- | **100.0%** |

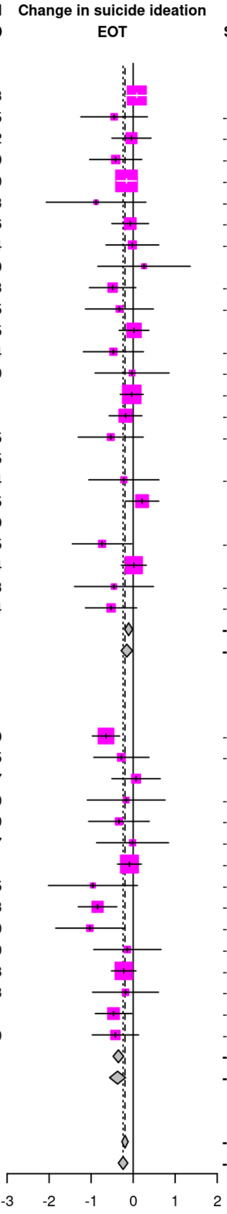

Heterogeneity: $I^2 = 24\%$, $\tau^2 = 0.0484$, $p = 0.10$
Test for subgroup differences (common effect): $\chi^2_1 = 9.25$, df = 1 ($p < 0.01$)
RE model$\chi^2_1 = 3.41$, df = 1 ($p = 0.06$)

#### Subgroup Analysis - Number of Doses

Results not provided for up to 72h because second doses are mostly given after a few days only.

### >72h up to 2w

| Study | Experimental Total | Experimental Mean | Experimental SD | Control Total | Control Mean | Control SD | SMD | 95%-CI | Weight (common) | Weight (random) |
|---|---|---|---|---|---|---|---|---|---|---|
| doses = multiple | | | | | | | | | | |
| Abbar 2022 | 73 | 3.50 | 7.18 | 80 | 8.00 | 11.30 | -0.47 | [-0.79; -0.15] | 5.6% | 5.1% |
| Ahmed 2023 | 18 | 75.72 | 20.58 | 18 | 80.67 | 13.15 | -0.28 | [-0.94; 0.38] | 1.3% | 1.9% |
| Anderson 2017 | 35 | 2.94 | 1.76 | 37 | 3.08 | 2.38 | -0.07 | [-0.53; 0.40] | 2.7% | 3.3% |
| Canuso 2018 | 29 | 1.76 | 2.03 | 26 | 1.54 | 1.68 | 0.12 | [-0.41; 0.65] | 2.1% | 2.7% |
| Chen 2023 | 117 | 0.25 | 0.60 | 111 | 0.23 | 0.61 | 0.03 | [-0.23; 0.29] | 8.6% | 6.2% |
| Colla 2024 | 17 | 1.18 | 0.87 | 10 | 1.60 | 0.95 | -0.45 | [-1.24; 0.34] | 0.9% | 1.4% |
| Daly 2018 PA | 40 | 0.20 | 0.55 | 32 | 0.22 | 0.42 | -0.04 | [-0.50; 0.43] | 2.7% | 3.3% |
| Daly 2018 PB | 20 | 0.15 | 0.49 | 21 | 0.38 | 0.59 | -0.41 | [-1.03; 0.20] | 1.5% | 2.1% |
| Fedgchin 2019 | 211 | 0.16 | 0.46 | 110 | 0.18 | 0.39 | -0.05 | [-0.28; 0.18] | 10.9% | 6.7% |
| Fu 2020 | 105 | 1.28 | 1.31 | 99 | 1.33 | 1.14 | -0.04 | [-0.32; 0.23] | 7.7% | 5.9% |
| George 2017 | 12 | 1.25 | 1.21 | 4 | 2.50 | 1.73 | -0.88 | [-2.07; 0.30] | 0.4% | 0.7% |
| Grabski 2022 | 41 | 0.10 | 0.30 | 44 | 0.00 | 0.00 | | | 0.0% | 0.0% |
| Ionescu 2019 | 13 | 1.30 | 1.40 | 13 | 1.50 | 1.40 | -0.14 | [-0.91; 0.63] | 1.0% | 1.5% |
| Ionescu 2020 | 92 | 0.97 | 1.19 | 100 | 1.35 | 1.34 | -0.30 | [-0.58; -0.01] | 7.1% | 5.7% |
| Loo 2016 | 9 | 2.00 | 1.66 | 5 | 1.60 | 0.89 | 0.26 | [-0.84; 1.36] | 0.5% | 0.8% |
| Ochs-Ross 2020 | 65 | 0.21 | 0.65 | 62 | 0.18 | 0.58 | 0.05 | [-0.30; 0.40] | 4.8% | 4.7% |
| Pattanaseri 2024 | 11 | 1.64 | 1.75 | 9 | 1.44 | 1.33 | 0.12 | [-0.76; 1.00] | 0.7% | 1.2% |
| Popova 2019 | 109 | 0.08 | 0.36 | 107 | 0.18 | 0.53 | -0.22 | [-0.49; 0.05] | 8.1% | 6.0% |
| Singh 2016 2x doses | 15 | 0.07 | 0.26 | 12 | 0.33 | 0.65 | -0.53 | [-1.31; 0.24] | 1.0% | 1.5% |
| Singh 2016 3x doses | 15 | 0.00 | 0.00 | 12 | 0.25 | 0.45 | | | 0.0% | 0.0% |
| Smith-Apeldoorn 2024 | 53 | 1.34 | 0.90 | 51 | 1.47 | 0.83 | -0.15 | [-0.53; 0.24] | 3.9% | 4.2% |
| Takahashi 2021 | 116 | 0.18 | 0.61 | 74 | 0.27 | 0.76 | -0.13 | [-0.43; 0.16] | 6.8% | 5.6% |
| Zhou 2024 | 27 | -2.80 | 2.10 | 23 | -2.20 | 2.50 | -0.26 | [-0.82; 0.30] | 1.9% | 2.5% |
| Common effect model | 1243 | | | 1060 | | | **-0.14** | **[-0.22; -0.05]** | **80.0%** | **--** |
| Random effects model | | | | | | | **-0.15** | **[-0.26; -0.03]** | **--** | **72.8%** |
| Heterogeneity: $I^2 = 0\%$, $\tau^2 = 0.0225$, $p = 0.73$ | | | | | | | | | | |
| doses = single | | | | | | | | | | |
| Burger 2016 | 3 | 2.40 | 13.61 | 7 | 6.70 | 8.93 | -0.38 | [-1.75; 0.99] | 0.3% | 0.5% |
| Domany 2020 | 9 | 7.40 | 18.00 | 9 | 1.40 | 2.20 | 0.45 | [-0.49; 1.38] | 0.7% | 1.0% |
| Domany 2021 | 15 | 3.10 | 14.10 | 15 | 11.40 | 15.20 | -0.55 | [-1.28; 0.18] | 1.1% | 1.6% |
| Duek 2023 | 14 | 0.14 | 0.36 | 13 | 0.08 | 0.28 | 0.20 | [-0.56; 0.95] | 1.0% | 1.5% |
| Fineberg 2023 | 10 | 4.30 | 7.10 | 12 | 5.67 | 7.04 | -0.19 | [-1.03; 0.65] | 0.8% | 1.3% |
| Hu 2016 | 12 | 0.90 | 0.90 | 14 | 1.00 | 0.50 | -0.14 | [-0.91; 0.64] | 1.0% | 1.5% |
| Lii 2023 | 19 | 0.32 | 0.58 | 20 | 0.40 | 0.77 | -0.11 | [-0.74; 0.51] | 1.5% | 2.1% |
| Murrough 2015 II | 12 | 7.92 | 8.73 | 12 | 10.08 | 9.39 | -0.23 | [-1.03; 0.57] | 0.9% | 1.4% |
| Price 2022 | 52 | 0.79 | 1.32 | 46 | 1.30 | 1.62 | -0.34 | [-0.74; 0.05] | 3.6% | 4.0% |
| Sos 2013 | 9 | 0.44 | 0.73 | 18 | 0.17 | 0.87 | 0.32 | [-0.49; 1.12] | 0.9% | 1.4% |
| Su 2017 | 47 | 0.98 | 0.87 | 24 | 1.29 | 0.81 | -0.36 | [-0.86; 0.13] | 2.4% | 3.0% |
| Su 2023 | 42 | 1.95 | 1.36 | 42 | 2.12 | 1.33 | -0.13 | [-0.55; 0.30] | 3.2% | 3.6% |
| Sumner 2019 | 17 | 0.94 | 0.97 | 16 | 1.63 | 0.96 | -0.70 | [-1.40; 0.01] | 1.2% | 1.7% |
| Zarate 2006 | 9 | 0.89 | 1.27 | 9 | 1.00 | 0.82 | -0.10 | [-1.02; 0.83] | 0.7% | 1.1% |
| Zarate 2012 | 11 | 2.09 | 1.38 | 17 | 2.12 | 1.05 | -0.02 | [-0.78; 0.74] | 1.0% | 1.5% |
| Common effect model | 281 | | | 274 | | | **-0.20** | **[-0.37; -0.03]** | **20.0%** | **--** |
| Random effects model | | | | | | | **-0.19** | **[-0.39; 0.01]** | **--** | **27.2%** |
| Heterogeneity: $I^2 = 0\%$, $\tau^2 = 0.0333$, $p = 0.85$ | | | | | | | | | | |
| **Common effect model** | **1524** | | | **1334** | | | **-0.15** | **[-0.23; -0.07]** | **100.0%** | **--** |
| **Random effects model** | | | | | | | **-0.16** | **[-0.26; -0.06]** | **--** | **100.0%** |

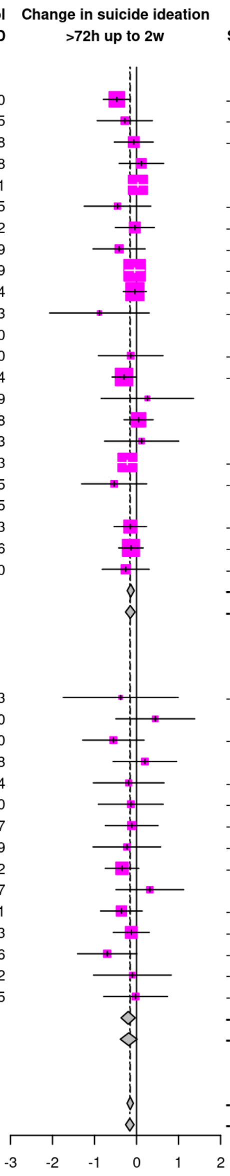

Heterogeneity: $I^2 = 0\%$, $\tau^2 = 0.0259$, $p = 0.90$
Test for subgroup differences (common effect): $\chi^2_1 = 0.39$, df = 1 ($p = 0.53$)
RE model$\chi^2_1 = 0.10$, df = 1 ($p = 0.75$)

# >2w up to 4w

| Study | Experimental Total | Experimental Mean | Experimental SD | Control Total | Control Mean | Control SD | SMD | 95%-CI | Weight (common) | Weight (random) |
|---|---|---|---|---|---|---|---|---|---|---|
| **doses = multiple** | | | | | | | | | | |
| Abbar 2022 | 73 | 2.83 | 5.67 | 80 | 6.43 | 9.66 | -0.45 | [-0.77; -0.13] | 7.4% | 7.1% |
| Canuso 2018 | 26 | 1.00 | 1.44 | 21 | 0.90 | 1.37 | 0.07 | [-0.51; 0.64] | 2.3% | 3.8% |
| Chen 2023 | 108 | 0.26 | 0.62 | 107 | 0.21 | 0.53 | 0.09 | [-0.18; 0.35] | 10.7% | 8.1% |
| Fedgchin 2019 | 204 | 0.11 | 0.37 | 107 | 0.17 | 0.40 | -0.16 | [-0.39; 0.08] | 13.9% | 8.7% |
| Fu 2020 | 100 | 0.97 | 1.27 | 93 | 1.08 | 1.21 | -0.09 | [-0.37; 0.19] | 9.6% | 7.8% |
| Grabski 2022 | 39 | 0.05 | 0.22 | 43 | 0.00 | 0.00 | | | 0.0% | 0.0% |
| Ionescu 2019 | 12 | 1.20 | 1.40 | 12 | 1.40 | 1.40 | -0.14 | [-0.94; 0.66] | 1.2% | 2.3% |
| Ionescu 2020 | 90 | 0.68 | 1.04 | 92 | 0.93 | 1.18 | -0.22 | [-0.52; 0.07] | 9.0% | 7.7% |
| Ochs-Ross 2020 | 62 | 0.18 | 0.51 | 61 | 0.17 | 0.45 | 0.02 | [-0.33; 0.37] | 6.1% | 6.6% |
| Ohtani 2024 | 15 | 1.40 | 1.08 | 16 | 1.94 | 1.14 | -0.47 | [-1.19; 0.25] | 1.5% | 2.8% |
| Popova 2019 | 103 | 0.07 | 0.29 | 102 | 0.08 | 0.31 | -0.03 | [-0.31; 0.24] | 10.2% | 8.0% |
| Smith-Apeldoorn 2024 | 50 | 1.42 | 0.83 | 49 | 1.39 | 1.02 | 0.03 | [-0.36; 0.43] | 4.9% | 5.9% |
| Takahashi 2021 | 111 | 0.15 | 0.65 | 72 | 0.14 | 0.54 | 0.02 | [-0.28; 0.31] | 8.7% | 7.6% |
| Zhou 2024 | 26 | -2.90 | 2.20 | 23 | -1.90 | 2.30 | -0.44 | [-1.01; 0.13] | 2.4% | 3.9% |
| Common effect model | **1019** | | | **878** | | | **-0.10** | **[-0.20; -0.01]** | **87.8%** | **--** |
| Random effects model | | | | | | | **-0.11** | **[-0.23; 0.01]** | **--** | **80.4%** |
| Heterogeneity: $I^2 = 0\%$, $\tau^2 = 0.0172$, $p = 0.47$ | | | | | | | | | | |
| **doses = single** | | | | | | | | | | |
| Domany 2021 | 15 | 3.00 | 14.50 | 15 | 8.30 | 16.00 | -0.34 | [-1.06; 0.38] | 1.5% | 2.8% |
| Duek 2023 | 13 | 0.31 | 0.48 | 11 | 0.18 | 0.41 | 0.27 | [-0.54; 1.08] | 1.2% | 2.3% |
| Fineberg 2023 | 10 | 7.10 | 6.01 | 11 | 4.73 | 5.75 | 0.39 | [-0.48; 1.25] | 1.0% | 2.1% |
| Hu 2016 | 12 | 0.40 | 0.90 | 14 | 0.20 | 0.40 | 0.29 | [-0.49; 1.06] | 1.3% | 2.5% |
| Price 2022 | 49 | 0.61 | 1.27 | 41 | 1.68 | 1.71 | -0.71 | [-1.14; -0.29] | 4.2% | 5.5% |
| Su 2017 | 47 | 1.00 | 0.75 | 24 | 1.33 | 1.01 | -0.39 | [-0.88; 0.11] | 3.1% | 4.6% |
| Common effect model | **146** | | | **116** | | | **-0.29** | **[-0.54; -0.04]** | **12.2%** | **--** |
| Random effects model | | | | | | | **-0.17** | **[-0.57; 0.22]** | **--** | **19.6%** |
| Heterogeneity: $I^2 = 51\%$, $\tau^2 = 0.1269$, $p = 0.07$ | | | | | | | | | | |
| **Common effect model** | **1165** | | | **994** | | | **-0.13** | **[-0.21; -0.04]** | **100.0%** | **--** |
| **Random effects model** | | | | | | | **-0.14** | **[-0.27; 0.00]** | **--** | **100.0%** |

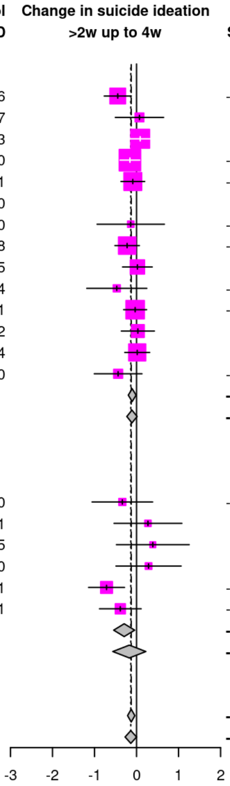

Heterogeneity: $I^2 = 25\%$, $\tau^2 = 0.0414$, $p = 0.16$
Test for subgroup differences (common effect): $\chi^2_1 = 1.96$, df = 1 ($p = 0.16$)
RE model$\chi^2_1 = 0.09$, df = 1 ($p = 0.76$)

## >4w

| Study | Experimental Total | Experimental Mean | Experimental SD | Control Total | Control Mean | Control SD | SMD | 95%-CI | Weight (common) | Weight (random) |
|---|---|---|---|---|---|---|---|---|---|---|
| doses = multiple | | | | | | | | | | |
| Abbar 2022 | 60 | 2.60 | 5.90 | 66 | 4.00 | 8.30 | -0.19 | [-0.54; 0.16] | 15.9% | 15.1% |
| Fu 2020 | 84 | 1.04 | 1.31 | 79 | 0.84 | 1.20 | 0.16 | [-0.15; 0.47] | 20.7% | 17.3% |
| Grabski 2022 | 38 | 0.05 | 0.23 | 43 | 0.07 | 0.26 | -0.07 | [-0.51; 0.37] | 10.3% | 11.5% |
| Ionescu 2020 | 80 | 0.49 | 1.08 | 86 | 0.66 | 0.98 | -0.16 | [-0.47; 0.14] | 21.0% | 17.5% |
| Pattanaseri 2024 | 11 | 1.64 | 1.50 | 9 | 1.89 | 1.27 | -0.17 | [-1.05; 0.71] | 2.5% | 3.7% |
| Smith-Apeldoorn 2024 | 48 | 1.67 | 0.75 | 46 | 1.35 | 0.82 | 0.40 | [-0.01; 0.81] | 11.7% | 12.5% |
| Zhou 2024 | 26 | -2.80 | 2.10 | 23 | -2.10 | 2.30 | -0.31 | [-0.88; 0.25] | 6.1% | 7.9% |
| Common effect model | **347** | | | **352** | | | **-0.02** | **[-0.17; 0.13]** | **88.2%** | **--** |
| Random effects model | | | | | | | **-0.02** | **[-0.23; 0.18]** | **--** | **85.4%** |
| Heterogeneity: $I^2 = 28\%$, $\tau^2 = 0.0318$, $p = 0.21$ | | | | | | | | | | |
| doses = single | | | | | | | | | | |
| Duek 2023 | 11 | 0.30 | 0.48 | 11 | 0.20 | 0.42 | 0.21 | [-0.63; 1.05] | 2.8% | 4.1% |
| Price 2022 | 36 | 0.56 | 0.97 | 35 | 0.60 | 0.85 | -0.04 | [-0.51; 0.42] | 9.0% | 10.5% |
| Common effect model | **47** | | | **46** | | | **0.02** | **[-0.39; 0.42]** | **11.8%** | **--** |
| Random effects model | | | | | | | **0.02** | **[-0.40; 0.44]** | **--** | **14.6%** |
| Heterogeneity: $I^2 = 0\%$, $\tau^2 = 0.0039$, $p = 0.60$ | | | | | | | | | | |
| **Common effect model** | **394** | | | **398** | | | **-0.01** | **[-0.15; 0.13]** | **100.0%** | **--** |
| **Random effects model** | | | | | | | **-0.01** | **[-0.20; 0.17]** | **--** | **100.0%** |

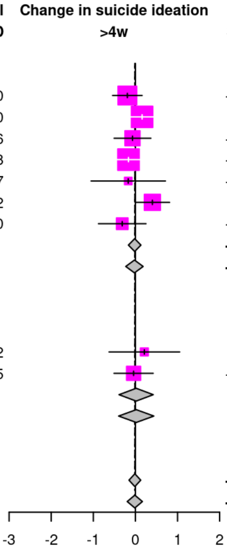

Heterogeneity: $I^2 = 8\%$, $\tau^2 = 0.0242$, $p = 0.37$
Test for subgroup differences (common effect): $\chi^2_1 = 0.02$, df = 1 ($p = 0.88$)
RE model$\chi^2_1 = 0.03$, df = 1 ($p = 0.86$)

#### End of Treatment

| Study | Experimental Total | Experimental Mean | Experimental SD | Control Total | Control Mean | Control SD | SMD | 95%-CI | Weight (common) | Weight (random) |
|---|---|---|---|---|---|---|---|---|---|---|
| **doses = multiple** | | | | | | | | | | |
| Abbar 2022 (day 2) | 73 | 3.00 | 6.05 | 80 | 8.50 | 10.19 | -0.65 | [-0.97; -0.32] | 5.4% | 4.4% |
| Ahmed 2023 (2w) | 18 | 75.72 | 20.58 | 18 | 80.67 | 13.15 | -0.28 | [-0.94; 0.38] | 1.3% | 2.1% |
| Canuso 2018 (25d) | 26 | 1.00 | 1.44 | 21 | 0.90 | 1.37 | 0.07 | [-0.51; 0.64] | 1.7% | 2.5% |
| Chen 2023 (4w) | 108 | 0.26 | 0.62 | 107 | 0.21 | 0.53 | 0.09 | [-0.18; 0.35] | 8.0% | 5.0% |
| Colla 2024 (2w) | 17 | 1.18 | 0.87 | 10 | 1.60 | 0.95 | -0.45 | [-1.24; 0.34] | 0.9% | 1.6% |
| Daly 2018 PA (7d) | 40 | 0.20 | 0.55 | 32 | 0.22 | 0.42 | -0.04 | [-0.50; 0.43] | 2.6% | 3.2% |
| Daly 2018 PB (7d) | 20 | 0.15 | 0.49 | 21 | 0.38 | 0.59 | -0.41 | [-1.03; 0.20] | 1.5% | 2.3% |
| Fedgchin 2019 (4w) | 204 | 0.11 | 0.37 | 107 | 0.17 | 0.40 | -0.16 | [-0.39; 0.08] | 10.4% | 5.3% |
| Fu 2020 (25d) | 100 | 0.97 | 1.27 | 93 | 1.08 | 1.21 | -0.09 | [-0.37; 0.19] | 7.1% | 4.8% |
| George 2017 (1w) | 12 | 1.25 | 1.21 | 4 | 2.50 | 1.73 | -0.88 | [-2.07; 0.30] | 0.4% | 0.8% |
| Grabski 2022 (1-3w) | 38 | 0.05 | 0.23 | 43 | 0.07 | 0.26 | -0.07 | [-0.51; 0.37] | 3.0% | 3.4% |
| Ionescu 2019 (3w) | 12 | 1.20 | 1.40 | 12 | 1.40 | 1.40 | -0.14 | [-0.94; 0.66] | 0.9% | 1.6% |
| Ionescu 2020 (25d) | 90 | 0.68 | 1.04 | 92 | 0.93 | 1.18 | -0.22 | [-0.52; 0.07] | 6.7% | 4.8% |
| Loo 2016 (1w-2w) | 9 | 2.00 | 1.66 | 5 | 1.60 | 0.89 | 0.26 | [-0.84; 1.36] | 0.5% | 0.9% |
| Ochs-Ross 2020 (4w) | 62 | 0.18 | 0.51 | 61 | 0.17 | 0.45 | 0.02 | [-0.33; 0.37] | 4.6% | 4.1% |
| Ohtani 2024 (2w) | 15 | 1.40 | 1.08 | 16 | 1.94 | 1.14 | -0.47 | [-1.19; 0.25] | 1.1% | 1.8% |
| Pattanaseri 2024 (3d) | 11 | 1.18 | 1.60 | 9 | 1.22 | 1.30 | -0.03 | [-0.91; 0.86] | 0.7% | 1.3% |
| Popova 2019 (4w) | 103 | 0.07 | 0.29 | 102 | 0.08 | 0.31 | -0.03 | [-0.31; 0.24] | 7.6% | 4.9% |
| Singh 2016 2x doses (2w) | 15 | 0.07 | 0.26 | 12 | 0.33 | 0.65 | -0.53 | [-1.31; 0.24] | 1.0% | 1.6% |
| Singh 2016 3x doses (2w) | 15 | 0.00 | 0.00 | 12 | 0.25 | 0.45 | | | 0.0% | 0.0% |
| Singh 2016 II (4d) | 18 | 0.56 | 1.29 | 8 | 0.88 | 1.64 | -0.22 | [-1.06; 0.61] | 0.8% | 1.5% |
| Smith-Apeldoorn 2024 (6w) | 50 | 1.60 | 0.83 | 48 | 1.42 | 0.85 | 0.21 | [-0.18; 0.61] | 3.6% | 3.7% |
| Takahashi 2021 (4w) | 111 | 0.15 | 0.65 | 72 | 0.14 | 0.54 | 0.02 | [-0.28; 0.31] | 6.5% | 4.7% |
| Zhou 2024 (6d) | 27 | -2.60 | 2.00 | 25 | -1.70 | 2.20 | -0.42 | [-0.97; 0.13] | 1.9% | 2.6% |
| Common effect model | **1194** | | | **1010** | | | **-0.13** | **[-0.21; -0.04]** | **78.2%** | **--** |
| Random effects model | | | | | | | **-0.14** | **[-0.27; -0.02]** | **--** | **69.1%** |
| Heterogeneity: $I^2 = 12\%$, $\tau^2 = 0.0330$, $p = 0.29$ | | | | | | | | | | |
| **doses = single** | | | | | | | | | | |
| Domany 2020 (24h) | 9 | 9.20 | 14.80 | 9 | 11.90 | 16.40 | -0.16 | [-1.09; 0.76] | 0.7% | 1.2% |
| Domany 2021 (24h) | 15 | 7.80 | 10.50 | 15 | 11.50 | 10.80 | -0.34 | [-1.06; 0.38] | 1.1% | 1.8% |
| Fineberg 2023 (24h) | 10 | 3.10 | 5.63 | 11 | 3.18 | 5.67 | -0.01 | [-0.87; 0.84] | 0.8% | 1.4% |
| Grunebaum 2017 (24h) | 7 | 4.29 | 5.74 | 9 | 10.11 | 5.75 | -0.96 | [-2.02; 0.10] | 0.5% | 1.0% |
| Grunebaum 2018 (24h) | 40 | 6.05 | 5.48 | 40 | 11.63 | 7.43 | -0.85 | [-1.30; -0.39] | 2.7% | 3.3% |
| Hu 2016 (24h) | 13 | 0.70 | 0.70 | 14 | 1.30 | 0.40 | -1.03 | [-1.84; -0.22] | 0.9% | 1.5% |
| Jones 2024 (24h) | 17 | 10.80 | 8.08 | 10 | 12.30 | 7.83 | -0.18 | [-0.96; 0.60] | 0.9% | 1.6% |
| Lii 2023 (24h) | 20 | 0.20 | 0.52 | 19 | 0.21 | 0.54 | -0.02 | [-0.65; 0.61] | 1.4% | 2.2% |
| Murrough 2013 (24h) | 26 | 0.85 | 1.12 | 26 | 1.46 | 1.33 | -0.49 | [-1.04; 0.06] | 1.9% | 2.6% |
| Murrough 2015 II (24h) | 12 | 10.83 | 8.48 | 12 | 14.00 | 10.15 | -0.33 | [-1.13; 0.48] | 0.9% | 1.5% |
| Price 2022 (24h) | 53 | 0.38 | 0.98 | 49 | 0.59 | 1.31 | -0.18 | [-0.57; 0.21] | 3.8% | 3.8% |
| Sos 2013 (24h) | 9 | 0.56 | 0.73 | . | 0.39 | 0.59 | | | 0.0% | 0.0% |
| Su 2023 (24h) | 42 | 1.48 | 1.42 | 42 | 2.10 | 1.21 | -0.47 | [-0.90; -0.03] | 3.0% | 3.4% |
| Sumner 2019 (24h) | 17 | 0.65 | 0.79 | 16 | 1.31 | 0.95 | -0.74 | [-1.45; -0.03] | 1.1% | 1.9% |
| Zarate 2006 (24h) | 9 | 0.44 | 0.53 | 9 | 0.78 | 0.83 | -0.46 | [-1.40; 0.48] | 0.6% | 1.2% |
| Zarate 2012 (24h) | 20 | 1.30 | 1.38 | 23 | 2.00 | 1.24 | -0.53 | [-1.14; 0.09] | 1.5% | 2.3% |
| Common effect model | **319** | | | **304** | | | **-0.44** | **[-0.60; -0.28]** | **21.8%** | **--** |
| Random effects model | | | | | | | **-0.44** | **[-0.64; -0.24]** | **--** | **30.9%** |
| Heterogeneity: $I^2 = 0\%$, $\tau^2 = 0.0428$, $p = 0.60$ | | | | | | | | | | |
| **Common effect model** | **1513** | | | **1314** | | | **-0.19** | **[-0.27; -0.12]** | **100.0%** | **--** |
| **Random effects model** | | | | | | | **-0.24** | **[-0.35; -0.13]** | **--** | **100.0%** |

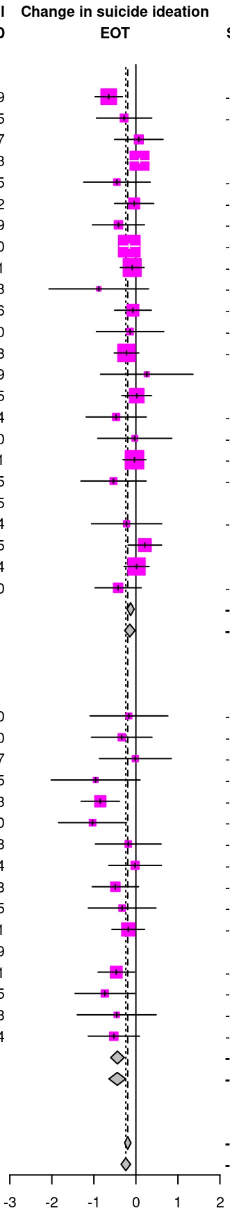

Heterogeneity: $I^2 = 24\%$, $\tau^2 = 0.0484$, $p = 0.10$
Test for subgroup differences (common effect): $\chi^2_1 = 11.33$, df = 1 ($p < 0.01$)
RE model$\chi^2_1 = 6.21$, df = 1 ($p = 0.01$)

#### Subgroup Analysis - Number of Doses, only trials with suicidal patients

Results not provided for up to 72h because second doses are mostly given after a few days only.

### >72h up to 2w

| Study | Experimental Total | Experimental Mean | Experimental SD | Control Total | Control Mean | Control SD | SMD | 95%-CI | Weight (common) | Weight (random) |
|---|---|---|---|---|---|---|---|---|---|---|
| doses = multiple | | | | | | | | | | |
| Abbar 2022 | 73 | 3.50 | 7.18 | 80 | 8.00 | 11.30 | -0.47 | [-0.79; -0.15] | 16.6% | 14.8% |
| Ahmed 2023 | 18 | 75.72 | 20.58 | 18 | 80.67 | 13.15 | -0.28 | [-0.94; 0.38] | 4.0% | 5.4% |
| Canuso 2018 | 29 | 1.76 | 2.03 | 26 | 1.54 | 1.68 | 0.12 | [-0.41; 0.65] | 6.1% | 7.6% |
| Fu 2020 | 105 | 1.28 | 1.31 | 99 | 1.33 | 1.14 | -0.04 | [-0.32; 0.23] | 22.8% | 17.5% |
| Ionescu 2019 | 13 | 1.30 | 1.40 | 13 | 1.50 | 1.40 | -0.14 | [-0.91; 0.63] | 2.9% | 4.1% |
| Ionescu 2020 | 92 | 0.97 | 1.19 | 100 | 1.35 | 1.34 | -0.30 | [-0.58; -0.01] | 21.2% | 16.9% |
| Zhou 2024 | 27 | -2.80 | 2.10 | 23 | -2.20 | 2.50 | -0.26 | [-0.82; 0.30] | 5.5% | 7.0% |
| Common effect model | **357** | | | **359** | | | **-0.22** | **[-0.37; -0.07]** | **79.2%** | **--** |
| Random effects model | | | | | | | **-0.22** | **[-0.40; -0.03]** | **--** | **73.3%** |
| Heterogeneity: $I^2 = 0\%$, $\tau^2 = 0.0155$, $p = 0.44$ | | | | | | | | | | |
| doses = single | | | | | | | | | | |
| Burger 2016 | 3 | 2.40 | 13.61 | 7 | 6.70 | 8.93 | -0.38 | [-1.75; 0.99] | 0.9% | 1.4% |
| Domany 2020 | 9 | 7.40 | 18.00 | 9 | 1.40 | 2.20 | 0.45 | [-0.49; 1.38] | 2.0% | 2.9% |
| Domany 2021 | 15 | 3.10 | 14.10 | 15 | 11.40 | 15.20 | -0.55 | [-1.28; 0.18] | 3.2% | 4.5% |
| Fineberg 2023 | 10 | 4.30 | 7.10 | 12 | 5.67 | 7.04 | -0.19 | [-1.03; 0.65] | 2.4% | 3.5% |
| Hu 2016 | 12 | 0.90 | 0.90 | 14 | 1.00 | 0.50 | -0.14 | [-0.91; 0.64] | 2.9% | 4.1% |
| Su 2023 | 42 | 1.95 | 1.36 | 42 | 2.12 | 1.33 | -0.13 | [-0.55; 0.30] | 9.4% | 10.4% |
| Common effect model | **91** | | | **99** | | | **-0.16** | **[-0.44; 0.13]** | **20.8%** | **--** |
| Random effects model | | | | | | | **-0.16** | **[-0.50; 0.18]** | **--** | **26.7%** |
| Heterogeneity: $I^2 = 0\%$, $\tau^2 = 0.0356$, $p = 0.73$ | | | | | | | | | | |
| **Common effect model** | **448** | | | **458** | | | **-0.21** | **[-0.34; -0.07]** | **100.0%** | **--** |
| **Random effects model** | | | | | | | **-0.20** | **[-0.37; -0.03]** | **--** | **100.0%** |

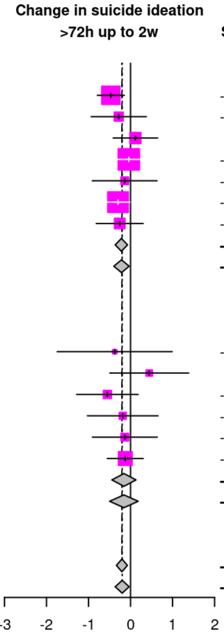

Heterogeneity: $I^2 = 0\%$, $\tau^2 = 0.0216$, $p = 0.72$
Test for subgroup differences (common effect): $\chi^2_1 = 0.14$, df = 1 ($p = 0.71$)
RE model$\chi^2_1 = 0.08$, df = 1 ($p = 0.77$)

## >2w up to 4w

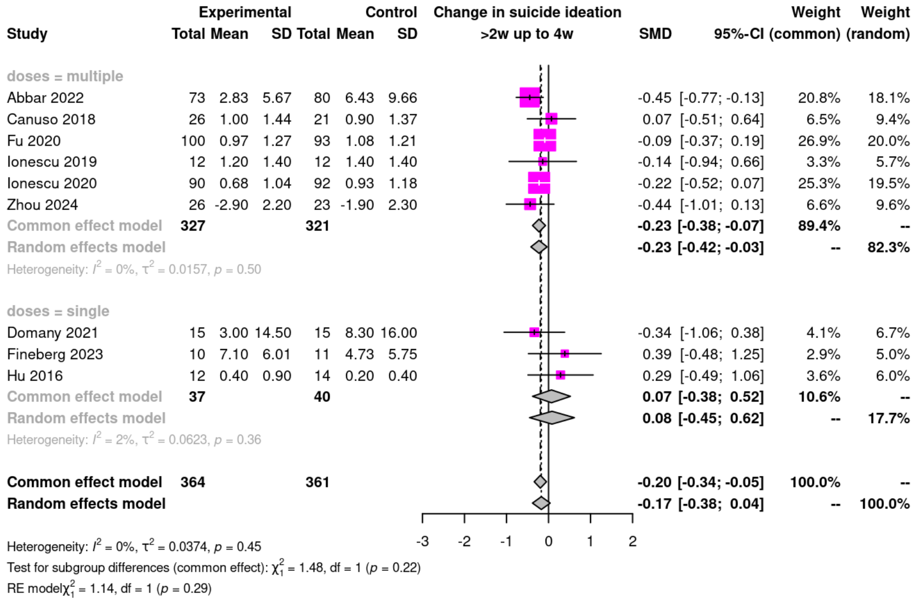
Study
Experimental Total Mean SD
Control Total Mean SD
Change in suicide ideation >2w up to 4w
SMD 95%-CI
Weight (common)
Weight (random)
doses = multiple
Abbar 2022 73 2.83 5.67 80 6.43 9.66 -0.45 [-0.77; -0.13] 20.8% 18.1%
Canuso 2018 26 1.00 1.44 21 0.90 1.37 0.07 [-0.51; 0.64] 6.5% 9.4%
Fu 2020 100 0.97 1.27 93 1.08 1.21 -0.09 [-0.37; 0.19] 26.9% 20.0%
Ionescu 2019 12 1.20 1.40 12 1.40 1.40 -0.14 [-0.94; 0.66] 3.3% 5.7%
Ionescu 2020 90 0.68 1.04 92 0.93 1.18 -0.22 [-0.52; 0.07] 25.3% 19.5%
Zhou 2024 26 -2.90 2.20 23 -1.90 2.30 -0.44 [-1.01; 0.13] 6.6% 9.6%
Common effect model 327 321 -0.23 [-0.38; -0.07] 89.4% --
Random effects model -0.23 [-0.42; -0.03] -- 82.3%
Heterogeneity: $I^2$ = 0%, $\tau^2$ = 0.0157, $p$ = 0.50
doses = single
Domany 2021 15 3.00 14.50 15 8.30 16.00 -0.34 [-1.06; 0.38] 4.1% 6.7%
Fineberg 2023 10 7.10 6.01 11 4.73 5.75 0.39 [-0.48; 1.25] 2.9% 5.0%
Hu 2016 12 0.40 0.90 14 0.20 0.40 0.29 [-0.49; 1.06] 3.6% 6.0%
Common effect model 37 40 0.07 [-0.38; 0.52] 10.6% --
Random effects model 0.08 [-0.45; 0.62] -- 17.7%
Heterogeneity: $I^2$ = 2%, $\tau^2$ = 0.0623, $p$ = 0.36
Common effect model 364 361 -0.20 [-0.34; -0.05] 100.0% --
Random effects model -0.17 [-0.38; 0.04] -- 100.0%
-3 -2 -1 0 1 2
Heterogeneity: $I^2$ = 0%, $\tau^2$ = 0.0374, $p$ = 0.45
Test for subgroup differences (common effect): $\chi^2_1$ = 1.48, df = 1 ($p$ = 0.22)
RE model$\chi^2_1$ = 1.14, df = 1 ($p$ = 0.29)

# >4w

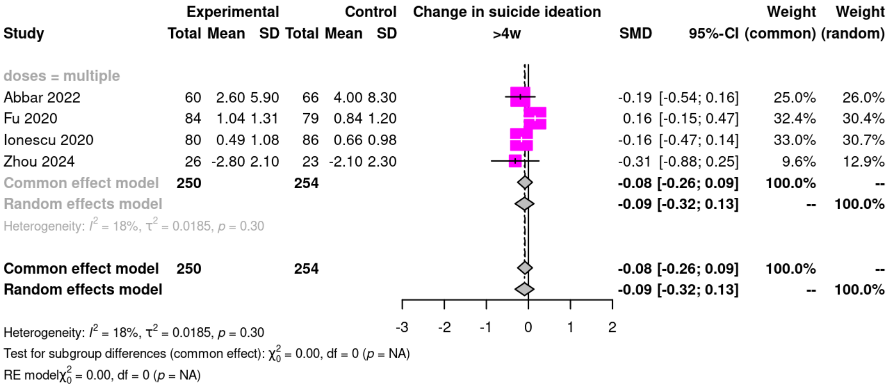

Experimental
Control
Change in suicide ideation
>4w
Study
Total Mean SD Total Mean SD
SMD
95%-CI
Weight (common)
Weight (random)
doses = multiple
Abbar 2022 60 2.60 5.90 66 4.00 8.30 -0.19 [-0.54; 0.16] 25.0% 26.0%
Fu 2020 84 1.04 1.31 79 0.84 1.20 0.16 [-0.15; 0.47] 32.4% 30.4%
Ionescu 2020 80 0.49 1.08 86 0.66 0.98 -0.16 [-0.47; 0.14] 33.0% 30.7%
Zhou 2024 26 -2.80 2.10 23 -2.10 2.30 -0.31 [-0.88; 0.25] 9.6% 12.9%
Common effect model 250 254 -0.08 [-0.26; 0.09] 100.0% --
Random effects model -0.09 [-0.32; 0.13] -- 100.0%
Heterogeneity: $I^2$ = 18%, $\tau^2$ = 0.0185, $p$ = 0.30
Common effect model 250 254 -0.08 [-0.26; 0.09] 100.0% --
Random effects model -0.09 [-0.32; 0.13] -- 100.0%
-3 -2 -1 0 1 2
Heterogeneity: $I^2$ = 18%, $\tau^2$ = 0.0185, $p$ = 0.30
Test for subgroup differences (common effect): $\chi^2_0$ = 0.00, df = 0 ($p$ = NA)
RE model$\chi^2_0$ = 0.00, df = 0 ($p$ = NA)

#### End of Treatment

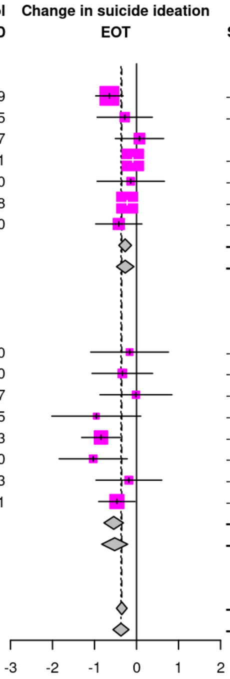

| Study | Experimental Total | Experimental Mean | Experimental SD | Control Total | Control Mean | Control SD | SMD | 95%-CI | Weight (common) | Weight (random) |
|---|---|---|---|---|---|---|---|---|---|---|
| doses = multiple | | | | | | | | | | |
| Abbar 2022 (day 2) | 73 | 3.00 | 6.05 | 80 | 8.50 | 10.19 | -0.65 | [-0.97; -0.32] | 15.1% | 11.3% |
| Ahmed 2023 (2w) | 18 | 75.72 | 20.58 | 18 | 80.67 | 13.15 | -0.28 | [-0.94; 0.38] | 3.7% | 5.6% |
| Canuso 2018 (25d) | 26 | 1.00 | 1.44 | 21 | 0.90 | 1.37 | 0.07 | [-0.51; 0.64] | 4.8% | 6.6% |
| Fu 2020 (25d) | 100 | 0.97 | 1.27 | 93 | 1.08 | 1.21 | -0.09 | [-0.37; 0.19] | 20.1% | 12.3% |
| Ionescu 2019 (3w) | 12 | 1.20 | 1.40 | 12 | 1.40 | 1.40 | -0.14 | [-0.94; 0.66] | 2.5% | 4.2% |
| Ionescu 2020 (25d) | 90 | 0.68 | 1.04 | 92 | 0.93 | 1.18 | -0.22 | [-0.52; 0.07] | 18.8% | 12.1% |
| Zhou 2024 (6d) | 27 | -2.60 | 2.00 | 25 | -1.70 | 2.20 | -0.42 | [-0.97; 0.13] | 5.3% | 7.0% |
| Common effect model | **346** | | | **341** | | | **-0.27** | **[-0.42; -0.12]** | **70.3%** | **--** |
| Random effects model | | | | | | | **-0.27** | **[-0.48; -0.06]** | **--** | **59.1%** |
| Heterogeneity: $I^2$ = 30%, $\tau^2$ = 0.0278, $p$ = 0.20 | | | | | | | | | | |
| doses = single | | | | | | | | | | |
| Domany 2020 (24h) | 9 | 9.20 | 14.80 | 9 | 11.90 | 16.40 | -0.16 | [-1.09; 0.76] | 1.9% | 3.4% |
| Domany 2021 (24h) | 15 | 7.80 | 10.50 | 15 | 11.50 | 10.80 | -0.34 | [-1.06; 0.38] | 3.1% | 4.9% |
| Fineberg 2023 (24h) | 10 | 3.10 | 5.63 | 11 | 3.18 | 5.67 | -0.01 | [-0.87; 0.84] | 2.2% | 3.8% |
| Grunebaum 2017 (24h) | 7 | 4.29 | 5.74 | 9 | 10.11 | 5.75 | -0.96 | [-2.02; 0.10] | 1.4% | 2.7% |
| Grunebaum 2018 (24h) | 40 | 6.05 | 5.48 | 40 | 11.63 | 7.43 | -0.85 | [-1.30; -0.39] | 7.6% | 8.5% |
| Hu 2016 (24h) | 13 | 0.70 | 0.70 | 14 | 1.30 | 0.40 | -1.03 | [-1.84; -0.22] | 2.4% | 4.1% |
| Jones 2024 (24h) | 17 | 10.80 | 8.08 | 10 | 12.30 | 7.83 | -0.18 | [-0.96; 0.60] | 2.6% | 4.4% |
| Su 2023 (24h) | 42 | 1.48 | 1.42 | 42 | 2.10 | 1.21 | -0.47 | [-0.90; -0.03] | 8.5% | 9.0% |
| Common effect model | **153** | | | **150** | | | **-0.54** | **[-0.78; -0.31]** | **29.7%** | **--** |
| Random effects model | | | | | | | **-0.52** | **[-0.83; -0.21]** | **--** | **40.9%** |
| Heterogeneity: $I^2$ = 0%, $\tau^2$ = 0.0683, $p$ = 0.43 | | | | | | | | | | |
| **Common effect model** | **499** | | | **491** | | | **-0.35** | **[-0.48; -0.22]** | **100.0%** | **--** |
| **Random effects model** | | | | | | | **-0.37** | **[-0.56; -0.18]** | **--** | **100.0%** |

Heterogeneity: $I^2$ = 27%, $\tau^2$ = 0.0558, $p$ = 0.15
Test for subgroup differences (common effect): $\chi^2_1$ = 3.73, df = 1 ($p$ = 0.05)
RE model$\chi^2_1$ = 1.74, df = 1 ($p$ = 0.19)

#### Subgroup Analysis - Clinician vs. Patient Ratings

## Up to 4h

| Study | Experimental Total | Experimental Mean | Experimental SD | Control Total | Control Mean | Control SD | SMD | 95%-CI | Weight (common) | Weight (random) |
|---|---|---|---|---|---|---|---|---|---|---|
| scale_mode = clinician | | | | | | | | | | |
| Abbar 2022 | 73 | 6.17 | 8.70 | 83 | 14.33 | 10.11 | -0.86 | [-1.19; -0.53] | 11.9% | 6.3% |
| Canuso 2018 | 35 | 4.26 | 2.09 | 31 | 4.81 | 1.70 | -0.28 | [-0.77; 0.20] | 5.4% | 5.5% |
| Fineberg 2023 | 10 | 4.40 | 5.23 | 12 | 3.42 | 5.71 | 0.17 | [-0.67; 1.01] | 1.8% | 3.8% |
| Fu 2020 | 110 | 2.49 | 1.65 | 112 | 2.74 | 1.44 | -0.16 | [-0.42; 0.10] | 18.5% | 6.6% |
| George 2017 | 12 | 2.17 | 1.53 | 4 | 1.75 | 1.50 | 0.26 | [-0.88; 1.40] | 1.0% | 2.7% |
| Glue 2024b | 17 | 0.41 | 0.71 | 8 | 0.25 | 0.46 | 0.24 | [-0.60; 1.08] | 1.8% | 3.8% |
| Grunebaum 2017 | 7 | 5.71 | 6.10 | 9 | 5.89 | 5.58 | -0.03 | [-1.02; 0.96] | 1.3% | 3.2% |
| Grunebaum 2018 | 40 | 5.00 | 5.18 | 40 | 9.85 | 7.21 | -0.77 | [-1.22; -0.31] | 6.2% | 5.7% |
| Ionescu 2019 | 13 | 1.90 | 1.40 | 13 | 1.50 | 1.40 | 0.28 | [-0.50; 1.05] | 2.2% | 4.1% |
| Ionescu 2020 | 113 | 2.25 | 1.47 | 112 | 2.92 | 1.32 | -0.48 | [-0.74; -0.21] | 18.3% | 6.6% |
| Jones 2024 | 17 | 8.50 | 7.99 | 11 | 14.20 | 8.63 | -0.67 | [-1.45; 0.11] | 2.1% | 4.0% |
| Loo 2016 | 9 | 1.44 | 1.31 | 6 | 0.83 | 0.75 | 0.51 | [-0.55; 1.56] | 1.2% | 3.0% |
| Singh 2016 2x doses | 18 | 0.11 | 0.32 | 16 | 0.75 | 1.18 | -0.74 | [-1.44; -0.04] | 2.6% | 4.4% |
| Singh 2016 3x doses | 17 | 0.12 | 0.49 | 16 | 0.50 | 1.03 | -0.46 | [-1.16; 0.23] | 2.7% | 4.5% |
| Su 2023 | 42 | 1.57 | 1.41 | 42 | 2.36 | 1.23 | -0.59 | [-1.03; -0.16] | 6.7% | 5.8% |
| Sumner 2019 | 17 | 0.41 | 0.87 | 16 | 1.00 | 0.89 | -0.65 | [-1.36; 0.05] | 2.6% | 4.4% |
| Zarate 2006 | 9 | 0.12 | 0.35 | 9 | 0.56 | 0.73 | -0.72 | [-1.68; 0.24] | 1.4% | 3.3% |
| Zarate 2012 | 19 | 1.10 | 1.24 | 22 | 1.59 | 1.22 | -0.39 | [-1.01; 0.23] | 3.3% | 4.8% |
| Common effect model | **578** | | | **562** | | | **-0.42** | **[-0.54; -0.31]** | **91.1%** | **--** |
| Random effects model | | | | | | | **-0.38** | **[-0.59; -0.17]** | **--** | **82.5%** |
| Heterogeneity: $I^2 = 40\%$, $\tau^2 = 0.0991$, $p = 0.04$ | | | | | | | | | | |
| scale_mode = self | | | | | | | | | | |
| Ahmed 2023 | 18 | 83.89 | 19.55 | 18 | 80.56 | 13.02 | 0.20 | [-0.46; 0.85] | 3.0% | 4.6% |
| Burger 2016 | 3 | 9.40 | 11.75 | 7 | 22.70 | 11.65 | -1.03 | [-2.50; 0.44] | 0.6% | 1.9% |
| Domany 2020 | 9 | 13.10 | 14.20 | 9 | 12.30 | 14.70 | 0.05 | [-0.87; 0.98] | 1.5% | 3.4% |
| Domany 2021 | 15 | 8.60 | 9.30 | 15 | 13.60 | 8.60 | -0.54 | [-1.27; 0.19] | 2.4% | 4.3% |
| Hu 2016 | 13 | 0.30 | 0.50 | 14 | 1.30 | 0.40 | -2.15 | [-3.13; -1.18] | 1.4% | 3.2% |
| Common effect model | **58** | | | **63** | | | **-0.47** | **[-0.85; -0.09]** | **8.9%** | **--** |
| Random effects model | | | | | | | **-0.64** | **[-1.48; 0.20]** | **--** | **17.5%** |
| Heterogeneity: $I^2 = 77\%$, $\tau^2 = 0.6911$, $p < 0.01$ | | | | | | | | | | |
| **Common effect model** | **636** | | | **625** | | | **-0.43** | **[-0.54; -0.31]** | **100.0%** | **--** |
| **Random effects model** | | | | | | | **-0.40** | **[-0.63; -0.16]** | **--** | **100.0%** |

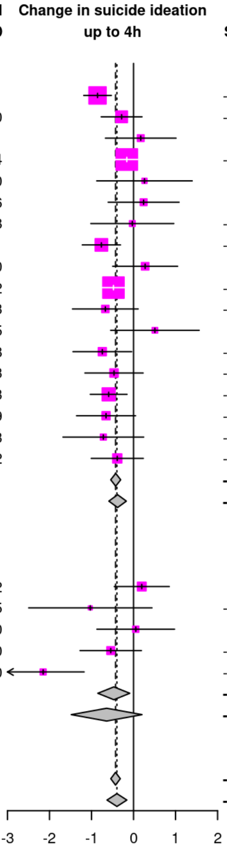

Heterogeneity: $I^2 = 52\%$, $\tau^2 = 0.2017$, $p < 0.01$
Test for subgroup differences (common effect): $\chi^2_1 = 0.05$, df = 1 ($p = 0.82$)
RE model$\chi^2_1 = 0.35$, df = 1 ($p = 0.56$)

## >12h up to 24h

| Study | Experimental Total | Experimental Mean | Experimental SD | Control Total | Control Mean | Control SD | SMD | 95%-CI | Weight (common) | Weight (random) |
|---|---|---|---|---|---|---|---|---|---|---|
| **scale_mode = clinician** | | | | | | | | | | |
| Abbar 2022 | 73 | 5.77 | 9.68 | 83 | 11.97 | 10.41 | -0.61 | [-0.93; -0.29] | 10.8% | 7.8% |
| Canuso 2018 | 35 | 3.29 | 2.47 | 30 | 4.47 | 1.57 | -0.55 | [-1.05; -0.06] | 4.5% | 5.3% |
| Fineberg 2023 | 10 | 3.10 | 5.63 | 11 | 3.18 | 5.67 | -0.01 | [-0.87; 0.84] | 1.5% | 2.5% |
| Fu 2020 | 110 | 2.23 | 1.67 | 112 | 2.43 | 1.63 | -0.12 | [-0.38; 0.14] | 16.1% | 8.8% |
| Grunebaum 2017 | 7 | 4.29 | 5.74 | 9 | 10.11 | 5.75 | -0.96 | [-2.02; 0.10] | 1.0% | 1.8% |
| Grunebaum 2018 | 40 | 6.05 | 5.48 | 40 | 11.63 | 7.43 | -0.85 | [-1.30; -0.39] | 5.3% | 5.7% |
| Ionescu 2020 | 110 | 2.22 | 1.51 | 110 | 2.44 | 1.57 | -0.14 | [-0.41; 0.12] | 16.0% | 8.7% |
| Jones 2024 | 17 | 10.80 | 8.08 | 10 | 12.30 | 7.83 | -0.18 | [-0.96; 0.60] | 1.8% | 2.9% |
| Lii 2023 | 20 | 0.20 | 0.52 | 19 | 0.21 | 0.54 | -0.02 | [-0.65; 0.61] | 2.8% | 4.0% |
| Loo 2016 | 9 | 1.78 | 1.20 | 6 | 1.33 | 1.03 | 0.37 | [-0.67; 1.42] | 1.0% | 1.8% |
| Murrough 2013 | 26 | 0.85 | 1.12 | 26 | 1.46 | 1.33 | -0.49 | [-1.04; 0.06] | 3.7% | 4.7% |
| Pattanaseri 2024 | 11 | 1.64 | 1.57 | 9 | 2.11 | 1.16 | -0.32 | [-1.21; 0.57] | 1.4% | 2.4% |
| Price 2022 | 53 | 0.38 | 0.98 | 49 | 0.59 | 1.31 | -0.18 | [-0.57; 0.21] | 7.4% | 6.7% |
| Singh 2016 II | 18 | 0.61 | 1.42 | 8 | 0.88 | 1.64 | -0.18 | [-1.01; 0.66] | 1.6% | 2.6% |
| Sos 2013 | 9 | 0.56 | 0.73 | 18 | 0.39 | 0.59 | 0.26 | [-0.55; 1.06] | 1.7% | 2.8% |
| Su 2023 | 42 | 1.48 | 1.42 | 42 | 2.10 | 1.21 | -0.47 | [-0.90; -0.03] | 6.0% | 6.1% |
| Sumner 2019 | 17 | 0.65 | 0.79 | 16 | 1.31 | 0.95 | -0.74 | [-1.45; -0.03] | 2.2% | 3.4% |
| Zarate 2006 | 9 | 0.44 | 0.53 | 9 | 0.78 | 0.83 | -0.46 | [-1.40; 0.48] | 1.3% | 2.2% |
| Zarate 2012 | 20 | 1.30 | 1.38 | 23 | 2.00 | 1.24 | -0.53 | [-1.14; 0.09] | 3.0% | 4.1% |
| Zhou 2024 | 27 | -2.40 | 2.20 | 27 | -1.50 | 2.00 | -0.42 | [-0.96; 0.12] | 3.8% | 4.8% |
| **Common effect model** | **663** | | | **657** | | | **-0.33** | **[-0.44; -0.22]** | **93.1%** | **--** |
| **Random effects model** | | | | | | | **-0.35** | **[-0.51; -0.19]** | **--** | **89.0%** |
| Heterogeneity: $I^2 = 17\%$, $\tau^2 = 0.0519$, $p = 0.24$ | | | | | | | | | | |
| **scale_mode = self** | | | | | | | | | | |
| Domany 2020 | 9 | 9.20 | 14.80 | 9 | 11.90 | 16.40 | -0.16 | [-1.09; 0.76] | 1.3% | 2.2% |
| Domany 2021 | 15 | 7.80 | 10.50 | 15 | 11.50 | 10.80 | -0.34 | [-1.06; 0.38] | 2.2% | 3.3% |
| Hu 2016 | 13 | 0.70 | 0.70 | 14 | 1.30 | 0.40 | -1.03 | [-1.84; -0.22] | 1.7% | 2.7% |
| Murrough 2015 II | 12 | 10.83 | 8.48 | 12 | 14.00 | 10.15 | -0.33 | [-1.13; 0.48] | 1.7% | 2.8% |
| **Common effect model** | **49** | | | **50** | | | **-0.47** | **[-0.88; -0.07]** | **6.9%** | **--** |
| **Random effects model** | | | | | | | **-0.47** | **[-0.94; -0.01]** | **--** | **11.0%** |
| Heterogeneity: $I^2 = 0\%$, $\tau^2 = 0.0570$, $p = 0.47$ | | | | | | | | | | |
| **Common effect model** | **712** | | | **707** | | | **-0.34** | **[-0.44; -0.23]** | **100.0%** | **--** |
| **Random effects model** | | | | | | | **-0.36** | **[-0.52; -0.21]** | **--** | **100.0%** |

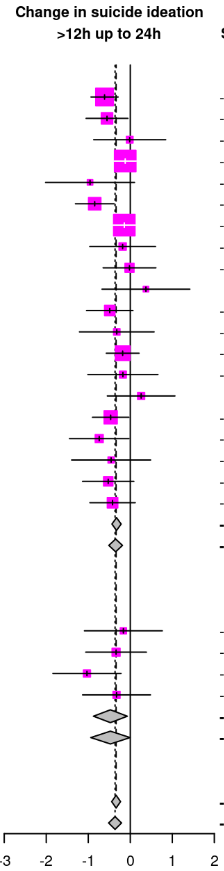

Heterogeneity: $I^2 = 11\%$, $\tau^2 = 0.0519$, $p = 0.30$

Test for subgroup differences (common effect): $\chi^2_1 = 0.47$, df = 1 ($p = 0.49$)

RE model$\chi^2_1 = 0.24$, df = 1 ($p = 0.62$)

## >24h up to 72h

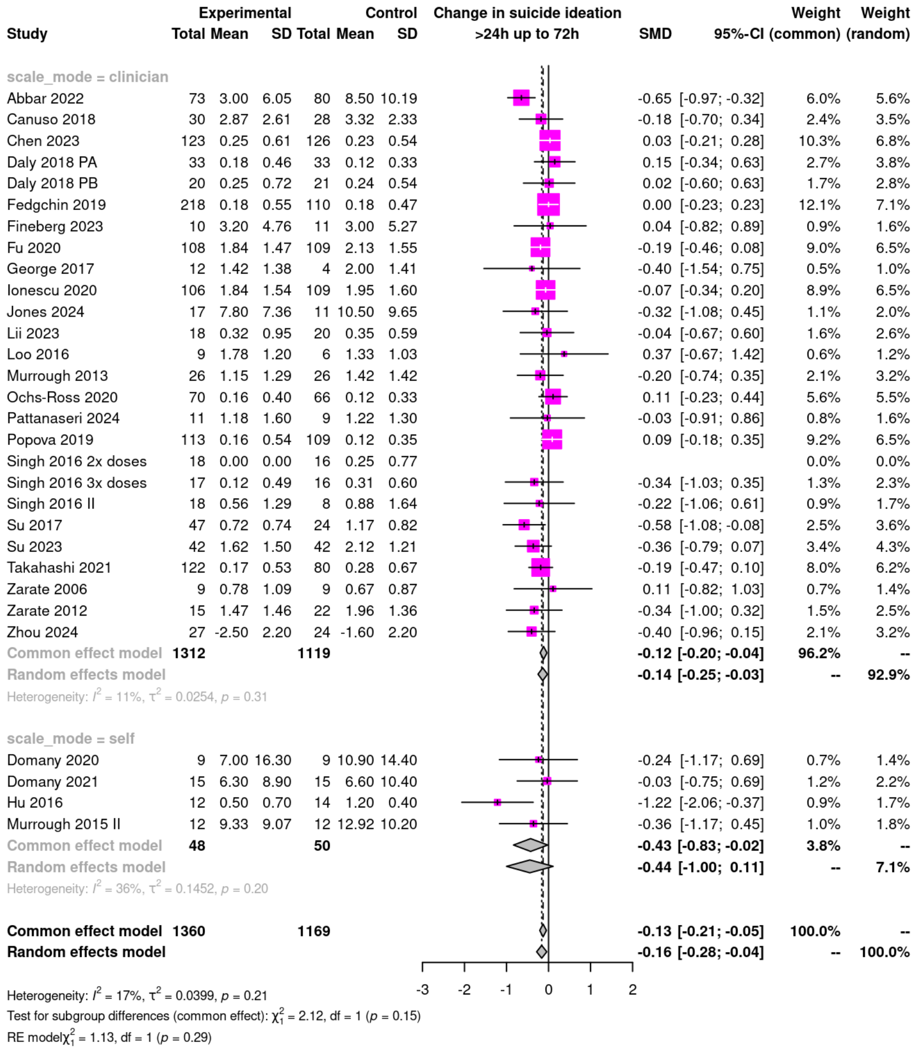
Study
Experimental Total Mean SD
Control Total Mean SD
Change in suicide ideation >24h up to 72h
SMD 95%-CI
Weight (common)
Weight (random)
scale_mode = clinician
Abbar 2022 73 3.00 6.05 80 8.50 10.19 -0.65 [-0.97; -0.32] 6.0% 5.6%
Canuso 2018 30 2.87 2.61 28 3.32 2.33 -0.18 [-0.70; 0.34] 2.4% 3.5%
Chen 2023 123 0.25 0.61 126 0.23 0.54 0.03 [-0.21; 0.28] 10.3% 6.8%
Daly 2018 PA 33 0.18 0.46 33 0.12 0.33 0.15 [-0.34; 0.63] 2.7% 3.8%
Daly 2018 PB 20 0.25 0.72 21 0.24 0.54 0.02 [-0.60; 0.63] 1.7% 2.8%
Fedgchin 2019 218 0.18 0.55 110 0.18 0.47 0.00 [-0.23; 0.23] 12.1% 7.1%
Fineberg 2023 10 3.20 4.76 11 3.00 5.27 0.04 [-0.82; 0.89] 0.9% 1.6%
Fu 2020 108 1.84 1.47 109 2.13 1.55 -0.19 [-0.46; 0.08] 9.0% 6.5%
George 2017 12 1.42 1.38 4 2.00 1.41 -0.40 [-1.54; 0.75] 0.5% 1.0%
Ionescu 2020 106 1.84 1.54 109 1.95 1.60 -0.07 [-0.34; 0.20] 8.9% 6.5%
Jones 2024 17 7.80 7.36 11 10.50 9.65 -0.32 [-1.08; 0.45] 1.1% 2.0%
Lii 2023 18 0.32 0.95 20 0.35 0.59 -0.04 [-0.67; 0.60] 1.6% 2.6%
Loo 2016 9 1.78 1.20 6 1.33 1.03 0.37 [-0.67; 1.42] 0.6% 1.2%
Murrough 2013 26 1.15 1.29 26 1.42 1.42 -0.20 [-0.74; 0.35] 2.1% 3.2%
Ochs-Ross 2020 70 0.16 0.40 66 0.12 0.33 0.11 [-0.23; 0.44] 5.6% 5.5%
Pattanaseri 2024 11 1.18 1.60 9 1.22 1.30 -0.03 [-0.91; 0.86] 0.8% 1.6%
Popova 2019 113 0.16 0.54 109 0.12 0.35 0.09 [-0.18; 0.35] 9.2% 6.5%
Singh 2016 2x doses 18 0.00 0.00 16 0.25 0.77 0.0% 0.0%
Singh 2016 3x doses 17 0.12 0.49 16 0.31 0.60 -0.34 [-1.03; 0.35] 1.3% 2.3%
Singh 2016 II 18 0.56 1.29 8 0.88 1.64 -0.22 [-1.06; 0.61] 0.9% 1.7%
Su 2017 47 0.72 0.74 24 1.17 0.82 -0.58 [-1.08; -0.08] 2.5% 3.6%
Su 2023 42 1.62 1.50 42 2.12 1.21 -0.36 [-0.79; 0.07] 3.4% 4.3%
Takahashi 2021 122 0.17 0.53 80 0.28 0.67 -0.19 [-0.47; 0.10] 8.0% 6.2%
Zarate 2006 9 0.78 1.09 9 0.67 0.87 0.11 [-0.82; 1.03] 0.7% 1.4%
Zarate 2012 15 1.47 1.46 22 1.96 1.36 -0.34 [-1.00; 0.32] 1.5% 2.5%
Zhou 2024 27 -2.50 2.20 24 -1.60 2.20 -0.40 [-0.96; 0.15] 2.1% 3.2%
Common effect model 1312 1119 -0.12 [-0.20; -0.04] 96.2% --
Random effects model -0.14 [-0.25; -0.03] -- 92.9%
Heterogeneity: $I^2$ = 11%, $\tau^2$ = 0.0254, $p$ = 0.31
scale_mode = self
Domany 2020 9 7.00 16.30 9 10.90 14.40 -0.24 [-1.17; 0.69] 0.7% 1.4%
Domany 2021 15 6.30 8.90 15 6.60 10.40 -0.03 [-0.75; 0.69] 1.2% 2.2%
Hu 2016 12 0.50 0.70 14 1.20 0.40 -1.22 [-2.06; -0.37] 0.9% 1.7%
Murrough 2015 II 12 9.33 9.07 12 12.92 10.20 -0.36 [-1.17; 0.45] 1.0% 1.8%
Common effect model 48 50 -0.43 [-0.83; -0.02] 3.8% --
Random effects model -0.44 [-1.00; 0.11] -- 7.1%
Heterogeneity: $I^2$ = 36%, $\tau^2$ = 0.1452, $p$ = 0.20
Common effect model 1360 1169 -0.13 [-0.21; -0.05] 100.0% --
Random effects model -0.16 [-0.28; -0.04] -- 100.0%
-3 -2 -1 0 1 2
Heterogeneity: $I^2$ = 17%, $\tau^2$ = 0.0399, $p$ = 0.21
Test for subgroup differences (common effect): $\chi^2_1$ = 2.12, df = 1 ($p$ = 0.15)
RE model$\chi^2_1$ = 1.13, df = 1 ($p$ = 0.29)

## >72h up to 2w

| Study | Experimental Total | Experimental Mean | Experimental SD | Control Total | Control Mean | Control SD | SMD | 95%-CI | Weight (common) | Weight (random) |
|---|---|---|---|---|---|---|---|---|---|---|
| **scale_mode = clinician** | | | | | | | | | | |
| Abbar 2022 | 73 | 3.50 | 7.18 | 80 | 8.00 | 11.30 | -0.47 | [-0.79; -0.15] | 5.8% | 5.3% |
| Anderson 2017 | 35 | 2.94 | 1.76 | 37 | 3.08 | 2.38 | -0.07 | [-0.53; 0.40] | 2.8% | 3.4% |
| Canuso 2018 | 29 | 1.76 | 2.03 | 26 | 1.54 | 1.68 | 0.12 | [-0.41; 0.65] | 2.1% | 2.8% |
| Chen 2023 | 117 | 0.25 | 0.60 | 111 | 0.23 | 0.61 | 0.03 | [-0.23; 0.29] | 8.9% | 6.4% |
| Daly 2018 PA | 40 | 0.20 | 0.55 | 32 | 0.22 | 0.42 | -0.04 | [-0.50; 0.43] | 2.8% | 3.4% |
| Daly 2018 PB | 20 | 0.15 | 0.49 | 21 | 0.38 | 0.59 | -0.41 | [-1.03; 0.20] | 1.6% | 2.2% |
| Fedgchin 2019 | 211 | 0.16 | 0.46 | 110 | 0.18 | 0.39 | -0.05 | [-0.28; 0.18] | 11.3% | 6.9% |
| Fineberg 2023 | 10 | 4.30 | 7.10 | 12 | 5.67 | 7.04 | -0.19 | [-1.03; 0.65] | 0.8% | 1.3% |
| Fu 2020 | 105 | 1.28 | 1.31 | 99 | 1.33 | 1.14 | -0.04 | [-0.32; 0.23] | 8.0% | 6.1% |
| George 2017 | 12 | 1.25 | 1.21 | 4 | 2.50 | 1.73 | -0.88 | [-2.07; 0.30] | 0.4% | 0.7% |
| Ionescu 2019 | 13 | 1.30 | 1.40 | 13 | 1.50 | 1.40 | -0.14 | [-0.91; 0.63] | 1.0% | 1.6% |
| Ionescu 2020 | 92 | 0.97 | 1.19 | 100 | 1.35 | 1.34 | -0.30 | [-0.58; -0.01] | 7.4% | 5.9% |
| Lii 2023 | 19 | 0.32 | 0.58 | 20 | 0.40 | 0.77 | -0.11 | [-0.74; 0.51] | 1.5% | 2.2% |
| Loo 2016 | 9 | 2.00 | 1.66 | 5 | 1.60 | 0.89 | 0.26 | [-0.84; 1.36] | 0.5% | 0.8% |
| Ochs-Ross 2020 | 65 | 0.21 | 0.65 | 62 | 0.18 | 0.58 | 0.05 | [-0.30; 0.40] | 5.0% | 4.8% |
| Pattanaseri 2024 | 11 | 1.64 | 1.75 | 9 | 1.44 | 1.33 | 0.12 | [-0.76; 1.00] | 0.8% | 1.2% |
| Popova 2019 | 109 | 0.08 | 0.36 | 107 | 0.18 | 0.53 | -0.22 | [-0.49; 0.05] | 8.4% | 6.2% |
| Price 2022 | 52 | 0.79 | 1.32 | 46 | 1.30 | 1.62 | -0.34 | [-0.74; 0.05] | 3.8% | 4.1% |
| Singh 2016 2x doses | 15 | 0.07 | 0.26 | 12 | 0.33 | 0.65 | -0.53 | [-1.31; 0.24] | 1.0% | 1.6% |
| Singh 2016 3x doses | 15 | 0.00 | 0.00 | 12 | 0.25 | 0.45 | | | 0.0% | 0.0% |
| Sos 2013 | 9 | 0.44 | 0.73 | 18 | 0.17 | 0.87 | 0.32 | [-0.49; 1.12] | 0.9% | 1.5% |
| Su 2017 | 47 | 0.98 | 0.87 | 24 | 1.29 | 0.81 | -0.36 | [-0.86; 0.13] | 2.4% | 3.1% |
| Su 2023 | 42 | 1.95 | 1.36 | 42 | 2.12 | 1.33 | -0.13 | [-0.55; 0.30] | 3.3% | 3.8% |
| Sumner 2019 | 17 | 0.94 | 0.97 | 16 | 1.63 | 0.96 | -0.70 | [-1.40; 0.01] | 1.2% | 1.8% |
| Takahashi 2021 | 116 | 0.18 | 0.61 | 74 | 0.27 | 0.76 | -0.13 | [-0.43; 0.16] | 7.1% | 5.8% |
| Zarate 2006 | 9 | 0.89 | 1.27 | 9 | 1.00 | 0.82 | -0.10 | [-1.02; 0.83] | 0.7% | 1.1% |
| Zarate 2012 | 11 | 2.09 | 1.38 | 17 | 2.12 | 1.05 | -0.02 | [-0.78; 0.74] | 1.0% | 1.6% |
| Zhou 2024 | 27 | -2.80 | 2.10 | 23 | -2.20 | 2.50 | -0.26 | [-0.82; 0.30] | 1.9% | 2.6% |
| Common effect model | **1330** | | | **1141** | | | **-0.15** | **[-0.23; -0.07]** | **92.5%** | **--** |
| Random effects model | | | | | | | **-0.16** | **[-0.27; -0.05]** | **--** | **88.5%** |
| Heterogeneity: $I^2$ = 0%, $\tau^2$ = 0.0252, $p$ = 0.77 | | | | | | | | | | |
| **scale_mode = self** | | | | | | | | | | |
| Ahmed 2023 | 18 | 75.72 | 20.58 | 18 | 80.67 | 13.15 | -0.28 | [-0.94; 0.38] | 1.4% | 2.0% |
| Burger 2016 | 3 | 2.40 | 13.61 | 7 | 6.70 | 8.93 | -0.38 | [-1.75; 0.99] | 0.3% | 0.6% |
| Colla 2024 | 17 | 1.18 | 0.87 | 10 | 1.60 | 0.95 | -0.45 | [-1.24; 0.34] | 1.0% | 1.5% |
| Domany 2020 | 9 | 7.40 | 18.00 | 9 | 1.40 | 2.20 | 0.45 | [-0.49; 1.38] | 0.7% | 1.1% |
| Domany 2021 | 15 | 3.10 | 14.10 | 15 | 11.40 | 15.20 | -0.55 | [-1.28; 0.18] | 1.1% | 1.7% |
| Duek 2023 | 14 | 0.14 | 0.36 | 13 | 0.08 | 0.28 | 0.20 | [-0.56; 0.95] | 1.0% | 1.6% |
| Grabski 2022 | 41 | 0.10 | 0.30 | 44 | 0.00 | 0.00 | | | 0.0% | 0.0% |
| Hu 2016 | 12 | 0.90 | 0.90 | 14 | 1.00 | 0.50 | -0.14 | [-0.91; 0.64] | 1.0% | 1.6% |
| Murrough 2015 II | 12 | 7.92 | 8.73 | 12 | 10.08 | 9.39 | -0.23 | [-1.03; 0.57] | 0.9% | 1.5% |
| Common effect model | **141** | | | **142** | | | **-0.19** | **[-0.47; 0.10]** | **7.5%** | **--** |
| Random effects model | | | | | | | **-0.18** | **[-0.50; 0.13]** | **--** | **11.5%** |
| Heterogeneity: $I^2$ = 0%, $\tau^2$ = 0.0392, $p$ = 0.74 | | | | | | | | | | |
| **Common effect model** | **1471** | | | **1283** | | | **-0.15** | **[-0.23; -0.07]** | **100.0%** | **--** |
| **Random effects model** | | | | | | | **-0.16** | **[-0.27; -0.06]** | **--** | **100.0%** |

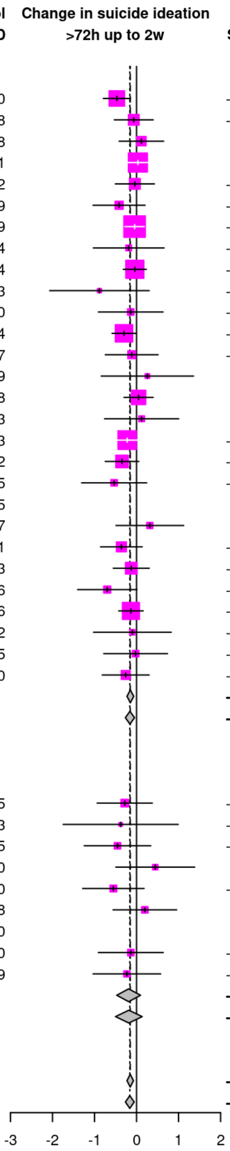

Heterogeneity: $I^2$ = 0%, $\tau^2$ = 0.0271, $p$ = 0.87
Test for subgroup differences (common effect): $\chi^2_1$ = 0.07, df = 1 ($p$ = 0.79)
RE model$\chi^2_1$ = 0.02, df = 1 ($p$ = 0.88)

## >2w up to 4w

| Study | Experimental Total | Experimental Mean | Experimental SD | Control Total | Control Mean | Control SD | SMD | 95%-CI | Weight (common) | Weight (random) |
|---|---|---|---|---|---|---|---|---|---|---|
| scale_mode = clinician | | | | | | | | | | |
| Abbar 2022 | 73 | 2.83 | 5.67 | 80 | 6.43 | 9.66 | -0.45 | [-0.77; -0.13] | 7.9% | 7.8% |
| Canuso 2018 | 26 | 1.00 | 1.44 | 21 | 0.90 | 1.37 | 0.07 | [-0.51; 0.64] | 2.5% | 4.2% |
| Chen 2023 | 108 | 0.26 | 0.62 | 107 | 0.21 | 0.53 | 0.09 | [-0.18; 0.35] | 11.4% | 8.8% |
| Fedgchin 2019 | 204 | 0.11 | 0.37 | 107 | 0.17 | 0.40 | -0.16 | [-0.39; 0.08] | 14.9% | 9.5% |
| Fineberg 2023 | 10 | 7.10 | 6.01 | 11 | 4.73 | 5.75 | 0.39 | [-0.48; 1.25] | 1.1% | 2.3% |
| Fu 2020 | 100 | 0.97 | 1.27 | 93 | 1.08 | 1.21 | -0.09 | [-0.37; 0.19] | 10.2% | 8.5% |
| Ionescu 2019 | 12 | 1.20 | 1.40 | 12 | 1.40 | 1.40 | -0.14 | [-0.94; 0.66] | 1.3% | 2.6% |
| Ionescu 2020 | 90 | 0.68 | 1.04 | 92 | 0.93 | 1.18 | -0.22 | [-0.52; 0.07] | 9.6% | 8.4% |
| Ochs-Ross 2020 | 62 | 0.18 | 0.51 | 61 | 0.17 | 0.45 | 0.02 | [-0.33; 0.37] | 6.5% | 7.2% |
| Popova 2019 | 103 | 0.07 | 0.29 | 102 | 0.08 | 0.31 | -0.03 | [-0.31; 0.24] | 10.9% | 8.7% |
| Price 2022 | 49 | 0.61 | 1.27 | 41 | 1.68 | 1.71 | -0.71 | [-1.14; -0.29] | 4.4% | 6.0% |
| Su 2017 | 47 | 1.00 | 0.75 | 24 | 1.33 | 1.01 | -0.39 | [-0.88; 0.11] | 3.3% | 5.1% |
| Takahashi 2021 | 111 | 0.15 | 0.65 | 72 | 0.14 | 0.54 | 0.02 | [-0.28; 0.31] | 9.3% | 8.3% |
| Zhou 2024 | 26 | -2.90 | 2.20 | 23 | -1.90 | 2.30 | -0.44 | [-1.01; 0.13] | 2.5% | 4.3% |
| Common effect model | **1021** | | | **846** | | | **-0.14** | **[-0.23; -0.05]** | **95.8%** | **--** |
| Random effects model | | | | | | | **-0.15** | **[-0.30; -0.01]** | **--** | **91.7%** |
| Heterogeneity: $I^2$ = 35%, $\tau^2$ = 0.0406, $p$ = 0.10 | | | | | | | | | | |
| scale_mode = self | | | | | | | | | | |
| Domany 2021 | 15 | 3.00 | 14.50 | 15 | 8.30 | 16.00 | -0.34 | [-1.06; 0.38] | 1.6% | 3.0% |
| Duek 2023 | 13 | 0.31 | 0.48 | 11 | 0.18 | 0.41 | 0.27 | [-0.54; 1.08] | 1.3% | 2.6% |
| Grabski 2022 | 39 | 0.05 | 0.22 | 43 | 0.00 | 0.00 | | | 0.0% | 0.0% |
| Hu 2016 | 12 | 0.40 | 0.90 | 14 | 0.20 | 0.40 | 0.29 | [-0.49; 1.06] | 1.4% | 2.7% |
| Common effect model | **79** | | | **83** | | | **0.05** | **[-0.39; 0.49]** | **4.2%** | **--** |
| Random effects model | | | | | | | **0.05** | **[-0.45; 0.56]** | **--** | **8.3%** |
| Heterogeneity: $I^2$ = 0%, $\tau^2$ = 0.0468, $p$ = 0.42 | | | | | | | | | | |
| **Common effect model** | **1100** | | | **929** | | | **-0.13** | **[-0.22; -0.04]** | **100.0%** | **--** |
| **Random effects model** | | | | | | | **-0.14** | **[-0.28; 0.01]** | **--** | **100.0%** |

Change in suicide ideation >2w up to 4w

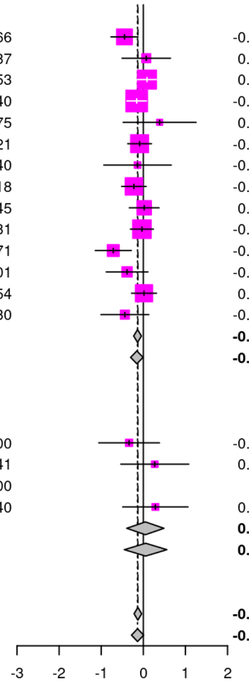

Heterogeneity: $I^2$ = 29%, $\tau^2$ = 0.0430, $p$ = 0.13
Test for subgroup differences (common effect): $\chi^2_1$ = 0.64, df = 1 ($p$ = 0.42)
RE model$\chi^2_1$ = 0.60, df = 1 ($p$ = 0.44)

**>4w**

| Study | Experimental Total | Experimental Mean | Experimental SD | Control Total | Control Mean | Control SD | SMD | 95%-CI | Weight (common) | Weight (random) |
|---|---|---|---|---|---|---|---|---|---|---|
| **scale_mode = clinician** | | | | | | | | | | |
| Abbar 2022 | 60 | 2.60 | 5.90 | 66 | 4.00 | 8.30 | -0.19 | [-0.54; 0.16] | 18.0% | 17.7% |
| Fu 2020 | 84 | 1.04 | 1.31 | 79 | 0.84 | 1.20 | 0.16 | [-0.15; 0.47] | 23.4% | 21.6% |
| Ionescu 2020 | 80 | 0.49 | 1.08 | 86 | 0.66 | 0.98 | -0.16 | [-0.47; 0.14] | 23.8% | 21.8% |
| Pattanaseri 2024 | 11 | 1.64 | 1.50 | 9 | 1.89 | 1.27 | -0.17 | [-1.05; 0.71] | 2.8% | 3.5% |
| Price 2022 | 36 | 0.56 | 0.97 | 35 | 0.60 | 0.85 | -0.04 | [-0.51; 0.42] | 10.2% | 11.2% |
| Zhou 2024 | 26 | -2.80 | 2.10 | 23 | -2.10 | 2.30 | -0.31 | [-0.88; 0.25] | 6.9% | 8.0% |
| **Common effect model** | **297** | | | **298** | | | **-0.08** | **[-0.24; 0.08]** | **85.2%** | **--** |
| **Random effects model** | | | | | | | **-0.09** | **[-0.27; 0.10]** | **--** | **83.7%** |
| Heterogeneity: $I^2 = 0\%$, $\tau^2 = 0.0095$, $p = 0.59$ | | | | | | | | | | |
| **scale_mode = self** | | | | | | | | | | |
| Duek 2023 | 11 | 0.30 | 0.48 | 11 | 0.20 | 0.42 | 0.21 | [-0.63; 1.05] | 3.1% | 3.8% |
| Grabski 2022 | 38 | 0.05 | 0.23 | 43 | 0.07 | 0.26 | -0.07 | [-0.51; 0.37] | 11.6% | 12.5% |
| **Common effect model** | **49** | | | **54** | | | **-0.01** | **[-0.40; 0.38]** | **14.8%** | **--** |
| **Random effects model** | | | | | | | **-0.01** | **[-0.41; 0.40]** | **--** | **16.3%** |
| Heterogeneity: $I^2 = 0\%$, $\tau^2 = 0.0058$, $p = 0.56$ | | | | | | | | | | |
| **Common effect model** | **346** | | | **352** | | | **-0.07** | **[-0.22; 0.08]** | **100.0%** | **--** |
| **Random effects model** | | | | | | | **-0.07** | **[-0.24; 0.10]** | **--** | **100.0%** |

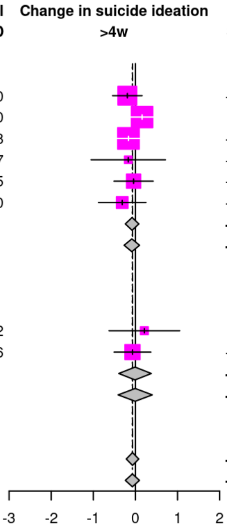

Heterogeneity: $I^2 = 0\%$, $\tau^2 = 0.0095$, $p = 0.76$

Test for subgroup differences (common effect): $\chi^2_1 = 0.11$, df = 1 ($p = 0.74$)

RE model$\chi^2_1 = 0.12$, df = 1 ($p = 0.73$)

#### End of Treatment

| Study | Experimental Total | Experimental Mean | Experimental SD | Control Total | Control Mean | Control SD | SMD | 95%-CI | Weight (common) | Weight (random) |
|---|---|---|---|---|---|---|---|---|---|---|
| scale_mode = clinician | | | | | | | | | | |
| Abbar 2022 (day 2) | 73 | 3.00 | 6.05 | 80 | 8.50 | 10.19 | -0.65 | [-0.97; -0.32] | 5.6% | 4.7% |
| Canuso 2018 (25d) | 26 | 1.00 | 1.44 | 21 | 0.90 | 1.37 | 0.07 | [-0.51; 0.64] | 1.8% | 2.6% |
| Chen 2023 (4w) | 108 | 0.26 | 0.62 | 107 | 0.21 | 0.53 | 0.09 | [-0.18; 0.35] | 8.4% | 5.4% |
| Daly 2018 PA (7d) | 40 | 0.20 | 0.55 | 32 | 0.22 | 0.42 | -0.04 | [-0.50; 0.43] | 2.8% | 3.4% |
| Daly 2018 PB (7d) | 20 | 0.15 | 0.49 | 21 | 0.38 | 0.59 | -0.41 | [-1.03; 0.20] | 1.6% | 2.4% |
| Fedgchin 2019 (4w) | 204 | 0.11 | 0.37 | 107 | 0.17 | 0.40 | -0.16 | [-0.39; 0.08] | 10.9% | 5.8% |
| Fineberg 2023 (24h) | 10 | 3.10 | 5.63 | 11 | 3.18 | 5.67 | -0.01 | [-0.87; 0.84] | 0.8% | 1.5% |
| Fu 2020 (25d) | 100 | 0.97 | 1.27 | 93 | 1.08 | 1.21 | -0.09 | [-0.37; 0.19] | 7.5% | 5.2% |
| George 2017 (1w) | 12 | 1.25 | 1.21 | 4 | 2.50 | 1.73 | -0.88 | [-2.07; 0.30] | 0.4% | 0.8% |
| Grunebaum 2017 (24h) | 7 | 4.29 | 5.74 | 9 | 10.11 | 5.75 | -0.96 | [-2.02; 0.10] | 0.5% | 1.0% |
| Grunebaum 2018 (24h) | 40 | 6.05 | 5.48 | 40 | 11.63 | 7.43 | -0.85 | [-1.30; -0.39] | 2.8% | 3.4% |
| Ionescu 2019 (3w) | 12 | 1.20 | 1.40 | 12 | 1.40 | 1.40 | -0.14 | [-0.94; 0.66] | 0.9% | 1.6% |
| Ionescu 2020 (25d) | 90 | 0.68 | 1.04 | 92 | 0.93 | 1.18 | -0.22 | [-0.52; 0.07] | 7.0% | 5.1% |
| Jones 2024 (24h) | 17 | 10.80 | 8.08 | 10 | 12.30 | 7.83 | -0.18 | [-0.96; 0.60] | 1.0% | 1.7% |
| Lii 2023 (24h) | 20 | 0.20 | 0.52 | 19 | 0.21 | 0.54 | -0.02 | [-0.65; 0.61] | 1.5% | 2.3% |
| Loo 2016 (1w-2w) | 9 | 2.00 | 1.66 | 5 | 1.60 | 0.89 | 0.26 | [-0.84; 1.36] | 0.5% | 1.0% |
| Murrough 2013 (24h) | 26 | 0.85 | 1.12 | 26 | 1.46 | 1.33 | -0.49 | [-1.04; 0.06] | 2.0% | 2.8% |
| Ochs-Ross 2020 (4w) | 62 | 0.18 | 0.51 | 61 | 0.17 | 0.45 | 0.02 | [-0.33; 0.37] | 4.8% | 4.4% |
| Pattanaseri 2024 (3d) | 11 | 1.18 | 1.60 | 9 | 1.22 | 1.30 | -0.03 | [-0.91; 0.86] | 0.8% | 1.4% |
| Popova 2019 (4w) | 103 | 0.07 | 0.29 | 102 | 0.08 | 0.31 | -0.03 | [-0.31; 0.24] | 8.0% | 5.3% |
| Price 2022 (24h) | 53 | 0.38 | 0.98 | 49 | 0.59 | 1.31 | -0.18 | [-0.57; 0.21] | 3.9% | 4.1% |
| Singh 2016 2x doses (2w) | 15 | 0.07 | 0.26 | 12 | 0.33 | 0.65 | -0.53 | [-1.31; 0.24] | 1.0% | 1.7% |
| Singh 2016 3x doses (2w) | 15 | 0.00 | 0.00 | 12 | 0.25 | 0.45 | | | 0.0% | 0.0% |
| Singh 2016 II (4d) | 18 | 0.56 | 1.29 | 8 | 0.88 | 1.64 | -0.22 | [-1.06; 0.61] | 0.9% | 1.5% |
| Sos 2013 (24h) | 9 | 0.56 | 0.73 | . | 0.39 | 0.59 | | | 0.0% | 0.0% |
| Su 2023 (24h) | 42 | 1.48 | 1.42 | 42 | 2.10 | 1.21 | -0.47 | [-0.90; -0.03] | 3.2% | 3.6% |
| Sumner 2019 (24h) | 17 | 0.65 | 0.79 | 16 | 1.31 | 0.95 | -0.74 | [-1.45; -0.03] | 1.2% | 2.0% |
| Takahashi 2021 (4w) | 111 | 0.15 | 0.65 | 72 | 0.14 | 0.54 | 0.02 | [-0.28; 0.31] | 6.8% | 5.0% |
| Zarate 2006 (24h) | 9 | 0.44 | 0.53 | 9 | 0.78 | 0.83 | -0.46 | [-1.40; 0.48] | 0.7% | 1.3% |
| Zarate 2012 (24h) | 20 | 1.30 | 1.38 | 23 | 2.00 | 1.24 | -0.53 | [-1.14; 0.09] | 1.6% | 2.4% |
| Zhou 2024 (6d) | 27 | -2.60 | 2.00 | 25 | -1.70 | 2.20 | -0.42 | [-0.97; 0.13] | 2.0% | 2.8% |
| **Common effect model** | **1326** | | | **1129** | | | **-0.20** | **[-0.28; -0.12]** | **90.8%** | -- |
| **Random effects model** | | | | | | | **-0.24** | **[-0.37; -0.11]** | -- | **86.1%** |
| Heterogeneity: $I^2 = 28\%$, $\tau^2 = 0.0480$, $p = 0.08$ | | | | | | | | | | |
| scale_mode = self | | | | | | | | | | |
| Ahmed 2023 (2w) | 18 | 75.72 | 20.58 | 18 | 80.67 | 13.15 | -0.28 | [-0.94; 0.38] | 1.4% | 2.2% |
| Colla 2024 (2w) | 17 | 1.18 | 0.87 | 10 | 1.60 | 0.95 | -0.45 | [-1.24; 0.34] | 1.0% | 1.7% |
| Domany 2020 (24h) | 9 | 9.20 | 14.80 | 9 | 11.90 | 16.40 | -0.16 | [-1.09; 0.76] | 0.7% | 1.3% |
| Domany 2021 (24h) | 15 | 7.80 | 10.50 | 15 | 11.50 | 10.80 | -0.34 | [-1.06; 0.38] | 1.1% | 1.9% |
| Grabski 2022 (1-3w) | 38 | 0.05 | 0.23 | 43 | 0.07 | 0.26 | -0.07 | [-0.51; 0.37] | 3.1% | 3.6% |
| Hu 2016 (24h) | 13 | 0.70 | 0.70 | 14 | 1.30 | 0.40 | -1.03 | [-1.84; -0.22] | 0.9% | 1.6% |
| Murrough 2015 II (24h) | 12 | 10.83 | 8.48 | 12 | 14.00 | 10.15 | -0.33 | [-1.13; 0.48] | 0.9% | 1.6% |
| **Common effect model** | **122** | | | **121** | | | **-0.30** | **[-0.56; -0.05]** | **9.2%** | -- |
| **Random effects model** | | | | | | | **-0.33** | **[-0.63; -0.03]** | -- | **13.9%** |
| Heterogeneity: $I^2 = 0\%$, $\tau^2 = 0.0361$, $p = 0.62$ | | | | | | | | | | |
| **Common effect model** | **1448** | | | **1250** | | | **-0.21** | **[-0.28; -0.13]** | **100.0%** | -- |
| **Random effects model** | | | | | | | **-0.25** | **[-0.37; -0.14]** | -- | **100.0%** |

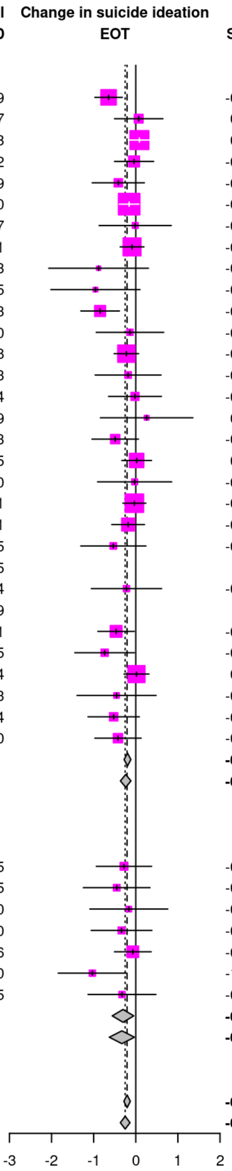

Heterogeneity: $I^2 = 20\%$, $\tau^2 = 0.0456$, $p = 0.15$
Test for subgroup differences (common effect): $\chi^2_1 = 0.62$, df = 1 ($p = 0.43$)
RE model$\chi^2_1 = 0.30$, df = 1 ($p = 0.58$)

#### Subgroup Analysis - Active vs. Non-Active Placebo

## Up to 4h

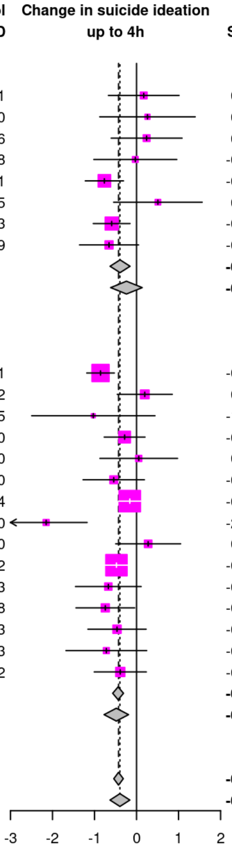

| Study | Experimental Total | Experimental Mean | Experimental SD | Control Total | Control Mean | Control SD | SMD | 95%-CI | Weight (common) | Weight (random) |
|---|---|---|---|---|---|---|---|---|---|---|
| actplac = active | | | | | | | | | | |
| Fineberg 2023 | 10 | 4.40 | 5.23 | 12 | 3.42 | 5.71 | 0.17 | [-0.67; 1.01] | 1.8% | 3.8% |
| George 2017 | 12 | 2.17 | 1.53 | 4 | 1.75 | 1.50 | 0.26 | [-0.88; 1.40] | 1.0% | 2.7% |
| Glue 2024b | 17 | 0.41 | 0.71 | 8 | 0.25 | 0.46 | 0.24 | [-0.60; 1.08] | 1.8% | 3.8% |
| Grunebaum 2017 | 7 | 5.71 | 6.10 | 9 | 5.89 | 5.58 | -0.03 | [-1.02; 0.96] | 1.3% | 3.2% |
| Grunebaum 2018 | 40 | 5.00 | 5.18 | 40 | 9.85 | 7.21 | -0.77 | [-1.22; -0.31] | 6.2% | 5.7% |
| Loo 2016 | 9 | 1.44 | 1.31 | 6 | 0.83 | 0.75 | 0.51 | [-0.55; 1.56] | 1.2% | 3.0% |
| Su 2023 | 42 | 1.57 | 1.41 | 42 | 2.36 | 1.23 | -0.59 | [-1.03; -0.16] | 6.7% | 5.8% |
| Sumner 2019 | 17 | 0.41 | 0.87 | 16 | 1.00 | 0.89 | -0.65 | [-1.36; 0.05] | 2.6% | 4.4% |
| **Common effect model** | **154** | | | **137** | | | **-0.39** | **[-0.63; -0.15]** | **22.6%** | **--** |
| **Random effects model** | | | | | | | **-0.24** | **[-0.62; 0.14]** | **--** | **32.3%** |
| Heterogeneity: $I^2 = 44\%$, $\tau^2 = 0.1414$, $p = 0.09$ | | | | | | | | | | |
| actplac = non-active | | | | | | | | | | |
| Abbar 2022 | 73 | 6.17 | 8.70 | 83 | 14.33 | 10.11 | -0.86 | [-1.19; -0.53] | 11.9% | 6.3% |
| Ahmed 2023 | 18 | 83.89 | 19.55 | 18 | 80.56 | 13.02 | 0.20 | [-0.46; 0.85] | 3.0% | 4.6% |
| Burger 2016 | 3 | 9.40 | 11.75 | 7 | 22.70 | 11.65 | -1.03 | [-2.50; 0.44] | 0.6% | 1.9% |
| Canuso 2018 | 35 | 4.26 | 2.09 | 31 | 4.81 | 1.70 | -0.28 | [-0.77; 0.20] | 5.4% | 5.5% |
| Domany 2020 | 9 | 13.10 | 14.20 | 9 | 12.30 | 14.70 | 0.05 | [-0.87; 0.98] | 1.5% | 3.4% |
| Domany 2021 | 15 | 8.60 | 9.30 | 15 | 13.60 | 8.60 | -0.54 | [-1.27; 0.19] | 2.4% | 4.3% |
| Fu 2020 | 110 | 2.49 | 1.65 | 112 | 2.74 | 1.44 | -0.16 | [-0.42; 0.10] | 18.5% | 6.6% |
| Hu 2016 | 13 | 0.30 | 0.50 | 14 | 1.30 | 0.40 | -2.15 | [-3.13; -1.18] | 1.4% | 3.2% |
| Ionescu 2019 | 13 | 1.90 | 1.40 | 13 | 1.50 | 1.40 | 0.28 | [-0.50; 1.05] | 2.2% | 4.1% |
| Ionescu 2020 | 113 | 2.25 | 1.47 | 112 | 2.92 | 1.32 | -0.48 | [-0.74; -0.21] | 18.3% | 6.6% |
| Jones 2024 | 17 | 8.50 | 7.99 | 11 | 14.20 | 8.63 | -0.67 | [-1.45; 0.11] | 2.1% | 4.0% |
| Singh 2016 2x doses | 18 | 0.11 | 0.32 | 16 | 0.75 | 1.18 | -0.74 | [-1.44; -0.04] | 2.6% | 4.4% |
| Singh 2016 3x doses | 17 | 0.12 | 0.49 | 16 | 0.50 | 1.03 | -0.46 | [-1.16; 0.23] | 2.7% | 4.5% |
| Zarate 2006 | 9 | 0.12 | 0.35 | 9 | 0.56 | 0.73 | -0.72 | [-1.68; 0.24] | 1.4% | 3.3% |
| Zarate 2012 | 19 | 1.10 | 1.24 | 22 | 1.59 | 1.22 | -0.39 | [-1.01; 0.23] | 3.3% | 4.8% |
| **Common effect model** | **482** | | | **488** | | | **-0.44** | **[-0.57; -0.31]** | **77.4%** | **--** |
| **Random effects model** | | | | | | | **-0.48** | **[-0.77; -0.19]** | **--** | **67.7%** |
| Heterogeneity: $I^2 = 57\%$, $\tau^2 = 0.2110$, $p < 0.01$ | | | | | | | | | | |
| **Common effect model** | **636** | | | **625** | | | **-0.43** | **[-0.54; -0.31]** | **100.0%** | **--** |
| **Random effects model** | | | | | | | **-0.40** | **[-0.63; -0.16]** | **--** | **100.0%** |

Heterogeneity: $I^2 = 52\%$, $\tau^2 = 0.2017$, $p < 0.01$

Test for subgroup differences (common effect): $\chi^2_1 = 0.11$, df = 1 ($p = 0.74$)

RE model$\chi^2_1 = 0.99$, df = 1 ($p = 0.32$)

# >12h up to 24h

| Study | Experimental Total | Experimental Mean | Experimental SD | Control Total | Control Mean | Control SD | SMD | 95%-CI | Weight (common) | Weight (random) |
|---|---|---|---|---|---|---|---|---|---|---|
| **actplac = active** | | | | | | | | | | |
| Fineberg 2023 | 10 | 3.10 | 5.63 | 11 | 3.18 | 5.67 | -0.01 | [-0.87; 0.84] | 1.5% | 2.5% |
| Grunebaum 2017 | 7 | 4.29 | 5.74 | 9 | 10.11 | 5.75 | -0.96 | [-2.02; 0.10] | 1.0% | 1.8% |
| Grunebaum 2018 | 40 | 6.05 | 5.48 | 40 | 11.63 | 7.43 | -0.85 | [-1.30; -0.39] | 5.3% | 5.6% |
| Loo 2016 | 9 | 1.78 | 1.20 | 6 | 1.33 | 1.03 | 0.37 | [-0.67; 1.42] | 1.0% | 1.8% |
| Murrough 2013 | 26 | 0.85 | 1.12 | 26 | 1.46 | 1.33 | -0.49 | [-1.04; 0.06] | 3.6% | 4.6% |
| Murrough 2015 II | 12 | 10.83 | 8.48 | 12 | 14.00 | 10.15 | -0.33 | [-1.13; 0.48] | 1.7% | 2.8% |
| Pattanaseri 2024 | 11 | 1.64 | 1.57 | 9 | 2.11 | 1.16 | -0.32 | [-1.21; 0.57] | 1.4% | 2.4% |
| Ray-Griffith 2017 | 8 | 2.87 | 0.99 | 8 | 2.62 | 0.74 | 0.27 | [-0.72; 1.26] | 1.1% | 2.0% |
| Su 2023 | 42 | 1.48 | 1.42 | 42 | 2.10 | 1.21 | -0.47 | [-0.90; -0.03] | 5.9% | 5.9% |
| Sumner 2019 | 17 | 0.65 | 0.79 | 16 | 1.31 | 0.95 | -0.74 | [-1.45; -0.03] | 2.2% | 3.3% |
| Zhou 2024 | 27 | -2.40 | 2.20 | 27 | -1.50 | 2.00 | -0.42 | [-0.96; 0.12] | 3.8% | 4.7% |
| Common effect model | **209** | | | **206** | | | **-0.47** | **[-0.67; -0.28]** | **28.5%** | **--** |
| Random effects model | | | | | | | **-0.43** | **[-0.70; -0.16]** | **--** | **37.5%** |
| Heterogeneity: $I^2 = 0\%$, $\tau^2 = 0.0808$, $p = 0.44$ | | | | | | | | | | |
| **actplac = non-active** | | | | | | | | | | |
| Abbar 2022 | 73 | 5.77 | 9.68 | 83 | 11.97 | 10.41 | -0.61 | [-0.93; -0.29] | 10.7% | 7.4% |
| Canuso 2018 | 35 | 3.29 | 2.47 | 30 | 4.47 | 1.57 | -0.55 | [-1.05; -0.06] | 4.5% | 5.2% |
| Domany 2020 | 9 | 9.20 | 14.80 | 9 | 11.90 | 16.40 | -0.16 | [-1.09; 0.76] | 1.3% | 2.2% |
| Domany 2021 | 15 | 7.80 | 10.50 | 15 | 11.50 | 10.80 | -0.34 | [-1.06; 0.38] | 2.1% | 3.3% |
| Fu 2020 | 110 | 2.23 | 1.67 | 112 | 2.43 | 1.63 | -0.12 | [-0.38; 0.14] | 16.0% | 8.3% |
| Hu 2016 | 13 | 0.70 | 0.70 | 14 | 1.30 | 0.40 | -1.03 | [-1.84; -0.22] | 1.7% | 2.7% |
| Ionescu 2020 | 110 | 2.22 | 1.51 | 110 | 2.44 | 1.57 | -0.14 | [-0.41; 0.12] | 15.8% | 8.3% |
| Jones 2024 | 17 | 10.80 | 8.08 | 10 | 12.30 | 7.83 | -0.18 | [-0.96; 0.60] | 1.8% | 2.9% |
| Lii 2023 | 20 | 0.20 | 0.52 | 19 | 0.21 | 0.54 | -0.02 | [-0.65; 0.61] | 2.8% | 3.9% |
| Price 2022 | 53 | 0.38 | 0.98 | 49 | 0.59 | 1.31 | -0.18 | [-0.57; 0.21] | 7.3% | 6.5% |
| Singh 2016 II | 18 | 0.61 | 1.42 | 8 | 0.88 | 1.64 | -0.18 | [-1.01; 0.66] | 1.6% | 2.6% |
| Sos 2013 | 9 | 0.56 | 0.73 | 18 | 0.39 | 0.59 | 0.26 | [-0.55; 1.06] | 1.7% | 2.8% |
| Zarate 2006 | 9 | 0.44 | 0.53 | 9 | 0.78 | 0.83 | -0.46 | [-1.40; 0.48] | 1.3% | 2.2% |
| Zarate 2012 | 20 | 1.30 | 1.38 | 23 | 2.00 | 1.24 | -0.53 | [-1.14; 0.09] | 3.0% | 4.1% |
| Common effect model | **511** | | | **509** | | | **-0.27** | **[-0.40; -0.15]** | **71.5%** | **--** |
| Random effects model | | | | | | | **-0.30** | **[-0.48; -0.11]** | **--** | **62.5%** |
| Heterogeneity: $I^2 = 11\%$, $\tau^2 = 0.0425$, $p = 0.33$ | | | | | | | | | | |
| **Common effect model** | **720** | | | **715** | | | **-0.33** | **[-0.44; -0.23]** | **100.0%** | **--** |
| **Random effects model** | | | | | | | **-0.35** | **[-0.51; -0.19]** | **--** | **100.0%** |

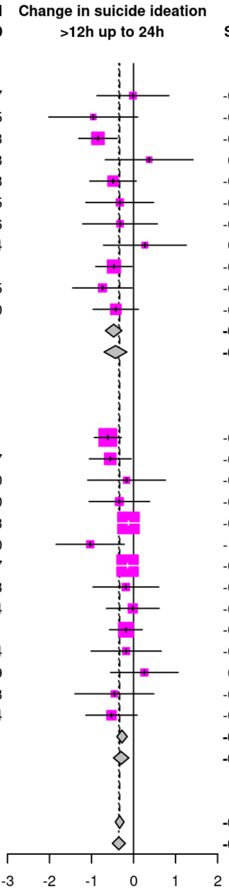

Heterogeneity: $I^2 = 12\%$, $\tau^2 = 0.0573$, $p = 0.29$
Test for subgroup differences (common effect): $\chi^2_1 = 2.82$, df = 1 ($p = 0.09$)
RE model$\chi^2_1 = 0.64$, df = 1 ($p = 0.42$)

## >24h up to 72h

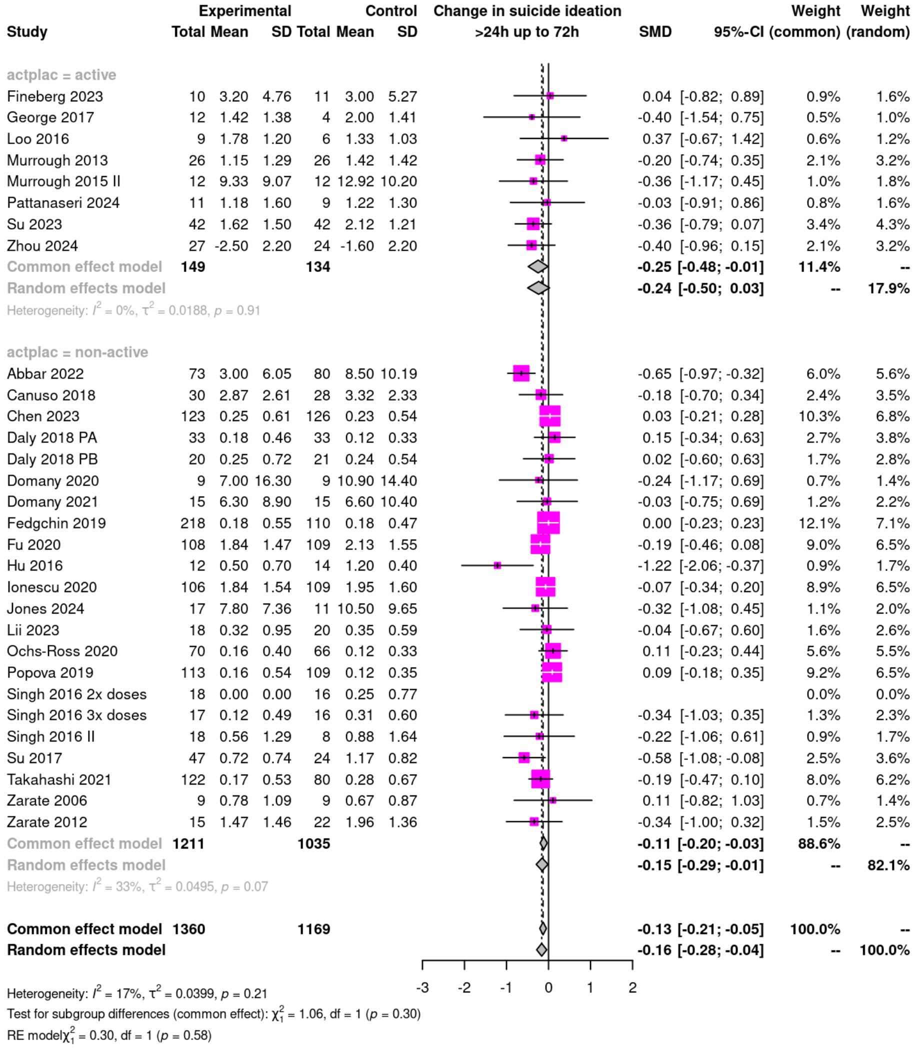
Study
Experimental Total Mean SD
Control Total Mean SD
Change in suicide ideation >24h up to 72h
SMD 95%-CI
Weight (common)
Weight (random)
actplac = active
Fineberg 2023 10 3.20 4.76 11 3.00 5.27 0.04 [-0.82; 0.89] 0.9% 1.6%
George 2017 12 1.42 1.38 4 2.00 1.41 -0.40 [-1.54; 0.75] 0.5% 1.0%
Loo 2016 9 1.78 1.20 6 1.33 1.03 0.37 [-0.67; 1.42] 0.6% 1.2%
Murrough 2013 26 1.15 1.29 26 1.42 1.42 -0.20 [-0.74; 0.35] 2.1% 3.2%
Murrough 2015 II 12 9.33 9.07 12 12.92 10.20 -0.36 [-1.17; 0.45] 1.0% 1.8%
Pattanaseri 2024 11 1.18 1.60 9 1.22 1.30 -0.03 [-0.91; 0.86] 0.8% 1.6%
Su 2023 42 1.62 1.50 42 2.12 1.21 -0.36 [-0.79; 0.07] 3.4% 4.3%
Zhou 2024 27 -2.50 2.20 24 -1.60 2.20 -0.40 [-0.96; 0.15] 2.1% 3.2%
Common effect model 149 134 -0.25 [-0.48; -0.01] 11.4% --
Random effects model -0.24 [-0.50; 0.03] -- 17.9%
Heterogeneity: $I^2$ = 0%, $\tau^2$ = 0.0188, $p$ = 0.91
actplac = non-active
Abbar 2022 73 3.00 6.05 80 8.50 10.19 -0.65 [-0.97; -0.32] 6.0% 5.6%
Canuso 2018 30 2.87 2.61 28 3.32 2.33 -0.18 [-0.70; 0.34] 2.4% 3.5%
Chen 2023 123 0.25 0.61 126 0.23 0.54 0.03 [-0.21; 0.28] 10.3% 6.8%
Daly 2018 PA 33 0.18 0.46 33 0.12 0.33 0.15 [-0.34; 0.63] 2.7% 3.8%
Daly 2018 PB 20 0.25 0.72 21 0.24 0.54 0.02 [-0.60; 0.63] 1.7% 2.8%
Domany 2020 9 7.00 16.30 9 10.90 14.40 -0.24 [-1.17; 0.69] 0.7% 1.4%
Domany 2021 15 6.30 8.90 15 6.60 10.40 -0.03 [-0.75; 0.69] 1.2% 2.2%
Fedgchin 2019 218 0.18 0.55 110 0.18 0.47 0.00 [-0.23; 0.23] 12.1% 7.1%
Fu 2020 108 1.84 1.47 109 2.13 1.55 -0.19 [-0.46; 0.08] 9.0% 6.5%
Hu 2016 12 0.50 0.70 14 1.20 0.40 -1.22 [-2.06; -0.37] 0.9% 1.7%
Ionescu 2020 106 1.84 1.54 109 1.95 1.60 -0.07 [-0.34; 0.20] 8.9% 6.5%
Jones 2024 17 7.80 7.36 11 10.50 9.65 -0.32 [-1.08; 0.45] 1.1% 2.0%
Lii 2023 18 0.32 0.95 20 0.35 0.59 -0.04 [-0.67; 0.60] 1.6% 2.6%
Ochs-Ross 2020 70 0.16 0.40 66 0.12 0.33 0.11 [-0.23; 0.44] 5.6% 5.5%
Popova 2019 113 0.16 0.54 109 0.12 0.35 0.09 [-0.18; 0.35] 9.2% 6.5%
Singh 2016 2x doses 18 0.00 0.00 16 0.25 0.77 0.0% 0.0%
Singh 2016 3x doses 17 0.12 0.49 16 0.31 0.60 -0.34 [-1.03; 0.35] 1.3% 2.3%
Singh 2016 II 18 0.56 1.29 8 0.88 1.64 -0.22 [-1.06; 0.61] 0.9% 1.7%
Su 2017 47 0.72 0.74 24 1.17 0.82 -0.58 [-1.08; -0.08] 2.5% 3.6%
Takahashi 2021 122 0.17 0.53 80 0.28 0.67 -0.19 [-0.47; 0.10] 8.0% 6.2%
Zarate 2006 9 0.78 1.09 9 0.67 0.87 0.11 [-0.82; 1.03] 0.7% 1.4%
Zarate 2012 15 1.47 1.46 22 1.96 1.36 -0.34 [-1.00; 0.32] 1.5% 2.5%
Common effect model 1211 1035 -0.11 [-0.20; -0.03] 88.6% --
Random effects model -0.15 [-0.29; -0.01] -- 82.1%
Heterogeneity: $I^2$ = 33%, $\tau^2$ = 0.0495, $p$ = 0.07
Common effect model 1360 1169 -0.13 [-0.21; -0.05] 100.0% --
Random effects model -0.16 [-0.28; -0.04] -- 100.0%
-3 -2 -1 0 1 2
Heterogeneity: $I^2$ = 17%, $\tau^2$ = 0.0399, $p$ = 0.21
Test for subgroup differences (common effect): $\chi^2_1$ = 1.06, df = 1 ($p$ = 0.30)
RE model$\chi^2_1$ = 0.30, df = 1 ($p$ = 0.58)

## >72h up to 2w

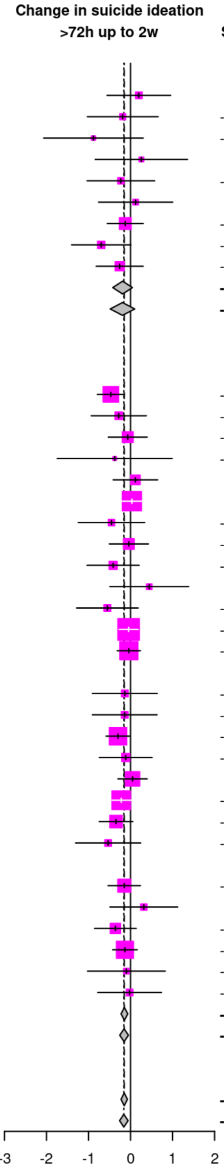

| Study | Experimental Total | Experimental Mean | Experimental SD | Control Total | Control Mean | Control SD | SMD | 95%-CI | Weight (common) | Weight (random) |
|---|---|---|---|---|---|---|---|---|---|---|
| actplac = active | | | | | | | | | | |
| Duek 2023 | 14 | 0.14 | 0.36 | 13 | 0.08 | 0.28 | 0.20 | [-0.56; 0.95] | 1.0% | 1.5% |
| Fineberg 2023 | 10 | 4.30 | 7.10 | 12 | 5.67 | 7.04 | -0.19 | [-1.03; 0.65] | 0.8% | 1.3% |
| George 2017 | 12 | 1.25 | 1.21 | 4 | 2.50 | 1.73 | -0.88 | [-2.07; 0.30] | 0.4% | 0.7% |
| Loo 2016 | 9 | 2.00 | 1.66 | 5 | 1.60 | 0.89 | 0.26 | [-0.84; 1.36] | 0.5% | 0.8% |
| Murrough 2015 II | 12 | 7.92 | 8.73 | 12 | 10.08 | 9.39 | -0.23 | [-1.03; 0.57] | 0.9% | 1.4% |
| Pattanaseri 2024 | 11 | 1.64 | 1.75 | 9 | 1.44 | 1.33 | 0.12 | [-0.76; 1.00] | 0.7% | 1.2% |
| Su 2023 | 42 | 1.95 | 1.36 | 42 | 2.12 | 1.33 | -0.13 | [-0.55; 0.30] | 3.2% | 3.6% |
| Sumner 2019 | 17 | 0.94 | 0.97 | 16 | 1.63 | 0.96 | -0.70 | [-1.40; 0.01] | 1.2% | 1.7% |
| Zhou 2024 | 27 | -2.80 | 2.10 | 23 | -2.20 | 2.50 | -0.26 | [-0.82; 0.30] | 1.9% | 2.5% |
| Common effect model | **154** | | | **136** | | | **-0.19** | **[-0.42; 0.05]** | **10.5%** | **--** |
| Random effects model | | | | | | | **-0.19** | **[-0.48; 0.10]** | **--** | **14.7%** |
| Heterogeneity: $I^2 = 0\%$, $\tau^2 = 0.0542$, $p = 0.69$ | | | | | | | | | | |
| actplac = non-active | | | | | | | | | | |
| Abbar 2022 | 73 | 3.50 | 7.18 | 80 | 8.00 | 11.30 | -0.47 | [-0.79; -0.15] | 5.6% | 5.1% |
| Ahmed 2023 | 18 | 75.72 | 20.58 | 18 | 80.67 | 13.15 | -0.28 | [-0.94; 0.38] | 1.3% | 1.9% |
| Anderson 2017 | 35 | 2.94 | 1.76 | 37 | 3.08 | 2.38 | -0.07 | [-0.53; 0.40] | 2.7% | 3.3% |
| Burger 2016 | 3 | 2.40 | 13.61 | 7 | 6.70 | 8.93 | -0.38 | [-1.75; 0.99] | 0.3% | 0.5% |
| Canuso 2018 | 29 | 1.76 | 2.03 | 26 | 1.54 | 1.68 | 0.12 | [-0.41; 0.65] | 2.1% | 2.7% |
| Chen 2023 | 117 | 0.25 | 0.60 | 111 | 0.23 | 0.61 | 0.03 | [-0.23; 0.29] | 8.6% | 6.2% |
| Colla 2024 | 17 | 1.18 | 0.87 | 10 | 1.60 | 0.95 | -0.45 | [-1.24; 0.34] | 0.9% | 1.4% |
| Daly 2018 PA | 40 | 0.20 | 0.55 | 32 | 0.22 | 0.42 | -0.04 | [-0.50; 0.43] | 2.7% | 3.3% |
| Daly 2018 PB | 20 | 0.15 | 0.49 | 21 | 0.38 | 0.59 | -0.41 | [-1.03; 0.20] | 1.5% | 2.1% |
| Domany 2020 | 9 | 7.40 | 18.00 | 9 | 1.40 | 2.20 | 0.45 | [-0.49; 1.38] | 0.7% | 1.0% |
| Domany 2021 | 15 | 3.10 | 14.10 | 15 | 11.40 | 15.20 | -0.55 | [-1.28; 0.18] | 1.1% | 1.6% |
| Fedgchin 2019 | 211 | 0.16 | 0.46 | 110 | 0.18 | 0.39 | -0.05 | [-0.28; 0.18] | 10.9% | 6.7% |
| Fu 2020 | 105 | 1.28 | 1.31 | 99 | 1.33 | 1.14 | -0.04 | [-0.32; 0.23] | 7.7% | 5.9% |
| Grabski 2022 | 41 | 0.10 | 0.30 | 44 | 0.00 | 0.00 | | | 0.0% | 0.0% |
| Hu 2016 | 12 | 0.90 | 0.90 | 14 | 1.00 | 0.50 | -0.14 | [-0.91; 0.64] | 1.0% | 1.5% |
| Ionescu 2019 | 13 | 1.30 | 1.40 | 13 | 1.50 | 1.40 | -0.14 | [-0.91; 0.63] | 1.0% | 1.5% |
| Ionescu 2020 | 92 | 0.97 | 1.19 | 100 | 1.35 | 1.34 | -0.30 | [-0.58; -0.01] | 7.1% | 5.7% |
| Lii 2023 | 19 | 0.32 | 0.58 | 20 | 0.40 | 0.77 | -0.11 | [-0.74; 0.51] | 1.5% | 2.1% |
| Ochs-Ross 2020 | 65 | 0.21 | 0.65 | 62 | 0.18 | 0.58 | 0.05 | [-0.30; 0.40] | 4.8% | 4.7% |
| Popova 2019 | 109 | 0.08 | 0.36 | 107 | 0.18 | 0.53 | -0.22 | [-0.49; 0.05] | 8.1% | 6.0% |
| Price 2022 | 52 | 0.79 | 1.32 | 46 | 1.30 | 1.62 | -0.34 | [-0.74; 0.05] | 3.6% | 4.0% |
| Singh 2016 2x doses | 15 | 0.07 | 0.26 | 12 | 0.33 | 0.65 | -0.53 | [-1.31; 0.24] | 1.0% | 1.5% |
| Singh 2016 3x doses | 15 | 0.00 | 0.00 | 12 | 0.25 | 0.45 | | | 0.0% | 0.0% |
| Smith-Apeldoorn 2024 | 53 | 1.34 | 0.90 | 51 | 1.47 | 0.83 | -0.15 | [-0.53; 0.24] | 3.9% | 4.2% |
| Sos 2013 | 9 | 0.44 | 0.73 | 18 | 0.17 | 0.87 | 0.32 | [-0.49; 1.12] | 0.9% | 1.4% |
| Su 2017 | 47 | 0.98 | 0.87 | 24 | 1.29 | 0.81 | -0.36 | [-0.86; 0.13] | 2.4% | 3.0% |
| Takahashi 2021 | 116 | 0.18 | 0.61 | 74 | 0.27 | 0.76 | -0.13 | [-0.43; 0.16] | 6.8% | 5.6% |
| Zarate 2006 | 9 | 0.89 | 1.27 | 9 | 1.00 | 0.82 | -0.10 | [-1.02; 0.83] | 0.7% | 1.1% |
| Zarate 2012 | 11 | 2.09 | 1.38 | 17 | 2.12 | 1.05 | -0.02 | [-0.78; 0.74] | 1.0% | 1.5% |
| Common effect model | **1370** | | | **1198** | | | **-0.15** | **[-0.23; -0.07]** | **89.5%** | **--** |
| Random effects model | | | | | | | **-0.15** | **[-0.26; -0.05]** | **--** | **85.3%** |
| Heterogeneity: $I^2 = 0\%$, $\tau^2 = 0.0198$, $p = 0.83$ | | | | | | | | | | |
| **Common effect model** | **1524** | | | **1334** | | | **-0.15** | **[-0.23; -0.07]** | **100.0%** | **--** |
| **Random effects model** | | | | | | | **-0.16** | **[-0.26; -0.06]** | **--** | **100.0%** |

Heterogeneity: $I^2 = 0\%$, $\tau^2 = 0.0259$, $p = 0.90$
Test for subgroup differences (common effect): $\chi^2_1 = 0.12$, df = 1 ($p = 0.73$)
RE model$\chi^2_1 = 0.06$, df = 1 ($p = 0.81$)

## >2w up to 4w

| Study | Experimental Total | Experimental Mean | Experimental SD | Control Total | Control Mean | Control SD | SMD | 95%-CI | Weight (common) | Weight (random) |
|---|---|---|---|---|---|---|---|---|---|---|
| **actplac = active** | | | | | | | | | | |
| Duek 2023 | 13 | 0.31 | 0.48 | 11 | 0.18 | 0.41 | 0.27 | [-0.54; 1.08] | 1.2% | 2.3% |
| Fineberg 2023 | 10 | 7.10 | 6.01 | 11 | 4.73 | 5.75 | 0.39 | [-0.48; 1.25] | 1.0% | 2.1% |
| Zhou 2024 | 26 | -2.90 | 2.20 | 23 | -1.90 | 2.30 | -0.44 | [-1.01; 0.13] | 2.4% | 3.9% |
| Common effect model | **49** | | | **45** | | | **-0.07** | **[-0.48; 0.34]** | **4.6%** | **--** |
| Random effects model | | | | | | | **-0.00** | **[-0.56; 0.56]** | **--** | **8.2%** |
| Heterogeneity: $I^2 = 41\%$, $\tau^2 = 0.1054$, $p = 0.19$ | | | | | | | | | | |
| **actplac = non-active** | | | | | | | | | | |
| Abbar 2022 | 73 | 2.83 | 5.67 | 80 | 6.43 | 9.66 | -0.45 | [-0.77; -0.13] | 7.4% | 7.1% |
| Canuso 2018 | 26 | 1.00 | 1.44 | 21 | 0.90 | 1.37 | 0.07 | [-0.51; 0.64] | 2.3% | 3.8% |
| Chen 2023 | 108 | 0.26 | 0.62 | 107 | 0.21 | 0.53 | 0.09 | [-0.18; 0.35] | 10.7% | 8.1% |
| Domany 2021 | 15 | 3.00 | 14.50 | 15 | 8.30 | 16.00 | -0.34 | [-1.06; 0.38] | 1.5% | 2.8% |
| Fedgchin 2019 | 204 | 0.11 | 0.37 | 107 | 0.17 | 0.40 | -0.16 | [-0.39; 0.08] | 13.9% | 8.7% |
| Fu 2020 | 100 | 0.97 | 1.27 | 93 | 1.08 | 1.21 | -0.09 | [-0.37; 0.19] | 9.6% | 7.8% |
| Grabski 2022 | 39 | 0.05 | 0.22 | 43 | 0.00 | 0.00 | | | 0.0% | 0.0% |
| Hu 2016 | 12 | 0.40 | 0.90 | 14 | 0.20 | 0.40 | 0.29 | [-0.49; 1.06] | 1.3% | 2.5% |
| Ionescu 2019 | 12 | 1.20 | 1.40 | 12 | 1.40 | 1.40 | -0.14 | [-0.94; 0.66] | 1.2% | 2.3% |
| Ionescu 2020 | 90 | 0.68 | 1.04 | 92 | 0.93 | 1.18 | -0.22 | [-0.52; 0.07] | 9.0% | 7.7% |
| Ochs-Ross 2020 | 62 | 0.18 | 0.51 | 61 | 0.17 | 0.45 | 0.02 | [-0.33; 0.37] | 6.1% | 6.6% |
| Ohtani 2024 | 15 | 1.40 | 1.08 | 16 | 1.94 | 1.14 | -0.47 | [-1.19; 0.25] | 1.5% | 2.8% |
| Popova 2019 | 103 | 0.07 | 0.29 | 102 | 0.08 | 0.31 | -0.03 | [-0.31; 0.24] | 10.2% | 8.0% |
| Price 2022 | 49 | 0.61 | 1.27 | 41 | 1.68 | 1.71 | -0.71 | [-1.14; -0.29] | 4.2% | 5.5% |
| Smith-Apeldoorn 2024 | 50 | 1.42 | 0.83 | 49 | 1.39 | 1.02 | 0.03 | [-0.36; 0.43] | 4.9% | 5.9% |
| Su 2017 | 47 | 1.00 | 0.75 | 24 | 1.33 | 1.01 | -0.39 | [-0.88; 0.11] | 3.1% | 4.6% |
| Takahashi 2021 | 111 | 0.15 | 0.65 | 72 | 0.14 | 0.54 | 0.02 | [-0.28; 0.31] | 8.7% | 7.6% |
| Common effect model | **1116** | | | **949** | | | **-0.13** | **[-0.22; -0.04]** | **95.4%** | **--** |
| Random effects model | | | | | | | **-0.14** | **[-0.28; -0.01]** | **--** | **91.8%** |
| Heterogeneity: $I^2 = 27\%$, $\tau^2 = 0.0339$, $p = 0.15$ | | | | | | | | | | |
| **Common effect model** | **1165** | | | **994** | | | **-0.13** | **[-0.21; -0.04]** | **100.0%** | **--** |
| **Random effects model** | | | | | | | **-0.14** | **[-0.27; 0.00]** | **--** | **100.0%** |

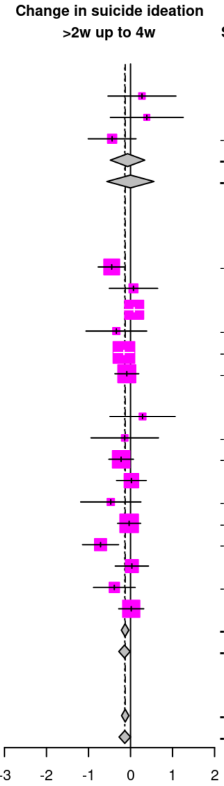

Heterogeneity: $I^2 = 25\%$, $\tau^2 = 0.0414$, $p = 0.16$

Test for subgroup differences (common effect): $\chi^2_1 = 0.07$, df = 1 ($p = 0.78$)

RE model$\chi^2_1 = 0.23$, df = 1 ($p = 0.63$)

## >4w

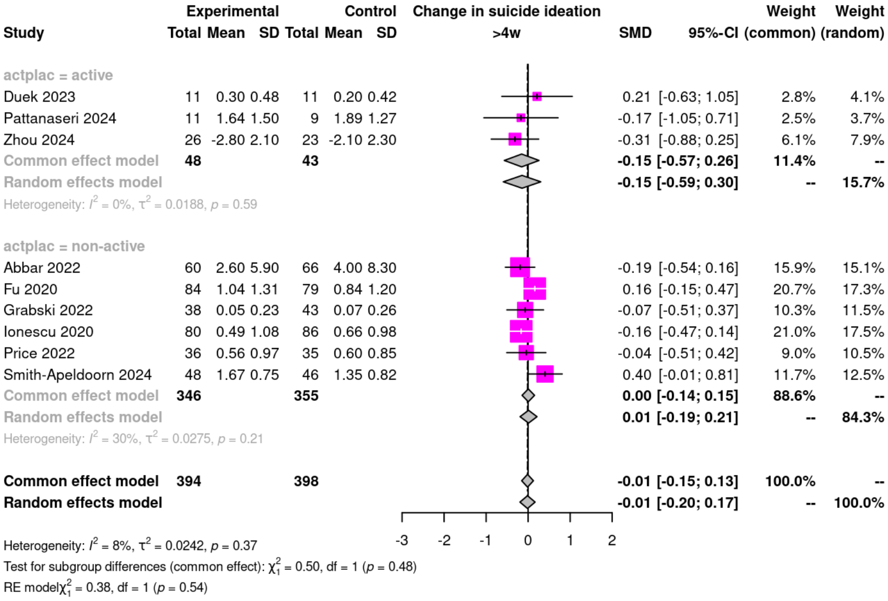
Experimental
Control
Change in suicide ideation
Weight
Weight
Study
Total Mean SD Total Mean SD
>4w
SMD
95%-CI (common) (random)
actplac = active
Duek 2023 11 0.30 0.48 11 0.20 0.42 0.21 [-0.63; 1.05] 2.8% 4.1%
Pattanaseri 2024 11 1.64 1.50 9 1.89 1.27 -0.17 [-1.05; 0.71] 2.5% 3.7%
Zhou 2024 26 -2.80 2.10 23 -2.10 2.30 -0.31 [-0.88; 0.25] 6.1% 7.9%
Common effect model 48 43 -0.15 [-0.57; 0.26] 11.4% --
Random effects model -0.15 [-0.59; 0.30] -- 15.7%
Heterogeneity: $I^2$ = 0%, $\tau^2$ = 0.0188, $p$ = 0.59
actplac = non-active
Abbar 2022 60 2.60 5.90 66 4.00 8.30 -0.19 [-0.54; 0.16] 15.9% 15.1%
Fu 2020 84 1.04 1.31 79 0.84 1.20 0.16 [-0.15; 0.47] 20.7% 17.3%
Grabski 2022 38 0.05 0.23 43 0.07 0.26 -0.07 [-0.51; 0.37] 10.3% 11.5%
Ionescu 2020 80 0.49 1.08 86 0.66 0.98 -0.16 [-0.47; 0.14] 21.0% 17.5%
Price 2022 36 0.56 0.97 35 0.60 0.85 -0.04 [-0.51; 0.42] 9.0% 10.5%
Smith-Apeldoorn 2024 48 1.67 0.75 46 1.35 0.82 0.40 [-0.01; 0.81] 11.7% 12.5%
Common effect model 346 355 0.00 [-0.14; 0.15] 88.6% --
Random effects model 0.01 [-0.19; 0.21] -- 84.3%
Heterogeneity: $I^2$ = 30%, $\tau^2$ = 0.0275, $p$ = 0.21
Common effect model 394 398 -0.01 [-0.15; 0.13] 100.0% --
Random effects model -0.01 [-0.20; 0.17] -- 100.0%
-3 -2 -1 0 1 2
Heterogeneity: $I^2$ = 8%, $\tau^2$ = 0.0242, $p$ = 0.37
Test for subgroup differences (common effect): $\chi^2_1$ = 0.50, df = 1 ($p$ = 0.48)
RE model$\chi^2_1$ = 0.38, df = 1 ($p$ = 0.54)

#### End of Treatment

| Study | Experimental Total | Experimental Mean | Experimental SD | Control Total | Control Mean | Control SD | SMD | 95%-CI | Weight (common) | Weight (random) |
|---|---|---|---|---|---|---|---|---|---|---|
| **actplac = active** | | | | | | | | | | |
| Fineberg 2023 (24h) | 10 | 3.10 | 5.63 | 11 | 3.18 | 5.67 | -0.01 | [-0.87; 0.84] | 0.8% | 1.4% |
| George 2017 (1w) | 12 | 1.25 | 1.21 | 4 | 2.50 | 1.73 | -0.88 | [-2.07; 0.30] | 0.4% | 0.8% |
| Grunebaum 2017 (24h) | 7 | 4.29 | 5.74 | 9 | 10.11 | 5.75 | -0.96 | [-2.02; 0.10] | 0.5% | 1.0% |
| Grunebaum 2018 (24h) | 40 | 6.05 | 5.48 | 40 | 11.63 | 7.43 | -0.85 | [-1.30; -0.39] | 2.7% | 3.3% |
| Loo 2016 (1w-2w) | 9 | 2.00 | 1.66 | 5 | 1.60 | 0.89 | 0.26 | [-0.84; 1.36] | 0.5% | 0.9% |
| Murrough 2013 (24h) | 26 | 0.85 | 1.12 | 26 | 1.46 | 1.33 | -0.49 | [-1.04; 0.06] | 1.9% | 2.6% |
| Murrough 2015 II (24h) | 12 | 10.83 | 8.48 | 12 | 14.00 | 10.15 | -0.33 | [-1.13; 0.48] | 0.9% | 1.5% |
| Pattanaseri 2024 (3d) | 11 | 1.18 | 1.60 | 9 | 1.22 | 1.30 | -0.03 | [-0.91; 0.86] | 0.7% | 1.3% |
| Su 2023 (24h) | 42 | 1.48 | 1.42 | 42 | 2.10 | 1.21 | -0.47 | [-0.90; -0.03] | 3.0% | 3.4% |
| Sumner 2019 (24h) | 17 | 0.65 | 0.79 | 16 | 1.31 | 0.95 | -0.74 | [-1.45; -0.03] | 1.1% | 1.9% |
| Zhou 2024 (6d) | 27 | -2.60 | 2.00 | 25 | -1.70 | 2.20 | -0.42 | [-0.97; 0.13] | 1.9% | 2.6% |
| **Common effect model** | **213** | | | **199** | | | **-0.51** | **[-0.71; -0.31]** | **14.4%** | **--** |
| **Random effects model** | | | | | | | **-0.48** | **[-0.74; -0.22]** | **--** | **20.8%** |
| Heterogeneity: $I^2$ = 0%, $\tau^2$ = 0.0615, $p$ = 0.61 | | | | | | | | | | |
| **actplac = non-active** | | | | | | | | | | |
| Abbar 2022 (day 2) | 73 | 3.00 | 6.05 | 80 | 8.50 | 10.19 | -0.65 | [-0.97; -0.32] | 5.4% | 4.4% |
| Ahmed 2023 (2w) | 18 | 75.72 | 20.58 | 18 | 80.67 | 13.15 | -0.28 | [-0.94; 0.38] | 1.3% | 2.1% |
| Canuso 2018 (25d) | 26 | 1.00 | 1.44 | 21 | 0.90 | 1.37 | 0.07 | [-0.51; 0.64] | 1.7% | 2.5% |
| Chen 2023 (4w) | 108 | 0.26 | 0.62 | 107 | 0.21 | 0.53 | 0.09 | [-0.18; 0.35] | 8.0% | 5.0% |
| Colla 2024 (2w) | 17 | 1.18 | 0.87 | 10 | 1.60 | 0.95 | -0.45 | [-1.24; 0.34] | 0.9% | 1.6% |
| Daly 2018 PA (7d) | 40 | 0.20 | 0.55 | 32 | 0.22 | 0.42 | -0.04 | [-0.50; 0.43] | 2.6% | 3.2% |
| Daly 2018 PB (7d) | 20 | 0.15 | 0.49 | 21 | 0.38 | 0.59 | -0.41 | [-1.03; 0.20] | 1.5% | 2.3% |
| Domany 2020 (24h) | 9 | 9.20 | 14.80 | 9 | 11.90 | 16.40 | -0.16 | [-1.09; 0.76] | 0.7% | 1.2% |
| Domany 2021 (24h) | 15 | 7.80 | 10.50 | 15 | 11.50 | 10.80 | -0.34 | [-1.06; 0.38] | 1.1% | 1.8% |
| Fedgchin 2019 (4w) | 204 | 0.11 | 0.37 | 107 | 0.17 | 0.40 | -0.16 | [-0.39; 0.08] | 10.4% | 5.3% |
| Fu 2020 (25d) | 100 | 0.97 | 1.27 | 93 | 1.08 | 1.21 | -0.09 | [-0.37; 0.19] | 7.1% | 4.8% |
| Grabski 2022 (1-3w) | 38 | 0.05 | 0.23 | 43 | 0.07 | 0.26 | -0.07 | [-0.51; 0.37] | 3.0% | 3.4% |
| Hu 2016 (24h) | 13 | 0.70 | 0.70 | 14 | 1.30 | 0.40 | -1.03 | [-1.84; -0.22] | 0.9% | 1.5% |
| Ionescu 2019 (3w) | 12 | 1.20 | 1.40 | 12 | 1.40 | 1.40 | -0.14 | [-0.94; 0.66] | 0.9% | 1.6% |
| Ionescu 2020 (25d) | 90 | 0.68 | 1.04 | 92 | 0.93 | 1.18 | -0.22 | [-0.52; 0.07] | 6.7% | 4.8% |
| Jones 2024 (24h) | 17 | 10.80 | 8.08 | 10 | 12.30 | 7.83 | -0.18 | [-0.96; 0.60] | 0.9% | 1.6% |
| Lii 2023 (24h) | 20 | 0.20 | 0.52 | 19 | 0.21 | 0.54 | -0.02 | [-0.65; 0.61] | 1.4% | 2.2% |
| Ochs-Ross 2020 (4w) | 62 | 0.18 | 0.51 | 61 | 0.17 | 0.45 | 0.02 | [-0.33; 0.37] | 4.6% | 4.1% |
| Ohtani 2024 (2w) | 15 | 1.40 | 1.08 | 16 | 1.94 | 1.14 | -0.47 | [-1.19; 0.25] | 1.1% | 1.8% |
| Popova 2019 (4w) | 103 | 0.07 | 0.29 | 102 | 0.08 | 0.31 | -0.03 | [-0.31; 0.24] | 7.6% | 4.9% |
| Price 2022 (24h) | 53 | 0.38 | 0.98 | 49 | 0.59 | 1.31 | -0.18 | [-0.57; 0.21] | 3.8% | 3.8% |
| Singh 2016 2x doses (2w) | 15 | 0.07 | 0.26 | 12 | 0.33 | 0.65 | -0.53 | [-1.31; 0.24] | 1.0% | 1.6% |
| Singh 2016 3x doses (2w) | 15 | 0.00 | 0.00 | 12 | 0.25 | 0.45 | | | 0.0% | 0.0% |
| Singh 2016 II (4d) | 18 | 0.56 | 1.29 | 8 | 0.88 | 1.64 | -0.22 | [-1.06; 0.61] | 0.8% | 1.5% |
| Smith-Apeldoorn 2024 (6w) | 50 | 1.60 | 0.83 | 48 | 1.42 | 0.85 | 0.21 | [-0.18; 0.61] | 3.6% | 3.7% |
| Sos 2013 (24h) | 9 | 0.56 | 0.73 | . | 0.39 | 0.59 | | | 0.0% | 0.0% |
| Takahashi 2021 (4w) | 111 | 0.15 | 0.65 | 72 | 0.14 | 0.54 | 0.02 | [-0.28; 0.31] | 6.5% | 4.7% |
| Zarate 2006 (24h) | 9 | 0.44 | 0.53 | 9 | 0.78 | 0.83 | -0.46 | [-1.40; 0.48] | 0.6% | 1.2% |
| Zarate 2012 (24h) | 20 | 1.30 | 1.38 | 23 | 2.00 | 1.24 | -0.53 | [-1.14; 0.09] | 1.5% | 2.3% |
| **Common effect model** | **1300** | | | **1115** | | | **-0.14** | **[-0.22; -0.06]** | **85.6%** | **--** |
| **Random effects model** | | | | | | | **-0.17** | **[-0.28; -0.05]** | **--** | **79.2%** |
| Heterogeneity: $I^2$ = 11%, $\tau^2$ = 0.0328, $p$ = 0.30 | | | | | | | | | | |
| **Common effect model** | **1513** | | | **1314** | | | **-0.19** | **[-0.27; -0.12]** | **100.0%** | **--** |
| **Random effects model** | | | | | | | **-0.24** | **[-0.35; -0.13]** | **--** | **100.0%** |

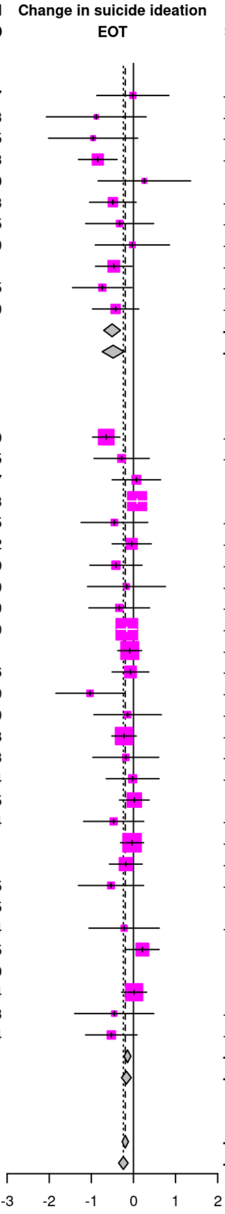

Heterogeneity: $I^2$ = 24%, $\tau^2$ = 0.0484, $p$ = 0.10

Test for subgroup differences (common effect): $\chi^2_1$ = 11.09, df = 1 ($p$ < 0.01)

RE model$\chi^2_1$ = 4.71, df = 1 ($p$ = 0.03)

##### Subgroup Analysis - Only studies with adequate blinding

This subgroup analysis only included studies where ketamine/esketamine were administered alongside an anesthetic procedure so that blinding was guaranteed. This was also tested in two of the studies (Lii 2023 and Anderson et al. 2017) but was also assumed for Ray-Griffith 2017.

**Up to 4h** No study results available

**>12h up to 24h**

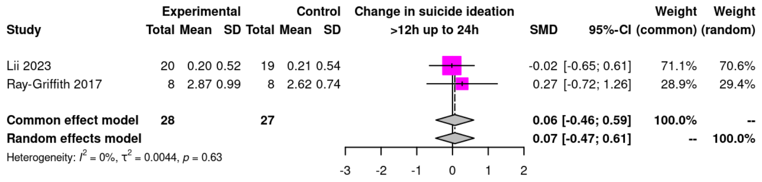

## >24h up to 72h

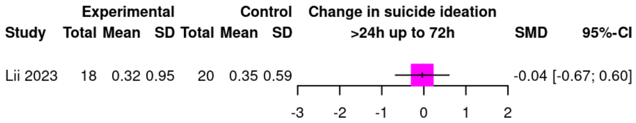
Experimental
Control
Change in suicide ideation
Study
Total
Mean
SD
Total
Mean
SD
>24h up to 72h
SMD
95%-CI
Lii 2023
18
0.32
0.95
20
0.35
0.59
-0.04
[-0.67; 0.60]
-3
-2
-1
0
1
2

## >72h up to 2w

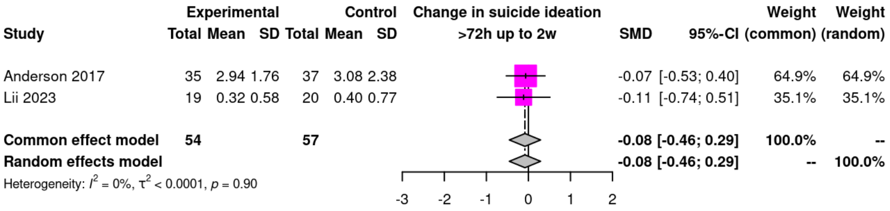

**>2w up to 4w** No study results available

**>4w** No study results available

#### End of Treatment

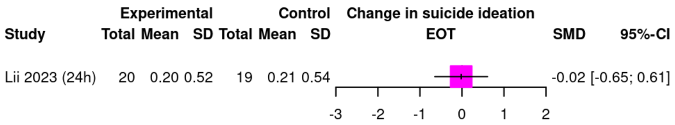

#### Funnel Plots, Suicide Ideation Meta-Analyses

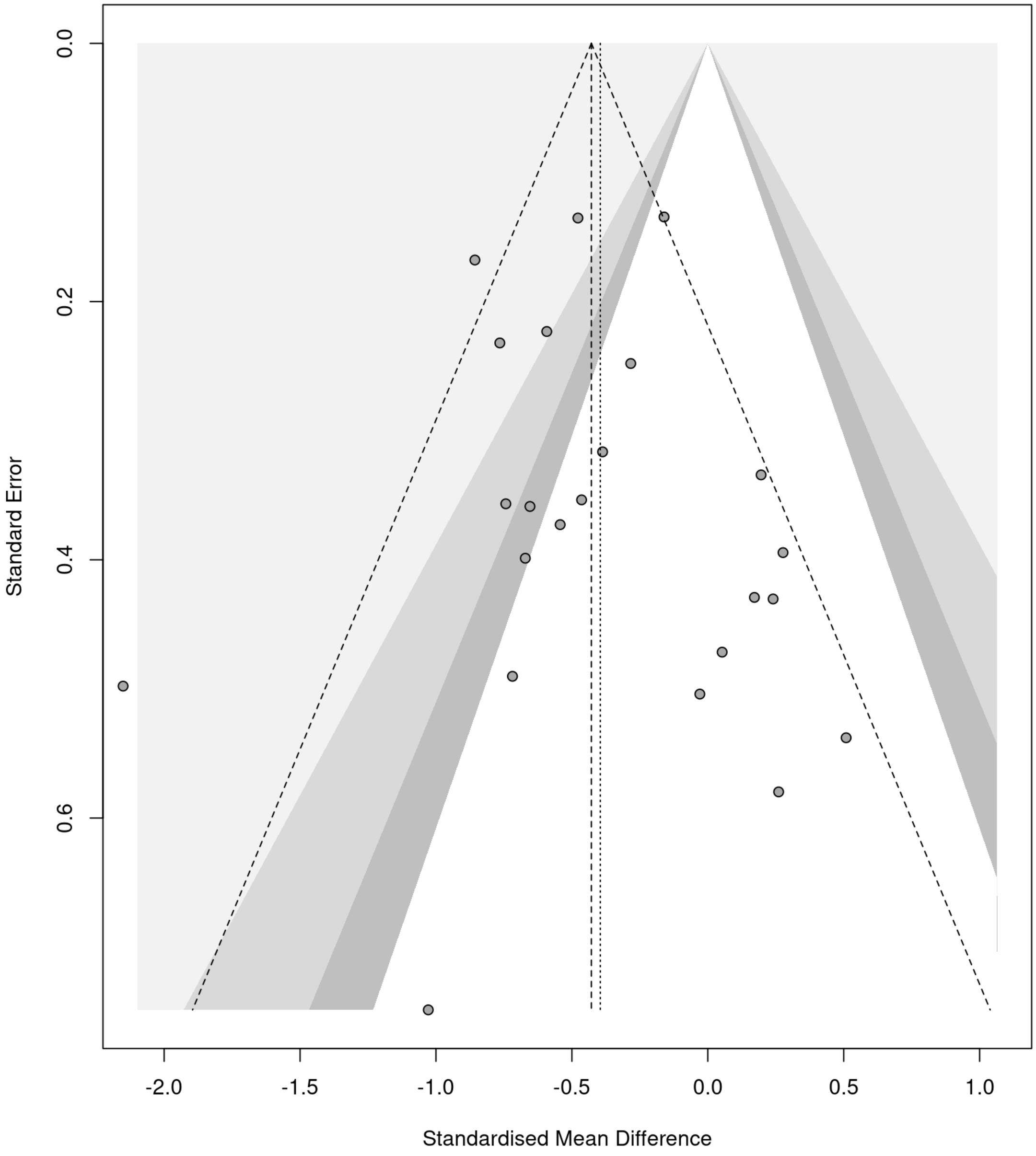

Figure S3: Funnel Plot, Up to 4h

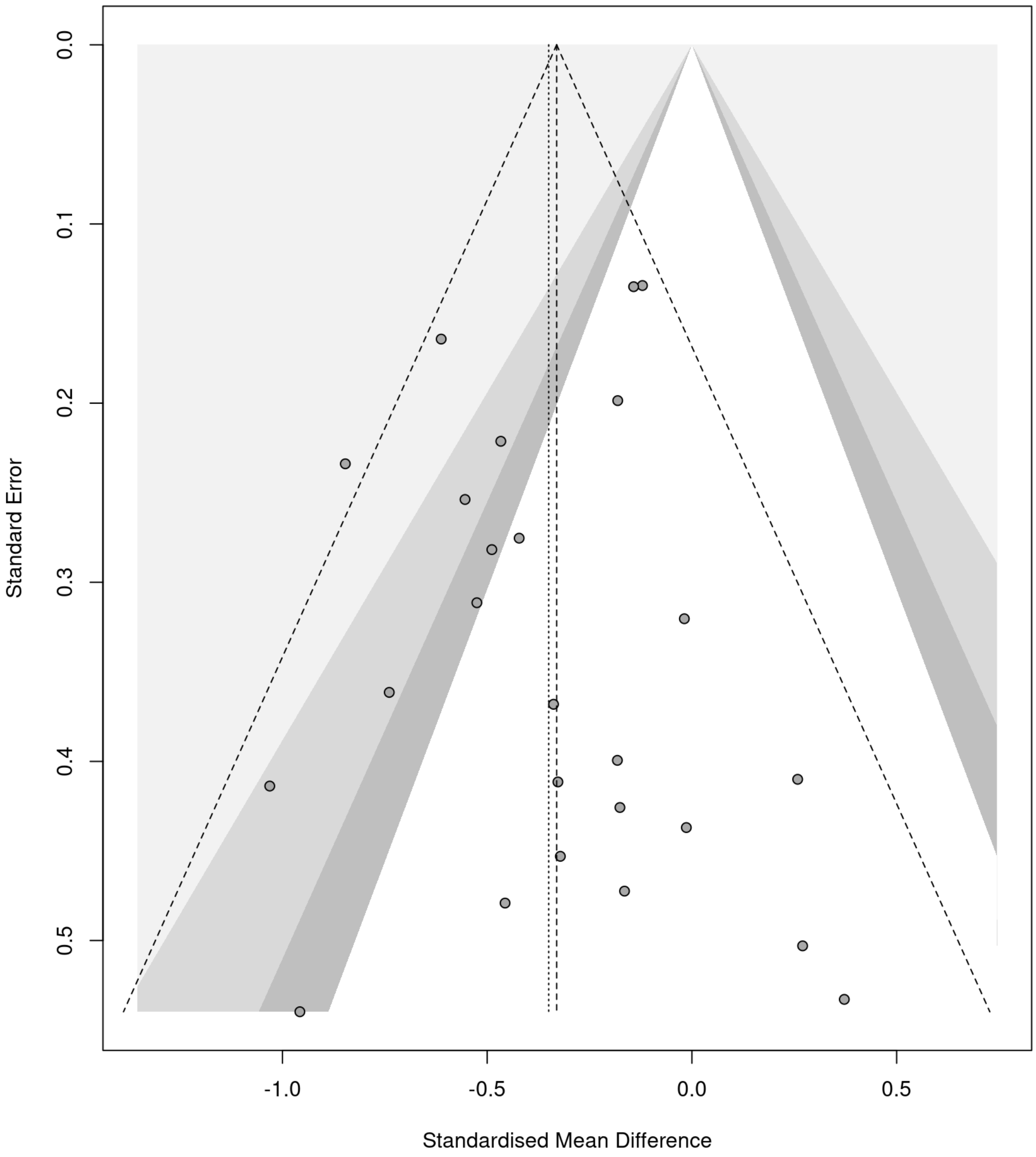

Figure S4: Funnel Plot, >12h up to 24h

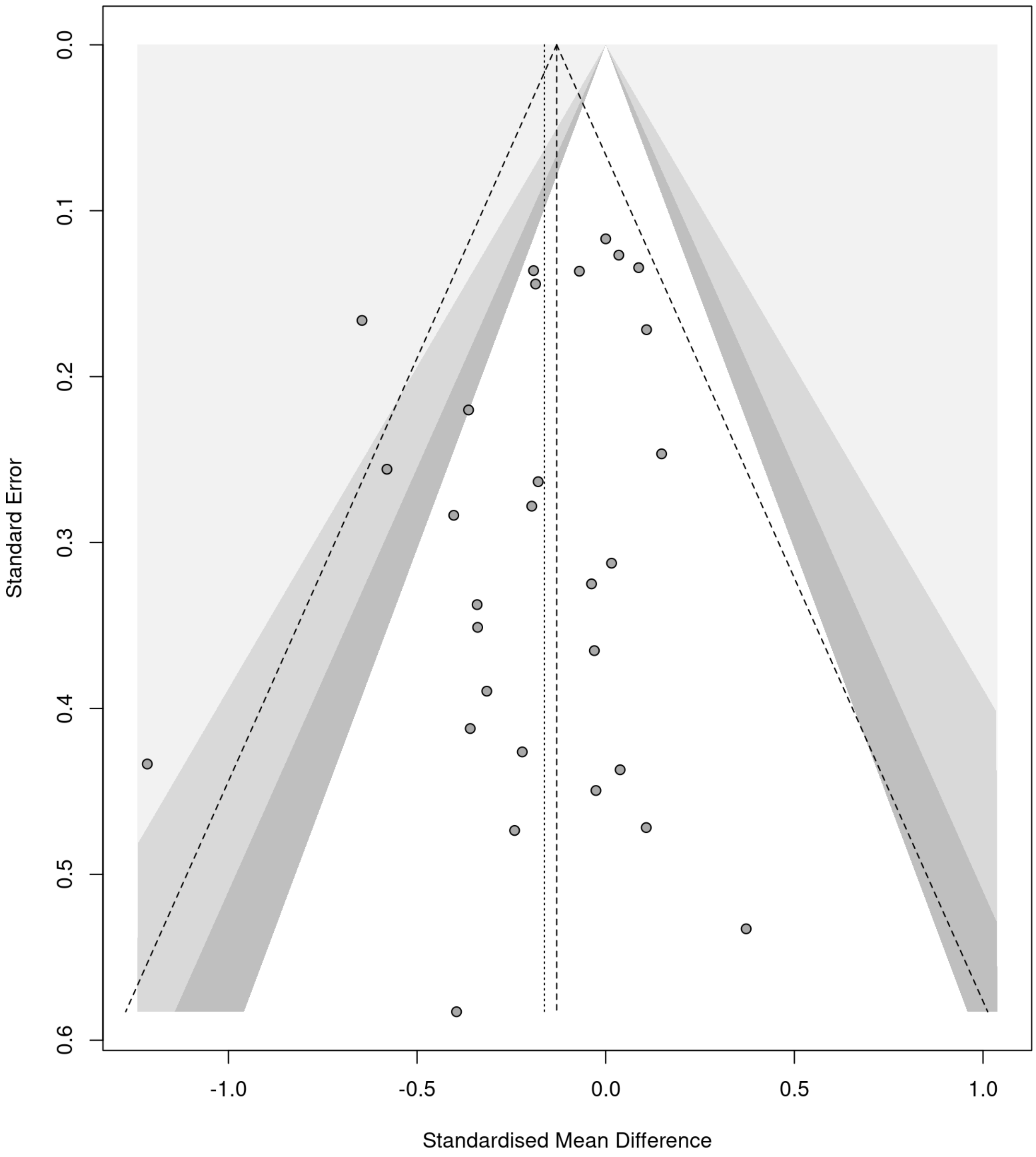

Figure S5: Funnel Plot, >24h up to 72h

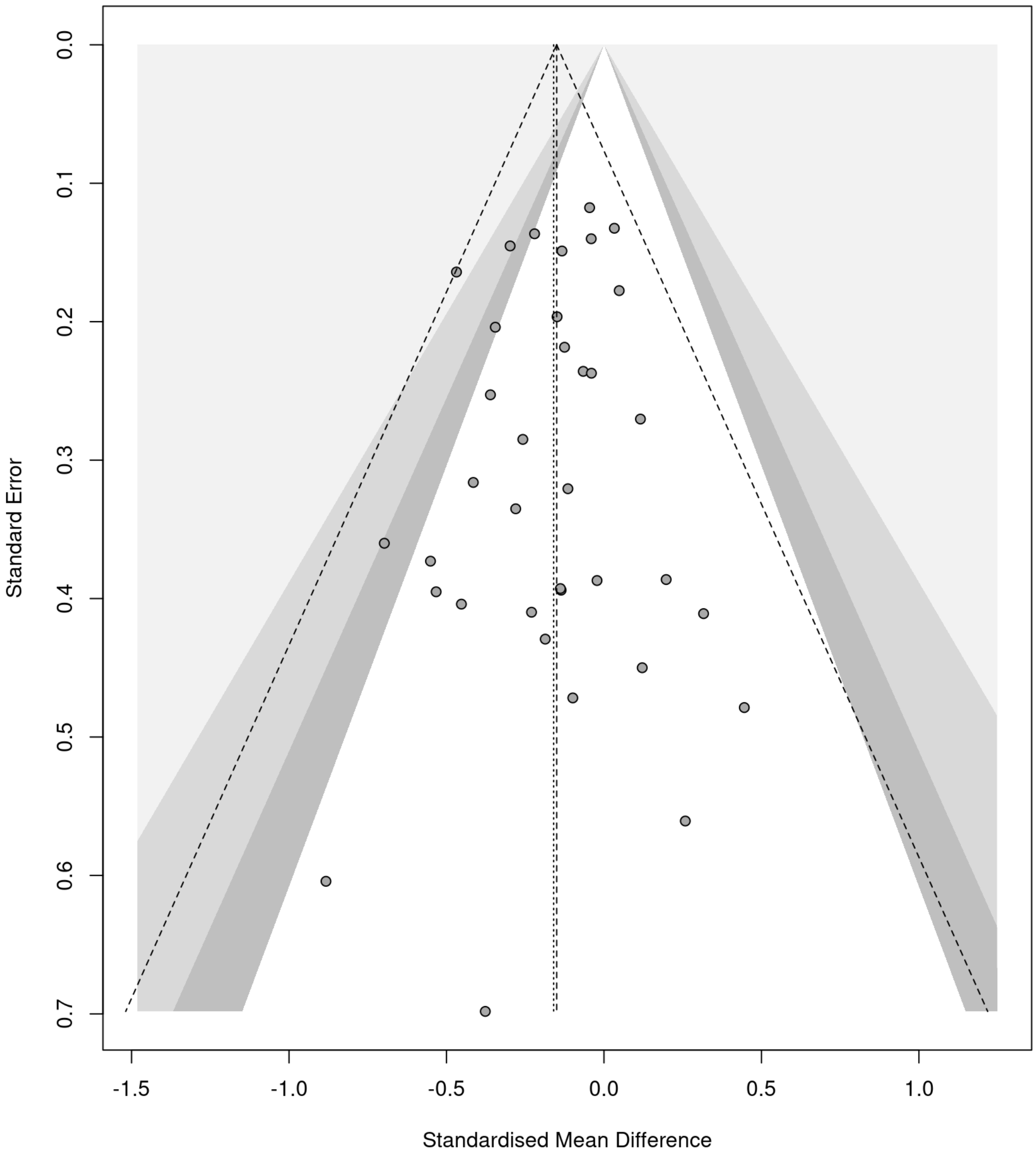

Figure S6: Funnel Plot, >72h up to 2w

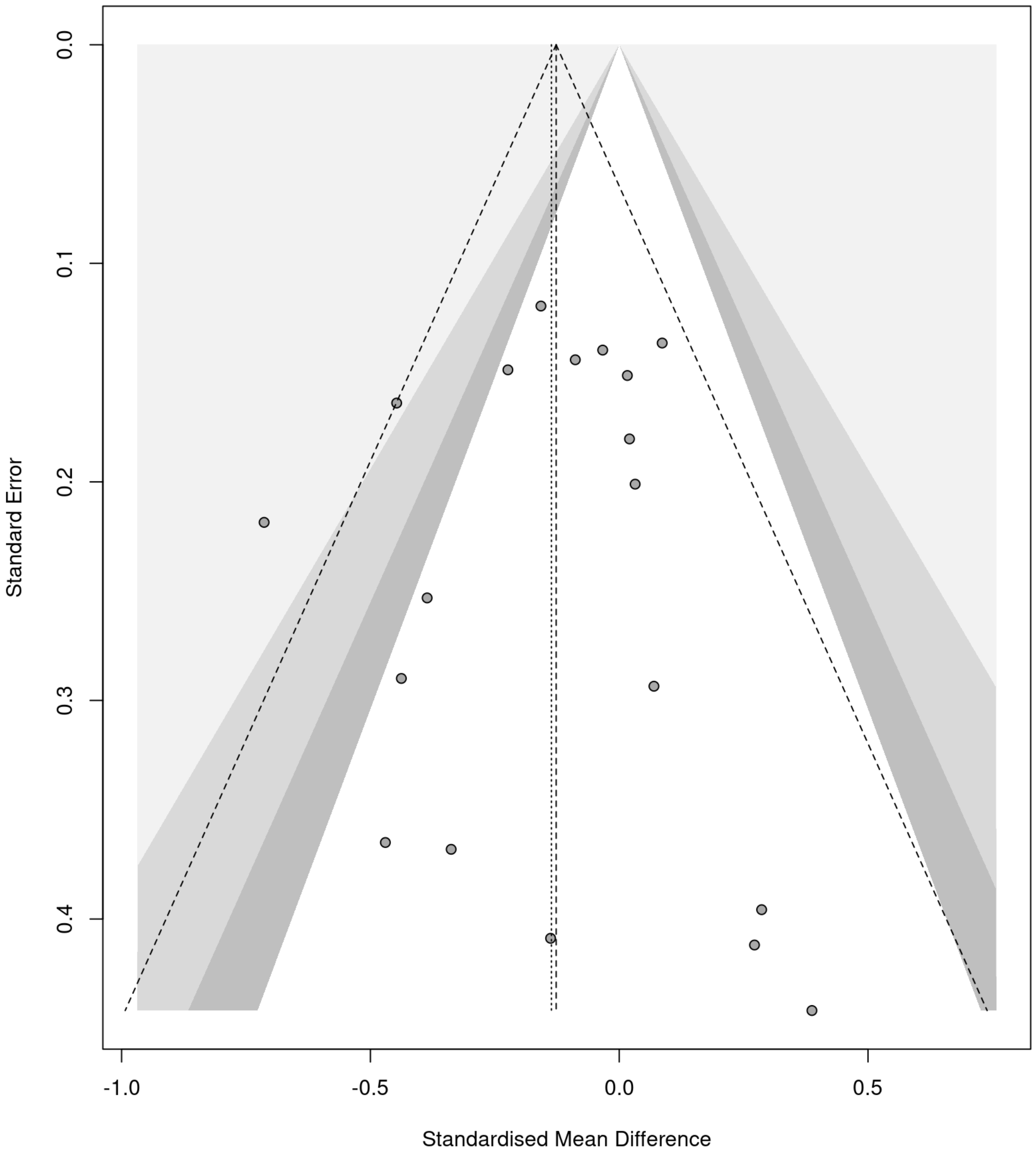

Figure S7: Funnel Plot, >2w up to 4w

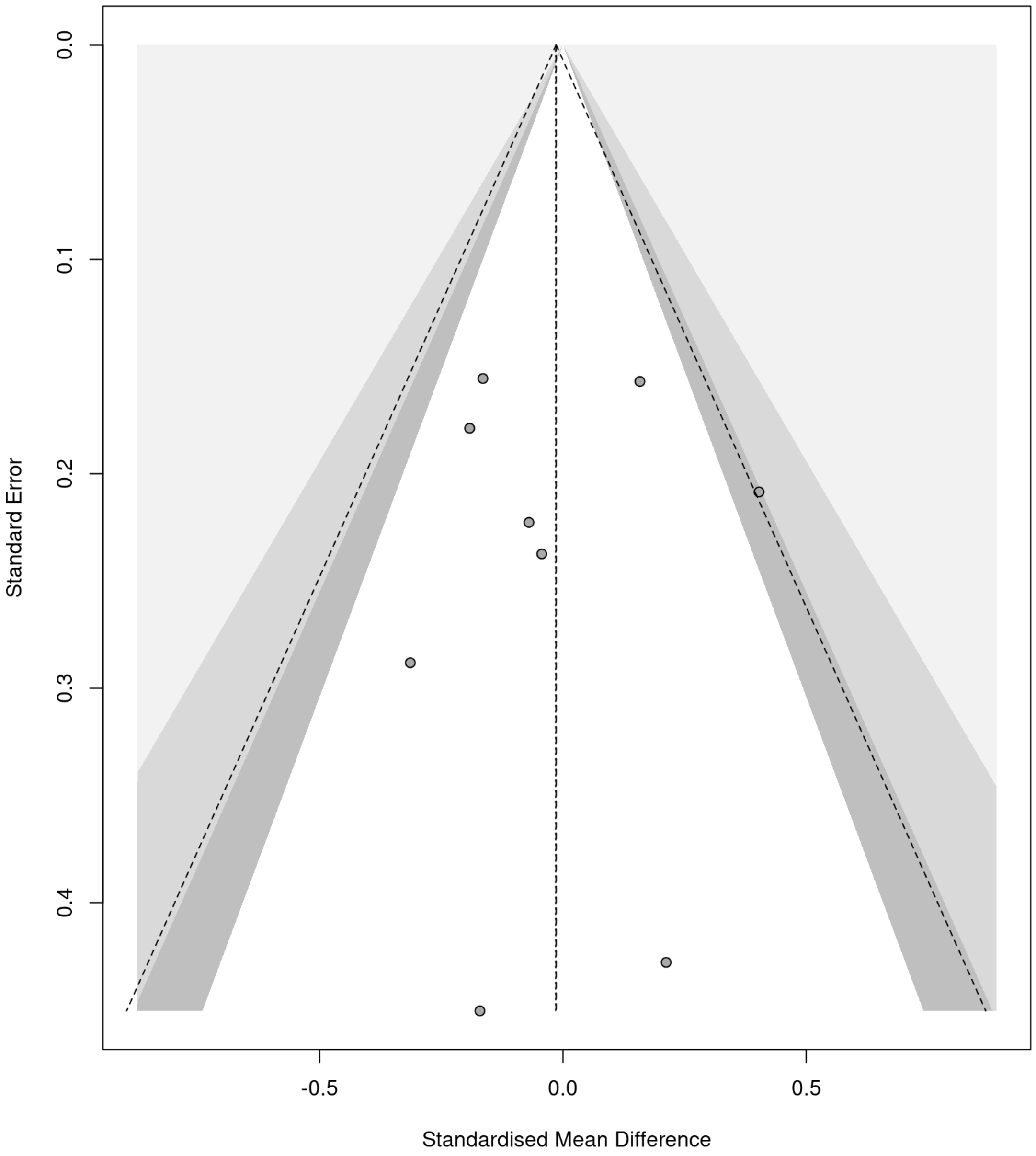

Figure S8: Funnel Plot, >4w

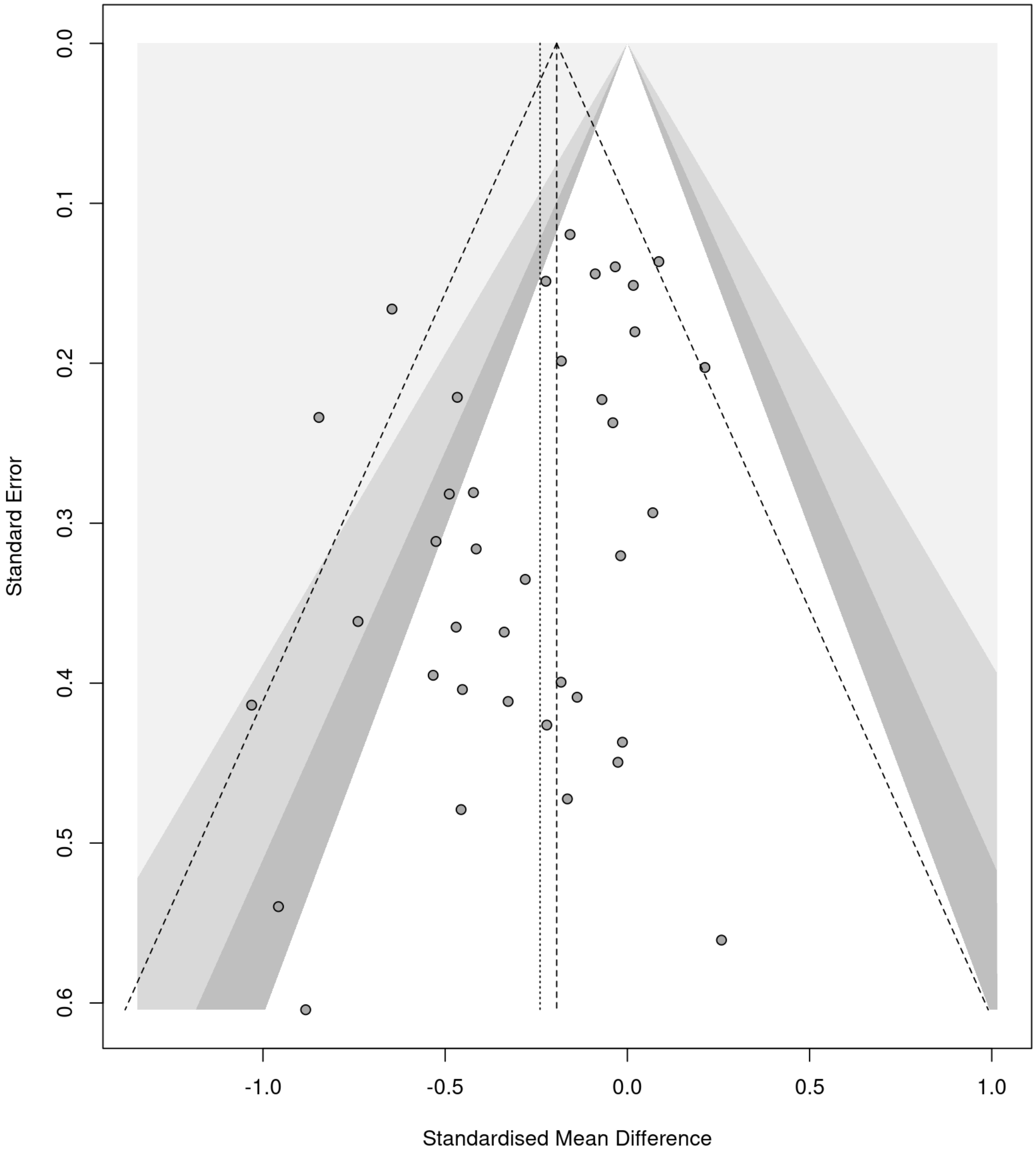

Figure S9: Funnel Plot, EOT
